# Efficacy of the NVX-CoV2373 Covid-19 Vaccine Against the B.1.1.7 Variant

**DOI:** 10.1101/2021.05.13.21256639

**Authors:** Paul T. Heath, Eva P. Galiza, David Neil Baxter, Marta Boffito, Duncan Browne, Fiona Burns, David R. Chadwick, Rebecca Clark, Catherine Cosgrove, James Galloway, Anna L. Goodman, Amardeep Heer, Andrew Higham, Shalini Iyengar, Arham Jamal, Christopher Jeanes, Philip A. Kalra, Christina Kyriakidou, Daniel F. McAuley, Agnieszka Meyrick, Angela M. Minassian, Jane Minton, Patrick Moore, Imrozia Munsoor, Helen Nicholls, Orod Osanlou, Jonathan Packham, Carol H. Pretswell, Alberto San Francisco Ramos, Dinesh Saralaya, Ray P. Sheridan, Richard Smith, Roy L. Soiza, Pauline A. Swift, Emma C. Thomson, Jeremy Turner, Marianne Elizabeth Viljoen, Gary Albert, Iksung Cho, Filip Dubovsky, Greg Glenn, Joy Rivers, Andreana Robertson, Kathy Smith, Seth Toback

**Author notes:** **Corresponding author:** Seth Toback, MD, Novavax, Inc., 21 Firstfield Rd., Gaithersburg, MD, 20878.

## Abstract

**Background:** Covid-19 vaccines are urgently needed, especially against emerging variants. NVX-CoV2373 is a recombinant severe acute respiratory syndrome coronavirus 2 (SARS-CoV-2 rS) nanoparticle vaccine containing trimeric full-length SARS-CoV-2 spike glycoprotein and Matrix-M adjuvant.

**Methods:** A phase 3, randomized, observer-blinded, placebo-controlled trial was conducted in adults 18-84 years old who received two intramuscular 5-µg doses, 21 days apart, of NVX-CoV2373 or placebo (1:1) across 33 sites in the United Kingdom. The primary efficacy endpoint was virologically confirmed symptomatic Covid-19 with onset 7 days after second vaccination in serologically negative participants.

**Results:** A total of 15,187 participants were randomized, of whom 7569 received NVX-CoV2373 and 7570 received placebo; 27.2% were 65 years or older, 44.7% had comorbidities and 4.2% had baseline serological evidence of SARS-CoV-2. There were 10 cases of Covid-19 among NVX-CoV2373 recipients and 96 cases among placebo recipients, with symptom onset at least 7 days after second vaccination; NVX-CoV2373 was 89.7% (95% confidence interval, 80.2 to 94.6) effective in preventing Covid-19, with no hospitalizations or deaths reported. There were five cases of severe Covid-19, all in the placebo group. Post hoc analysis revealed efficacies of 96.4% (73.8 to 99.5) and 86.3% (71.3 to 93.5) against the prototype strain and B.1.1.7 variant, respectively. Vaccine efficacy was similar across subgroups, including participants with comorbidities and those ≥65 years old. Reactogenicity was generally mild and transient. The incidence of serious adverse events was low and similar in the two groups.

**Conclusion:** A two-dose regimen of NVX-CoV2373 conferred 89.7% protection against a blend of prototype and variant Covid-19, demonstrated high efficacy against the B.1.1.7 variant, and had a reassuring safety profile.

(Funded by Novavax, Inc. EudraCT number, 2020-004123-16).

## INTRODUCTION

More than a year after its emergence as a global pandemic, coronavirus disease 2019 (Covid-19) caused by the severe acute respiratory syndrome coronavirus 2 (SARS-CoV-2) infection remains a devastating disease with more than 140 million cases and over 3 million deaths, as of April 19, 2021.^1^ To control this pandemic, development efforts were accelerated to find safe and effective vaccines against SARS-CoV-2 by targeting the spike glycoprotein of the prototype strain.^2^ By the beginning of 2021, several vaccine candidates have emerged as safe and effective in preventing Covid-19, supporting their emergency use around the world.^3-6^

Recent reports from the United Kingdom (UK), Brazil, and South Africa in late 2020 have confirmed the emergence of SARS-CoV2 variants (B.1.1.7, P.1 and B.1.351, respectively) that have been associated with increased transmission, more severe disease, and varying degrees of immune avoidance to Covid-19 vaccines.^7-9^ In the UK, the B.1.1.7 variant was identified from genomic sequencing of samples from patients with Covid-19 in the southeast of England in early October 2020. During December 2020, this new variant spread from the southeast to London and the rest of the UK. Subsequently, the prevalence of the B.1.1.7 variant has been increasing rapidly in Europe and the United States.^7^ These new variants may threaten our current attempts at controlling Covid-19, resulting in further health and socioeconomic devastation.

Early clinical data from the SARS-CoV-2 recombinant nanoparticle spike protein vaccine with Matrix-M adjuvant have shown that a two-dose regimen of 5 µg SARS-CoV-2 rS with 50 µg Matrix-M adjuvant (hereafter referred to as NVX-CoV2373), administered 21 days apart, was well tolerated and associated with robust immune responses in healthy adult participants 18 to 84 years of age.^10,11^ Here, we present efficacy and safety data from a phase 3 trial evaluating the efficacy, immunogenicity, and safety of NVX-CoV2373, in preventing Covid-19 in participants 18 to 84 years old in the UK. This trial was initiated in September 2020 at a time when the B.1.1.7 variant was starting to circulate more widely.

## METHODS

### Trial Design and Participants

We assessed the safety and efficacy of two 5-µg doses of NVX-CoV2373 or placebo, administered intramuscularly 21 days apart. This phase 3 trial was conducted at 33 sites in the UK. Eligible participants were men and non-pregnant women 18 to 84 years old (inclusive) who were healthy or had stable chronic medical conditions, including but not limited to human immunodeficiency virus (and receiving highly active antiretroviral therapy) and cardiac and respiratory diseases. Key exclusion criteria included a history of documented Covid-19, treatment with immunosuppressive therapy, or diagnosis with an immunodeficient condition. (Details of the trial design, conduct, oversight, and analyses are provided in the protocol, statistical analysis plan, and supplemental appendix will be available with the full text of this article upon publication).

All participants provided written informed consent before enrollment in the trial. The trial was designed and funded by Novavax, Inc. (Gaithersburg, MD, USA). The trial protocol was approved by the North West—Greater Manchester Central Research Ethics Committee (Ref 20/NW/03/99) and was performed in accordance with the International Council for Harmonisation Good Clinical Practice guidelines (see protocol in the Supplementary Appendix).

Safety oversight for specific vaccination pause rules was performed by an independent safety monitoring committee (SMC). All trial data were available to all the authors. The authors assume responsibility for the accuracy and completeness of the data and for the fidelity of the trial to the protocol.

### Trial Procedures

Participants were randomly assigned in a 1:1 ratio via block randomization to receive two doses of NVX-CoV2373 or placebo (normal saline), 21 days apart, using a centralized Interactive Response Technology system according to pre-generated randomization schedules.

Randomization was stratified by site and by age ≥65 years. In a 400-person sub-study, participants received a concomitant dose of seasonal influenza vaccine with the first dose (results of this sub-study will be represented separately).

This was an observer-blinded study. Only unblinded site personnel managed study vaccine logistics and preparation and they were not involved in study-related assessments or had participant contact for data collection following vaccine administration.

### Safety

After each vaccination, participants were observed for at least 30 minutes to monitor for the presence of any acute reactions. Solicited local and systemic adverse events were collected via an electronic diary for 7 days after each dose in a subgroup of participants (solicited adverse event subgroup). All participants were assessed for unsolicited adverse events from the first dose through 28 days after the second dose; serious adverse events, adverse events of special interest, and medically attended adverse events are being assessed from the first dose through 1 year after the second dose. Unsolicited adverse event terms that matched solicited adverse event terms for 7 days after each dose were excluded. Safety data are reported for all participants who received at least one dose of vaccine or placebo.

### Efficacy

The primary endpoint was the efficacy of NVX-CoV2373 against the first occurrence of virologically confirmed symptomatic mild, moderate, or severe Covid-19, with onset at least 7 days after second vaccination in participants who were seronegative at baseline. Symptomatic Covid-19 was defined according to US Food and Drug Administration (FDA) criteria (see Tables S1 and S2 in the Supplementary Appendix). Symptoms of suspected Covid-19 were monitored throughout the trial and collected using an electronic symptom diary (InFLUenza Patient-Reported Outcome [FLU-PRO^©^] questionnaire) for at least 10 days. At the onset of suspected symptoms, respiratory specimens from the nose and throat were collected daily over a 3-day period to confirm SARS-CoV-2 infection and a clinical assessment was performed. Virological confirmation was performed using polymerase chain reaction (PCR) testing (UK DHSC laboratories) with the TaqPath™ system (Thermo Fisher Scientific, Waltham, MA, USA) (see Supplementary Appendix for details).

### Statistical Analysis

#### Safety Analysis

Safety was analyzed in all participants who received at least one dose of NVX-CoV2373 or placebo and summarized descriptively. Solicited local and systemic adverse events were summarized by FDA toxicity grading criteria (see protocol in the Supplementary Appendix) and duration after each injection. Unsolicited adverse events were coded by preferred term and system organ class using the *Medical Dictionary for Regulatory Activities* (MedDRA), version 23.1, and summarized by severity and relationship to study vaccine.

#### Efficacy Analysis

The trial was designed and driven by the total number of events expected to achieve statistical significance for the primary endpoint – a target of 100 mild, moderate, or severe Covid-19 cases. The target number of 100 cases for the final analysis provides >95% power for 70% or higher vaccine efficacy. A single interim analysis of efficacy was conducted based on the accumulation of approximately 50% (50 events) of the total anticipated primary endpoints using Pocock boundary conditions. The main (hypothesis testing) event-driven analysis for the interim and final analyses of the primary objective was carried out at an overall one-sided type I error rate of 0.025 for the primary endpoint. The primary endpoint was analyzed in participants who were seronegative at baseline, received both doses of study vaccine or placebo, had no major protocol deviations affecting the primary endpoint, and had no confirmed cases of symptomatic Covid-19 from the first dose until 6 days after the second dose (per-protocol efficacy population). Vaccine efficacy was defined as VE (%) = (1 – RR) × 100, where RR = relative risk of incidence rates between the two study groups (NVX-CoV2373 or placebo). Mean disease incidence rate was reported as incidence rate per year in 1000 people. The estimated RR and its confidence interval (CI) were derived using Poisson regression with robust error variance. Hypothesis testing of the primary endpoint was carried out against the null hypothesis: H0: vaccine efficacy ≤30%. The success criterion required rejection of the null hypothesis to demonstrate a statistically significant vaccine efficacy.

## RESULTS

### Participants

Between September 28 and November 28, 2020, a total of 16,645 participants were screened and 15,187 participants were randomized (Figure 1). A total of 15,139 participants received at least one dose of NVX-CoV2373 (7569) or placebo (7570), with 14,039 participants (7020 in the NVX-CoV2373 group and 7019 in the placebo group) meeting the criteria for the per-protocol efficacy population. Baseline demographics were well balanced between the NVX-CoV2373 and placebo groups in the per-protocol efficacy population, where 48.4% were female, 94.5% were White, 2.9% were Asian, 0.4% were Black or African American, and 44.6% had at least one comorbid condition (based on Centers for Disease Control and Prevention [CDC] definitions.^12^ The median age was 56 years, and 27.9% were ≥65 years old (Table 1).

**Table 1.**
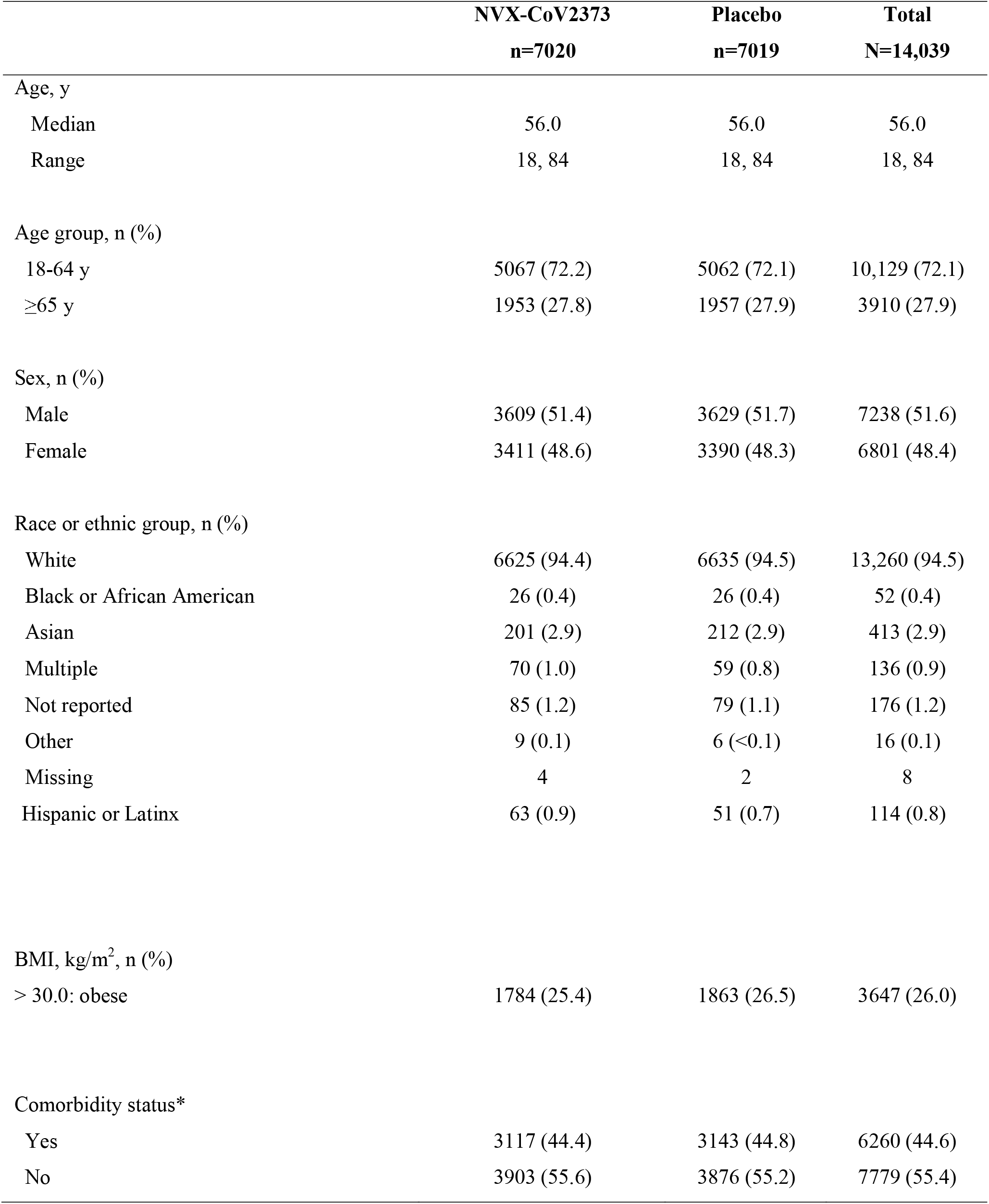
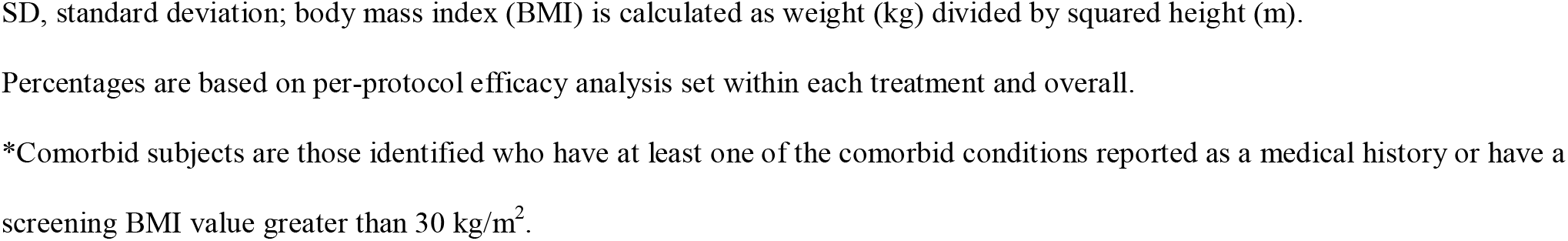
Demographics and Baseline Characteristics (Per-Protocol Efficacy Analysis)

|  | **NVX-CoV2373**<br>**n=7020** | **Placebo**<br>**n=7019** | **Total**<br>**N=14,039** |
| --- | --- | --- | --- |
| Age, y |  |  |  |
| Median | 56.0 | 56.0 | 56.0 |
| Range | 18, 84 | 18, 84 | 18, 84 |
| Age group, n (%) |  |  |  |
| 18-64 y | 5067 (72.2) | 5062 (72.1) | 10,129 (72.1) |
| ≥65 y | 1953 (27.8) | 1957 (27.9) | 3910 (27.9) |
| Sex, n (%) |  |  |  |
| Male | 3609 (51.4) | 3629 (51.7) | 7238 (51.6) |
| Female | 3411 (48.6) | 3390 (48.3) | 6801 (48.4) |
| Race or ethnic group, n (%) |  |  |  |
| White | 6625 (94.4) | 6635 (94.5) | 13,260 (94.5) |
| Black or African American | 26 (0.4) | 26 (0.4) | 52 (0.4) |
| Asian | 201 (2.9) | 212 (2.9) | 413 (2.9) |
| Multiple | 70 (1.0) | 59 (0.8) | 136 (0.9) |
| Not reported | 85 (1.2) | 79 (1.1) | 176 (1.2) |
| Other | 9 (0.1) | 6 (<0.1) | 16 (0.1) |
| Missing | 4 | 2 | 8 |
| Hispanic or Latinx | 63 (0.9) | 51 (0.7) | 114 (0.8) |
| BMI, kg/m<sup>2</sup>, n (%) |  |  |  |
| > 30.0: obese | 1784 (25.4) | 1863 (26.5) | 3647 (26.0) |
| Comorbidity status* |  |  |  |
| Yes | 3117 (44.4) | 3143 (44.8) | 6260 (44.6) |
| No | 3903 (55.6) | 3876 (55.2) | 7779 (55.4) |

SD, standard deviation; body mass index (BMI) is calculated as weight (kg) divided by squared height (m).
Percentages are based on per-protocol efficacy analysis set within each treatment and overall.
\*Comorbid subjects are those identified who have at least one of the comorbid conditions reported as a medical history or have a screening BMI value greater than 30 kg/m<sup>2</sup>.

**Figure 1.**
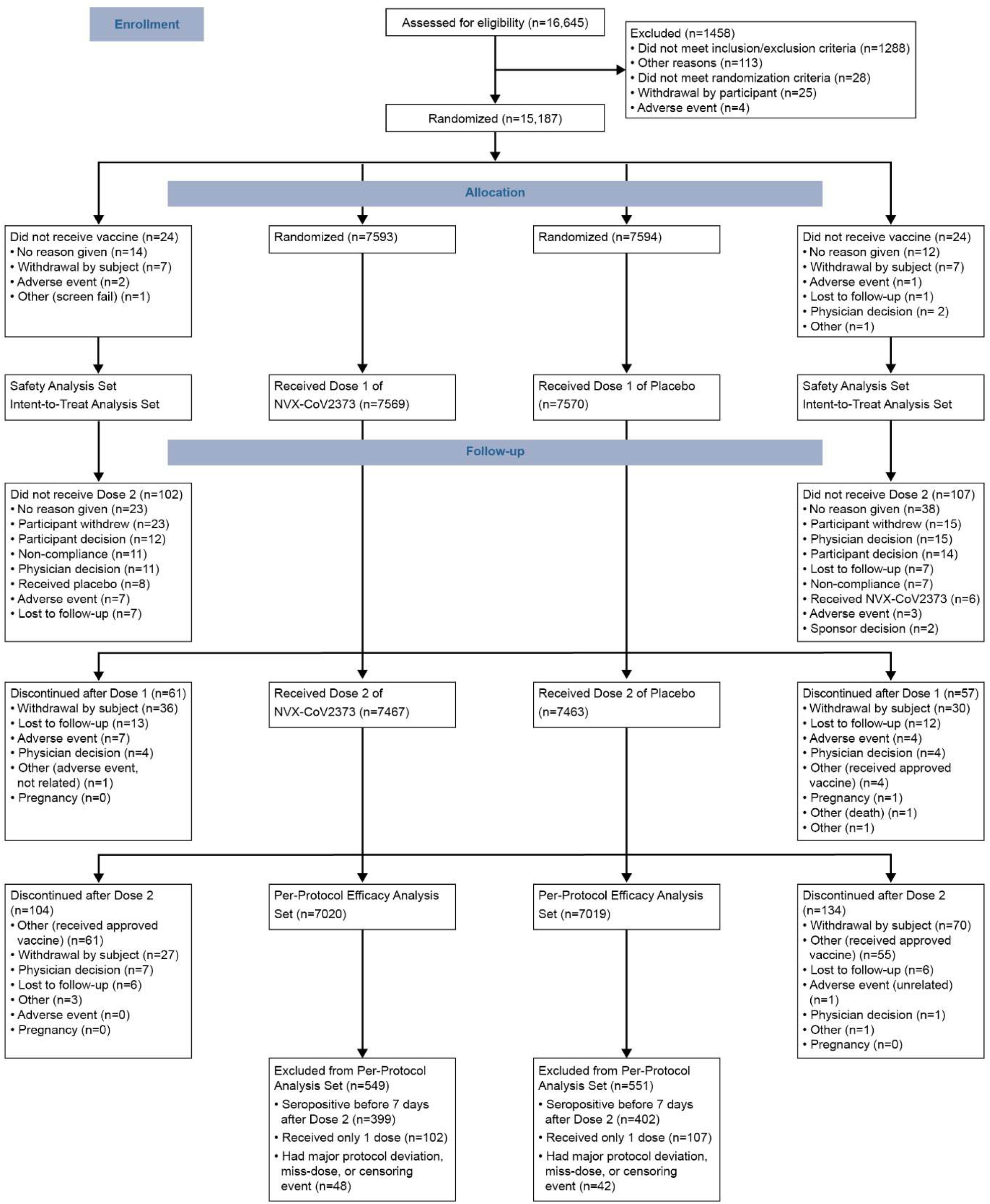
Disposition of Participants in the Trial. The full analysis set included all participants who were randomly assigned to treatment and received at least one dose, regardless of protocol violations or missing data, and are analyzed according to the trial vaccine group as randomized. IgG, immunoglobulin G; PCR, polymerase chain reaction; SARS-CoV-2, severe acute respiratory syndrome coronavirus 2.

### Safety

The solicited adverse event subgroup included 2310 participants. Overall, NVX-CoV2373 recipients reported higher frequencies of solicited local adverse events than placebo recipients after both the first dose (57.6% vs. 17.9%) and the second dose (79.6% vs. 16.4%) (Figure 2). Among NVX-CoV2373 recipients, the most commonly reported local adverse events were injection site tenderness and pain after both the first dose (53.3% and 29.3%) and the second dose (76.4% and 51.2%), with most events being grade 1 (mild) or 2 (moderate) in severity and of short mean duration (2.3 and 1.7 days after the first dose and 2.8 and 2.2 days after the second dose). Solicited local adverse events were reported more frequently among younger NVX-CoV2373 recipients (18 to 64 years) than older NVX-CoV2373 recipients (≥65 years).

**Figure 2.**
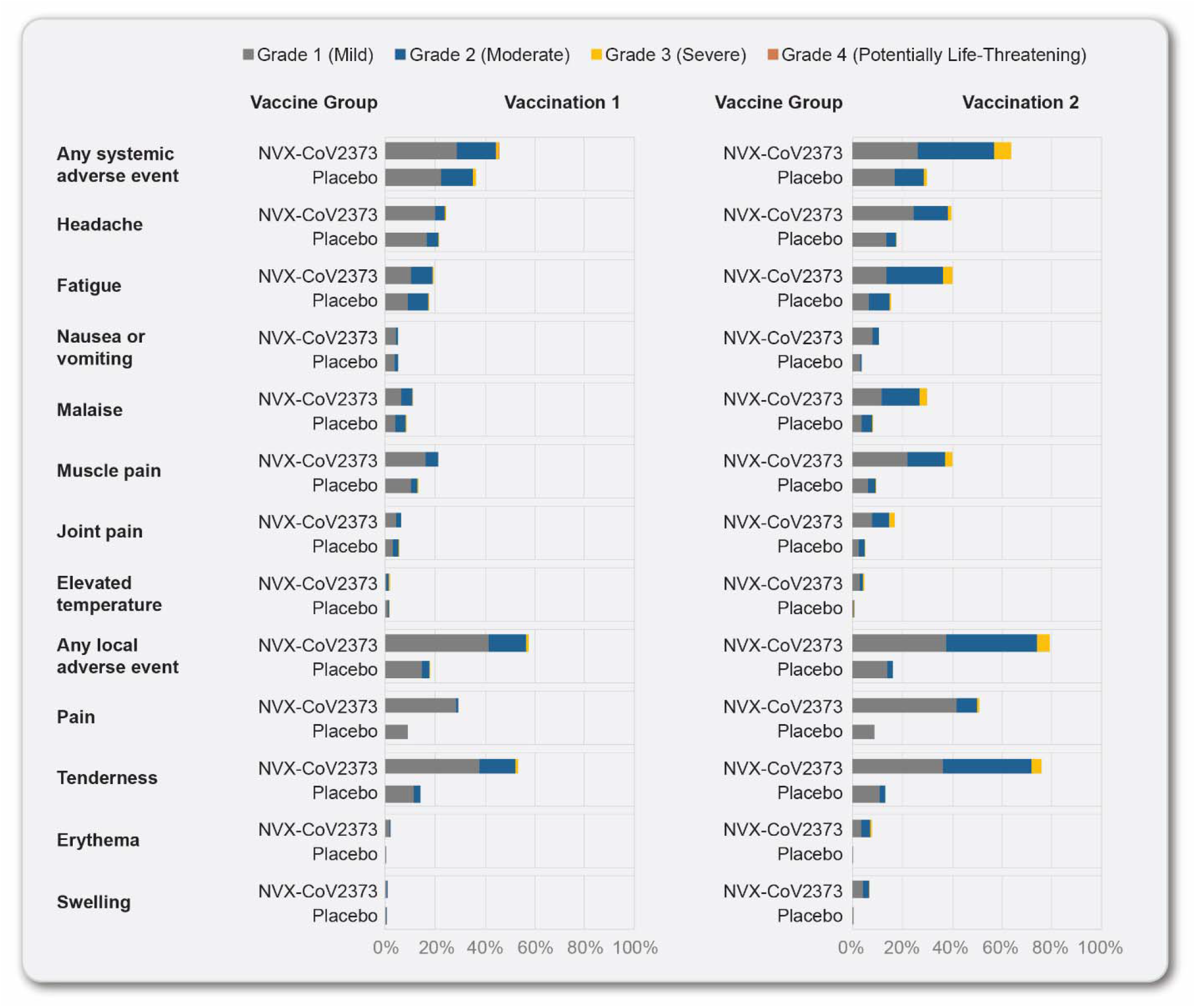
Solicited Local and Systemic Adverse Events. The percentage of participants in each treatment group with solicited local and systemic adverse events during the 7 days after each vaccination is plotted according to the maximum toxicity grade (mild, moderate, severe, or potentially life-threatening) (excludes participants in the seasonal influenza vaccine sub-study).

Overall, NVX-CoV2373 recipients reported higher frequencies of solicited systemic adverse events than placebo recipients after both the first dose (45.7% vs. 36.3%) and the second dose (64.0% vs. 30.0%) (Figure 2). Among NVX-CoV2373 recipients, the most commonly reported systemic adverse events were headache, muscle pain, and fatigue after both the first dose (24.5%, 21.4%, and 19.4%) and the second dose (40.0%, 40.3%, and 40.3%), with most events being grade 1 or 2 in severity and of short mean duration (1.6, 1.6, and 1.8 days after the first dose and 2.0, 1.8, and 1.9 days after the second dose). Grade 4 systemic adverse events were reported in two NVX-CoV2373 participants after the first dose and in one NVX-CoV2373 participant after the second dose. Systemic adverse events were reported more often by younger vaccine recipients than by older vaccine recipients and more often after dose 2 than dose 1. Notably, fever (temperature ≥38°C) was reported in 2.0% and 4.8% of NVX-CoV2373 participants after the first and second doses, with grade 3 fever (39-40°C) in 0.4% and 0.6% of participants after the first and second doses, respectively; one grade 4 fever (>40°C) was reported after each dose of vaccine.

All 15,139 participants who received at least one dose of vaccine or placebo through the data cutoff date of the final efficacy analysis were assessed for unsolicited adverse events. The frequency of unsolicited adverse events was higher among NVX-CoV2373 recipients than among placebo recipients (25.3% vs. 20.5%), with similar frequencies of severe adverse events (1.0% vs. 0.8%), serious adverse events (0.5% vs. 0.5%), medically attended adverse events (3.8% vs. 3.9%), adverse events leading to dose (0.3% vs. 0.3%) or study (0.2% vs. 0.2%) discontinuation, potentially immune-mediated medical conditions (<0.1% vs. <0.1%), and adverse events of special interest relevant to Covid-19 (0.1% vs. 0.3%). One related serious adverse event was reported in an NVX-CoV2373 recipient (myocarditis), which occurred 3 days after the second dose and was considered a potentially immune-mediated condition; an independent SMC considered the event most likely a viral myocarditis. The participant recovered. There were no episodes of anaphylaxis, and no evidence of vaccine-associated enhanced Covid-19. Two Covid-19-related deaths were reported, one in the NVX-CoV2373 group, with onset of symptoms 7 days after receiving a single vaccine dose, and one in the placebo group.

### Efficacy

Among 14,039 participants in the per-protocol efficacy population, there were 10 cases of virologically confirmed, symptomatic mild, moderate or severe Covid-19 with onset at least 7 days after the second dose among vaccine recipients (6.53 per 1000 person-years; 95% CI: 3.32 to 12.85) and 96 cases among placebo recipients (63.43 per 1000 person-years; 95% CI: 45.19 to 89.03) for a vaccine efficacy of 89.7% (95% CI, 80.2 to 94.6; Figure 3). Of the 10 cases ≥65 years old who had mild, moderate, or severe Covid-19, one had received NVX-CoV2373 and nine had received placebo (Figure 4). Severe Covid-19 occurred in five participants, of whom none had received NVX-CoV2373 and five had received placebo. There were no hospitalizations or deaths among per-protocol vaccine recipients.

**Figure 3.**
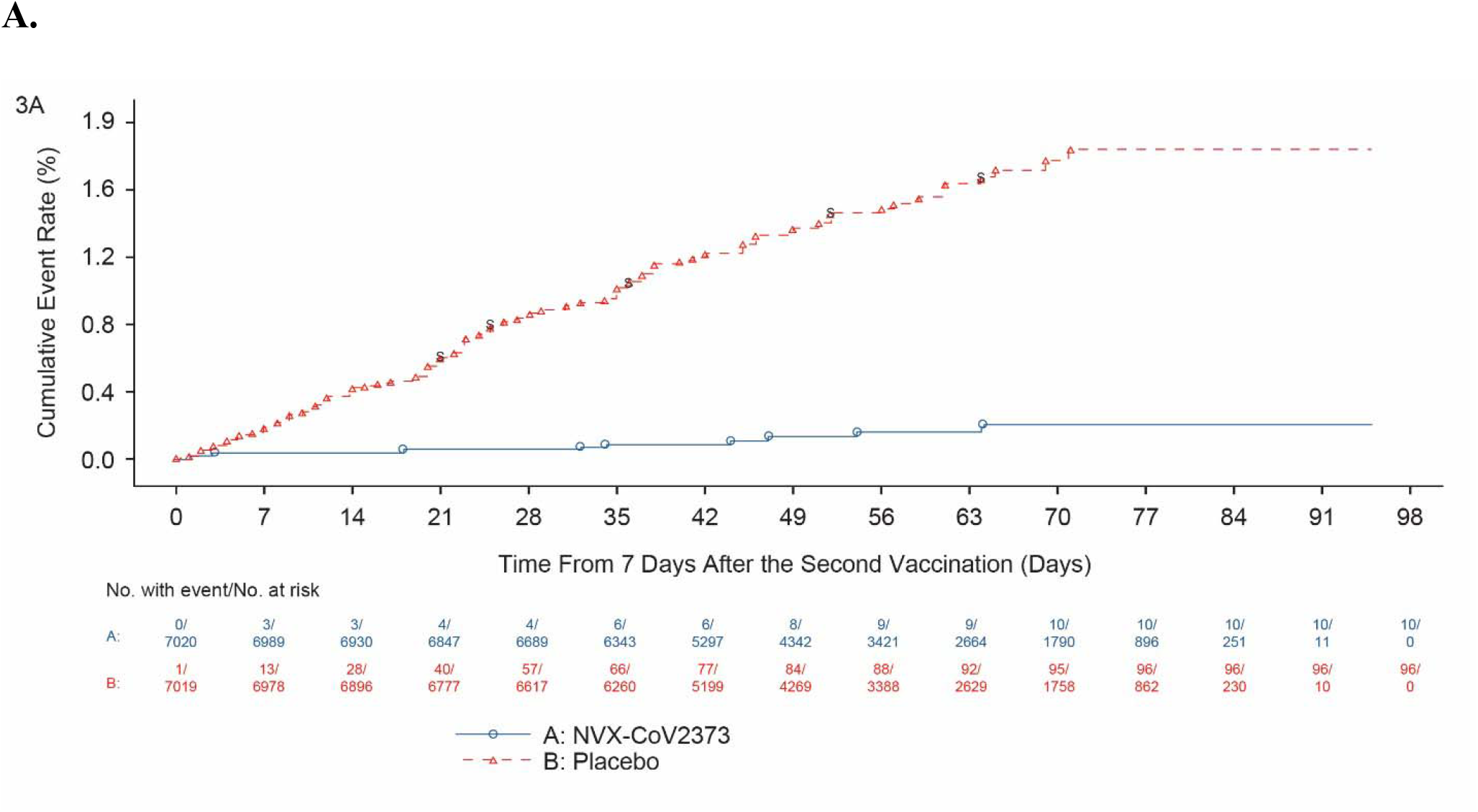
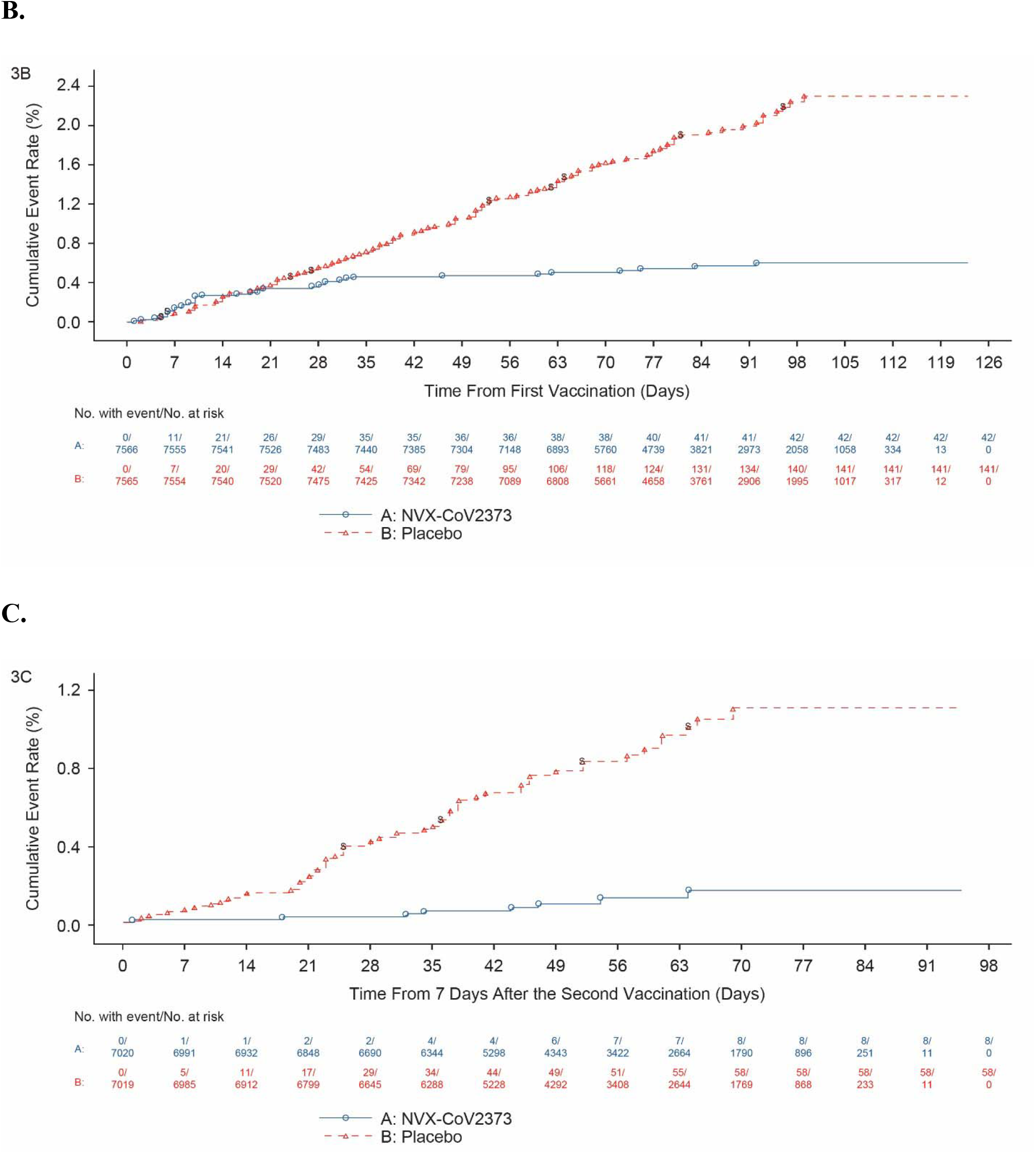
Kaplan-Meyer Plots of Efficacy of NVX-CoV2373 Against Symptomatic Covid-19 in the Per-Protocol and Intention-to-Treat Analysis Sets and due to the B.1.1.7 Variant. Shown is the cumulative incidence of symptomatic Covid-19. The time period for surveillance of per-protocol symptomatic Covid-19 cases was from at least 7 days after the second dose (i.e., Day 28) of NVX-CoV2373 or placebo through the first approximate 3 months of follow-up. A) All participants, baseline seronegative, per-protocol population; B) All participants, intention-to-treat population; C) All participants, baseline seronegative, per-protocol population with endpoints due to the B.1.1.7 variant.

**Figure 4.**
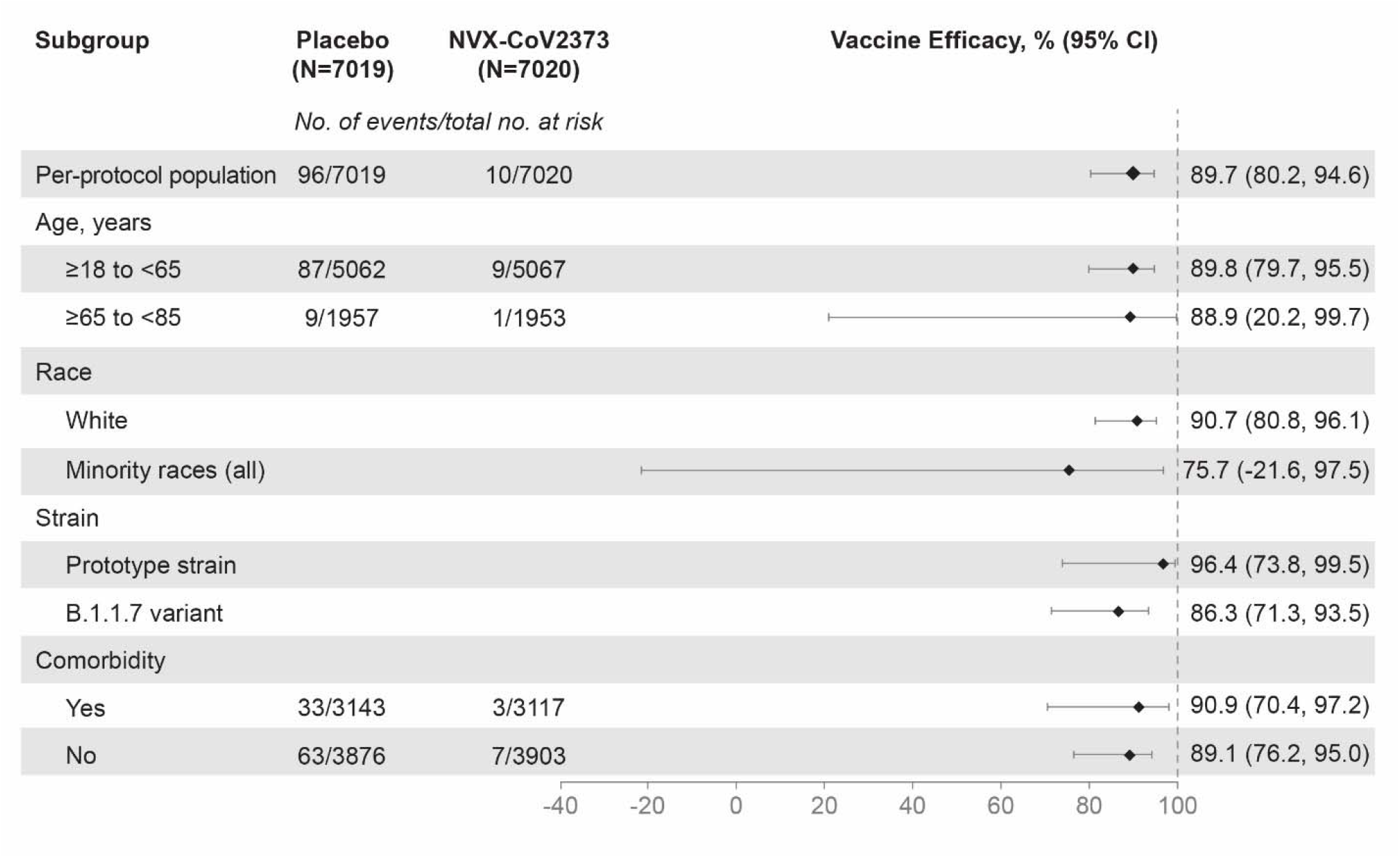
Vaccine Efficacy of NVX-CoV2373 in Specific Subgroups. The efficacy of NVX-2373 in preventing Covid-19 in various subgroups within the per-protocol population. Vaccine efficacy was defined as 1 minus the relative risk (NVX-CoV2373 vs. placebo) and 95% confidence intervals were derived using Poisson regression with robust error variance. Minority races included ethnic minorities and multiple races (only 2 endpoints in the NVX-Cov2373 group and 8 endpoints in the placebo group). Data from minority and multiple races were pooled to ensure that the subpopulations would be large enough for meaningful analyses. Comorbidity assessment is based on the Centers for Disease Control and Prevention definitions^12^ of those at increased risk for Covid-19.

Additional efficacy analyses (among subgroups defined by age, race, and presence of comorbid conditions) are detailed in Figure 4. Of note, vaccine efficacy among participants ≥65 years was 88.9% (95% CI, 12.8 to 98.6) and efficacy from 14 days after dose 1 was 83.4% (95% CI, 73.6 to 89.5). A post hoc analysis of the primary endpoint identified 29, 66, and 11 cases of Covid-19 where the isolated strain was the prototype strain, B.1.1.7 variant, or unknown, respectively.

Unknown samples were those where the PCR tests were performed with a non-DHSC PCR test (e.g., at a local hospital laboratory) where variant determination was not performed. Vaccine efficacy against the prototype strain was 96.4% (95% CI, 73.8 to 99.4), while efficacy against the B.1.1.7 variant was 86.3% (95% CI, 71.3 to 93.5).

## DISCUSSION

A two-dose regimen of NVX-CoV2373, given 21 days apart, was found to be safe and 89.7% effective against symptomatic Covid-19 caused by both prototype and B1.1.7 variants. The timing of accumulated cases in this study (see Table S5 in the Supplementary Appendix) allowed for a post hoc assessment of vaccine efficacy against different strains, including the B.1.1.7 variant, which is now circulating widely outside of the United Kingdom and is soon expected to be the most prominent strain in the United States.^13^ This variant is known to be more transmissible and to be associated with a higher case fatality rate than previous strains.^7^ This is the first vaccine to demonstrate high vaccine efficacy (86.3%) against the B.1.1.7 variant in a phase 3 trial. Immunogenicity data will be reported elsewhere.

Though the study was not powered to assess efficacy for individual SARS-CoV-2 strains, it is reassuring to demonstrate significant efficacy against all strains detected in trial participants. In particular, the 96.4% point estimate of efficacy determined against the prototype strain is similar to that reported against this strain for the BNT161b2 mRNA vaccine (95.0%) and the mRNA-1273 vaccine (94.1%)^3,4^ and greater than that demonstrated by the adenoviral vector vaccines.^5,6^ Finally, as assessed elsewhere, the NVX-CoV2373 vaccine is also the first vaccine to show efficacy against the B.1.351 variant, albeit at lower levels than those shown for the B.1.1.7 and prototype strains.^14^ This emphasizes the need to consider additional strategies, including the use of modified vaccines and booster doses, in order to target significant variants as they emerge.

Prevention of severe disease (including hospitalization, intensive care admission, and death) is an important objective of a vaccination program, and the two-dose regimen of NVX-CoV2373 demonstrated very high efficacy against this endpoint, similar to that reported for other licensed Covid-19 vaccines.^3-6^ In addition, though not designed to assess efficacy after a single dose, NVX-CoV2373 provided levels of protection after the first dose in a range similar to that of other Covid-19 vaccines.^3-6^ The favorable safety profile observed during phase 1/2 studies of NVX-CoV2373 was confirmed in this phase 3 trial. Reactogenicity was generally mild or moderate, and reactions were less common and milder in older subjects and more common after the second dose. Injection site tenderness and pain, fatigue, headache, and muscle pain were the most commonly reported local and systemic adverse events. The incidence of serious adverse events was similar in the vaccine and placebo groups (0.5% in each) and no deaths were attributable to receipt of the vaccine.

This trial has several limitations. Although approximately 7500 participants received NVX-CoV2373, it is not possible to exclude the occurrence of rare adverse events. Such events may, however, be captured through the ongoing follow-up of participants, which is planned to continue until 12 months after the second dose of vaccine and will also be assessed in the larger PREVENT-19 phase 3 trial.^15^ Similarly, the efficacy estimates reported here are derived from a relatively short duration of observation (median 3 months after dose 2). The ongoing follow-up will therefore allow determination of the durability of vaccine efficacy, continued assessment of severe cases, and the assessment of efficacy against asymptomatic disease.

A further limitation is the lack of sequencing data on study isolates, although the use of S gene target failure, as detected by the Thermo Fisher TaqPath™ assay used in DHS Public Health England has proved to be a good proxy for the B.1.1.7 variants.^16^ Additionally, a convenience sample of isolates from this study were sequenced by the Covid-19 genomics UK consortium and demonstrated a similar proportion of B.1.1.7 variants as detected by PCR.^17^

The results of this trial provide further evidence that both prototype and B.1.1.7 Covid-19 can be prevented by immunization, providing the first evidence for a protein-based, adjuvanted vaccine. NVX-CoV2373 can be stored at standard refrigerator temperatures and has the potential to induce a broad epitope response to the spike protein antigen. Both of these attributes are vitally important for the efficient implementation of this vaccine globally and the continued need to vaccinate against emerging variants. We propose that NVX-CoV2373 will be a valuable new tool in our battle to control the Covid-19 pandemic.

Funded by Novavax, Inc. EudraCT number, 2020-004123-16.

Disclosure forms provided by the authors will be available with the full text of this article upon publication.

A data sharing statement provided by the authors will be available with the full text of this article upon publication.

We thank all the participants who volunteered for this study, the members of the safety monitoring committee, and the NVX-CoV2373 team members, who are listed in the Supplementary Appendix.

We are grateful to the UK National Institute for Health Research (NIHR) Clinical Research Network and the UK Vaccine Task Force.

The views expressed in this publication are those of the author(s) and not necessarily those of the NIHR or the Department of Health and Social Care.

## Supporting information

Trial Approval letter

Redacted protocol and SAP

## Data Availability

Limited data will be available on the following websites:

https://clinicaltrials.gov/ct2/show/NCT04583995

https://www.clinicaltrialsregister.eu/ctr-search/trial/2020-004123-16/GB

## REFERENCES

1. World Health Organization. WHO Coronavirus Disease (COVID-19) Dashboard. https://covid19.who.int/. Accessed April 19, 2021.

2. Lan J, Ge J, Yu J, et al. Structure of the SARS-CoV-2 spike receptor-binding domain bound to the ACE2 receptor. Nature 2020;581-215-220.

3. Baden LR, El Sahly HM, Essink B, et al; COVE Study Group. Efficacy and safety of the mRNA-1273 SARS-CoV-2 vaccine. N Engl J Med 2021;384:403–16.

4. Polack FP, Thomas SJ, Kitchin N, et al. Safety and efficacy of the BNT162b2 mRNA Covid-19 vaccine. N Engl J Med 2020;383:2603–15.

5. Voysey M, Costa Clemens SA, Madhi SA, et al. Safety and efficacy of the ChAdOx1 nCoV-19 vaccine (AZD1222) against SARS-CoV-2: an interim analysis of four randomised controlled trials in Brazil, South Africa, and the UK. Lancet 2021;397:99–111.

6. Janssen Biotech, Inc. FDA Briefing Document: Janssen Ad26.COV2.S Vaccine for the Prevention of COVID-19. Presented at Vaccines and Related Biological Products Advisory Committee Meeting, February 26, 2021. https://www.fda.gov/media/146217/download. Accessed April 7, 2021.

7. Challen R, Brooks-Pollock E, Read JM, et al. Risk of mortality in patients infected with SARS-CoV-2 variant of concern 202012/1: matched cohort study. BMJ 2021;372:579 http://dx.doi.org/10.1136/bmj.n579.

8. Walensky RP, Walke HT, Fauci AS. SARS-CoV-2 variants of concern in the United States – challenges and opportunities. JAMA 2021;325:1037–8.

9. Callaway E. Could new COVID variants undermine vaccines? Labs scramble to find out. Nature 2021;589:177–8.

10. Keech C, Albert G, Cho I, et al. Phase 1-2 trial of a SARS-CoV-2 recombinant spike protein nanoparticle vaccine. N Engl J Med 2020;383:2320–32.

11. Formica N, Mallory R, Albert G, et al. Evaluation of a SARS-CoV-2 vaccine NVX-CoV2373 in younger and older adults. medRxiv 2021.02.26.21252482. doi: https://doi.org/10.1101/2021.02.26.21252482.

12. Centers for Disease Control and Prevention. People with certain medical conditions. Updated March 29, 2021. https://www.cdc.gov/coronavirus/2019-ncov/need-extra-precautions/people-with-medical-conditions.html. Accessed April 7, 2021.

13. Centers for Disease Control and Prevention. About variants of the virus that causes Covid-19. Updated April 2, 2021. https://www.cdc.gov/coronavirus/2019-ncov/transmission/variant.html. Accessed April 7, 2021.

14. Public Heath England. Investigation of novel SARS-CoV-2 variant. December 2020. PHE gateway number: GOV-7132. https://assets.publishing.service.gov.uk/government/uploads/system/uploads/attachment_data/file/959361/Technical_Briefing_VOC202012-2_Briefing_2.pdf. Accessed April 7, 2021.

15. Shinde V, Bhikha S, Hoosain Z, et al. Preliminary efficacy of the NVX-CoV2373 Covid-19 vaccine against the B.1.351 variant. medRxiv 2021.02.25.21252477; doi: https://doi.org/10.1101/2021.02.25.21252477.

16. US National Library of Medicine. A Study Looking at the Efficacy, Immune Response, and Safety of a COVID-19 Vaccine in Adults at Risk for SARS-CoV-2. February 24, 2021. https://clinicaltrials.gov/ct2/show/NCT04611802?term=PREVENT-19&draw=2&rank=1. Accessed April 7, 2021.

17. Novavax, Inc. Data on file.

