## Supplemental Appendix for "Efficacy of the NVX-CoV2373 Covid-19 Vaccine Against the B.1.1.7 Variant"

This appendix has been provided by the authors to give readers additional information about their work.

Supplementary Appendix to Manuscript Entitled

**Efficacy of the NVX-CoV2373 Covid-19 Vaccine Against the B.1.1.7 Variant**

**Table of Contents**

NVX-CoV2373 Study Team Members …………………………………………………………….……………………………….	3

Supplemental Methods ………………………………………………………………………………….………………………………	6	Vaccination Pause Rules …………………………………………………………………………….………………………………	6

RT-PCR Testing Assays ………………………………………………………………………………….……………………………	6

Swabbing …………………………………………………………………………………………………………………….…………….	7

Supplemental Tables and Figure …………………………………………………………………………………….…………….	8

Primary Efficacy Endpoint …………………………………………………………………………………………….…………..	8

Table S1. Endpoint Definitions of Covid-19 Severity …………………………………………………….……	8

Table S2. Qualifying Symptoms of Suspected Covid-19 ………………………………………………….….	9

Unsolicited Adverse Events ……………………………………………………………………………………………………….	10

Table S3. Overall Summary of Unsolicited Adverse Events — Excluding Reactogenicity

Adverse Events (Specific Preferred Terms) and Events Reported Post Unblinding or

Post Receipt of an Approved or Deployed SARS-CoV-2 Vaccine (Safety Analysis Set) ….…….	10

Table S4. Summary of Unsolicited Treatment-Emergent Adverse Events (Excluding

Reactogenicity Adverse Events) Reported in ≥1% of Participants in Any Group

(Safety Analysis Set) ………………………………………………………………………………………………………….	11

Efficacy Against UK Variant ……………………………………………………………………………….….…….……….……	11

Table S5. Vaccine Efficacy Against PCR-Confirmed Symptomatic Mild, Moderate, or Severe

Covid-19 With an Onset at Least 7 Days After Second Study Vaccination in Serologically

Negative Adult Participants by SARS-CoV-2 Strain (PP-EFF Analysis Set) ……………………………	12

Demographics: Intention-to-Treat Population …………………………………………………………………………..	13

Table S6. Demographics and Baseline Characteristics (Intention-to-Treat Analysis Set) …….	13

Table S7. Summary of Unsolicited Serious Treatment-Emergent Adverse Events

(Safety Analysis Set) ………………………………………………………………………………………….………………	14	B1.1.1.7 Variant .…………………………………………………………………………………………………………….............	16

Figure S1. Evolution of Covid-19 Variants During the Endpoint Period ………………………………	17

References ……………………………………………………………………………………………………………………………………..	17

### NVX-CoV2373 Study Team Members

The NVX-CoV2373 clinical trial was a collective group effort across multiple institutions and locations. Below is a list of sites and staff that significantly contributed to the implementation and conduct of the NVX-CoV2373-2019nCoV-302 clinical trial.

| **Site** | **Investigators** |
| --- | --- |
| Aberdeen Royal Infirmary, NHS Grampian | Robin Brittain-Long, Chiara Scicluna, Carole Edwards, Lynn Mackay, Mariella D’Allesandro, Amy Nicol, Karen Norris, Sandra Mann, Heather Lawrence, Ruth Valentine |
| Accelerated Enrollment Solutions | Nick Richards, Helen Price, Bridie Rowbotham, Danielle Bird, Karen Smith, Olga Littler, Kirsty Fielding, Anna Townsend-Rose, Karen Miller, Jessica Davis, Alison Elliot-Garwood, Lauren Trottier, Paul Edwards |
| Belfast Health and Trust | Margaret McFarland |
| Betsi Cadwaladr University Health Board | Laura Longshaw, Jane Stockport, Victoria Saul, Alice Thomas, Lynne Grundy |
| Blandford Group Practice | Katharine Lucy Broad |
| Bradford Teaching Hospitals NHS Foundation Trust | Karen Regan, Kim Storton, Declan Ryan-Wakeling, Brad Wilson, Malathy Munisamy, John Wright, Anil Shenoy, Beverley English, Lucy Brear |
| Centre for Clinical Vaccinology and Tropical Medicine, University of Oxford | Paola Cicconi |
| Chelsea and Westminster Hospital | Ana Milinkovic, Ruth Byrne, Roya Movahedi, Rosalie Housman |
| County Durham & Darlington NHS Foundation Trust | Naveed Kara, Ellen Brown |
| CRN Thames Valley and South Midlands, Oxford University Hospitals NHS Foundation Trust | Helen Collins |
| Department of Psychiatry, University of Oxford | Andrea Cipriani, Mary-Jane Attenburrow, Katharine A Smith |
| Dorset Research Hub, Royal Bournemouth Hospital, University Hospitals Dorset NHS Foundation Trust | Geoff Sparrow |
| East Suffolk and North Essex NHS Foundation Trust | Josephine M Rosier, Khalid Saja, Nyasha Nago, Brian Camilleri, Anita Immanuel, Mike Hamblin, Rawlings Osagie, Mahalakshmi Mohan |
| Epsom and St Helier University Hospitals NHS Trust | Hilary Floyd, Suzanne Goddard, Sanjay Mutgi, John Evans, Sean McKeon, Neringa Vilimiene, Rosavic Chicano, Rachel Hayre, Alice Pandaan |
| Health and Care Research Wales | Nicola Williams, Jayne Goodwin |
| Highcliffe Medical Centre | Zelda Cheng |
| Keele University | Toby Helliwell, Adrian Chudyk |
| Kings College London | Rafaela Giemza, John Lord Villajin, Noah Yogo, Esther Makanju, Pearl Dulawan, Deepak Nagra, April Buazon, Alice Russell, Georgie Bird |
| Lakeside Healthcare Research, Lakeside Surgery | Rex Sarmiento, Balraj Sanghera, Melanie Mullin, Adam Champion, Aisling Bevan, Kinzah Iqbal, Alshia Johnson |
| Layton Medical Centre | Sarah Shaw, Steven Shaw, Amanda Chalk, Martin Lovatt, Caroline Lillicrap, Angela Parker, Jan Hansel, Zhi Wong, Galvin Gan, Eyad Tuma |
| Leeds Teaching Hospitals NHS Trust | Jennifer Murira, Razan Saman, Alistair Hall, Kyra Holliday, Zara Khan, James Calderwood, George Twigg, Helena Baker, Julie Corrigan, Katy Houseman |
| Midlands Partnership NHS Foundation Trust | Subhra Raguvanshi, Dominic Heining, Jake Weddell, Liz Glaves, Kim Thompson, Francis Davies, Ruth Lambley Burke |
| Guy's and St Thomas' NHS Foundation Trust NIHR BRC | Sonia Serrano, Andrea Mazzella, Thurkka Rajeswaran, Moncy Mathew, Karen Bisnauthsing, Laura Bremner, Henry Fok, Franca Morselli, Paola Cinardo, Blair Merrick, Lucy Sowole |
| NIHR Clinical Research Facility, University Hospital Southampton NHS Foundation Trust | Lisa Berry |
| NIHR Southampton Clinical Research Facility and NIHR Wessex Local Clinical Research Network, University Hospital Southampton NHS Foundation Trust | Mihaela Pacurar, Saul N Faust |
| NIHR Clinical Research Network, Thames Valley and South Midlands | Nancy Hopewell, Leigh Gerdes |
| Norfolk and Norwich University Hospital NHS Foundation Trust | Adele Cooper, Jocelyn Keshet-Price, Lou Coke, Melissa Cambell-Kelly, Ketan Dhatariya, Claire Williams , Georgina Marks, James Sudbury, Lisa Rodolico |
| Northern Ireland Clinical Research Facility, Queen's University Belfast and Belfast Health and Social Care Trust | Judy Bradley, Sharon Carr, Roisin Martin, Angelina Madden, Paul Biagioni, Sonia McKenna, Alison Clinton |
| Northern Ireland Clinical Research Network | Maurice O’Kane |
| North Tees and Hartlepool NHS Foundation Trust | Justin Carter, Matthew Dewhurst, Bill Wetherill |
| Oxford Health NHS Foundation Trust, Warneford Hospital | Catherine Henshall, Jennifer Potts, Sarah McCartney, Katrina Lennon Collins, Marie Chowdhury, Adil Nathoo, Anna Heinen, Orla MacDonald, Claudia Hurducas, Liliana Cifuentes, Thomas Rawlinson, Harjeevan Gill |
| Quadram Institute | Jane Ewing |
| Queen Elizabeth University Hospital, NHS Greater Glasgow and Clyde | Guy Mollett, Rachel Blacow, John Haughney, Jonathan MacDonald, John Paul Seenan, Stewart Webb, Colin O'Leary, Scott Muir, Beth White, Neil Ritchie |
| Queen's University Belfast and Belfast Health and Social Care Trust | Jonathan Stewart |
| Research and Development, NHS Grampian | Chiara Scicluna, Mariella D'Alessandro, Carole Edwards, Lynn MacKay, Amy Nicol, Karen Norris, Heather Lawrence, Sandra Mann, Ruth Valentine |
| Royal Bournemouth Hospital, University Hospitals Dorset NHS Foundation Trust | Nicki Lakeman, Laura Purandare |
| Royal Cornwall Hospital NHS Trust | David Tucker, Peter Luck, Angharad Everden, Lisa Trembath, Michael Visick, Nick Morley, Laura Reid, Helen Chenoweth, Kirsty Maclean |
| Royal Devon and Exeter Hospital | Tom Burden, Craig Francis Lunt, Shirley Todd, Stephanie Estcourt, Jasmine Marie Pearce, Suzanne Wilkins, Cathryn Love-Rouse |
| Royal Free London NHS Foundation Trust | Eva Torok-Pollok, Mike Youle, Sara Madge, Danielle Solomon, Melissa Chowdhury, Aarti Nandani, Janet North, Nargis Hemat, Suluma Mohamed |
| Royal Oldham Hospital, Northern Care Alliance, Greater Manchester | Rachel Newport |
| Salford Royal Hospital, Northern Care Alliance, Greater Manchester | Chukwuma Chukwu, Olivia Wickens, Vikki O'loughlin, Hema Mistry, Louise Harrison, Robert Oliver, Anne-Marie Peers, Jess Zadik, Katie Doyle |
| South Tees Hospitals NHS Foundation Trust | Kerry Colling, Caroline Wroe, Marie Branch, Alison Chilvers, Sarah Essex |
| Stafford Town Primary Care Network | Mark Stone |
| Vaccine Institute, St George’s University of London & St. George's University Hospitals NHS Foundation Trust | Emily Beales, Olivia Bird, Zsofia Danos, Hazel Fofie, Cecilia Hultin, Sabina Ikram, Fran Mabesa, Aoife Mescall, Josyanne Pereira, Jennifer Pearce, Natalina Sutton |
| St Helens and Knowsley Teaching Hospitals NHS Trust | Emma Snashall |
| Stockport NHS Foundation Trust, Stepping Hill Hospital | Sara Bennett, Debbie Suggitt, Kerry Hughes, Wiesia Woodyatt, Lynsey Beacon, Alissa Kent, Chris Cooper, Milan Rudic, Simon Tunstall, Matthew Jackson |
| Swanage Medical Practice | Claire Hombersley |
| The Adam Practice | Rebecca Cutts |
| University College London | Danielle Solomon, Janet M. North |
| University Hospitals of Morecambe Bay NHS Foundation Trust | Marwan Bukhari, Mohamed Elnaggar, Michelle Glover, Fiona Richardson, Alexandra Dent, Shahzeb Mirza, Rajiv Ark, Jennie Han |
| University of Exeter Medical School, William Wright House, Royal Devon and Exeter Hospital | Suzy V Hope, Philip J Mitchelmore |
| University of Liverpool | Rostam Osanlou, Thomas Heseltine |

### Supplemental Methods

#### Vaccination Pause Rules

Study vaccination pause rules based on reactogenicity, adverse events (AEs), and serious adverse events (SAEs) related to study participation were in place to monitor participant safety for the initial set of vaccinations only.

AEs meeting any one of the following criteria resulted in a hold being placed on subsequent study vaccinations pending further review by the safety monitoring committee (SMC) at the direction of the SMC chair:

- Any toxicity grade 3 or higher (severe or potentially life-threatening) solicited (local or systemic) single AE term occurring in ≥10% of participants (after a minimum of 100 subjects are enrolled) in the SARS-CoV-2 rS with Matrix-M1 adjuvant group within the first 7 days after study vaccination.
- Any severe unsolicited single AE preferred term that the investigator assesses as related that occurs in ≥5% of participants (after a minimum of 100 subjects are enrolled) in the NVX2373 group within the first 7 days after study vaccination.

In addition, any SAE assessed as related to vaccine (final assessment by the Sponsor) was reported by the Sponsor to the SMC Chair as soon as possible, and within 24 hours of the Sponsor’s awareness of the event. Based on this initial report of the event to the SMC Chair, the Chair could advise the Sponsor to immediately pause enrollment and further dosing in either some or all participants in the study and to convene an ad hoc meeting or make alternative recommendations. The SMC Charter defines processes for how this review occurred and how the Chair’s recommendations were documented.

The Sponsor, along with the Medical Monitor, were able to request an SMC review for any safety concerns that arose in the study, even if they were not associated with any specific pause rule, for example, any SAE for which causality was at least possibly related.

#### RT-PCR Testing Assays

Virological confirmation was performed using polymerase chain reaction (PCR) testing at the United Kingdom (UK) Department of Health and Social Care (DHSC) laboratories, which utilized the TaqPath™ system (Thermo Fisher Scientific, Waltham, Massachusetts, USA). Testing using this system in the UK has shown a close correlation between VOC-202012/1 cases confirmed by genomic sequencing and TaqPath™ PCR results where the spike protein gene PCR target has not been detected but other PCR targets (N gene and ORF1ab gene) have been detected. Such a result is referred to as S gene negative, or S gene target failure, and has a strong association to infection with the B.1.1.7 variant in the UK. S gene negative results are therefore used as a proxy for this variant.

#### Swabbing

Education and assistance with the self-swab technique was provided at the initial study visit. An online National Health Service (NHS) video guide (<https://www.youtube.com/watch?v=zCqo7MhQT6U>) was also recommended to participants and played at some sites when available. Swabbing was to be performed by participants and swabs mailed to the DHSC testing laboratories. The nose/throat swab involved swabbing the back of the throat for 10 seconds and then swabbing the nose at a depth of approximately 2.5 cm for 10 to 15 seconds. All kits were registered online prior to posting. Participants were directly informed of their results by text or email from the NHS.

### Supplemental Tables and Figure

Primary Efficacy Endpoint
The primary efficacy endpoint was the first occurrence of virologically confirmed (by PCR to SARS-CoV-2), symptomatic mild, moderate, or severe Covid-19 (see definitions in Table S1), with onset at least 7 days after second study vaccination in serologically negative (to SARS-CoV-2) participants at baseline.

| Table S1. Endpoint Definitions of Covid-19 Severity | |
| --- | --- |
| **Covid-19 Severity** | **Endpoint Definitions** |
|  | First episode of PCR-positive nasal swab and ≥1 of symptomatic mild, moderate, or severe Covid-19: |
| **Mild** | Virologically confirmed SARS-CoV-2 infection plus ≥1 of:   - Fever (defined by subjective or objective measure, regardless of use of anti-pyretic medications) - New onset cough - ≥2 additional Covid-19 symptoms:   - New onset or worsening of shortness of breath or difficulty breathing compared to baseline.   - New onset fatigue.   - New onset generalized muscle or body aches.   - New onset headache.   - New loss of taste or smell.   - Acute onset of sore throat, congestion, or runny nose.   - New onset nausea, vomiting, or diarrhea. |
| **Moderate** | - High fever (≥38.4°C) for ≥3 days (regardless of use of anti-pyretic medications, need not be contiguous days). - Any evidence of significant lower respiratory tract infection (LRTI):   - Shortness of breath (or breathlessness or difficulty breathing) with or without exertion (greater than baseline).   - Tachypnea: 20 to 29 breaths per minute at rest.   - SpO_2_: 94% to 95% on room air.   - Abnormal chest X-ray or chest computerized tomography (CT) consistent with pneumonia or LRTI. - Adventitious sounds on lung auscultation (eg, crackles/rales, wheeze, rhonchi, pleural rub, stridor). |
| **Severe** | - Tachypnea: ≥30 breaths per minute at rest. - Resting heart rate ≥125 beats per minute. - SpO_2_: ≤93% on room air or PaO_2_/FiO_2_ <300 mmHg. - High flow oxygen (O_2_) therapy or non-invasive ventilation (NIV)/non-invasive positive pressure ventilation (NIPPV) (eg, continuous positive airway pressure [CPAP] or bilevel positive airway pressure [BiPAP]). - Mechanical ventilation or extracorporeal membrane oxygenation (ECMO). - One or more major organ system dysfunction or failure to be defined by diagnostic testing/clinical syndrome/interventions, including any of the following:   - Acute respiratory failure, including acute respiratory distress syndrome (ARDS).   - Acute renal failure.   - Acute hepatic failure.   - Acute right or left heart failure.   - Septic or cardiogenic shock (with shock defined as systolic blood pressure [SBP] <90 mm Hg OR diastolic blood pressure [DBP] <60 mm Hg).   - Acute stroke (ischemic or hemorrhagic).   - Acute thrombotic event: acute myocardial infarction (AMI), deep vein thrombosis (DVT), pulmonary embolism (PE).   - Requirement for: vasopressors, systemic corticosteroids, or hemodialysis. - Admission to an intensive care unit (ICU). - Death. |

Abbreviations: AMI = acute myocardial infarction; ARDS = acute respiratory distress syndrome; BiPAP = bi-level positive airway pressure; CPAP = continuous positive air pressure; CT = computed tomography; DBP = diastolic blood pressure; DVT = deep vein thrombosis; ECMO = extracorporeal membrane oxygenation; FiO_2_ = fraction of inspired oxygen; ICU = intensive care unit; LRTI = lower respiratory tract infection; NIV = non-invasive ventilation; NIPPV = non-invasive positive pressure ventilation; PAO_2_ = partial pressure of oxygen in the alveolus; PE = pulmonary embolism; SBP = systolic blood pressure; SpO_2_ = oxygen saturation.

*Participants with a single vital sign abnormality placing them in the moderate or severe categories must also meet the criteria for mild Covid-19.

Table S2 shows the qualifying symptoms of suspected Covid-19.

Table S2. Qualifying Symptoms of Suspected Covid-19

- Fever (body temperature >38°C, in the absence of other symptoms) or chills
- New onset or worsening of cough compared with baseline
- New onset or worsening of shortness of breath or difficulty breathing over baseline
- Severe fatigue
- New onset generalized muscle or body aches
- Headache
- New loss of taste or smell
- Sore throat
- Congestion or runny nose
- Nausea or vomiting
- Diarrhea

Abbreviations: Covid-19 = coronavirus disease 2019.

#### Unsolicited Adverse Events

Unsolicited TEAEs classified as severe, medically attended, serious, leading to vaccination or study discontinuation, potential immune-mediated medical conditions, or adverse events of special interest were reported at similar frequencies between the NVX-CoV2373 and placebo groups (Table S3).

| **Table S3. Overall Summary of Unsolicited Adverse Events — Excluding Reactogenicity Adverse Events (Specific Preferred Terms) and Events Reported Post Unblinding or Post Receipt of an Approved or Deployed SARS-CoV-2 Vaccine (Safety Analysis Set)** | | | | |
| --- | --- | --- | --- | --- |
| **Parameters** | **NVX-CoV2373 N=7569** | | **Placebo N=7570** | |
|  | **n (%)** | **[E]** | **n (%)** | **[E]** |
| Any TEAEs | 1912 (25.3) | [2916] | 1551 (20.5) | [2514] |
| Any severe TEAEs | 76 ( 1.0) | [92] | 64 ( 0.8) | [88] |
| Any treatment-related TEAEs | 824 (10.9) | [1071] | 344 ( 4.5) | [464] |
| Any severe treatment-related TEAEs | 13 ( 0.2) | [14] | 3 (<0.1) | [3] |
| Any MAAEs | 285 ( 3.8) | [335] | 295 ( 3.9) | [355] |
| Any treatment-related MAAEs | 33 ( 0.4) | [41] | 13 ( 0.2) | [15] |
| Any serious TEAEs | 41 ( 0.5) | [50] | 41 ( 0.5) | [50] |
| Any treatment-related serious TEAEs | 1 (<0.1) | [1] | 0 | 0 |
| Any TEAEs leading to vaccination discontinuation | 23 ( 0.3) | [27] | 22 ( 0.3) | [43] |
| Any treatment-related TEAEs leading to vaccination discontinuation | 7 (<0.1) | [9] | 8 ( 0.1) | [9] |
| Any TEAEs leading to study discontinuation | 17 ( 0.2) | [17] | 16 ( 0.2) | [16] |
| Any treatment-related TEAEs leading to study discontinuation | 5 (<0.1) | [5] | 2 (<0.1) | [2] |
| Any PIMMCs | 5 (<0.1) | [5] | 7 (<0.1) | [7] |
| Any AESIs: relevant to Covid-19 | 8 ( 0.1) | [12] | 22 ( 0.3) | [32] |

Abbreviations: AESIs = adverse event(s) of special interest; Covid-19 = coronavirus disease 2019; [E] = number of adverse events; MAAE = medically attended adverse event; PIMMC = potential immune-mediated medical conditions; SARS-CoV-2 = severe acute respiratory coronavirus 2; TEAE = treatment-emergent adverse event.

All counts exclude reactogenicity adverse events (selected preferred terms) where the event start date is on the day of either vaccination or within the 6 calendar days that follow either vaccination.

Any event that occurred after unblinding or after receipt of an approved or deployed SARS-CoV-2 vaccine was excluded from the analysis.

Unsolicited TEAEs reported for all participants who received at least one dose of NVX-CoV2373 or placebo during the entire study were predominantly mild or moderate in severity and occurred in 25.4% of participants receiving active vaccine and 20.8% of participants receiving placebo (Table S4). No unsolicited TEAE in the NVX-CoV2373 group occurred at a frequency that was one percentage point higher than in the placebo group. This analysis excludes reactogenicity TEAEs (selected preferred terms) with an event start date on the day of either vaccination or within 6 days after each vaccination.

Table S4. Summary of Unsolicited Treatment-Emergent Adverse Events (Excluding Reactogenicity Adverse Events) Reported in ≥1% of Participants in Any Group (Safety Analysis Set)

| **System Organ Class  Preferred Term** | **NVX-CoV2373**  **N=7569 n (%)** | **Placebo N=7570 n (%)** |
| --- | --- | --- |
| Any TEAE | 1925 (25.4) | 1574 (20.8) |
| General disorders and administration site conditions | 461 ( 6.1) | 223 ( 2.9) |
| Pain | 93 ( 1.2) | 25 ( 0.3) |
| Nervous system disorders | 325 ( 4.3) | 313 ( 4.1) |
| Headache | 109 ( 1.4) | 131 ( 1.7) |
| Lethargy | 77 ( 1.0) | 32 ( 0.4) |
| Respiratory, thoracic, and mediastinal disorders | 305 ( 4.0) | 316 ( 4.2) |
| Oropharyngeal pain | 114 ( 1.5) | 116 ( 1.5) |
| Rhinorrhea | 70 ( 0.9) | 92 ( 1.2) |
| Cough | 66 ( 0.9) | 87 ( 1.1) |
| Infections and infestations | 282 ( 3.7) | 328 ( 4.3) |
| Musculoskeletal and connective tissue disorders | 219 ( 2.9) | 203 ( 2.7) |
| Gastrointestinal disorders | 217 ( 2.9) | 188 ( 2.5) |
| Diarrhea | 82 ( 1.1) | 66 ( 0.9) |
| Skin and subcutaneous tissue disorders | 157 ( 2.1) | 110 ( 1.5) |
| Injury, poisoning, and procedural complications | 121 ( 1.6) | 92 ( 1.2) |
| Vascular disorders | 86 ( 1.1) | 53 ( 0.7) |
| Blood and lymphatic system disorders | 72 ( 1.0) | 61 ( 0.8) |

Abbreviations: MedDRA = *Medical Dictionary for Regulatory Activities;* TEAE = treatment-emergent adverse event.

Overall includes TEAE recorded during the entire study duration. If any solicited AE extended beyond 6 days after vaccination (toxicity grade ≥1), then it was recorded as an unsolicited TEAE with the start date as the 7th day following the relevant study vaccination and followed to resolution. A TEAE is defined as any event not present before exposure to study vaccination or any event already present that worsened in intensity after exposure. At each level of subject summarization, a participant is counted once if the participant reported one or more events.

n represents the number of participants at each level of summarization. Percentages were based on the number of participants in the safety analysis set within each treatment and total.

Adverse events were coded using MedDRA, version 23.1.

System organ class is displayed in descending order of frequency for the total column and then alphabetically.

Within class, preferred term is displayed in descending order of frequency for total and then alphabetically.

#### Efficacy Against UK Variant

A two-dose regimen of NVX-CoV2373 given 21 days apart was found to be safe and 89.7% effective against symptomatic Covid-19. The timing of accumulated cases in this study allowed for a post hoc assessment of vaccine efficacy against the prototype strain and the B.1.1.7 variant, which is now circulating widely outside of the UK and is soon expected to be the most prominent strain in the United States.^1^ This variant is known to be more transmissible and to be associated with a higher case fatality rate than previous strains,^1^ emphasizing the need for an effective vaccine. High vaccine efficacy was demonstrated against the prototype strain (96.4%) and the B.1.1.7 variant (86.3%) (Table S5). This is the first vaccine to demonstrate high vaccine efficacy (86.3%) against the B.1.1.7 variant in a phase 3 trial.

| Table S5. Vaccine Efficacy Against PCR-Confirmed Symptomatic Mild, Moderate, or Severe  Covid-19 With an Onset at Least 7 Days After Second Study Vaccination in Serologically Negative Adult Participants by SARS-CoV-2 Strain (PP-EFF Analysis Set) | | |
| --- | --- | --- |
| Parameter | Final Analysis | |
|  | NVX-CoV2373  N=7020 | Placebo  N=7019 |
| **UK (Kent) Variant Strain (B.1.1.7)** | | |
| Participants in PP-EFF | 7019^*^ | 7009^*^ |
| Participants with first occurrence of event, n (%) | 8 (0.1) | 58 (0.8) |
| Severity of first occurrence, n (%) | | |
| Mild | 1 (<0.1) | 15 (0.2) |
| Moderate | 7 (<0.1) | 39 (0.6) |
| Severe | 0 | 4 (<0.1) |
| Median surveillance time (days) | 56 | 55 |
| Log-linear model using modified Poisson regression | | |
| Mean disease incidence rate per year in 1000 people | 4.94 | 36.11 |
| 95% CI | 2.33, 10.48 | 23.15, 56.32 |
| Relative risk | 0.137 | |
| 95% CI | 0.065, 0.287 | |
| Vaccine efficacy (%) | 86.3 | |
| 95% CI | 71.3, 93.5 | |
| **Ancestral (Wuhan) Strain** | | |
| Participants in PP-EFF | 7019^*^ | 7009^*^ |
| Participants with first occurrence of event, n (%) | 1 (<0.1) | 28 (0.4) |
| Severity of first occurrence, n (%) | | |
| Mild | 0 | 9 (0.1) |
| Moderate | 1 (<0.1) | 18 (0.3) |
| Severe | 0 | 1 (<0.1) |
| Median surveillance time (days) | 56 | 56 |
| Log-linear model using modified Poisson regression | | |
| Mean disease incidence rate per year in 1000 people | 0.43 | 12.15 |
| 95% CI | 0.05, 3.79 | 4.23, 34.92 |
| Relative risk | 0.036 | |
| 95% CI | 0.005, 0.262 | |
| Vaccine efficacy (%) | 96.4 | |
| 95% CI | 73.8, 99.5 | |
| Abbreviations: CI = confidence interval; Covid-19 = coronavirus disease 2019; NVX CoV2373 = SARS-CoV-2 rS (5 μg) + Matrix-M1 adjuvant (50 μg); PCR = polymerase chain reaction; PP = per-protocol; SARS-CoV-2 = severe acute respiratory syndrome coronavirus 2; SARS-CoV-2 rS = severe acute respiratory syndrome coronavirus 2 recombinant nanoparticle spike protein vaccine.  *Excludes 1 participant in the NVX-CoV2373 group and 10 participants in the placebo group who had no sequence data for either SARS-CoV-2 strain. | | |

#### Demographics: Intention-to-Treat Population

**Table S6. Demographics and Baseline Characteristics (Intention-to-Treat Analysis Set)**

| **Parameter** | **NVX-CoV2373**  **N=7569** | **Placebo**  **N=7570** | **Total**  **N=15,139** |
| --- | --- | --- | --- |
| Age, yr  Median  Range | 55.0  18, 84 | 55.0  18, 84 | 55.0  18, 84 |
| Age group, n (%)  18-64 yr  ≥65 yr | 5503 (72.7)  2066 (27.3) | 5511 (72.8)  2059 (27.2) | 11014 (72.8)  4125 (27.2) |
| Sex, n (%)  Male  Female | 3890 (51.4)  3679 (48.6) | 3918 (51.8)  3652 (48.2) | 7808 (51.6)  7331 (48.4) |
| Race or ethnic group, n (%)  White  Black or African American  Asian  American Indian or Alaska native  Native Hawaiian or other Pacific Islander  Multiple  Not reported  Other  Missing  Hispanic or Latinx | 7127 (94.2)  31 (0.4)  230 (3.0)  5 (<0.1)  1 (<0.1)  75 (1.0)  91 (1.2)  5 (<0.1)  4  69 (0.9) | 7153 (94.5)  29 (0.4)  232 (3.1)  0  0  61 (0.8)  85 (1.1)  6 (<0.1)  4  56 (0.7) | 14280 (94.3)  60 (0.4)  462 (3.1)  5 (<0.1)  1 (<0.1  136 (0.9)  176 (1.2)  11 (<0.1)  8  125 (0.8) |
| SARS-CoV-2 serostatus, n (%)  Negative  Positive  Missing | 7180 (94.9)  330 (4.4)  59 | 7182 (94.9)  313 (4.1)  75 | 14362 (94.9)  643 (4.2)  134 |
| BMI, kg/m^2^, n (%)  ≥30.0: obese | 344 (4.5) | 353 (4.7) | 697 (4.6) |
| Comorbidity status*  Yes  No | 3368 (44.5)  4201 (55.5) | 3399 (44.9)  4171 (55.1) | 6767 (44.7)  8372 (55.3) |

SD = standard deviation. Body mass index (BMI) is calculated as weight (kg) divided by squared height (m).

Percentages are based on per-protocol efficacy analysis set within each treatment and overall.

*Comorbid subjects are those identified who have at least one of the comorbid conditions reported as a medical history or have a screening BMI value greater than 30 kg/m^2^.

**Table S7. Summary of Unsolicited Serious Treatment-Emergent Adverse Events (Safety Analysis Set)**

| **System Organ Class  Preferred Term  Severity** | **NVX-CoV2373**  **N=7569 n (%)** | **Placebo N=7570 n (%)** | **Total N=15139 n (%)** |
| --- | --- | --- | --- |
| Any Serious TEAE | 44 ( 0.6) | 44 ( 0.6) | 88 ( 0.6) |
| Infections and infestations | 6 (<0.1) | 11 ( 0.1) | 17 ( 0.1) |
| Covid-19 pneumonia | 1 (<0.1) | 3 (<0.1) | 4 (<0.1) |
| Appendicitis | 1 (<0.1) | 2 (<0.1) | 3 (<0.1) |
| Covid-19 | 2 (<0.1) | 0 | 2 (<0.1) |
| Pneumonia | 0 | 2 (<0.1) | 2 (<0.1) |
| Appendicitis perforated | 1 (<0.1) | 0 | 1 (<0.1) |
| Bacterial sepsis | 0 | 1 (<0.1) | 1 (<0.1) |
| Diverticulitis | 0 | 1 (<0.1) | 1 (<0.1) |
| Epiglottitis | 0 | 1 (<0.1) | 1 (<0.1) |
| Gastroenteritis | 0 | 1 (<0.1) | 1 (<0.1) |
| Intestinal gangrene | 1 (<0.1) | 0 | 1 (<0.1) |
| Otitis externa | 0 | 1 (<0.1) | 1 (<0.1) |
| Pharyngeal abscess | 0 | 1 (<0.1) | 1 (<0.1) |
| Postoperative wound infection | 1 (<0.1) | 0 | 1 (<0.1) |
| Wound infection | 1 (<0.1) | 0 | 1 (<0.1) |
| Injury, poisoning, and procedural complications | 11 ( 0.1) | 6 (<0.1) | 17 ( 0.1) |
| Ankle fracture | 3 (<0.1) | 0 | 3 (<0.1) |
| Femoral neck fracture | 0 | 3 (<0.1) | 3 (<0.1) |
| Cervical vertebral fracture | 1 (<0.1) | 0 | 1 (<0.1) |
| Fall | 0 | 1 (<0.1) | 1 (<0.1) |
| Femur fracture | 1 (<0.1) | 0 | 1 (<0.1) |
| Intentional overdose | 1 (<0.1) | 0 | 1 (<0.1) |
| Joint dislocation | 1 (<0.1) | 0 | 1 (<0.1) |
| Limb injury | 1 (<0.1) | 0 | 1 (<0.1) |
| Overdose | 0 | 1 (<0.1) | 1 (<0.1) |
| Poisoning deliberate | 1 (<0.1) | 0 | 1 (<0.1) |
| Radius fracture | 1 (<0.1) | 0 | 1 (<0.1) |
| Skin laceration | 1 (<0.1) | 0 | 1 (<0.1) |
| Ulna fracture | 1 (<0.1) | 0 | 1 (<0.1) |
| Wrist fracture | 0 | 1 (<0.1) | 1 (<0.1) |
| Cardiac disorders | 7 (<0.1) | 5 (<0.1) | 12 (<0.1) |
| Atrioventricular block  complete | 1 (<0.1) | 1 (<0.1) | 2 (<0.1) |
| Acute coronary syndrome | 1 (<0.1) | 0 | 1 (<0.1) |
| Acute myocardial infarction | 1 (<0.1) | 0 | 1 (<0.1) |
| Angina pectoris | 0 | 1 (<0.1) | 1 (<0.1) |
| Arrhythmia | 0 | 1 (<0.1) | 1 (<0.1) |
| Atrial fibrillation | 0 | 1 (<0.1) | 1 (<0.1) |
| Atrial flutter | 0 | 1 (<0.1) | 1 (<0.1) |
| Atrial tachycardia | 1 (<0.1) | 0 | 1 (<0.1) |
| Cardiac failure acute | 1 (<0.1) | 0 | 1 (<0.1) |
| Myocardial infarction | 1 (<0.1) | 0 | 1 (<0.1) |
| Myocarditis | 1 (<0.1) | 0 | 1 (<0.1) |
| Palpitations | 1 (<0.1) | 0 | 1 (<0.1) |
| Neoplasms benign, malignant, and  unspecified (incl cysts and  polyps) | 5 (<0.1) | 5 (<0.1) | 10 (<0.1) |
| Breast cancer | 2 (<0.1) | 0 | 2 (<0.1) |
| Adenocarcinoma of appendix | 0 | 1 (<0.1) | 1 (<0.1) |
| Bladder cancer | 1 (<0.1) | 0 | 1 (<0.1) |
| Glioblastoma | 0 | 1 (<0.1) | 1 (<0.1) |
| Intraductal proliferative  breast lesion | 0 | 1 (<0.1) | 1 (<0.1) |
| Lung neoplasm malignant | 1 (<0.1) | 0 | 1 (<0.1) |
| Ovarian cancer | 0 | 1 (<0.1) | 1 (<0.1) |
| Squamous cell carcinoma of skin | 1 (<0.1) | 0 | 1 (<0.1) |
| Squamous cell carcinoma of the tongue | 0 | 1 (<0.1) | 1 (<0.1) |
| Nervous system disorders | 5 (<0.1) | 3 (<0.1) | 8 (<0.1) |
| Migraine | 3 (<0.1) | 0 | 3 (<0.1) |
| Lumbar radiculopathy | 0 | 1 (<0.1) | 1 (<0.1) |
| Migraine with aura | 0 | 1 (<0.1) | 1 (<0.1) |
| Presyncope | 1 (<0.1) | 0 | 1 (<0.1) |
| Sciatica | 1 (<0.1) | 0 | 1 (<0.1) |
| Transient ischemic attack | 0 | 1 (<0.1) | 1 (<0.1) |
| Gastrointestinal disorders | 2 (<0.1) | 4 (<0.1) | 6 (<0.1) |
| Abdominal pain lower | 1 (<0.1) | 0 | 1 (<0.1) |
| Ascites | 0 | 1 (<0.1) | 1 (<0.1) |
| Gastro-esophageal reflux  disease | 1 (<0.1) | 0 | 1 (<0.1) |
| Intestinal perforation | 0 | 1 (<0.1) | 1 (<0.1) |
| Obstructive pancreatitis | 0 | 1 (<0.1) | 1 (<0.1) |
| Small intestinal obstruction | 0 | 1 (<0.1) | 1 (<0.1) |
| Upper gastrointestinal  hemorrhage | 1 (<0.1) | 0 | 1 (<0.1) |
| Renal and urinary disorders | 2 (<0.1) | 3 (<0.1) | 5 (<0.1) |
| Acute kidney injury | 1 (<0.1) | 1 (<0.1) | 2 (<0.1) |
| Nephrolithiasis | 0 | 1 (<0.1) | 1 (<0.1) |
| Urethral dilatation | 0 | 1 (<0.1) | 1 (<0.1) |
| Urinary retention | 1 (<0.1) | 0 | 1 (<0.1) |
| Metabolism and nutrition  disorders | 1 (<0.1) | 3 (<0.1) | 4 (<0.1) |
| Dehydration | 1 (<0.1) | 0 | 1 (<0.1) |
| Diabetic ketoacidosis | 0 | 1 (<0.1) | 1 (<0.1) |
| Diabetic ketosis | 0 | 1 (<0.1) | 1 (<0.1) |
| Hypoalbuminemia | 0 | 1 (<0.1) | 1 (<0.1) |
| Respiratory, thoracic, and  mediastinal disorders | 1 (<0.1) | 3 (<0.1) | 4 (<0.1) |
| Pulmonary embolism | 1 (<0.1) | 2 (<0.1) | 3 (<0.1) |
| Epistaxis | 0 | 1 (<0.1) | 1 (<0.1) |
| Blood and lymphatic system  disorders | 0 | 3 (<0.1) | 3 (<0.1) |
| Anemia | 0 | 1 (<0.1) | 1 (<0.1) |
| Hemolytic anemia | 0 | 1 (<0.1) | 1 (<0.1) |
| Iron deficiency anemia | 0 | 1 (<0.1) | 1 (<0.1) |
| General disorders and administration site conditions | 2 (<0.1) | 0 | 2 (<0.1) |
| Mass | 1 (<0.1) | 0 | 1 (<0.1) |
| Non-cardiac chest pain | 1 (<0.1) | 0 | 1 (<0.1) |
| Hepatobiliary disorders | 1 (<0.1) | 1 (<0.1) | 2 (<0.1) |
| Cholecystitis | 1 (<0.1) | 0 | 1 (<0.1) |
| Liver injury | 0 | 1 (<0.1) | 1 (<0.1) |
| Reproductive system and breast disorders | 2 (<0.1) | 0 | 2 (<0.1) |
| Endometriosis | 1 (<0.1) | 0 | 1 (<0.1) |
| Vaginal prolapse | 1 (<0.1) | 0 | 1 (<0.1) |
| Vascular disorders | 0 | 2 (<0.1) | 2 (<0.1) |
| Hypertension | 0 | 1 (<0.1) | 1 (<0.1) |
| Peripheral ischemia | 0 | 1 (<0.1) | 1 (<0.1) |
| Investigations | 0 | 1 (<0.1) | 1 (<0.1) |
| Blood pressure systolic  increased | 0 | 1 (<0.1) | 1 (<0.1) |
| Musculoskeletal and connective  tissue disorders | 0 | 1 (<0.1) | 1 (<0.1) |
| Osteoarthritis | 0 | 1 (<0.1) | 1 (<0.1) |
| Pregnancy, puerperium, and  perinatal conditions | 1 (<0.1) | 0 | 1 (<0.1) |
| Abortion spontaneous | 1 (<0.1) | 0 | 1 (<0.1) |
| Surgical and medical procedures | 1 (<0.1) | 0 | 1 (<0.1) |
| Cholecystectomy | 1 (<0.1) | 0 | 1 (<0.1) |
| UNCODED | 1 (<0.1) | 1 (<0.1) | 2 (<0.1) |

Vaccination Dose 1 includes serious TEAE with start date on or after first vaccination dose to end of study or second vaccination dose, whichever occurs earlier. Vaccination Dose 2 includes serious TEAE with start date on or after second vaccination dose to end of study. Overall includes TEAE recorded during the entire study duration. If any solicited AE extended beyond 6 days after vaccination (toxicity grade ≥1), then it is recorded as an unsolicited AE with the start date the 7th day following the relevant study vaccination and followed to resolution.

n represents the number of subjects at each level of summarization. Percentages are based on the number of subjects in the safety analysis set within each treatment and total.

SAEs were coded using MedDRA, version 23.1.

System organ class is displayed in descending order of frequency for the total column and then alphabetically.

Within class, preferred term is displayed in descending order of frequency for total and then alphabetically.

*B1.1.1.7 Variant*

The B.1.1.7 variant was first detected in the UK in September 2020 and has since been found in more than 100 countries. It has 23 mutations in its genetic code; some of these changes have increased its ability to spread. It is estimated that the B.1.1.7 variant is 40% to 70% more transmissible than previously dominant circulating coronavirus variants.^2^ Figure S1 shows the growth of the B.1.1.7 variant over the course of the current study in the UK.

**Figure S1. Evolution of Covid-19 Variants During the Endpoint Period.**
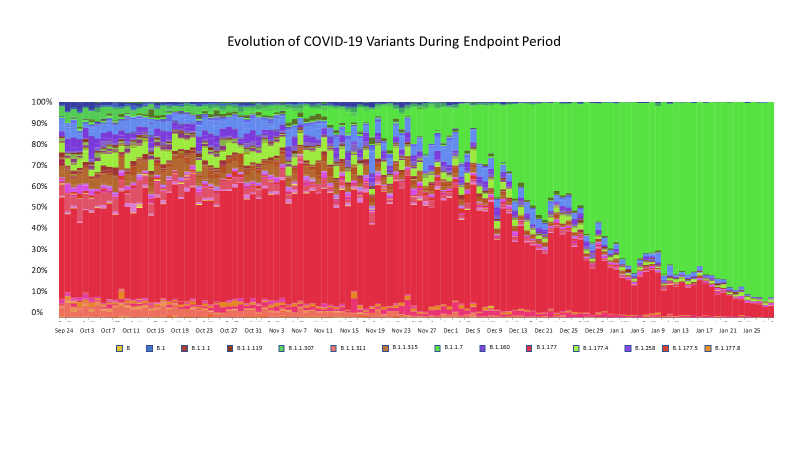

### Microreact has been developed by the [Centre for Genomic Pathogen Surveillance](https://urldefense.com/v3/__http:/www.pathogensurveillance.net/__;!!IfJP2Nwhk5Z0yJ43lA!fM99cBFzkIadivaYJ4jlfyPiQUB4M6xpV0y4yvOIMlex0Lu6x317rJlW2yYfDQl7$) at Imperial College London and the Wellcome Genome Campus. Argimón S et al, 2016.^3^

### References

1. Centers for Disease Control and Prevention. About variants of the virus that causes Covid-19. Updated April 2, 2021. <https://www.cdc.gov/coronavirus/2019-ncov/transmission/variant.html>. Accessed April 7, 2021.
2. Challen R, Brooks-Pollock E, Read JM, et al. Risk of mortality in patients infected with SARS-CoV-2 variant of concern 202012/1: matched cohort study. BMJ 2021;372:n579. <http://dx.doi.org/10.1136/bmj.n579>.
3. Argimón S, Abudahab K, Goater RJE, et al. Microreact: visualizing and sharing data for genomic epidemiology and phylogedoography. Microb Genom 2016;2:e000093. <http://dx.doi.org/10.1099/mgen.0.000093>.
