## Supplementary material for "Efficacy of the NVX-CoV2373 Covid-19 Vaccine Against the B.1.1.7 Variant": Trial Approval letter

Health Research Authority
**North West - Greater Manchester Central Research Ethics Committee**
3rd Floor
Barlow House
4 Minshull Street
Manchester
M1 3DZ

**Please note: This is the favourable opinion of the REC only and does not allow you to start your study at NHS sites in England until you receive HRA Approval**

21 September 2020

Professor Paul Heath
Vaccine Institute, St George's University Hospitals NHS Foundation Trust
Blackshaw Road, Tooting
London
SW17 0QT

Dear Professor Heath

| | |
|---|---|
| **Study title:** | **A Phase 3, Randomised, Observer-Blinded, Placebo-Controlled Trial to Evaluate the Efficacy and Safety of a SARS-CoV-2 Recombinant Spike Protein Nanoparticle Vaccine (SARS-CoV-2 rS) with Matrix-M1â„¢ Adjuvant in Adult Participants 18-84 Years of Age in the United Kingdom** |
| **REC reference:** | **20/NW/0399** |
| **Protocol number:** | **2019nCoV-302** |
| **EudraCT number:** | **2020-004123-16** |
| **IRAS project ID:** | **288793** |

Thank you for your email of 21 September 2020, responding to the Committee's request for further information on the above research. The further information has been considered on behalf of the Committee by the Chair.

### Confirmation of ethical opinion

On behalf of the Committee, I am pleased to confirm a favourable ethical opinion for the above research on the basis described in the application form, protocol and supporting documentation as revised, subject to the conditions specified below.

Confirmation of Capacity and Capability (in England, Northern Ireland and Wales) or NHS management permission (in Scotland) should be sought from all NHS organisations involved in the study in accordance with NHS research governance arrangements. Each NHS organisation must confirm through the signing of agreements and/or other documents that it has given permission for the research to proceed (except where explicitly specified otherwise).

Guidance on applying for HRA and HCRW Approval (England and Wales)/ NHS permission for research is available in the Integrated Research Application System.

For non-NHS sites, site management permission should be obtained in accordance with the procedures of the relevant host organisation.

Sponsors are not required to notify the Committee of management permissions from host organisations

Registration of Clinical Trials

It is a condition of the REC favourable opinion that **all clinical trials are registered** on a publicly accessible database. For this purpose, 'clinical trials' are defined as the first four project categories in IRAS project filter question 2. Registration is a legal requirement for clinical trials of investigational medicinal products (CTIMPs), except for phase I trials in healthy volunteers (these must still register as a condition of the REC favourable opinion).

Registration should take place as early as possible and within six weeks of recruiting the first research participant at the latest. Failure to register is a breach of these approval conditions, unless a deferral has been agreed by or on behalf of the Research Ethics Committee ( see here for more information on requesting a deferral: https://www.hra.nhs.uk/planning-and-improving-research/research-planning/research-registration-research-project-identifiers/

As set out in the UK Policy Framework, research sponsors are responsible for making information about research publicly available before it starts e.g. by registering the research project on a publicly accessible register. Further guidance on registration is available at: https://www.hra.nhs.uk/planning-and-improving-research/research-planning/transparency-responsibilities/

You should notify the REC of the registration details. We will audit these as part of the annual progress reporting process.

Publication of Your Research Summary

We will publish your research summary for the above study on the research summaries section of our website, together with your contact details, no earlier than three months from the date of this favourable opinion letter.

Should you wish to provide a substitute contact point, make a request to defer, or require further information, please visit: https://www.hra.nhs.uk/planning-and-improving-research/application-summaries/research-summaries/

**N.B. If your study is related to COVID-19 we will aim to publish your research summary within 3 days rather than three months**.

During this public health emergency, it is vital that everyone can promptly identify all relevant research related to COVID-19 that is taking place globally. If you haven't already done so, please register your study on a public registry as soon as possible and provide the HRA with the registration detail, which will be posted alongside other information relating to your project. We are also asking sponsors not to request deferral of publication of research summary for any projects relating to COVID-19. In addition, to facilitate finding and extracting studies related to COVID-19 from public databases, please enter the WHO official acronym for the coronavirus disease (COVID-19) in the full title of your study. Approved COVID-19 studies can be found at: https://www.hra.nhs.uk/covid-19-research/approved-covid-19-research/

Clinical trial authorisation must be obtained from the Medicines and Healthcare products Regulatory Agency (MHRA).

**It is the responsibility of the sponsor to ensure that all the conditions are complied with before the start of the study or its initiation at a particular site (as applicable).**

**After ethical review: Reporting requirements**

The attached document "After ethical review – guidance for researchers" gives detailed guidance on reporting requirements for studies with a favourable opinion, including:

- Notifying substantial amendments
- Adding new sites and investigators
- Notification of serious breaches of the protocol
- Progress and safety reports
- Notifying the end of the study, including early termination of the study
- Final report

The latest guidance on these topics can be found at https://www.hra.nhs.uk/approvals-amendments/managing-your-approval/.

**Ethical review of research sites**

NHS/HSC sites

The favourable opinion applies to all NHS/HSC sites listed in the application subject to confirmation of Capacity and Capability (in England, Northern Ireland and Wales) or management permission (in Scotland) being obtained from the NHS/HSC R&D office prior to the start of the study (see "Conditions of the favourable opinion" below).

Non-NHS/HSC sites

I am pleased to confirm that the favourable opinion applies to any non-NHS/HSC sites listed in the application, subject to site management permission being obtained prior to the start of the study at the site.

**Approved documents**

The final list of documents reviewed and approved by the Committee is as follows:

| *Document* | *Version* | *Date* |
|---|---|---|
| Copies of advertisement materials for research participants [NVX-040CTM_2019nCoV-302_UK_NIHR_BeAPartOfResearch_Approved_Vaccine_website_text_V1.0_15Sep2020] | 1.0 | 15 September 2020 |
| Copies of advertisement materials for research participants [NVX-041CTM_2019nCoV-302_UK_NIHR_BeAPartOfResearch_Email_Wording_V1.0_15Sep2020] | 1.0 | 15 September 2020 |
| Copies of advertisement materials for research participants [NVX-042CTM_2019nCoV-302_UK_NIHR_BeAPartOfResearch_website_text_V1.0_15Sep2020] | 1.0 | 15 September 2020 |
| Copies of advertisement materials for research participants [NVX-039CTM_2019nCoV-302 Newsletter Bulletin Social Media text ALL SITES 10Sep2020] | 1.0 | 10 September 2020 |
| Covering letter on headed paper [Cover letter] | | 11 September 2020 |
| Evidence of Sponsor insurance or indemnity (non- NHS Sponsors only) [Insurance certificate] | Final | 27 August 2020 |
| GP/consultant information sheets or letters [GP Letter] | 1.0 | 31 August 2020 |
| GP/consultant information sheets or letters [NVX-037CTM_2019nCoV-302_Phase3_MD to MD letter UK Version 1.0_09Sep2020] | 1.0 | 09 September 2020 |
| Instructions for use of medical device [Digital thermometer instructions] | 1.0 | 10 September 2020 |
| Instructions for use of medical device [Digital Thermometer description] | Final | 03 April 2020 |
| Instructions for use of medical device [NHS guidelines] | 1.0 | 10 September 2020 |
| Investigator's brochure / IMP Dossier [Investigator Brochure] | 3.0 | 13 August 2020 |
| IRAS Application Form [IRAS_Form_11092020] | | 11 September 2020 |
| IRAS Checklist XML [Checklist_11092020] | | 11 September 2020 |
| Letter of invitation to participant ]NVX-038CTM_2019nCoV-302_Participant Invitation Letter v 1.0 09Sep2020] | 1.0 | 09 September 2020 |
| Other [Delegation of authority] | Final | 09 April 2020 |
| Other [EU legal Rep] | | 11 September 2020 |
| Other [Response cover letter] | | 17 September 2020 |
| Participant information sheet (PIS) [Novavax 2019nCoV-302_Anti S protein serology sub-study and neutralisation assay substudy_UK_ICF_v2.1.0 15Sep2020_Clean] | 2.1.0 | 15 September 2020 |
| Participant information sheet (PIS) [Novavax 2019nCoV-302_Cell Mediated Testing_UKICF_v2.1.0_15Sep2020_Clean] | 2.1.0 | 15 September 2020 |
| Participant information sheet (PIS) [Novavax 2019nCoV-302_Influenza Co-ad Substudy_UK_ICF_v1.2.0 15Sep2020_Clean] | 1.2.0 | 15 September 2020 |
| Participant information sheet (PIS) [Novavax 2019nCoV-302_Solicited_AE_UK_ICF_v1.2.0_15Sep2020_Clean] | 1.2.0 | 15 September 2020 |
| Participant information sheet (PIS) [Novavax_2019nCoV-302_Main_UK_ICF_v2.2.0_14Sep2020_Clean] | 2.2.0 | 14 September 2020 |
| Research protocol or project proposal [Protocol] | 1.1 | 17 September 2020 |
| Response to Request for Further Information | | 21 September 2020 |
| Sample diary card/patient card [Patient card] | 1.0 | 07 September 2020 |

| | | |
|---|---|---|
| Sample diary card/patient card [e-Diary participant text] | 1.0 | 10 September 2020 |
| Summary CV for Chief Investigator (CI) [CI CV] | 03Sep2020 | 03 September 2020 |
| Summary of product characteristics (SmPC) [SmPC for placebo] | | 01 October 2018 |

**Statement of compliance**

This Committee is recognised by the United Kingdom Ethics Committee Authority under the Medicines for Human Use (Clinical Trials) Regulations 2004 and is authorised to carry out the ethical review of clinical trials of investigational medicinal products.

The Committee is fully compliant with the Regulations as they relate to ethics committees and the conditions and principles of good clinical practice.

The Committee is constituted in accordance with the Governance Arrangements for Research Ethics Committees and complies fully with the Standard Operating Procedures for Research Ethics Committees in the UK.

**User Feedback**

The Health Research Authority is continually striving to provide a high quality service to all applicants and sponsors. You are invited to give your view of the service you have received and the application procedure. If you wish to make your views known please use the feedback form available on the HRA website: http://www.hra.nhs.uk/about-the-hra/governance/quality-assurance/

**HRA Learning**

We are pleased to welcome researchers and research staff to our HRA Learning Events and online learning opportunities– see details at: https://www.hra.nhs.uk/planning-and-improving-research/learning/

**IRAS project ID: 288793      Please quote this number on all correspondence**

With the Committee's best wishes for the success of this project.

Yours sincerely

Elaine Hutchings

pp
**Dr George Gkimpas**
**Chair**

*Enclosure:* "After ethical review – guidance for researchers"

*Copy to:* Ms Ellie Pugh-Tymms
