## Supplementary material for "Efficacy of the NVX-CoV2373 Covid-19 Vaccine Against the B.1.1.7 Variant": Redacted protocol and SAP

This trial protocol has been provided by the authors to give readers additional information about their work.

Protocol for: Heath PT, Galiza EP, Baxter DN, et al. Efficacy of the NVX-CoV2373 Covid-19 Vaccine Against the B.1.1.7 Variant. N Engl J Med. DOI:

This supplement contains the following items:

1. Original protocol v1.0 (redacted) and final protocol v.4.0 (redacted)
2. Table of Amendments to protocol
3. Original statistical analysis plan v1.0 (redacted) and final statistical analysis plan v4.0 (redacted)
4. Table of Amendments to statistical analysis plan

### CLINICAL STUDY PROTOCOL

### A PHASE 3, RANDOMISED, OBSERVER-BLINDED, PLACEBO-CONTROLLED TRIAL TO EVALUATE THE EFFICACY AND SAFETY OF A SARS-COV-2 RECOMBINANT SPIKE PROTEIN NANOPARTICLE VACCINE (SARS-COV-2 RS) WITH MATRIX-M1™ ADJUVANT IN ADULT PARTICIPANTS 18-84 YEARS OF AGE IN THE UNITED KINGDOM

| | |
|---|---|
| **Investigational Materials:** | SARS-CoV-2 rS with Matrix-M1™ adjuvant |
| **Protocol Number:** | 2019nCoV-302 |
| **EudraCT Number:** | 2020-004123-16 |
| **Sponsor:** | Novavax, Inc.<br>21 Firstfield Road<br>Gaithersburg, MD 20878<br>United States |
| **Responsible Clinical Operations Manager:** | [redacted] Clinical Operations<br>Telephone: [redacted] |
| **Responsible Clinical Development Lead:** | [redacted] Clinical Development<br>Telephone: [redacted] |
| **Chief Investigator:** | [redacted]<br>Telephone: [redacted] |
| **Version – Date:** | Version 1.0 – 24 August 2020 |
| **Prior Versions:** | Not applicable |

### CONFIDENTIAL

The information in this document is considered privileged and confidential by Novavax, Inc. and may not be disclosed to others except to the extent necessary to obtain Independent Ethics Committee (IEC) approval and informed consent, or as required by national and local laws. Persons to whom this information is disclosed must be informed that this information is privileged and confidential and that it should not be further disclosed.

The study will be conducted according to the International Council for Harmonisation of Technical Requirements for Pharmaceuticals for Human Use Guideline E6(R2): Good Clinical Practice.

### SIGNATURE PAGE

**PROTOCOL TITLE:** A Phase 3, Randomised, Observer-Blinded, Placebo-Controlled Trial to Evaluate the Efficacy and Safety of a SARS-CoV-2 Recombinant Spike Protein Nanoparticle Vaccine (SARS-CoV-2 rS) with Matrix-M1™ Adjuvant in Adult Participants 18-84 Years of Age in the United Kingdom

**PROTOCOL NUMBER:** 2019-nCoV-302

**EUDRACT NUMBER:** 2020-004123-16

[redacted] 25-Aug-20 | 10:30 EDT

[redacted]

[redacted] Clinical Developme[redacted]

Novavax, Inc.

25-Aug-20 | 10:22 EDT

[redacted]

[redacted] Clinical Operation[redacted]

Novavax, Inc.

[redacted]

25-AUG-20

[redacted]

Date

### INVESTIGATOR PROTOCOL AGREEMENT PAGE

I agree to conduct the study as outlined in the protocol titled "A Phase 3, Randomised, Observer-Blinded, Placebo-Controlled Trial to Evaluate the Efficacy and Safety of a SARS-CoV-2 Recombinant Spike Protein Nanoparticle Vaccine (SARS-CoV-2 rS) with Matrix-M1™ Adjuvant in Adult Participants 18-84 Years of Age in the United Kingdom" in accordance with all guidelines, including International Council for Harmonisation of Technical Requirements for Pharmaceuticals for Human Use guidelines, and all applicable government regulations. I have read and understand all sections of the protocol.

Signature of Investigator

Date

Printed Name of Investigator

### TABLE OF CONTENTS

**TITLE PAGE** 1
**SIGNATURE PAGE** 3
**INVESTIGATOR PROTOCOL AGREEMENT PAGE** 4
**TABLE OF CONTENTS** 5
**LIST OF TABLES** 9
**LIST OF FIGURES** 10
**PROTOCOL SYNOPSIS** 11
**1. INTRODUCTION** 26
1.1 BACKGROUND 26
1.2 NON-CLINICAL EXPERIENCE 27
1.2.1 Nonclinical Data from SARS-CoV-2 Spike Protein Constructs that Support SARS-CoV-2 rS Development 27
1.2.2 Nonclinical Data from Other Baculovirus-Sf9-Produced Nanoparticle Vaccines that Support SARS-CoV-2 rS Development 28
1.3 CLINICAL EXPERIENCE 29
1.4 RATIONALE FOR STUDY 29
1.5 RATIONALE FOR DOSE SELECTION 30
1.6 BENEFIT-RISK ASSESSMENT 31
**2. STUDY OBJECTIVES AND ENDPOINTS** 33
2.1 STUDY OBJECTIVES 33
2.1.1 Primary Objective 33
2.1.2 Secondary Objectives 33
2.1.3 Exploratory Objective 33
2.2 STUDY ENDPOINTS 34
2.2.1 Primary Endpoints 34
2.2.2 Secondary Endpoints 36
2.2.3 Exploratory Endpoints 36
**3. STUDY DESIGN** 38
3.1 STUDY VACCINATION PAUSE RULES 41
3.2 SCHEDULE OF EVENTS (SOE) 42
**4. STUDY POPULATION** 45
4.1 INCLUSION CRITERIA 45
4.2 EXCLUSION CRITERIA 46
4.3 OTHER CONSIDERATIONS 48
4.4 WITHDRAWAL OF PARTICIPANTS FROM THE STUDY 49
4.4.1 Reasons for Withdrawal 49
4.4.2 Handling of Withdrawals 49

6.4.1.4 Assessment of Severity ... 71
6.4.1.5 Assessment of Causality ... 71
6.4.1.6 Follow-up of Adverse Events ... 72
6.4.2 Vital Sign Measurements ... 72
6.4.3 Physical Examinations ... 72
6.4.4 Safety Monitoring ... 72
**7. STATISTICAL ANALYSIS PLANS ... 73**
7.1 SAMPLE SIZE CALCULATIONS ... 73
7.2 ANALYSIS SETS ... 75
7.3 STATISTICAL ANALYSIS ... 76
7.3.1 Efficacy Analyses ... 77
7.3.2 Immunogenicity Analysis and Correlates of Risk ... 78
7.3.3 Safety Analyses ... 78
7.3.3.1 Safety: Study Vaccine-Associated Enhanced Disease ... 79
7.4 HANDLING OF MISSING DATA ... 79
7.5 PLANNED ANALYSES PRIOR TO STUDY COMPLETION ... 79
**8. REFERENCE LIST ... 80**
**9. APPENDICES ... 82**
9.1 APPENDIX 1: LIST OF ABBREVIATIONS ... 82
9.2 APPENDIX 2: STUDY GOVERNANCE ... 84
9.2.1 Data Quality Assurance ... 84
9.2.2 Investigator Obligations ... 84
9.2.2.1 Confidentiality ... 84
9.2.2.2 Institutional Review ... 85
9.2.2.3 Participant Consent ... 85
9.2.2.4 Study Reporting Requirements ... 85
9.2.2.5 Financial Disclosure and Obligations ... 86
9.2.2.6 Investigator Documentation ... 86
9.2.2.7 Study Conduct ... 86
9.2.2.8 Case Report Forms and Source Documents ... 87
9.2.2.9 Adherence to Protocol ... 87
9.2.2.10 Reporting Adverse Events ... 87
9.2.2.11 Investigator's Final Report ... 88
9.2.2.12 Records Retention ... 88
9.2.2.13 Publications ... 88
9.2.3 Study Management ... 89
9.2.3.1 Monitoring ... 89
9.2.3.2 Management of Protocol Amendments and Deviations ... 89

### LIST OF FIGURES

Figure 1: Trial Schema ........................................ 39

### PROTOCOL SYNOPSIS

**PROTOCOL NO.:** 2019nCoV-302

**TITLE:** A Phase 3, Randomised, Observer-Blinded, Placebo-Controlled Trial to Evaluate the Efficacy and Safety of a SARS-CoV-2 Recombinant Spike Protein Nanoparticle Vaccine (SARS-CoV-2 rS) with Matrix-M1™ Adjuvant in Adult Participants 18-84 Years of Age in the United Kingdom

**STUDY PHASE:** Phase 3

**STUDY SITES:** Up to 18 regions across the United Kingdom (UK).

**OBJECTIVES:**

- **The primary objective is:**
  - To demonstrate the efficacy of SARS-CoV-2 rS with Matrix-M1 adjuvant in the prevention of virologically confirmed (by polymerase chain reaction [PCR]) to severe acute respiratory syndrome coronavirus 2 (SARS-CoV-2), symptomatic coronavirus disease 2019 (COVID-19), when given as a 2-dose vaccination regimen, as compared to placebo, in serologically negative (to SARS-CoV-2) adults.
- **The secondary objectives are:**
  - To demonstrate the efficacy of SARS-CoV-2 rS with Matrix-M1 adjuvant in the prevention of virologically confirmed [by PCR to SARS-CoV-2], symptomatic COVID-19, when given as a 2-dose vaccination regimen, as compared to placebo, in adults regardless of their serostatus at baseline.
  - To assess the efficacy of SARS-CoV-2 rS with Matrix-M1 adjuvant on SARS-CoV-2 seropositive adult participants requiring specific medical interventions as compared to placebo.
  - To assess the efficacy of SARS-CoV-2 rS with Matrix-M1 adjuvant on mild COVID-19 symptoms as compared to placebo.
  - To assess the efficacy of SARS-CoV-2 rS with Matrix-M1 adjuvant on occurrence of asymptomatic or undetected infections with SARS-CoV-2 as compared to placebo.
  - In a subset of adult participants, to evaluate the immunogenicity of SARS-CoV-2 rS with Matrix-M1 adjuvant as compared to placebo.
  - To evaluate safety in terms of serious adverse events (SAEs) and medically attended adverse events (MAAEs) in all adult participants during the entire study period.
  - To evaluate safety in terms of adverse events of special interest (AESI), which encompasses potential immune-mediated medical conditions (PIMMCs) and

AESIs relevant to COVID-19 including possible vaccine-enhanced disease, in all adult participants at any time after the first dose.

- In a subset of adult participants, to evaluate the safety and reactogenicity in terms of solicited local and systemic adverse events (AEs) for 7 days after each study vaccination, and in terms of unsolicited AEs for 21 days after first study vaccination and 28 days after second study vaccination.

- **The exploratory objectives are:**
  - In a subset of adult participants, to evaluate the safety and immunogenicity of SARS-CoV-2 rS with Matrix-M1 adjuvant when co-administered with a licensed seasonal influenza vaccine.

**ENDPOINTS**

- **The primary endpoints are**:

  There are 2 independent primary endpoints.

  - **FIRST PRIMARY ENDPOINT:** First occurrence of virologically confirmed (by PCR to SARS-CoV-2), symptomatic COVID-19 with onset at least 7 days after second study vaccination (e.g., Day 28) in serologically negative (to SARS-CoV-2) adult participants at baseline until the endpoint-driven efficacy analysis is triggered by the occurrence of a prespecified number of blinded endpoints.
  - **SECOND PRIMARY ENDPOINT:** First occurrence of virologically confirmed (by PCR to SARS-CoV-2), symptomatic moderate or severe COVID-19 with onset at least 7 days after second study vaccination (e.g., Day 28) in serologically negative (to SARS-CoV-2) adult participants at baseline until the endpoint-driven efficacy analysis is triggered by the occurrence of a prespecified number of blinded endpoints.

- **The secondary endpoints are**:
  - First occurrence of virologically confirmed (by PCR to SARS-CoV-2), symptomatic COVID-19, with onset at least 7 days after second study vaccination (e.g., Day 28) in adult participants regardless of their serostatus at baseline.
  - First occurrence of laboratory-confirmed (by PCR or serology to SARS-CoV-2) symptomatic or asymptomatic COVID-19 with onset at least 7 days after second study vaccination (e.g., Day 28) in adult participants regardless of their serostatus at baseline.
  - First occurrence of COVID-19 requiring hospitalisation, intensive care unit (ICU) admission or mechanical ventilation linked to any virologically confirmed (by PCR to SARS-CoV-2) COVID-19 with onset at least 7 days after second study vaccination (e.g., Day 28) in adult participants regardless of their serostatus at baseline.
  - First occurrence of virologically confirmed (by PCR to SARS-CoV-2), mild COVID-19 (with no progression to moderate or severe COVID-19 during the course of the COVID-19 episode) with onset at least 7 days after second study

vaccination (e.g., Day 28) in adult participants, regardless of their serostatus at baseline.

  - Analysis of antibodies binding to the SARS-CoV-2 spike (S) protein by enzyme-linked immunosorbent assay (ELISA) at Day 0 (baseline), Day 21 (21 days after first study vaccination), and Day 35 (14 days after second study vaccination).
  - The occurrence and relationship to study vaccination of SAEs and MAAEs (in all adult participants) during the entire study period.

- **Exploratory endpoints are:**
  - First occurrence of virologically confirmed (by PCR to SARS-CoV-2), symptomatic COVID-19 with onset at least 14 days after first study vaccination (e.g., Day 14) in serologically negative (to SARS-CoV-2) adult participants at baseline until the endpoint-driven efficacy analysis is triggered by the occurrence of a prespecified number of blinded endpoints.
  - Any occurrence of serologic conversion (by serology to SARS-CoV-2 nucleocapsid (N) protein) between baseline and 1 year after last study vaccination in adult participants seronegative at baseline.
  - Analysis of the proportion of adult participants with evidence of seroconversion as demonstrated by binding antibodies to the SARS-CoV-2 S protein by ELISA at Day 35 (14 days after second study vaccination).
  - Analysis of the cell-mediated immune responses (for example, as measured by enzyme-linked immune absorbent spot (ELISpot) ± intracellular cytokine staining) on Days 0 (baseline) and 35 (14 days after second study vaccination).
  - SARS-CoV-2 neutralisation as measured by virus neutralisation assay (VNA; wild-type virus and/or pseudovirion expressing SARS-CoV-2 S protein) at Day 0 (baseline) and Day 35 (14 days after second study vaccination).
  - Analysis of the safety and immunogenicity of SARS-CoV-2 rS with Matrix-M1 adjuvant when co-administered with seasonal influenza vaccine in a subset population.

**STUDY DESIGN:**

This is a Phase 3, multi-centre, randomised, observer-blind, placebo-controlled study evaluating the efficacy, safety, and immunogenicity of SARS-CoV-2 rS with Matrix-M1 adjuvant in adult participants 18 to 84 years of age (inclusive) in the UK. Every effort will be made to identify regions of high SARS-CoV-2 activity, and populations within these regions who are at high risk of exposure to the virus will be enrolled such that the incidence of COVID-19 in the study will be higher than 1% and the sample size and enrolment period may be reduced.

After signing informed consent, participants may be screened within a window of up to approximately 30 days. During the screening period, mucosal samples (nasal or throat swabs or saliva based on test availability) may be taken to detect PCR for SARS-CoV-2, if the participant has any COVID-19 symptoms or significant exposure history.

Approximately 9,000 male and female adult participants 18 to 84 years of age (inclusive) with and without relevant comorbidities is planned for the study. An effort will be made to enrol a target of at least 25% of participants who are ≥ 65 years of age as well as prioritising other groups that are most affected by COVID-19, including racial and ethnic minorities. This will be under the assumption that the annual incidence of symptomatic COVID-19 will be approximately 1% to 5%. Enrolment of participants ≥ 65 years of age will not be initiated until approval from the Medicines and Healthcare products Regulatory Agency (MHRA) following their review of Phase 2 study safety data in this age group.

The actual sample size for the study will be selected at the operational cut-off date before initiation of the study, based on estimated incidences for the targeted study regions and populations at that time. The sample size may be adjusted by the sponsor during the study, based on blinded data, to ensure sufficient power at the time of the primary analysis. Details on the possible sample-size reassessment will be described in the statistical analysis plan (SAP).

On Day 0, mucosal samples (nasal or throat swabs or saliva based on test availability) will be taken to detect PCR for SARS-CoV-2 and blood samples will be taken to detect any existing SARS-CoV-2 antibodies at baseline. Participants will be randomised in a 1:1 ratio via block randomisation to receive 2 intramuscular (IM) injections of the SARS-CoV-2 rS with Matrix-M1 adjuvant or placebo, as described in Table S1-1. Randomisation will be stratified by site and by age ≥ 65 years. In addition, there will not be specific age sub-stratification recruitment targets within the ≥ 65-year age group, but through site recruitment; the aim is to allow a generally representative age distribution overall.

**Table S1-1 Study Vaccine Groups**

| Study Vaccine Groups | Number of Randomised Participants | 2 Vaccinations | |
|---|---|---|---|
| | | Day 0 | Day 21 (+ 7 days) |
| SARS-CoV-2 rS (5 µg) + Matrix-M1 adjuvant (50 µg) | N = 4,500 | X | X |
| Placebo | N = 4,500 | X | X |

The study will consist of the screening period (Days -30 to 0); study vaccination days (Days 0 and 21 (+7-day window); and outpatient study visits on Day 0, Day 21 (+ 7 days), and Day 35 (14 days minimum after second study vaccination [+ 7 days]); and at 3, 6, and 12 months (± 15 days) after last study vaccination.

The duration of individual participation, including screening, will be a maximum of 1 year after last study vaccination (Day 386 ± 15 days). This will be an event-driven study for the assessment of vaccine efficacy (VE) and will end when a sufficient number of events have been observed, yet all participants will be followed for the entire study duration for safety endpoints.

A licensed seasonal influenza co-administration sub-study will be conducted in the first 400 participants who meet the additional inclusion criteria for this study. Participants may be enrolled at select study sites due to the availability of seasonal influenza vaccine. After being randomised to receive IM injections of SARS-CoV-2 rS with Matrix-M1 adjuvant or placebo, sub-study participants will receive a licensed seasonal influenza vaccine on Day 0 in the opposite deltoid. These participants will be part of the solicited AE safety subset analysis.

Given that this study and others will enrol during the influenza virus season and that the highest at-risk population, namely the elderly and adults with comorbid conditions for influenza mirrors that of SARS-CoV-2, a co-vaccination sub-study is included. This sub-group will undergo the same safety and efficacy evaluations as the entire study population. In addition, the entire sub-study will have reactogenicity assessed. This group must not have had a current season licensed influenza vaccine and have no prior history of allergy or severe reaction to seasonal influenza vaccine. To assess the possible impact of the study vaccine on the immunogenicity of the influenza vaccine, this group will also have a hemagglutination inhibition assay (HAI) performed.

A global Safety Monitoring Committee (SMC) was convened to oversee the safety of the ongoing Phase 1/2 study (Protocol 2019nCoV-101) and will oversee one or more additional studies across the SARS-CoV-2 rS vaccine program. A separate SMC will be convened for this study. The designated SMC will monitor the safety of participants in the study and will follow an SMC charter. The SMC will review blinded and unblinded safety and reactogenicity data. The SMC will also review sufficient clinical event data to ensure detection of any evidence of enhanced disease. The SMC will convene to perform safety reviews on a scheduled basis, for immediate concerns regarding safety observations during this study, and as needed.

**Study Vaccination Pause Rules:**

Study vaccination pause rules based on reactogenicity, AEs, and SAEs related to study participation will be in place to monitor participant safety during the study.

AEs meeting any one of the following criteria will result in a hold being placed on subsequent study vaccinations pending further review by the SMC and the sponsor:

- The occurrence of 1 or more related SAEs (final assessment by the sponsor per United States Food and Drug Administration Center for Biologics Evaluation and Research Guidance) in a given Medical Dictionary for Regulatory Activities (MedDRA) system organ class within the first 7 days after study vaccination with the exception of hypersensitivity (anaphylaxis), which would require 2 occurrences.
- Any toxicity grade 3 (severe) solicited (local or systemic) single AE term occurring in $\geq$ 10% of participants in the SARS-CoV-2 rS with Matrix-M1 adjuvant group within the first 7 days after study vaccination.
- Any grade 3 (severe) unsolicited single AE preferred term for which the investigator assesses as related that occurs in $\geq$ 5% of participants in the SARS-CoV-2 rS with Matrix-M1 adjuvant group, within the first 7 days after study vaccination.

The sponsor, along with medical monitor, may request an SMC review for any safety concerns that may arise in the study, even if they are not associated with any specific pause rule.

**STUDY POPULATION:**

**Inclusion Criteria:**

Each participant must meet all of the following criteria to be enrolled in this study:

1. Adult males or females aged 18 to 84 years (inclusive) at screening.
2. Able and willing (in the investigator's opinion) to comply with all study requirements.
3. Willing to allow the investigators to discuss the volunteer's medical history with their General Practitioner and access all medical records when relevant to study procedures.
4. Willing and able to give informed consent prior to study enrolment.
5. Female participants of childbearing potential (defined as any female who has experienced menarche and who is NOT surgically sterile [i.e., hysterectomy, bilateral tubal ligation, or bilateral oophorectomy] or postmenopausal [defined as amenorrhea at least 12 consecutive months or > 1 documented plasma follicle-stimulating hormone level ≥ 40 mIU/mL]) must agree to be heterosexually inactive from at least 28 days prior to enrolment and through 3 months after the last study vaccination OR agree to consistently use any of the following methods of contraception from at least 28 days prior to enrolment and through 3 months after the last study vaccination:
   a. Condoms (male or female) with spermicide
   b. Diaphragm with spermicide
   c. Cervical cap with spermicide
   d. Intrauterine device
   e. Oral or patch contraceptives
   f. Norplant®, Depo-Provera®, or other in country regulatory-approved contraceptive method that is designed to protect against pregnancy
   g. Abstinence, as a form of contraception, is acceptable if in line with the participant's lifestyle

   **NOTE**: Periodic abstinence (e.g., calendar, ovulation, symptothermal, post-ovulation methods) and withdrawal are not acceptable methods of contraception.

**Seasonal Influenza Vaccine Co-Administration Sub-Study Only**

6. Participant should not have received a current season influenza vaccine, have no contraindication to the specific vaccine to be administered in the study, and no prior history of allergy or severe reaction to seasonal influenza vaccines.

**Exclusion Criteria:**

Participants meeting any of the following criteria will be excluded from the study:

1. Participation in COVID-19 prophylactic drug trials for the duration of the study.
2. Participation in SARS-CoV-2 serological surveys where participants are informed of their serostatus for the duration of the study.
3. Participation in research involving an investigational product (drug/biologic/device) within 45 days prior to first study vaccination.
4. History of laboratory-confirmed (by PCR or serology to SARS-CoV-2) COVID-19 infection at any time prior to randomisation.
5. Administration of immunoglobulins and/or any blood products within the 3 months preceding the planned administration of the study vaccine candidate.
6. Any confirmed or suspected immunosuppressive or immunodeficient state; chronic administration (defined as more than 14 continuous days) of immunosuppressant medication within the past 3 months, except topical steroids or short-term oral steroids (course lasting ≤ 14 days).

   **NOTE:** An immunosuppressant dose of glucocorticoid is defined as a systemic dose ≥ 10 mg of prednisone per day or equivalent. The use of topical, inhaled, and nasal glucocorticoids will be permitted if other chronic disease conditions are not exclusionary.
7. History of allergic disease or reactions likely to be exacerbated by any component of the study vaccines.
8. Any history of anaphylaxis to any prior vaccine.
9. Pregnancy, lactation or willingness/intention to become pregnant during the study.
10. Current diagnosis of or treatment for cancer (except basal cell carcinoma of the skin and cervical carcinoma in situ, at the discretion of the investigator).
11. Bleeding disorder (e.g., factor deficiency, coagulopathy or platelet disorder), or prior history of significant bleeding or bruising following IM injections or venepuncture.
12. Continuous use of anticoagulants, such as coumarins and related anticoagulants (i.e., warfarin) or novel oral anticoagulants/anti-platelet agents.

    **NOTE:** The use of ≤ 325 mg of aspirin per day as prophylaxis is permitted.
13. Suspected or known current alcohol or drug dependency.
14. Study team member or first-degree relative of any study team member (inclusive of sponsor, contract research organisation (CRO), and site personnel involved in the study).
15. Participants who are having any current workup of undiagnosed illness within the last 8 weeks that is either participant-reported or has been clinician-assessed, which could lead to a new condition or diagnosis.
16. Received any live vaccine within 4 weeks or **any vaccine** (excluding influenza) within 2 weeks prior to first study vaccination or any licensed influenza vaccine within 1 week prior to first study vaccination. Plans to receive any vaccine from these time periods until 28 days after second study vaccination (with the exception of

participants in the seasonal influenza co-administration sub-study; a licensed seasonal influenza vaccine may be received as soon as 7 days after second study vaccination).

**NOTE:** An influenza co-administration sub-study will occur in which 400 participants will receive a single dose of seasonal influenza vaccine at the same time as first study vaccination.

17. Have clinically significant chronic cardiovascular, endocrine, gastrointestinal, hepatic, renal, neurological, respiratory, psychiatric or other medical disorders not excluded by other exclusion criteria, that are assessed by the investigator as being clinically unstable within the prior 4 weeks as evidenced by:
    a. Hospitalisation for the condition, including day surgical interventions.
    b. New significant organ function deterioration.
    c. Needing addition of new treatments or major dose adjustments of current treatments (mild or moderate well-controlled comorbidities are allowed).
18. History of chronic neurological disorders that have required prior specialist physician review for diagnosis and management (such as multiple sclerosis, dementia, transient ischemic attacks, Parkinson's disease, degenerative neurological conditions, neuropathy, and epilepsy) or a history of stroke or previous neurological disorder within 12 months with residual symptoms. Participants with a history of migraine or chronic headaches or nerve root compression that have been stable on treatment for the last 4 weeks are not excluded.
19. Any autoimmune or immunodeficiency disease/condition (iatrogenic or congenital).

    **NOTE:** Stable endocrine disorders that have a confirmed autoimmune etiology (e.g., thyroid, pancreatic) are allowed.
20. Any other significant disease, disorder or finding that, in the opinion of the investigator, may significantly increase the risk to the volunteer because of participation in the study, affect the ability of the volunteer to participate in the study, or impair interpretation of the study data.

**Other Considerations:**

Participants meeting any of the following criteria may have planned study vaccination deferred for a later date, but these criteria are not exclusionary for study enrolment.

- Respiratory symptoms in the past 3 days (i.e., cough, sore throat, difficulty breathing). Participant may be vaccinated once all symptoms have been resolved for > 3 days. Out-of-window study vaccination is allowed for this reason (**see NOTE**).
- Temperature of > 38°C within 24 hours of planned study vaccination (site measured or participant measured). Participant may be vaccinated once the fever has resolved and there has not been any temperature measured as being > 38°C for > 3 days. Out-of-window study vaccination is allowed for this reason (**see NOTE**).

  **NOTE:** PCR testing for SARS-CoV-2 is likely to be indicated for either of the above reasons or if COVID-19 is suspected based on other symptoms, potential exposure to SARS-CoV-2 infection through either close contacts or based on local epidemiology.

If a participant has a positive PCR for SARS-CoV-2 prior to enrolment that participant will not be eligible and will be considered a screen failure.

- Participants having any symptoms or signs of possible COVID-19 infection (Table 2-2) that may also be due to post-vaccination reactogenicity within 7 days of either study vaccine dose will NOT be required to be tested for SARS-CoV-2 PCR.
- Any participant with a new positive PCR-confirmed SARS-CoV-2 infection occurring from Screening and prior to second study vaccination will not be removed from the study but must meet health requirements before receiving second study vaccination.
- Any participant who has a positive PCR for SARS-CoV-2 between the first and second study vaccination should also have PCR testing for SARS-CoV-2 on the day of the subsequent study vaccination, but the results of the PCR test are not needed before study vaccination can be given.
- Any participant with a new positive PCR-confirmed SARS-CoV-2 infection occurring from Screening and prior to the end of immunogenicity assessments will be removed from applicable immunogenicity analyses as defined in the SAP.
- Any acute illness (cardiovascular, endocrine, gastrointestinal, hepatic, renal, neurological, respiratory, or other medical disorders) that is actively causing symptoms that could, in the opinion of the investigator, impact the assessment of reactogenicity or other study assessments may have planned study vaccination deferred for a later date, but these criteria are not exclusionary for study enrolment. Participant may be vaccinated once symptoms have resolved or are stabilised for > 3 days. Out-of-window study vaccination is allowed for this reason.

**STUDY VACCINES:**

Study vaccinations (5-µg SARS-CoV-2 rS with 50-µg Matrix-M1 adjuvant or placebo [saline]) will comprise 2 IM injections on Days 0 and 21, ideally in alternating deltoids. For blinding purposes, all participants will be vaccinated using the same injection volume (i.e., 0.5 mL).

The seasonal influenza vaccine co-administration sub-study will comprise a single IM injection (0.5 mL) of a licensed influenza vaccine on Day 0, ideally in the opposite deltoid of the study vaccine.

All study vaccinations will be administered on an outpatient basis by designated site personnel in a way to maintain the blind. The influenza vaccine will not require blinding. Any pharmacy preparation with unblinded product will require unblinded site personnel who will not otherwise be involved in the study procedures or observation of participants.

**STUDY PROCEDURES:**

Study procedures, including efficacy, immunogenicity, and safety assessments are listed in the schedule of events (SOE) in Table 3-1.

**Efficacy Assessments:**

**Mucosal Testing for SARS-CoV-2 Detection and Confirmation:**

Mucosal samples (nasal/throat swabs or saliva) for virus detection will be taken at the study visits described in the SOE (Table 3-1).

Mucosal sampling (nasal/throat swabs or saliva) will be performed to virologically confirm (by PCR to SARS-CoV-2) the presence of SARS-CoV-2 beginning on Day 0 until the EOS, yet only those SARS-CoV-2 cases detected 7 days after second study vaccination will be utilised in study efficacy endpoints.

**Monitoring for COVID-19:**

Identification and laboratory confirmation of SARS-CoV-2 infection and symptomatic COVID-19 will be performed throughout the study.

On study enrolment, participants will be given details of a 24/7 on-call number to contact the study team. Participants will be instructed to contact the study team within 24 hours via this number if they self-assess COVID-19 symptoms in Table 2-2 or are clinically concerned. Newly discovered symptoms of suspected COVID-19 will trigger a COVID-19 Surveillance Visit. Please refer to Table 2-2 for the qualifying symptoms of suspected COVID-19 disease. Participants will receive weekly reminders (email or text messages) to immediately contact the study team if they have experienced any new symptoms or health concerns that could be related to infection with SARS-CoV-2. Newly discovered symptoms of suspected COVID-19 will trigger a COVID-19 Surveillance Visit. A participant with suspected or confirmed COVID-19 will be asked to complete the COVID-19 symptom diary. Participants should start the diary on their first day of symptoms and complete this diary daily for a minimum of 10 days even if their symptoms have resolved and regardless of any SARS-CoV-2 PCR results. They should continue the diary beyond 10 days if they have any ongoing symptoms related to the episode and should continue to do so until all symptoms have resolved.

The occurrence of COVID-19-related hospitalisation and COVID-19-related complications (such as, but not limited to, pneumonia, neurological or vascular complications, severe pneumonia, severe neurological or vascular events, acute respiratory distress syndrome, renal complications, sepsis, septic shock, death) will be monitored throughout the study. Every episode of a new onset of symptoms of suspected COVID-19 will trigger a COVID-19 Surveillance Visit (Initial and Follow-Up). A “new onset” episode will require at least a 7-day period of being symptom-free prior to the event to differentiate a specific episode from any prior illness.

**Immunogenicity Assessments:**

Blood will be collected from all participants for humoral immunogenicity according to the time points specified in the SOE (Table 3-1).

Immune measurements (ELISA) will be conducted on serum (immunoglobulin G [IgG]) for SARS-CoV-2 anti-S protein serology in approximately 900 participants in the Anti-S Protein

Serology Subset and on anti-N protein serology in all participants. The immunologic test for SARS-CoV-2 serology based on a non-S protein (e.g., SARS-CoV-2 N protein) will be performed to identify cases of asymptomatic infection. Additional immunogenicity assessments specific to SARS-CoV-2 (or related variants) will include a neutralising antibody assay, which will be performed in approximately 900 participants in a Neutralisation Assay Subset. Cell-mediated immune responses, as assessed by ELISpot ± intracellular cytokine staining, will be performed in approximately 450 participants in the Cell-mediated Assay Subset. In addition, an HAI will be performed in the approximately 400 participants in the seasonal influenza vaccine co-administration sub-study.

**Safety Assessments:**

The timing and frequency of all safety assessments are listed in the SOE (Table 3-1).

After each study vaccination, including the licensed seasonal influenza vaccine for participants in the seasonal influenza vaccine co-administration sub-study, participants will remain under observation at the study site for at least 30 minutes for the presence of any acute reactions and solicited AEs. Solicited AEs, collected through an electronic diary (e-Diary), will be recorded from the time of study vaccination until 7 days after study vaccination in all participants in the seasonal influenza vaccine co-administration sub-study and a subgroup of all other participants.

All participants will be assessed for unsolicited AEs from the time of first study vacation until Day 49; SAEs will be assessed from the time the informed consent is signed until completion of the participant's last study-related procedure; all MAAEs will be reported from the time of first study vaccination until Day 35; MAAEs related to study vaccination and AESIs will be reported from the time of first study vaccination until completion of the participant's last study-related procedure. All AEs will be followed until resolution or until clinically stable.

Safety assessments will include monitoring and recording of solicited (local and systemic reactogenicity events) and unsolicited AEs; MAAEs; AESI; SAEs; vital sign measurements on day of study vaccinations; and physical examination findings. COVID-19 severity will be categorised as virologically confirmed, mild, moderate, or severe according to protocol-specified criteria (Table 2-1). Recording of solicited and unsolicited AEs may be conducted by electronic data capture (EDC)/reporting. Potential immune-mediated medical conditions (PIMMC) and AESIs specific to potential disease enhancement for COVID-19 will also be monitored (see Section 9.4 [Appendix 4] for details).

**STATISTICAL ANALYSIS PLANS:**
**Sample Size:**

This study is designed to enrol approximately 9,000 participants, randomised 1:1 into the 2 study vaccine groups.

Power calculations were performed for each primary endpoint using the two-sided 97.5% confidence intervals (CIs; i.e., one-side alpha of 0.0125) to be conservative (Table 7-11). This is designed to provide at least 90% power for each of the 2 primary endpoints based on the following assumptions:

1. A symptomatic mild, moderate, or severe COVID-19 incident rate of 3% in the placebo group and a VE of 50%.
2. A symptomatic moderate or severe COVID-19 incident rate of 2% in the placebo group and a VE of 70%.
3. 90% evaluability rate for the per-protocol efficacy (PP-EFF) population (assuming 10% unevaluable due to attrition and/or baseline-seropositive participants).

Based on the above conservative assumptions, the minimum target numbers of endpoints needed are 92 symptomatic mild, moderate, or severe COVID-19 endpoints or 43 symptomatic moderate or severe COVID-19 endpoints. However, in order to increase the precision around VE (i.e., narrow the CIs), the target numbers for the final analysis may be increased up to 114 symptomatic mild, moderate, or severe COVID-19 endpoints or 55 symptomatic moderate or severe COVID-19 endpoints. These increased target numbers of events will provide 80% power at one-sided Type I error rate of 0.025 to demonstrate a lower bound CI > 30% if the VE is 60% for symptomatic mild, moderate, or severe COVID-19 and 70% for symptomatic moderate or severe COVID-19. The minimum sample size equals 3,633/group (7,266 in total). In order to expedite the time to target number of endpoints needed or lower the expected endpoint accrual rate the study will enrol approximately 9,000 participants. The actual sample size for the study will be selected at the operational cut-off date before initiation of the study, based on estimated incidences for the targeted study regions and population at that time.

**Analysis Sets:**

The intent-to-treat efficacy (ITT-EFF) and immunogenicity (ITT-IMM) analysis sets will include all participants who are randomised and receive at least 1 dose study vaccine (SARS-CoV-2 rS with Matrix-M1 adjuvant or placebo), regardless of protocol violations or missing data. The intent-to-treat (ITT) analysis set will be used as a supportive analysis population for the immunogenicity and efficacy analyses and will be analysed according to the study vaccine group as randomised.

The safety analysis set will include all participants who receive at least 1 dose of study vaccine (SARS-CoV-2 rS with Matrix-M1 adjuvant or placebo). Participants in the safety analysis set will be analysed according to the study vaccine actually received.

The PP-EFF will include seronegative participants who receive both doses of study vaccine (SARS-CoV-2 rS with Matrix-M1 adjuvant or placebo) and have no major protocol deviations affecting the primary efficacy outcome as assessed by the sponsor prior to unblinding. All analyses of the PP-EFF population will exclude any illness episodes with positive SARS-CoV-2 by any validated PCR or antibody test occurring before 14 days after second study vaccination (e.g., Day 35).

The per-protocol immunogenicity (PP-IMM) analysis set for each post-randomisation visit will include participants who receive scheduled dose(s) of study vaccine (SARS-CoV-2 rS with Matrix-M1 adjuvant or placebo), have at least a baseline and the serum sample result for the visit available after study vaccination, and have no major protocol violations that are considered clinically likely to impact the immunogenicity response at the corresponding study visit as assessed by the sponsor prior to unblinding. For each visit, the SARS-CoV-2 unexposed population will also exclude any illness episodes with positive SARS-CoV-2 by any validated PCR or antibody test prior to each visit. Prior exposed participants will be determined using baseline SARS-CoV-2 immunity defined as positive SARS-CoV-2 by PCR or serum IgG antibody at baseline, or positive SARS-CoV-2 by qualitative PCR through Day 21 or Day 35, according to the specified analysis. Analysis will be performed to assess if immune responses differ between exposed and unexposed individuals (i.e., whether prior exposure alters dosing regimen considerations in a pandemic response).

The review and determination for exclusion from the PP populations will be carried out in a blinded fashion by a study clinician prior to unblinding for each interim analysis based on all available information from the locked database.

**Efficacy Analyses:**

The study will have 2 key decision time points for efficacy: 1) evaluation of an early efficacy signal to guide a potential request for conditional approval based on accumulating events and 2) the primary analysis to evaluate the primary and secondary objectives of the study.

The primary endpoints will be analysed on the PP-EFF Analysis Set and supported by analysis of the ITT-EFF Set. Conclusions concerning declaration of attainment of the primary endpoint at the completion of the study will only be based on the PP-EFF population. In addition, supportive analyses based on the ITT-EFF population will also be performed.

VE is defined as VE (%) = (1 – RR) × 100, where RR = relative risk of incidence rates between the 2 study vaccine groups (SARS-CoV-2 rS with Matrix-M1 adjuvant or placebo). The final analysis for the primary objective in the PP-EFF population will be carried out at the overall one-sided Type I error rate of 0.025 overall for the 2 primary endpoints. The family-wise type 1 error rate among these 2 analyses will be controlled using the Hochberg approach. Since the demonstration of the primary objective can be achieved if at least 1 of the 2 primary endpoints meets the pre-specified success criteria, the statistical analysis will be performed in the following order:

a. Construct two-sided 95% CIs for the 2 primary endpoints. If both lower bounds are > 0%, declare that both endpoints have met the success criteria. Otherwise, proceed to the next step.

b. Construct two-sided 97.5% CIs for the 2 primary endpoints. If the higher of the lower bounds is > 0%, declare that only the endpoint associated with lower bound > 0% has met the success criterion. Otherwise, both endpoints did not meet the success criteria.

The estimated RR and its CI will be derived using Poisson regression with robust error variance [Zou 2004]. The explanatory variables in the model will include study vaccine group. The dependent variable will be the incidence rate of the endpoint of interest. The robust error variances will be estimated using the repeated statement and the participant identifier. The Poisson distribution will be used with a logarithm link function.

As the primary analysis, hypothesis testing of each primary efficacy endpoint will be carried out against H0: VE ≤ 0%. Rejection of the null hypothesis, H0: VE ≤ 0% demonstrates a statistically significant vaccine effect for either primary endpoint. The study will continue for the intended duration to measure immunogenicity and safety endpoints, regardless of primary endpoint success. The final analysis of the primary endpoints will be triggered when at least 92 PP-EFF participants with symptomatic mild, moderate, or severe COVID-19 or approximately 43 PP-EFF participants with symptomatic moderate or severe COVID-19 have accrued. Also, in order to be able to respond to the unexpected and rapidly evolving COVID-19 pandemic situation globally, other factors such as requests by government or public health agencies may also be factored into the decision-making to unblind the study for the final analysis, but this always occurs in consultation with lead regulatory agencies.

In order to address the uncertainty around the COVID-19 circulation in the study population, an active monitoring of the blinded endpoint accruals for the 2 primary endpoints will be performed by the sponsor. The sponsor may choose to switch 1 of the 2 primary endpoints to a secondary or exploratory endpoint prior to unblinding, if it is determined that the accrual for the endpoint is likely to fall well short of the target number. The decision will be based solely on the total numbers of blinded endpoints accrued without consideration of treatment group assignment. The decision will not be based on unblinded VE estimates. If this option is triggered, the remaining primary endpoint will be analysed using one-sided Type I error rate of 0.025 (i.e., two-sided 95% CI). The details for the criteria to be used to act on this option will be included in the SAP.

The final analysis will be performed when the last participant completes the last visit (12 months after the last study vaccination) or discontinues earlier. The EOS analysis will be performed when all participants in the Neutralisation Assay Subset have completed the last study visit or discontinued earlier.

**Immunogenicity Analyses:**

The primary and secondary immunogenicity analyses will be performed using the PP-IMM and ITT-IMM analysis populations.

For the serum antibody levels measured by microneutralization and HAI assays, the geometric mean at each study visit, the geometric mean fold rises (GMFRs) comparing to Day 0 (baseline) at each follow-up study visit, along with 95% CI will be summarised by study vaccine group. The 95% CI will be calculated based on the t distribution of the log-transformed values for geometric means or GMFRs, then back transformed to the original scale for presentation. The seroconversion rate, proportion of participants with ≥ 4 fold rises if naïve at baseline, along with 95% CIs based on the Clopper-Pearson method will be summarised by study vaccine group at each follow-up study visit.

Similar summaries will be generated for the other immunogenicity endpoints and other assays if conducted.

Correlates of risk will be assessed where immune responses are measured in all participants, and the results compared between participants who experience a SARS-CoV-2 event and participants who do not. The goal of this analysis is to assess correlates of risk of SARS-

CoV-2 infection (and potential other secondary endpoints) in the study vaccine group by comparing study vaccine-induced immune responses associated with COVID-19.

**Safety Analyses:**

Solicited AEs (e.g., local and general systemic reactogenicity symptoms) will be captured in the Safety Subset of approximately 2,000 participants and in the seasonal influenza vaccine co-administration sub-study of approximately 400 participants.

Numbers and percentages (with 95% CIs based on the Clopper-Pearson method) of participants with solicited local and systemic AEs through 7 days after each study vaccination will be summarised by study vaccine group and the maximum toxicity grade over 7 days after each study vaccination. The duration of solicited local and systemic AEs after each study vaccination will also be summarised by treatment group.

Unsolicited AEs will be coded by preferred term and system organ class using the latest version of the MedDRA and summarised by study vaccine group as well as by severity and relationship to study vaccine. AEs through 35 days after first study vaccination; all MAAEs and MAAEs related to study vaccine; SAEs; or AESI through EOS will be listed separately and summarised by study vaccine group.

Vital sign measurements will be summarised by study vaccine group using descriptive statistics at baseline and following study vaccination.

Concomitant medications will be provided in a data listing with preferred drug name as coded using the World Health Organisation drug dictionary.

**Planned Analyses Prior to Study Completion:**

Analysis of the data will be provided on an ongoing basis for confirming success and to review safety as the study progresses. The SAP will outline the sequential nature of these reviews.

### 1. INTRODUCTION

#### 1.1 Background

Coronaviruses are medium sized, enveloped, positive-stranded ribonucleic acid (RNA) viruses, with a characteristic crown-like appearance in electron micrographs due to circumferential studding of the viral envelope with projections comprising the spike (S) protein. There are 4 different strains (229E, OC43, NL63, and HKU1), which are ubiquitous in humans and generally result in mild upper respiratory illnesses and other common cold symptoms including malaise, headache, nasal discharge, sore throat, fever, and cough [Su 2016]. In addition, other coronavirus strains are widespread in animals, where they typically cause enteric disease. These zoonotic coronaviruses have been known to evolve into strains that can infect humans with serious consequences including severe acute respiratory syndrome coronavirus (SARS-CoV) from 2002 to 2003, Middle Eastern Respiratory Syndrome (MERS)-CoV since 2012, and most recently, the novel SARS-CoV-2 since 2019 [Habibzadeh 2020].

In late December of 2019, an outbreak of respiratory disease caused by novel coronavirus (2019 nCoV) was detected in Wuhan, Hubei province, China. The virus' rapidly discerned genetic relationship with the 2002-2003 SARS-CoV has resulted in adoption of the name "SARS-CoV-2," with the disease being referred to as coronavirus disease 2019 (COVID-19). Despite containment efforts since the start of the outbreak, the SARS-CoV-2 has spread rapidly with over 214 countries/territories/areas outside of China reporting laboratory confirmed COVID-19 cases as of 15 May 2020 [WHO, 2020]. On 30 January 2020, the International Health Regulations Emergency Committee of the World Health Organisation (WHO) designated the outbreak as a public health emergency of international concern (PHEIC) and subsequently declared a pandemic on 11 March 2020.

Novavax, Inc. is developing a recombinant vaccine adjuvanted with the saponin-based Matrix-M1™ for the prevention of disease caused by SARS-CoV-2. SARS-CoV-2 recombinant (r) spike (S) protein nanoparticle vaccine (SARS-CoV-2 rS) is constructed from the full-length, wild-type SARS-CoV-2 S glycoprotein (GP) based upon the GenBank gene sequence MN908947, nucleotides 21563-25384, from the 2019 SARS-CoV-2 genome. The S protein is a type 1 trimeric glycoprotein of 1,273 amino acids that is produced as an inactive S0 precursor. The S-gene was codon optimised for expression in *Spodoptera frugiperda* (Sf9) insect cells. The SARS-CoV-2 rS nanoparticle vaccine is intended for administration with Matrix-M1 adjuvant, which is a saponin-based adjuvant that has previously been shown to enhance the immunogenicity of other nanoparticle vaccines in nonclinical and clinical studies.

#### 1.2 Non-Clinical Experience

In support of the development of SARS-CoV-2 rS, Novavax has obtained nonclinical pharmacology data concerning several SARS-CoV-2 S protein variants, toxicity data concerning SARS-CoV-2 rS with Matrix-M1 adjuvant, and prior toxicity data concerning other viral glycoproteins manufactured in the baculovirus-Sf9 system and formulated with Matrix-M1 adjuvant.

##### 1.2.1 Nonclinical Data from SARS-CoV-2 Spike Protein Constructs that Support SARS-CoV-2 rS Development

Mouse immunogenicity studies were conducted to evaluate several SARS-CoV-2 S protein variants and to select the vaccine candidate. The selected vaccine candidate, BV2373 (3Q-2P), was demonstrated to be immunogenic and elicited functional antibodies. For the tested constructs, shallow dose responses with Matrix-M1 adjuvant were observed, suggesting that the adjuvant may be significantly antigen-sparing in large animals and humans.

The candidate SARS-CoV-2 rS vaccine, based on the BV2373 construct, is also being evaluated in dose-titration studies in [redacted] cynomolgus macaques, and baboons. [redacted] In cynomolgus macaques, 2 dose regimens of 5 or 25 µg SARS-CoV-2 rS/25 or 50 µg Matrix-Ml adjuvant were also highly immunogenic, resulting in high anti-S IgG levels, high hACE2 binding inhibition titers, and high neutralising antibody responses. The 5 and 25 µg antigen doses gave generally similar responses when administered twice with 50 µg of Matrix-Ml adjuvant. In baboons, which may be more predictive of responses in humans, 5 and 25 µg SARS-CoV-2 rS/50 µg Matrix-Ml adjuvant induced high levels of anti-S IgG, hACE2-binding inhibiting antibodies, and neutralising antibodies. Matrix-Ml adjuvant provided antigen-sparing, and supported induction of functional antibodies. Importantly, Matrix-Ml adjuvanted SARS-CoV-2 rS also appeared to induce strong T helper 1 (Thl) type $CD4^+$ T-cell responses to SARS-CoV-2 S protein that included polyfunctional effector phenotypes. Current data in this small baboon study confirms that doses of 5 µg and 25 µg with 50 µg Matrix-M1 are the correct doses to test clinically, with Matrix-M appearing critical for maximum responses.

Virus challenge studies were performed in mice [redacted] and cynomolgus macaques. In 2 mouse challenge models, immunisation with 1 or 2 doses of SARS-CoV-2 rS/Matrix-M1 adjuvant suppressed viral replication, reduced lung inflammation, and reduced systemic morbidity (weight loss) after SARS-CoV-2 live virus challenge and were not associated with any obvious exacerbation of the inflammatory response to the virus or worsening of clinical outcomes. [redacted]

[redacted] In cynomolgus macaques, administered with human doses of 5 or 25 µg SARS-CoV-2 rS adjuvanted with 50 µg Matrix-M1, high and comparable levels of anti-S IgG titers and hACE2 receptor binding inhibition titers were detected 21 days after the first immunisation. All of the macaques immunised with any dose or regimen of SARS-CoV-2 rS/Matrix-M1 adjuvant were protected against live virus challenge as evidenced by the reduction of total viral RNA and subgenomic RNA to below the limit of quantitation in bronchoalveolar lavages and nasal swabs.

[redacted]

##### 1.2.2 Nonclinical Data from Other Baculovirus-Sf9-Produced Nanoparticle Vaccines that Support SARS-CoV-2 rS Development

The immunogenicity and protective efficacy of 2002-2003 SARS-CoV S protein and chimeric influenza/SARS-CoV virus-like particle (VLP) vaccines produced in the baculovirus-Sf9 system and administered with and without aluminum hydroxide adjuvants was demonstrated in a mouse challenge study [Liu 2011]. Robust neutralising antibody titers were observed following vaccination, although both antigens required adsorption to aluminum hydroxide for optimal responses. The immunogenicity and protective efficacy of a MERS-CoV S nanoparticle vaccine with and without Matrix-M1 adjuvant was demonstrated in a mouse challenge study [Coleman 2017]. Following vaccination, the MERS-CoV S nanoparticle was immunogenic across all active treatment groups; however, the presence of Matrix-M induced a 3- to > 10-fold enhancement of the binding and neutralising antibody responses. In addition, Matrix-M1 adjuvant essentially eliminated the antigen dose-response, suggesting the potential for major antigen-sparing and consequent improved manufacturing efficiency and timeliness [Coleman 2017]. The Matrix-M1 adjuvant was also shown to enhance antibody, cellular, and protective immune responses in Balb/c mice administered Zaire ebolavirus (EBOV) GP vaccine with or without Matrix-M1 or aluminum phosphate adjuvants [Bengtsson 2016].

In addition, 3 GLP-compliant toxicology studies in NZW rabbits have been performed with 4 different antigens (influenza hemagglutinin [HA] ± respiratory syncytial virus [RSV] F, Zika virus envelope dimers [ZIKV EnvD], and EBOV GP), in which up to 100 µg Matrix-M1 adjuvant alone or with antigen was evaluated. These toxicological investigations indicated that baculovirus-Sf9-produced antigens (up to 240 µg total nanoparticle dose) with Matrix-M1 adjuvant (up to 100 µg) were well tolerated in the animals tested with no evidence of toxicity suggestive of any unusual risk or target organ for toxicity. Non-adverse findings, including local injection site inflammation, enlargement of the lymph nodes draining the injection sites, and elevated serum markers of inflammation (including C-reactive protein), were transient and were considered consistent with immune system stimulation consequent to immunisation.

Further details are provided in the SARS-CoV-2 rS Investigator Brochure (IB).

#### 1.3 Clinical Experience

The first clinical study with SARS-CoV-2 rS nanoparticle vaccine is 2019nCoV-101, which is a 2-part, randomised, observer-blinded, placebo-controlled, Phase 1/2 trial designed to evaluate the immunogenicity and safety of SARS-CoV-2 rS nanoparticle vaccine with or without Matrix-M1 adjuvant in healthy participants ≥ 18 to ≤ 59 years of age. Results of an interim analysis at Day 35 showed that SARS-CoV-2 rS with Matrix-M1 adjuvant was well tolerated and elicited robust immune responses [Keech 2020]. There were no serious adverse events (SAEs) or adverse events of special interest (AESI). Reactogenicity was mainly mild in severity and of short duration (mean ≤ 2 days), with second vaccinations inducing greater local and systemic reactogenicity. The adjuvant significantly enhanced immune responses (anti-S IgG, hACE2 receptor binding inhibition antibody, and neutralising antibody) and was antigen dose-sparing, and the 2 dose 5µg SARS-CoV-2 rS/Matrix-M1 adjuvant induced mean anti-S IgG and neutralising antibody responses that exceeded the mean responses in convalescent sera from COVID-19 patients with clinically significant illnesses. The vaccine also induced antigen-specific T cells with a largely Th1 phenotype.

Novavax has, in its internally-sponsored clinical trials, tested baculovirus-Sf9-produced nanoparticle vaccines in 14,848 participants comprising older adults, young adults, and a limited number of children 2 to 5 years of age; and also including 3,075 pregnant women, with acceptable safety. Matrix-M adjuvant has been given to 4,311 humans (of which, approximately 2,657 humans received nanoparticle vaccine) with acceptable short-term reactogenicity, and an unremarkable long-term safety profile.

Further details on the clinical experience of the study vaccine can be found in the SARS-CoV-2 rS IB.

#### 1.4 Rationale for Study

Both nonclinical and early clinical data to date have supported clinical development of SARS-CoV-2 rS and Matrix-M1 adjuvant as a potential vaccine against SARS-CoV-2. In rodent and nonhuman primate (NHP) challenge models, SARS-CoV-2 rS and Matrix-M1 adjuvant induced high titers of antibodies measured against anti-S protein and hACE2 receptor binding and achieved neutralisation of wild-type virus that exceeded the magnitude of responses measured in COVID-19 human convalescent sera and provided protection against SARS-CoV-2 challenge [Tian 2020; Mandolesi 2020; Guebre-Xabier 2020]. Notably in NHP studies, clinical doses of vaccine (5- and 25-µg SARS-CoV-2 rS/50-µg Matrix-M1) resulted in sterile immunity in the lungs and nasal passage following wild-type virus challenge, suggesting that the vaccine may both protect against upper and lower respiratory tract disease and interrupt transmission [Guebre-Xabier 2020]. Results from a Day 35 interim analysis of Part 1 (Phase 1) of Study 2019nCoV-101 indicate that in healthy adult participants 18 to 59 years of age two-dose regimens of 5- and 25-µg SARS-CoV-2 rS/50 µg Matrix-M1 (on Days 0 and 21) were well tolerated and induced the most robust immune responses with high levels of neutralising antibodies that closely correlated with anti-spike IgG [Keech 2020]. Furthermore, neutralising antibody responses following second vaccination were of the magnitude seen in convalescent serum from symptomatic COVID-19 patients and exceeded overall convalescent sera geometric mean titers (GMTs) by four-fold. The benefit of Matrix-M1 adjuvant was clear in the magnitude of the antibody and T-cell response, induction of functional antibodies, and dose sparing. A Phase 2 clinical program is underway and will provide safety and immunogenicity results in older participants (> 60 years of age) and participants with comorbidities, as well as preliminary efficacy results. Combining the current nonclinical and clinical data with positive Phase 1/2 data provide the impetus for early initiation of the Phase 3 clinical development program in the context of the current public health pandemic crisis.

The purpose of this study is to evaluate the efficacy, safety, and immunogenicity of SARS-CoV-2 rS with Matrix-M1 adjuvant in adults 18-84 years of age (inclusive). Enrolment of participants ≥ 65 years of age will not be initiated until approval from the Medicines and Healthcare products Regulatory Agency (MHRA) following their review of Phase 2 study safety data in this age group. The study will be conducted at anticipated high COVID-19 transmission areas in the United Kingdom (UK). The information provided in this study will inform progression of the study vaccine, to determine efficacy of the study vaccine to prevent COVID-19 in the general population, in participants regardless of serostatus, in participants who have required medical intervention, and in participants with mild or asymptomatic infections. The study will determine the safety of the study vaccine to use in the general population and to ensure that it elicits a robust immune response.

#### 1.5 Rationale for Dose Selection

As previously described, clinical doses of vaccine and adjuvant (5- and 25-µg SARS-CoV-2 rS/ 50-µg Matrix-M1) resulted in sterile immunity in the lungs and nasal passage following wild-type virus challenge in NHP, suggesting that the vaccine may both protect against upper and lower respiratory tract disease and interrupt transmission [Guebre-Xabier 2020]. These doses are being evaluated in Part 1 of Study 2019nCoV-101 in 131 healthy adult participants ≥ 18 to ≤ 59 years of age and in Part 2 of Study 2019nCoV-101 in up to 1,500 participants ≥ 18 to ≤ 84 years of age, including participants with comorbidities. Results from the Part 1 Day 35 interim analysis support either dose of SARS-CoV-2 rS/Matrix-M1 in terms of safety and immunology, with the lower dose (5 µg) offering advantages in regards to dose sparing.

All study vaccinations will be administered on an outpatient basis by designated site personnel in a way to maintain the blind. Any pharmacy preparation with unblinded product will require unblinded site personnel who will not otherwise be involved in the study procedures or observation of participants.

#### 1.6 Benefit-Risk Assessment

The SARS-CoV-2 rS nanoparticle vaccine contains purified protein antigens. It cannot replicate, nor can it cause COVID-19. However, in common with all vaccines produced in cell culture or other systems, the SARS-CoV-2 rS nanoparticle vaccine contains residual non-vaccine proteins derived from the production system, and sensitisation to these, or the SARS-CoV-2 S protein itself, may theoretically occur. While the occurrence of immediate hypersensitivity is possible with the administration of any vaccine, whether licensed or in development, no such reactions have been observed in any of these clinical trials to date. As clinical data become available with increased exposure, it is possible that this profile may change.

The risk for enhanced COVID-19 in immunised participants is a theoretical risk. Enhanced disease in coronavirus vaccine-immunised animals after live virus challenge has been demonstrated in nonclinical studies of several, but not all, coronavirus vaccine candidates. There is currently no evidence for immunoenhancement in nonclinical testing of SARS-CoV-2 rS or other Novavax baculovirus-Sf9-based vaccines taken into nonclinical evaluation or clinical trials.

No risks have been identified in nonclinical or early clinical testing of SARS-CoV-2 or other coronavirus vaccines (SARS-CoV and MERS-CoV) developed using the baculovirus-Sf9 system to date. In supportive toxicology studies with other viral GP nanoparticle vaccines developed using the baculovirus-Sf9 system with different antigens, findings were generally consistent with an immune response to the vaccine formulations. These toxicological investigations indicated that baculovirus-Sf9-produced antigens (up to 240 µg total nanoparticle dose) with Matrix-M1 adjuvant (up to 100 µg) were well tolerated in the animal and antigen system tested with no evidence of toxicity suggestive of any unusual risk or target organ for toxicity. Non-adverse findings, including local injection site inflammation and serum chemical markers of inflammation (such as C-reactive protein), were transient and considered consistent with immune system stimulation consequent to immunisation.

Findings to date suggest that SARS-CoV-2 rS when administered with or without Matrix-M1 adjuvant can be reasonably expected to demonstrate an acceptable safety profile in healthy adult participants aged ≤ 59 years. Novavax baculovirus-Sf9-produced nanoparticle vaccines comprising viral glycoproteins, with and without Matrix-M1 or aluminum adjuvants, have been shown to induce robust and protective immune responses in relevant animal models to influenza HAs, RSV F protein, SARS-CoV and MERS-CoV S proteins, rabies GP, and EBOV GP. In addition, the Novavax SARS-CoV-2 candidate adsorbed to aluminum phosphate has induced antibodies in pregnant women which, when transferred transplacentally, were associated with reduced rates of SARS-CoV-2 lower respiratory tract infections in their infants during the first 90 to 180 days of life. The goal of this program is to investigate the efficacy, safety, and immunogenicity of the SARS-CoV-2 rS and Matrix-M1 adjuvant.

Further details are provided in the SARS-CoV-2 rS IB.

### 2. STUDY OBJECTIVES AND ENDPOINTS

#### 2.1 Study Objectives

##### 2.1.1 Primary Objective

- To demonstrate the efficacy of SARS-CoV-2 rS with Matrix-M1 adjuvant in the prevention of virologically confirmed (by polymerase chain reaction [PCR]) to SARS-CoV-2, symptomatic COVID-19, when given as a 2-dose vaccination regimen, as compared to placebo, in serologically negative (to SARS-CoV-2) adult participants.

##### 2.1.2 Secondary Objectives

- To demonstrate the efficacy of SARS-CoV-2 rS with Matrix-M1 adjuvant in the prevention of virologically confirmed [by PCR to SARS-CoV-2], symptomatic COVID-19, when given as a 2-dose vaccination regimen, as compared to placebo, in adult participants regardless of their serostatus at baseline.
- To assess the efficacy of SARS-CoV-2 rS with Matrix-M1 adjuvant on SARS-CoV-2 seropositive adult participants requiring specific medical interventions as compared to placebo.
- To assess the efficacy of SARS-CoV-2 rS with Matrix-M1 adjuvant on mild COVID-19 symptoms as compared to placebo.
- To assess the efficacy of SARS-CoV-2 rS with Matrix-M1 adjuvant on occurrence of asymptomatic or undetected infections with SARS-CoV-2 as compared to placebo.
- In a subset of adult participants, to evaluate the immunogenicity of SARS-CoV-2 rS with Matrix-M1 adjuvant as compared to placebo.
- To evaluate safety in terms of SAEs and medically attended adverse events (MAAEs) in all adult participants during the entire study period.
- To evaluate safety in terms of AESI, which encompasses potential immune-mediated medical conditions (PIMMCs) and AESIs relevant to COVID-19 including possible vaccine-enhanced disease, in all adult participants at any time after the first dose.
- In a subset of adult participants, to evaluate the safety and reactogenicity in terms of solicited local and systemic adverse events (AEs) for 7 days after each study vaccination, and in terms of unsolicited AEs for 21 days after first study vaccination and 28 days after second study vaccination.

##### 2.1.3 Exploratory Objective

- In a subset of adult participants, to evaluate the safety and immunogenicity of SARS-CoV-2 rS with Matrix-M1 adjuvant when co-administered with a licensed seasonal influenza vaccine.

#### 2.2 Study Endpoints

##### 2.2.1 Primary Endpoints

There are 2 independent primary endpoints.

- **FIRST PRIMARY ENDPOINT:** First occurrence of virologically confirmed (by PCR to SARS-CoV-2), symptomatic COVID-19 (Table 2-1) with onset at least 7 days after second study vaccination (e.g., Day 28) in serologically negative (to SARS-CoV-2) adult participants at baseline until the endpoint-driven efficacy analysis is triggered by the occurrence of a prespecified number of blinded endpoints.
- **SECOND PRIMARY ENDPOINT:** First occurrence of virologically confirmed (by PCR to SARS-CoV-2), symptomatic moderate or severe COVID-19 (Table 2-1) with onset at least 7 days after second study vaccination (e.g., Day 28) in serologically negative (to SARS-CoV-2) adult participants at baseline until the endpoint-driven efficacy analysis is triggered by the occurrence of a prespecified number of blinded endpoints.

**Table 2-1: Endpoint Definitions of COVID-19 Severity**

| COVID-19 Severity | Endpoint Definitions |
|---|---|
| **Virologically Confirmed** | ≥ 1 COVID-19 disease symptom in Table 2-2<br>**AND**<br>Does not meet criteria for mild, moderate, or severe disease |
| **Mild** | ≥ 1 of:<br>• Fever (defined by subjective or objective measure, regardless of use of anti-pyretic medications)<br>• New onset cough<br>• ≥ 2 COVID-19 respiratory/non-respiratory symptoms in Table 2-2<br>**AND**<br>• Does not meet criteria for moderate or severe disease |
| **Moderate** | ≥ 1 of:<br>• Fever (defined by subjective or objective measure, regardless of use of anti-pyretic medications) + any 2 COVID-19 symptoms in Table 2-2 for ≥ 3 days (need not be contiguous days)<br>• High fever (≥ 38.4°C) for ≥ 3 days (need not be contiguous days)<br>• Any evidence of significant LRTI:<br>– Shortness of breath (or breathlessness or difficulty breathing) with or without exertion (greater than baseline)<br>– Tachypnea: 20 to 29 breaths per minute at rest<br>– SpO2: 94% to 95% on room air<br>– Abnormal chest x-ray or chest CT consistent with pneumonia or LRTI<br>– Adventitious sounds on lung auscultation (e.g., crackles/rales, wheeze, rhonchi, pleural rub, stridor)<br>**AND** |

**Table 2-1: Endpoint Definitions of COVID-19 Severity**

| COVID-19 Severity | Endpoint Definitions |
|---|---|
| | • Does not meet criteria for severe disease |
| **Severe** | ≥ 1 of:<br>• Tachypnea: ≥ 30 breaths per minute at rest<br>• Resting heart rate ≥ 125 beats per minute<br>• $SpO_2$: ≤ 93% on room air or $PAO_2/FiO_2$ < 300<br>• High flow oxygen therapy or NIV/NIPPV (e.g., CPAP or BiPAP)<br>• Mechanical ventilation or ECMO<br>• One or more major organ system dysfunction or failure (e.g., cardiac/circulatory, pulmonary, renal, hepatic, and/or neurological, to be defined by diagnostic testing/clinical syndrome/interventions), including any of the following:<br>o ARDS<br>o Acute renal failure<br>o Acute hepatic failure<br>o Acute right or left heart failure<br>o Septic or cardiogenic shock (with shock defined as SBP < 90 mm Hg OR DBP < 60 mm Hg<br>o Acute stroke (ischemic or hemorrhagic)<br>o Acute thrombotic event: AMI, DVT, PE<br>o Requirement for: vasopressors, systemic corticosteroids, or hemodialysis.<br>• Admission to an ICU<br>• Death |

Abbreviations: AMI = acute myocardial infarction; ARDS = acute respiratory distress syndrome; BiPAP = bi-level positive airway pressure; CPAP = continuous positive air pressure; CT = computed tomography; DBP = diastolic blood pressure; DVT = deep vein thrombosis; ECMO = extracorporeal membrane oxygenation; $FiO_2$ = fraction of inspired oxygen; ICU = intensive care unit; LRTI = lower respiratory tract infection; NIV = non-invasive ventilation; NIPPV = non-invasive positive pressure ventilation; $PAO_2$ = partial pressure of oxygen in the alveolus; PE = pulmonary embolism; SBP = systolic blood pressure; $SpO_2$ = oxygen saturation.

**Table 2-2: Qualifying Symptoms of Suspected COVID-19**

| |
|---|
| • Fever or chills<br>• Cough<br>• Shortness of breath or difficulty breathing<br>• Fatigue<br>• Muscle or body aches<br>• Headache<br>• New loss of taste or smell<br>• Sore throat<br>• Congestion or runny nose<br>• Nausea or vomiting<br>• Diarrhea |

Abbreviations: COVID-19 = coronavirus disease 2019.

##### 2.2.2 Secondary Endpoints

- First occurrence of virologically confirmed (by PCR to SARS-CoV-2), symptomatic COVID-19, with onset at least 7 days after second study vaccination (e.g., Day 28) in adult participants regardless of their serostatus at baseline.
- First occurrence of laboratory-confirmed (by PCR or serology to SARS-CoV-2) symptomatic or asymptomatic COVID-19 with onset at least 7 days after second study vaccination (e.g., Day 28) in adult participants regardless of their serostatus at baseline.
- First occurrence of COVID-19 requiring hospitalisation, intensive care unit (ICU) admission or mechanical ventilation linked to any virologically confirmed (by PCR to SARS-CoV-2) COVID-19 with onset at least 7 days after second study vaccination (e.g., Day 28) in adult participants regardless of their serostatus at baseline.
- First occurrence of virologically confirmed (by PCR to SARS-CoV-2), mild COVID-19 (with no progression to moderate or severe COVID-19 during the course of the COVID-19 episode) with onset at least 7 days after second study vaccination (e.g., Day 28) in adult participants regardless of their serostatus at baseline.
- Analysis of antibodies binding to the SARS-CoV-2 spike (S) protein by enzyme-linked immunosorbent assay (ELISA) at Day 0 (baseline), Day 21 (21 days after first study vaccination), and Day 35 (14 days after second study vaccination).
- The occurrence and relationship to study vaccination of SAEs and MAAEs (in all adult participants) during the entire study period.

##### 2.2.3 Exploratory Endpoints

- First occurrence of virologically confirmed (by PCR to SARS-CoV-2), symptomatic COVID-19 with onset at least 14 days after first study vaccination (e.g., Day 14) in serologically negative (to SARS-CoV-2) adult participants at baseline until the endpoint-driven efficacy analysis is triggered by the occurrence of a prespecified number of blinded endpoints.
- Any occurrence of serologic conversion (by serology to SARS-CoV-2 nucleocapsid (N) protein) between baseline and 1 year after last study vaccination in adult participants seronegative at baseline.
- Analysis of the proportion of adult participants with evidence of seroconversion as demonstrated by antibodies to the SARS-CoV-2 S protein by ELISA at Day 35 (14 days after second study vaccination).
- Analysis of the cell-mediated immune responses (for example, as measured by enzyme-linked immune absorbent spot (ELISpot) ± intracellular cytokine staining) on Days 0 (baseline) and 35 (14 days after second study vaccination).

- SARS-CoV-2 neutralisation as measured by virus neutralisation assay (VNA; wild-type virus and/or pseudovirion expressing SARS-CoV-2 S protein) at Day 0 (baseline) and Day 35 (14 days after second study vaccination).
- Analysis of the safety and immunogenicity of SARS-CoV-2 rS with Matrix-M1 adjuvant when co-administered with seasonal influenza vaccine in a subset population.

### 3. STUDY DESIGN

This is a Phase 3, multi-centre, randomised, observer-blind, placebo-controlled study evaluating the efficacy, safety, and immunogenicity of SARS-CoV-2 rS with Matrix-M1 adjuvant in adult participants 18 to 84 years of age (inclusive) in the UK. Every effort will be made to identify regions of high SARS-CoV-2 activity, and populations within these regions who are at high risk of exposure to the virus will be enrolled such that the incidence of COVID-19 in the study will be higher than 1% and the sample size and enrolment period may be reduced.

After signing informed consent, participants may be screened within a window of up to approximately 30 days. Mucosal samples (nasal or throat swabs or saliva based on test availability) may be taken during the screening period to detect PCR for SARS-CoV-2, if the participant has any COVID-19 symptoms or significant exposure history.

Approximately 9,000 male and female adult participants 18 to 84 years of age (inclusive) with and without relevant comorbidities is planned for the study. An effort will be made to enrol a target of at least 25% of participants who are ≥ 65 years of age as well as prioritising other groups that are most affected by COVID-19, including racial and ethnic minorities. This will be under the assumption that the annual incidence of symptomatic COVID-19 will be approximately 1% to 5%. Enrolment of participants ≥ 65 years of age will not be initiated until approval from the Medicines and Healthcare products Regulatory Agency (MHRA) following their review of Phase 2 study safety data in this age group.

The actual sample size for the study will be selected at the operational cut-off date before initiation of the study, based on estimated incidences for the targeted study regions and populations at that time. The sample size may be adjusted by the sponsor during the study, based on blinded data, to ensure sufficient power at the time of the primary analysis. Details on the possible sample-size reassessment will be described in the statistical analysis plan (SAP).

On Day 0, mucosal samples (nasal or throat swabs or saliva based on test availability) will be taken to detect PCR for SARS-CoV-2 and blood samples will be taken to detect any existing SARS-CoV-2 antibodies at baseline. Participants will be randomised in a 1:1 ratio via block randomisation to receive 2 intramuscular (IM) injections of the SARS-CoV-2 rS with Matrix-M1 adjuvant or placebo, as described in Figure 1. Randomisation will be stratified by site and by age ≥ 65 years. In addition, there will not be specific age sub-stratification recruitment targets within the ≥ 65-year age group, but through site recruitment; the aim is to allow a generally representative age distribution overall.

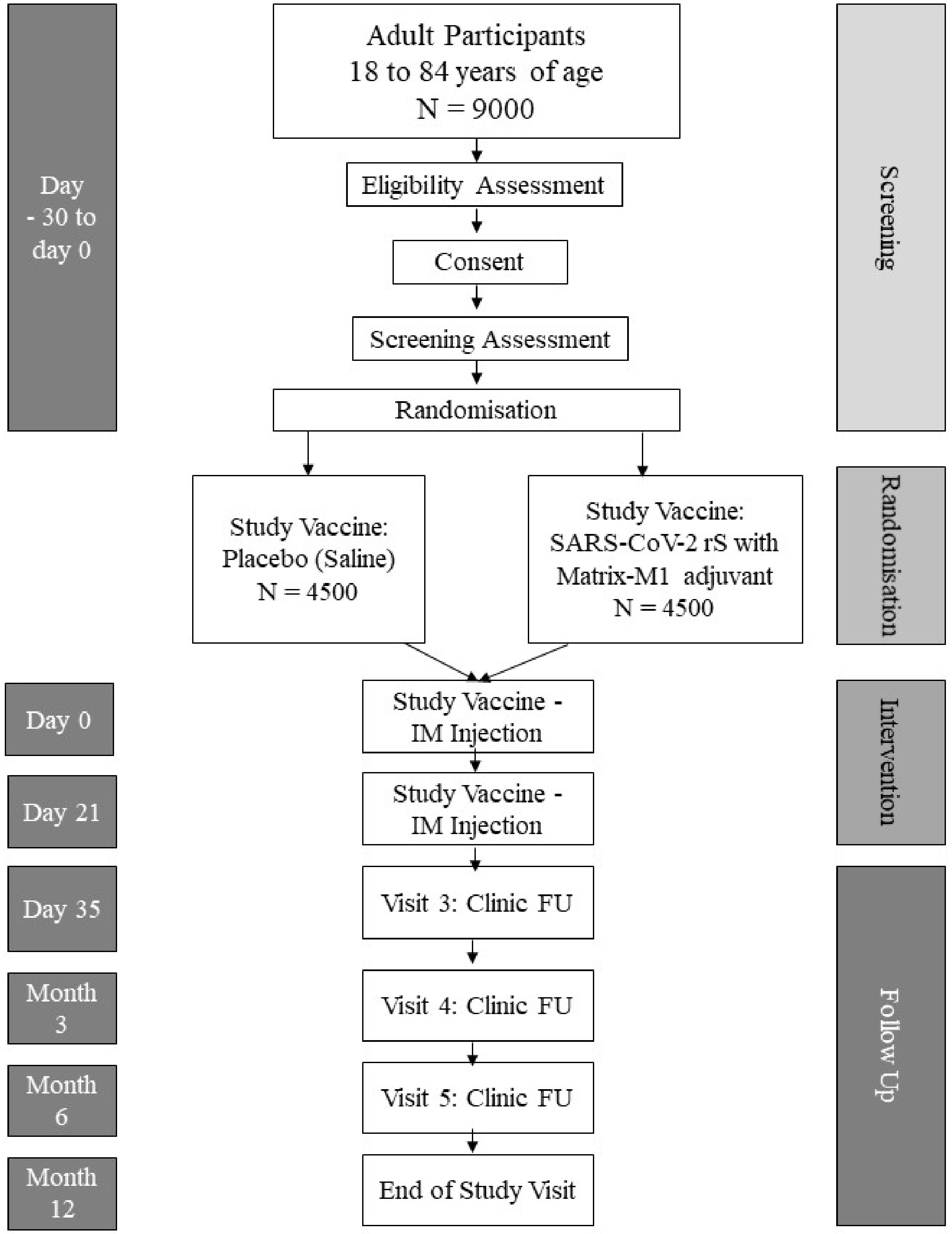

**Figure 1: Trial Schema**

FU = follow-up; IM = intramuscular; N = number of participants.

The study will consist of the screening period (Days -30 to 0); study vaccination Days 0 and 21 (+7-day window); and outpatient study visits on Day 0, Day 21 (+ 7 days), and Day 35 (14 days minimum after second study vaccination [+ 7 days]); and at 3, 6, and 12 months (± 15 days) after last study vaccination.

The duration of individual participation, including screening, will be a maximum of 1 year after second study vaccination (Day 386 ± 15 days). This will be an event-driven study for the assessment of vaccine efficacy (VE) and will end when a sufficient number of events have been observed, yet all participants will be followed for the entire study duration for safety endpoints.

A licensed seasonal influenza co-administration sub-study will be conducted in the first 400 participants who meet the additional inclusion criteria for this study. Participants may be enrolled at select study sites due to the availability of seasonal influenza vaccine. After being randomised to receive IM injections of SARS-CoV-2 rS with Matrix-M1 adjuvant or placebo, sub-study participants will receive a licensed seasonal influenza vaccine on Day 0 in the opposite deltoid. These participants will be part of the solicited AE safety subset analysis.

Given that this study and others will enrol during the influenza virus season and that the highest at-risk population, namely the elderly and adults with comorbid conditions for influenza mirrors that of SARS-CoV-2, a co-vaccination sub-study is included. This sub-group will undergo the same safety and efficacy evaluations as the entire study population. In addition, the entire sub-study will have reactogenicity assessed. This group must not have had a current season licensed influenza vaccine and have no prior history of allergy or severe reaction to seasonal influenza vaccine. To assess the possible impact of the study vaccine on the immunogenicity of the influenza vaccine, this group will also have a hemagglutination inhibition assay (HAI) performed.

A global Safety Monitoring Committee (SMC) was convened to oversee the safety of the ongoing Phase 1/2 study (Protocol 2019nCoV-101) and will oversee one or more additional studies across the SARS-CoV-2 rS vaccine program. A separate SMC will be convened for this study. The designated SMC will monitor the safety of participants in the study and will follow an SMC charter. The SMC will review blinded and unblinded safety and reactogenicity data. The SMC will also review sufficient clinical event data to ensure detection of any evidence of enhanced disease. The SMC will convene to perform safety reviews on a scheduled basis, for immediate concerns regarding safety observations during this study, and as needed.

#### 3.1 Study Vaccination Pause Rules

Study vaccination pause rules based on reactogenicity, AEs, and SAEs related to study participation will be in place to monitor participant safety during the study.

AEs meeting any one of the following criteria will result in a hold being placed on subsequent study vaccinations pending further review by the SMC and the sponsor:

- The occurrence of 1 or more related SAEs (final assessment by the sponsor per United States Food and Drug Administration Center for Biologics Evaluation and Research Guidance) in a given Medical Dictionary for Regulatory Activities (MedDRA) system organ class within the first 7 days after study vaccination with the exception of hypersensitivity (anaphylaxis), which would require 2 occurrences.
- Any toxicity grade 3 (severe) solicited (local or systemic) single AE term occurring in ≥ 10% of participants in the SARS-CoV-2 rS with Matrix-M1 adjuvant group within the first 7 days after study vaccination.
- Any grade 3 (severe) unsolicited single AE preferred term for which the investigator assesses as related that occurs in ≥ 5% of participants in the SARS-CoV-2 rS with Matrix-M1 adjuvant group, within the first 7 days after study vaccination.

The sponsor, along with medical monitor, may request an SMC review for any safety concerns that may arise in the study, even if they are not associated with any specific pause rule.

#### 3.2 Schedule of Events (SOE)

Table 3-1 lists the study procedures that will be performed during the study. Detailed descriptions of each visit are presented in Section 6.1.

**Table 3-1 Schedule of Events**

| **Study Period:** | **Screening Period** [a] | **Clinic Visits** | | | | **Months After Last Study Vaccination** | | |
|---|---|---|---|---|---|---|---|---|
| **Study Day:** | **–30 to 0** | **0**[a] | **21** | **35** | **COVID-19 Surveillance Visits (Unscheduled)** | **3** | **6** | **12** |
| **Window (days):**[b] | **–** | **0** | **+ 7** | **+ 7** | | **± 15** | **± 15** | **± 15** |
| **Minimum days following most recent study vaccination:**[b] | **–** | **0** | **21** | **14** | | **–** | **–** | **–** |
| **Study Visit:** | **Screening** | **1** | **2** | **3** | | **4** | **5** | **EOS** [c] |
| Informed consent | X | | | | | | | |
| Medical history [d] | X | | | | X | | | |
| Inclusion/exclusion criteria [e] | X | X [f] | X [f] | | | | | |
| Demographics [g] | X | | | | | | | |
| Prior/concomitant medications [h] | X | X [f] | X [f] | X | X | X | X | X |
| Vital sign measurements [i] | X | X | X | | X | | | |
| Urine pregnancy test (WOCBP) [j] | X | X [f] | X [f] | | | | | |
| Physical examination (targeted) [k] | X | X [f] | X [f] | X | X | | | |
| Mucosal testing for SARS-CoV-2 (PCR) | X | X [f] | X [f] | | X [y] | | | |
| Blood sampling for SARS-CoV-2 (ELISA for anti N-protein serology) | | X [f] | | X | | X | X | X |
| Blood sampling for SARS-CoV-2 (ELISA for anti S-protein serology – subset of participants) [l] | | X [f] | | X | | | | |
| Blood sampling for SARS-CoV-2 neutralisation assay (subset) [m] | | X [f] | | X | | | | |
| Blood sampling for HAI (influenza co-administration subset) [n] | | X [f] | X | | | | | |
| Cell-mediated assessments (subset of participants) [o] | | X [f] | | X | | | | |
| Randomisation | | X | | | | | | |
| Study vaccination [p] | | X | X | | | | | |
| Reactogenicity (subset of participants) [q] | | X | X | | | | | |
| Monitoring for COVID-19 [r] | | COVID-19 case ascertainment will commence from Day 0 until EOS | | | | | | |
| COVID-19 Symptom Diary [s] | | | | | X | | | |

**Table 3-1 Schedule of Events**

| Study Period: | Screening Period [a] | Clinic Visits | | | | Months After Last Study Vaccination | | |
|---|---|---|---|---|---|---|---|---|
| Study Day: | –30 to 0 | 0[a] | 21 | 35 | COVID-19 Surveillance Visits (Unscheduled) | 3 | 6 | 12 |
| Window (days):[b] | – | 0 | + 7 | + 7 | | ± 15 | ± 15 | ± 15 |
| Minimum days following most recent study vaccination:[b] | – | 0 | 21 | 14 | | – | – | – |
| Study Visit: | Screening | 1 | 2 | 3 | | 4 | 5 | EOS [c] |
| All unsolicited AEs [t] | | X | X | X | | | | |
| MAAEs [u] | | X | X | X | X | X | X | X |
| SAEs [v] | X | X | X | X | X | X | X | X |
| AESI [w] | | X | X | X | X | X | X | X |
| EOS form [x] | | | | | | | | X |

Abbreviations: AE = adverse event; AESI = adverse event(s) of special interest; COVID-19 = coronavirus disease 2019; ELISA = enzyme-linked immunosorbent assay; ELISpot = enzyme-linked immune sorbent spot; EOS = end of study; HAI = hemagglutinin assay inhibition; HEENT = head, eye, ear, nose, and throat (exam); ID = identification; MAAE = medically attended adverse event; N = nucleocapsid; PCR = polymerase chain reaction; PIMMC = potential immune-mediated medical conditions; S = spike protein; SAE = serious adverse event; SARS-CoV-2 = severe acute respiratory syndrome coronavirus 2; WOCBA = women of childbearing age.

[a] The Screening visit and Day 0 visit may be combined if feasible at any given study site.

[b] Days relative to study vaccination are only estimates because the window allowance is not inclusive. Should a study pause occur, visits/windows will be adjusted to allow participants to continue without protocol deviation. Visit schedules after second study vaccination are calculated relative to the day the study vaccinations were received.

[c] EOS telephone call. Should participants decide to terminate early, an EOS telephone call will occur to collect the maximum safety data possible.

[d] Including prior and concomitant medical conditions, recent vaccinations (≤ 90 days), and significant surgical procedures.

[e] Specific exclusions to study vaccination will be assessed before any vaccination. Waivers to enrolling participants with exclusions will not be given.

[f] Performed prior to study vaccination.

[g] Screening only. Including date of birth (day, month, and year), sex, race, ethnicity, weight, and height.

[h] Recent and current medications at the time of screening to be reviewed to ensure eligibility criteria are fulfilled. Concomitant medications include all medications (including vaccines) taken by the participant.

[i] Including respiratory rate, blood pressure, pulse rate, pulse oximetry and oral temperature (or via forehead/ear reader). On study vaccination days, vital sign measurements will be collected once before study vaccination to ensure participant has no evidence of fever prior to study vaccination. Vital sign measurements will also be collected once again, approximately 15 to 30 minutes after study vaccination, to check for any reactions to the vaccine.

[j] Women of childbearing potential only. A urine pregnancy test will be performed at Screening and prior to each study vaccination. A positive urine pregnancy test at any time will result in the participant not receiving any further study vaccination. A positive urine pregnancy test at Screening will result in screen failure.

[k] Examination at screening to include height and weight; HEENT, neck, thyroid, lungs, heart, cardiovascular, abdomen, lymph nodes of the upper extremities and neck; and musculoskeletal system/extremities to allow for study vaccination; symptom-directed (targeted) physical examination at all other scheduled time points but always to include lymphatic assessment of injected upper extremity on study vaccination days. Physical examination on study vaccination visits must be done prior to vaccination. Interim physical examinations will be performed at any unscheduled visit at the discretion of the investigator, if necessary.

[l] The ELISA for anti-S protein serology will be performed in the approximately 900 participants in the Anti-S Protein Serology Subset.

[m] The neutralising antibody assay will be performed in the approximately 900 participants in the Neutralisation Assay Subset.

**Table 3-1 Schedule of Events**

| Study Period: | Screening Period [a] | Clinic Visits | | | | Months After Last Study Vaccination | | |
|---|---|---|---|---|---|---|---|---|
| Study Day: | –30 to 0 | 0[a] | 21 | 35 | COVID-19 Surveillance Visits (Unscheduled) | 3 | 6 | 12 |
| Window (days):[b] | – | 0 | + 7 | + 7 | | ± 15 | ± 15 | ± 15 |
| Minimum days following most recent study vaccination:[b] | – | 0 | 21 | 14 | | – | – | – |
| Study Visit: | Screening | 1 | 2 | 3 | | 4 | 5 | EOS [c] |

[n] The HAI assay will be performed in the approximately 400 participants in the licensed seasonal influenza co-administration sub-study.

[o] Cell-mediated immune responses, as assessed by ELISpot ± intracellular cytokine staining, will be performed in approximately 450 participants in the Cell-mediated Assay Subset.

[p] Study vaccination on Day 0 will consist of study vaccine plus, in the seasonal influenza vaccine co-administration sub-study, a single dose of licensed influenza vaccine. Study vaccination on Day 21 will consist of study vaccine.

[q] Solicited AEs (e.g., local and general systemic reactogenicity symptoms) will be captured in the Safety Subset of approximately 2,000 participants and in the first approximately 400 participants who qualify for the seasonal influenza vaccine co-administration sub-study. On study vaccination days, participants will remain in clinic for at least 30 minutes to be monitored for any severe reactogenicity. Severe reactions will be noted as AEs on day of study vaccination.

[r] Participants will monitor for COVID-19 symptoms. This will include provision of a Study ID Card that provides details on study participation, study site contact information and assessment of symptoms of suspected COVID-19 (see Table 2-2). Instructions to participants to contact the study site within 24 hours for symptoms of suspected COVID-19 will be given. A participant with suspected or confirmed COVID-19 will be asked to complete a COVID-19 symptom diary for 10 days. The Study ID Card should be presented to healthcare providers not affiliated with the study who encounter participants with symptoms of suspected COVID-19.

[s] A participant with suspected or confirmed COVID-19 will be asked to complete the COVID-19 symptom diary. Participants should start the diary on their first day of symptoms and complete this diary daily for a minimum of 10 days even if their symptoms have resolved and regardless of any SARS-CoV-2 PCR results. They should continue the diary beyond 10 days if they have any ongoing symptoms related to the episode and should continue to do so until all symptoms have resolved.

[t] All unsolicited AEs are to be reported from the time of first study vaccination until 21 days after first study vaccination and 28 days after second study vaccination in all participants.

[u] MAAEs are to be collected from the time of first study vaccination until Day 35, and MAAEs assessed as related to study vaccination from the time of first study vaccination until completion of the participant's last study-related procedure.

[v] SAEs are to be collected from the time of informed consent until completion of the participant's last study-related procedure.

[w] AESI: To include PIMMC and AEs related to COVID-19 are to be collected from the time of first study vaccination until completion of the participant's last study-related procedure.

[x] EOS form will be completed for all participants, including participants who are terminated early.

[y] Samples will be collected in an effort to determine if the current symptoms are due to SARS-CoV-2 infection. Approximately 24 hours after the onset of symptoms the participants will be tested daily for up to 3 days but only until the point that 1 of the samples tests positive for SARS-CoV-2. Testing may occur at home or in the clinic as part of a COVID-19 Surveillance Visit. If the participant self-tested at home and has a clinic visit that same day, they should be sampled again by a member of the study staff. At Follow-up COVID-19 Surveillance Visits, participants with a prior positive PCR will not need to be tested again while participants with negative tests should be tested again.

### 4. STUDY POPULATION

Approximately 9,000 male and female adult participants 18 to 84 years of age (inclusive) with and without relevant comorbidities will be randomised in a 1:1 ratio via block randomisation to receive 2 IM injections of SARS-CoV-2 rS with Matrix-M1 adjuvant or placebo in a blinded fashion in up to 18 regions across the UK.

#### 4.1 Inclusion Criteria

Each participant must meet all of the following criteria to be enrolled in this study:

1. Adult males or females aged 18 to 84 years (inclusive) at screening.
2. Able and willing (in the investigator's opinion) to comply with all study requirements.
3. Willing to allow the investigators to discuss the volunteer's medical history with their General Practitioner and access all medical records when relevant to study procedures.
4. Willing and able to give informed consent prior to study enrolment.
5. Female participants of childbearing potential (defined as any female who has experienced menarche and who is NOT surgically sterile [i.e., hysterectomy, bilateral tubal ligation, or bilateral oophorectomy] or postmenopausal [defined as amenorrhea at least 12 consecutive months or > 1 documented plasma follicle-stimulating hormone level ≥ 40 mIU/mL]) must agree to be heterosexually inactive from at least 28 days prior to enrolment and through 3 months after the last study vaccination OR agree to consistently use any of the following methods of contraception from at least 28 days prior to enrolment and through 3 months after the last study vaccination:
   a. Condoms (male or female) with spermicide
   b. Diaphragm with spermicide
   c. Cervical cap with spermicide
   d. Intrauterine device
   e. Oral or patch contraceptives
   f. Norplant®, Depo-Provera®, or other in country regulatory-approved contraceptive method that is designed to protect against pregnancy
   g. Abstinence, as a form of contraception, is acceptable if in line with the participant's lifestyle

      **NOTE**: Periodic abstinence (e.g., calendar, ovulation, symptothermal, post-ovulation methods) and withdrawal are not acceptable methods of contraception.

**Seasonal Influenza Vaccine Co-Administration Sub-Study Only**

6. Participant should not have received a current season influenza vaccine, have no contraindication to the specific vaccine to be administered in the study, and no prior history of allergy or severe reaction to seasonal influenza vaccines.

#### 4.2 Exclusion Criteria

Participants meeting any of the following criteria will be excluded from the study:

1. Participation in COVID-19 prophylactic drug trials for the duration of the study.
2. Participation in SARS-CoV-2 serological surveys where participants are informed of their serostatus for the duration of the study.
3. Participation in research involving an investigational product (drug/biologic/device) within 45 days prior to first study vaccination.
4. History of laboratory-confirmed (by PCR or serology to SARS-CoV-2) COVID-19 infection at any time prior to randomisation.
5. Administration of immunoglobulins and/or any blood products within the 3 months preceding the planned administration of the study vaccine candidate.
6. Any confirmed or suspected immunosuppressive or immunodeficient state; chronic administration (defined as more than 14 continuous days) of immunosuppressant medication within the past 3 months, except topical steroids or short-term oral steroids (course lasting ≤ 14 days).

   **NOTE:** An immunosuppressant dose of glucocorticoid is defined as a systemic dose ≥ 10 mg of prednisone per day or equivalent. The use of topical, inhaled, and nasal glucocorticoids will be permitted if other chronic disease conditions are not exclusionary.
7. History of allergic disease or reactions likely to be exacerbated by any component of the study vaccines.
8. Any history of anaphylaxis to any prior vaccine.
9. Pregnancy, lactation or willingness/intention to become pregnant during the study.
10. Current diagnosis of or treatment for cancer (except basal cell carcinoma of the skin and cervical carcinoma in situ, at the discretion of the investigator).
11. Bleeding disorder (e.g., factor deficiency, coagulopathy or platelet disorder), or prior history of significant bleeding or bruising following IM injections or venepuncture.
12. Continuous use of anticoagulants, such as coumarins and related anticoagulants (i.e., warfarin) or novel oral anticoagulants/anti-platelet agents.

    **NOTE:** The use of ≤ 325 mg of aspirin per day as prophylaxis is permitted.
13. Suspected or known current alcohol or drug dependency.

14. Study team member or first-degree relative of any study team member (inclusive of sponsor, contract research organisation (CRO), and site personnel involved in the study).
15. Participants who are having any current workup of undiagnosed illness within the last 8 weeks that is either participant-reported or has been clinician-assessed, which could lead to a new condition or diagnosis.
16. Received any live vaccine within 4 weeks or **any vaccine** (excluding influenza) within 2 weeks prior to first study vaccination or any licensed influenza vaccine within 1 week prior to first study vaccination. Plans to receive any vaccine from these time periods until 28 days after second study vaccination (with the exception of participants in the seasonal influenza co-administration sub-study; a licensed seasonal influenza vaccine may be received as soon as 7 days after second study vaccination).

    **NOTE:** An influenza co-administration sub-study will occur in which 400 participants will receive a single dose of seasonal influenza vaccine at the same time as first study vaccination.
17. Have clinically significant chronic cardiovascular, endocrine, gastrointestinal, hepatic, renal, neurological, respiratory, psychiatric or other medical disorders not excluded by other exclusion criteria, that are assessed by the investigator as being clinically unstable within the prior 4 weeks as evidenced by:
    a. Hospitalisation for the condition, including day surgical interventions.
    b. New significant organ function deterioration.
    c. Needing addition of new treatments or major dose adjustments of current treatments (mild or moderate well-controlled comorbidities are allowed).
18. History of chronic neurological disorders that have required prior specialist physician review for diagnosis and management (such as multiple sclerosis, dementia, transient ischemic attacks, Parkinson’s disease, degenerative neurological conditions, neuropathy, and epilepsy) or a history of stroke or previous neurological disorder within 12 months with residual symptoms. Participants with a history of migraine or chronic headaches or nerve root compression that have been stable on treatment for the last 4 weeks are not excluded.
19. Any autoimmune or immunodeficiency disease/condition (iatrogenic or congenital).

    **NOTE:** Stable endocrine disorders that have a confirmed autoimmune etiology (e.g., thyroid, pancreatic) are allowed.
20. Any other significant disease, disorder or finding that, in the opinion of the investigator, may significantly increase the risk to the volunteer because of participation in the study, affect the ability of the volunteer to participate in the study, or impair interpretation of the study data.

#### 4.3 Other Considerations

Participants meeting any of the following criteria may have planned study vaccination deferred for a later date, but these criteria are not exclusionary for study enrolment.

- Respiratory symptoms in the past 3 days (i.e., cough, sore throat, difficulty breathing). Participant may be vaccinated once all symptoms have been resolved for > 3 days. Out-of-window study vaccination is allowed for this reason (**see NOTE**).
- Temperature of > 38°C within 24 hours of planned study vaccination (site measured or participant measured). Participant may be vaccinated once the fever has resolved and there has not been any temperature measured as being > 38°C for > 3 days. Out-of-window study vaccination is allowed for this reason (**see NOTE**).

**NOTE:** PCR testing for SARS-CoV-2 is likely to be indicated for either of the above reasons or if COVID-19 is suspected based on other symptoms, potential exposure to SARS-CoV-2 infection through either close contacts or based on local epidemiology. If a participant has a positive PCR for SARS-CoV-2 prior to enrolment that participant will not be eligible and will be considered a screen failure.

- Participants having any symptoms or signs of possible COVID-19 infection (Table 2-2) that may also be due to post-vaccination reactogenicity within 7 days of either study vaccine dose will NOT be required to be tested for SARS-CoV-2 PCR.
- Any participant with a new positive PCR-confirmed SARS-CoV-2 infection occurring from Screening and prior to second study vaccination will not be removed from the study but must meet health requirements before receiving the second study vaccination.
- Any participant who has a positive PCR for SARS-CoV-2 between the first and second study vaccination should also have PCR testing for SARS-CoV-2 on the day of the subsequent study vaccination, but the results of the PCR test are not needed before study vaccination can be given.
- Any participant with a new positive PCR-confirmed SARS-CoV-2 infection occurring from Screening and prior to the end of immunogenicity assessments will be removed from applicable immunogenicity analyses as defined in the SAP.
- Any acute illness (cardiovascular, endocrine, gastrointestinal, hepatic, renal, neurological, respiratory, or other medical disorders) that is actively causing symptoms that could, in the opinion of the investigator, impact the assessment of reactogenicity or other study assessments may have planned study vaccination deferred for a later date, but these criteria are not exclusionary for study enrolment. Participant may be vaccinated once symptoms have resolved or are

stabilised for > 3 days. Out-of-window study vaccination is allowed for this reason.

#### 4.4 Withdrawal of Participants from the Study

##### 4.4.1 Reasons for Withdrawal

Participants can withdraw consent and discontinue from the study at any time, for any reason. Participants may refuse further procedures (including study vaccination) but are encouraged to remain in the study for safety follow-up. In such cases where only safety is being conducted, participant contact could be managed via telemedicine contact (e.g., telephone, web chat, video, FaceTime).

The investigator may **withhold** further study vaccination from a participant in the study if the participant:

1. Is noncompliant with the protocol.
2. Experiences an SAE or intolerable AE(s) for which study vaccination is not advised by the investigator.

The investigator will withhold further study vaccination from a participant in the study if the participant:

3. Becomes pregnant (discontinuation of further study vaccination required).

The investigator can also withdraw a participant upon the request of the sponsor or if the sponsor terminates the study. Upon the occurrence of an SAE or intolerable AE, the investigator may confer with the sponsor before future study vaccination.

##### 4.4.2 Handling of Withdrawals

Participants are free to withdraw from the study at any time upon request. Participant participation in the study may be stopped at any time at the discretion of the investigator or at the request of the sponsor.

When a participant withdraws from the study, the reason(s) for withdrawal shall be recorded by the investigator on the relevant page of the electronic case report form (eCRF). Whenever possible, any participant who withdraws from the study prematurely will undergo all EOS assessments. Any participant who fails to return for final assessments will be contacted by the site in an attempt to have them comply with the protocol. The status of participants who fail to complete final assessments will be documented in the eCRF.

##### 4.4.3 Replacements

Participants who withdraw, are withdrawn or terminated from this study, or are lost to follow-up after signing the ICF but prior to first study vaccination may be replaced.

Participants who receive study vaccine and subsequently withdraw, are discontinued from further study vaccination, are terminated from the study, or are lost to follow-up will not be replaced.

### 5. TEST ARTICLES

#### 5.1 Study Vaccines Administered

Study vaccinations (SARS-CoV-2 rS with Matrix-M1 adjuvant or placebo) will comprise 2 IM injections (Day 0 and Day 21), ideally in alternating deltoids. For blinding purposes, all participants will be vaccinated using the same injection volume (i.e., 0.5 mL). The dose level will be 5 µg SARS-CoV-2 rS with 50 µg Matrix-M1 adjuvant (co-formulated in a single vial); placebo will be saline for injection. Study vaccinations will be administered on an outpatient basis by designated site personnel in a way to maintain the blind. Any pharmacy preparation with unblinded product will require unblinded site personnel who will not otherwise be involved in the study procedures or observation of participants.

The seasonal influenza vaccine co-administration sub-study will comprise a single IM injection (0.5 mL) of a licensed influenza vaccine on Day 0, ideally in the opposite deltoid of the study vaccine. Seasonal influenza vaccine will be administered in an open-label manner.

#### 5.2 Investigational Products

The following supplies will be used for vaccination in the study:

| Investigational Product | Supplied Formulation |
|---|---|
| SARS-CoV-2 rS with Matrix-M1 adjuvant | Solution for preparation for injection, at a concentration of 5 µg antigen and 50 µg adjuvant. |
| Placebo | Sodium chloride injection (BP, sterile), 0.9% |
| **Seasonal Influenza Vaccine Co-Administration Sub-Study** | |
| Licensed seasonal influenza vaccine | Single or multi-dose vial of licensed seasonal influenza vaccine |
| Abbreviations: BP = British Pharmacopoeia; SARS-CoV-2 rS = severe acute respiratory syndrome coronavirus 2 recombinant spike protein nanoparticle vaccine. | |

It is anticipated that the product will be available in a co-formulated single vial.

Further details on the study vaccine can be found in the SARS-CoV-2 rS IB.

##### 5.2.1 Investigational Product Packaging and Storage

Novavax, Inc. will provide adequate quantities and appropriate labelling of SARS-CoV-2 rS with Matrix-M1 adjuvant and PPD will ensure distribution to the clinical sites from a designated depot. Sodium chloride injection (British Pharmacopoeia, sterile) and licensed seasonal influenza vaccine are commercially available and will be supplied by PPD. The clinical unit pharmacy will prepare the study vaccines for each participant. Detailed instructions for the preparation of study vaccine will be provided in a separate pharmacy manual.

All investigational products must be stored according to the labelled instructions in a secure cabinet or room with access restricted to necessary clinic personnel. The site will be required to keep a temperature log to establish a record of compliance with storage conditions.

##### 5.2.2 Investigational Product Accountability

The investigator (or delegate) will maintain accurate records of receipt of all investigational product, including dates of receipt. Accurate records will be kept regarding when and how much investigational product is dispensed and used by each participant in the study. Reasons for departure from the expected dispensing regimen must also be recorded. At the completion of the study, and to satisfy regulatory requirements regarding investigational product accountability, all investigational product will be reconciled and retained or destroyed according to applicable regulations. No investigational product will be destroyed until authorised in writing by the sponsor.

#### 5.3 Method of Assigning Participants to Study Vaccine Groups

Participants will be randomly assigned in a blinded manner using the centralised Interactive Response Technology (IRT) according to pre-generated randomisation schedules. Participants will be randomised in a 1:1 ratio via block randomisation to receive 2 IM injections of the SARS-CoV-2 rS with Matrix-M1 adjuvant or placebo. Randomisation will be stratified by site and by age ≥ 65 years. The first approximately 400 participants who qualify for the seasonal influenza vaccine co-administration sub-study (which may be at select study locations only) will be assigned prior to randomisation. These participants will be part of the solicited AE safety subset analysis. Details regarding the IRT process will be provided separately to the sites.

##### 5.3.1 Blinding Procedures

This is an observer-blinded study. To maintain the blind, placebo vaccination via IM route will be included and unblinded site personnel will manage study vaccine logistics, preparation, and administration so as to maintain the blind from the remainder of the site personnel and participants. The unblinded site personnel will not be involved in study-related assessments or have participant contact for data collection following study vaccine administration.

Seasonal influenza vaccine will be administered in an open-label manner.

##### 5.3.2 Breaking the Blind

A participant's study vaccine assignment will not be broken until the end of the study for the clinical site study team unless medical treatment of the participant depends on knowing the study vaccine the participant received. In the event that the blind needs to be broken because of a medical emergency, the investigator may unblind an individual participant's study vaccine allocation.

Whenever possible, the investigator should contact the medical monitor to discuss the medical emergency and the reason for revealing the actual study vaccine received by that participant. In the event that the investigator cannot contact the medical monitor in a timely manner the blind may be broken by the investigator. The medical monitor should be contacted as soon as feasible after the unblinding. The study vaccine assignment will be unblinded through IRT. Reasons for study vaccine unblinding must be clearly explained and justified in the eCRF. The date on which the code was broken together with the identity of the person responsible must also be documented.

In the event that a safe and effective vaccine against SARS-CoV-2 is licensed and made widely available during the course of the study, a discussion with appropriate regulatory agencies will take place to discuss if the blind should be broken to offer vaccine to placebo participants.

The blind may also be broken in the event of a Suspected Unexpected Serious Adverse Reaction (SUSAR) to determine regulatory reporting.

In addition to the aforementioned situations where the blind may be broken, the data will also be unblinded to a statistical team at specified time points for planned analyses prior to study completion, as outlined in Section 7.5.

#### 5.4 Study Vaccine Compliance

All doses of the study vaccine should be administered in the clinical unit under direct observation of clinic personnel and recorded in the eCRF but may need to occur outside of the clinical site depending on the pandemic situation (e.g., home study vaccinations). Home study vaccination visits must have adequate oversight for issues associated with immediate severe reactions. Clinic personnel will confirm that the participant has received the entire dose.

The location (right or left arm), date, and timing of all doses of study vaccine will be recorded in the participants' eCRF. If a participant is not administered study vaccine, the reason for the missed dose will be recorded.

#### 5.5 Concomitant Medications and Prohibitive Therapy

##### 5.5.1 Concomitant Medications

Administration of medications, therapies, or vaccines will be recorded in the eCRF. Concomitant medications will include all medications (including vaccines) taken by the participant from the time of signing the ICF through EOS (or through the early termination visit if prior to that time). Prescription and over-the-counter (OTC) drugs, as well as herbals, vitamins, and supplements, will be included.

##### 5.5.2 Prohibitive Therapy

- No live vaccine will be allowed within 4 weeks of first study vaccination until 28 days after second study vaccination (Day 49).
- No vaccine (except for a licensed seasonal influenza vaccine and participants in the seasonal influenza co-administration sub-study) will be allowed within 2 weeks of first study vaccination until 28 days after second study vaccination (Day 49).
- No influenza vaccine (except participants in the seasonal influenza co-administration sub-study) will be allowed within 1 week of first study vaccination until 7 days after second vaccination (Day 28).

  **NOTE:** Participants in the seasonal influenza co-administration sub-study will be allowed to have the co-administration of a licensed seasonal influenza vaccine at the same time as first study vaccination.
- No unlicensed vaccine should be given within 45 days prior to first study vaccination until after the last study visit.
- No investigational product (drug/biologic/device) within 45 days prior to first study vaccination until after the last study visit.
- No chronic administration (defined as more than 14 continuous days) of any immunosuppressant medication within 3 months of first study vaccination until the last study visit (except topical steroids or short-term oral steroids (course lasting ≤ 14 days). The use of topical, inhaled, and nasal glucocorticoids will be permitted if other chronic disease conditions are not exclusionary.
- No continuous use of anticoagulants, such as coumarins and related anticoagulants (i.e., warfarin) or novel oral anticoagulants/anti-platelet agents. Use of ≤ 325 mg of aspirin per day as prophylaxis is permitted.

### 6. STUDY PROCEDURES

Written informed consent will be obtained after explanation of the aims, benefits and all safety concerns of the trial as detailed in the information sheet BEFORE any trial specific procedures are performed. They should take as much time as they need to consider joining the study. Signed consent will be kept by the investigator and documented in medical notes and a copy given to the participant, as described in Section 9.2.2.3 (Appendix 2).

Due to the ongoing pandemic, recent national regulatory and local Ethics Committee and public health guidance will be applied at the site locations regarding alternations in the ability of study participants to attend an investigational site for protocol-specified visits, with the site's investigator being allowed to conduct safety assessments (e.g., telephone contact, alternative location for assessment, including local laboratories or imaging centers) when necessary and feasible, as long as such visits are sufficient to assure the safety of study participants. Serum samples may be drawn using local phlebotomy services, home health, or other modalities if site visits cannot occur. Study vaccination visits must have adequate oversight for issues associated with immediate severe reactions but may need to occur outside of the clinical site depending on the pandemic situation (e.g., home study vaccinations).

#### 6.1 Study Visit Procedures

##### 6.1.1 Days -30 to 0 – Screening

The following procedures will be performed within 30 days of first study vaccination. The Screening visit and Day 0 visit may be combined, if feasible, at any given study site.

- Written informed consent will be obtained in conformance with Section 9.2.2.3 of this protocol.
- Review of medical history, including prior and concomitant medical conditions, recent vaccinations (≤ 90 days), and significant surgical procedures.
- Inclusion and exclusion criteria review consistent with Section 4. Specific exclusions to study vaccination will be assessed before any vaccination. Waivers to enrolling participants with exclusions will not be given.
- Demographics, including date of birth (day, month, and year), sex, race, ethnicity, weight, and height.
- Prior and concomitant medications, including recent and current medications at the time of screening to be reviewed to ensure eligibility criteria are fulfilled. Concomitant medications include all medications (including vaccines) taken by the participant.
- Vital sign measurements, including respiratory rate, blood pressure, pulse rate, pulse oximetry, and oral temperature (or via forehead/ear reader).

- Urine pregnancy test for women of childbearing potential only. A positive urine pregnancy test at Screening will result in screen failure.
- Physical examination to include height and weight; head, eyes, ears, nose, and throat (HEENT), neck, thyroid, lungs, heart, cardiovascular, abdomen, lymph nodes of the upper extremities and neck; and musculoskeletal system/extremities to allow for study vaccination.
- Mucosal samples (nasal or throat swabs or saliva based on test availability) will be collected to test for SARS-CoV-2 (PCR) only if the participant has any COVID-19 symptoms or significant exposure history. If a participant has a positive PCR for SARS-CoV-2 prior to enrolment that participant will not be eligible and will be considered a screen failure.
- Assessment of SAEs, starting from the time of informed consent.

##### 6.1.2 Day 0 – First Study Vaccination

The Screening and Day 0 Visits may be combined whenever feasible.

All participants with confirmed eligibility will have the following procedures performed:

- Inclusion and exclusion criteria review consistent with Section 4. Specific exclusions to study vaccination will be assessed before any study vaccination.
- Prior and concomitant medications, including recent and current medications to be reviewed to ensure eligibility criteria are fulfilled. Concomitant medications include all medications (including vaccines) taken by the participant.
- Urine pregnancy test for women of childbearing potential only. A urine pregnancy test will be performed prior to each study vaccination. A positive urine pregnancy test at any time will result in the participant not receiving any further study vaccination.
- Symptom-directed (targeted) physical examination, including lymphatic assessment of injected upper extremity. Physical examination must be done prior to study vaccination.
- Blood sampling for SARS-CoV-2 prior to study vaccination (ELISA for anti-N-protein serology) in all participants.
- Blood sampling for SARS-CoV-2 prior to study vaccination (ELISA for anti-S-protein serology) in approximately 900 participants in the Anti-S Protein Serology Subset.
- Blood sampling for SARS-CoV-2 neutralisation assay prior to study vaccination in approximately 900 participants in the Neutralisation Assay Subset.
- Blood sampling for HAI prior to study vaccination for participants who received licensed seasonal influenza vaccine on Day 0 in the approximately 400 participants in the licensed seasonal influenza co-administration sub-study.

- Blood sampling for cell-mediated assessments prior to study vaccination, as measured by ELISpot ± intracellular cytokine staining, in approximately 450 participants in the Cell-mediated Assay Subset.
- Mucosal samples (nasal or throat swabs or saliva based on test availability) will be collected to test for SARS-CoV-2 (PCR) to determine current infection with SARS-CoV-2 and to demonstrate the methods required for mucosal sample collection; participants will be taught how to self-sample at this time. Participants with possible COVID-19 symptoms will not be randomised but may return when symptoms have resolved per protocol. Participants found to be SARS-CoV-2 positive on Day 0 will be excluded from the immunogenicity analyses of the study as per the SAP.
- Randomisation.
- Pre-injection vital sign collection (respiratory rate, blood pressure, pulse rate, pulse oximetry, and oral temperature [or via forehead/ear reader]) before study vaccination to ensure participant has no evidence of fever prior to study vaccination.
- Alcohol swab cleansing of the injection sites for both study vaccine (SARS-CoV-2 rS with Matrix-M1 adjuvant or placebo) and licensed seasonal influenza vaccination (first approximately 400 eligible participants).
- Vaccination of study vaccine as an IM injection into the deltoid muscle. The first approximately 400 eligible participants will also receive an IM injection of a licensed seasonal influenza vaccine in the opposite deltoid following study vaccination.
- Monitoring for reactogenicity, participants will remain in clinic for at least 30 minutes to be monitored for any severe reactogenicity. Severe reactions will be noted as AEs on day of study vaccination.
- Post-injection vital sign collection (respiratory rate, blood pressure, pulse rate, pulse oximetry, and oral temperature [or via forehead/ear reader]) at approximately 15 to 30 minutes after study vaccination to check for any reactions to the study vaccine.
- Monitoring for COVID-19. Participants will monitor for COVID-19 symptoms. This will include provision of a Study Identification (ID) Card that provides details on study participation, study site contact information, and assessment of symptoms of suspected COVID-19 (see Table 2-2). Instructions to participants to contact the study site within 24 hours for symptoms of suspected COVID-19 will be given. A participant with suspected or confirmed COVID-19 will be asked to complete a COVID-19 symptom diary for 10 days. The Study ID Card should be presented to healthcare providers not affiliated with the study who encounter participants with symptoms of suspected COVID-19. Symptoms of reactogenicity within 7 days post-vaccination may not trigger the actions associated with COVID-19 monitoring.
- Assessment of unsolicited AEs, MAAEs (all and related to study vaccination), SAEs, and AESIs.

##### 6.1.3 Day 21 – Second Study Vaccination (+ 7 days)

All participants will have the following procedures performed:

- Prior and concomitant medications. Concomitant medications include all medications (including vaccines) taken by the participant.
- Urine pregnancy test for women of childbearing potential only. A urine pregnancy test will be performed prior to each study vaccination. A positive urine pregnancy test at any time will result in the participant not receiving any further study vaccination.
- Symptom-directed (targeted) physical examination, including lymphatic assessment of injected upper extremity. Physical examination must be done prior to study vaccination.
- Blood sampling for HAI for participants who received licensed seasonal influenza vaccine on Day 0 in the approximately 400 participants in the licensed seasonal influenza co-administration sub-study.
- Mucosal samples (nasal or throat swabs or saliva based on test availability) will be collected to test for SARS-CoV-2 (PCR) only if the participant has any COVID-19 symptoms or significant exposure history between Days 0 and 21. Results of that test are not required for study vaccination, but participants who are symptomatic may be have second study vaccination delayed per protocol. Participants found to be SARS-CoV-2 positive on Day 21 will be excluded from the immunogenicity analyses of the study as per the SAP.
- Pre-injection vital sign collection (respiratory rate, blood pressure, pulse rate, pulse oximetry, and oral temperature [or via forehead/ear reader]) before study vaccination to ensure participant has no evidence of fever prior to study vaccination.
- Alcohol swab cleansing of the injection site for study vaccine (SARS-CoV-2 rS with Matrix-M1 adjuvant or placebo)
- Vaccination of study vaccine as an IM injection into the opposite deltoid muscle of the Day 0 study vaccine.
- Monitoring for reactogenicity, participants will remain in clinic for at least 30 minutes to be monitored for any severe reactogenicity. Severe reactions will be noted as AEs on day of study vaccination.
- Post-injection vital sign collection (respiratory rate, blood pressure, pulse rate, pulse oximetry, and oral temperature [or via forehead/ear reader]) at approximately 15 to 30 minutes after study vaccination to check for any reactions to the study vaccine.
- Monitoring for COVID-19. Participants will monitor for COVID-19 symptoms. This will include provision of a Study ID Card that provides details on study participation, study site contact information, and assessment of symptoms of suspected COVID-19 (see Table 2-2). Instructions to participants to contact the study site within 24 hours for symptoms of suspected COVID-19 will be given. A participant with suspected or confirmed COVID-19 will be asked to complete a COVID-19 symptom diary for 10 days. The Study ID Card should be presented to healthcare providers not affiliated

with the study who encounter participants with symptoms of suspected COVID-19. Symptoms of reactogenicity within 7 days post-vaccination may not trigger the actions associated with COVID-19 monitoring.

- Assessment of unsolicited AEs, MAAEs (all and related to study vaccination), SAEs, and AESIs.

##### 6.1.4 Day 35 – Follow-up Visit (+ 7 days)

All participants will have the following procedures performed:

- Prior and concomitant medications. Concomitant medications include all medications (including vaccines) taken by the participant.
- Symptom-directed (targeted) physical examination, including lymphatic assessment of injected upper extremity.
- Blood sampling for SARS-CoV-2 prior to study vaccination (ELISA for anti-N-protein serology) in all participants.
- Blood sampling for SARS-CoV-2 prior to study vaccination (ELISA for anti-S-protein serology) in approximately 900 participants in the Anti-S Protein Serology Subset.
- Blood sampling for SARS-CoV-2 neutralisation assay prior to study vaccination in approximately 900 participants in the Neutralisation Assay Subset.
- Blood sampling for cell-mediated assessments prior to study vaccination, as measured by ELISpot ± intracellular cytokine staining, in approximately 450 participants in the Cell-mediated Assay Subset.
- Monitoring for COVID-19. Participants will monitor for COVID-19 symptoms. This will include provision of a Study ID Card that provides details on study participation, study site contact information, and assessment of symptoms of suspected COVID-19 (see Table 2-2). Instructions to participants to contact the study site within 24 hours for symptoms of suspected COVID-19 will be given. A participant with suspected or confirmed COVID-19 will be asked to complete a COVID-19 symptom diary for 10 days. The Study ID Card should be presented to healthcare providers not affiliated with the study who encounter participants with symptoms of suspected COVID-19.
- Assessment of unsolicited AEs, MAAEs (all and related to study vaccination), SAEs, and AESIs.

##### 6.1.5 COVID-19 Surveillance Visits (Unscheduled)

###### 6.1.5.1 Initial COVID-19 Surveillance Visit

All participants will have the following procedures performed:

- Review of medical history, including prior and concomitant medical conditions, recent vaccinations (≤ 90 days), and significant surgical procedures.
- Prior and concomitant medications. Concomitant medications include all medications (including vaccines) taken by the participant.
- Vital sign measurements, including respiratory rate, blood pressure, pulse rate, pulse oximetry, and oral temperature (or via forehead/ear reader).
- Symptom-directed (targeted) physical examination, including lymphatic assessment of injected upper extremity.
- Mucosal samples (nasal or throat swabs or saliva based on test availability) will be collected to test for SARS-CoV-2 (PCR) in an effort to determine if the current symptoms are due to SARS-CoV-2 infection. Testing may occur at home or in the clinic as part of a COVID-19 Surveillance Visit. If the participant self-tested at home and has a clinic visit that same day they should be sampled again by a member of the study staff.
- COVID-19 Symptom Diary. A participant with suspected or confirmed COVID-19 will be asked to complete the COVID-19 symptom diary. Participants should start the diary on their first day of symptoms and complete this diary daily for a minimum of 10 days even if their symptoms have resolved and regardless of any SARS-CoV-2 PCR results. They should continue the diary beyond 10 days if they have any ongoing symptoms related to the episode and should continue to do so until all symptoms have resolved.
- Assessment of MAAEs (related to study vaccination), SAEs, and AESIs.

###### 6.1.5.2 Follow-up COVID-19 Surveillance Visit

A Follow-Up COVID-19 Surveillance Visit will occur approximately 7 (+ 3) days after the Initial COVID-19 Surveillance Visit to ascertain worsening/progression of COVID-19 symptoms.

All participants will have the following procedures performed:

- Review of medical history, including prior and concomitant medical conditions, recent vaccinations (≤ 90 days), and significant surgical procedures.
- Prior and concomitant medications. Concomitant medications include all medications (including vaccines) taken by the participant.
- Vital sign measurements, including respiratory rate, blood pressure, pulse rate, pulse oximetry, and oral temperature (or via forehead/ear reader).

- Symptom-directed (targeted) physical examination, including lymphatic assessment of injected upper extremity.
- Mucosal samples (nasal or throat swabs or saliva based on test availability) will be collected to test for SARS-CoV-2 (PCR), **only in participants testing negative in all self-testing and Initial COVID-19 Surveillance Visit tests**.
- COVID-19 Symptom Diary. A participant with suspected or confirmed COVID-19 will be asked to complete the COVID-19 symptom diary. Participants should start the diary on their first day of symptoms and complete this diary daily for a minimum of 10 days even if their symptoms have resolved and regardless of any SARS-CoV-2 PCR results. They should continue the diary beyond 10 days if they have any ongoing symptoms related to the episode and should continue to do so until all symptoms have resolved.
- Assessment of MAAEs (related to study vaccination), SAEs, and AESIs.

##### 6.1.6 3 Months (± 15 days) After Second Study Vaccination

All participants will have the following procedures performed:

- Prior and concomitant medications. Concomitant medications include all medications (including vaccines) taken by the participant.
- Blood sampling for SARS-CoV-2 prior to study vaccination (ELISA for anti-N-protein serology) in all participants.
- Monitoring for COVID-19. Participants will monitor for COVID-19 symptoms. This will include provision of a Study ID Card that provides details on study participation, study site contact information, and assessment of symptoms of suspected COVID-19 (see Table 2-2). Instructions to participants to contact the study site within 24 hours for symptoms of suspected COVID-19 will be given. A participant with suspected or confirmed COVID-19 will be asked to complete a COVID-19 symptom diary for 10 days. The Study ID Card should be presented to healthcare providers not affiliated with the study who encounter participants with symptoms of suspected COVID-19.
- Assessment of MAAEs (related to study vaccination), SAEs, and AESIs.

##### 6.1.7 6 Months (± 15 days) After Second Study Vaccination

All participants will have the following procedures performed:

- Prior and concomitant medications. Concomitant medications include all medications (including vaccines) taken by the participant.
- Blood sampling for SARS-CoV-2 prior to study vaccination (ELISA for anti-N-protein serology) in all participants.
- Monitoring for COVID-19. Participants will monitor for COVID-19 symptoms. This will include provision of a Study ID Card that provides details on study participation, study site contact information, and assessment of symptoms of suspected COVID-19

(see Table 2-2). Instructions to participants to contact the study site within 24 hours for symptoms of suspected COVID-19 will be given. A participant with suspected or confirmed COVID-19 will be asked to complete a COVID-19 symptom diary for 10 days. The Study ID Card should be presented to healthcare providers not affiliated with the study who encounter participants with symptoms of suspected COVID-19.

- Assessment of MAAEs (related to study vaccination), SAEs, and AESIs.

##### 6.1.8 12 Months (± 15 days) After Second Study Vaccination

All participants will have the following procedures performed:

- Prior and concomitant medications. Concomitant medications include all medications (including vaccines) taken by the participant.
- Blood sampling for SARS-CoV-2 prior to study vaccination (ELISA for anti-N-protein serology) in all participants.
- Monitoring for COVID-19. Participants will monitor for COVID-19 symptoms. This will include provision of a Study ID Card that provides details on study participation, study site contact information, and assessment of symptoms of suspected COVID-19 (see Table 2-2). Instructions to participants to contact the study site within 24 hours for symptoms of suspected COVID-19 will be given. A participant with suspected or confirmed COVID-19 will be asked to complete a COVID-19 symptom diary for 10 days. The Study ID Card should be presented to healthcare providers not affiliated with the study who encounter participants with symptoms of suspected COVID-19.
- Assessment of MAAEs (related to study vaccination), SAEs, and AESIs.

#### 6.2 Efficacy assessments

##### 6.2.1 Mucosal Samples for Virus Detection

Mucosal samples (nasal/throat swabs or saliva) for virus detection will be taken at the study visits described in the SOE (Table 3-1).

- Mucosal samples will be not be taken at Screening unless participants have symptoms or significant exposure to SARS-CoV-2. If a participant has a positive PCR for SARS-CoV-2 prior to enrolment that participant will not be eligible and will be considered a screen failure.
- Mucosal samples will be taken on Day 0 to determine current infection with SARS-CoV-2 and to demonstrate the methods required for mucosal sample collection. Participants will be taught how to self-sample at this time. Participants with possible COVID-19 symptoms will not be randomised but may return when symptoms have resolved per protocol. Participants found to be SARS-CoV-2 positive on Day 0 will be excluded from the immunogenicity analyses of the study as per the SAP.
- Mucosal samples will be not be routinely taken on Day 21. Participants with possible COVID-19 symptoms that develop between Day 0 and Day 21 may have a SARS-

CoV-2 PCR test performed prior to second study vaccination on Day 21. Results of that test are not required for vaccination, but participants who are symptomatic may be have the second study vaccination delayed per protocol. Participants found to be SARS-CoV-2 positive on Day 21 will be excluded from the immunogenicity analyses of the study as per the SAP.

##### 6.2.2 Virologic Confirmation of SARS-CoV-2

Mucosal sampling (nasal/throat swabs or saliva) will be performed to virologically confirm (by PCR to SARS-CoV-2) the presence of SARS-CoV-2 beginning on Day 0 until the EOS, yet only those SARS-CoV-2 cases detected 7 days after second study vaccination will be utilised in study efficacy endpoints.

If a participant experiences any symptom in Table 2-2, this will trigger:

- Mucosal self-sampling once daily for 3 consecutive days (self-sampling on any single day may be replaced by sampling by a healthcare provider [HCP]), but if self-sampling has occurred in the same day before the Initial COVID-19 Surveillance Visit the HCP sampling should still be performed. If any test is found positive for before 3 consecutive days of testing is performed, the full 3 consecutive tests may not be required. Participants will self-sample based on the training given on Day 0.
- Mucosal sampling will be started approximately 24 hours after the first symptom(s) from Table 2-2 are reported.

The logistics and processing of these samples is being coordinated by the Department of Health and Social Care (DHSC) (UK government) as part of the national community testing programme.

##### 6.2.3 Monitoring for Suspected COVID-19

Identification and laboratory confirmation of SARS-CoV-2 infection and symptomatic COVID-19 will be performed throughout the study.

On study enrolment, participants will be given details of a 24/7 on-call number to contact the study team. Participants will be instructed to contact the study team within 24 hours via this number if they self-assess COVID-19 symptoms in Table 2-2 or are clinically concerned. Newly discovered symptoms of suspected COVID-19 will trigger a COVID-19 Surveillance Visit.

Please refer to Table 2-2 for the qualifying symptoms of suspected COVID-19 disease.

Participants will receive weekly reminders (email or text messages) to immediately contact the study team if they have experienced any new symptoms or health concerns that could be related to infection with SARS-CoV-2. Newly discovered symptoms of suspected COVID-19 will trigger a COVID-19 Surveillance Visit.

A participant with suspected or confirmed COVID-19 will be asked to complete the COVID-19 symptom diary. Participants should start the diary on their first day of symptoms and complete this diary daily for a minimum of 10 days even if their symptoms have resolved and regardless of any SARS-CoV-2 PCR results. They should continue the diary beyond 10 days if they have any ongoing symptoms related to the episode and should continue to do so until all symptoms have resolved.

The occurrence of COVID-19-related hospitalisation and COVID-19-related complications (such as, but not limited to, pneumonia, neurological or vascular complications, severe pneumonia, severe neurological or vascular events, acute respiratory distress syndrome, renal complications, sepsis, septic shock, death) will be monitored throughout the study.

Every episode of a new onset of symptoms of suspected COVID-19 will trigger a COVID-19 Surveillance Visit (Initial and Follow-Up). A “new onset” episode will require at least a 7-day period of being symptom-free prior to the event to differentiate a specific episode from any prior illness.

###### 6.2.3.1 Severity of COVID-19 Symptoms

COVID-19 symptoms will be categorised as virologically confirmed, mild, moderate, or severe as described in Table 2-1.

###### 6.2.3.2 COVID-19 Surveillance Visit (Initial and Follow-up)

A COVID-19 Surveillance Visit (Initial and Follow-Up) will be triggered by symptoms of suspected COVID-19 captured by surveillance.

When a participant is determined to have a new onset of symptoms, the participant will contact the study team immediately, begin their COVID-19 symptom diary and begin the 3 consecutive days of PCR self-testing (beginning approximately 24 hours after the start of symptoms) as above. Participants will be asked to attend an Initial COVID-19 Surveillance Visit at the study clinic or will be seen at an in-home visit by study staff depending on local conditions.

###### 6.2.3.2.1 Initial COVID-19 Surveillance Visit

An Initial COVID-19 Surveillance Visit will be performed at the study site (or home) and will occur as soon as possible within approximately **1-3 days** of new symptom onset (however, data from specimens obtained up to 14 days will be accepted). The visit will consist of the following:

- Review and confirmation of the history of COVID-19 symptoms, including approximate date of onset of illness and solicitation of each symptom (see Table 2-2).

- Vital signs, including resting respiratory rate (on room air or the participant's basal level of chronic supplemental oxygen use) and pulse oximetry, will be captured as numerical values. Lung auscultation (exam) will be performed.
- Ascertainment of any unscheduled healthcare visit by the participant (or home visit by a healthcare provider) in response to symptoms of suspected COVID-19.
- Ascertainment of new concomitant medications (especially antibiotics) or altered doses/frequencies of existing concomitant medications resulting from symptoms of suspected COVID-19.
- Collection of a specimen of upper respiratory secretions via nasal mid-turbinate swab for qualitative PCR detection of SARS-CoV-2.

###### 6.2.3.2.2 Follow-Up COVID-19 Surveillance Visit

A Follow-Up COVID-19 Surveillance Visit will occur approximately 7 (+ 3) days after the Initial COVID-19 Surveillance Visit. This visit will consist of the following:

- If any prior mucosal sample obtained after the onset of symptoms tests **positive** for SARS-CoV-2 virus by PCR, the participant may have a clinic or home visit. Study staff will conduct the Follow-Up COVID-19 Surveillance Visit approximately 7 (+ 3) days after the Initial COVID-19 Surveillance Visit to ascertain worsening/ progression of COVID-19 symptoms. This follow-up visit by study staff will include all of the same procedures outlined in this section, including review of symptom history, measurement of vital signs (including pulse oximetry), and lung auscultation; however, a repeat mucosal sampling will NOT be obtained since the participant has already tested positive.
- If all prior mucosal samples obtained after the onset of symptoms test **negative** for SARS-CoV-2 by qualitative PCR or the mucosal sample is inadequate for analysis, the participant will be re-evaluated in the clinic (or home) approximately 7 (+ 3) days after the Initial COVID-19 Surveillance Visit to ascertain worsening/progression of suspected COVID-19 symptoms AND to obtain an additional sample for PCR detection of SARS-CoV-2. This re-evaluation will include all of the same procedures outlined in this section, including review of symptom history, measurement of vital signs (including pulse oximetry), and lung auscultation.

After the Follow-Up COVID-19 Surveillance Visit, participants will continue to receive telephone contacts approximately every week for ascertainment of COVID-19 symptom status until resolution of symptoms. Subsequent calls will document resolution or return to baseline of COVID-19 symptoms in order to calculate illness duration (date of symptom onset to day of symptom resolution) and will collect any additional healthcare visits, hospitalisations, and/or concomitant medications due to the suspected COVID-19.

Should a participant visit an emergency room, be admitted to the hospital or a COVID-19 ward, and PCR sampling is missed, then the local public health COVID-19 diagnostic test (or COVID-19 diagnostic test performed in the healthcare setting) will be taken as a valid result.

Importantly, clinical data on symptoms, vital signs, exam findings, COVID-19 and other diagnostic testing, diagnoses, and complications occurring during the suspected COVID-19 emergency room and/or hospitalisation episode will be collected from available medical records on a study specific hospitalisation/emergency room data collection form in order to assess severity.

Participants will be notified of positive SARS-CoV-2 results as soon as locally practicable due to requirements of self-isolation and potential transmission.

Note that PCR-positive COVID-19 symptoms and events captured as efficacy endpoints will NOT be doubly recorded in the AE eCRF, unless a particular illness fulfils the definition of an SAE. Immunogenicity assessments

#### 6.3 Immunogenicity Assessments

Blood will be collected from all participants for humoral immunogenicity according to the time points specified in the SOE (Table 3-1).

Immune measurements (ELISA) will be conducted on serum (IgG) for SARS-CoV-2 anti-S protein serology in approximately 900 participants in the Anti-S Protein Serology Subset and on anti-N protein serology in all participants. The immunologic test for SARS-CoV-2 serology based on a non-S protein (e.g., SARS-CoV-2 N protein) will be performed to identify cases of asymptomatic infection. Additional immunogenicity assessments specific to SARS-CoV-2 (or related variants) will include a neutralising antibody assay, which will be performed in approximately 900 participants in a Neutralisation Assay Subset. Cell-mediated immune responses, as assessed by ELISpot ± intracellular cytokine staining, will be performed in approximately 450 participants in the Cell-mediated Assay Subset. In addition, an HAI will be performed in the approximately 400 participants in the seasonal influenza vaccine co-administration sub-study.

Details on the handling, processing, and shipping of immunogenicity samples will be provided separately in a laboratory manual.

Participants will be asked to provide consent for the use of samples for future testing or assay development specific to SARS-CoV-2 (or related variants). Aliquots of all collected samples from this study may be retained for additional testing with antigens specific to SARS-CoV-2 (or related variants) for a maximum of 25 years (starting from the date at which the last participant had the last study visit), unless local rules, regulations, or guidelines require different timeframes or different procedures, in accord with participant consent.

#### 6.4 Safety Assessments

The timing and frequency of all safety assessments are listed in the SOE (Table 3-1).

After each study vaccination, including the licensed seasonal influenza vaccine for participants in the seasonal influenza vaccine co-administration sub-study, participants will remain under observation at the study site for at least 30 minutes for the presence of any acute reactions and solicited AEs. Solicited AEs, collected through an electronic diary (e-Diary), will be recorded at each study vaccination visit from the time of study vaccination until 7 days after study vaccination in all participants in the seasonal influenza vaccine co-administration sub-study and a subgroup of all other participants.

All participants will be assessed for unsolicited AEs from the time of first study vacation until Day 49; SAEs will be assessed from the time the informed consent is signed until completion of the participant's last study-related procedure; all MAAEs will be reported from the time of first study vaccination until Day 35; MAAEs related to study vaccination and AESIs will be reported from the time of first study vaccination until completion of the participant's last study-related procedure. All AEs will be followed until resolution or until clinically stable.

Safety assessments will include monitoring and recording of solicited (local and systemic reactogenicity events) and unsolicited AEs; MAAEs; AESI; SAEs; vital sign measurements on day of study vaccinations; and physical examination findings. COVID-19 severity will be categorised as virologically confirmed, mild, moderate, or severe according to protocol-specified criteria (Table 2-1). Recording of solicited and unsolicited AEs may be conducted by electronic data capture (EDC)/reporting. Potential immune-mediated medical conditions (PIMMC) and AESIs specific to potential disease enhancement for COVID-19 will also be monitored (see Section 9.4 [Appendix 4] for details).

##### 6.4.1 Adverse Events

AEs will be assessed during the study as described in the SOE (Table 3-1) and should be followed until they are resolved, stable, or judged by the investigator to be not clinically significant. AEs will be captured after the first dose of study vaccine administered with the exception of an AE related to study procedure or one that causes a delay in study vaccine administration (e.g., acute illness).

The investigator is responsible for ensuring that all AEs and SAEs are recorded in the eCRF and reported to the sponsor, regardless of their relationship to study vaccine or clinical significance. If there is any doubt as to whether a clinical observation is an AE, the event should be reported.

###### 6.4.1.1 Adverse Event Definitions

The investigator is responsible for reporting all AEs that are observed or reported during the study, regardless of their relationship to study vaccination or their clinical significance.

An AE is defined as any untoward medical occurrence in a participant enrolled into this study regardless of its causal relationship to study vaccination. Participants will be instructed to contact the investigator at any time after randomisation if any symptoms develop.

###### 6.4.1.1.1 Serious Adverse Events

An SAE is defined as any event that

- results in death
- is immediately life threatening
- requires inpatient hospitalisation or prolongation of existing hospitalisation
- results in persistent or significant disability/incapacity
- is a congenital anomaly/birth defect

Important medical events that may not result in death, be life threatening, or require hospitalisation may be considered SAEs when, based upon appropriate medical judgment, they may jeopardise the participant or may require medical or surgical intervention to prevent one of the outcomes listed in this definition. Examples of such medical events include allergic bronchospasm requiring intensive treatment in an emergency room or at home, blood dyscrasias or convulsions that do not result in inpatient hospitalisation, or the development of drug dependency or drug abuse.

###### 6.4.1.1.2 Local and General Systemic Reactogenicity Symptoms

Solicited AEs (e.g., local and general systemic reactogenicity symptoms) will be captured in the Safety Subset of approximately 2,000 participants and in the seasonal influenza vaccine co-administration sub-study of approximately 400 participants. Participants will record all local reactogenicity symptoms for each injection at each location (ideally in opposite deltoids) while recording of general systemic reactogenicity symptoms may not be assigned to either injection. Solicited AEs, collected through an electronic diary (e-Diary), will be recorded from the time of study vaccination until 7 days after study vaccination.

Site-specific local (arm) and general systemic reactogenicity reactions including start and stop dates will be recorded and the investigator will apply a standard toxicology grading at the subsequent study visit (Section 9.3, Appendix 3). Should any reactogenicity event extend beyond 7 days after study vaccination and be clinically significant by toxicity grade 1 or greater, then it will be recorded as an unsolicited AE with a start date on the 8$^{th}$ day following study vaccination and followed to resolution.

###### 6.4.1.1.3 Adverse Events of Special Interest

Participants will be assessed for diagnosis of an AESI at all study visits. AESIs include PIMMCs, AEs specific to COVID-19, or other potential AEs that may be determined at any time by regulatory authorities as additional information concerning COVID-19 is obtained. Given the concern for cytokine storm, an AESI of cytokine release syndrome will be included as an AE specific to COVID-19. Listings of AESI are presented in Section 9.4, Appendix 4.

###### 6.4.1.1.4 Medically Attended Adverse Events

MAAEs are defined as AEs with medically-attended visits including hospital, emergency room, urgent care clinic, or other visits to or from medical personnel for any reason. Routine study visits (including COVID-19 Surveillance Visits) will not be considered medically attended visits. MAAEs are to be reported from the time of first study vaccination until Day 35. MAAEs related to study vaccination are to be reported from the time of first study vaccination until completion of the participant's last study-related procedure, which may include contact for safety follow-up.

###### 6.4.1.1.5 Pregnancy

Pregnancy is not considered an AE unless there is a suspicion that an investigational vaccine may have interfered with the effectiveness of a contraceptive medication. Any pregnancy that occurs during study participation must be reported using a clinical study pregnancy form. To ensure participant safety, each pregnancy must be reported to Novavax, Inc. within 2 weeks of learning of its occurrence. If pregnancy occurs, further study vaccination will be discontinued. The pregnancy must be followed up to determine outcome (including spontaneous miscarriage, elective termination, normal birth, or congenital abnormality) and the status of both mother and child, even if the participant was discontinued from the study. Pregnancy complications and elective terminations for medical reasons must be reported as an AE or SAE. Spontaneous miscarriages must be reported as an SAE.

Any pregnancy brought to the investigator's attention after the participant has completed the study but occurring while the participant was in the study must be promptly reported to:

Sponsor Safety Monitor: [redacted]

###### 6.4.1.2 Eliciting and Documenting Adverse Events

At every study visit, participants will be asked a standard question to elicit any medically-related changes in their well-being. They will also be asked if they have been hospitalised, had any accidents, used any new medications, or changed concomitant medication regimens (both prescription and OTC medications).

In addition to participant observations, AEs will be documented from any data collected on the AE page of the eCRF or other documents that are relevant to participant safety.

###### 6.4.1.3 Reporting Adverse Events

All AEs reported or observed during the study will be recorded on the AE page of the eCRF. Information to be collected includes study vaccine, type of event, time of onset, dosage, investigator-specified assessment of severity and relationship to study vaccine and/or study procedure, time of resolution of the event, seriousness, any required treatment or evaluations, and outcome. Any AEs resulting from concurrent illnesses, reactions to concurrent illnesses, reactions to concurrent medications, or progression of disease must also be reported. All AEs will be followed until they are resolved, stable, or judged by the investigator to be not clinically significant. MedDRA will be used to code all AEs.

Any medical condition that is present at the time that the participant is screened but does not deteriorate should not be reported as an AE. However, if it deteriorates at any time during the study, it should be recorded as an AE.

Any AE that is considered serious by the investigator or that meets SAE criteria (Section 6.4.1.1.1) must be reported to the sponsor within 24 hours after the investigator has confirmed the occurrence of the SAE. The investigator will provide a causality assessment (whether there is a reasonable possibility that the study vaccine caused the event) to the study vaccine. The sponsor will be responsible for notifying the relevant regulatory authorities of any SAE, in compliance with health authority requirements, as outlined in the relevant clinical study guidelines.

For this study, the following contact information will be used for SAE reporting:

Phone: [redacted]
Fax: [redacted]
Email: [redacted]

###### 6.4.1.4 Assessment of Severity

The severity (or intensity) of an AE refers to the extent to which it affects the participant's daily activities and will be classified as mild, moderate, or severe using the following criteria:

- Mild (grade 1): These events require minimal or no treatment and do not interfere with the participant's daily activities.
- Moderate (grade 2): These events result in a low level of inconvenience or require minor therapeutic measures. Moderate events may cause some interference with normal functioning.
- Severe (grade 3): These events interrupt a participant's usual daily activity and may require systemic drug therapy or other treatment. Severe events are usually incapacitating.

If the severity of an AE changes, the most intense severity should be reported. An AE characterised as intermittent does not require documentation of the onset and duration of each episode.

###### 6.4.1.5 Assessment of Causality

The investigator's assessment of an AE's relationship to study vaccine is part of the documentation process, but it is not a factor in determining what is or is not reported in the study. If there is any doubt as to whether a clinical observation is an AE, the event should be reported.

The investigator will assess causality (i.e., whether there is a reasonable possibility that the study vaccine caused the event) for all AEs and SAEs (solicited reactions are to be considered as being related to study vaccination). The relationship will be classified as follows:

- Not related: There is not a reasonable possibility of relationship to study vaccine. The AE does not follow a reasonable temporal sequence from administration of study vaccine or can be reasonably explained by the participant's clinical state or other factors (e.g., disease under study, concurrent diseases, and concomitant medications).
- Related: There is a reasonable possibility of relationship to study vaccine. The AE follows a reasonable temporal sequence from administration of study vaccine and cannot be reasonably explained by the participant's clinical state or other factors (e.g., disease under study, concurrent diseases, or concomitant medications), represents a known reaction to study vaccine or other vaccines in its class, is consistent with the known pharmacological properties of the study vaccine, and/or resolves with discontinuation of the study vaccine (and/or recurs with re-challenge, if applicable).

###### 6.4.1.6 Follow-up of Adverse Events

All AEs must be reported in detail on the appropriate page of the eCRF and followed until they are resolved, stable, or judged by the investigator to be not clinically significant.

##### 6.4.2 Vital Sign Measurements

Vital sign measurements will include oral temperature (or via forehead/ear reader), pulse rate and diastolic and systolic blood pressure (after participant is seated for at least 5 minutes), and pulse oximetry. Temperature will be recorded and graded during general systemic reactogenicity evaluation (Section 6.4.1.1.2). The other vital sign measurements will be recorded as continuous variables prior to each study vaccination. Pulse oximetry and other vital signs may be taken at a participant's baseline oxygen utilisation level which should be noted in the eCRF.

On study vaccination days, vital sign measurements will be collected once before study vaccination to ensure participant has controlled blood pressure and heart rate and no evidence of fever prior to study vaccination and once more, at approximately 15 to 30 minutes after study vaccination, to check for any reactions to the study vaccine. The investigator will only apply standard toxicology grading on the day of study vaccination, both before and after study vaccination (Section 9.3, Appendix 3). If individual vital sign measurements are considered clinically significant by the investigator, study vaccination may be withheld that day, and participants may return on a subsequent day for re-evaluation and study vaccination, ideally, within the time window specified in the SOE (Table 3-1).

##### 6.4.3 Physical Examinations

A physical examination will be performed at screening (at minimum, assessment of skin, head, ears, eyes, nose, throat, neck, thyroid, lungs, heart, cardiovascular, abdomen, lymph nodes of the upper extremities and neck, and musculoskeletal system/extremities). Height and weight will be measured at screening only.

A targeted or symptom-directed physical examination will be performed at the time points specified in the SOE (Table 3-1).

##### 6.4.4 Safety Monitoring

Safety oversight will be conducted by an SMC during the course of the study. The SMC is an independent group of experts that monitors participant safety and advises the sponsor. The SMC members will be separate and independent of site personnel participating in this study and should not have a scientific, financial, or other conflict of interest related to this study or the sponsor. The SMC will consist of members with appropriate expertise to contribute to the interpretation of the data from this study.

A global SMC was convened to oversee the safety of the ongoing Phase 1/2 study (Protocol 2019nCoV-101) and will oversee 1 or more additional studies across the SARS-CoV-2 rS vaccine program. A separate SMC will be convened for this study. The designated SMC will monitor the safety of participants in the study and will follow an SMC charter. The SMC will review blinded and unblinded safety and reactogenicity data. The SMC will also review sufficient clinical event data to ensure detection of evidence of enhanced disease. The SMC will convene to perform safety reviews on a scheduled basis as per the SMC charter; for immediate concerns regarding safety observations during this study; and as needed.

The SMC will operate under the rules of a sponsor-approved charter that will be approved at the organisational meeting of the SMC. At this time, each data element that the SMC needs to assess will be clearly defined. Procedures for SMC reviews/meetings will be defined in the charter. The SMC will review applicable data for safety assessments (AEs by classifications) and any clinical data that may be of significance to this review (e.g., demographics, study vaccination timing, and medications). Additional data may be requested by the SMC, and interim statistical reports may be generated as deemed necessary and appropriate for this review. The SMC may receive data in aggregate and presented by study vaccine group. The SMC may also be provided with expected and observed rates of the expected AEs in an unblinded fashion and may request the study vaccine assignment be unblinded for an individual participant if required for safety assessment.

### 7. STATISTICAL ANALYSIS PLANS

#### 7.1 Sample Size Calculations

This study is designed to enrol approximately 9,000 participants, randomised 1:1 into the 2 study vaccine groups.

Power calculations were performed for each primary endpoint using the two-sided 97.5% confidence intervals (CIs; i.e., one-side alpha of 0.0125) to be conservative (Table 7-1). This is designed to provide at least 90% power for each of the 2 primary endpoints based on the following assumptions:

1. A symptomatic mild, moderate, or severe COVID-19 incident rate of 3% in the placebo group and a VE of 50%.
2. A symptomatic moderate or severe COVID-19 incident rate of 2% in the placebo group and a VE of 70%.
3. 90% evaluability rate for the per-protocol efficacy (PP-EFF) population (assuming 10% unevaluable due to attrition and/or baseline-seropositive participants).

**Table 7-1: Sample Size Needed for 90% for Each of 2 Primary Endpoints**

| Independent Primary Endpoints [a] | Placebo Attack Rate | Vaccine Efficacy | LBCI Success Criteria | Enrolled Sample Size [b] | | |
|---|---|---|---|---|---|---|
| | | | | Per Group Size | Total Size | Number of Events |
| Mild, Moderate, or Severe COVID-19 Symptoms | 4.0% | 30% | > 0% | 6289 | 12578 | 323 |
| | 4.0% | 40% | | 3336 | 6672 | 161 |
| | 4.0% | 50% | | 2004 | 4008 | 91 |
| | 4.0% | 60% | | 1301 | 2602 | 55 |
| | 4.0% | 70% | | 889 | 1778 | 35 |
| | 4.0% | 80% | | 629 | 1258 | 23 |
| | 3.5% | 30% | | 7219 | 14438 | 324 |
| | 3.5% | 40% | | 3828 | 7656 | 162 |
| | 3.5% | 50% | | 2300 | 4600 | 91 |
| | 3.5% | 60% | | 1492 | 2984 | 55 |
| | 3.5% | 70% | | 1020 | 2040 | 35 |
| | 3.5% | 80% | | 721 | 1442 | 23 |
| | 3.0% | 30% | | 8459 | 16918 | 326 |
| | 3.0% | 40% | | 4484 | 8968 | 163 |
| | **3.0%** | **50%** | | **2694** | **5388** | **92** |
| | 3.0% | 60% | | 1748 | 3496 | 55 |
| | 3.0% | 70% | | 1193 | 2386 | 35 |
| | 3.0% | 80% | | 844 | 1688 | 23 |
| Moderate or Severe COVID-19 Symptoms | 2.0% | 50% | > 0% | 4073 | 8146 | 110 |
| | 2.0% | 60% | | 2642 | 5284 | 67 |
| | 2.0% | 70% | | 1803 | 3606 | 42 |
| | 2.0% | 80% | | 1276 | 2552 | 28 |
| | 1.5% | 50% | | 5452 | 10904 | 110 |
| | 1.5% | 60% | | 3536 | 7072 | 67 |
| | 1.5% | 70% | | 2413 | 4826 | 42 |
| | 1.5% | 80% | | 1707 | 3414 | 28 |
| | 1.0% | 50% | | 8210 | 16420 | 111 |
| | 1.0% | 60% | | 5323 | 10646 | 67 |
| | **1.0%** | **70%** | | **3633** | **7266** | **43** |
| | 1.0% | 80% | | 2568 | 5136 | 28 |

Abbreviation: CI = confidence interval; LBCI = lower bound of the confidence interval; PP-EFF = per-protocol efficacy.

[a] Each endpoint to be tested at one-sided alpha of 0.0125 (i.e., lower bound of two-sided 97.5% CI).

[b] Accounting for 90% evaluability rate for the PP-EFF population.

Based on the above conservative assumptions, the minimum target numbers of endpoints needed are 92 symptomatic mild, moderate, or severe COVID-19 endpoints and 43 symptomatic moderate or severe COVID-19 endpoints. Table 7-2 summarizes the power calculations for the 2 primary endpoints based on the proposed multiplicity adjustment using Hochberg method, and the single primary endpoint without an adjustment based on the target numbers of endpoints determined. The following power calculations were performed by 10,000 simulated trials that were created under various assumptions of VEs.

**Table 7-2: Power For 92 Mild, Moderate, or Severe COVID-19 Endpoints or 43 Moderate or Severe COVID-19 Endpoints**

| Assume Vaccine Efficacy | | Power For 2 Primary Endpoints using Hochberg Method | | | Power for Single Primary Endpoint Without Multiplicity Adjustment | |
|---|---|---|---|---|---|---|
| **Mild, Moderate, or Severe COVID-19 Symptoms** | **Moderate or Severe COVID-19 Symptoms** | **Mild, Moderate, or Severe COVID-19 Symptoms** | **Moderate or Severe COVID-19 Symptoms** | **At least 1 Endpoint** | **Mild, Moderate, or Severe COVID-19 Symptoms** | **Moderate or Severe COVID-19 Symptoms** |
| 50% | 70% | 89.20% | 94.52% | 97.93% | 89.66% | 94.52% |
| 60% | 70% | 98.70% | 94.69% | 99.49% | 98.91% | 94.49% |
| 50% | 60% | 87.56% | 75.53% | 92.39% | 89.35% | 75.53% |
| 60% | 80% | 98.77% | 99.68% | 99.90% | 98.80% | 99.68% |

Abbreviations: COVID-19 = coronavirus disease 2019.

In order to increase the precision around VE (i.e., narrow the CIs), the target numbers for the final analysis may be increased up to 114 symptomatic mild, moderate, or severe COVID-19 endpoints or 55 symptomatic moderate or severe COVID-19 endpoints. These increased target numbers of events will provide 80% power at one-sided Type I error rate of 0.025 to demonstrate a lower bound CI > 30% if the VE is 60% for symptomatic mild, moderate, or severe COVID-19 and 70% for symptomatic moderate or severe COVID-19. The minimum sample size equals 3,633/group (7,266 in total). In order to expedite the time to target number of endpoints needed or lower the expected endpoint accrual rate the study will enrol approximately 9,000 participants. The actual sample size for the study will be selected at the operational cut-off date before initiation of the study, based on estimated incidences for the targeted study regions and population at that time.

#### 7.2 Analysis Sets

The intent-to-treat efficacy (ITT-EFF) and immunogenicity (ITT-IMM) analysis sets will include all participants who are randomised and receive at least 1 dose study vaccine (SARS-CoV-2 rS with Matrix-M1 adjuvant or placebo), regardless of protocol violations or missing data. The intent-to-treat (ITT) analysis set will be used as a supportive analysis population for the immunogenicity and efficacy analyses and will be analysed according to the study vaccine group as randomised.

The safety analysis set will include all participants who receive at least 1 dose of study vaccine (SARS-CoV-2 rS with Matrix-M1 adjuvant or placebo). Participants in the safety analysis set will be analysed according to the study vaccine actually received.

The PP-EFF will include seronegative participants who receive both doses of study vaccine (SARS-CoV-2 rS with Matrix-M1 adjuvant or placebo) and have no major protocol deviations affecting the primary efficacy outcome as assessed by the sponsor prior to unblinding. All analyses of the PP-EFF population will exclude any illness episodes with positive SARS-CoV-2 by any validated PCR or antibody test occurring before 14 days after second study vaccination (e.g., Day 35).

The per-protocol immunogenicity (PP-IMM) analysis set for each post-randomisation visit will include participants who receive scheduled dose(s) of study vaccine (SARS-CoV-2 rS with Matrix-M1 adjuvant or placebo), have at least a baseline and the serum sample result for the visit available after study vaccination, and have no major protocol violations that are considered clinically likely to impact the immunogenicity response at the corresponding study visit as assessed by the sponsor prior to unblinding. For each visit, the SARS-CoV-2 unexposed population will also exclude any illness episodes with positive SARS-CoV-2 by any validated PCR or antibody test prior to each visit. Prior exposed participants will be determined using baseline SARS-CoV-2 immunity defined as positive SARS-CoV-2 by PCR or serum IgG antibody at baseline, or positive SARS-CoV-2 by qualitative PCR through Day 21 or Day 35, according to the specified analysis. Analysis will be performed to assess if immune responses differ between exposed and unexposed individuals (i.e., whether prior exposure alters dosing regimen considerations in a pandemic response).

The review and determination for exclusion from the PP populations will be carried out in a blinded fashion by a study clinician prior to unblinding for each interim analysis based on all available information from the locked database.

#### 7.3 Statistical Analysis

Details of all statistical analyses will be described in the SAP.

All data collected will be presented in data listings. Data from participants excluded from an analysis set will be presented in the data listings but not included in the calculation of summary statistics for that analysis set.

For categorical variables, frequencies and percentages will be presented. Continuous variables will be summarised using descriptive statistics (number of participants, mean, median, minimum, and maximum).

Baseline demographic and background variables will be summarised by study vaccine group. The number of participants who enrol in the study and the number and percentage of participants who complete the study will be presented. Frequency and percentage of participants who withdraw or discontinue from the study, and the reason for withdrawal or discontinuation, will also be summarised.

##### 7.3.1 Efficacy Analyses

The study will have 2 key decision time points for efficacy: 1) evaluation of an early efficacy signal to guide a potential request for conditional approval based on accumulating events and 2) the primary analysis to evaluate the primary and secondary objectives of the study.

The primary endpoints will be analysed on the PP-EFF Analysis Set and supported by analysis of the ITT-EFF Set. Conclusions concerning declaration of attainment of the primary endpoint at the completion of the study will only be based on the PP-EFF population. In addition, supportive analyses based on the ITT-EFF population will also be performed.

VE is defined as VE (%) = (1 – RR) × 100, where RR = relative risk of incidence rates between the 2 study vaccine groups (SARS-CoV-2 rS with Matrix-M1 adjuvant or placebo). The final analysis for the primary objective in the PP-EFF population will be carried out at the overall one-sided Type I error rate of 0.025 overall for the 2 primary endpoints. The family-wise type 1 error rate among these 2 analyses will be controlled using the Hochberg approach. Since the demonstration of the primary objective can be achieved if at least 1 of the 2 primary endpoints meets the pre-specified success criteria, the statistical analysis will be performed in the following order:

a. Construct two-sided 95% CIs for the 2 primary endpoints. If both lower bounds are > 0%, declare that both endpoints have met the success criteria. Otherwise, proceed to the next step.
b. Construct two-sided 97.5% CIs for the 2 primary endpoints. If the higher of the lower bounds is > 0%, declare that only the endpoint associated with lower bound > 0% has met the success criterion. Otherwise, both endpoints did not meet the success criteria.

The estimated RR and its CI will be derived using Poisson regression with robust error variance [Zou 2004]. The explanatory variables in the model will include study vaccine group. The dependent variable will be the incidence rate of the endpoint of interest. The robust error variances will be estimated using the repeated statement and the participant identifier. The Poisson distribution will be used with a logarithm link function.

As the primary analysis, hypothesis testing of each primary efficacy endpoint will be carried out against H0: VE ≤ 0%. Rejection of the null hypothesis, H0: VE ≤ 0% demonstrates a statistically significant vaccine effect for either primary endpoint. The study will continue for the intended duration to measure immunogenicity and safety endpoints, regardless of primary endpoint success. The final analysis of the primary endpoints will be triggered when at least 92 PP-EFF participants with symptomatic mild, moderate, or severe COVID-19 or approximately 43 PP-EFF participants with symptomatic moderate or severe COVID-19 have accrued. Also, in order to be able to respond to the unexpected and rapidly evolving COVID-19 pandemic situation globally, other factors such as requests by government or public health agencies may also be factored into the decision-making to unblind the study for the final analysis, but this always occurs in consultation with lead regulatory agencies.

In order to address the uncertainty around the COVID-19 circulation in the study population, an active monitoring of the blinded endpoint accruals for the 2 primary endpoints will be performed by the sponsor. The sponsor may choose to switch 1 of the 2 primary endpoints to a secondary or exploratory endpoint prior to unblinding, if it is determined that the accrual for the endpoint is likely to fall well short of the target number. The decision will be based solely on the total numbers of blinded endpoints accrued without consideration of treatment group assignment. The decision will not be based on unblinded VE estimates. If this option is triggered, the remaining primary endpoint will be analysed using one-sided Type I error rate of 0.025 (i.e., two-sided 95% CI). The details for the criteria to be used to act on this option will be included in the SAP.

The final analysis will be performed when the last participant completes the last visit (12 months after the last study vaccination) or discontinues earlier. The EOS analysis will be performed when all participants in the Neutralisation Assay Subset have completed the last study visit or discontinued earlier.

##### 7.3.2 Immunogenicity Analysis and Correlates of Risk

The primary and secondary immunogenicity analyses will be performed using the PP-IMM and ITT-IMM analysis populations.

For the serum antibody levels measured by microneutralization and HAI assays, the geometric mean at each study visit, the geometric mean fold rises (GMFRs) comparing to Day 0 (baseline) at each follow-up study visit, along with 95% CI will be summarised by study vaccine group. The 95% CI will be calculated based on the t distribution of the log-transformed values for geometric means or GMFRs, then back transformed to the original scale for presentation. The seroconversion rate, proportion of participants with ≥ 4 fold rises if naïve at baseline, along with 95% CIs based on the Clopper-Pearson method will be summarised by study vaccine group at each follow-up study visit.

Similar summaries will be generated for the other immunogenicity endpoints and other assays if conducted.

Correlates of risk will be assessed where immune responses are measured in all participants, and the results compared between participants who experience a SARS-CoV-2 event and participants who do not. The goal of this analysis is to assess correlates of risk of SARS-CoV-2 infection (and potential other secondary endpoints) in the study vaccine group by comparing study vaccine-induced immune responses associated with COVID-19.

##### 7.3.3 Safety Analyses

Solicited AEs (e.g., local and general systemic reactogenicity symptoms) will be captured in the Safety Subset of approximately 2,000 participants and in the seasonal influenza vaccine co-administration sub-study of approximately 400 participants.

Numbers and percentages (with 95% CIs based on the Clopper-Pearson method) of participants with solicited local and systemic AEs through 7 days after each study vaccination will be summarised by study vaccine group and the maximum toxicity grade over 7 days after each study vaccination. The duration of solicited local and systemic AEs after each study vaccination will also be summarised by treatment group.

Unsolicited AEs will be coded by preferred term and system organ class using the latest version of the MedDRA and summarised by study vaccine group as well as by severity and relationship to study vaccine. AEs through 35 days after first study vaccination; all MAAEs and MAAEs related to study vaccine; SAEs; or AESI through EOS will be listed separately and summarised by study vaccine group.

Vital sign measurements will be summarised by study vaccine group using descriptive statistics at baseline and following study vaccination.

Concomitant medications will be provided in a data listing with preferred drug name as coded using the World Health Organisation drug dictionary.

###### 7.3.3.1 Safety: Study Vaccine-Associated Enhanced Disease

Continuous monitoring for study vaccine-associated enhanced disease will be performed through the CRO and sponsor medical monitors. These events will be monitored in real-time and after each confirmed respective case. The SMC will review this data at scheduled SMC meetings throughout the study or at an ad hoc meeting if the medical monitors would like a more immediate review of the data.

#### 7.4 Handling of Missing Data

For calculating geometric means and GMFR, immunogenicity values reported as below the lower level of quantification (LLOQ) will be replaced by 0.5 × LLOQ. Values that are greater than the upper level of quantitation (ULOQ) will be replaced by the ULOQ. Missing results will not be imputed.

#### 7.5 Planned Analyses Prior to Study Completion

Analysis of the data will be provided on an ongoing basis for confirming success and to review safety as the study progresses. The SAP will outline the sequential nature of these reviews.

### 8. REFERENCE LIST

Bengtsson KL, Song H, Stertman L, et al. Matrix-M adjuvant enhances antibody, cellular and protective immune responses of a Zaire Ebola/Makona virus glycoprotein (GP) nanoparticle vaccine in mice. Vaccine. 2016;34(16):1927-35.

Coleman CM, Venkataraman T, Liu YV, et al. MERS-CoV spike nanoparticles protect mice from MERS-CoV infection. Vaccine. 2017;35:1586-9.

Department of Health and Human Services (DHHS), Food and Drug Administration, Center for Biologics Evaluation and Research (US). Guidance for industry: Toxicity grading scale for healthy adult and adolescent volunteers enrolled in preventive vaccine clinical trials. September 2007 [cited 2020 Apr 01]. Available from: https://www.fda.gov/media/73679/download

Division of AIDS (DAIDS), National Institute of Allergy and Infectious Diseases, National Institutes of Health, US Department of Health and Human Services. Division of AIDS (DAIDS) table for grading the severity of adult and pediatric adverse events. July 2017 [cited 2020 Apr 01]. Available from: https://rsc.niaid.nih.gov/sites/default/files/daidsgradingcorrectedv21.pdf

NVX-CoV2373 vaccine protects cynomolgus macaque upper and lower airways against SARS-CoV-2 challenge

Guebre-Xabier M, Patel N, Tian J-H, et al. NVX-CoV2373 vaccine protects cynomolgus macaque upper and lower airways against SARS-CoV-2 challenge. bioRxiv 2020.08.18.256578; doi: https://doi.org/10.1101/2020.08.18.256578.

Habibzadeh P, Stoneman EK. The novel coronavirus: a bird's eye view. Int J Occup Environ Med. 2020;11(2):65-71.

Keech C, Albert G, Reed P, et al, First-in-Human Trial of a SARS-CoV-2 Recombinant Spike Protein Nanoparticle Vaccine. medRxiv 2020.08.05.20168435; doi: https://doi.org/10.1101/2020.08.05.20168435.

Liu YV, Massare MJ, Barnard DL, et al. Chimeric severe acute respiratory syndrome coronavirus (SARS-CoV) S glycoprotein and influenza matrix 1 efficiently form virus-like particles (VLPs) that protect mice against challenge with SARS-CoV. Vaccine. 2011;29(38):6606 13.

Mandolesi M, Sheward DJ, Hanke L, et al. SARS-CoV-2 protein subunit vaccination elicits potent neutralizing antibody responses. bioRxiv 2020:2020.07.31.228486.

SPEAC: Safety Platform for Emergency Vaccines. D2.3 Priority List of Adverse Events of Special Interest: COVID-19. V2.0; May 25, 2020.

Su S, Wong G, Shi W, et al. Epidemiology, genetic recombination, and pathogenesis of coronaviruses. Trends Microbiol. 2016;24(6):490-502.

Tian J-H, Patel N, Haupt R, et al. SARS-CoV-2 spike glycoprotein vaccine candidate NVX-CoV2373 elicits immunogenicity in baboons and protection in mice. bioRxiv 2020:2020.06.29.178509.

World Health Organization (WHO)b. Coronavirus disease 2019 (COVID-19) situation report 203. 2020 August 10. Retrieved 10 August 2020, from https://www.who.int/docs/default-source/coronaviruse/situation-reports/20200810-covid-19-sitrep-203.pdf?sfvrsn=aa050308_4.

Zou G. A modified poisson regression approach to prospective studies with binary data. Am J Epidemiol. 2004;159(7):702-706.

### 9. APPENDICES

#### 9.1 Appendix 1: List of Abbreviations

| Abbreviation | Term |
|---|---|
| ACE2 | Angiotensin-converting enzyme 2 |
| AE | Adverse event |
| AESI | Adverse event(s) of special interest |
| CFR | Code of Federal Regulations |
| CI | Confidence interval |
| COVID-19 | Coronavirus disease 2019 |
| EBOV GP | Ebolavirus glycoprotein |
| eCRF | Electronic case report form |
| EDC | Electronic data capture |
| ELISA | Enzyme-linked immunosorbent assay |
| EOS | End of study |
| FDA | United States Food and Drug Administration |
| GCP | Good Clinical Practice |
| GMEU | Geometric mean ELISA unit |
| GMFR | Geometric mean fold rise |
| GMT | Geometric mean titer |
| GOLD | Global Initiative for Chronic Obstructive Lung Disease |
| IB | Investigator's Brochure |
| ICF | Informed consent form |
| ICH | International Council for Harmonisation of Technical Requirements for Pharmaceuticals for Human Use |
| ID | Identification |
| IgG | Immunoglobulin G |
| IM | Intramuscular |
| IRT | Interactive Response Technology |
| ITT | Intent-to-treat |
| ITT-EFF | ITT efficacy |
| ITT-IMM | ITT immunogenicity |
| LLOQ | Lower limit of quantification |
| MAAE | Medically attended adverse event |
| MedDRA | Medical Dictionary for Regulatory Activities |

| **Abbreviation** | **Term** |
|---|---|
| N-protein | Nucleocapsid |
| OTC | Over-the-counter |
| PCR | Polymerase chain reaction |
| PIMMC | Potential immune-mediated medical conditions |
| PP | Per-protocol |
| PP-EFF | PP efficacy |
| PP-IMM | PP immunogenicity |
| REC | Research Ethics Committee |
| RR | Relative risk |
| RSV F | Respiratory syncytial virus fusion (protein) |
| S-protein | Spike |
| SAE | Serious adverse event |
| SAP | Statistical analysis plan |
| SARS-CoV | Severe acute respiratory syndrome coronavirus |
| SARS-CoV-2 | Severe acute respiratory syndrome coronavirus 2 |
| SARS-CoV-2 rS | SARS-CoV-2 recombinant spike (S) protein nanoparticle vaccine |
| Sf9 | *Spodoptera frugiperda* (insect cells) |
| SMC | Safety Monitoring Committee |
| SOE | Schedule of Events |
| ULOQ | Upper limit of quantitation |
| VE | Vaccine efficacy |

#### 9.2 Appendix 2: Study Governance

##### 9.2.1 Data Quality Assurance

This study will be conducted using the quality processes described in applicable procedural documents. The quality management approach to be implemented will be documented and will comply with current International Council for Harmonisation of Technical Requirements for Pharmaceuticals for Human Use (ICH) guidance on quality and risk management. All aspects of the study will be monitored for compliance with applicable government regulatory requirements, current Good Clinical Practice (GCP), the protocol, and standard operating procedures. The monitor will maintain current personal knowledge of the study through observation, review of study records and source documentation, and discussion of the conduct of the study with the investigator and personnel. eCRFs and EDC will be utilised. The EDC system is validated and compliant with US Title 21 Code of Federal Regulations (CFR) Part 11 and local regulations. Each person involved with the study will have an individual identification code and password that allows for record traceability.

Important protocol deviations, should they occur during the study, will be presented in Section 10.2 of the clinical study report.

##### 9.2.2 Investigator Obligations

The following administrative items are meant to guide the investigator in the conduct of the study and may be subject to change based on industry and government standard operating procedures, working practice documents, or guidelines. Changes will be reported to the Research Ethics Committee (REC) but will not result in protocol amendments.

###### 9.2.2.1 Confidentiality

All laboratory specimens, evaluation forms, reports, and other records will be identified in a manner designed to maintain participant confidentiality. All records will be kept in a secure storage area with limited access. Clinical information will not be released without the written permission of the participant, except as necessary for monitoring and auditing by the sponsor, its designee, relevant regulatory authority(ies), or the REC.

The investigator and all employees and co-workers involved with this study may not disclose or use for any purpose other than performance of the study, any data, record, or other unpublished, confidential information disclosed to those individuals for the purpose of the study. Prior written agreement from the sponsor or its designee must be obtained for the disclosure of any said confidential information to other parties.

###### 9.2.2.2 Institutional Review

Prior to initiation of a study site, regulatory authority regulations and the ICH E6(R2) guidelines require that approval be obtained from the REC before participation of human participants in research studies. Before study onset, the protocol, informed consent, advertisements to be used for the recruitment of study participants, and any other written information regarding this study to be provided to the participant must be approved by the REC. Documentation of all REC approvals and of the REC compliance with the ICH E6(R2) guidelines will be maintained by the study site and will be available for review by the sponsor or its designee.

All REC approvals should be signed by the REC chairman or designee and must identify the REC name and address, the clinical protocol by title or protocol number or both and the date approval or a favorable opinion was granted.

###### 9.2.2.3 Participant Consent

Written informed consent in compliance with US Title 21 CFR Part 50 and local regulatory authority requirements shall be obtained from each participant before he or she enters the study or before any unusual or nonroutine procedure that involves risk to the participant is performed. If any institution-specific modifications to study-related procedures are proposed or made by the study site, the consent should be reviewed by the sponsor or its designee or both before REC submission. Once reviewed, the investigator will submit the ICF to the REC for review and approval before the start of the study. If the ICF is revised during the course of the study, all active participating participants must sign the revised form.

Before recruitment and enrolment, each prospective participant will be given a full explanation of the study and be allowed to read the approved ICF. Once the investigator is assured that the participant understands the implications of participating in the study, the participant will be asked to give his or her consent to participate in the study by signing the ICF.

The investigator or designee will provide a copy of the ICF to the participant. The original form shall be maintained in the participant's medical records at the study site.

###### 9.2.2.4 Study Reporting Requirements

By participating in this study, the investigator agrees to submit reports of SAEs according to the timeline and method outlined in this protocol. In addition, the investigator agrees to submit annual reports to his or her REC as appropriate.

###### 9.2.2.5 Financial Disclosure and Obligations

The investigator is required to provide financial disclosure information to allow the sponsor to submit the complete and accurate certification or disclosure statements required under US Title 21 CFR Part 54 and local regulations. In addition, the investigator must provide to the sponsor a commitment to promptly update this information if any relevant changes occur during the course of the investigation and for 1 year following the completion of the study.

Neither the sponsor nor PPD nor the study site is financially responsible for further testing or treatment of any medical condition that may be detected during the screening process. In addition, in the absence of specific arrangements, neither the sponsor nor PPD nor the study site is financially responsible for further treatment of the disease under study.

###### 9.2.2.6 Investigator Documentation

Prior to beginning the study, the investigator will be asked to comply with ICH E6(R2) Section 8.2, US Title 21 of the CFR, and local regulations by providing essential documents, including but not limited to, the following:

- REC approval.
- An original investigator-signed investigator agreement page of the protocol.
- Curriculum vitae for the principal investigator and each sub-investigator. Current licensure must be noted on the curriculum vitae. They will be signed and dated by the principal investigators and sub-investigators at study start-up, indicating that they are accurate and current.
- Financial disclosure information to allow the sponsor to submit complete and accurate certification or disclosure statements required under US Title 21 CFR Part 54 and local regulations. In addition, the investigators must provide to the sponsor a commitment to promptly update this information if any relevant changes occur during the course of the investigation and for 1 year after the completion of the study.
- An REC-approved ICF, samples of study site advertisements for recruitment for this study, and any other written information about this study that is to be provided to the participant.
- Laboratory certifications and reference ranges for any local laboratories used by the study site, in accordance with US Title 42 CFR Part 493 and local regulations.

###### 9.2.2.7 Study Conduct

The investigator agrees that the study will be conducted according to the principles of ICH E6(R2). The investigator will conduct all aspects of this study in accordance with all national, state, and local laws or regulations. The study will be conducted in compliance with the protocol, current GCP guidelines – adopting the principles of the Declaration of Helsinki – and all applicable regulatory requirements.

Prior to study initiation, the protocol and the informed consent documents will be reviewed and approved by the sponsor and an appropriate ethics committee. Any amendment to the protocol or consent materials must also be approved by the study sponsor and REC and must be submitted, notified, or approved to the regulatory authority, as required, before they are implemented.

###### 9.2.2.8 Case Report Forms and Source Documents

Site personnel will maintain source documentation, enter participant data into the eCRF as accurately as possible, and will rapidly respond to any reported discrepancies.

eCRFs and EDC will be utilised. The EDC system is validated and compliant with US Title 21 CFR Part 11 and local regulations. Each person involved with the study will have an individual identification code and password that allows for record traceability. Thus, the system, and any subsequent investigative reviews, can identify coordinators, investigators, and individuals who have entered or modified records, as well as the time and date of any modifications. There may be an internal quality review audit of the data and additional reviews by the clinical monitor.

Each eCRF is presented as an electronic copy, allowing data entry by site personnel, who can add and edit data, add new participants, identify and resolve discrepancies, and view records. This system provides immediate direct data transfer to the database, as well as immediate detection of discrepancies, enabling site coordinators to resolve and manage discrepancies in a timely manner.

Paper copies of the eCRFs and other database reports may be printed and signed by the investigator. This system provides site personnel, monitors, and reviewers with access to hardcopy audits, discrepancy reviews, and investigator comment information.

###### 9.2.2.9 Adherence to Protocol

The investigator agrees to conduct the study as outlined in this protocol, in accordance with ICH E6(R2) and all applicable guidelines and regulations.

###### 9.2.2.10 Reporting Adverse Events

By participating in this study, the investigator agrees to submit reports of SAEs according to the timeline and method outlined in this protocol. In addition, the investigator agrees to submit annual reports to his or her REC as appropriate. The investigator also agrees to provide the sponsor with an adequate report, if applicable, shortly after completion of the investigator's participation in the study.

###### 9.2.2.11 Investigator's Final Report

Upon completion of the study, the investigator, where applicable, should inform the institution; the investigator/institution should provide the REC with a summary of the study's outcome and the sponsor and regulatory authority(ies) with any reports as required. Interim reports are expected to be provided to regulatory authorities to allow study vaccine development advancement given the pandemic situation. These reports are planned to be aggregate and at the study vaccine level unless the SMC deems additional data at the individual level (e.g., select listings of select participants) will be beneficial. In such a case, a firewall will be in place to maintain the blind for those individuals involved in the study conduct to ensure unbiased assessment continue.

###### 9.2.2.12 Records Retention

Essential documents should be retained until at least 2 years after the last approval of a marketing application in an ICH region and until there are no pending or contemplated marketing applications in an ICH region or until at least 2 years have elapsed since the formal discontinuation of clinical development of the study vaccine or per local regulation, whichever is longer. These documents should be retained for a longer period, however, if required by applicable regulatory requirements or by an agreement with the sponsor. It is the sponsor's responsibility to inform the investigator/institution as to when these documents are no longer need to be retained.

###### 9.2.2.13 Publications

After completion of the study, the data may be considered for reporting at a scientific meeting or for publication in a scientific journal. In these cases, the sponsor will be responsible for these activities and will work with the investigators to determine how the manuscript is written and edited, the number and order of authors, the publication to which it will be submitted, and any other related issues. The sponsor has final approval authority over all such issues.

Data are the property of the sponsor and cannot be published without their prior authorisation, but data and any publication thereof will not be unduly withheld.

##### 9.2.3 Study Management

###### 9.2.3.1 Monitoring

###### 9.2.3.1.1 Monitoring of the Study

The clinical research organisation clinical monitor, as a representative of the sponsor, is obligated to follow the study closely. In doing so, the monitor will visit the investigator and study site at periodic intervals in addition to maintaining necessary telephone and email contact. The monitor will maintain current personal knowledge of the study through observation, review of study records and source documentation, and discussion of the conduct of the study with the investigator and personnel. The monitor will be blinded to study vaccine assignment. A separate unblinded study monitor will be responsible for drug accountability.

All aspects of the study will be carefully monitored by the sponsor or its designee for compliance with applicable government regulation with respect to current ICH E6(R2) guidelines and standard operating procedures.

###### 9.2.3.1.2 Inspection of Records

The investigator and institution involved in the study will permit study-related monitoring, audits, REC review, and regulatory inspections by providing direct access to all study records. In the event of an audit, the investigator agrees to allow the sponsor, their representatives, or the regulatory authority access to all study records.

The investigator should promptly notify the sponsor of any audits scheduled by any regulatory authorities and promptly forward copies of any audit reports received to the sponsor.

###### 9.2.3.2 Management of Protocol Amendments and Deviations

###### 9.2.3.2.1 Modification of the Protocol

This is a Phase 3 study to evaluate the efficacy, immunogenicity, and safety of SARS-CoV-2 rS with Matrix-M1 adjuvant. This protocol is written with some flexibility to accommodate the evolving pandemic and urgency for efficacious vaccine availability. Modifications to the dose, dosing regimen, and/or clinical or laboratory procedures currently outlined below may be required to achieve the scientific goals of the study objectives and/or to ensure appropriate safety monitoring of the study participants:

- The timing of procedures for assessment of safety procedures may be modified based on newly available safety and tolerability data or evolving COVID-19 data.

- Up to an additional 25 mL of blood may be drawn for safety or immunogenicity analyses. The total blood volume withdrawn from any single participant will not exceed the maximum allowable volume during his or her participation in the entire study.
- Additional database freezes may occur as the study evolves and should the ongoing epidemic progression warrant rapid decision-making on product manufacturing. The study will continue in a blinded fashion (at the participant level) until the EOS.
- Rapid diagnostic testing for SARS-CoV-2 by point-of-care tests may be available and substituted for centralised testing if accepted by regulatory authorities as a secondary endpoint in this study and hold validity for study vaccine advancement.

It is understood that the current study may employ some or none of the alterations described above. Any changes in this research activity, except those necessary to remove an apparent immediate hazard to the participant, must be reviewed and approved by the sponsor or designee. Amendments to the protocol must be approved by the REC, and regulatory authority where applicable, before participants can be enrolled into an amended protocol.

###### 9.2.3.2.2 Protocol Deviations

The investigator or designee must document and explain in the participant's source documentation any deviation from the approved protocol. The investigator may implement a deviation from, or a change to, the protocol to eliminate an immediate hazard to study participants without prior REC approval. As soon as possible after such an occurrence, the implemented deviation or change, the reasons for it, and any proposed protocol amendments should be submitted to the REC for review and approval, to the sponsor for agreement, and to the regulatory authorities, if required.

A protocol deviation is any change, divergence, or departure from the study design or procedures defined in the protocol. An important deviation (sometimes referred to as a major or significant deviation) is a subset of protocol deviations that leads to a participant being discontinued from the study or significantly affects the participant's rights, safety, or well-being and/or the completeness, accuracy, and reliability of the study data. An important deviation can include nonadherence to inclusion or exclusion criteria or nonadherence to regulatory authority including ICH E6(R2) guidelines.

Protocol deviations will be documented by the clinical monitor throughout the course of monitoring visits. The investigator will be notified in writing by the monitor of deviations. The REC should be notified of all protocol deviations, if appropriate, in a timely manner.

##### 9.2.3.3 Study Termination

Although the sponsor has every intention of completing the study, they reserve the right to discontinue it at any time for clinical or administrative reasons.

The end of the study is defined as the date on which the last participant completes the last study visit (including the EOS visit and any additional long-term follow-up). Any additional long-term follow-up that is required for monitoring of the resolution of an AE or finding may be appended to the clinical study report.

##### 9.2.3.4 Final Report

Regardless of whether the study is completed or prematurely terminated, the sponsor will ensure that clinical study reports are prepared and provided to regulatory agency(ies) as required by the applicable regulatory requirement(s). The sponsor will also ensure that clinical study reports in marketing applications meet the standards of the ICH E3: Structure and Content of Clinical Study Reports.

Where required by applicable regulatory requirements, an investigator signatory will be identified for the approval of the clinical study report. The investigator will be provided reasonable access to statistical tables, figures, and relevant reports and will have the opportunity to review complete study results.

#### 9.3 Appendix 3: FDA Toxicity Grading Scales

##### Table 9-1 FDA Toxicity Grading Scale for Clinical Abnormalities (Local and General Systemic Reactogenicity)

| **Local Reaction to Injectable Product** | | | | |
|---|---|---|---|---|
| **Clinical Abnormality** | **Mild (Grade 1)** | **Moderate (Grade 2)** | **Severe (Grade 3)** | **Potentially Life Threatening (Grade 4)** |
| Pain | Does not interfere with activity | Repeated use of nonnarcotic pain reliever >24 hours or interferes with activity | Any use of narcotic pain reliever or prevents daily activity | ER visit or hospitalization |
| Tenderness | Mild discomfort to touch | Discomfort with movement | Significant discomfort at rest | ER visit or hospitalization |
| Erythema/redness[a] | 2.5 – 5 cm | 5.1 – 10 cm | > 10 cm | Necrosis or exfoliative dermatitis |
| Induration/swelling[b] | 2.5 – 5 cm and does not interfere with activity | 5.1 – 10 cm or interferes with activity | > 10 cm or prevents daily activity | Necrosis |
| **Systemic (General)** | | | | |
| **Clinical Abnormality** | **Mild (Grade 1)** | **Moderate (Grade 2)** | **Severe (Grade 3)** | **Potentially Life Threatening (Grade 4)** |
| Fever (°C)<br>(°F) | 38.0 – 38.4<br>100.4 – 101.1 | 38.5 – 38.9<br>101.2 – 102.0 | 39.0 – 40<br>102.1 – 104 | > 40<br>> 104 |
| Nausea/vomiting | No interference with activity or 1 – 2 episodes/24 hours | Some interference with activity or >2 episodes/24 hours | Prevents daily activity, or requires outpatient IV hydration | ER visit or hospitalization for hypotensive shock |
| Headache | No interference with activity | Repeated use of nonnarcotic pain reliever >24 hours or some interference with activity | Significant; any use of narcotic pain reliever or prevents daily activity | ER visit or hospitalization |
| Fatigue/Malaise | No interference with activity | Some interference with activity | Significant; prevents daily activity | ER visit or hospitalization |
| Myalgia | No interference with activity | Some interference with activity | Significant; prevents daily activity | ER visit or hospitalization |
| Arthralgia | No interference with activity | Some interference with activity | Significant; prevents daily activity | ER visit or hospitalization |

Abbreviations: DHHS = Department of Health and Human Services; ER = emergency room; FDA = United States Food and Drug Administration.

[a] In addition to grading the measured local reaction at the greatest single diameter, the measurement should be recorded as a continuous variable.

[b] Induration/swelling should be evaluated and graded using the functional scale as well as the actual measurement.

[c] Oral temperature; no recent hot or cold beverages.

Source: DHHS 2007.

**Table 9-2 FDA Toxicity Grading Scale for Clinical Abnormalities (Vital Signs)**

| Vital Signs | Mild (Grade 1) | Moderate (Grade 2) | Severe (Grade 3) | Potentially Life Threatening (Grade 4) |
|---|---|---|---|---|
| Tachycardia (bpm) | 101 – 115 | 116 – 130 | > 130 | ER visit or hospitalization for arrhythmia |
| Bradycardia (bpm)[a] | 50 – 54 | 45 – 49 | < 45 | ER visit or hospitalization for arrhythmia |
| Hypertension (systolic) (mm Hg) | 141 – 150 | 151 – 155 | > 155 | ER visit or hospitalization for malignant hypertension |
| Hypertension (diastolic) (mm Hg) | 91 – 95 | 96 – 100 | > 100 | ER visit or hospitalization for malignant hypertension |
| Hypotension (systolic) (mm Hg) | 85 – 89 | 80 – 84 | < 80 | ER visit or hospitalization for hypotensive shock |
| Respiratory Rate (breaths per minute) | 17 – 20 | 21 – 25 | > 25 | Intubation |

Abbreviations: DHHS = Department of Health and Human Services; ER = emergency room; FDA = United States Food and Drug Administration.

Note: Participant should be at rest for all vital sign measurements, with toxicity scored on day of study vaccination (pre- and post-dose).

[a] When resting heart rate is between 60 – 100 bpm. Use clinical judgement when characterising bradycardia among some healthy participant populations (e.g., conditioned athletes).

Source: DHHS 2007.

#### 9.4 Appendix 4: Listings of Adverse Events of Special Interest

Because it has been hypothesised that immunisations with or without adjuvant may be associated with autoimmunity, regulatory authorities have requested that Novavax instruct investigators to be especially vigilant regarding the PIMMC listed in Table 9-3.

**Table 9-3 Potential Immune-Mediated Medical Conditions (PIMMC)**

| Categories | Diagnoses (as MedDRA Preferred Terms) |
|---|---|
| Neuro-inflammatory Disorders: | Acute disseminated encephalomyelitis (including site specific variants: e.g., noninfectious encephalitis, encephalomyelitis, myelitis, myeloradiculomyelitis), cranial nerve disorders including paralyses/paresis (e.g., Bell's palsy), generalised convulsion, Guillain-Barre syndrome (including Miller Fisher syndrome and other variants), immune-mediated peripheral neuropathies and plexopathies (including chronic inflammatory demyelinating polyneuropathy, multifocal motor neuropathy and polyneuropathies associated with monoclonal gammopathy), myasthenia gravis, multiple sclerosis, narcolepsy, optic neuritis, transverse myelitis, uveitis |
| Musculoskeletal and Connective Tissue Disorders: | Antisynthetase syndrome, dermatomyositis, juvenile chronic arthritis (including Still's disease), mixed connective tissue disorder, polymyalgia rheumatic, polymyositis, psoriatic arthropathy, relapsing polychondritis, rheumatoid arthritis, scleroderma (including diffuse systemic form and CREST syndrome), spondyloarthritis (including ankylosing spondylitis, reactive arthritis [Reiter's Syndrome] and undifferentiated spondyloarthritis), systemic lupus erythematosus, systemic sclerosis, Sjogren's syndrome |
| Vasculidities: | Large vessels vasculitis (including giant cell arteritis such as Takayasu's arteritis and temporal arteritis), medium sized and/or small vessels vasculitis (including polyarteritis nodosa, Kawasaki's disease, microscopic polyangiitis, Wegener's granulomatosis, Churg–Strauss syndrome [allergic granulomatous angiitis], Buerger's disease [thromboangiitis obliterans], necrotising vasculitis and anti-neutrophil cytoplasmic antibody [ANCA] positive vasculitis [type unspecified], Henoch-Schonlein purpura, Behcet's syndrome, leukocytoclastic vasculitis) |
| Gastrointestinal Disorders: | Crohn's disease, celiac disease, ulcerative colitis, ulcerative proctitis |
| Hepatic Disorders: | Autoimmune hepatitis, autoimmune cholangitis, primary sclerosing cholangitis, primary biliary cirrhosis |
| Renal Disorders: | Autoimmune glomerulonephritis (including IgA nephropathy, glomerulonephritis rapidly progressive, membranous glomerulonephritis, membranoproliferative glomerulonephritis, and mesangioproliferative glomerulonephritis. |
| Cardiac Disorders: | Autoimmune myocarditis/cardiomyopathy |
| Skin Disorders: | Alopecia areata, psoriasis, vitiligo, Raynaud's phenomenon, erythema nodosum, autoimmune bullous skin diseases (including pemphigus, pemphigoid and dermatitis herpetiformis), cutaneous lupus erythematosus, morphoea, lichen planus, Stevens-Johnson syndrome, Sweet's syndrome |
| Haematologic Disorders: | Autoimmune hemolytic anemia, autoimmune thrombocytopenia, antiphospholipid syndrome, thrombocytopenia |
| Metabolic Disorders: | Autoimmune thyroiditis, Grave's or Basedow's disease, Hashimoto thyroiditis[a], diabetes mellitus type 1, Addison's disease |

| Categories | Diagnoses (as MedDRA Preferred Terms) |
|---|---|
| Other Disorders: | Goodpasture syndrome, idiopathic pulmonary fibrosis, pernicious anemia, sarcoidosis |

Abbreviations: ANCA = anti-neutrophil cytoplasmic antibody; IgA = immunoglobulin A; MedDRA = Medical Dictionary for Regulatory Activities.
[a] For Hashimoto thyroiditis: new onset only.

AESIs relevant to COVID-19 are listed in Table 9-4. The list is not intended to be exhaustive, nor does it exclude the possibility that other diagnoses may be AESI. It is anticipated that additional AESI may be associated with COVID-19. Investigators should stay updated regarding such public health notifications.

**Table 9-4 Adverse Events of Special Interest Relevant to COVID-19**

| Body System | Diagnoses[a] |
|---|---|
| Immunologic | Enhanced disease following immunisation, cytokine release syndrome related to COVID-19[b], Multisystem inflammatory syndrome in children (MIS-C) |
| Respiratory | Acute respiratory distress syndrome (ARDS) |
| Cardiac | Acute cardiac injury including:<br>• Microangiopathy<br>• Heart failure and cardiogenic shock<br>• Stress cardiomyopathy<br>• Coronary artery disease<br>• Arrhythmia<br>• Myocarditis, pericarditis |
| Haematologic | Coagulation disorder<br>• Deep vein thrombosis<br>• Pulmonary embolus<br>• Cerebrovascular stroke<br>• Limb ischemia<br>• Hemorrhagic disease<br>• Thrombotic complications |
| Renal | Acute kidney injury |
| Gastrointestinal | Liver injury |
| Neurologic | Guillain-Barré Syndrome, anosmia, ageusia, meningoencephalitis |
| Dermatologic | Chilblain-like lesions, single organ cutaneous vasulitis, erythema multiforme |

Abbreviations: COVID-19 = coronavirus disease 2019; DAIDS = Division of AIDS.
[a] COVID-19 manifestations associated with more severe presentation and decompensation with consideration of enhanced disease potential (SPEAC 2020).
[b] Cytokine release syndrome related to COVID-19 infection is a disorder characterised by nausea, headache, tachycardia, hypotension, rash, and/or shortness of breath (DAIDS 2017).

### CLINICAL STUDY PROTOCOL

### A PHASE 3, RANDOMISED, OBSERVER-BLINDED, PLACEBO-CONTROLLED TRIAL TO EVALUATE THE EFFICACY AND SAFETY OF A SARS-COV-2 RECOMBINANT SPIKE PROTEIN NANOPARTICLE VACCINE (SARS-COV-2 RS) WITH MATRIX-M1™ ADJUVANT IN ADULT PARTICIPANTS 18-84 YEARS OF AGE IN THE UNITED KINGDOM

| | |
|---|---|
| **Investigational Materials:** | SARS-CoV-2 rS with Matrix-M1™ adjuvant |
| **Protocol Number:** | 2019nCoV-302 |
| **EudraCT Number:** | 2020-004123-16 |
| **Sponsor:** | Novavax, Inc.<br>21 Firstfield Road<br>Gaithersburg, MD 20878<br>United States |
| **Responsible Clinical Operations Manager:** | [redacted] Clinical Operations<br>Telephone: [redacted] |
| **Responsible Clinical Development Lead:** | [redacted] Clinical Development<br>Telephone: [redacted] |
| **Chief Investigator:** | [redacted]<br>Telephone: [redacted] |
| **Version – Date:** | Version 4.0 – 25 February 2021 |

**Prior Versions:**

Version 1.0 – 24 August 2020
Version 1.1 – 17 September 2020
Version 1.2 – 21 September 2020
Version 2.0 – 23 October 2020
Version 3.0 – 23 December 2020

**CONFIDENTIAL**

The information in this document is considered privileged and confidential by Novavax, Inc. and may not be disclosed to others except to the extent necessary to obtain Independent Ethics Committee (IEC) approval and informed consent, or as required by national and local laws. Persons to whom this information is disclosed must be informed that this information is privileged and confidential and that it should not be further disclosed.

The study will be conducted according to the International Council for Harmonisation of Technical Requirements for Pharmaceuticals for Human Use Guideline E6(R2): Good Clinical Practice.

### SIGNATURE PAGE

| | |
|---|---|
| **PROTOCOL TITLE:** | A Phase 3, Randomised, Observer-Blinded, Placebo-Controlled Trial to Evaluate the Efficacy and Safety of a SARS-CoV-2 Recombinant Spike Protein Nanoparticle Vaccine (SARS-CoV-2 rS) with Matrix-M1™ Adjuvant in Adult Participants 18-84 Years of Age in the United Kingdom |
| **PROTOCOL NUMBER:** | 2019-nCoV-302 |
| **EUDRACT NUMBER:** | 2020-004123-16 |

______________________________ ______________________________
[redacted]
Novavax, Inc.
Date

______________________________ ______________________________
[redacted] Clinical Operations
Novavax, Inc.
Date

______________________________ ______________________________
[redacted]
Date

### INVESTIGATOR PROTOCOL AGREEMENT PAGE

I agree to conduct the study as outlined in the protocol titled "A Phase 3, Randomised, Observer-Blinded, Placebo-Controlled Trial to Evaluate the Efficacy and Safety of a SARS-CoV-2 Recombinant Spike Protein Nanoparticle Vaccine (SARS-CoV-2 rS) with Matrix-M1™ Adjuvant in Adult Participants 18-84 Years of Age in the United Kingdom" in accordance with all guidelines, including International Council for Harmonisation of Technical Requirements for Pharmaceuticals for Human Use guidelines, and all applicable government regulations. I have read and understand all sections of the protocol.

____________________________________ _______________________

Signature of Investigator Date

____________________________________

Printed Name of Investigator

### LIST OF FIGURES

Figure 1a: Trial Schema ........................................................................................................ 43

Figure 2b: Original Trial Schema ........................................................................................... 44

### PROTOCOL SYNOPSIS

**PROTOCOL NO.:** 2019nCoV-302

**TITLE:** A Phase 3, Randomised, Observer-Blinded, Placebo-Controlled Trial to Evaluate the Efficacy and Safety of a SARS-CoV-2 Recombinant Spike Protein Nanoparticle Vaccine (SARS-CoV-2 rS) with Matrix-M1™ Adjuvant in Adult Participants 18-84 Years of Age in the United Kingdom

**STUDY PHASE:** Phase 3

**STUDY SITES:** 33 sites across the United Kingdom (UK).

**OBJECTIVES:**

- **The primary objective is:**
  - To demonstrate the efficacy of SARS-CoV-2 rS with Matrix-M1 adjuvant in the prevention of virologically confirmed (by polymerase chain reaction [PCR] to severe acute respiratory syndrome coronavirus 2 [SARS-CoV-2]), symptomatic coronavirus disease 2019 (COVID-19), when given as a 2-dose vaccination regimen, as compared to placebo, in serologically negative (to SARS-CoV-2) adults.
- **The secondary objectives are:**
  - To demonstrate the efficacy of SARS-CoV-2 rS with Matrix-M1 adjuvant in the prevention of virologically confirmed (by PCR to SARS-CoV-2), symptomatic COVID-19, when given as a 2-dose vaccination regimen, as compared to placebo, in adults regardless of their serostatus at baseline.
  - To assess the efficacy of SARS-CoV-2 rS with Matrix-M1 adjuvant in adult participants requiring specific medical interventions as compared to placebo.
  - To assess the efficacy of SARS-CoV-2 rS with Matrix-M1 adjuvant on mild COVID-19 symptoms as compared to placebo.
  - To assess the efficacy of SARS-CoV-2 rS with Matrix-M1 adjuvant on occurrence of asymptomatic or undetected infections with SARS-CoV-2 as compared to placebo.
  - In a subset of adult participants, to evaluate the immunogenicity of SARS-CoV-2 rS with Matrix-M1 adjuvant as compared to placebo.
  - To evaluate safety in terms of serious adverse events (SAEs) and medically attended adverse events (MAAEs) related to study vaccination in all adult participants during the entire study period.
  - To evaluate safety in terms of adverse events of special interest (AESIs), which encompasses potential immune-mediated medical conditions (PIMMCs) and AESIs relevant to COVID-19 including possible vaccine-enhanced disease, in all adult participants at any time after the first dose. In a subset of adult participants,

to evaluate the safety and reactogenicity in terms of solicited local and systemic adverse events (AEs) after the initial set of study vaccinations.

- To evaluate safety in terms of all MAAEs for 14 days after second vaccination and unsolicited AEs for 21 days after first study vaccination and 28 days after second study vaccination in the initial set of vaccinations.
- To assess the duration of vaccine efficacy (measured by all efficacy endpoints) in initial active vaccine recipients vs. crossover (delayed) active vaccine recipients.

- **The exploratory objectives are:**
  - In a subset of adult participants, to evaluate the safety and immunogenicity of SARS-CoV-2 rS with Matrix-M1 adjuvant in the initial set of vaccinations when co-administered with a licensed seasonal influenza vaccine.
  - In a subset of adult participants unblinded before the crossover, to explore the efficacy and safety of SARS-CoV-2 rS with Matrix-M1 adjuvant in the initial set of vaccinations and/or an approved or deployed SARS-CoV-2 vaccine.

**ENDPOINTS**

- **The primary endpoint is**:
  - First occurrence of virologically confirmed (by PCR to SARS-CoV-2), symptomatic mild, moderate, or severe COVID-19 with onset at least 7 days after second study vaccination (e.g., Day 28) in the initial set of vaccinations in serologically negative (to SARS-CoV-2) adult participants at baseline until the endpoint-driven efficacy analysis is triggered by the occurrence of a prespecified number of blinded endpoints.
- **The key secondary endpoint is:**
  - First occurrence of virologically confirmed (by PCR to SARS-CoV-2), symptomatic moderate or severe COVID-19 with onset at least 7 days after second study vaccination (e.g., Day 28) in the initial set of vaccinations in serologically negative (to SARS-CoV-2) adult participants at baseline until the endpoint-driven efficacy analysis is triggered by the occurrence of a prespecified number of blinded endpoints.
- **The other secondary endpoints are**:
  - First occurrence of virologically confirmed (by PCR to SARS-CoV-2), symptomatic severe COVID-19 with onset at least 7 days after second study vaccination (e.g., Day 28) in the initial set of vaccinations in serologically negative (to SARS-CoV-2) adult participants at baseline until the endpoint-driven efficacy analysis is triggered by the occurrence of a prespecified number of blinded endpoints.
  - First occurrence of virologically confirmed (by PCR to SARS-CoV-2), symptomatic mild, moderate, or severe COVID-19, with onset at least 7 days after second study vaccination (e.g., Day 28) in the initial set of vaccinations in adult participants regardless of their serostatus at baseline.

- First occurrence of COVID-19 requiring hospitalisation, intensive care unit (ICU) admission or mechanical ventilation linked to any virologically confirmed (by PCR to SARS-CoV-2) COVID-19 with onset at least 7 days after second study vaccination (e.g., Day 28) in the initial set of vaccinations in adult participants regardless of their serostatus at baseline.
- First occurrence of virologically confirmed (by PCR to SARS-CoV-2), symptomatic mild COVID-19 (with no progression to moderate or severe COVID-19 during the course of the COVID-19 episode) with onset at least 7 days after second study vaccination (e.g., Day 28) in the initial set of vaccinations in adult participants, regardless of their serostatus at baseline.
- First occurrence of laboratory-confirmed (by PCR or nucleocapsid [N]-protein serology to SARS-CoV-2) symptomatic or asymptomatic COVID-19 with onset at least 7 days after second study vaccination (e.g., Day 28) in the initial set of vaccinations in adult participants with negative serostatus at baseline.
- Analysis of antibodies binding to the SARS-CoV-2 spike (S) protein by enzyme-linked immunosorbent assay (ELISA) at Day 0 (baseline) and Day 35 (14 days after second study vaccination).
- Analysis of antibodies binding to the SARS-CoV-2 S protein by ELISA at Crossover Day 0 visit (baseline) and Crossover Day 35 visit (14 days after second study vaccination).
- The occurrence and relationship to study vaccination of SAEs and MAAEs related to study vaccination (in all adult participants) during the entire study period.
- The occurrence and relationship to study vaccination of AESIs and PIMMCs (in all adult participants) during the entire study period.
- The occurrence and severity of reactogenicity in terms of solicited local and systemic AEs (in sub-study participants) after the initial set of vaccinations.
- The occurrence, severity, and relationship to study vaccination of all MAAEs for 14 days after second vaccination and unsolicited AEs (in all adult participants) for 21 days after first study vaccination and 28 days after second study vaccination following the initial set of vaccinations.
- Relative vaccine efficacy (measured by all efficacy endpoints) in initial active vaccine recipients vs. crossover (delayed) active vaccine recipients.

• **Exploratory endpoints are:**

- First occurrence of virologically confirmed (by PCR to SARS-CoV-2), symptomatic mild, moderate, or severe COVID-19 with onset at least 14 days after first study vaccination (e.g., Day 14) in the initial set of vaccinations in serologically negative (to SARS-CoV-2) adult participants at baseline until the endpoint-driven efficacy analysis is triggered by the occurrence of a prespecified number of blinded endpoints.

- Any occurrence of serologic conversion (by serology to SARS-CoV-2 N protein) between baseline and 1 year after last study vaccination in the initial set of vaccinations in adult participants seronegative at baseline.
- Any occurrence of serologic conversion (by serology to SARS-CoV-2 N protein) between Crossover Day 0 visit and the end of study (EOS) visit in the second set of vaccinations in adult participants seronegative at baseline.
- Analysis of the proportion of adult participants with evidence of seroconversion as demonstrated by binding antibodies to the SARS-CoV-2 S protein by ELISA at Day 35 (14 days after second study vaccination) and Crossover Day 35 visit (14 days after the Crossover Day 21visit).
- Analysis of the cell-mediated immune responses (for example, as measured by enzyme-linked immune absorbent spot (ELISpot) ± intracellular cytokine staining) on Days 0 (baseline) and 35 (14 days after second study vaccination) in the initial set of vaccinations.
- SARS-CoV-2 neutralisation as measured by virus neutralisation assay (VNA; wild-type virus and/or pseudovirion expressing SARS-CoV-2 S protein) at Day 0 (baseline) and Day 35 (14 days after second study vaccination) in the initial set of vaccinations.
- Analysis of the safety and immunogenicity of SARS-CoV-2 rS with Matrix-M1 adjuvant when co-administered with seasonal influenza vaccine in a subset population.
- Analysis of the efficacy and safety of SARS-CoV-2 rS with Matrix-M1 adjuvant and/or an approved or deployed SARS-CoV-2 vaccine in a subset population.
- Analysis of the efficacy of SARS-CoV-2 rS with Matrix-M1 adjuvant in participants with asymptomatic COVID-19 who test positive for the disease by SARS-CoV-2 N protein serology but have no accompanying symptoms.
- First occurrence of virologically confirmed (by PCR to SARS-CoV-2), symptomatic mild, moderate, or severe COVID-19, with onset at least 7 days after second study vaccination (e.g., Day 28) in the initial set of vaccinations in seronegative adult participants by age (<65 and ≥65), in racial and ethnic minorities, and in those with co-morbid conditions.
- The occurrence, severity, and relationship to study vaccination of unsolicited AEs (in all adult participants) for 21 days after first study vaccination and 28 days after second study vaccination following the initial set of vaccinations with the adjustment to remove reactogenicity events that were recorded as unsolicited AEs within 7 days of each dose in the initial set of vaccinations.

**STUDY DESIGN:**

This is a Phase 3, multi-centre, randomised, observer-blind, placebo-controlled study evaluating the efficacy, safety, and immunogenicity of SARS-CoV-2 rS with Matrix-M1 adjuvant in adult participants 18 to 84 years of age (inclusive) in the UK. Following permission by regulatory bodies and based on achieving the primary efficacy endpoint and an acceptable safety profile, participants will be revaccinated with 2 injections 21 days apart of the alternate study vaccine ("blinded crossover"). That is, initial recipients of placebo will receive active SARS-CoV-2 rS with Matrix-M1 and initial recipients of active vaccine will receive placebo. Every effort will be made to identify sites of high SARS-CoV-2 activity, and populations within these sites who are at high risk of exposure to the virus will be enrolled such that the incidence of COVID-19 in the study will be higher than 1% and the sample size and enrolment period may be reduced.

After signing informed consent, participants may be screened within a window of up to approximately 30 days. During the screening period, nose/throat samples may be taken to detect SARS-CoV-2 by PCR, if the participant has any COVID-19 symptoms or significant exposure history.

Approximately 15,000 male and female adult participants 18 to 84 years of age (inclusive) with and without relevant comorbidities are planned for the study. An effort will be made to enrol a target of at least 25% of participants who are ≥ 65 years of age as well as prioritising other groups that are most affected by COVID-19, including racial and ethnic minorities. This will be under the assumption that the annual incidence of symptomatic COVID-19 will be approximately 1% to 5%.

The sample size may be adjusted by the Sponsor during the study, based on blinded data, to ensure sufficient power at the time of the primary analysis. Details on the possible sample-size reassessment will be described in the statistical analysis plan (SAP).

On Day 0 and Crossover Day 0, nose/throat samples will be taken to detect PCR for SARS-CoV-2 and blood samples will be taken to detect any existing SARS-CoV-2 antibodies at baseline. Participants will be randomised in a 1:1 ratio via block randomisation to receive 2 intramuscular (IM) injections of the SARS-CoV-2 rS with Matrix-M1 adjuvant or placebo, as described in Table S1-1.

**Table S1-1 Study Vaccine Groups**

| Study Vaccine Groups | Number of Randomised Participants | 2 Vaccinations | |
|---|---|---|---|
| | | Day 0 | Day 21 (+ 7 days) |
| SARS-CoV-2 rS (5 µg) + Matrix-M1 adjuvant (50 µg) | N = 7,500 | X | X |
| Placebo | N = 7,500 | X | X |

Randomisation will be stratified by site and by age ≥ 65 years. In addition, there will not be specific age sub-stratification recruitment targets within the ≥ 65-year age group, but through site recruitment; the aim is to allow a generally representative age distribution overall.

The study will consist of the screening period (Days -30 to 0); initial vaccination days (Days 0 and 21 [+7-day window]); and outpatient study visits on Day 0, Day 21 (+ 7 days), and Day 35 (14 days minimum after second study vaccination [+ 7 days]) after last study vaccination. Visits for the purpose of obtaining lab tests prior to unblinding may occur if feasible. Additional study visits for blood draws and blinded crossover injections will occur after achieving the primary efficacy endpoint and an acceptable safety profile.

The duration of individual participation, including screening, will be a maximum of 1 year (Day 386 ± 15 days) from the initial set of vaccinations. This will be an event-driven study for the assessment of vaccine efficacy (VE) and will end when a sufficient number of events have been observed, yet all participants will be followed for the entire study duration for an assessment of duration of vaccine efficacy (placebo-controlled and actively controlled) and for safety endpoints.

A licensed seasonal influenza co-administration sub-study will be conducted in the first approximately 400 participants who meet the additional inclusion criteria for this study. Participants may be enrolled at selected study sites due to the availability of seasonal influenza vaccine. After being randomised to receive intramuscular (IM) injections of SARS-CoV-2 rS with Matrix-M1 adjuvant or placebo, sub-study participants will receive a licensed seasonal influenza vaccine on Day 0 in the opposite deltoid. These participants will be part of the solicited AE safety subset analysis.

Given that this study and others will enrol during the influenza virus season and that the highest at-risk population, namely the elderly and adults with comorbid conditions for influenza mirrors that of SARS-CoV-2, a co-vaccination sub-study is included. This sub-group will undergo the same safety and efficacy evaluations as the entire study population. In addition, the entire sub-study will have reactogenicity assessed. This group must not have had a current season licensed influenza vaccine and have no prior history of allergy or severe reaction to seasonal influenza vaccine. To assess the possible impact of the study vaccine on the immunogenicity of the influenza vaccine, this group will also have a hemagglutination inhibition assay (HAI) performed.

A global Safety Monitoring Committee (SMC) was convened to oversee the safety of the ongoing Phase 1/2 study (Protocol 2019nCoV-101) and will oversee one or more additional studies across the SARS-CoV-2 rS vaccine program. A separate SMC will be convened for this study. The designated SMC will monitor the safety of participants in the study and will follow an SMC charter. The SMC will review blinded and unblinded safety and reactogenicity data. The SMC will also review sufficient clinical event data to ensure detection of any evidence of enhanced disease. The SMC will convene to perform safety reviews on a scheduled basis, for immediate concerns regarding safety observations during this study, and as needed.

Following achievement of the primary efficacy endpoint, blinded participants will be revaccinated with 2 injections 21 days apart of the alternate study vaccine (“blinded crossover”). That is, initial recipients of placebo will receive active SARS-CoV-2 rS with Matrix-M1 and initial recipients of active vaccine will receive placebo. Because the blinded crossover will provide all study participants with active SARS-CoV-2 rS vaccine, either initially or at the time of blinded crossover, unblinding of participants to allow receipt of another active vaccine is discouraged after the crossover. Yet all participants will be reminded that they always have the option to become unblinded for a deployed vaccine or withdraw from the study at any time for any reason. Previously unblinded participants still in the study will not be eligible for crossover vaccinations. Participants who are unblinded and receive another active vaccine outside of this protocol in this manner will be censored in the final analysis at the time of unblinding but will be strongly encouraged to remain in safety follow-up as defined in this protocol.

**Study Vaccination Pause Rules:**

Study vaccination pause rules based on reactogenicity, AEs, and SAEs related to study participation will be in place to monitor participant safety for the initial set of vaccinations only.

AEs meeting any one of the following criteria will result in a hold being placed on subsequent study vaccinations pending further review by the SMC at the direction of the SMC chair:

- Any toxicity grade 3 or higher (severe or potentially life-threatening) solicited (local or systemic) single AE term occurring in ≥ 10% of participants (after a minimum of 100 subjects are enroled) in the SARS-CoV-2 rS with Matrix-M1 adjuvant group within the first 7 days after study vaccination.
- Any severe unsolicited single AE preferred term for which the investigator assesses as related that occurs in ≥ 5% of participants (after a minimum of 100 subjects are enroled) in the SARS-CoV-2 rS with Matrix-M1 adjuvant group, within the first 7 days after study vaccination.

In addition, any SAE assessed as related to vaccine (final assessment by the Sponsor) will be reported by the Sponsor to the SMC Chair as soon as possible, and within 24 hours of the sponsor's awareness of the event. Based on this initial report of the event to the SMC Chair, the Chair may advise the Sponsor to immediately pause enrolment and further dosing in either some or all participants in the study and to convene an ad hoc meeting, or make alternative recommendations. The SMC Charter defines processes for how this review will occur and how the Chair's recommendations will be documented.

The sponsor, along with medical monitor, may request an SMC review for any safety concerns that may arise in the study, even if they are not associated with any specific pause rule; for example, any SAE for which causality is at least possibly related.

The 400 subject influenza vaccine co-administration study will utilize the same pause rules with one difference due to the expectation of greater reactogenicity and the small size:

- Any grade 3 or higher (severe or potentially life-threatening) solicited (local or systemic) single AE term occurring in ≥ 20% of participants in the SARS-CoV-2 rS with Matrix-M1 adjuvant plus seasonal influenza vaccine group, within the first 7 days after study vaccination.

**STUDY POPULATION:**

**Inclusion Criteria:**

Each participant must meet all of the following criteria to be enrolled in this study:

1. Adult males or females aged 18 to 84 years (inclusive) at screening.
2. Able and willing (in the investigator's opinion) to comply with all study requirements.
3. Willing to allow the investigators to discuss the volunteer's medical history with their General Practitioner and access all medical records when relevant to study procedures.
4. Willing and able to give informed consent prior to study enrolment.
5. Female participants of childbearing potential (defined as any female who has experienced menarche and who is NOT surgically sterile [i.e., hysterectomy, bilateral tubal ligation, or bilateral oophorectomy] or postmenopausal [defined as amenorrhea at least 12 consecutive months or > 1 documented plasma follicle-stimulating hormone level ≥ 40 mIU/mL]) must agree to be heterosexually inactive from at least 28 days prior to enrolment and through 3 months after the last study vaccination OR agree to consistently use any of the following methods of contraception from at least 28 days prior to enrolment and through 3 months after the last study vaccination:
    a. Condoms (male or female)
    b. Diaphragm with spermicide
    c. Cervical cap with spermicide
    d. Intrauterine device
    e. Oral or patch contraceptives
    f. Norplant®, Depo-Provera®, or other in country regulatory-approved contraceptive method that is designed to protect against pregnancy
    g. Abstinence, as a form of contraception, is acceptable if in line with the participant's lifestyle (other approaches to abstinence are not acceptable)

       **NOTE**: Periodic abstinence (e.g., calendar, ovulation, symptothermal, post-ovulation methods) and withdrawal are not acceptable methods of contraception.
6. Room air oxygen saturation > 95% at Screening/Day 0.

**Seasonal Influenza Vaccine Co-Administration Sub-Study Only**

7. Participant should not have received a current season influenza vaccine, have no contraindication to the specific vaccine to be administered in the study, and no prior history of allergy or severe reaction to seasonal influenza vaccines.

**Exclusion Criteria:**

Participants meeting any of the following criteria will be excluded from the study:

1. Participation in COVID-19 prophylactic drug trials for the duration of the study.
2. Future participation in SARS-CoV-2 serological surveys where participants are informed of their serostatus for the duration of the study.
3. Participation in research involving an investigational product (drug/biologic/device) within 45 days prior to first study vaccination.
4. History of laboratory-confirmed (by PCR or serology to SARS-CoV-2) COVID-19 infection at any time prior to randomisation.
5. Administration of immunoglobulins and/or any blood products within the 3 months preceding the planned administration of the study vaccine candidate.
6. Any confirmed or suspected immunosuppressive or immunodeficient state; chronic administration (defined as more than 14 continuous days) of immunosuppressant medication within the past 3 months, except topical steroids or short-term oral steroids (course lasting ≤ 14 days).

   **NOTE:** An immunosuppressant dose of glucocorticoid is defined as a systemic dose ≥ 10 mg of prednisone per day or equivalent. The use of topical, inhaled, and nasal glucocorticoids will be permitted if other chronic disease conditions are not exclusionary. Human immunodeficiency syndrome (HIV)-positive participants receiving highly active antiretroviral therapy and a history within 6 months of screening of viral load < 1000 copies/mL or CD4 count > 300 cells/$mm^3$ would be eligible.
7. History of allergic disease or reactions likely to be exacerbated by any component of the study vaccines.
8. Any history of anaphylaxis to any prior vaccine.
9. Pregnancy, lactation, or willingness/intention to become pregnant within 3 months following the last study vaccination.
10. Current diagnosis of or treatment for cancer (except basal cell carcinoma of the skin and cervical carcinoma in situ, at the discretion of the investigator).
11. Bleeding disorder (e.g., factor deficiency, coagulopathy or platelet disorder), or prior history of significant bleeding or bruising following IM injections or venepuncture.
12. Continuous use of anticoagulants, such as coumarins and related anticoagulants (i.e., warfarin) or novel oral anticoagulants/anti-platelet agents.

    **NOTE:** The use of ≤ 325 mg of aspirin per day as prophylaxis is permitted.
13. Suspected or known current alcohol or drug dependency.
14. Study team member or first-degree relative of any study team member (inclusive of Sponsor, contract research organisation (CRO), and site personnel involved in the study).
15. Participants who are having any current workup of undiagnosed illness within the last 8 weeks that is either participant-reported or has been clinician-assessed, which could lead to a new condition or diagnosis.

16. Received any live vaccine within 4 weeks or **any vaccine** (excluding influenza) within 2 weeks prior to first study vaccination or any licensed influenza vaccine within 1 week prior to first study vaccination or plans to receive any vaccine from these time periods until 28 days after second study vaccination.

    **NOTE:** An influenza co-administration sub-study will occur in which 400 participants will receive a single dose of seasonal influenza vaccine at the same time as first study vaccination. In addition, a licensed seasonal influenza vaccine may be given 7 days after each vaccination but should not be given within 7 days prior to second vaccination.

17. Have clinically significant chronic cardiovascular, endocrine, gastrointestinal, hepatic (including hepatitis B and C), renal, neurological, respiratory, psychiatric or other medical disorders not excluded by other exclusion criteria, that are assessed by the investigator as being clinically unstable within the prior 4 weeks as evidenced by:
    a. Hospitalisation for the condition, including day surgical interventions.
    b. New significant organ function deterioration.
    c. Needing addition of new treatments or major dose adjustments of current treatments (mild or moderate well-controlled comorbidities are allowed).

18. History of chronic neurological disorders that have required prior specialist physician review for diagnosis and management (such as multiple sclerosis, dementia, transient ischemic attacks, Parkinson’s disease, degenerative neurological conditions, and neuropathy) or a history of stroke or previous neurological disorder within 12 months with residual symptoms. Participants with a history of migraine or chronic headaches or nerve root compression that have been stable on treatment for the last 4 weeks are not excluded.

19. Any autoimmune disease/condition (iatrogenic or congenital) listed in Table 9-3 or being treated with a biologic therapy.

    **NOTE:** The Skin and Metabolic Disorders listed in Table 9-3 are eligible at the discretion of the investigator.

20. Any other significant disease, disorder or finding that, in the opinion of the investigator, may significantly increase the risk to the volunteer because of participation in the study, affect the ability of the volunteer to participate in the study, or impair interpretation of the study data.

21. Participant requires the use of continuous oxygen therapy or any oxygen therapy while awake or is anticipated to require daytime oxygen therapy during the course of the study.

NOTE: Nocturnal oxygen use only is acceptable for study inclusion.

NOTE: Inclusion and exclusion criteria are applied at study entry and should not be used to determine whether to provide a second dose. Decision making regarding refraining from a second dose should be based on specific conditions such as pregnancy or anaphylaxis to the prior study dose or medical contraindications per the judgment of the investigator or medical monitor.

**Other Considerations:**

Participants meeting any of the following criteria may have planned study vaccination deferred for a later date, but these criteria are not exclusionary for study enrolment.

- Respiratory symptoms in the past 3 days (i.e., cough, sore throat, difficulty breathing). Participant may be vaccinated once all symptoms have been resolved for > 3 days. Out-of-window study vaccination is allowed for this reason (**see NOTE**).
- Temperature of > 38°C within 24 hours of planned study vaccination (site measured or participant measured). Participant may be vaccinated once the fever has resolved and there has not been any temperature measured as being > 38°C for > 3 days. Out-of-window study vaccination is allowed for this reason (**see NOTE**).

  **NOTE:** PCR testing for SARS-CoV-2 is likely to be indicated for either of the above reasons or if COVID-19 is suspected based on other symptoms, potential exposure to SARS-CoV-2 infection through either close contacts or based on local epidemiology. If a participant has a positive PCR for SARS-CoV-2 prior to enrolment that participant will not be eligible and will be considered a screen failure.

- Participants having any symptoms or signs of possible COVID-19 infection (Table 2-2) that may also be due to post-vaccination reactogenicity within 7 days of either study vaccine dose will NOT be required to be tested for SARS-CoV-2 PCR.
- Any participant with a new positive PCR-confirmed SARS-CoV-2 infection occurring from Screening and prior to second study vaccination will not be removed from the study but must meet health requirements before receiving second study vaccination.
- Any participant who has a positive PCR for SARS-CoV-2 between the first and second study vaccination should also have PCR testing for SARS-CoV-2 on the day of the subsequent study vaccination, but the results of the PCR test are not needed before study vaccination can be given.
- Any participant with a new positive PCR-confirmed SARS-CoV-2 infection occurring from Screening and prior to the end of immunogenicity assessments will be removed from applicable immunogenicity analyses as defined in the SAP.
- Any acute illness (cardiovascular, endocrine, gastrointestinal, hepatic, renal, neurological, respiratory, or other medical disorders) that is actively causing symptoms that could, in the opinion of the investigator, impact the assessment of reactogenicity or other study assessments may have planned study vaccination deferred for a later date, but these criteria are not exclusionary for study enrolment. Participant may be vaccinated once symptoms have resolved or are stabilised for > 3 days. Out-of-window study vaccination is allowed for this reason.
- Any participant who is otherwise eligible with a blood pressure of ≥ 160/100 mmHg may be retested onsite several times over a 3-hour interval to achieve a lower blood pressure. If the blood pressure remains ≥ 160/100 mmHg, study vaccination should be deferred for a later date if the baseline blood pressure is found to be < 160/100 mmHg.

**STUDY VACCINES:**

Study vaccinations (5-µg SARS-CoV-2 rS with 50-µg Matrix-M1 adjuvant or placebo [saline]) will comprise 2 IM injections on Days 0 and 21, ideally in alternating deltoids. For blinding purposes, all participants will be vaccinated using the same injection volume (i.e., 0.5 mL).

At the time of the blinded crossover, participants will receive the alternate study material in 2 IM injections 21 + 7 days apart.

The seasonal influenza vaccine co-administration sub-study will comprise a single IM injection (0.5 mL) of a licensed influenza vaccine on Day 0, ideally in the opposite deltoid of the study vaccine. Flucelvax Quadrivalent will be given to those 18 to 64 years of age, and an adjuvanted trivalent influenza vaccine will be given to those ≥ 65 years of age.

Whenever possible the RIGHT deltoid will be used for the influenza vaccine and the LEFT deltoid for the study vaccine.

All study vaccinations will be administered on an outpatient basis by designated site personnel in a way to maintain the blind. The influenza vaccine will not require blinding. Any pharmacy preparation with unblinded product will require unblinded site personnel who will not otherwise be involved in the study procedures or observation of participants.

**STUDY PROCEDURES:**

Study procedures, including efficacy, immunogenicity, and safety assessments are listed in the schedule of events (SOE) in Table 3-1.

**Efficacy Assessments:**

**Nose/Throat Testing for SARS-CoV-2 Detection and Confirmation:**

Nose/throat samples for virus detection will be taken at the study visits described in the SOE (Table 3-1).

**Monitoring for COVID-19:**

Identification and laboratory confirmation of SARS-CoV-2 infection and symptomatic COVID-19 will be performed throughout the study.

On study enrolment, participants will be given details of a 24/7 on-call number to contact the study team. Participants will be instructed to contact the study team within 24 hours via this number if they self-assess COVID-19 symptoms in Table 2-2 or are clinically concerned. Newly discovered symptoms of suspected COVID-19 will trigger a COVID-19 Surveillance Visit. Please refer to Table 2-2 for the qualifying symptoms of suspected COVID-19 disease. Participants will receive weekly reminders (email or text messages) to immediately contact the study team if they have experienced any new symptoms or health concerns that could be related to infection with SARS-CoV-2. Newly discovered symptoms of suspected COVID-19 will trigger a COVID-19 Surveillance Visit. If the participant is known to be COVID-19 positive at the time of scheduling for the Initial Surveillance Visit and cannot attend the visit due to extenuating circumstances (e.g., local government restrictions), then a phone call or phone calls can be substituted to assess the participant’s status. However, the participant should make every effort to attend the Follow-up Surveillance Visit (as per protocol timeline) when government restrictions or other circumstances allow. If a participant has made an Initial Surveillance phone contact or visit and is found to be COVID-19 negative on all swabs taken for this episode, then a phone call can be substituted for the Follow-up Surveillance Visit. A participant with suspected or confirmed COVID-19 will be asked to complete the COVID-19 symptom diary. Participants should start the diary on their first day of symptoms and complete this diary daily for a minimum of 10 days even if their symptoms have resolved and regardless of any SARS-CoV-2 PCR results. They should continue the diary beyond 10 days if they have any ongoing symptoms related to the episode and should continue to do so until all symptoms have resolved.

The occurrence of COVID-19-related hospitalisation and COVID-19-related complications (such as, but not limited to, pneumonia, neurological or vascular complications, severe pneumonia, severe neurological or vascular events, acute respiratory distress syndrome, renal complications, sepsis, septic shock, death) will be monitored throughout the study. Every episode of a new onset of symptoms of suspected COVID-19 will trigger a COVID-19 Surveillance Visit (Initial and Follow-Up) with exceptions per protocol. A "new onset" episode will require at least a 7-day period of being symptom-free prior to the event to differentiate a specific episode from any prior illness.

**Immunogenicity Assessments:**

Blood will be collected from all participants for humoral immunogenicity according to the time points specified in the SOE (Table 3-1).

Immune measurements (ELISA) will be conducted on serum (immunoglobulin G [IgG]) for SARS-CoV-2 anti-S protein serology in approximately 900 participants in the Anti-S Protein Serology Subset (after both sets of vaccinations) and on anti-N protein serology in all participants. The immunologic test for SARS-CoV-2 serology based on a non-S protein (e.g., SARS-CoV-2 N protein) will be performed to identify cases of asymptomatic infection. Additional immunogenicity assessments specific to SARS-CoV-2 (or related variants) will include a neutralising antibody assay, which will be performed in approximately 900 participants in a Neutralisation Assay Subset. Cell-mediated immune responses, as assessed by ELISpot ± intracellular cytokine staining, will be performed in approximately 450 participants in the Cell-mediated Assay Subset. In addition, an HAI will be performed in the approximately 400 participants in the seasonal influenza vaccine co-administration sub-study.

Anti-S protein serology in the Anti-S Protein Serology Subset will be collected after the initial and second set of vaccinations.

**Safety Assessments:**

The timing and frequency of all safety assessments are listed in the SOE (Table 3-1).

After each study vaccination, including the licensed seasonal influenza vaccine for participants in the seasonal influenza vaccine co-administration sub-study, participants will remain under observation at the study site for at least 30 minutes for the presence of any acute reactions and solicited AEs. Solicited AEs, collected through an electronic diary (e-Diary), will be recorded from the time of study vaccination until 7 days after study vaccination in all participants in the seasonal influenza vaccine co-administration sub-study and a subgroup of all other participants after the initial set of vaccinations. As solicited AEs are only being recorded in a subset of participants, it is expected that typical solicited AEs will be captured as unsolicited AEs in those participants who are not in the sub-study. An assessment of the impact of this decision will be part of an exploratory analysis. Participants in the licensed seasonal influenza vaccine co-administration sub-study will record local reactogenicity for the study vaccine injection site only.

All participants will be assessed for unsolicited AEs from the time of first study vaccination until Day 49 after the initial set of vaccinations; SAEs will be assessed from the time the informed consent is signed until completion of the participant's last study-related procedure; all MAAEs will be reported from the time of first study vaccination in the initial set of vaccinations until Day 35; MAAEs related to study vaccination and AESIs will be reported from the time of first study vaccination until completion of the participant's last study-related procedure. All AEs will be followed until resolution or until clinically stable.

Separate analyses of safety events received with respect to approved or deployed SARS-CoV-2 vaccines will be performed.

Safety assessments will include monitoring and recording of solicited (local and systemic reactogenicity events) and unsolicited AEs; MAAEs; AESI; SAEs; vital sign measurements on day of study vaccinations; and physical examination findings. Local and systemic reactogenicity events will not be recorded for the second set of vaccinations. COVID-19 severity will be categorised as mild, moderate, or severe according to protocol-specified criteria (Table 2-1). Recording of solicited and unsolicited AEs may be conducted by electronic data capture (EDC)/reporting. Potential immune-mediated medical conditions (PIMMC) and AESIs specific to potential disease enhancement for COVID-19 will also be monitored (see Section 9.5 [Appendix 4] for details).

**STATISTICAL ANALYSIS PLANS:**
**Sample Size:**

This study is designed to enrol approximately 15,000 participants, who will be initially randomised 1:1 into the 2 study vaccine groups. The sample size is driven by the total number of events expected to achieve statistical significance for the primary efficacy endpoint – a target of 100 mild, moderate, or severe COVID-19 cases. The target number of events of 100 was chosen to provide > 95% power for 70% or higher vaccine efficacy (VE). A single interim analysis of efficacy will be conducted based on the accumulation of approximately 50% (50 events) of the total anticipated primary endpoints using Pocock boundary conditions. Power calculations were performed using 10,000 simulated trials that were created under various assumptions of VEs and analysed using methods described in the "efficacy analysis" section without covariates. A 90% evaluability rate for the per-protocol (PP) efficacy (PP-EFF) population (assuming 10% unevaluable due to attrition and/or baseline-seropositive participants) was used for the power calculations. All simulations were performed in SAS V9.4.

**Analysis Sets:**

The all-randomised set will include all participants who were randomised regardless of the receipt of any study vaccine (SARS-CoV-2 rS with Matrix-M1 adjuvant or placebo). The all-randomised set will be used for the subject disposition summaries.

The intent-to-treat (ITT) analysis set will include all participants who are randomised and receive at least 1 dose study vaccine (SARS-CoV-2 rS with Matrix-M1 adjuvant or placebo), regardless of protocol violations or missing data. The ITT analysis set will be used as a supportive analysis population for the immunogenicity and efficacy analyses and will be analysed according to the study vaccine group as randomised.

The safety analysis set will include all participants who receive at least 1 dose of study vaccine (SARS-CoV-2 rS with Matrix-M1 adjuvant or placebo). Participants in the safety analysis set will be analysed according to the study vaccine actually received.

The PP-EFF will include baseline seronegative participants who receive both doses of study vaccine (SARS-CoV-2 rS with Matrix-M1 adjuvant or placebo) and have no major protocol deviations that occur before the first COVID-19 episode affecting the primary efficacy outcome (i.e., participants will be censored at the time of the protocol deviation) as assessed by the Sponsor prior to unblinding. All analyses of the PP-EFF population will also exclude any illness episodes with positive SARS-CoV-2 by any validated PCR or antibody test occurring 6 days or less after the second study vaccination (e.g., Day 28).

The per-protocol immunogenicity (PP-IMM) analysis set for each post-randomisation visit will include participants who receive scheduled dose(s) of study vaccine (SARS-CoV-2 rS with Matrix-M1 adjuvant or placebo), have at least a baseline and the serum sample result for the visit available after study vaccination, and have no major protocol violations that are considered clinically likely to impact the immunogenicity response at the corresponding study visit as assessed by the sponsor prior to unblinding. For each visit, the SARS-CoV-2 unexposed population will also exclude any illness episodes with positive SARS-CoV-2 by any validated PCR or antibody test prior to each visit. Prior exposed participants will be determined using baseline SARS-CoV-2 immunity defined as positive SARS-CoV-2 by PCR or antibody test at baseline, or positive SARS-CoV-2 by qualitative PCR through Day 35, according to the specified analysis. Analysis will be performed to assess if immune responses differ between exposed and unexposed individuals (i.e., whether prior exposure alters dosing regimen considerations in a pandemic response).

The review and determination for exclusion from the PP populations will be carried out in a blinded fashion by a study clinician prior to unblinding for each interim analysis based on all available information from the locked database. Both PP populations will be analysed according to the study vaccine group as randomised.

**Efficacy Analyses:**

The primary endpoint will be analysed on the PP-EFF Analysis Set and supported by analysis of the ITT Analysis Set. Conclusions concerning declaration of attainment of the primary endpoint will only be based on the PP-EFF population.

Primary analysis of the primary and key secondary efficacy endpoints will be performed based on the data generated prior to the blinded crossover. The analysis of data generated after the blinded crossover or the combined analyses of both pre- and post-blinded crossover will be performed using the approach described by Follmann et al (2020). Additional details on the analytical approach will be described in the Statistical Analysis Plan.

VE is defined as VE (%) = (1 – RR) × 100, where RR = relative risk of incidence rates between the 2 study vaccine groups (SARS-CoV-2 rS with Matrix-M1 adjuvant or placebo). The interim and final analyses for the primary objective in the PP-EFF population will be carried out at the overall one-sided Type I error rate of 0.025. The nominal alpha to be spent for the final analysis will be recalculated using the Lan-DeMets alpha spending function based on the actual numbers of events used for the interim analysis and the numbers of endpoints to be used for the final analysis. The estimated RR and its CI will be derived using Poisson regression with robust error variance [Zou 2004]. The explanatory variables in the model will include study vaccine group. The dependent variable will be the incidence rate of the endpoint of interest. The robust error variances will be estimated using the repeated statement and the participant identifier. The Poisson distribution will be used with a logarithm link function. The site (depending on the distribution of endpoints) and the age strata will be included in the model as covariates. To assess incidence rates rather than absolute counts of cases, accounting for differences in follow-up times starting with 7 days after the second vaccination among participants, an offset will be utilized in the Poisson regression. In case the total number of events to be analysed may be too low for an asymptotic method proposed (i.e., ≤ 5 events in either treatment group), an alternative method based on the single sample exact binomial distribution may be used for the analysis. This method is based on the proportion of the events in the SARS-CoV-2 rS with Matrix-M1 adjuvant group among the total number of events observed in both treatment groups after adjusting for the differential number of subjects (or inclusive of differential lengths of follow-up) between the 2 treatment groups.

Hypothesis testing of the primary efficacy endpoint will be carried out against H0: VE ≤ 30%. Rejection of the null hypothesis, H0: VE ≤ 30% demonstrates a statistically significant vaccine effect (i.e., alpha-adjusted lower bound confidence interval [LBCI] > 30%) will be considered meeting the pre-specified study success criterion. The study will also continue for the intended duration to measure efficacy, immunogenicity, and safety endpoints, regardless of primary endpoint success at the interim or final analysis. The final analysis of the primary efficacy endpoint will be triggered when approximately 100 PP-EFF participants with symptomatic mild, moderate, or severe COVID-19 endpoints have accrued. Also, in order to be able to respond to the unexpected and rapidly evolving COVID-19 pandemic situation globally, other factors such as requests by government or public health agencies may also be factored into the decision-making to unblind the study for the final analysis, but this always occurs in consultation with lead regulatory agencies.

The secondary and exploratory efficacy endpoints will be analysed using the same method as the primary efficacy analysis described above. Analysis of secondary and exploratory efficacy endpoints will be performed without adjustment for multiple comparisons (i.e., two-sided alpha of 0.05). The final interpretation of the overall vaccine efficacy will be based on the totality of statistical evidence, including immunogenicity results and the clinical importance in discussions with the regulatory agencies and scientific communities.

The EOS analysis will be performed when the last participant completes the last visit (12 months after the last study vaccination) or discontinues earlier.

**Immunogenicity Analyses:**

The primary and secondary immunogenicity analyses will be performed using the PP-IMM and ITT analysis populations.

For the SARS-CoV-2 rS serum antibody levels measured by microneutralization and ELISA assays, the geometric mean at each study visit, geometric mean fold rises (GMFRs) comparing to Day 0 (baseline) at each follow-up study visit, along with 95% CI will be summarised by study vaccine group. The 95% CI will be calculated based on the t distribution of the log-transformed values for geometric means or GMFRs, then back transformed to the original scale for presentation. The seroconversion rate (SCR), proportion of participants with ≥ 4 fold rises if naïve at baseline, along with 95% CIs based on the Clopper-Pearson method will be summarised by study vaccine group at each follow-up study visit. Immunogenicity analyses performed after the second set of vaccinations will be conducted in a similar fashion.

For the subset of participants who receives the influenza vaccine concurrently with the study vaccines, comparisons of strain-specific immune responses to influenza vaccine as measured by HAI will be performed. The treatment comparison will be made by comparing the strain-specific geometric mean titres (GMTs) and the SCRs. The SCR is defined as the proportion of subjects with either a baseline reciprocal (Day 0) titre of < 10 and a post-vaccination reciprocal titre ≥ 40, or a baseline titre of ≥ 10 and a post-vaccination titre ≥ 4-fold higher.

For influenza strain-specific GMTs in each treatment group, titres reported below the lower limit of quantitation (LLOQ; i.e., below the starting dilution of assay reported as "< 10") will be set to half that limit (i.e., 10 / 2 = 5). Strain-specific GMTs will be summarized by treatment group and visit day along with the corresponding two-sided 95% CIs, by exponentiating the corresponding log-transformed means and their 95% CIs. The ratio of GMTs between treatment groups post-vaccination and the corresponding two-sided 95% CI will be calculated on log-transformed titres using the analysis of covariance (ANCOVA) with treatment group and baseline (Day 0) measurement as the covariate.

For influenza strain-specific SCRs, the rate in percent and the corresponding two-sided exact binomial 95% CIs will be calculated using the Clopper-Pearson method. The two-sided 95% CIs for the absolute rate difference between the 2 treatment groups will be constructed using the Newcombe method.

Similar summaries will be generated for the other immunogenicity endpoints and other assays if conducted.

Correlates of risk will be assessed where immune responses are measured in all available participants, and the results compared between participants who experience a SARS-CoV-2 event and participants who do not. The goal of this analysis is to assess correlates of risk of SARS-CoV-2 infection (and potential other secondary endpoints) in the study vaccine group by comparing study vaccine-induced immune responses associated with COVID-19.

**Safety Analyses:**

Solicited AEs (e.g., local and general systemic reactogenicity symptoms) will be captured in the Safety Subset of approximately 2,000 participants and in the seasonal influenza vaccine co-administration sub-study of approximately 400 participants after the initial set of vaccinations only.

Numbers and percentages (with 95% CIs based on the Clopper-Pearson method) of participants with solicited local and systemic AEs through 7 days after each study vaccination will be summarised by study vaccine group and the maximum toxicity grade over 7 days after each study vaccination. The duration of solicited local and systemic AEs after each study vaccination will also be summarised by treatment group.

Unsolicited AEs will be coded by preferred term and system organ class using the latest version of the Medical Dictionary for Regulatory Activities and summarised by study vaccine group as well as by severity and relationship to study vaccine. AEs through 49 days after first study vaccination for the initial set of vaccinations; all MAAEs through 35 days after first study vaccination for the initial set of vaccinations; and MAAEs related to study vaccine; SAEs; or AESIs through EOS will be listed separately and summarised by study vaccine group. As solicited AEs are only being recorded in a subset of participants, it is expected that typical solicited AEs will be captured as unsolicited AEs in those participants who are not in the sub-study if they occur through Day 49 after the initial set of vaccinations. An assessment of the impact of this decision will be part of an exploratory analysis and will be summarised.

Separate analyses of safety events received with respect to approved or deployed SARS-CoV-2 vaccines will be performed.

Vital sign measurements will be summarised by study vaccine group using descriptive statistics at baseline and following study vaccination.

Concomitant medications will be provided in a data listing with preferred drug name as coded using the World Health Organisation (WHO) drug dictionary.

**Interim Analyses**

Prior to the final analysis, a single interim analysis of efficacy will be conducted based on the accumulation of approximately 50% (50 events) of the total target number of the primary endpoint (100 events). For this analysis, the data needed to perform the analysis of the primary efficacy endpoint will be cleaned. The interim analysis will be performed by an unblinded Biostatistics and Programming team (PPD), and the unblinded statistician will communicate the results of the analysis to the Sponsor in terms of fulfillment or nonfulfillment of the pre-defined success criterion (yes/no). Novavax will be unblinded at the participant level at the time of the primary 100-event analysis. If the pre-defined success criterion of the interim analysis is unfulfilled (no), then the study will remain blinded to treatment assignment until the final analysis.

If the pre-defined success criterion of the interim analysis is fulfilled (yes), then the Sponsor may unblind selected accrued data at the treatment group level and continue the study while maintaining the blind to achieve a more robust safety and efficacy data package. The unblinded Biostatistics and Programming team PPD will be isolated (by firewall) from study personnel. They will complete a review independent of the study team and Sponsor. The interim analysis will follow standard group-sequential design using the Lan-DeMets alpha-spending function for Pocock boundary conditions. Table 7-2 summarizes the timing, number of endpoints, and statistical success boundaries at the planned interim and final analyses.

**Pre Blinded Crossover**

Prior to the blinded crossover, an assessment of safety and efficacy will be made while there is a placebo-controlled comparator.

**Post Blinded Crossover**

Following blinded crossover, follow-up to assess safety and efficacy endpoints (assessment of MAAEs related to study vaccine, SAEs, and AESIs, blood testing for SARS-CoV-2 and vaccine efficacy) and will continue through study completion.

**Planned Analyses Prior to Study Completion:**

Analysis of the data will be provided on an ongoing basis for confirming success and to review safety as the study progresses. The statistical analysis plan (SAP) will outline the sequential nature of these reviews.

### 1. INTRODUCTION

#### 1.1 Background

Coronaviruses are medium sized, enveloped, positive-stranded ribonucleic acid (RNA) viruses, with a characteristic crown-like appearance in electron micrographs due to circumferential studding of the viral envelope with projections comprising the spike (S) protein. There are 4 different strains (229E, OC43, NL63, and HKU1), which are ubiquitous in humans and generally result in mild upper respiratory illnesses and other common cold symptoms including malaise, headache, nasal discharge, sore throat, fever, and cough [Su 2016]. In addition, other coronavirus strains are widespread in animals, where they typically cause enteric disease. These zoonotic coronaviruses have been known to evolve into strains that can infect humans with serious consequences including severe acute respiratory syndrome coronavirus (SARS-CoV) from 2002 to 2003, Middle Eastern Respiratory Syndrome (MERS)-CoV since 2012, and most recently, the novel SARS-CoV-2 since 2019 [Habibzadeh 2020].

In late December of 2019, an outbreak of respiratory disease caused by novel coronavirus (2019 nCoV) was detected in Wuhan, Hubei province, China. The virus' rapidly discerned genetic relationship with the 2002-2003 SARS-CoV has resulted in adoption of the name "SARS-CoV-2," with the disease being referred to as coronavirus disease 2019 (COVID-19). Despite containment efforts since the start of the outbreak, the SARS-CoV-2 has spread rapidly with over 214 countries/territories/areas outside of China reporting laboratory confirmed COVID-19 cases as of 15 May 2020 [WHO 2020]. On 30 January 2020, the International Health Regulations Emergency Committee of the World Health Organisation (WHO) designated the outbreak as a public health emergency of international concern (PHEIC) and subsequently declared a pandemic on 11 March 2020.

In December of 2020, the first SARS-CoV-2 vaccine was authorised for conditional use by the Medicines and Healthcare products Regulatory Authority (MHRA). Vaccination programs throughout the United Kingdom (UK) began on 08 December 2020, which resulted in questions concerning ongoing clinical trials for other SARS-CoV-2 vaccines. The following week the National Institute for Health Research (NIHR), with input from the MHRA and Health Research Authority [HRA]), released guidance to vaccine sponsors on conducting vaccine trials with a deployed vaccine in the UK. It is anticipated that additional SARS-CoV-2 vaccines will be authorised for conditional use or approved during the course of this study with increasing availability over time. This protocol has been amended to accommodate this change in the COVID-19 landscape.

Novavax, Inc. is developing a recombinant vaccine adjuvanted with the saponin-based Matrix-M1™ adjuvant for the prevention of disease caused by SARS-CoV-2. SARS-CoV-2 recombinant (r) spike (S) protein nanoparticle vaccine (SARS-CoV-2 rS) is constructed from the full-length, wild-type SARS-CoV-2 S glycoprotein (GP) based upon the GenBank gene sequence MN908947, nucleotides 21563-25384, from the 2019 SARS-CoV-2 genome. The S protein is a type 1 trimeric glycoprotein of 1,273 amino acids that is produced as an inactive S0 precursor. The S-gene was codon optimised for expression in *Spodoptera frugiperda* (Sf9) insect cells. The SARS-CoV-2 rS nanoparticle vaccine is intended for administration with Matrix-M1 adjuvant, which is a saponin-based adjuvant that has previously been shown to enhance the immunogenicity of other nanoparticle vaccines in nonclinical and clinical studies.

#### 1.2 Nonclinical Experience

In support of the development of SARS-CoV-2 rS, Novavax has obtained nonclinical pharmacology data concerning several SARS-CoV-2 S protein variants, toxicity data concerning SARS-CoV-2 rS with Matrix-M1 adjuvant, and prior toxicity data concerning other viral glycoproteins manufactured in the baculovirus-Sf9 system and formulated with Matrix-M1 adjuvant.

##### 1.2.1 Nonclinical Data from SARS-CoV-2 Spike Protein Constructs that Support SARS-CoV-2 rS Development

Mouse immunogenicity studies were conducted to evaluate several SARS-CoV-2 S protein variants and to select the vaccine candidate [Tian 2020]. The selected vaccine candidate, BV2373 (3Q-2P), was demonstrated to be immunogenic and elicited functional antibodies. For the tested constructs, shallow dose responses with Matrix-M1 adjuvant were observed, suggesting that the adjuvant may be significantly antigen-sparing in large animals and humans.

The candidate SARS-CoV-2 rS vaccine, based on the BV2373 construct, has been evaluated in dose-titration studies in [redacted] cynomolgus macaques, and baboons. [redacted] In cynomolgus macaques, 2 dose regimens of 5 or 25 µg SARS-CoV-2 rS/25 or 50 µg Matrix-Ml adjuvant were also highly immunogenic, resulting in high anti-S IgG levels, high hACE2 binding inhibition titres, and high neutralising antibody responses. The 5 and 25 µg antigen doses gave generally similar responses when administered twice with 50 µg of Matrix-Ml adjuvant. In baboons, which may be more predictive of responses in humans, 5 and 25 µg SARS-CoV-2 rS/50 µg Matrix-Ml adjuvant induced high levels of anti-S IgG, hACE2-binding inhibiting antibodies, and neutralising antibodies. Matrix-Ml adjuvant provided antigen-sparing, and supported induction of functional antibodies. Importantly, Matrix-Ml adjuvanted SARS-CoV-2 rS also appeared to induce strong T helper 1 (Thl) type $CD4^+$ T-cell responses to SARS-CoV-2 S protein that included polyfunctional effector phenotypes. Current data in this small baboon study confirms that doses of 5 µg and 25 µg with 50 µg Matrix-M1 are the correct doses to test clinically, with Matrix-M1 adjuvant appearing critical for maximum responses.

Virus challenge studies were performed in mice [redacted] and cynomolgus macaques. In 2 mouse challenge models, immunisation with 1 or 2 doses of SARS-CoV-2 rS/Matrix-M1 adjuvant suppressed viral replication, reduced lung inflammation, and reduced systemic morbidity (weight loss) after SARS-CoV-2 live virus challenge and were not associated with any obvious exacerbation of the inflammatory response to the virus or worsening of clinical outcomes. [redacted] In cynomolgus macaques, administered with human doses of 5 or 25 µg SARS-CoV-2 rS adjuvanted with 50 µg Matrix-M1, high and comparable levels of anti-S IgG titres and hACE2 receptor binding inhibition titres were detected 21 days after the first immunisation. All of the macaques immunised with any dose or regimen of SARS-CoV-2 rS/Matrix-M1 adjuvant were protected against live virus challenge as evidenced by the reduction of total viral RNA and subgenomic RNA to below the limit of quantitation in bronchoalveolar lavages and nasal swabs.

[redacted]

##### 1.2.2 Nonclinical Data from Other Baculovirus-Sf9-Produced Nanoparticle Vaccines that Support SARS-CoV-2 rS Development

The immunogenicity and protective efficacy of 2002-2003 SARS-CoV S protein and chimeric influenza/SARS-CoV virus-like particle (VLP) vaccines produced in the baculovirus-Sf9 system and administered with and without aluminum hydroxide adjuvants was demonstrated in a mouse challenge study [Liu 2011]. Robust neutralising antibody titres were observed following vaccination, although both antigens required adsorption to aluminum hydroxide for optimal responses. The immunogenicity and protective efficacy of a MERS-CoV S nanoparticle vaccine with and without Matrix-M1 adjuvant was demonstrated in a mouse challenge study [Coleman 2017]. Following vaccination, the MERS-CoV S nanoparticle was immunogenic across all active treatment groups; however, the presence of Matrix-M1 adjuvant induced a 3- to > 10-fold enhancement of the binding and neutralising antibody responses. In addition, Matrix-M1 adjuvant essentially eliminated the antigen dose-response, suggesting the potential for major antigen-sparing and consequent improved manufacturing efficiency and timeliness [Coleman 2017]. The Matrix-M1 adjuvant was also shown to enhance antibody, cellular, and protective immune responses in Balb/c mice administered Zaire ebolavirus (EBOV) GP vaccine with or without Matrix-M1 or aluminum phosphate adjuvants [Bengtsson 2016].

In addition, 3 GLP-compliant toxicology studies in NZW rabbits have been performed with 4 different antigens (influenza hemagglutinin [HA] ± respiratory syncytial virus [RSV] F, Zika virus envelope dimers [ZIKV EnvD], and EBOV GP), in which up to 100 µg Matrix-M1 adjuvant alone or with antigen was evaluated. These toxicological investigations indicated that baculovirus-Sf9-produced antigens (up to 240 µg total nanoparticle dose) with Matrix-M1 adjuvant (up to 100 µg) were well tolerated in the animals tested with no evidence of toxicity suggestive of any unusual risk or target organ for toxicity. Non-adverse findings, including local injection site inflammation, enlargement of the lymph nodes draining the injection sites, and elevated serum markers of inflammation (including C-reactive protein), were transient and were considered consistent with immune system stimulation consequent to immunisation.

Further details are provided in the SARS-CoV-2 rS Investigator Brochure (IB).

#### 1.3 Clinical Experience

The first clinical study with SARS-CoV-2 rS nanoparticle vaccine is 2019nCoV-101, which is a 2-part, randomised, observer-blinded, placebo-controlled, Phase 1/2 trial. Part 1 (Phase 1) is designed to evaluate the immunogenicity and safety of SARS-CoV-2 rS nanoparticle vaccine with or without Matrix-M1 adjuvant in 131 healthy participants ≥ 18 to ≤ 59 years of age. Results of an interim analysis at Day 35 showed that SARS-CoV-2 rS with Matrix-M1 adjuvant was well tolerated and elicited robust immune responses [Keech 2020]. There were no serious adverse events (SAEs) or adverse events of special interest (AESIs). Reactogenicity was mainly mild in severity and of short duration (mean ≤ 2 days), with second vaccinations inducing greater local and systemic reactogenicity. The adjuvant significantly enhanced immune responses (anti-S IgG, hACE2 receptor binding inhibition antibody, and neutralising antibody) and was antigen dose-sparing, and the 2 dose 5µg SARS-CoV-2 rS/Matrix-M1 adjuvant induced mean anti-S IgG and neutralising antibody responses that exceeded the mean responses in convalescent sera from COVID-19 patients with clinically significant illnesses. The vaccine also induced antigen-specific T cells with a largely Th1 phenotype.

Part 2 (Phase 2) is designed to evaluate the immunogenicity, safety, and preliminary efficacy of SARS-CoV-2 rS and Matrix-M1 adjuvant in up to 1,500 healthy adults ≥ 18 to ≤ 84 years of age with more co-morbidities than the participant population in Part 1 of the study. An interim 5-day reactogenicity analysis was conducted on 846 participants following the first dose of study vaccine to support initiation of the Phase 3 study. This analysis comprised 607 participants aged 18 to 59 years (the same age range of Part 1 of the study) and

239 participants aged 60 to 84 years, with data presented in masked groups to maintain the integrity of the study. Overall, local and systemic reactogenicity data from this analysis were consistent with the reactogenicity data in Part 1 of the study, with no safety concerns between the younger and older age cohorts. Both local and systemic reactogenicity events occurred less frequently in older adults.

Novavax has, in its internally sponsored clinical trials, tested baculovirus-Sf9-produced nanoparticle vaccines in 14,848 participants comprising older adults, young adults, and a limited number of children 2 to 5 years of age; and also including 3,075 pregnant women, with acceptable safety. Matrix-M adjuvant has been given to 4,311 humans (of which, approximately 2,657 humans received nanoparticle vaccine) with acceptable short-term reactogenicity, and an unremarkable long-term safety profile. Additionally, interim results from the current study have indicated that the SARS-CoV-2 rS and Matrix-M1 adjuvant vaccine met the primary endpoint for the study and achieved an acceptable safety profile.

Further details on the clinical experience of the study vaccine can be found in the SARS-CoV-2 rS IB.

#### 1.4 Rationale for Study

Both nonclinical and early clinical data to date have supported clinical development of SARS-CoV-2 rS and Matrix-M1 adjuvant as a potential vaccine against SARS-CoV-2. In rodent and nonhuman primate (NHP) challenge models, SARS-CoV-2 rS and Matrix-M1 adjuvant induced high titres of antibodies measured against anti-S protein and hACE2 receptor binding and achieved neutralisation of wild-type virus that exceeded the magnitude of responses measured in COVID-19 human convalescent sera and provided protection against SARS-CoV-2 challenge [Tian 2020; Mandolesi 2020; Guebre-Xabier 2020]. Notably in NHP studies, clinical doses of vaccine (5- and 25-µg SARS-CoV-2 rS/50-µg Matrix-M1) resulted in sterile immunity in the lungs and nasal passage following wild-type virus challenge, suggesting that the vaccine may both protect against upper and lower respiratory tract disease and interrupt transmission [Guebre-Xabier 2020].

Results from a Day 35 interim analysis of Part 1 (Phase 1) of Study 2019nCoV-101 indicate that in healthy adult participants 18 to 59 years of age two-dose regimens of 5- and 25-µg SARS-CoV-2 rS/50 µg Matrix-M1 (on Days 0 and 21) were well tolerated and induced the most robust immune responses with high levels of neutralising antibodies that closely correlated with anti-spike IgG [Keech 2020]. Furthermore, neutralising antibody responses following second vaccination were of the magnitude seen in convalescent serum from symptomatic COVID-19 patients and exceeded overall convalescent sera geometric mean titres (GMTs) by four-fold. The benefit of Matrix-M1 adjuvant was clear in the magnitude of the antibody and T-cell response, induction of functional antibodies, and dose sparing.

A Phase 2 clinical program is underway and will provide safety and immunogenicity results in older participants (> 60 years of age) and participants with comorbidities. Reactogenicity data following the first dose indicate that the reactogenicity profile between adults 18 to 59 years and older adults ≥ 60 years are comparable, with older adults generally reporting solicited events less frequently. Combining the current nonclinical and clinical data with positive Phase 1/2 data provide the impetus for early initiation of the Phase 3 clinical development program in the context of the current public health pandemic crisis.

The purpose of this study is to evaluate the efficacy, safety, and immunogenicity of SARS-CoV-2 rS with Matrix-M1 adjuvant in adults 18-84 years of age (inclusive). The study will be conducted at anticipated high COVID-19 transmission areas in the United Kingdom (UK). The information provided in this study will inform progression of the study vaccine, to determine efficacy of the study vaccine to prevent COVID-19 in the general population, in participants regardless of serostatus, in participants who have required medical intervention, and in participants with mild or asymptomatic infections. The study will determine the safety of the study vaccine to use in the general population and to ensure that it elicits a robust immune response.

#### 1.5 Rationale for Dose Selection

As previously described, clinical doses of vaccine and adjuvant (5- and 25-µg SARS-CoV-2 rS/50-µg Matrix-M1) resulted in sterile immunity in the lungs and nasal passage following wild-type virus challenge in NHP, suggesting that the vaccine may both protect against upper and lower respiratory tract disease and interrupt transmission [Guebre-Xabier 2020]. These doses are being evaluated in Part 1 of Study 2019nCoV-101 in 131 healthy adult participants ≥ 18 to ≤ 59 years of age and in Part 2 of Study 2019nCoV-101 in up to 1,500 participants ≥ 18 to ≤ 84 years of age, including participants with comorbidities. Results from the Part 1 Day 35 interim analysis support either dose of SARS-CoV-2 rS/Matrix-M1 in terms of safety and immunology, with the lower dose (5 µg) offering advantages in regards to dose sparing. All study vaccinations will be administered on an outpatient basis by designated site personnel in a way to maintain the blind. Any pharmacy preparation with unblinded product will require unblinded site personnel who will not otherwise be involved in the study procedures or observation of participants.

#### 1.6 Benefit-Risk Assessment

The SARS-CoV-2 rS nanoparticle vaccine contains purified protein antigens. It cannot replicate, nor can it cause COVID-19. However, in common with all vaccines produced in cell culture or other systems, the SARS-CoV-2 rS nanoparticle vaccine contains residual non-vaccine proteins derived from the production system, and sensitisation to these, or the SARS-CoV-2 S protein itself, may theoretically occur. While the occurrence of immediate hypersensitivity is possible with the administration of any vaccine, whether licensed or in development, no such reactions have been observed in any of these clinical trials to date. As clinical data become available with increased exposure, it is possible that this profile may change.

The risk for enhanced COVID-19 in immunised participants is a theoretical risk. Enhanced disease in coronavirus vaccine-immunised animals after live virus challenge has been demonstrated in nonclinical studies of several, but not all, coronavirus vaccine candidates. There is currently no evidence for immunoenhancement in nonclinical testing of SARS-CoV-2 rS or other Novavax baculovirus-Sf9-based vaccines taken into nonclinical evaluation or clinical trials.

No risks have been identified in nonclinical or early clinical testing of SARS-CoV-2 or other coronavirus vaccines (SARS-CoV and MERS-CoV) developed using the baculovirus-Sf9 system to date. In supportive toxicology studies with other viral GP nanoparticle vaccines developed using the baculovirus-Sf9 system with different antigens, findings were generally consistent with an immune response to the vaccine formulations. These toxicological investigations indicated that baculovirus-Sf9-produced antigens (up to 240 µg total nanoparticle dose) with Matrix-M1 adjuvant (up to 100 µg) were well tolerated in the animal and antigen system tested with no evidence of toxicity suggestive of any unusual risk or target organ for toxicity. Non-adverse findings, including local injection site inflammation and serum chemical markers of inflammation (such as C-reactive protein), were transient and considered consistent with immune system stimulation consequent to immunisation.

Findings to date suggest that SARS-CoV-2 rS when administered with or without Matrix-M1 adjuvant can be reasonably expected to demonstrate an acceptable safety profile in healthy adult participants aged ≤ 59 years. Novavax baculovirus-Sf9-produced nanoparticle vaccines comprising viral glycoproteins, with and without Matrix-M1 or aluminum adjuvants, have been shown to induce robust and protective immune responses in relevant animal models to influenza HAs, RSV F protein, SARS-CoV and MERS-CoV S proteins, rabies GP, and EBOV GP. In addition, the Novavax SARS-CoV-2 candidate adsorbed to aluminum phosphate has induced antibodies in pregnant women which, when transferred transplacentally, were associated with reduced rates of SARS-CoV-2 lower respiratory tract infections in their infants during the first 90 to 180 days of life. The goal of this program is to investigate the efficacy, safety, and immunogenicity of the SARS-CoV-2 rS and Matrix-M1 adjuvant.

Further details are provided in the SARS-CoV-2 rS IB.

### 2. STUDY OBJECTIVES AND ENDPOINTS

#### 2.1 Study Objectives

##### 2.1.1 Primary Objective

- To demonstrate the efficacy of SARS-CoV-2 rS with Matrix-M1 adjuvant in the prevention of virologically confirmed (by polymerase chain reaction [PCR] to SARS-CoV-2), symptomatic COVID-19, when given as a 2-dose vaccination regimen, as compared to placebo, in serologically negative (to SARS-CoV-2) adults.

##### 2.1.2 Secondary Objectives

- To demonstrate the efficacy of SARS-CoV-2 rS with Matrix-M1 adjuvant in the prevention of virologically confirmed (by PCR to SARS-CoV-2), symptomatic COVID-19, when given as a 2-dose vaccination regimen, as compared to placebo, in adults regardless of their serostatus at baseline.
- To assess the efficacy of SARS-CoV-2 rS with Matrix-M1 adjuvant in adult participants requiring specific medical interventions as compared to placebo.
- To assess the efficacy of SARS-CoV-2 rS with Matrix-M1 adjuvant on mild COVID-19 symptoms as compared to placebo.
- To assess the efficacy of SARS-CoV-2 rS with Matrix-M1 adjuvant on occurrence of asymptomatic or undetected infections with SARS-CoV-2 as compared to placebo.
- In a subset of adult participants, to evaluate the immunogenicity of SARS-CoV-2 rS with Matrix-M1 adjuvant as compared to placebo.
- To evaluate safety in terms of SAEs and medically attended adverse events (MAAEs) related to study vaccination in all adult participants during the entire study period.
- To evaluate safety in terms of AESIs, which encompasses potential immune-mediated medical conditions (PIMMCs) and AESIs relevant to COVID-19, including possible vaccine-enhanced disease, in all adult participants at any time after the first dose.
- In a subset of adult participants, to evaluate the safety and reactogenicity in terms of solicited local and systemic adverse events (AEs) after the initial set of study vaccinations.
- To evaluate safety in terms of all MAAEs for 14 days after second vaccination and unsolicited AEs for 21 days after first study vaccination and 28 days after second study vaccination in the initial set of vaccinations.
- To assess the duration of vaccine efficacy (measured by all efficacy endpoints) in initial active vaccine recipients vs. crossover (delayed) active vaccine recipients.

##### 2.1.3 Exploratory Objectives

- In a subset of adult participants, to evaluate the safety and immunogenicity of SARS-CoV-2 rS with Matrix-M1 adjuvant in the initial set of vaccinations when co-administered with a licensed seasonal influenza vaccine.

- In a subset of adult participants unblinded before the crossover, to explore the efficacy and safety of SARS-CoV-2 rS with Matrix-M1 adjuvant in the initial set of vaccinations and/or an approved or deployed SARS-CoV-2 vaccine.

#### 2.2 Study Endpoints

##### 2.2.1 Primary Endpoint

- First occurrence of virologically confirmed (by PCR to SARS-CoV-2), symptomatic mild, moderate, or severe COVID-19 (Table 2-1) with onset at least 7 days after second study vaccination (e.g., Day 28) in the initial set of vaccinations in serologically negative (to SARS-CoV-2) adult participants at baseline until the endpoint-driven efficacy analysis is triggered by the occurrence of a prespecified number of blinded endpoints.

**Table 2-1: Endpoint Definitions of COVID-19 Severity**

| COVID-19 Severity | Endpoint Definitions |
|---|---|
| **Mild** | ≥ 1 of:<br>• Fever (defined by subjective or objective measure, regardless of use of anti-pyretic medications)<br>• New onset cough<br>• ≥ 2 COVID-19 respiratory/non-respiratory symptoms in Table 2-2<br>**AND**<br>• Does not meet criteria for moderate or severe disease |
| **Moderate** | ≥ 1 of:<br>• Fever (defined by subjective or objective measure, regardless of use of anti-pyretic medications) + any 2 COVID-19 symptoms in Table 2-2 for ≥ 3 days (need not be contiguous days)<br>• High fever (≥ 38.4°C) for ≥ 3 days (need not be contiguous days)<br>• Any evidence of significant LRTI:<br>– Shortness of breath (or breathlessness or difficulty breathing) with or without exertion (greater than baseline)<br>– Tachypnea: 20 to 29 breaths per minute at rest*<br>– SpO2: 94% to 95% on room air*<br>– Abnormal chest x-ray or chest CT consistent with pneumonia or LRTI<br>– Adventitious sounds on lung auscultation (e.g., crackles/rales, wheeze, rhonchi, pleural rub, stridor)<br>**AND**<br>• Does not meet criteria for severe disease |

**Table 2-1: Endpoint Definitions of COVID-19 Severity**

| COVID-19 Severity | Endpoint Definitions |
|---|---|
| **Severe** | ≥ 1 of:<br>• Tachypnea: ≥ 30 breaths per minute at rest*<br>• Resting heart rate ≥ 125 beats per minute*<br>• $SpO_2$: ≤ 93% on room air or $PAO_2/FiO_2$ < 300*<br>• High flow oxygen therapy or NIV/NIPPV (e.g., CPAP or BiPAP)<br>• Mechanical ventilation or ECMO<br>• One or more major organ system dysfunction or failure (e.g., cardiac/circulatory, pulmonary, renal, hepatic, and/or neurological, to be defined by diagnostic testing/clinical syndrome/interventions), including any of the following:<br>o ARDS<br>o Acute renal failure<br>o Acute hepatic failure<br>o Acute right or left heart failure<br>o Septic or cardiogenic shock (with shock defined as SBP < 90 mm Hg OR DBP < 60 mm Hg<br>o Acute stroke (ischemic or hemorrhagic)<br>o Acute thrombotic event: AMI, DVT, PE<br>o Requirement for: vasopressors, systemic corticosteroids, or hemodialysis.<br>• Admission to an ICU<br>• Death |

Abbreviations: AMI = acute myocardial infarction; ARDS = acute respiratory distress syndrome; BiPAP = bi-level positive airway pressure; CPAP = continuous positive air pressure; CT = computed tomography; DBP = diastolic blood pressure; DVT = deep vein thrombosis; ECMO = extracorporeal membrane oxygenation; $FiO_2$ = fraction of inspired oxygen; ICU = intensive care unit; LRTI = lower respiratory tract infection; NIV = non-invasive ventilation; NIPPV = non-invasive positive pressure ventilation; $PAO_2$ = partial pressure of oxygen in the alveolus; PE = pulmonary embolism; SBP = systolic blood pressure; $SpO_2$ = oxygen saturation.

*Participants with a single vital sign abnormality placing them in the moderate or severe categories must also meet the criteria for mild COVID-19.

**Table 2-2: Qualifying Symptoms of Suspected COVID-19**

| |
|---|
| • Fever<br>• New onset cough<br>• New onset or worsening of shortness of breath or difficulty breathing compared to baseline<br>• New onset fatigue<br>• New onset generalised muscle or body aches<br>• New onset headache<br>• New loss of taste or smell<br>• Acute onset of sore throat, congestion, and runny nose<br>• New onset nausea, vomiting, or diarrhea |

Abbreviations: COVID-19 = coronavirus disease 2019.

##### 2.2.2 Secondary Endpoints

###### 2.2.2.1 Key Secondary Endpoint

- First occurrence of virologically confirmed (by PCR to SARS-CoV-2), symptomatic moderate or severe COVID-19 with onset at least 7 days after second study vaccination (e.g., Day 28) in the initial set of vaccinations in serologically negative (to SARS-CoV-2) adult participants at baseline until the endpoint-driven efficacy analysis is triggered by the occurrence of a prespecified number of blinded endpoints.

###### 2.2.2.2 Other Secondary Endpoints

- First occurrence of virologically confirmed (by PCR to SARS-CoV-2), symptomatic severe COVID-19 with onset at least 7 days after second study vaccination (e.g., Day 28) in the initial set of vaccinations in serologically negative (to SARS-CoV-2) adult participants at baseline until the endpoint-driven efficacy analysis is triggered by the occurrence of a prespecified number of blinded endpoints.
- First occurrence of virologically confirmed (by PCR to SARS-CoV-2), symptomatic mild, moderate, or severe COVID-19, with onset at least 7 days after second study vaccination (e.g., Day 28) in the initial set of vaccinations in adult participants regardless of their serostatus at baseline.
- First occurrence of COVID-19 requiring hospitalisation, intensive care unit (ICU) admission or mechanical ventilation linked to any virologically confirmed (by PCR to SARS-CoV-2) COVID-19 with onset at least 7 days after second study vaccination (e.g., Day 28) in the initial set of vaccinations in adult participants regardless of their serostatus at baseline.
- First occurrence of virologically confirmed (by PCR to SARS-CoV-2), symptomatic mild COVID-19 (with no progression to moderate or severe COVID-19 during the course of the COVID-19 episode) with onset at least 7 days after second study vaccination (e.g., Day 28) in the initial set of vaccinations in adult participants, regardless of their serostatus at baseline.
- First occurrence of laboratory-confirmed (by PCR or nucleocapsid [N] protein serology to SARS-CoV-2) symptomatic or asymptomatic COVID-19 with onset at least 7 days after second study vaccination (e.g., Day 28) in the initial set of vaccinations in adult participants with negative serostatus at baseline.
- Analysis of antibodies binding to the SARS-CoV-2 spike (S) protein by enzyme-linked immunosorbent assay (ELISA) at Day 0 (baseline) and Day 35 (14 days after second study vaccination).
- Analysis of antibodies binding to the SARS-CoV-2 S protein by ELISA at Crossover Day 0 visit (baseline) and Crossover Day 35 visit (14 days after second study vaccination).

- The occurrence and relationship to study vaccination of SAEs and MAAEs related to study vaccination (in all adult participants) during the entire study period.
- The occurrence and relationship to study vaccination of AESIs and PIMMCs (in all adult participants) during the entire study period.
- The occurrence and severity of reactogenicity in terms of solicited local and systemic AEs (in all sub-study participants) after the initial set of vaccinations.
- The occurrence, severity, and relationship to study vaccination of all MAAEs for 14 days after second vaccination and unsolicited AEs (in all adult participants) for 21 days after first study vaccination and 28 days after second study vaccination following the initial set of vaccinations.
- Relative vaccine efficacy (measured by all efficacy endpoints) in initial active vaccine recipients vs. crossover (delayed) active vaccine recipients.

##### 2.2.3 Exploratory Endpoints

- First occurrence of virologically confirmed (by PCR to SARS-CoV-2), symptomatic mild, moderate, or severe COVID-19 with onset at least 14 days after first study vaccination (e.g., Day 14) in the initial set of vaccinations in serologically negative (to SARS-CoV-2) adult participants at baseline until the endpoint-driven efficacy analysis is triggered by the occurrence of a prespecified number of blinded endpoints.
- Any occurrence of serologic conversion (by serology to SARS-CoV-2 N protein) between baseline and 1 year after last study vaccination in the initial set of vaccinations in adult participants seronegative at baseline.
- Any occurrence of serologic conversion (by serology to SARS-CoV-2 N protein) between Crossover Day 0 visit and the end of study (EOS) visit in the second set of vaccinations in adult participants seronegative at baseline.
- Analysis of the proportion of adult participants with evidence of seroconversion as demonstrated by binding antibodies to the SARS-CoV-2 S protein by ELISA at Day 35 (14 days after second study vaccination) and Crossover Day 35 visit (14 days after the Crossover Day 21 visit).
- Analysis of the cell-mediated immune responses (for example, as measured by enzyme-linked immune absorbent spot (ELISpot) ± intracellular cytokine staining) on Days 0 (baseline) and 35 (14 days after second study vaccination) in the initial set of vaccinations.
- SARS-CoV-2 neutralisation as measured by virus neutralisation assay (VNA; wild-type virus and/or pseudovirion expressing SARS-CoV-2 S protein) at Day 0 (baseline) and Day 35 (14 days after second study vaccination) in the initial set of vaccinations.

- Analysis of the safety and immunogenicity of SARS-CoV-2 rS with Matrix-M1 adjuvant when co-administered with seasonal influenza vaccine in a subset population.
- Analysis of the efficacy and safety of SARS-CoV-2 rS with Matrix-M1 adjuvant and/or an approved or deployed SARS-CoV-2 vaccine in a subset population.
- Analysis of the efficacy of SARS-CoV-2 rS with Matrix-M1 adjuvant in participants with asymptomatic COVID-19 who test positive for the disease by SARS-CoV-2 N protein serology but have no accompanying symptoms.
- First occurrence of virologically confirmed (by PCR to SARS-CoV-2), symptomatic mild, moderate, or severe COVID-19, with onset at least 7 days after second study vaccination (e.g., Day 28) in the initial set of vaccinations in seronegative adult participants by age (<65 and ≥65), in racial and ethnic minorities, and in those with comorbid conditions.
- The occurrence, severity, and relationship to study vaccination of unsolicited AEs (in all adult participants) for 21 days after first study vaccination and 28 days after second study vaccination following the initial set of vaccinations with the adjustment to remove reactogenicity events that were recorded as unsolicited AEs within 7 days of each dose in the initial set of vaccinations.

### 3. STUDY DESIGN

This is a Phase 3, multi-centre, randomised, observer-blind, placebo-controlled study evaluating the efficacy, safety, and immunogenicity of SARS-CoV-2 rS with Matrix-M1 adjuvant in adult participants 18 to 84 years of age (inclusive) in the UK. Following permission by regulatory bodies based on achieving the primary efficacy endpoint and an acceptable safety profile, participants will be revaccinated with 2 injections 21 days apart of the alternate study vaccine ("blinded crossover"). That is, initial recipients of placebo will receive active SARS-CoV-2 rS with Matrix-M1 and initial recipients of active vaccine will receive placebo. Every effort will be made to identify sites of high SARS-CoV-2 activity, and populations within these sites who are at high risk of exposure to the virus will be enrolled such that the incidence of COVID-19 in the study will be higher than 1% and the sample size and enrolment period may be reduced.

After signing informed consent, participants may be screened within a window of up to approximately 30 days. Nose/throat samples may be taken during the screening period to detect SARS-CoV-2 by PCR, if the participant has any COVID-19 symptoms or significant exposure history.

Approximately 15,000 male and female adult participants 18 to 84 years of age (inclusive) with and without relevant comorbidities are planned for the study. An effort will be made to enrol a target of at least 25% of participants who are ≥ 65 years of age as well as prioritising other groups that are most affected by COVID-19, including racial and ethnic minorities. This will be under the assumption that the annual incidence of symptomatic COVID-19 will be approximately 1% to 5%.

The sample size may be adjusted by the Sponsor during the study, based on blinded data, to ensure sufficient power at the time of the primary analysis. Details on the possible sample-size reassessment will be described in the statistical analysis plan (SAP).

On Day 0 and Crossover Day 0, nose/throat samples will be taken to detect PCR for SARS-CoV-2 and blood samples will be taken to detect any existing SARS-CoV-2 antibodies at baseline. Participants will be randomised in a 1:1 ratio via block randomisation to receive 2 intramuscular (IM) injections of the SARS-CoV-2 rS with Matrix-M1 adjuvant or placebo, as described in Figure 1a. Participants who are unblinded and ineligible for the blinded crossover will continue to follow the original study design depicted in Figure 1b.

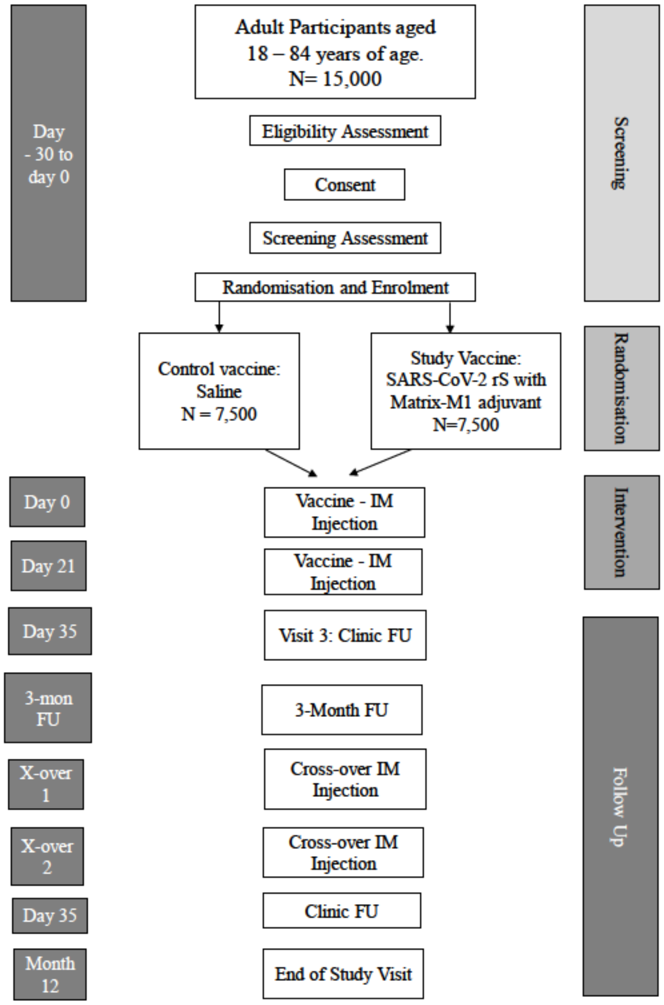

#### Figure 1a: Trial Schema with Blinded Crossover

FU = follow-up; IM = intramuscular; N = number of participants.

Note: The 3-month follow-up and Day 1 Crossover visits can be consolidated if they occur within 30 days of each other. The Day 35 Crossover visit is only for participants in the anti-S immunogenicity subgroup.

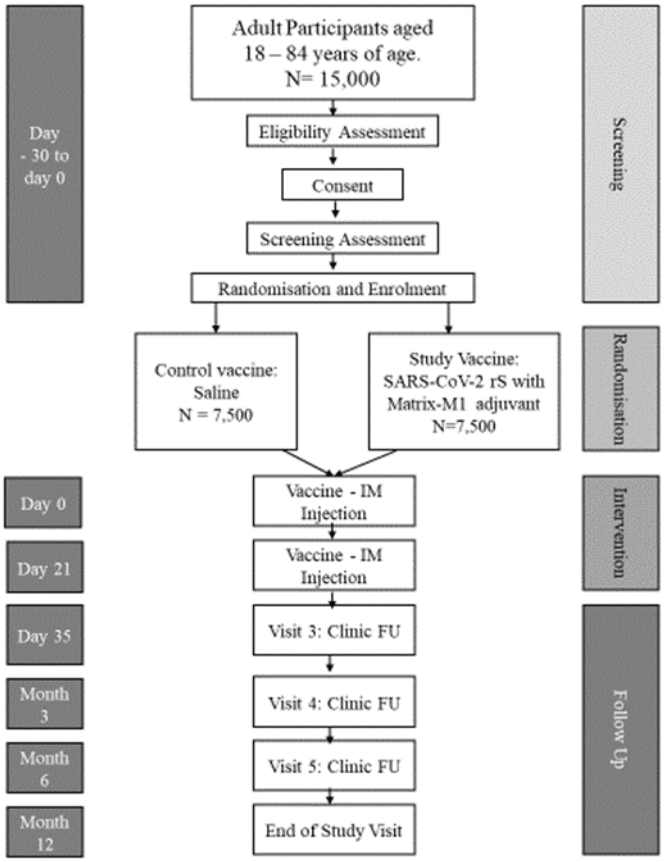

**Figure 1b: Original Trial Schema**

FU = follow-up; IM = intramuscular; N = number of participants.

Randomisation will be stratified by site and by age ≥ 65 years. In addition, there will not be specific age sub-stratification recruitment targets within the ≥ 65-year age group, but through site recruitment; the aim is to allow a generally representative age distribution overall.

The study will consist of the screening period (Days -30 to 0); initial vaccination Days 0 and 21 (+7-day window); and outpatient study visits on Day 0, Day 21 (+ 7 days), and Day 35 (14 days minimum after second study vaccination [+ 7 days]) after last study vaccination. Visits for the purpose of obtaining lab tests prior to unblinding may occur if feasible. Additional study visits for blood draws and blinded crossover injections will occur after achieving the primary efficacy endpoint and an acceptable safety profile.

The duration of individual participation, including screening, will be a maximum of 1 year (Day 386 ± 15 days) from the initial set of vaccinations. This will be an event-driven study for the assessment of vaccine efficacy (VE) and will end when a sufficient number of events have been observed, yet all participants will be followed for the entire study duration for an assessment of duration of vaccine efficacy (placebo-controlled and actively controlled) and for safety endpoints.

A licensed seasonal influenza co-administration sub-study will be conducted in the first approximately 400 participants who meet the additional inclusion criteria for this study. Participants may be enrolled at selected study sites due to the availability of seasonal influenza vaccine. After being randomised to receive IM injections of SARS-CoV-2 rS with Matrix-M1 adjuvant or placebo, sub-study participants will receive a licensed seasonal influenza vaccine on Day 0 in the opposite deltoid. These participants will be part of the solicited AE safety subset analysis.

Given that this study and others will enrol during the influenza virus season and that the highest at-risk population, namely the elderly and adults with comorbid conditions for influenza mirrors that of SARS-CoV-2, a co-vaccination sub-study is included. This sub-group will undergo the same safety and efficacy evaluations as the entire study population. In addition, the entire sub-study will have reactogenicity assessed. This group must not have had a current season licensed influenza vaccine and have no prior history of allergy or severe reaction to seasonal influenza vaccine. To assess the possible impact of the study vaccine on the immunogenicity of the influenza vaccine, this group will also have a hemagglutination inhibition assay (HAI) performed.

A global Safety Monitoring Committee (SMC) was convened to oversee the safety of the ongoing Phase 1/2 study (Protocol 2019nCoV-101) and will oversee one or more additional studies across the SARS-CoV-2 rS vaccine program. A separate SMC will be convened for this study. The designated SMC will monitor the safety of participants in the study and will follow an SMC charter. The SMC will review blinded and unblinded safety and reactogenicity data. The SMC will also review sufficient clinical event data to ensure detection of any evidence of enhanced disease. The SMC will convene to perform safety reviews on a scheduled basis, for immediate concerns regarding safety observations during this study, and as needed.

Following achievement of the primary efficacy endpoint, blinded participants will be revaccinated with 2 injections 21 days apart of the alternate study vaccine ("blinded crossover"). That is, initial recipients of placebo will receive active SARS-CoV-2 rS with Matrix-M1 and initial recipients of active vaccine will receive placebo. Because the blinded crossover will provide all study participants with active SARS-CoV-2 rS vaccine, either initially or at the time of blinded crossover, unblinding of participants to allow receipt of another active vaccine is discouraged after the crossover. Yet all participants will be reminded that they always have the option to become unblinded for a deployed vaccine or withdraw from the study at any time for any reason. Previously unblinded participants still in the study will not be eligible for crossover vaccinations. Participants who are unblinded and receive another active vaccine outside of this protocol in this manner will be censored in the final analysis at the time of unblinding but will be strongly encouraged to remain in safety follow-up as defined in this protocol.

#### 3.1 Study Vaccination Pause Rules

Study vaccination pause rules based on reactogenicity, AEs, and SAEs related to study participation will be in place to monitor participant safety after the initial set of vaccinations only.

AEs meeting any one of the following criteria will result in a hold being placed on subsequent study vaccinations pending further review by the SMC at the direction of the SMC chair:

- Any toxicity grade 3 or higher (severe or potentially life-threatening) solicited (local or systemic) single AE term occurring in ≥ 10% of participants (after a minimum of 100 subjects are enroled) in the SARS-CoV-2 rS with Matrix-M1 adjuvant group within the first 7 days after study vaccination.
- Any severe unsolicited single AE preferred term for which the investigator assesses as related that occurs in ≥ 5% of participants (after a minimum of 100 subjects are enroled) in the SARS-CoV-2 rS with Matrix-M1 adjuvant group, within the first 7 days after study vaccination.

In addition, any SAE assessed as related to vaccine (final assessment by the sponsor) will be reported by the sponsor to the SMC Chair as soon as possible, and within 24 hours of the sponsor's awareness of the event. Based on this initial report of the event to the SMC Chair, the Chair may advise the sponsor to immediately pause enrolment and further dosing in either some or all subjects in the study and to convene an ad hoc meeting or make alternative recommendations. The SMC Charter defines processes for how this review will occur and how the Chair's recommendations will be documented.

The sponsor, along with medical monitor, may request an SMC review for any safety concerns that may arise in the study, even if they are not associated with any specific pause rule; for example, any SAE for which causality is at least possibly related.

The 400 subject influenza vaccine co-administration study will utilize the same pause rules with one difference due to the expectation of greater reactogenicity and the small size:

- Any grade 3 or higher (severe or potentially life-threatening) solicited (local or systemic) single AE term occurring in ≥ 20% of participants in the SARS-CoV-2 rS with Matrix-M1 adjuvant plus seasonal influenza vaccine group, within the first 7 days after study vaccination.

#### 3.2 Schedule of Events (SOE)

Table 3-1 lists the study procedures that will be performed during the study for participants who will participate in the blinded crossover. Detailed descriptions of each visit and details for those not participating in the blinded crossover are presented in Section 6.1.

**Table 3-1 Schedule of Events**

| **Study Period:** | **Screening Period [a]** | **Clinic Visits** | | | | **Unblinding Visit[d]** | **Months After Last Study Vaccination** | **Crossover Vaccination Period[e]** | | | **Last Follow-up Visit** |
|---|---|---|---|---|---|---|---|---|---|---|---|
| **Study Day:** | **–30 to 0** | **0[a]** | **21** | **35** | **COVID-19 Surveillance Visits (Unscheduled)[c]** | | **3** | **CO0** | **CO21** | **CO35** | **12** |
| **Window (days):[b]** | – | **0** | **+ 7** | **+ 7** | | | **± 15** | **± 30** | **+7** | **+7** | **±15** |
| **Minimum days following most recent study vaccination:[b]** | – | **0** | **21** | **14** | | | – | – | **21** | **35** | -- |
| **Study Visit:** | **Screening** | **1** | **2** | **3** | | | **4** | **5** | **6** | **7** | **EOS[f]** |
| Informed consent | X | | | | | X[e] | | X | | | |
| Medical history [g] | X | | | | X | | | | | | |
| Inclusion/exclusion criteria [h] | X | X[i] | X[i] | | | | | X[h] | X[h] | | |
| Demographics [j] | X | | | | | | | | | | |
| Prior/concomitant medications [k] | X | X[i] | X[i] | X | X | X | X | X[i] | X[i] | X | X |
| Vital sign measurements[l] | X | X | X | | X | | | X | X | | |
| Urine pregnancy test (WOCBP) [m] | X | X[i] | X[i] | | | | | X[i] | X[i] | | |
| Physical examination (targeted) [n] | X | X[i] | X[i] | X | X | | | X[o] | X[o] | | |
| Nose/throat testing for SARS-CoV-2 (PCR) [p] | X | X[i] | X[i] | | X | | | X[i] | X[q] | | |
| Blood sampling for SARS-CoV-2 (ELISA for anti N-protein serology)[r] | | X[i] | | X | | X[d] | X | X[i] | | | X |
| Blood sampling for SARS-CoV-2 (ELISA for anti S-protein serology – subset of participants) [s] | | X[i] | | X | | X[d] | | X[i] | | X | |
| Blood sampling for SARS-CoV-2 neutralisation assay (subset) [t] | | X[i] | | X | | X[d] | | | | | |

**Table 3-1 Schedule of Events**

| **Study Period:** | **Screening Period [a]** | **Clinic Visits** | | | | | **Months After Last Study Vaccination** | **Crossover Vaccination Period[e]** | | | **Last Follow-up Visit** |
|---|---|---|---|---|---|---|---|---|---|---|---|
| **Study Day:** | **–30 to 0** | **0[a]** | **21** | **35** | | | **3** | **CO0** | **CO21** | **CO35** | **12** |
| **Window (days):[b]** | – | 0 | + 7 | + 7 | **COVID-19 Surveillance Visits (Unscheduled)[c]** | | ± 15 | ± 30 | +7 | +7 | ±15 |
| **Minimum days following most recent study vaccination:[b]** | – | 0 | 21 | 14 | | | – | – | 21 | 35 | -- |
| **Study Visit:** | **Screening** | **1** | **2** | **3** | | **Unblinding Visit[d]** | **4** | **5** | **6** | **7** | **EOS[f]** |
| Blood sampling for HAI (influenza co-administration subset) [u] | | X [i] | X | | | X[d] | | | | | |
| Cell-mediated assessments (subset of participants)[v] | | X [i] | | X | | | | | | | |
| Randomisation | | X | | | | | | | | | |
| Study vaccination [w] | | X | X | | | | | X | X | | |
| Reactogenicity (subset of participants) [x] | | X | X | | | | | | | | |
| Monitoring for COVID-19 [y] | | | COVID-19 case ascertainment will commence from Day 0 until EOS | | | | | | | | |
| COVID-19 Symptom Diary [z] | | | | | X | | | | | | |
| All unsolicited AEs [aa] | | X | X | X | | | | | | | |
| MAAEs [bb] | | X | X | X | X | X | X | X | X | X | X |
| SAEs [cc] | X | X | X | X | X | X | X | X | X | X | X |
| AESI [dd] | | X | X | X | X | X | X | X | X | X | X |
| EOS form [ee] | | | | | | | | | | | X |

Abbreviations: AE = adverse event; AESI = adverse event(s) of special interest; COVID-19 = coronavirus disease 2019; CO0= Crossover Day 0 visit. CO21= Crossover Day 21 visit; CO35 = Crossover Day 35 visit; EDC = electronic data capture; ELISA = enzyme-linked immunosorbent assay; ELISpot = enzyme-linked immune sorbent spot; EOS = end of study; HAI = hemagglutinin assay inhibition; HEENT = head, eye, ear, nose, and throat (exam); ID = identification; MAAE = medically attended adverse event; N = nucleocapsid; PCR = polymerase chain reaction; PIMMC = potential immune-mediated medical conditions; S = spike protein; SAE = serious adverse event; SAP = statistical analysis plan; SARS-CoV-2 = severe acute respiratory syndrome coronavirus 2; WOCBA = women of childbearing age.

[a] The Screening visit and Day 0 visit may be combined if feasible at any given study site.

[b] Days relative to study vaccination are only estimates because the window allowance is not inclusive. Should a study pause occur, visits/windows will be adjusted to allow participants to continue without protocol deviation. Visit schedules after second study vaccination are calculated relative to the day the study vaccinations were received. The precise timing of the first crossover visit is dependent on the timing of the protocol amendment and available crossover vaccination doses; however, it is expected that the blinded crossover will be implemented approximately 3-4 months after the Day 21 visit for the majority of participants.

[c] COVID-19 Surveillance Visits will occur during the initial vaccination period as well as during the crossover period. If the participant is known to be COVID-19 positive at the time of scheduling for the Initial Surveillance Visit and cannot attend the visit due to extenuating circumstances (e.g., local government restrictions), then a phone call or

**Table 3-1 Schedule of Events**

| Study Period: | Screening Period [a] | Clinic Visits | | | | | Months After Last Study Vaccination | Crossover Vaccination Period[e] | | | Last Follow-up Visit |
|---|---|---|---|---|---|---|---|---|---|---|---|
| Study Day: | –30 to 0 | 0[a] | 21 | 35 | COVID-19 Surveillance Visits (Unscheduled)[c] | Unblinding Visit[d] | 3 | CO0 | CO21 | CO35 | 12 |
| Window (days):[b] | – | 0 | + 7 | + 7 | | | ± 15 | ± 30 | +7 | +7 | ±15 |
| Minimum days following most recent study vaccination:[b] | – | 0 | 21 | 14 | | | – | – | 21 | 35 | -- |
| Study Visit: | Screening | 1 | 2 | 3 | | | 4 | 5 | 6 | 7 | EOS[f] |

phone calls can be substituted to assess the participant's status. However, the participant should make every effort to attend the Follow-up Surveillance Visit (as per protocol timeline) when government restrictions or other circumstances allow. If the participant has made an Initial Surveillance phone contact or visit and is found to be COVID-19 negative on all swabs taken for this episode, then a phone call can be substituted for the Follow-up Surveillance Visit.

d An Unscheduled Unblinding Visit should be arranged (where possible and practical to do so) for those who are offered and are willing to accept an approved or deployed SARS-CoV-2 approved or deployed vaccine. Serology will be obtained as per Section 6.1.6. Visits on Days 21 and 35 may be skipped for those who have been unblinded as per Section 6.1.6. Participants who are unblinded are not eligible for participation in the blinded crossover.

e Crossover Day 0 visit and Crossover Day 21 visit will be in lieu of the 3- and 6-Month visit unless the participant has already had a 3-Month visit; in this case, Crossover Day 0 visit should occur as soon as feasible and Crossover Day 21 visit will replace the 6-Month visit. These visits will not be 3 months apart but will be 21 +7 days apart. The Crossover Day 35 visit will only be for those in the anti-S serology subset and will occur approximately 35 days after the second dose 1. For participants who have been unblinded and are ineligible for the blinded crossover, follow the schedule for the 3- and 6-Month Visits in Section 6.1.

f EOS visit. Should participants decide to terminate early, an EOS telephone call will occur to collect the maximum safety data possible.

g Including prior and concomitant medical conditions (as appropriate), recent vaccinations (≤ 90 days), and significant surgical procedures.

h Specific exclusions to study vaccination (e.g., anaphylaxis to dose 1, pregnancy) will be assessed before any vaccination. Waivers to enrolling participants with exclusions will not be given.

i Performed prior to study vaccination (each set).

j Screening only. Including date of birth (day, month, and year), sex, race, ethnicity, weight, and height.

k Recent and current medications at the time of screening to be reviewed to ensure eligibility criteria are fulfilled. Concomitant medications include all medications (including vaccines) taken by the participant. Concomitant medications will be collected through Day 49 only unless related to an SAE, AESI, related MAAE, or is an approved or deployed COVID-19 vaccine.

l Including respiratory rate, blood pressure, pulse rate, pulse oximetry and oral temperature (or via forehead/ear reader). On all study vaccination days, vital sign measurements will be collected once before study vaccination to ensure participant has no evidence of fever prior to study vaccination. Vital sign measurements will also be collected once again, approximately 15 to 30 minutes after study vaccination, to check for any reactions to the vaccine.

m Women of childbearing potential only. A urine pregnancy test will be performed at Screening and prior to each study vaccination. A positive urine pregnancy test at any of the vaccination visits will result in the participant not receiving any further study vaccination. A positive urine pregnancy test at Screening will result in screen failure.

n Examination at screening to include height and weight; HEENT, neck, thyroid, lungs, heart, cardiovascular, abdomen, lymph nodes of the upper extremities and neck; and musculoskeletal system/extremities to allow for study vaccination; symptom-directed (targeted) physical examination at all other scheduled time points but always to include lymphatic assessment of injected upper extremity on study vaccination days. Physical examination on study vaccination visits must be done prior to vaccination. Interim physical examinations will be performed at any unscheduled visit at the discretion of the investigator, if necessary.

o Crossover Day 0 and Day 21 visits—targeted PE is optional and may be conducted if participants have specific complaints or as per investigator judgment.

**Table 3-1 Schedule of Events**

| Study Period: | Screening Period [a] | Clinic Visits | | | COVID-19 Surveillance Visits (Unscheduled)[c] | Unblinding Visit[d] | Months After Last Study Vaccination | Crossover Vaccination Period[e] | | | Last Follow-up Visit |
|---|---|---|---|---|---|---|---|---|---|---|---|
| Study Day: | –30 to 0 | 0[a] | 21 | 35 | | | 3 | CO0 | CO21 | CO35 | 12 |
| Window (days):[b] | – | 0 | + 7 | + 7 | | | ± 15 | ± 30 | +7 | +7 | ±15 |
| Minimum days following most recent study vaccination:[b] | – | 0 | 21 | 14 | | | – | – | 21 | 35 | -- |
| Study Visit: | Screening | 1 | 2 | 3 | | | 4 | 5 | 6 | 7 | EOS[f] |

[p] Samples will be collected at Screening only if the participant has any COVID-19 symptoms or significant exposure history. If a participant has a positive PCR prior to enrolment, they will be considered a screen failure. Samples will be collected on Day 0 and the method of collection will be taught. Participants found to be SARS-CoV-2 positive on Day 0 will be excluded from some analyses of the study as per the SAP. Samples may be collected on Day 21 only if the participant has any COVID-19 symptoms or significant exposure history between Days 0 and 21. Results of that test are not required for study vaccination, but participants who are symptomatic may have the second study vaccination delayed per protocol. Participants found to be SARS-CoV-2 positive on Day 21 will be excluded from some analyses of the study as per the SAP. A sample will be taken at Crossover Day 0 visit. The sample bar code should be put in EDC as soon as is feasible.

[q] PCR to be done for symptomatic cases. If a sample is taken the sample bar code should be put in EDC as soon as feasible.

[r] The ELISA for anti-S protein serology will be performed in the approximately 900 participants in the Anti-S Protein Serology Subset after each set of vaccinations. These participants will have an extra visit the Crossover Day 35 visit, which will occur approximately 35 days after the second dose 1.

[s] The neutralising antibody assay will be performed in the approximately 900 participants in the Neutralisation Assay Subset.

[t] The HAI assay will be performed in the approximately 400 participants in the licensed seasonal influenza co-administration sub-study.

[u] Cell-mediated immune responses, as assessed by ELISpot ± intracellular cytokine staining, will be performed in approximately 450 participants in the Cell-mediated Assay Subset.

[v] Study vaccination on Day 0 will consist of study vaccine plus, in the seasonal influenza vaccine co-administration sub-study, a single dose of licensed influenza vaccine. Study vaccination on Day 21 will consist of study vaccine.

[w] Solicited AEs (e.g., local and general systemic reactogenicity symptoms) will be captured in the Safety Subset of approximately 2,000 participants and in the first approximately 400 participants who qualify for the seasonal influenza vaccine co-administration sub-study after the initial set of vaccinations only. On study vaccination days, participants will remain in clinic for at least 30 minutes to be monitored for any severe reactogenicity. Severe reactions will be noted as AEs on day of study vaccination.

[x] Participants will monitor for COVID-19 symptoms. This will include provision of a Study ID Card that provides details on study participation, study site contact information and assessment of symptoms of suspected COVID-19 (see Table 2-2). Instructions to participants to contact the study site within 24 hours for symptoms of suspected COVID-19 will be given. A participant with suspected or confirmed COVID-19 will be asked to complete a COVID-19 symptom diary for 10 days. The Study ID Card should be presented to healthcare providers not affiliated with the study who encounter participants with symptoms of suspected COVID-19.

[y] Samples will be self-collected by the participants in an effort to determine if the current symptoms are due to SARS-CoV-2 infection. Approximately 24 hours after the onset of symptoms the participants will swab themselves daily for up to 3 days but only until the point that 1 of the samples tests positive for SARS-CoV-2.

[z] A participant with suspected or confirmed COVID-19 will be asked to complete the COVID-19 symptom diary. Participants should start the diary on their first day of symptoms and complete this diary daily for a minimum of 10 days even if their symptoms have resolved and regardless of any SARS-CoV-2 PCR results. They should continue the diary beyond 10 days if they have any ongoing symptoms related to the episode and should continue to do so until all symptoms have resolved.

[aa] All unsolicited AEs are to be reported from the time of first study vaccination until 21 days after first study vaccination and 28 days after second study vaccination in all participants after the initial set of vaccinations only.

**Table 3-1 Schedule of Events**

| Study Period: | Screening Period [a] | Clinic Visits | | | | Unblinding Visit[d] | Months After Last Study Vaccination | Crossover Vaccination Period[e] | | | Last Follow-up Visit |
|---|---|---|---|---|---|---|---|---|---|---|---|
| Study Day: | –30 to 0 | 0[a] | 21 | 35 | COVID-19 Surveillance Visits (Unscheduled)[c] | | 3 | CO0 | CO21 | CO35 | 12 |
| Window (days):[b] | – | 0 | + 7 | + 7 | | | ± 15 | ± 30 | +7 | +7 | ±15 |
| Minimum days following most recent study vaccination:[b] | – | 0 | 21 | 14 | | | – | – | 21 | 35 | -- |
| Study Visit: | Screening | 1 | 2 | 3 | | | 4 | 5 | 6 | 7 | EOS[f] |

[bb] MAAEs are to be collected from the time of first study vaccination until Day 35 in the initial set of vaccinations, and MAAEs assessed as related to study vaccination from the time of first study vaccination until completion of the participant's last study-related procedure.

[cc] SAEs are to be collected from the time of informed consent until completion of the participant's last study-related procedure.

[dd] AESI: To include PIMMC and AEs related to COVID-19 are to be collected from the time of first study vaccination until completion of the participant's last study-related procedure.

[ee] EOS form will be completed for all participants, including participants who are terminated early.

### 4. STUDY POPULATION

Approximately 15,000 male and female adult participants 18 to 84 years of age (inclusive) with and without relevant comorbidities will be randomised in a 1:1 ratio via block randomisation to receive 2 intramuscular (IM) injections of SARS-CoV-2 rS with Matrix-M1 adjuvant or placebo in a blinded fashion in up to 28 sites across the UK.

#### 4.1 Inclusion Criteria

Each participant must meet all of the following criteria to be enrolled in this study:

1. Adult males or females aged 18 to 84 years (inclusive) at screening.
2. Able and willing (in the investigator's opinion) to comply with all study requirements.
3. Willing to allow the investigators to discuss the volunteer's medical history with their General Practitioner and access all medical records when relevant to study procedures.
4. Willing and able to give informed consent prior to study enrolment.
5. Female participants of childbearing potential (defined as any female who has experienced menarche and who is NOT surgically sterile [i.e., hysterectomy, bilateral tubal ligation, or bilateral oophorectomy] or postmenopausal [defined as amenorrhea at least 12 consecutive months or > 1 documented plasma follicle-stimulating hormone level ≥ 40 mIU/mL]) must agree to be heterosexually inactive from at least 28 days prior to enrolment and through 3 months after the last study vaccination OR agree to consistently use any of the following methods of contraception from at least 28 days prior to enrolment and through 3 months after the last study vaccination:
   a. Condoms (male or female)
   b. Diaphragm with spermicide
   c. Cervical cap with spermicide
   d. Intrauterine device
   e. Oral or patch contraceptives
   f. Norplant®, Depo-Provera®, or other in country regulatory-approved contraceptive method that is designed to protect against pregnancy
   g. Abstinence, as a form of contraception, is acceptable if in line with the participant's lifestyle (other approaches to abstinence are not acceptable)

      **NOTE**: Periodic abstinence (e.g., calendar, ovulation, symptothermal, post-ovulation methods) and withdrawal are not acceptable methods of contraception.
6. Room air oxygen saturation > 95% at Screening/Day 0.

**Seasonal Influenza Vaccine Co-Administration Sub-Study Only**

7. Participant should not have received a current season influenza vaccine, have no contraindication to the specific vaccine to be administered in the study, and no prior history of allergy or severe reaction to seasonal influenza vaccines.

#### 4.2 Exclusion Criteria

Participants meeting any of the following criteria will be excluded from the study:

1. Participation in COVID-19 prophylactic drug trials for the duration of the study.
2. Future participation in SARS-CoV-2 serological surveys where participants are informed of their serostatus for the duration of the study.
3. Participation in research involving an investigational product (drug/biologic/device) within 45 days prior to first study vaccination.
4. History of laboratory-confirmed (by PCR or serology to SARS-CoV-2) COVID-19 infection at any time prior to randomisation.
5. Administration of immunoglobulins and/or any blood products within the 3 months preceding the planned administration of the study vaccine candidate.
6. Any confirmed or suspected immunosuppressive or immunodeficient state; chronic administration (defined as more than 14 continuous days) of immunosuppressant medication within the past 3 months, except topical steroids or short-term oral steroids (course lasting ≤ 14 days).

   **NOTE:** An immunosuppressant dose of glucocorticoid is defined as a systemic dose ≥ 10 mg of prednisone per day or equivalent. The use of topical, inhaled, and nasal glucocorticoids will be permitted if other chronic disease conditions are not exclusionary. Human immunodeficiency syndrome (HIV)-positive participants receiving highly active antiretroviral therapy and a history within 6 months of screening of viral load < 1000 copies/mL or CD4 count > 300 cells/mm$^3$ would be eligible.
7. History of allergic disease or reactions likely to be exacerbated by any component of the study vaccines.
8. Any history of anaphylaxis to any prior vaccine.
9. Pregnancy, lactation, or willingness/intention to become pregnant within 3 months following the last study vaccination.
10. Current diagnosis of or treatment for cancer (except basal cell carcinoma of the skin and cervical carcinoma in situ, at the discretion of the investigator).
11. Bleeding disorder (e.g., factor deficiency, coagulopathy or platelet disorder), or prior history of significant bleeding or bruising following IM injections or venepuncture.

12. Continuous use of anticoagulants, such as coumarins and related anticoagulants (i.e., warfarin) or novel oral anticoagulants/anti-platelet agents.

    **NOTE:** The use of ≤ 325 mg of aspirin per day as prophylaxis is permitted.

13. Suspected or known current alcohol or drug dependency.
14. Study team member or first-degree relative of any study team member (inclusive of sponsor, contract research organisation (CRO), and site personnel involved in the study).
15. Participants who are having any current workup of undiagnosed illness within the last 8 weeks that is either participant-reported or has been clinician-assessed, which could lead to a new condition or diagnosis.
16. Received any live vaccine within 4 weeks or **any vaccine** (excluding influenza) within 2 weeks prior to first study vaccination or any licensed influenza vaccine within 1 week prior to first study vaccination or plans to receive any vaccine from these time periods until 28 days after second study vaccination.

    **NOTE:** An influenza co-administration sub-study will occur in which 400 participants will receive a single dose of seasonal influenza vaccine at the same time as first study vaccination. In addition, a licensed seasonal influenza vaccine may be given 7 days after each vaccination but should not be given within 7 days prior to second vaccination.

17. Have clinically significant chronic cardiovascular, endocrine, gastrointestinal, hepatic (including hepatitis B and C), renal, neurological, respiratory, psychiatric or other medical disorders not excluded by other exclusion criteria, that are assessed by the investigator as being clinically unstable within the prior 4 weeks as evidenced by:
    a. Hospitalisation for the condition, including day surgical interventions.
    b. New significant organ function deterioration.
    c. Needing addition of new treatments or major dose adjustments of current treatments (mild or moderate well-controlled comorbidities are allowed).
18. History of chronic neurological disorders that have required prior specialist physician review for diagnosis and management (such as multiple sclerosis, dementia, transient ischemic attacks, Parkinson's disease, degenerative neurological conditions, and neuropathy) or a history of stroke or previous neurological disorder within 12 months with residual symptoms. Participants with a history of migraine or chronic headaches or nerve root compression that have been stable on treatment for the last 4 weeks are not excluded.
19. Any autoimmune disease/condition (iatrogenic or congenital) listed in Table 9-3 or being treated with a biologic therapy.

    **NOTE:** The Skin and Metabolic Disorders listed in Table 9-3 are eligible at the discretion of the investigator.

20. Any other significant disease, disorder or finding that, in the opinion of the investigator, may significantly increase the risk to the volunteer because of participation in the study, affect the ability of the volunteer to participate in the study, or impair interpretation of the study data.
21. Participant requires the use of continuous oxygen therapy or any oxygen therapy while awake or is anticipated to require daytime oxygen therapy during the course of the study.

    **NOTE**: Nocturnal oxygen use only is acceptable for study inclusion.

NOTE: Inclusion and exclusion criteria are applied at study entry and should not be used to determine whether to provide a second dose. Decision making regarding refraining from a second dose should be based on specific conditions such as pregnancy or anaphylaxis to the prior study dose or medical contraindications per the judgment of the investigator or medical monitor.

#### 4.3 Other Considerations

Participants meeting any of the following criteria may have planned study vaccination deferred for a later date, but these criteria are not exclusionary for study enrolment.

- Respiratory symptoms in the past 3 days (i.e., cough, sore throat, difficulty breathing). Participant may be vaccinated once all symptoms have been resolved for > 3 days. Out-of-window study vaccination is allowed for this reason (**see NOTE**).
- Temperature of > 38°C within 24 hours of planned study vaccination (site measured or participant measured). Participant may be vaccinated once the fever has resolved and there has not been any temperature measured as being > 38°C for > 3 days. Out-of-window study vaccination is allowed for this reason (**see NOTE**).

  **NOTE:** PCR testing for SARS-CoV-2 is likely to be indicated for either of the above reasons or if COVID-19 is suspected based on other symptoms, potential exposure to SARS-CoV-2 infection through either close contacts or based on local epidemiology. If a participant has a positive PCR for SARS-CoV-2 prior to enrolment that participant will not be eligible and will be considered a screen failure.
- Participants having any symptoms or signs of possible COVID-19 infection (Table 2-2) that may also be due to post-vaccination reactogenicity within 7 days of either study vaccine dose will NOT be required to be tested for SARS-CoV-2 PCR.
- Any participant with a new positive PCR-confirmed SARS-CoV-2 infection occurring from Screening and prior to second study vaccination will not be removed from the study but must meet health requirements before receiving the second study vaccination.
- Any participant who has a positive PCR for SARS-CoV-2 between the first and second study vaccination should also have PCR testing for SARS-CoV-2 on the day

of the subsequent study vaccination, but the results of the PCR test are not needed before study vaccination can be given.

- Any participant with a new positive PCR-confirmed SARS-CoV-2 infection occurring from Screening and prior to the end of immunogenicity assessments will be removed from applicable immunogenicity analyses as defined in the SAP.
- Any acute illness (cardiovascular, endocrine, gastrointestinal, hepatic, renal, neurological, respiratory, or other medical disorders) that is actively causing symptoms that could, in the opinion of the investigator, impact the assessment of reactogenicity or other study assessments may have planned study vaccination deferred for a later date, but these criteria are not exclusionary for study enrolment. Participant may be vaccinated once symptoms have resolved or are stabilised for > 3 days. Out-of-window study vaccination is allowed for this reason.
- Any participant who is otherwise eligible with a blood pressure of ≥ 160/100 mmHg may be retested onsite several times over a 3-hour interval to achieve a lower blood pressure. If the blood pressure remains ≥ 160/100 mmHg, study vaccination should be deferred for a later date if the baseline blood pressure is found to be < 160/100 mmHg.

#### 4.4 Withdrawal of Participants from the Study

##### 4.4.1 Reasons for Withdrawal

Participants can withdraw consent and discontinue from the study at any time, for any reason. Participants may refuse further procedures (including study vaccination) but are encouraged to remain in the study for safety follow-up. In such cases where only safety is being conducted, participant contact could be managed via telemedicine contact (e.g., telephone, web chat, video, FaceTime).

The investigator will **withhold** further study vaccination from a participant in the study if the participant:

1. Is noncompliant with the protocol.
2. Experiences an SAE or intolerable AE(s) for which study vaccination is not advised by the investigator.
3. Becomes pregnant (discontinuation of further study vaccination required).

The investigator can also withdraw a participant upon the request of the sponsor or if the sponsor terminates the study.

Vaccination with an approved or deployed SARS-CoV-2 vaccine alone will not be considered a withdrawal from the study.

##### 4.4.2 Handling of Withdrawals

Participants are free to withdraw from the study at any time upon request. Participant participation in the study may be stopped at any time at the discretion of the investigator or at the request of the sponsor.

When a participant withdraws from the study, the reason(s) for withdrawal shall be recorded by the investigator on the relevant page of the electronic case report form (eCRF). Whenever possible, any participant who withdraws from the study prematurely will undergo all end of study (EOS) assessments. Any participant who fails to return for final assessments will be contacted by the site in an attempt to have them comply with the protocol. The status of participants who fail to complete final assessments will be documented in the eCRF.

##### 4.4.3 Replacements

Participants who withdraw, are withdrawn or terminated from this study, or are lost to follow-up after signing the informed consent form (ICF) but prior to first study vaccination may be replaced. Participants who receive study vaccine and subsequently withdraw, are discontinued from further study vaccination, are terminated from the study, or are lost to follow-up will not be replaced.

### 5. TEST ARTICLES

#### 5.1 Study Vaccines Administered

Study vaccinations (SARS-CoV-2 rS with Matrix-M1 adjuvant or placebo [saline]) will comprise 2 IM injections (Day 0 and Day 21), ideally in alternating deltoids. For blinding purposes, all participants will be vaccinated using the same injection volume (i.e., 0.5 mL). The dose level will be 5 µg SARS-CoV-2 rS with 50 µg Matrix-M1 adjuvant (co-formulated in a single vial); placebo will be saline for injection. Study vaccinations will be administered on an outpatient basis by designated site personnel in a way to maintain the blind. Any pharmacy preparation with unblinded product will require unblinded site personnel who will not otherwise be involved in the study procedures or observation of participants.

At the time of the blinded crossover, participants will receive the alternate study material in 2 IM injections 21 + 7 days apart.

The seasonal influenza vaccine co-administration sub-study will comprise a single IM injection (0.5 mL) of a licensed influenza vaccine on Day 0, ideally in the opposite deltoid of the study vaccine. Seasonal influenza vaccine will be administered in an open-label manner. Flucelvax Quadrivalent will be given to those 18 to 64 years of age, and an adjuvanted trivalent influenza vaccine will be given to those ≥ 65 years of age.

Whenever possible the RIGHT deltoid will be used for the influenza vaccine and the LEFT deltoid for the study vaccine.

#### 5.2 Investigational Products

The following supplies will be used for vaccination in the study:

| Investigational Product | Supplied Formulation |
|---|---|
| SARS-CoV-2 rS with Matrix-M1 adjuvant | Solution for preparation for injection, at a concentration of 5 µg antigen and 50 µg adjuvant. |
| Placebo | Sodium chloride injection (BP, sterile), 0.9% |
| **Seasonal Influenza Vaccine Co-Administration Sub-Study** | |
| Flucelvax Quadrivalent seasonal influenza vaccine | Single-dose pre-filled syringe (0.5 mL) or multi-dose vial |
| Adjuvanted trivalent seasonal influenza vaccine | Single-dose pre-filled syringe (0.5 mL) |

Abbreviations: BP = British Pharmacopoeia; SARS-CoV-2 rS = severe acute respiratory syndrome coronavirus 2 recombinant spike protein nanoparticle vaccine.

It is anticipated that the product will be available in a co-formulated single vial.

Further details on the study vaccine can be found in the SARS-CoV-2 rS IB.

##### 5.2.1 Investigational Product Packaging and Storage

Novavax, Inc., will provide adequate quantities and appropriate labelling of SARS-CoV-2 rS with Matrix-M1 adjuvant and PPD will ensure distribution to the clinical sites from a designated depot. Sodium chloride injection (British Pharmacopoeia, sterile) and licensed seasonal influenza vaccine are commercially available and will be supplied by PPD. The clinical unit pharmacy will prepare the study vaccines for each participant. Detailed instructions for the preparation of study vaccine will be provided in a separate pharmacy manual.

All investigational products must be stored according to the labelled instructions in a secure cabinet or room with access restricted to necessary clinic personnel. The site will be required to keep a temperature log to establish a record of compliance with storage conditions.

##### 5.2.2 Investigational Product Accountability

The investigator (or delegate) will maintain accurate records of receipt of all investigational product, including dates of receipt. Accurate records will be kept regarding when and how much investigational product is dispensed and used by each participant in the study. Reasons for departure from the expected dispensing regimen must also be recorded. At the completion of the study, and to satisfy regulatory requirements regarding investigational product accountability, all investigational product will be reconciled and retained or destroyed according to applicable regulations. No investigational product will be destroyed until authorised in writing by the sponsor.

#### 5.3 Method of Assigning Participants to Study Vaccine Groups

Participants will be randomly assigned in a blinded manner using the centralised Interactive Response Technology (IRT) according to pre-generated randomisation schedules. Participants will be randomised in a 1:1 ratio via block randomisation to receive 2 IM injections of the SARS-CoV-2 rS with Matrix-M1 adjuvant or placebo. Randomisation will be stratified by site and by age ≥ 65 years. The first approximately 400 participants who qualify for the seasonal influenza vaccine co-administration sub-study (which may be at select study locations only) will be assigned prior to randomisation. These participants will be part of the solicited AE safety subset analysis. Details regarding the IRT process will be provided separately to the sites. The IRT system will be utilized to assign the crossover doses in a blinded fashion.

##### 5.3.1 Blinding Procedures

This is an observer-blinded study. To maintain the blind, placebo vaccination via IM route will be included and unblinded site personnel will manage study vaccine logistics, preparation, and administration so as to maintain the blind from the remainder of the site personnel and participants. The unblinded site personnel will not be involved in study-related assessments or have participant contact for data collection following study vaccine administration.

Unblinding of treatment assignment may occur in order to allow a participant to make an informed decision regarding receipt of an approved or deployed SARS-CoV-2 vaccine. Participants who choose to receive an approved or deployed SARS-CoV-2 vaccine as per UK government guidance will be encouraged to remain in the study for scheduled safety assessments. At the time of implementation of the blinded crossover process, a similar procedure will be employed to ensure that all study participants and personnel remain blinded as to initial and subsequent treatment assignment.

Seasonal influenza vaccine will be administered in an open-label manner.

##### 5.3.2 Breaking the Blind

A participant's study vaccine assignment will not be broken until the end of the study for the clinical site study team unless medical treatment of the participant depends on knowing the study vaccine the participant received. In the event that the blind needs to be broken because of a medical emergency or because the participant chooses to receive an approved or deployed SARS-CoV-2 vaccine, the investigator may unblind an individual participant's study vaccine allocation.

Whenever possible, the investigator should contact the medical monitor to discuss the medical emergency and the reason for revealing the actual study vaccine received by that participant. In the event that the investigator cannot contact the medical monitor in a timely manner the blind may be broken by the investigator. The medical monitor should be contacted as soon as feasible after the unblinding. The study vaccine assignment will be unblinded through IRT. Reasons for study vaccine unblinding must be clearly explained and justified in the eCRF. The date on which the code was broken together with the identity of the person responsible must also be documented.

As of December 2020, an approved SARS-CoV-2 vaccine has been deployed in the UK and regulatory agencies have issued advice concerning unblinding of treatment assignment. Those who are eligible (as per the UK government prioritisation strategy) and have been invited to receive an approved or deployed SARS-CoV-2 vaccine may request to be unblinded. Participants should only request to be unblinded if they are willing to receive an approved or deployed SARS-CoV-2 vaccine and may wish to discuss this decision with the investigator and others so as to make an informed choice. Participants who decline the approved or deployed vaccine should remain blinded to treatment assignment for the entire duration of the study or until other study continuation decisions are made in accordance with the appropriate regulatory agencies. All participants will be encouraged to remain in the study regardless of the UK government approved combination of SARS-CoV-2 vaccines received. For non-emergency unblinding, there is no requirement to seek approval or to contact the medical monitor.

An Unscheduled Visit should be arranged (where possible and practical to do so) for those who are offered and are willing to accept an approved or deployed SARS-CoV-2 vaccine where a blood sample for immunogenicity assessment may be drawn, regardless of which arm of the study they have been allocated to as outlined in Section 6.1.6.

The blind may also be broken in the event of a Suspected Unexpected Serious Adverse Reaction (SUSAR) to determine regulatory reporting.

In addition to the aforementioned situations where the blind may be broken, the data will also be unblinded to a statistical team at specified time points for planned analyses prior to study completion, as outlined in Section 7.6.

#### 5.4 Study Vaccine Compliance

All doses of the study vaccine should be administered in the clinical unit under direct observation of clinic personnel and recorded in the eCRF. Clinic personnel will confirm that the participant has received the entire dose.

The location (right or left arm), date, and timing of all doses of study vaccine will be recorded in the participants' eCRF. If a participant is not administered study vaccine, the reason for the missed dose will be recorded.

#### 5.5 Concomitant Medications and Prohibitive Therapy

##### 5.5.1 Concomitant Medications

Administration of medications, therapies, or vaccines will be recorded in the eCRF. Concomitant medications will include all medications (including vaccines) taken by the participant from the time of signing the ICF through 49 days after dose 1 in the initial and second set of vaccinations (or through the early termination visit if prior to that time). Prescription and over-the-counter (OTC) drugs, as well as herbals, vitamins, and supplements, will be included.

Concomitant medications will be collected through EOS for all related MAAEs, SAEs, or AESIs. Receipt of any approved or deployed COVID-19 vaccine should also be recorded through EOS.

Participants will be asked to record the date(s) and brand of the approved or deployed SARS-CoV-2 vaccine received.

##### 5.5.2 Prohibitive Therapy

- No live vaccine will be allowed within 4 weeks of first study vaccination until 28 days after second study vaccination (Day 49).

- No vaccine (except for a licensed seasonal influenza vaccine or an approved or deployed SARS-CoV-2 vaccine and vaccines given to participants in the seasonal influenza co-administration sub-study) will be allowed within 2 weeks of first study vaccination until 28 days after second study vaccination (Day 49). Any approved or deployed SARS-CoV-2 vaccine should only be given at least 21 days after the most recent study vaccination.
- No influenza vaccine (except participants in the seasonal influenza co-administration sub-study) will be allowed 7 days before each vaccination.

  **NOTE:** Participants in the seasonal influenza co-administration sub-study will be allowed to have the co-administration of a licensed seasonal influenza vaccine at the same time as first study vaccination.
- No unlicensed vaccine should be given within 45 days prior to first study vaccination until after the last study visit.
- No investigational product (drug/biologic/device) within 45 days prior to first study vaccination until after the last study visit.
- No chronic administration (defined as more than 14 continuous days) of any immunosuppressant medication within 3 months of first study vaccination until the last study visit (except topical steroids or short-term oral steroids with course lasting ≤ 14 days). The use of topical, inhaled, and nasal glucocorticoids will be permitted if other chronic disease conditions are not exclusionary.
- No continuous use of anticoagulants, such as coumarins and related anticoagulants (i.e., warfarin) or novel oral anticoagulants/anti-platelet agents. Use of ≤ 325 mg of aspirin per day as prophylaxis is permitted.

### 6. STUDY PROCEDURES

Written informed consent will be obtained after explanation of the aims, benefits and all safety concerns of the trial as detailed in the information sheet BEFORE any trial specific procedures are performed. They should take as much time as they need to consider joining the study. Signed consent will be kept by the investigator and documented in medical notes and a copy given to the participant, as described in Section 9.3.2.3 (Appendix 2).

Due to the ongoing pandemic, recent national regulatory and local Ethics Committee and public health guidance will be applied at the site locations regarding alternations in the ability of study participants to attend an investigational site for protocol-specified visits, with the site's investigator being allowed to conduct safety assessments (e.g., telephone contact, alternative location for assessment, including local laboratories or imaging centres) when necessary and feasible, as long as such visits are sufficient to assure the safety of study participants. Serum samples may be drawn using local phlebotomy services, home health, or other modalities if site visits cannot occur. Study vaccination visits must have adequate oversight for issues associated with immediate severe reactions.

#### 6.1 Study Visit Procedures

##### 6.1.1 Days -30 to 0 – Screening

The following procedures will be performed within 30 days of first study vaccination. The Screening visit and Day 0 visit may be combined, if feasible, at any given study site.

- Written informed consent will be obtained in conformance with Section 9.3.2.3 of this protocol.
- Review of medical history, including prior and concomitant medical conditions, recent vaccinations (≤ 90 days), and significant surgical procedures.
- Inclusion and exclusion criteria review consistent with Section 4. Specific exclusions to study vaccination will be assessed before any vaccination. Waivers to enrolling participants with exclusions will not be given.
- Demographics, including date of birth (day, month, and year), sex, race, ethnicity, weight, and height.
- Prior and concomitant medications, including recent and current medications at the time of screening to be reviewed to ensure eligibility criteria are fulfilled. Concomitant medications include all medications (including vaccines) taken by the participant.
- Vital sign measurements, including respiratory rate, blood pressure, pulse rate, pulse oximetry, and oral temperature (or via forehead/ear reader).
- Urine pregnancy test for women of childbearing potential only. A positive urine pregnancy test at Screening will result in screen failure.

- Physical examination to include height and weight; head, eyes, ears, nose, and throat (HEENT), neck, thyroid, lungs, heart, cardiovascular, abdomen, lymph nodes of the upper extremities and neck; and musculoskeletal system/extremities to allow for study vaccination.
- Nose/throat samples will be collected to test for SARS-CoV-2 (PCR) only if the participant has any COVID-19 symptoms or significant exposure history. If a participant has a positive PCR for SARS-CoV-2 prior to enrolment that participant will not be eligible and will be considered a screen failure.
- Assessment of SAEs, starting from the time of informed consent.

##### 6.1.2 Day 0 – First Study Vaccination

The Screening and Day 0 Visits may be combined whenever feasible.

All participants with confirmed eligibility will have the following procedures performed:

- Inclusion and exclusion criteria review consistent with Section 4. Specific exclusions to study vaccination will be assessed before any study vaccination.
- Prior and concomitant medications, including recent and current medications to be reviewed to ensure eligibility criteria are fulfilled. Concomitant medications include all medications (including vaccines) taken by the participant.
- Vital sign measurements, including respiratory rate, blood pressure, pulse rate, pulse oximetry, and oral temperature (or via forehead/ear reader).
- Urine pregnancy test for women of childbearing potential only. A urine pregnancy test will be performed prior to each study vaccination. A positive urine pregnancy test at any time will result in the participant not receiving any further study vaccination.
- Symptom-directed (targeted) physical examination, including lymphatic assessment of injected upper extremity. Physical examination must be done prior to study vaccination.
- Nose/throat samples will be collected to test for SARS-CoV-2 (PCR) to determine current infection with SARS-CoV-2 and to demonstrate the methods required for nose/throat sample collection; participants will be taught how to self-sample at this time. Participants with possible COVID-19 symptoms will not be randomised but may return when symptoms have resolved per protocol. Participants found to be SARS-CoV-2 positive on Day 0 will be excluded from some analyses of the study as per the SAP.
- Blood sampling for SARS-CoV-2 prior to study vaccination (ELISA for anti-N-protein serology) in all participants.
- Blood sampling for SARS-CoV-2 prior to study vaccination (ELISA for anti-S-protein serology) in approximately 900 participants in the Anti-S Protein Serology Subset.

- Blood sampling for SARS-CoV-2 neutralisation assay prior to study vaccination in approximately 900 participants in the Neutralisation Assay Subset.
- Blood sampling for HAI prior to study vaccination for participants who received licensed seasonal influenza vaccine on Day 0 in the approximately 400 participants in the licensed seasonal influenza co-administration sub-study.
- Blood sampling for cell-mediated assessments prior to study vaccination, as measured by enzyme-linked immune absorbent spot (ELISpot) ± intracellular cytokine staining, in approximately 450 participants in the Cell-mediated Assay Subset.
- Randomisation.
- Pre-injection vital sign collection (respiratory rate, blood pressure, pulse rate, pulse oximetry, and oral temperature [or via forehead/ear reader]) before study vaccination to ensure participant has no evidence of fever prior to study vaccination.
- Alcohol swab cleansing of the injection sites for both study vaccine (SARS-CoV-2 rS with Matrix-M1 adjuvant or placebo) and licensed seasonal influenza vaccination (first approximately 400 eligible participants).
- Vaccination of study vaccine as an IM injection into the deltoid muscle. The first approximately 400 eligible participants will also receive an IM injection of a licensed seasonal influenza vaccine in the opposite deltoid following study vaccination.
- Monitoring for reactogenicity, participants will remain in clinic for at least 30 minutes to be monitored for any severe reactogenicity. Severe reactions will be noted as AEs on day of study vaccination.
- Post-injection vital sign collection (respiratory rate, blood pressure, pulse rate, pulse oximetry, and oral temperature [or via forehead/ear reader]) at approximately 15 to 30 minutes after study vaccination to check for any reactions to the study vaccine.
- Monitoring for COVID-19. Participants will monitor for COVID-19 symptoms. This will include provision of a Study Identification (ID) Card that provides details on study participation, study site contact information, and assessment of symptoms of suspected COVID-19 (see Table 2-2). Instructions to participants to contact the study site within 24 hours for symptoms of suspected COVID-19 will be given. A participant with suspected or confirmed COVID-19 will be asked to complete a COVID-19 symptom diary for 10 days. The Study ID Card should be presented to healthcare providers not affiliated with the study who encounter participants with symptoms of suspected COVID-19. Symptoms of reactogenicity within 7 days post-vaccination may not trigger the actions associated with COVID-19 monitoring.
- Assessment of unsolicited AEs, MAAEs (all and related to study vaccination), SAEs, and AESIs.

##### 6.1.3 Day 21 – Second Study Vaccination (+ 7 days)

All participants will have the following procedures performed:

- Inclusion and exclusion criteria review consistent with Section 4. Specific exclusions to study vaccination will be assessed before any study vaccination.
- Prior and concomitant medications. Concomitant medications include all medications (including vaccines) taken by the participant.
- Urine pregnancy test for women of childbearing potential only. A urine pregnancy test will be performed prior to each study vaccination. A positive urine pregnancy test at any time will result in the participant not receiving any further study vaccination.
- Symptom-directed (targeted) physical examination, including lymphatic assessment of injected upper extremity. Physical examination must be done prior to study vaccination.
- Nose/throat samples will be collected to test for SARS-CoV-2 (PCR) only if the participant has any COVID-19 symptoms or significant exposure history between Days 0 and 21. Results of that test are not required for study vaccination, but participants who are symptomatic may have the second study vaccination delayed per protocol. Participants found to be SARS-CoV-2 positive on Day 21 will be excluded from the immunogenicity analyses of the study as per the SAP.
- Blood sampling for HAI for participants who received licensed seasonal influenza vaccine on Day 0 in the approximately 400 participants in the licensed seasonal influenza co-administration sub-study.
- Pre-injection vital sign collection (respiratory rate, blood pressure, pulse rate, pulse oximetry, and oral temperature [or via forehead/ear reader]) before study vaccination to ensure participant has no evidence of fever prior to study vaccination.
- Alcohol swab cleansing of the injection site for study vaccine (SARS-CoV-2 rS with Matrix-M1 adjuvant or placebo).
- Vaccination of study vaccine as an IM injection into the opposite deltoid muscle of the Day 0 study vaccine.
- Monitoring for reactogenicity, participants will remain in clinic for at least 30 minutes to be monitored for any severe reactogenicity. Severe reactions will be noted as AEs on day of study vaccination.
- Post-injection vital sign collection (respiratory rate, blood pressure, pulse rate, pulse oximetry, and oral temperature [or via forehead/ear reader]) at approximately 15 to 30 minutes after study vaccination to check for any reactions to the study vaccine.
- Monitoring for COVID-19. Participants will monitor for COVID-19 symptoms. This will include provision of a Study ID Card that provides details on study participation, study site contact information, and assessment of symptoms of suspected COVID-19 (see Table 2-2). Instructions to participants to contact the study site within 24 hours for symptoms of suspected COVID-19 will be given. A participant with suspected or

confirmed COVID-19 will be asked to complete a COVID-19 symptom diary for 10 days. The Study ID Card should be presented to healthcare providers not affiliated with the study who encounter participants with symptoms of suspected COVID-19. Symptoms of reactogenicity within 7 days post-vaccination may not trigger the actions associated with COVID-19 monitoring.

- Assessment of unsolicited AEs, MAAEs (all and related to study vaccination), SAEs, and AESIs.

##### 6.1.4 Day 35 – Follow-up Visit (+ 7 days)

All participants will have the following procedures performed:

- Prior and concomitant medications. Concomitant medications include all medications (including vaccines) taken by the participant.
- Symptom-directed (targeted) physical examination may be performed if participant has any ongoing complaints.
- Blood sampling for SARS-CoV-2 (ELISA for anti-N-protein serology) in all participants unless the participant has had an Unblinding Visit in the prior 14 days where anti-N serology was drawn.
- Blood sampling for SARS-CoV-2 (ELISA for anti-S-protein serology) in approximately 900 participants in the Anti-S Protein Serology Subset.
- Blood sampling for SARS-CoV-2 neutralisation assay in approximately 900 participants in the Neutralisation Assay Subset.
- Blood sampling for cell-mediated assessments, as measured by ELISpot ± intracellular cytokine staining, in approximately 450 participants in the Cell-mediated Assay Subset.
- Monitoring for COVID-19. Participants will monitor for COVID-19 symptoms. This will include provision of a Study ID Card that provides details on study participation, study site contact information, and assessment of symptoms of suspected COVID-19 (see Table 2-2). Instructions to participants to contact the study site within 24 hours for symptoms of suspected COVID-19 will be given. A participant with suspected or confirmed COVID-19 will be asked to complete a COVID-19 symptom diary for 10 days. The Study ID Card should be presented to healthcare providers not affiliated with the study who encounter participants with symptoms of suspected COVID-19.
- Assessment of unsolicited AEs, MAAEs (all and related to study vaccination), SAEs, and AESIs.

##### 6.1.5 COVID-19 Surveillance Visits (Unscheduled)

###### 6.1.5.1 Initial COVID-19 Surveillance Visit

If the participant is known to be COVID-19 positive at the time of scheduling of the Initial Surveillance Visit and cannot attend the visit due to extenuating circumstances (e.g., local government restrictions), then a phone call or phone calls can be substituted to assess the participant's status.

All participants (including those who have been unblinded to treatment assignment due to receipt of an approved or deployed SARS-CoV-2 vaccine) will have the following procedures performed:

- Review of medical history, including prior and concomitant medical conditions, recent vaccinations (≤ 90 days), and significant surgical procedures.
- Prior and concomitant medications (as appropriate). Concomitant medications include all medications (including vaccines) taken by the participant.
- Vital sign measurements, including respiratory rate, blood pressure, pulse rate, pulse oximetry, and oral temperature (or via forehead/ear reader).
- Symptom-directed (targeted) physical examination, including a respiratory assessment.
- Nose/throat samples will be collected to test for SARS-CoV-2 (PCR) only if the participant has any COVID-19 symptoms or significant exposure history between Days 0 and 21. Results of that test are not required for study vaccination, but participants who are symptomatic may have the second study vaccination delayed per protocol. Participants found to be SARS-CoV-2 positive on Day 21 will be excluded from the immunogenicity analyses of the study as per the SAP.
- COVID-19 Symptom Diary. A participant with suspected or confirmed COVID-19 will be asked to complete the COVID-19 symptom diary. Participants should start the diary on their first day of symptoms and complete this diary daily for a minimum of 10 days even if their symptoms have resolved and regardless of any SARS-CoV-2 PCR results. They should continue the diary beyond 10 days if they have any ongoing symptoms related to the episode and should continue to do so until all symptoms have resolved.
- Assessment of MAAEs (related to study vaccination), SAEs, and AESIs.

###### 6.1.5.2 Follow-up COVID-19 Surveillance Visit

A Follow-Up COVID-19 Surveillance Visit will occur approximately 7 (+ 3) days after the Initial COVID-19 Surveillance Visit to ascertain worsening/progression of COVID-19 symptoms.

If the participant is known to be COVID-19 positive at the time of scheduling of the Initial Surveillance Visit and cannot attend the visit due to extenuating circumstances (e.g., local government restrictions), then a phone call or phone calls can be substituted to assess the participant's status. However, the participant should make every effort to attend the Follow-up Surveillance Visit (as per protocol timeline) when government restrictions or other circumstances allow. If a participant has made an Initial Surveillance phone contact or visit and is found to be COVID-19 negative on all swabs taken for this episode, then a phone call can be substituted for the Follow-up Surveillance Visit.

All participants (including those who have been unblinded to treatment assignment due to receipt of an approved or deployed SARS-CoV-2 vaccine) will have the following procedures performed:

- Review of medical history, including prior and concomitant medical conditions, recent vaccinations (≤ 90 days), and significant surgical procedures.
- Prior and concomitant medications (as appropriate). Concomitant medications include all medications (including vaccines) taken by the participant.
- Vital sign measurements, including respiratory rate, blood pressure, pulse rate, pulse oximetry, and oral temperature (or via forehead/ear reader).
- Symptom-directed (targeted) physical examination, including a respiratory assessment.
- Nose/throat samples will be collected to test for SARS-CoV-2 (PCR) only if the participant has any COVID-19 symptoms or significant exposure history between Days 0 and 21. Results of that test are not required for study vaccination, but participants who are symptomatic may have the second study vaccination delayed per protocol. Participants found to be SARS-CoV-2 positive on Day 21 will be excluded from the immunogenicity analyses of the study as per the SAP.
- COVID-19 Symptom Diary. A participant with suspected or confirmed COVID-19 will be asked to complete the COVID-19 symptom diary. Participants should start the diary on their first day of symptoms and complete this diary daily for a minimum of 10 days even if their symptoms have resolved and regardless of any SARS-CoV-2 PCR results. They should continue the diary beyond 10 days if they have any ongoing symptoms related to the episode and should continue to do so until all symptoms have resolved.
- Assessment of MAAEs (related to study vaccination), SAEs, and AESIs.

##### 6.1.6 Unblinding Visit

An Unblinding Visit should be arranged (where possible and practical to do so) for those who are offered and are willing to accept an approved or deployed SARS-CoV-2 vaccine.

If an unblinding visit occurs within 30 days of the scheduled 3- or 6-Month visit, the 3- or 6-Month visit may be skipped.

Unblinding after the blinded crossover is discouraged. If a participant has undergone both blinded crossover vaccinations and still desires to be unblinded and vaccinated with a deployed or approved vaccine, against national guidance, that participant should be withdrawn from the trial.

All participants will have the following procedures performed prior to unblinding of treatment assignment:

- Assessment of unsolicited AEs, MAAEs (all and related to study vaccination), SAEs, and AESIs.
- Prior and concomitant medications (as appropriate). Concomitant medications include all medications (including vaccines) taken by the participant.
- Blood sampling for SARS-CoV-2 (ELISA for anti-N-protein serology) in all participants. If a visit for anti-N-protein sampling has occurred 30 days before an unblinding visit this blood sampling may be skipped.

All participants should follow the following guidance after unblinding:

*For those who have received 1 or 2 doses of placebo:*

- The participant should be advised to receive the approved or deployed SARS-CoV-2 vaccine through the National Health Service (NHS). Participants will be asked to record the date and type of SARS-CoV-2 vaccine received.
- The participant should be advised that if they have not had a Day 21/35 Visit they may skip the Day 21 and Day 35 Visits and resume the protocol visits with the 3-Month Visit.

*For those who have received a single dose of the Sponsor's study vaccine:*

Those who are eligible to receive an approved or deployed SARS-CoV-2 vaccine and who, after unblinding to treatment assignment, are found to have received a single dose of study vaccine will be given an option to receive the second dose of study vaccine as per their current study vaccination schedule (in accordance with national policy). Alternatively, the participant may choose to receive a single dose of approved or deployed SARS-CoV-2 vaccine through the NHS. The safety and benefit of mixed SARS-CoV-2 vaccine schedules is unknown.

Any participant receiving the approved or deployed SARS-CoV-2 vaccine will be advised to allow a period of at least 3 weeks between vaccination with the Sponsor's study vaccine and the approved or deployed SARS-CoV-2 vaccine. Participants will be asked to record the date and type of vaccine received.

Participants who desire the option to receive the second dose of the Sponsor's study vaccine should attend the Day 21 Visit as scheduled and follow all protocol visits from that time forward.

Participants who would like a second dose of the approved or deployed SARS-CoV-2 vaccine should be vaccinated through the NHS; if they have not had a Day 21/35 Visit they may skip the Day 21 and Day 35 Visits and resume the protocol visits with the 3-Month Visit.

*For those who have received 2 doses of the Sponsor's study vaccine:*

In accordance with UK national policy, those participants who have received 2 doses of study vaccine will be advised to not receive the approved or deployed SARS-CoV-2 vaccine. The safety and benefit of mixed SARS-CoV-2 vaccine schedules is unknown. Any participant who still desires the approved or deployed SARS-CoV-2 vaccine after receiving 2 doses of study vaccine, against national policy, should be withdrawn from the study.

Any participant receiving the approved or deployed SARS-CoV-2 vaccine will be advised to allow a period of at least 3 weeks between vaccination with the Sponsor's study vaccine and the approved or deployed SARS-CoV-2 vaccine.

##### 6.1.7 3 Months (± 15 days) After Second Study Vaccination

This visit will apply to only those participants who have had this visit prior to enacting the blinded crossover or for those unblinded participants who are ineligible for the blinded crossover visits. If an unblinding visit has occurred within 30 days of the 3-Month visit, the 3-Month visit may be skipped.

This visit will be substituted for the Crossover Day 0 visit. In the event that the 3-Month visit falls within 30 days of the time the site is able to begin Crossover Day 0 visits, this visit may be skipped and the Crossover Day 0 visit will be scheduled as soon as is feasible.

All participants will have the following procedures performed:

- Prior and concomitant medications (as appropriate). Concomitant medications include all medications (including vaccines) taken by the participant.
- Blood sampling for SARS-CoV-2 (ELISA for anti-N-protein serology) in all participants unless the participant has had an Unblinding Visit in the prior 14 days where anti-N serology was drawn.
- Monitoring for COVID-19. Participants will monitor for COVID-19 symptoms. This will include provision of a Study ID Card that provides details on study participation, study site contact information, and assessment of symptoms of suspected COVID-19 (see Table 2-2). Instructions to participants to contact the study site within 24 hours for symptoms of suspected COVID-19 will be given. A participant with suspected or confirmed COVID-19 will be asked to complete a COVID-19 symptom diary for 10 days. The Study ID Card should be presented to healthcare providers not affiliated with the study who encounter participants with symptoms of suspected COVID-19.
- Assessment of MAAEs (related to study vaccination), SAEs, and AESIs.

##### 6.1.8 Crossover Day 0 Visit

This visit may take the place of the 3-Month visit and should occur approximately 3 months after the second dose of the initial set of vaccinations.

All blinded participants with no vaccination exclusions will have the following procedures performed:

- Written informed consent will be obtained in conformance with Section 9.3.2.3 of this protocol.
- Specific exclusions to study vaccination (e.g., anaphylaxis to previous doses, pregnancy) will be assessed before any study vaccination.
- Concomitant medications (as appropriate).
- Urine pregnancy test for women of childbearing potential only. A urine pregnancy test will be performed prior to each study vaccination. A positive urine pregnancy test at any time will result in the participant not receiving any further study vaccination.
- A symptom-directed (targeted) physical examination may be performed if the participant has any specific complaints or at investigator discretion.
- Blood sampling for SARS-CoV-2 prior to study vaccination (ELISA for anti-N-protein serology) in all participants.
- Blood sampling for SARS-CoV-2 prior to study vaccination (ELISA for anti-S-protein serology) in approximately 900 participants in the Anti-S Protein Serology Subset.
- Nose/throat samples will be collected to test for SARS-CoV-2 (PCR) to determine current infection with SARS-CoV-2. Participants with possible COVID-19 symptoms will not be dosed but may return when symptoms have resolved per protocol. Participants found to be SARS-CoV-2 positive will be excluded from some analyses of the study as per the SAP.
- IRT assignment of vaccination.
- Pre-injection vital sign collection (respiratory rate, blood pressure, pulse rate, pulse oximetry, and oral temperature [or via forehead/ear reader]) before study vaccination to ensure participant has no evidence of fever prior to study vaccination.
- Alcohol swab cleansing of the injection sites for study vaccine (SARS-CoV-2 rS with Matrix-M1 adjuvant or placebo).
- Vaccination of study vaccine as an IM injection into the deltoid muscle.
- Monitoring for reactogenicity, participants will remain in clinic for at least 30 minutes to be monitored for any severe reactogenicity. Severe reactions will be noted as AEs on day of study vaccination.

- Post-injection vital sign collection (respiratory rate, blood pressure, pulse rate, pulse oximetry, and oral temperature [or via forehead/ear reader]) at approximately 15 to 30 minutes after study vaccination to check for any reactions to the study vaccine.
- Monitoring for COVID-19. Participants will monitor for COVID-19 symptoms.
- Assessment of MAAEs (related to study vaccination), SAEs, and AESIs.

##### 6.1.9 Crossover Day 21 Visit (+7 days)

All blinded participants will have the following procedures performed:

- Concomitant medications (as appropriate).
- Urine pregnancy test for women of childbearing potential only. A urine pregnancy test will be performed prior to each study vaccination. A positive urine pregnancy test at any time will result in the participant not receiving any further study vaccination.
- A symptom-directed (targeted) physical examination may be performed if the participant has any specific complaints or at investigator discretion.
- Nose/throat samples will be collected to test for SARS-CoV-2 (PCR) only if the participant has any COVID-19 symptoms or significant exposure history between Crossover Day 0 and Crossover Day 21. Results of that test are not required for study vaccination, but participants who are symptomatic may have the second study vaccination delayed per protocol.
- Pre-injection vital sign collection (respiratory rate, blood pressure, pulse rate, pulse oximetry, and oral temperature [or via forehead/ear reader]) before study vaccination to ensure participant has no evidence of fever prior to study vaccination.
- Alcohol swab cleansing of the injection site for study vaccine (SARS-CoV-2 rS with Matrix-M1 adjuvant or placebo).
- Vaccination of study vaccine as an IM injection into the opposite deltoid muscle of the Crossover Day 0 study vaccine.
- Monitoring for reactogenicity, participants will remain in clinic for at least 30 minutes to be monitored for any severe reactogenicity. Severe reactions will be noted as AEs on day of study vaccination.
- Post-injection vital sign collection (respiratory rate, blood pressure, pulse rate, pulse oximetry, and oral temperature [or via forehead/ear reader]) at approximately 15 to 30 minutes after study vaccination to check for any reactions to the study vaccine.
- Monitoring for COVID-19. Participants will monitor for COVID-19 symptoms.
- Assessment of MAAEs (related to study vaccination), SAEs, and AESIs.

- .

##### 6.1.10 Crossover Day 35 Visit (+7 days)

Only for participants in Anti-S Protein Serology Subset. These participants will have the following procedures performed:

- Concomitant medications (as appropriate).
- Blood sampling for SARS-CoV-2 prior to study vaccination (ELISA for anti-S-protein serology)
- Monitoring for COVID-19. Participants will monitor for COVID-19 symptoms.
- Assessment of MAAEs (related to study vaccination), SAEs, and AESIs.

##### 6.1.11 6 Months (± 15 days) After Second Study Vaccination

This visit will apply to only those participants who have had this visit prior to enacting the blinded crossover or for those unblinded participants who are ineligible for the blinded crossover visits. If an unblinding visit has occurred within 30 days of the 6-Month visit, the 6-Month visit may be skipped.

- Concomitant medications (as appropriate)
- Blood sampling for SARS-CoV-2 (ELISA for anti-N-protein serology) in all participants.
- Monitoring for COVID-19. Participants will monitor for COVID-19 symptoms.
- Assessment of MAAEs (related to study vaccination), SAEs, and AESIs.

##### 6.1.12 12 Months (± 15 days) After Second Study Vaccination

All participants will have the following procedures performed:

- Prior and concomitant medications (as appropriate). Concomitant medications include all medications (including vaccines) taken by the participant.
- Blood sampling for SARS-CoV-2 (ELISA for anti-N-protein serology) in all participants.
- Assessment of MAAEs (related to study vaccination), SAEs, and AESIs.
- End of study form

#### 6.2 Efficacy assessments

##### 6.2.1 Nose/Throat Samples for Virus Detection

Nose/throat samples for virus detection will be taken at the study visits described in the schedule of events (SOE) (Table 3-1).

- Nose/throat samples will not be taken at Screening unless participants have symptoms or significant exposure to SARS-CoV-2. If a participant has a positive PCR for SARS-CoV-2 prior to enrolment that participant will not be eligible and will be considered a screen failure.
- Nose/throat samples will be taken on Day 0/Crossover Day 0 to determine current infection with SARS-CoV-2 and to demonstrate the methods required for nose/throat sample collection. Participants will be taught how to self-sample at this time. Participants with possible COVID-19 symptoms will not be randomised but may return when symptoms have resolved per protocol. Participants found to be SARS-CoV-2 positive on Day 0 will be excluded from the immunogenicity analyses of the study as per the SAP.
- Nose/throat samples will not be routinely taken on Day 21/Crossover Day 21. Participants with possible COVID-19 symptoms that develop between Day 0 and Day 21 or Crossover Day 0 and 21 may have a SARS-CoV-2 PCR test performed prior to second study vaccination on Day 21/Crossover Day 21. Results of that test are not required for vaccination, but participants who are symptomatic may have the second study vaccination delayed per protocol. Participants found to be SARS-CoV-2 positive on Day 21/Crossover Day 21 will be excluded from the immunogenicity analyses of the study as per the SAP.

##### 6.2.2 Monitoring for Suspected COVID-19

Identification and laboratory confirmation of SARS-CoV-2 infection and symptomatic COVID-19 will be performed throughout the study. This is the case for both blinded and unblinded participants.

On study enrolment, participants will be given details of a 24/7 on-call number to contact the study team. Participants will be instructed to contact the study team within 24 hours via this number if they self-assess COVID-19 symptoms in Table 2-2 or are clinically concerned. Newly discovered symptoms of suspected COVID-19 will trigger a COVID-19 Surveillance Visit.

Please refer to Table 2-2 for the qualifying symptoms of suspected COVID-19 disease.

Participants will receive weekly reminders (email or text messages) to immediately contact the study team if they have experienced any new symptoms or health concerns that could be related to infection with SARS-CoV-2. Newly discovered symptoms of suspected COVID-19 will trigger a COVID-19 Surveillance Visit.

A participant with suspected or confirmed COVID-19 will be asked to complete the COVID-19 symptom diary. Participants should start the diary on their first day of symptoms and complete this diary daily for a minimum of 10 days even if their symptoms have resolved and regardless of any SARS-CoV-2 PCR results. They should continue the diary beyond 10 days if they have any ongoing symptoms related to the episode and should continue to do so until all symptoms have resolved.

The occurrence of COVID-19-related hospitalisation and COVID-19-related complications (such as, but not limited to, pneumonia, neurological or vascular complications, severe pneumonia, severe neurological or vascular events, acute respiratory distress syndrome, renal complications, sepsis, septic shock, death) will be monitored throughout the study.

Every episode of a new onset of symptoms of suspected COVID-19 will trigger a COVID-19 Surveillance Visit (Initial and Follow-Up) with exceptions per protocol. A "new onset" episode will require at least a 7-day period of being symptom-free prior to the event to differentiate a specific episode from any prior illness.

###### 6.2.2.1 Severity of COVID-19 Symptoms

COVID-19 symptoms will be categorised as mild, moderate, or severe as described in Table 2-1. Participants with a single vital sign abnormality placing them in the moderate or severe COVID-19 severity categories must also meet the criteria for mild COVID-19.

###### 6.2.2.2 COVID-19 Surveillance Visit (Initial and Follow-up)

A COVID-19 Surveillance Visit (Initial and Follow-Up) will be triggered by symptoms of suspected COVID-19 captured by surveillance. See Section 6.1.5.1 for exceptions for participants affected by extenuating circumstances (e.g., local government restrictions).

When a participant is determined to have a new onset of symptoms, the participant will contact the study team immediately, begin their COVID-19 symptom diary and begin the 3 consecutive days of PCR self-testing (beginning approximately 24 hours after the start of symptoms) as above. Participants will be asked to attend an Initial COVID-19 Surveillance Visit at the study clinic or will be seen at an in-home visit by study staff depending on local conditions.

###### 6.2.2.2.1 Initial COVID-19 Surveillance Visit

An Initial COVID-19 Surveillance Visit will be performed at the study site (or home) and will occur as soon as possible within approximately **1-3 days** of new symptom onset (however, data from specimens obtained up to 14 days will be accepted). The visit will consist of the following:

- Review and confirmation of the history of COVID-19 symptoms, including approximate date of onset of illness and solicitation of each symptom (see Table 2-2).

- Vital signs, including resting respiratory rate (on room air) and pulse oximetry, will be captured as numerical values. Lung auscultation (exam) will be performed.
- Ascertainment of any unscheduled healthcare visit by the participant (or home visit by a healthcare provider) in response to symptoms of suspected COVID-19.
- Ascertainment of new concomitant medications as appropriate (especially antibiotics) or altered doses/frequencies of existing concomitant medications resulting from symptoms of suspected COVID-19.
- Proper documentation in the eCRF of the dates, bar codes, and results of all self-swabs taken in proximity of this visit.

###### 6.2.2.2.2 Follow-Up COVID-19 Surveillance Visit

A Follow-Up COVID-19 Surveillance Visit will occur approximately 7 (+ 3) days after the Initial COVID-19 Surveillance Visit. This visit will consist of the following:

- Study staff will conduct the Follow-Up COVID-19 Surveillance Visit approximately 7 (+ 3) days after the Initial COVID-19 Surveillance Visit to ascertain worsening/ progression of COVID-19 symptoms. This follow-up visit by study staff will include all of the same procedures outlined in this section, including review of symptom history, measurement of vital signs (including pulse oximetry), and lung auscultation.

After the Follow-Up COVID-19 Surveillance Visit, participants will continue to receive telephone contacts approximately every week for ascertainment of COVID-19 symptom status until resolution of symptoms. Subsequent calls will document resolution or return to baseline of COVID-19 symptoms in order to calculate illness duration (date of symptom onset to day of symptom resolution) and will collect any additional healthcare visits, hospitalisations, and/or concomitant medications due to the suspected COVID-19.

Should a participant visit an emergency room, be admitted to the hospital or a COVID-19 ward, and PCR sampling is missed, then the local public health COVID-19 diagnostic test (or COVID-19 diagnostic test performed in the healthcare setting) will be taken as a valid result. Importantly, clinical data on symptoms, vital signs, exam findings, COVID-19 and other diagnostic testing, diagnoses, and complications occurring during the suspected COVID-19 emergency room and/or hospitalisation episode will be collected from available medical records on a study specific hospitalisation/emergency room data collection form in order to assess severity.

Participants will be notified of positive SARS-CoV-2 results as soon as locally practicable due to requirements of self-isolation and potential transmission.

Note that PCR-positive COVID-19 symptoms and events captured as efficacy endpoints will NOT be doubly recorded in the AE eCRF, unless a particular illness fulfils the definition of an SAE.

#### 6.3 Immunogenicity Assessments

Blood will be collected from all participants for humoral immunogenicity according to the time points specified in the SOE (Table 3-1).

Immune measurements (ELISA) will be conducted on serum (IgG) for SARS-CoV-2 anti-S protein serology in approximately 900 participants in the Anti-S Protein Serology Subset (after both sets of vaccinations) and on anti-N protein serology in all participants. The immunologic test for SARS-CoV-2 serology based on a non-S protein (e.g., SARS-CoV-2 N protein) will be performed to identify cases of asymptomatic infection. Additional immunogenicity assessments specific to SARS-CoV-2 (or related variants) will include a neutralising antibody assay, which will be performed in approximately 900 participants in a Neutralisation Assay Subset. Cell-mediated immune responses, as assessed by ELISpot ± intracellular cytokine staining, will be performed in approximately 450 participants in the Cell-mediated Assay Subset. In addition, an HAI will be performed in the approximately 400 participants in the seasonal influenza vaccine co-administration sub-study.

Anti-S protein serology in the Anti-S Protein Serology Subset will be collected after the initial and second set of vaccinations.

Details on the handling, processing, and shipping of immunogenicity samples will be provided separately in a laboratory manual.

Participants will be asked to provide consent for the use of samples for future testing or assay development specific to SARS-CoV-2 (or related variants). Aliquots of all collected samples from this study may be retained for additional testing with antigens specific to SARS-CoV-2 (or related variants) for a maximum of 25 years (starting from the date at which the last participant had the last study visit), unless local rules, regulations, or guidelines require different timeframes or different procedures, in accord with participant consent.

#### 6.4 Safety Assessments

The timing and frequency of all safety assessments are listed in the SOE (Table 3-1).

After each study vaccination, including the licensed seasonal influenza vaccine for participants in the seasonal influenza vaccine co-administration sub-study, participants will remain under observation at the study site for at least 30 minutes for the presence of any acute reactions and solicited AEs. Solicited AEs, collected through an electronic diary (e-Diary), will be recorded from the time of study vaccination until 7 days after study vaccination in all participants in the seasonal influenza vaccine co-administration sub-study and a subgroup of all other participants after the initial set of vaccinations. As solicited AEs are only being recorded in a subset of participants, it is expected that typical solicited AEs will be captured as unsolicited AEs in those participants who are not in the sub-study. An assessment of the impact of this decision will be part of an exploratory analysis. Participants in the licensed seasonal influenza vaccine co-administration sub-study will record local reactogenicity for the study vaccine injection site only.

All participants will be assessed for unsolicited AEs from the time of first study vaccination until Day 49 after the initial set of vaccinations; SAEs will be assessed from the time the informed consent is signed until completion of the participant's last study-related procedure; all MAAEs will be reported from the time of first study vaccination until Day 35 in the initial set of vaccinations; MAAEs related to study vaccination and AESIs will be reported from the time of first study vaccination until completion of the participant's last study-related procedure. All AEs will be followed until resolution or until clinically stable.

Separate analyses of safety events received with respect to approved or deployed SARS-CoV-2 vaccines will be performed.

Safety assessments will include monitoring and recording of solicited (local and systemic reactogenicity events) and unsolicited AEs; MAAEs; AESI; SAEs; vital sign measurements on day of study vaccinations; and physical examination findings. Local and systemic reactogenicity events will not be recorded for the second set of vaccinations. COVID-19 severity will be categorised as mild, moderate, or severe according to protocol-specified criteria (Table 2-1). Recording of solicited and unsolicited AEs may be conducted by electronic data capture (EDC)/reporting. PIMMCs and AESIs specific to potential disease enhancement for COVID-19 will also be monitored (see Section 9.5 [Appendix 4] for details).

Participants who are unblinded to treatment assignment for the purpose of receiving an approved or deployed vaccine as per UK guidance will be encouraged to remain in the study for safety follow-up, and safety assessments will be performed via the timelines and mechanisms as described above and throughout Section 6.4. Unblinded participants should not be included in the blinded crossover.

In addition, investigators will be required to report any suspected adverse reactions to another manufacturer's approved or deployed SARS-CoV-2 vaccine to healthcare authorities via the Coronavirus Yellow Card reporting site: https://coronavirus-yellowcard.mhra.gov.uk/.

##### 6.4.1 Adverse Events

AEs will be assessed during the study as described in the SOE (Table 3-1) and should be followed until they are resolved, stable, or judged by the investigator to be not clinically significant. AEs will be captured after the first dose of study vaccine administered with the exception of an AE related to study procedure or one that causes a delay in study vaccine administration (e.g., acute illness).

The investigator is responsible for ensuring that all AEs and SAEs are recorded in the eCRF and reported to the sponsor, regardless of their relationship to study vaccine or clinical significance. If there is any doubt as to whether a clinical observation is an AE, the event should be reported.

###### 6.4.1.1 Adverse Event Definitions

The investigator is responsible for reporting all AEs that are observed or reported during the study, regardless of their relationship to study vaccination or their clinical significance.

An AE is defined as any untoward medical occurrence in a participant enrolled into this study regardless of its causal relationship to study vaccination. Participants will be instructed to contact the investigator at any time after randomisation if any symptoms develop.

###### 6.4.1.1.1 Serious Adverse Events

An SAE is defined as any event that

- results in death
- is immediately life threatening
- requires inpatient hospitalisation or prolongation of existing hospitalisation
- results in persistent or significant disability/incapacity
- is a congenital anomaly/birth defect

Important medical events that may not result in death, be life threatening, or require hospitalisation may be considered SAEs when, based upon appropriate medical judgment, they may jeopardise the participant or may require medical or surgical intervention to prevent one of the outcomes listed in this definition. Examples of such medical events include allergic bronchospasm requiring intensive treatment in an emergency room or at home, blood dyscrasias or convulsions that do not result in inpatient hospitalisation, or the development of drug dependency or drug abuse.

###### 6.4.1.1.2 Local and General Systemic Reactogenicity Symptoms

Solicited AEs (e.g., local and general systemic reactogenicity symptoms) will be captured in the Safety Subset of approximately 2,000 participants and in the seasonal influenza vaccine co-administration sub-study of approximately 400 participants after the initial set of vaccinations only. Participants will record all local reactogenicity symptoms for each injection of study vaccine at each location (ideally in opposite deltoids) while recording of general systemic reactogenicity symptoms may not be assigned to either injection site. Local reactogenicity symptoms should not be recorded for the influenza vaccine injection site. Solicited AEs, collected through an electronic diary (e-Diary), will be recorded from the time of study vaccination until 7 days after study vaccination.

Site-specific local (arm) and general systemic reactogenicity reactions including start and stop dates will be recorded and the investigator will apply a standard toxicology grading at the subsequent study visit (Section 9.4, Appendix 3). Should any reactogenicity event extend beyond 7 days after study vaccination and be clinically significant by toxicity grade 1 or greater, then it will be recorded as an unsolicited AE with a start date on the 8$^{th}$ day following study vaccination and followed to resolution.

Solicited AEs will not be captured for any approved or deployed SARS-CoV-2 vaccines.

###### 6.4.1.1.3 Adverse Events of Special Interest

Participants will be assessed for diagnosis of an AESI at all study visits. AESIs include PIMMCs, AEs specific to COVID-19, or other potential AEs that may be determined at any time by regulatory authorities as additional information concerning COVID-19 is obtained. Given the concern for cytokine storm, an AESI of cytokine release syndrome will be included as an AE specific to COVID-19. Listings of AESI are presented in Section 9.5, Appendix 4.

###### 6.4.1.1.4 Medically Attended Adverse Events

MAAEs are defined as AEs with medically attended visits including hospital, emergency room, urgent care clinic, or other visits to or from medical personnel for any reason. Routine study visits (including COVID-19 Surveillance Visits) will not be considered medically attended visits. MAAEs are to be reported from the time of first study vaccination until Day 35 for the initial set of vaccinations only. MAAEs related to study vaccination are to be reported from the time of first study vaccination until completion of the participant's last study-related procedure, which may include contact for safety follow-up.

###### 6.4.1.1.5 Pregnancy

Pregnancy is not considered an AE unless there is a suspicion that an investigational vaccine may have interfered with the effectiveness of a contraceptive medication. Any pregnancy that occurs during study participation must be reported using a clinical study pregnancy form. To ensure participant safety, each pregnancy must be reported to Novavax, Inc. within 2 weeks of learning of its occurrence. If pregnancy occurs, further study vaccination will be discontinued. The pregnancy must be followed up to determine outcome (including spontaneous miscarriage, elective termination, normal birth, or congenital abnormality) and the status of both mother and child, even if the participant was discontinued from the study. Pregnancy complications and elective terminations for medical reasons must be reported as an AE or SAE. Spontaneous miscarriages must be reported as an SAE.

Any pregnancy brought to the investigator's attention after the participant has completed the study but occurring while the participant was in the study must be promptly reported to:

Sponsor Safety Monitor: [redacted]

###### 6.4.1.2 Eliciting and Documenting Adverse Events

At every study visit, participants will be asked a standard question to elicit any medically related changes in their well-being. They will also be asked if they have been hospitalised, had any accidents, used any new medications, or changed concomitant medication regimens (both prescription and OTC medications).

In addition to participant observations, AEs will be documented from any data collected on the AE page of the eCRF or other documents that are relevant to participant safety.

###### 6.4.1.3 Reporting Adverse Events

All AEs reported or observed during the study will be recorded on the AE page of the eCRF. Information to be collected includes study vaccine, type of event, time of onset, dosage, investigator-specified assessment of severity and relationship to study vaccine and/or study procedure, time of resolution of the event, seriousness, any required treatment or evaluations, and outcome. Any AEs resulting from concurrent illnesses, reactions to concurrent illnesses, reactions to concurrent medications, or progression of disease must also be reported. All AEs will be followed until they are resolved, stable, or judged by the investigator to be not clinically significant. The Medical Dictionary for Regulatory Activities (MedDRA) will be used to code all AEs.

Any medical condition that is present at the time that the participant is screened but does not deteriorate should not be reported as an AE. However, if it deteriorates at any time during the study, it should be recorded as an AE.

Any AE that is considered serious by the investigator or that meets SAE criteria (Section 6.4.1.1.1) must be reported to the sponsor within 24 hours after the investigator has confirmed the occurrence of the SAE. The investigator will provide a causality assessment (whether there is a reasonable possibility that the study vaccine caused the event) to the study vaccine. The sponsor will be responsible for notifying the relevant regulatory authorities of any SAE, in compliance with health authority requirements, as outlined in the relevant clinical study guidelines. SAE reports that may be attributed to a combination of the Novavax vaccine and an approved or deployed SARS-CoV-2 vaccine will be reported to the regulatory authorities as applicable.

SAE reporting forms allow for the notation of other factors that may have impacted the investigator's assessment of causality. Investigators will be instructed to utilize this section of the reporting form to note the impact of an approved or deployed SARS-CoV-2 vaccine on the event, if applicable. Investigators will be required to report any suspected adverse reactions to another manufacturer's approved or deployed SARS-CoV-2 vaccine to health care authorities via the Coronavirus Yellow Card reporting site: https://coronavirus-yellowcard.mhra.gov.uk/.

For this study, the following contact information will be used for SAE reporting:

Phone: [redacted]
Fax: [redacted]
Email: [redacted]

###### 6.4.1.4 Assessment of Severity

The severity (or intensity) of an AE refers to the extent to which it affects the participant's daily activities and will be classified as mild, moderate, or severe using the following criteria:

- Mild (grade 1): These events require minimal or no treatment and do not interfere with the participant's daily activities.
- Moderate (grade 2): These events result in a low level of inconvenience or require minor therapeutic measures. Moderate events may cause some interference with normal functioning.
- Severe (grade 3): These events interrupt a participant's usual daily activity and may require systemic drug therapy or other treatment. Severe events are usually incapacitating.

If the severity of an AE changes, the most intense severity should be reported. An AE characterised as intermittent does not require documentation of the onset and duration of each episode.

###### 6.4.1.5 Assessment of Causality

The investigator's assessment of an AE's relationship to study vaccine is part of the documentation process, but it is not a factor in determining what is or is not reported in the study. If there is any doubt as to whether a clinical observation is an AE, the event should be reported.

The investigator will assess causality (i.e., whether there is a reasonable possibility that the study vaccine caused the event) for all AEs and SAEs (solicited reactions are to be considered as being related to study vaccination). The relationship will be classified as follows:

- Not related: There is not a reasonable possibility of relationship to study vaccine. The AE does not follow a reasonable temporal sequence from administration of study vaccine or can be reasonably explained by the participant's clinical state or other factors (e.g., disease under study, concurrent diseases, and concomitant medications).
- Related: There is a reasonable possibility of relationship to study vaccine. The AE follows a reasonable temporal sequence from administration of study vaccine and cannot be reasonably explained by the participant's clinical state or other factors (e.g., disease under study, concurrent diseases, or concomitant medications), represents a known reaction to study vaccine or other vaccines in its class, is consistent with the known pharmacological properties of the study vaccine, and/or resolves with discontinuation of the study vaccine (and/or recurs with re-challenge, if applicable).

###### 6.4.1.6 Follow-up of Adverse Events

All AEs must be reported in detail on the appropriate page of the eCRF and followed until they are resolved, stable, or judged by the investigator to be not clinically significant.

##### 6.4.2 Vital Sign Measurements

Vital sign measurements will include oral temperature (or via forehead/ear reader), pulse rate and diastolic and systolic blood pressure (after participant is seated for at least 5 minutes), and pulse oximetry. Temperature will be recorded and graded during general systemic reactogenicity evaluation (Section 6.4.1.1.2). The other vital sign measurements will be recorded as continuous variables prior to each study vaccination. Pulse oximetry and other vital signs will be taken on room air.

On study vaccination days, vital sign measurements will be collected once before study vaccination to ensure participant has controlled blood pressure and heart rate and no evidence of fever prior to study vaccination and once more, at approximately 15 to 30 minutes after study vaccination, to check for any reactions to the study vaccine. The investigator will only apply standard toxicology grading on the day of study vaccination, both before and after study vaccination (Section 9.4, Appendix 3). If individual vital sign measurements are considered clinically significant by the investigator, study vaccination may be withheld that day, and participants may return on a subsequent day for re-evaluation and study vaccination, ideally, within the time window specified in the SOE (Table 3-1).

##### 6.4.3 Physical Examinations

A physical examination will be performed at screening/Day 0 (at minimum, assessment of skin, head, ears, eyes, nose, throat, neck, thyroid, lungs, heart, cardiovascular, abdomen, lymph nodes of the upper extremities and neck, and musculoskeletal system/extremities). Height and weight will be measured at screening only.

A targeted or symptom-directed physical examination will be performed at the time points specified in the SOE (Table 3-1). Special attention should be made to examine the lymph nodes of the upper extremities on vaccination days for the initial set of vaccinations and the respiratory system at all COVID-19 Surveillance Visits.

##### 6.4.4 Safety Monitoring

Safety oversight will be conducted by an SMC during the course of the study. The SMC is an independent group of experts that monitors participant safety and advises the sponsor. The SMC members will be separate and independent of site personnel participating in this study and should not have a scientific, financial, or other conflict of interest related to this study or the sponsor. The SMC will consist of members with appropriate expertise to contribute to the interpretation of the data from this study.

A global SMC was convened to oversee the safety of the ongoing Phase 1/2 study (Protocol 2019nCoV-101) and will oversee 1 or more additional studies across the SARS-CoV-2 rS vaccine program. A separate SMC will be convened for this study. The designated SMC will monitor the safety of participants in the study and will follow an SMC charter. The SMC will review blinded and unblinded safety and reactogenicity data. The SMC will also review sufficient clinical event data to ensure detection of evidence of enhanced disease. The SMC will convene to perform safety reviews on a scheduled basis as per the SMC charter; for immediate concerns regarding safety observations during this study; and as needed.

The SMC will operate under the rules of a sponsor-approved charter that will be approved at the organisational meeting of the SMC. At this time, each data element that the SMC needs to assess will be clearly defined. Procedures for SMC reviews/meetings will be defined in the charter. The SMC will review applicable data for safety assessments (AEs by classifications) and any clinical data that may be of significance to this review (e.g., demographics, study vaccination timing, and medications). Additional data may be requested by the SMC, and interim statistical reports may be generated as deemed necessary and appropriate for this review. The SMC may receive data in aggregate and presented by study vaccine group. The SMC may also be provided with expected and observed rates of the expected AEs in an unblinded fashion and may request the study vaccine assignment be unblinded for an individual participant if required for safety assessment.

### 7. STATISTICAL ANALYSIS PLANS

#### 7.1 Sample Size Calculations

This study is designed to enrol approximately 15,000 participants, who will be initially randomised 1:1 into the 2 study vaccine groups. The sample size is driven by the total number of events expected to achieve statistical significance for the primary efficacy endpoint – a target of 100mild, moderate, or severe COVID-19 cases. The target number of events of 100 was chosen to provide > 95% power for 70% or higher vaccine efficacy (VE) (Table 7-1). A single interim analysis of efficacy will be conducted based on the accumulation of approximately 50% (50 events) of the total anticipated primary endpoints using Pocock boundary conditions. Power calculations were performed by 10,000 simulated trials that were created under various assumptions of VEs and analyzed using methods described in the "efficacy analysis" section without covariates. A 90% evaluability rate for the per-protocol (PP) efficacy (PP-EFF) population (assuming 10% unevaluable due to attrition and/or baseline-seropositive participants) was used for the power calculations. All simulations were performed in SAS V9.4.

**Table 7-1 Power Under Various Vaccine Efficacy Assumptions**

| Assumed Vaccine Efficacy | Estimated Power | | |
|---|---|---|---|
| Symptomatic COVID-19 Illness PCR-Confirmed SARS-CoV-2 Infection | At Planned Interim Analysis with 50 Events | At Final Analysis with 100 Events | Overall (At Interim Analysis or Final Analysis) |
| 60% | 29% | 39% | 68% |
| 65% | 45% | 41% | 87% |
| 70% | 64% | 32% | 96% |
| 75% | 81% | 18% | >99% |
| 80% | 94% | 6% | >99% |
| 85% | 99% | 1% | >99% |
| 90% | >99% | <10% | >99% |

Abbreviations: COVID-19 = coronavirus disease 2019; PCR = polymerase chain reaction; SARS-CoV-2 = severe acute respiratory syndrome coronavirus 2

#### 7.2 Analysis Sets

The all-randomised set will include all participants who were randomised regardless of the receipt of any study vaccine (SARS-CoV-2 rS with Matrix-M1 adjuvant or placebo). The all randomised set will be used for the subject disposition summaries.

The intent-to-treat (ITT) analysis set will include all participants who are randomised and receive at least 1 dose study vaccine (SARS-CoV-2 rS with Matrix-M1 adjuvant or placebo), regardless of protocol violations or missing data. The ITT analysis set will be used as a supportive analysis population for the immunogenicity and efficacy analyses and will be analysed according to the study vaccine group as randomised.

The safety analysis set will include all participants who receive at least 1 dose of study vaccine (SARS-CoV-2 rS with Matrix-M1 adjuvant or placebo). Participants in the safety analysis set will be analysed according to the study vaccine actually received.

The PP-EFF will include baseline seronegative participants who receive both doses of study vaccine (SARS-CoV-2 rS with Matrix-M1 adjuvant or placebo) and have no major protocol deviations that occur before the first COVID-19 episode affecting the primary efficacy outcome (i.e., participants will be censored at the time of the protocol deviation) as assessed by the sponsor prior to unblinding. All analyses of the PP-EFF population will also exclude any illness episodes with positive SARS-CoV-2 by any validated PCR or antibody test occurring 6 days or less after the second study vaccination (e.g., Day 28).

The per-protocol immunogenicity (PP-IMM) analysis set for each post-randomisation visit will include participants who receive scheduled dose(s) of study vaccine (SARS-CoV-2 rS with Matrix-M1 adjuvant or placebo), have at least a baseline and the serum sample result for the visit available after study vaccination, and have no major protocol violations that are considered clinically likely to impact the immunogenicity response at the corresponding study visit as assessed by the sponsor prior to unblinding. For each visit, the SARS-CoV-2 unexposed population will also exclude any illness episodes with positive SARS-CoV-2 by any validated PCR or antibody test prior to each visit. Prior exposed participants will be determined using baseline SARS-CoV-2 immunity defined as positive SARS-CoV-2 by PCR or antibody test at baseline, or positive SARS-CoV-2 by qualitative PCR through Day 35, according to the specified analysis. Analysis will be performed to assess if immune responses differ between exposed and unexposed individuals (i.e., whether prior exposure alters dosing regimen considerations in a pandemic response).

The review and determination for exclusion from the PP populations will be carried out in a blinded fashion by a study clinician prior to unblinding for each interim analysis based on all available information from the locked database. Both PP populations will be analysed according to the study vaccine group as randomised.

#### 7.3 Statistical Analysis

Details of all statistical analyses will be described in the SAP.

All data collected will be presented in data listings. Data from participants excluded from an analysis set will be presented in the data listings but not included in the calculation of summary statistics for that analysis set.

For categorical variables, frequencies and percentages will be presented. Continuous variables will be summarised using descriptive statistics (number of participants, mean, median, minimum, and maximum).

Baseline demographic and background variables will be summarised by study vaccine group. The number of participants who enrol in the study and the number and percentage of participants who complete the study will be presented. Frequency and percentage of participants who withdraw or discontinue from the study, and the reason for withdrawal or discontinuation, will also be summarised.

In order to manage unblinded participants, unblinding will result in censoring of all efficacy and immunogenicity endpoints. The main presentation of data based on the Safety Analysis set will include all participant data, with supporting presentations to exclude the data post unblinding. Sensitivity analysis of reactogenicity and unsolicited AEs will be conducted for the Safety analysis subset of participants not unblinded. Additional analysis of AEs collected after unblinding may be undertaken as exploratory safety analyses.

##### 7.3.1 Efficacy Analyses

The primary endpoint will be analysed on the PP-EFF Analysis Set and supported by analysis of the ITT Analysis Set. Conclusions concerning declaration of attainment of the primary endpoint will only be based on the PP-EFF population.

Primary analysis of the primary and key secondary efficacy endpoints will be performed based on the data generated prior to the blinded crossover. The analysis of data generated after the blinded crossover or the combined analyses of both pre- and post-blinded crossover will be performed using the approach described by Follmann et al (2020). Additional details on the analytical approach will be described in the Statistical Analysis Plan.

VE is defined as VE (%) = (1 – RR) × 100, where RR = relative risk of incidence rates between the 2 study vaccine groups (SARS-CoV-2 rS with Matrix-M1 adjuvant or placebo). The interim and final analyses for the primary objective in the PP-EFF population will be carried out at the overall one-sided Type I error rate of 0.025. The nominal alpha to be spent for the final analysis will be recalculated using the Lan-DeMets alpha spending function based on the actual numbers of events used for the interim analysis and the numbers of endpoints to be used for the final analysis. The estimated RR and its CI will be derived using Poisson regression with robust error variance [Zou 2004]. The explanatory variables in the model will include study vaccine group. The dependent variable will be the incidence rate of the endpoint of interest. The robust error variances will be estimated using the repeated statement and the participant identifier. The Poisson distribution will be used with a logarithm link function. The site (depending on the distribution of endpoints) and the age strata will be included in the model as covariates. To assess incidence rates rather than absolute counts of cases, accounting for differences in follow-up times starting with 7 days after the second vaccination among participants, an offset will be utilized in the Poisson regression. In case the total number of events to be analysed may be too low for an asymptotic method proposed (i.e., ≤ 5 events in either treatment group), an alternative method based on the single sample exact binomial distribution may be used for the analysis. This method is based on the proportion of the events in the SARS-CoV-2 rS with Matrix-M1 adjuvant group among the total number of events observed in both treatment groups after adjusting for the differential number of subjects (or inclusive of differential lengths of follow-up) between the 2 treatment groups.

Hypothesis testing of the primary efficacy endpoint will be carried out against H0: VE ≤ 30%. Rejection of the null hypothesis, H0: VE ≤ 30% demonstrates a statistically significant vaccine effect (i.e., alpha-adjusted lower bound confidence interval [LBCI] > 30%) will be considered meeting the prespecified study success criterion. The study will also continue for the intended duration to measure efficacy, immunogenicity, and safety endpoints, regardless of primary endpoint success at the interim or final analysis. The final analysis of the primary efficacy endpoint will be triggered when approximately 100 PP-EFF participants with symptomatic mild, moderate, or severe COVID-19 endpoints have accrued. Also, in order to be able to respond to the unexpected and rapidly evolving COVID-19 pandemic situation globally, other factors such as requests by government or public health agencies may also be factored into the decision-making to unblind the study for the final analysis, but this always occurs in consultation with lead regulatory agencies.

The secondary and exploratory efficacy endpoints will be analysed using the same method as the primary efficacy analysis described above. Analysis of secondary and exploratory efficacy endpoints will be performed without adjustment for multiple comparisons (i.e., two-sided alpha of 0.05). The final interpretation of the overall vaccine efficacy will be based on the totality of statistical evidence, including immunogenicity results and the clinical importance in discussions with the regulatory agencies and scientific communities. The EOS analysis will be performed when the last participant completes the last visit (12 months after the last study vaccination) or discontinues earlier.

##### 7.3.2 Immunogenicity Analysis and Correlates of Risk

The primary and secondary immunogenicity analyses will be performed using the PP-IMM and ITT analysis populations.

For the SARS-CoV-2 rS serum antibody levels measured by microneutralization and ELISA assays, geometric mean at each study visit, the geometric mean fold rises (GMFRs) comparing to Day 0 (baseline) at each follow-up study visit, along with 95% CI will be summarised by study vaccine group. The 95% CI will be calculated based on the t distribution of the log-transformed values for geometric means or GMFRs, then back transformed to the original scale for presentation. The seroconversion rate (SCR), proportion of participants with ≥ 4 fold rises if naïve at baseline, along with 95% CIs based on the Clopper-Pearson method will be summarised by study vaccine group at each follow-up study visit. Immunogenicity analyses performed after the second set of vaccinations will be conducted in a similar fashion.

For the subset of participants who receives the influenza vaccine concurrently with the study vaccines, comparisons of strain-specific immune responses to influenza vaccine as measured by HAI will be performed. The treatment comparison will be made by comparing the strain-specific GMTs and the SCRs. The SCR is defined as the proportion of subjects with either a baseline reciprocal (Day 0) titre of < 10 and a post-vaccination reciprocal titre ≥ 40, or a baseline titre of ≥ 10 and a post-vaccination titre ≥ 4-fold higher.

For influenza strain-specific GMTs in each treatment group, titres reported below the lower limit of quantitation (LLOQ; i.e., below the starting dilution of assay reported as "< 10") will be set to half that limit (i.e., 10 / 2 = 5). Strain-specific GMTs will be summarized by treatment group and visit day along with the corresponding two-sided 95% CIs, by exponentiating the corresponding log-transformed means and their 95% CIs. The ratio of GMTs between treatment groups post-vaccination and the corresponding two-sided 95% CI will be calculated on log-transformed titres using the analysis of covariance (ANCOVA) with treatment group and baseline (Day 0) measurement as the covariate.

For influenza strain-specific SCRs, the rate in percent and the corresponding two-sided exact binomial 95% CIs will be calculated using the Clopper-Pearson method. The two-sided 95% CIs for the absolute rate difference between the 2 treatment groups will be constructed using the Newcombe method.

Similar summaries will be generated for the other immunogenicity endpoints and other assays if conducted.

Correlates of risk will be assessed where immune responses are measured in all available participants, and the results compared between participants who experience a SARS-CoV-2 event and participants who do not. The goal of this analysis is to assess correlates of risk of SARS-CoV-2 infection (and potential other secondary endpoints) in the study vaccine group by comparing study vaccine-induced immune responses associated with COVID-19.

##### 7.3.3 Safety Analyses

Solicited AEs (e.g., local and general systemic reactogenicity symptoms) will be captured in the Safety Subset of approximately 2,000 participants and in the seasonal influenza vaccine co-administration sub-study of approximately 400 participants after the initial set of vaccinations only.

Numbers and percentages (with 95% CIs based on the Clopper-Pearson method) of participants with solicited local and systemic AEs through 7 days after each study vaccination will be summarised by study vaccine group and the maximum toxicity grade over 7 days after each study vaccination. The duration of solicited local and systemic AEs after each study vaccination will also be summarised by treatment group.

Unsolicited AEs will be coded by preferred term and system organ class using the latest version of the MedDRA and summarised by study vaccine group as well as by severity and relationship to study vaccine. AEs through 49 days after first study vaccination for the initial set of vaccinations; all MAAEs through 35 days after first study vaccination for the initial set of vaccinations; and MAAEs related to study vaccine; SAEs; or AESI through EOS will be listed separately and summarised by study vaccine group. As solicited AEs are only being recorded in a subset of participants, it is expected that typical solicited AEs will be captured as unsolicited AEs in those who are not in the sub-study if they occur through Day 49 after the initial set of vaccinations. An assessment of the impact of this decision will be part of an exploratory analysis and will be summarised.

Separate analyses of safety events received with respect to approved or deployed SARS-CoV-2 vaccines will be performed.

Vital sign measurements will be summarised by study vaccine group using descriptive statistics at baseline and following study vaccination.

Concomitant medications will be provided in a data listing with preferred drug name as coded using the World Health Organisation drug dictionary.

##### 7.3.3.1 Safety: Study Vaccine-Associated Enhanced Disease

Continuous monitoring for study vaccine-associated enhanced disease will be performed through the CRO and sponsor medical monitors. These events will be monitored in real-time and after each confirmed respective case. The SMC will review this data at scheduled SMC meetings throughout the study or at an ad hoc meeting if the medical monitors would like a more immediate review of the data.

#### 7.4 Handling of Missing Data

For calculating geometric means and GMFR, immunogenicity values reported as below the LLOQ will be replaced by 0.5 × LLOQ. Values that are greater than the upper level of quantitation (ULOQ) will be replaced by the ULOQ. Missing results will not be imputed.

#### 7.5 Interim Analyses

Prior to the final analysis, a single interim analysis of efficacy will be conducted based on the accumulation of approximately 50% (50 events) of the total anticipated target number of the primary endpoint (100 events). For this analysis, the data needed to perform the analysis of the primary efficacy endpoint will be cleaned. The interim analysis will be performed by an unblinded Biostatistics and Programming team (PPD), and the unblinded statistician will communicate the results of the analyses to the Sponsor in terms of fulfillment or nonfulfillment of the predefined success criterion (yes/no). Novavax will be unblinded at the participant level at the time of the primary 100-event analysis. If the pre-defined success criterion of the interim analysis is unfulfilled (no), then the study will remain blinded to treatment assignment until the final analysis.

If the pre-defined success criterion of the interim analysis is fulfilled (yes), then the Sponsor may unblind selected accrued data at the treatment group level and continue the study while maintaining the blind to achieve a more robust safety and efficacy data package. The unblinded Biostatistics and Programming team (PPD) will be isolated (by firewall) from study personnel. They will complete a review independent of the study team and Sponsor. The interim analysis will follow standard group-sequential design using the Lan-DeMets alpha-spending function for Pocock boundary conditions. Table 7-2 summarises the timing, number of endpoints, and statistical success boundaries at the planned interim and final analyses.

**Table 7-2: Interim and Final Boundaries Using Pocock Spending Function**

| Planned Information Fraction (% of total endpoints) | Planned Blinded Total Number of Endpoints | Planned One-Sided Nominal Alpha | VE Boundary for LBCI > 30% |
|---|---|---|---|
| Interim analysis at 50% | 50 | 0.01550 | ~68% |
| Final analysis at 100% | 100 | 0.01387 | ~57% |

Abbreviations: LBCI = lower bound confidence interval; VE = vaccine efficacy.

If an unplanned additional interim analysis is to be added or the timing of a planned analysis is modified, the Lan-DeMets alpha-spending function will be used to adjust the nominal alphas to maintain the pre-specified overall one-sided type I error at 0.025.

#### 7.6 Pre Blinded Crossover

Prior to the blinded crossover, assessment of safety and efficacy will be made while there is a placebo-controlled comparator.

#### 7.7 Post Blinded Crossover

Following blinded crossover, follow-up to assess safety and efficacy endpoints (assessment of MAAEs related to study vaccine, SAEs, and AESIs, blood testing for SARS-CoV-2 and vaccine efficacy) will continue through study completion.

#### 7.8 Planned Analyses Prior to Study Completion

Analysis of the data will be provided on an ongoing basis for confirming success and to review safety as the study progresses. The SAP will outline the sequential nature of these reviews.

### 8. REFERENCE LIST

Bengtsson KL, Song H, Stertman L, et al. Matrix-M adjuvant enhances antibody, cellular and protective immune responses of a Zaire Ebola/Makona virus glycoprotein (GP) nanoparticle vaccine in mice. Vaccine. 2016;34(16):1927-35.

Coleman CM, Venkataraman T, Liu YV, et al. MERS-CoV spike nanoparticles protect mice from MERS-CoV infection. Vaccine. 2017;35:1586-9.

Department of Health and Human Services (DHHS), Food and Drug Administration, Center for Biologics Evaluation and Research (US). Guidance for industry: Toxicity grading scale for healthy adult and adolescent volunteers enrolled in preventive vaccine clinical trials. September 2007 [cited 2020 Apr 01]. Available from: https://www.fda.gov/media/73679/download

Division of AIDS (DAIDS), National Institute of Allergy and Infectious Diseases, National Institutes of Health, US Department of Health and Human Services. Division of AIDS (DAIDS) table for grading the severity of adult and pediatric adverse events. July 2017 [cited 2020 Apr 01]. Available from: https://rsc.niaid.nih.gov/sites/default/files/daidsgradingcorrectedv21.pdf

Follmann D, Fintzi J, Fay MP, et al. Assessing durability of vaccine effect following blinded crossover in COVID-19 vaccine efficacy trials. medRxiv 2020.12.14.20248137; doi: https://doi.org/10.1101/2020.12.14.20248137. Preprint.

Guebre-Xabier M, Patel N, Tian J-H, et al. NVX-CoV2373 vaccine protects cynomolgus macaque upper and lower airways against SARS-CoV-2 challenge. bioRxiv 2020.08.18.256578; doi: https://doi.org/10.1101/2020.08.18.256578.

Habibzadeh P, Stoneman EK. The novel coronavirus: a bird's eye view. Int J Occup Environ Med. 2020;11(2):65-71.

Keech C, Albert G, Reed P, et al, First-in-Human Trial of a SARS-CoV-2 Recombinant Spike Protein Nanoparticle Vaccine. medRxiv 2020.08.05.20168435; doi: https://doi.org/10.1101/2020.08.05.20168435.

Liu YV, Massare MJ, Barnard DL, et al. Chimeric severe acute respiratory syndrome coronavirus (SARS-CoV) S glycoprotein and influenza matrix 1 efficiently form virus-like particles (VLPs) that protect mice against challenge with SARS-CoV. Vaccine. 2011;29(38):6606 13.

Mandolesi M, Sheward DJ, Hanke L, et al. SARS-CoV-2 protein subunit vaccination elicits potent neutralizing antibody responses. bioRxiv 2020:2020.07.31.228486.

SPEAC: Safety Platform for Emergency Vaccines. D2.3 Priority List of Adverse Events of Special Interest: COVID-19. V2.0; May 25, 2020.

Su S, Wong G, Shi W, et al. Epidemiology, genetic recombination, and pathogenesis of coronaviruses. Trends Microbiol. 2016;24(6):490-502.

Tian J-H, Patel N, Haupt R, et al. SARS-CoV-2 spike glycoprotein vaccine candidate NVX-CoV2373 elicits immunogenicity in baboons and protection in mice. bioRxiv 2020:2020.06.29.178509.

World Health Organization (WHO). Coronavirus disease 2019 (COVID-19) situation report 203. 2020 August 10. Retrieved 10 August 2020, from https://www.who.int/docs/default-source/coronaviruse/situation-reports/20200810-covid-19-sitrep-203.pdf?sfvrsn=aa050308_4.

Zou G. A modified poisson regression approach to prospective studies with binary data. Am J Epidemiol. 2004;159(7):702-706.

### 9. APPENDICES

#### 9.1 Appendix 1: Protocol Change History

**Protocol Version 4.0, 25 February 2021 (revised from Version 3.0, 23 December 2020)**

The following is a summary of the changes made from Version 3.0 (23 December 2020) to Version 4.0 (25 February 2021).

| Location of change | Change/Modification |
|---|---|
| **Changes Made from Version 3.0 (December 23, 2020) to Version 4.0 (25 February 2021)**<br>The primary purpose of this amendment is to add the blinded crossover design. All changes were made because of addition of crossover design except as noted below. | |
| Synopsis; Secondary Objectives (Section 2.1.2) | • Deleted "in SARS CoV-2 seropositive" from the second objective to correct error.<br>• Added "in the initial set of vaccinations" to the 9th secondary objective per MHRA recommendation.<br>• Changed safety and reactogenicity objectives from 7 days after each study vaccination to "the initial set of study vaccinations."<br>• Added the following secondary objective: "To assess the duration of vaccine efficacy (measured by all efficacy endpoints) in initial active vaccine recipients vs. crossover (delayed) active vaccine recipients. |
| Synopsis; Exploratory Objectives (Section 2.1.3) | • Added "in the initial set of vaccinations" to the first and second exploratory objective.<br>• Added "unblinded before the crossover" to second exploratory objective. |
| Synopsis; Primary Endpoint (Section 2.2.1) and Secondary Endpoints (Section 2.2.2) | • Added "in the initial set of vaccinations" to the primary endpoint and the first 6 secondary endpoints.<br>• Changed "First occurrence of laboratory-confirmed COVID-19 to participants with negative serostatus at baseline. |
| Synopsis; Other Secondary Endpoints (Section 2.2.2.2) | • Added analysis of antibodies endpoint for Crossover Day 0 and Day 35.<br>• Deleted "for 7 days" and added "the initial set of vaccinations" to reactogenicity endpoint.<br>• Added "following the initial set of vaccinations" to MAAE and unsolicited AE endpoint.<br>• Added the following secondary endpoint: "Relative vaccine efficacy (measured by all efficacy endpoints) in initial active vaccine recipients vs. crossover (delayed) active vaccine recipients. |
| Synopsis; Exploratory Endpoints (Section 2.2.3) | • Added "in the initial set of vaccinations" to the first 2 exploratory endpoints.<br>• Added the following endpoint: "Any occurrence of serologic conversion (by serology to SARS-CoV-2 N protein) between Crossover Day 0 visit and the end of study visit in the second set of vaccinations in adult participants seronegative at baseline."<br>• Added "and Crossover Day 35 visit (14 days after the Crossover Day 21 visit" to seroconversion and VNA endpoint.<br>• Added the following endpoint: First occurrence of virologically confirmed (by PCR to SARS-CoV-2), symptomatic mild, moderate, or severe COVID-19, with onset at least 7 days after second study vaccination (e.g., Day 28) in the initial set of vaccinations in seronegative adult participants by age (<65 and ≥65) in racial and ethnic minorities and in those with co-morbid conditions. |

| | |
|---|---|
| | • Added "The occurrence, severity, and relationship to study vaccination of unsolicited AEs (in all adult participants) for 21 days after first study vaccination and 28 days after second study vaccination following the initial set of vaccinations with the adjustment to remove reactogenicity events which were recorded as unsolicited AEs within 7 days of each dose in the initial set of vaccinations." |
| Synopsis; Study Design (Section 3) | • Added description of crossover design.<br>• Added "and Crossover Day 0 visit" for nose/throat samples.<br>• Updated paragraph about participants receiving an approved vaccine to include crossover portion; added a sentence about participants who are unblinded and receive another vaccine.<br>• Added "Participants who are unblinded and ineligible for the blinded crossover will continue to follow the original study design featured in Figure 1b.<br>• Clarified that the duration of individual participation is 1 year from the initial set of vaccinations to correct error.<br>• Added information about blood draws and assessment of vaccine efficacy.<br>• Added information about unblinded participants who receive another vaccine |
| Synopsis; Study Vaccination Pause Rules (Section 3.1) | • Added "for the first set of vaccinations only" to pause rules based on reactogenicity. |
| Synopsis; Exclusion Criteria (Section 4.2) | • Added statement about inclusion/exclusion criteria at crossover. |
| Synopsis ; Study Vaccine Administration (Section 5.1) | • Added "At the time of the blinded crossover, participants will receive the alternate study material in 2 IM injections 21 + 7 days apart" to description of study vaccines.<br>• Deleted description of nose/throat samples and referred to SOE. |
| Day21 (Section 6.1.3) | • Added sentence about assessment of exclusion criteria prior to second dose. |
| COVID Surveillance Visits (Section 6.1.5.1 and 6.1.5.2) | • Added "as appropriate" to collection of concomitant medications and added collection of nose/throat samples. |
| Synopsis; Immunogenetic Assessments (Section 6.3) | • Added "Anti-S protein serology in the Anti-S Protein Serology Subset will be collected after the first and second set of vaccinations." |
| Synopsis; Safety Assessments (Section 6.4) and Local and General Systemic Reactogenicity Symptoms (Section 6.4.1.1.2) | • Added "after the first set of vaccinations" and "in the initial set of vaccinations to Safety Assessments.<br>• Added "Local and systemic reactogenicity events will not be recorded for the second set of vaccinations."<br>• Added statement about unsolicited AEs in participants who are not in the sub-study.<br>• Deleted 'virologically confirmed" to correct error.<br>• Added "who will be initially" to description of randomization.<br>• Added "unblinded participants should not be included in the blinded crossover." |
| MAAEs (Section 6.4.1.1.4) | • Added "for the first set of vaccinations only." |
| Synopsis; Analysis Sets (Section 7.2) | • Changed 7 to 6 days or less for positive SARS-CoV-2 test to correct error.<br>• Deleted serum IgG antibody and changed to antibody tests to correct error. |
| Synopsis; Efficacy Analyses (Section 7.3.1) | • Added a paragraph about primary analysis of primary and key secondary efficacy endpoints pre and post crossover. |
| Synopsis; Immunogenicity Analysis (Section 7.3.2) | • Added description of immunogenicity analysis after the second set of vaccinations.<br>• Deleted sentence about EOS analysis. |
| Synopsis; Safety Analysis (Section 7.3.3) | • Added "after the initial set of vaccinations only" to capture of solicited AEs.<br>• Added clarification about solicited AEs in participants who are not part of the sub-study. |

| | |
|---|---|
| Synopsis; Interim Analyses (Section 7.5) | • Added "Novavax will be unblinded at the participant level at the time of the primary 100-event analysis." |
| Synopsis; Pre- and Post-Blinded Crossover (Sections 7.6 and 7.7) | • Added sections on pre- and post-blinded crossover. |
| Clinical Experience (Section 1.3) | • Added sentence about 302 interim analysis. |
| Figure 1a: Trial Design | • Updated figure to portray crossover design (Figure 1a) and changed Figure 1 to Figure 1b.<br>• Added footnote to Figure 1a to describe possible consolidation of crossover visits. |
| Table 3-1: Schedule of Events | • Updated to include crossover design |
| Method of Assigning Participants (Section 5.3) | • Added sentence about the use of the IRT system. |
| Blinding Procedures (Section 5.3.1) | • Added sentence about keeping study participants and personnel blinded.<br>• Added information about reconsenting participants at crossover. |
| Concomitant Medications (Section 5.5.1) | • Added clarification about concomitant medications based on new study design. |
| Initial COVID-19 Surveillance Visit (Section 6.1.5) and Follow-Up Visit (Section 6.1.5.2) | • Added "as appropriate" to collection of prior and concomitant medications.<br>• Added paragraph about nose/throat sampling. |
| Unblinding Visit (Section 6.1.6) | • Changed instructions for unblinding visit.<br>• Added written informed consent<br>• Added "as appropriate" to collection of prior and concomitant medication.<br>• Deleted "If prior to participant's Day 21 and Day 35 visit." |
| 3 Month Visit (Section 6.1.7) | • Added a paragraph changing the requirement for the 3-month visit.<br>• Added "as appropriate" to collection of prior and concomitant medications. |
| Crossover Day 0 visit (Section 6.1.8) | • Added section |
| Crossover Day 21 visit | • Added section.<br>• Deleted 6-month visit. |
| Crossover Day 35 visit | • Added section |
| 12 months After Second Vaccination (Section 6.1.11) | • Updated instructions for EOS visit. |
| Day 35—Follow-up Visit (Section 6.1.4) | • Deleted "Visit may be skipped as per unblinding visit below." |
| Nose/Throat Samples (Section 6.2.1) | • Added Crossover Day 0 and 21 for Nose/Throat samples. |
| Virologic Confirmation of SARS-CoV-2 (Section 6.2.2 | • Deleted section |
| Initial COVID-19 Surveillance Visit | • Added "as appropriate" to ascertainment of new concomitant medications. |
| Safety Assessment (Section 6.4) | • Added "unblinded participants should not be included in the blinded crossover." |
| MAAEs (Section 6.4.1.1.4 | • Added "after the first set of vaccinations only" to MAAE collection. |
| Statistical analysis (Section 7.3) | • Changed information about management of unblinded participants. |
| Reference List (Section 8) | • Updated based on new text. |
| **Changes Made from Version 2.0 (23 October 2020) to Version 3.0 (23 December 2020)** | |

| | |
|---|---|
| Synopsis | • Updated number of study sites from 28 to 33. |
| Synopsis; Exploratory Objectives (Section 2.1.3) | • Added an objective to exploratory endpoint to explore the efficacy and safety of SARS-CoV-2rS with Matrix-M1 adjuvant with an approved or deployed vaccine to take into account the changing vaccine landscape. |
| Synopsis; Secondary Endpoints (Section 2.2.2) | • Added a secondary endpoint to measure severe COVID-19 separately. |
| Synopsis; Exploratory Endpoints (Section 2.2.3) | • Added 2 exploratory endpoints, one due to changes in vaccine landscape and one to address asymptomatic disease. |
| Synopsis; Study Design (Section 3) | • Added "approximately" to description of 400 participants in substudy to allow for greater flexibility. |
| Synopsis; Section 1 (Introduction); Section 3 (Study Design); Section 5.3.2 (Breaking the Blind) | • Added advice from regulatory agencies concerning unblinding of treatment assignment for participants receiving an approved or deployed SARS-CoV-2 vaccine during the study. |
| Synopsis, Schedule of Events, Section 6.1.5.1 (Initial COVID-19 Surveillance Visit); Follow-up COVID-19 Surveillance Visit (Section 6.1.5.2); and 6.2.3.2 (COVID-19 Surveillance Visit [Initial and Follow-up]) | • Added information about replacing in-person COVID-19 Surveillance Visits with phone calls if government restrictions are in place. Also added "with exception per protocol" to accommodate this new recommendation. |
| Synopsis, Section 7.1 (Sample Size), Table 7-1 | • Reduced number of target endpoints from 152 to 100 based on higher than predicted vaccine efficacy observed in the recent Phase 3 trials by Pfizer and Moderna. This reduced number was chosen to provide > 95% power for 70% or higher vaccine efficacy.<br>• Reduced number of planned interim analyses from 2 to 1 and revised timing of the single interim and final analyses of the primary efficacy endpoint to 50 and 100 events, respectively. |
| Synopsis, Section 7.2 (Analysis Sets) | • Clarified the wording of the PP-EFF analysis set. |
| Synopsis, Section 7.3.1 (Efficacy Analysis) | • Clarified the wording of the primary efficacy analyses at the interim and final analyses of the primary efficacy endpoint. |
| Synopsis, Section 7.3.2 (Immunogenicity Analysis and Correlates of Risk) | • Changed HAI to ELISA for SARS-CoV-2 rS serum antibody levels to correct error and added influenza vaccine to later sentences for greater clarity. |
| Synopsis, Section 7.5 (Interim Analysis); Table 7-2 | • Reduced the number of interim analyses for the primary efficacy endpoint from 2 to 1, with the timing of the single interim analysis based on the accumulation of approximately 50% (50 events).<br>• Clarified that the unblinded Biostatistics and Programming team is from the CRO, PPD.<br>• Clarified the wording of the single interim and final analyses of the primary efficacy endpoint. |
| Synopsis (2 times); Section 6.4 (Safety | • Added "Separate analyses of safety events received with respect to approved or deployed SARS-CoV-2 vaccines will be performed" to |

| | |
|---|---|
| Assessments); Section 7.3.3. (Safety Analyses) | synopsis. Added specific information about safety reporting for participants who receive an approved or deployed SARS-CoV-2 vaccine. |
| Introduction | • Added a paragraph about changes due to approved or deployed SARS-CoV-2 vaccines. |
| Section 2.2.1 (Primary Endpoints); Table 2-1; Section 6.2.3 (Severity of COVID-19 Symptoms) | • Added the following footnote: "Participants with vital sign abnormalities in the moderate or severe categories must meet the criteria for mild COVID-19" and added the same information in text. |
| Section 2.2.1 (Primary Endpoints); Table 2-2 | • Removed duration of ≥ 48 hours for headache, nausea, vomiting, and diarrhea for consistency with other SARS-CoV-2 rS trials. |
| Section 4.4.1 (Reasons for Withdrawal) | • Added a statement that vaccination with an approved or deployed SARS-CoV-2 vaccine does not constitute a withdrawal, this is based on MHRA guidance. |
| Section 5.3.1 (Blinding Procedures); Section 5.3.2 (Breaking the Blind) | • Added specific information on unblinding if participants wish to receive an approved or deployed SARS-CoV-2 vaccine. |
| Section 5.5.1 (Concomitant Medications) | • Added that participants will be asked to record the date and brand of the approved or deployed vaccine that they receive. |
| Section 5.5.2 (Prohibitive Therapy) | • Added an exception to vaccine restrictions for participants who wish to receive an approved or deployed vaccine at least 21 days after study vaccine. |
| Section 6.1 (Study Visit Procedures) | • Added "Visit may be skipped as per the Unblinding Visit described below" in Sections 6.1.3 and 6.1.4.<br>• Clarified that a targeted physical examination may be performed if participant has any ongoing complaints in Section 6.1.4.<br>• Clarified that blood sampling for SARS-CoV-2 (ELISA for anti-N-protein serology) in all participants "unless the participant has had an Unblinding Visit in the prior 14 days where anti-N serology was drawn" in Sections 6.1.4, 6.1.7, and 6.1.8. |
| Section 6.1.5.1 (Initial COVID-19 Surveillance Visit) | • Added "including those who have been unblinded" to list of requirements at this visit. |
| Section 6.1.6 (Unscheduled Blinding Visit) | • Added section to describe unblinding process for people who wish to receive approved or deployed SARS-CoV-2 vaccine. |
| Section 6.2.2 (Virologic Confirmation of SARS-CoV-2) | • Clarified that nose/throat self-sampling may be skipped is a participant is known to be PCR positive to SARS-CoV-2 from the Screening or Day 0 PCR within 14 days of the positive test.<br>• Clarified that if a participant obtains a PCR test for SARS-CoV-2 outside of the study for any other reason than having suspected COVID-19 symptoms, then this result should not be noted or reported.<br>• Clarified that if a participant obtains a PCR test for SARS-CoV-2 outside of the study for suspected COVID-19 symptoms, then the participant should begin all protocol-required assessments for participants with suspected COVID-19 symptoms. |
| Section 6.2.3 (Monitoring for Suspected COVID-19) | • Added "This is the case for blinded and unblinded participants." |

| | |
|---|---|
| Section 6.2.3.2 (COVID-19 Surveillance Visit) | • Added "See Section 6.1.5.1 for exceptions for participants affected by extenuating circumstances (e.g., local government restrictions). |
| Section 6.2.3.2.1 (Initial COVID-19 Surveillance Visit) | • Added "Proper documentation in the eCRF of the dates, bar codes, and results of all self-swabs taken in proximity of this visit." |
| Section 6.4 (Safety Assessments) | • Added safety requirements for unblinded patients. |
| Section 6.4.1.1.2 (Local and General Systemic Reactogenicity Symptoms) | • Added "Solicited AEs will not be captured for any approved or deployed SARS-CoV-2 vaccine." |
| Section 6.4.1.3 (Reporting Adverse Events) | • Added information about SAE reporting for participants who receive an approved or deployed vaccine. |
| Section 7.3 (Statistical Analysis) | • Added "In order to manage unblinded participants, unblinding will result in censoring of all efficacy and immunogenicity endpoints. The main presentation of data based on the Safety Analysis set will include all participant data, with supporting presentations to exclude the data post unblinding. Sensitivity analysis of reactogenicity and unsolicited AEs will be conducted for the Safety analysis subset of participants not unblinded. Additional analysis of AEs collected after unblinding may be undertaken as exploratory safety analyses. ." |

Abbreviations: COVID-19 = coronavirus disease 2019; CRO = contract research organisation; ELISA = enzyme-linked immunosorbent assay; HAI = hemagglutination inhibition assay; PCR = polymerase chain reaction; PP-EFF = per-protocol efficacy; SARS-CoV-2 = severe acute respiratory syndrome coronavirus 2; SARS-CoV-2 rS = severe acute respiratory syndrome coronavirus 2 recombinant spike nanoparticle vaccine.

#### Protocol Version 2.0, 23 October 2020 (revised from Version 1.0, 17 September 2020; Version 1.1, 17 September 2020; and Version 1.2, 21 September 2020

The following is a summary of the changes made from Version 1.0 (24 August 2020) to Version 1.1 (17 September 2020), Version 1.1 (17 September 2020) to Version 1.2 (21 September 2020), and Version 1.2 (21 September 2020) to Version 2.0 (23 October 2020).

| Location of Change | Change/Modification |
|---|---|
| **Changes Made from Version 1.0 (17 September 2020) to Version 1.1 (17 September 2020)** | |
| Section 4.4.1 (Reasons for Withdrawal) | The following changes were made based on MHRA recommendations:<br>• Changed "may withhold" to "will withhold" further vaccinations.<br>• Deleted "The investigator will withhold further study vaccination from a participant in the study if the participant" prior to the bullet regarding pregnancy to indicate that all numbered items are required to withhold vaccination.<br>• Deleted "Upon the occurrence of an SAE or intolerable AE, the investigator may confer with the sponsor before future study vaccination" for more restrictive requirements. |
| **Changes Made from Version 1.1 (17 September 2020) to Version 1.2 (21 September 2020)** | |
| Synopsis (Study Vaccination Pause Rules); Section 3.1 | • Added: "for example, any SAE for which causality is at least possibly related" for greater clarity/MHRA recommendation. |

| Location of Change | Change/Modification |
|---|---|
| Section 6.1.4 [Day 35 – Follow-up Visit (+ 7 days)] | • Deleted “prior to study vaccination” from the blood sampling procedures on Day 35 since vaccination is only given on Days 0 and 21 (MHRA recommendation). |
| Section 6.1.6 [3 Months (± 15 days) After Second Study Vaccination] | • Deleted “prior to study vaccination” from the blood sampling procedure on Month 3 since vaccination is only given on Days 0 and 21 (MHRA recommendation). |
| Section 6.1.7 [6 Months (± 15 days) After Second Study Vaccination] | • Deleted “prior to study vaccination” from the blood sampling procedure on Month 6 since vaccination is only given on Days 0 and 21 (MHRA recommendation). |
| Section 6.1.8 [12 Months (± 15 days) After Second Study Vaccination] | • Deleted “prior to study vaccination” from the blood sampling procedure on Month 12 since vaccination is only given on Days 0 and 21 (MHRA recommendation). |
| **Changes Made from Version 1.2 (21 September 2020) to Version 2.0 (23October 2020)** | |
| Synopsis | • Changed “up to 18 regions” to “28 sites” across the United Kingdom (UK) based on updated information. |
| Synopsis (Secondary Objectives); Section 2.1.2 | • Added “related to study vaccination” to 6th secondary objective for clarity.<br>• Deleted “and in terms of unsolicited AEs for 21 days after first study vaccination and 28 days after second study vaccination” and added a separate bullet for MAAEs for greater clarity. |
| Synopsis (Primary Endpoints); Section 2.2.1 | • Changed to only one primary endpoint and moved the second primary endpoint to key secondary endpoint<br>• Added “mild, moderate, or severe” to COVID-19 description in primary endpoint for clarity |
| Synopsis (Secondary Endpoints); Section 2.2.2 | • Added “mild, moderate, or severe” to COVID-19 description to 2nd secondary endpoint for clarity.<br>• Added “symptomatic” to description of mild COVID19 to 4th primary endpoint for clarity.<br>• Added N-protein to serology endpoint for greater clarity.<br>• Eliminated ELISA testing from Day 21, as this was an oversight from a prior version.<br>• Added “related to study vaccination” to endpoint for SAEs and MAAEs for greater clarity.<br>• Added 3 safety endpoints (i.e., AESIs/PIMMCs, solicited AEs, and unsolicited AEs) to correlate with secondary objectives. |
| Synopsis (Exploratory Endpoints); Section 2.2.3 | • Added “mild, moderate, or severe” to COVID-19 description to first exploratory endpoint |
| Synopsis (Study Design); Section 3 | • Changed “regions” to “sites” in 2 places for clarity.<br>• Changed mucosal to nose/throat and eliminated saliva samples for COVID-19 at Screening and Day 0 due to lack of saliva test availability in UK.<br>• Changed study population from 9000 to 15,000 to increase accrual rate of COVID-19 endpoints.<br>• Deleted information about enrolment of participants ≥age 65 and operational cutoff based on updated information. |
| Synopsis (Table S1-1) | • Changed both groups from 4,500 to 7,500 to accommodate increased enrolment. |

| Location of Change | Change/Modification |
|---|---|
| Synopsis (Study Vaccination Pause Rules); Section 3.1 | • Allowed the SMC chair to review SAEs and decide on the need for a full SMC meeting before starting a study enrolment pause.<br>• Expanded requirement for solicited and unsolicited AEs to also include Grade 4 (potentially life-threatening), in addition to Grade 3, for greater clarity.<br>• Added "after a minimum of 100 participants were enrolled" to 5% and 10% cutoffs for solicited and unsolicited AEs to prevent a pause being triggered due to an early cluster of grade 3 events.<br>• Changed grade 3 (severe) to severe and added "after a minimum of 100 subjects are enrolled" to third bullet for clarity.<br>• Added "for example, any SAE for which causality is at least possibly related" to provide more clarity to pause rule.<br>• Added specific text about analysis of grade 3 or higher solicited AEs in 400-subject influenza substudy for greater clarity. |
| Synopsis (Inclusion Criteria); Section 4.1 | • Deleted "with spermicide" from condom criteria due to input from the UK ethics review.<br>• Added "Other approaches to abstinence are not acceptable" to clarify how this method can be used as contraception due to input from the UK ethics review. .<br>• Added "Room saturation of > 95% at Screening/Day 0 to inclusion criteria as this is aligned with the severity grading criteria which assigns a greater level of severity below 95%. |
| Synopsis (Exclusion Criteria); Section 4.2 | • Changed participation in serologic surveys to future participation to allow participation to those who participated prior to enrollment.<br>• Added text that allowed participants with controlled HIV to participate per investigator's suggestion.<br>• Added "within 3 months following the last study vaccine" to pregnancy/lactation criteria participate per investigator's suggestion.<br>• Added additional information to restrictions on administration of influenza vaccine for greater clarity.<br>• Added "including hepatitis B and C" to hepatic exclusion for greater clarity per investigator's suggestion.<br>• Deleted "and epilepsy" from neurological disorders per investigator's suggestion.<br>• Deleted "or immunodeficiency" and added Table 9-3 (specific conditions) and the need for biologic therapy for greater clarity as it was repetitious of a prior exclusion.<br>• Added "The Skin and Metabolic disorders listed in Table 9-3 are eligible at the discretion of the investigator" and eliminated caveat about endocrine disorders.<br>• Added the use of continuous oxygen therapy as an Exclusion Criteria (allowing for nocturnal oxygen) as it would not be possible to grade severity levels based on oxygen saturation if someone is on continuous oxygen.<br>Other Considerations:<br>Added a bullet point addressing participants with hypertension. |
| Synopsis (Study Vaccine Administration); Section 5.1 | • Added information about specific flu vaccines for participants 18-64 years of age and those ≥ 65 years of age. |

| Location of Change | Change/Modification |
|---|---|
| | • Added instructions for using the right deltoid for flu vaccine and the left deltoid for study vaccine on Day 0 whenever possible to allow for the majority of right handed people to more easily access the study vaccine injection site. |
| Synopsis (Efficacy Assessments); Section 6.1.1, 6.1.2, and 6.1.3 | • Replaced mucosal with nose/throat and removed "or saliva" from testing method for efficacy assessments due to lack of saliva test availability in UK. |
| Synopsis (Safety Assessments); Section 6.4 | • Added "Participants in the seasonal influenza vaccine co-administration substudy will record local reactogenicity for the study vaccine injection site only" as the symptom diary could not accommodate the measurement of 2 injection sites. |
| Synopsis (Statistical Analysis Plan); Section 7.1 | • Changed sample size from 9000 to 15,000 to increase the rate of endpoint accumulation.<br>• Updated the statistical analysis plan based on increased enrolment.<br>• Deleted information designed about VE—information moved to Efficacy Section 7.3.1.<br>• Deleted information about sample size based on updated information. |
| Synopsis (Analysis Sets); Section 7.2 | • Defined the all randomized set and ITT set—deleted ITT-EFF and ITT-IMM analysis sets for further clarity.<br>• Added "baseline" to describe seronegative participants in the PP-EFF set.<br>• Added "that occur before the first COVID-19 episode" to the restriction for no major protocol deviations for the PP-EFF population for greater clarity.<br>• Changed 14-day exclusion to 7 days for positive SARS-CoV-2 illness episodes occurring after the second vaccination (i.e., Day 35 to Day 28) for PP-EFF population analysis as this was a correction.<br>• Changed Day 21 or Day 35 to Day 35 only for PP-IMM analysis as this was a correction.<br>• Added "Both PP populations will be analysed according to the study vaccine group as randomized" for further clarity. |
| Synopsis (Efficacy Analysis); Section 7.3.1 | • Changed 2 key decision points to 4 potential decision points and described them for greater clarity.<br>• Changed ITT-EFF population to ITT analysis group.<br>• Added extensive statistical information about planned analyses for greater clarity.<br>• Moved EOS analysis to Immunogenicity section<br>• Deleted reference to SAP because of expanded text.<br>• VE definition was moved here. |
| Synopsis (Immunogenicity Analysis); Section 7.3.2 | • Removed reference to ITT-IMM population—changed to ITT. Added further statistical details to the immunogenicity analysis for the influenza sub-study for greater clarity.<br>• Moved EOS analysis in Neutralisation Assay subset to this section |
| Synopsis (Safety Analysis); 7.3.3 | • Changed AE reporting window to 49 days from 35.<br>• Added "through 35 days after first study vaccination" for greater clarity. |
| Synopsis (Interim Analysis); Section 7.5 | • Added new section describing statistical information about planned interim analysis. These were added to potentially increase the speed of detecting vaccine efficacy during this global pandemic. |

| Location of Change | Change/Modification |
|---|---|
| Section 1.2.1 (Nonclinical Data) | • Updated study data. |
| Section 1.3 (Clinical Experience) | • Updated Part 1 data from Phase 1/2 study and added information about Part 2.<br>• Specified number of healthy participants as 131. |
| Section 1.4 (Rationale for Study) | • Added information about results of 5-day reactogenicity data from the Phase 2 (Part 2) study.<br>• Deleted information about enrolment of participants ≥ age 65. |
| Section 2.2.1 (Table 2-1) | • Deleted reference to virologically confirmed COVID-19. |
| Section 2.2.2 (Table 2-2) | • Updated information about qualifying symptoms of COVID-19 to clarify trigger should be based on new onset of symptoms. |
| Section 3 (Study Design); Figure 1 | • Updated trial schema to include 15,000 participants to reflect increased enrolment. |
| Table 3-1 (Schedule of Events) | • Changed mucosal to nose/throat for greater clarity.<br>• Footnote c: Changed EOS telephone call to visit to correct an error.<br>• Footnote l: Added to provide further clarity on nose/throat testing.<br>• Footnote z: Edited for clarity that swabbing refers to self collection by participants. |
| Section 4 (Study Population) | • Updated introduction from 9000 to 15,000 participants to increase the rate of endpoint accumulation. |
| Section 5.2 (Investigational Products) | • Added 2 choices of flu vaccine depending on age group to table and bullet points. This information was just made available. |
| Section 5.4 (Study Vaccine Compliance) | • Deleted information about home study vaccinations, as study sites were not able to perform home vaccinations. |
| Section 5.5.2 Prohibitive Therapy | • Added information to bullet point about administration of influenza vaccine for greater clarity. |
| Section 6 (Study Procedures) | • Deleted information about home vaccinations as study sites were not able to perform home vaccinations. |
| Section 6.1.5.1 (Initial COVID-19 Surveillance Visit) | • Deleted requirement for mucosal samples at this visit.<br>• Changed target physical examination to respiratory from lymphatic and thus removed "of injected upper extremity." This was done at Surveillance visits to stress the importance of the respiratory exam for participants with possible COVID and not the need to assess the vaccination site unless symptomatic. |
| Section 6.1.5.2 (Follow-up Surveillance Visit) | • Deleted requirements for mucosal samples at this visit as these samples have a very high capture rate of RNA when shedding.<br>• Changed target physical examination to respiratory from lymphatic and thus removed "of injected upper extremity." This was done at Surveillance visits to stress the importance of the respiratory exam for participants with possible COVID and not the need to assess the vaccination site unless symptomatic. |
| Section 6.2.1 (Nose/Throat Samples for Virus Detection) | • Changed mucosal to nose/throat and removed option for saliva testing for virus detection based on test availability in UK. |
| Section 6.2.2 (Virologic Confirmation) | • Changed mucosal to nose/throat and removed option for saliva testing for virus detection. |

| Location of Change | Change/Modification |
|---|---|
| | • Deleted HCP sampling option for mucosal sampling as these samples have a very high capture rate of RNA when shedding. |
| Section 6.2.3.2.1 (Initial COVID-19 Surveillance Visit) | • Deleted "or the participant's basal level of chronic supplemental oxygen use" from resting respiratory rate, eliminating the use of supplemental oxygen. This is aligned with the exclusion of those on basal oxygen treatment.<br>• Deleted reference to nasal turbinate swab. |
| Section 6.2.3.2.2 (Follow-up COVID-19 Surveillance Visit) | • Changed mucosal to nose/throat and removed option for saliva testing for virus detection due to saliva test availability in UK.<br>• Deleted "however, a repeat mucosal sampling will NOT be obtained since the participant has already tested positive." This was no longer needed, as sampling for this visit was removed.<br>• Deleted paragraph reference nose/throat samples |
| Section 6.4.1.1.2 (Local and General Systemic Reactogenicity Symptoms) | • Added "of study vaccine" after injection for greater clarity.<br>• Added "Local reactogenicity symptoms should not be recorded for the influenza vaccine injection site." |
| Section 6.4.14 (Medically Attended Adverse Events) | • Added "potentially life threatening (Grade 3)" to AE criteria |
| Section 6.4.2 (Vital Sign Measurements) | • Deleted text referring to supplemental oxygen due to new exclusion criteria. |
| Section 6.4.3 (Physical Examinations) | • Added Day 0 to physical exam.<br>• Special attention should made to examine the lymph nodes of the upper extremities on vaccination days and the respiratory system at all Surveillance visits. |
| Section 7.1 (Sample Size Calculations and Table 7-1) | • Updated enrolment from 9000 to 15,000 participants.<br>• Changed number of events based on increased enrollment.<br>• Updated text to match new enrolment and revised statistical analysis. |
| Section 7.1 (Table 7-2) | • Revised table based on revised enrolment and statistical analysis.<br>• Deleted information about increasing precision around VE |
| Section 7.5 (Interim Analysis; Tables 7-3 and 7-4) | • Revised numbers in tables and added text about an interim analysis. |
| Appendix 1 | • Added Protocol Change History |
| Appendix 2 | • Updated List of Abbreviations to accommodate text changes |
| Table 9-4 | • Added a footnote: "To be recorded as AESIs relevant to COVID-19, these complications should be associated with a positive PCR test for SARS-CoV-2" for greater clarity. |

Abbreviations: AE = adverse event; AESI = adverse event of special interest; COVID-19 = coronavirus disease 2019; ELISA = enzyme-linked immunosorbent assay; EOS = end of study; HIV = human immunodeficiency virus; IM = intramuscular; ITT = intent-to-treat; ITT-EFF = intent-to-treat efficacy; ITT-IMM = intent-to-treat immunogenicity; MAAE = medically attended adverse event; MHRA = Medicines and Healthcare products Regulatory Agency; N-protein = nucleocapsid; PCR = polymerase chain reaction; PIMMC = potential immune-mediated medical conditions; PP = per-protocol; PP-EFF = per-protocol efficacy; PP-IMM = per-protocol immunogenicity; SAE = serious adverse event; SAP = statistical analysis plan; SARS-CoV-2 = severe acute respiratory syndrome coronavirus 2; SMC = safety monitoring committee; UK = United Kingdom; VE = vaccine efficacy.

#### 9.2 Appendix 2: List of Abbreviations

| Abbreviation | Term |
|---|---|
| ACE2 | Angiotensin-converting enzyme 2 |
| AE | Adverse event |
| AESI | Adverse event(s) of special interest |
| ANCOVA | Analysis of covariance |
| CFR | Code of Federal Regulations |
| CI | Confidence interval |
| COVID-19 | Coronavirus disease 2019 |
| CRO | Clinical research organization |
| CT | Computed tomography |
| DHSC | Department of Health and Social Care |
| EBOV GP | Ebolavirus glycoprotein |
| eCRF | Electronic case report form |
| EDC | Electronic data capture |
| ELISA | Enzyme-linked immunosorbent assay |
| ELISpot | Enzyme-linked immune absorbent spot |
| EOS | End of study |
| FDA | United States Food and Drug Administration |
| GCP | Good Clinical Practice |
| GLP | Good Laboratory Practice |
| GMEU | Geometric mean ELISA unit |
| GMFR | Geometric mean fold rise |
| GMT | Geometric mean titre |
| GP | Glycoprotein |
| hACE2 | Human angiotensin-converting enzyme 2 |
| HAI | Hemagglutination inhibition assay |
| HEENT | Head, eyes, ears, nose, and throat |
| HIV | Human immunodeficiency syndrome |
| IB | Investigator's Brochure |
| ICF | Informed consent form |
| ICH | International Council for Harmonisation of Technical Requirements for Pharmaceuticals for Human Use |
| ICU | Intensive care unit |
| ID | Identification |

| Abbreviation | Term |
|---|---|
| IgG | Immunoglobulin G |
| IM | Intramuscular |
| IRT | Interactive Response Technology |
| ITT | Intent-to-treat |
| LBCI | Lower bound confidence interval |
| LLOQ | Lower limit of quantification |
| LRTI | Lower respiratory tract infection |
| MAAE | Medically attended adverse event |
| MedDRA | Medical Dictionary for Regulatory Activities |
| MERS | Middle Eastern Respiratory Syndrome |
| MHRA | Medicines and Healthcare products Regulatory Agency |
| NHP | Nonhuman primate |
| NHS | National Health Service |
| N-protein | Nucleocapsid |
| NZW | New Zealand White |
| OTC | Over-the-counter |
| PCR | Polymerase chain reaction |
| PHEIC | Public health emergency of international concern |
| PIMMC | Potential immune-mediated medical conditions |
| PP | Per-protocol |
| PP-EFF | PP efficacy |
| PP-IMM | PP immunogenicity |
| RR | Relative risk |
| RSV F | Respiratory syncytial virus fusion (protein) |
| S-protein | Spike |
| SAE | Serious adverse event |
| SAP | Statistical analysis plan |
| SARS-CoV | Severe acute respiratory syndrome coronavirus |
| SARS-CoV-2 | Severe acute respiratory syndrome coronavirus 2 |
| SARS-CoV-2 rS | SARS-CoV-2 recombinant spike (S) protein nanoparticle vaccine |
| SCR | Seroconversion rate |
| Sf9 | *Spodoptera frugiperda* (insect cells) |
| SMC | Safety Monitoring Committee |
| SOE | Schedule of Events |

| Abbreviation | Term |
|---|---|
| SUSAR | Suspected Unexpected Serious Adverse Reaction |
| UK | United Kingdom |
| ULOQ | Upper limit of quantitation |
| VE | Vaccine efficacy |
| VLP | Virus-like particle |
| VNA | Virus neutralisation assay |
| WHO | World Health Organisation |
| ZIKA EnvD | Zika virus envelope dimers |

### 9.3 Appendix 3: Study Governance

#### 9.3.1 Data Quality Assurance

This study will be conducted using the quality processes described in applicable procedural documents. The quality management approach to be implemented will be documented and will comply with current International Council for Harmonisation of Technical Requirements for Pharmaceuticals for Human Use (ICH) guidance on quality and risk management. All aspects of the study will be monitored for compliance with applicable government regulatory requirements, current Good Clinical Practice (GCP) guidelines, the protocol, and standard operating procedures. The monitor will maintain current personal knowledge of the study through observation, review of study records and source documentation, and discussion of the conduct of the study with the investigator and personnel. eCRFs and EDC will be utilised. The EDC system is validated and compliant with US Title 21 Code of Federal Regulations (CFR) Part 11 and local regulations. Each person involved with the study will have an individual identification code and password that allows for record traceability.

Important protocol deviations, should they occur during the study, will be presented in Section 10.2 of the clinical study report.

#### 9.3.2 Investigator Obligations

The following administrative items are meant to guide the investigator in the conduct of the study and may be subject to change based on industry and government standard operating procedures, working practice documents, or guidelines. Changes will be reported to the Research Ethics Committee (REC) but will not result in protocol amendments.

##### 9.3.2.1 Confidentiality

All laboratory specimens, evaluation forms, reports, and other records will be identified in a manner designed to maintain participant confidentiality. All records will be kept in a secure storage area with limited access. Clinical information will not be released without the written permission of the participant, except as necessary for monitoring and auditing by the sponsor, its designee, relevant regulatory authority(ies), or the REC.

The investigator and all employees and co-workers involved with this study may not disclose or use for any purpose other than performance of the study, any data, record, or other unpublished, confidential information disclosed to those individuals for the purpose of the study. Prior written agreement from the sponsor or its designee must be obtained for the disclosure of any said confidential information to other parties.

###### 9.3.2.2 Institutional Review

Prior to initiation of a study site, regulatory authority regulations and the ICH E6(R2) guidelines require that approval be obtained from the REC before participation of human participants in research studies. Before study onset, the protocol, informed consent, advertisements to be used for the recruitment of study participants, and any other written information regarding this study to be provided to the participant must be approved by the REC. Documentation of all REC approvals and of the REC compliance with the ICH E6(R2) guidelines will be maintained by the study site and will be available for review by the sponsor or its designee.

All REC approvals should be signed by the REC chairman or designee and must identify the REC name and address, the clinical protocol by title or protocol number or both and the date approval or a favourable opinion was granted.

###### 9.3.2.3 Participant Consent

Written informed consent in compliance with US Title 21 CFR Part 50 and local regulatory authority requirements shall be obtained from each participant before he or she enters the study or before any unusual or nonroutine procedure that involves risk to the participant is performed. If any institution-specific modifications to study-related procedures are proposed or made by the study site, the consent should be reviewed by the sponsor or its designee or both before REC submission. Once reviewed, the investigator will submit the ICF to the REC for review and approval before the start of the study. If the ICF is revised during the course of the study, all active participating participants must sign the revised form.

Before recruitment and enrolment, each prospective participant will be given a full explanation of the study and be allowed to read the approved ICF. Once the investigator is assured that the participant understands the implications of participating in the study, the participant will be asked to give his or her consent to participate in the study by signing the ICF.

The investigator or designee will provide a copy of the ICF to the participant. The original form shall be maintained in the participant's medical records at the study site.

###### 9.3.2.4 Study Reporting Requirements

By participating in this study, the investigator agrees to submit reports of SAEs according to the timeline and method outlined in this protocol. In addition, the investigator agrees to submit annual reports to his or her REC as appropriate.

###### 9.3.2.5 Financial Disclosure and Obligations

The investigator is required to provide financial disclosure information to allow the sponsor to submit the complete and accurate certification or disclosure statements required under US Title 21 CFR Part 54 and local regulations. In addition, the investigator must provide to the sponsor a commitment to promptly update this information if any relevant changes occur during the course of the investigation and for 1 year following the completion of the study.

Neither the sponsor nor PPD nor the study site is financially responsible for further testing or treatment of any medical condition that may be detected during the screening process. In addition, in the absence of specific arrangements, neither the sponsor nor PPD nor the study site is financially responsible for further treatment of the disease under study.

###### 9.3.2.6 Investigator Documentation

Prior to beginning the study, the investigator will be asked to comply with ICH E6(R2) Section 8.2, US Title 21 of the CFR, and local regulations by providing essential documents, including but not limited to, the following:

- REC approval.
- An original investigator-signed investigator agreement page of the protocol.
- Curriculum vitae for the principal investigator and each sub-investigator. Current licensure must be noted on the curriculum vitae. They will be signed and dated by the principal investigators and sub-investigators at study start-up, indicating that they are accurate and current.
- Financial disclosure information to allow the sponsor to submit complete and accurate certification or disclosure statements required under US Title 21 CFR Part 54 and local regulations. In addition, the investigators must provide to the sponsor a commitment to promptly update this information if any relevant changes occur during the course of the investigation and for 1 year after the completion of the study.
- An REC-approved ICF, samples of study site advertisements for recruitment for this study, and any other written information about this study that is to be provided to the participant.
- Laboratory certifications and reference ranges for any local laboratories used by the study site, in accordance with US Title 42 CFR Part 493 and local regulations.

###### 9.3.2.7 Study Conduct

The investigator agrees that the study will be conducted according to the principles of ICH E6(R2). The investigator will conduct all aspects of this study in accordance with all national, state, and local laws or regulations. The study will be conducted in compliance with the protocol, current GCP guidelines – adopting the principles of the Declaration of Helsinki – and all applicable regulatory requirements.

Prior to study initiation, the protocol and the informed consent documents will be reviewed and approved by the sponsor and an appropriate ethics committee. Any amendment to the protocol or consent materials must also be approved by the study sponsor and REC and must be submitted, notified, or approved to the regulatory authority, as required, before they are implemented.

###### 9.3.2.8 Case Report Forms and Source Documents

Site personnel will maintain source documentation, enter participant data into the eCRF as accurately as possible, and will rapidly respond to any reported discrepancies.

eCRFs and EDC will be utilised. The EDC system is validated and compliant with US Title 21 CFR Part 11 and local regulations. Each person involved with the study will have an individual identification code and password that allows for record traceability. Thus, the system, and any subsequent investigative reviews, can identify coordinators, investigators, and individuals who have entered or modified records, as well as the time and date of any modifications. There may be an internal quality review audit of the data and additional reviews by the clinical monitor.

Each eCRF is presented as an electronic copy, allowing data entry by site personnel, who can add and edit data, add new participants, identify and resolve discrepancies, and view records. This system provides immediate direct data transfer to the database, as well as immediate detection of discrepancies, enabling site coordinators to resolve and manage discrepancies in a timely manner.

Paper copies of the eCRFs and other database reports may be printed and signed by the investigator. This system provides site personnel, monitors, and reviewers with access to hardcopy audits, discrepancy reviews, and investigator comment information.

###### 9.3.2.9 Adherence to Protocol

The investigator agrees to conduct the study as outlined in this protocol, in accordance with ICH E6(R2) and all applicable guidelines and regulations.

###### 9.3.2.10 Reporting Adverse Events

By participating in this study, the investigator agrees to submit reports of SAEs according to the timeline and method outlined in this protocol. In addition, the investigator agrees to submit annual reports to his or her REC as appropriate. The investigator also agrees to provide the sponsor with an adequate report, if applicable, shortly after completion of the investigator’s participation in the study.

##### 9.3.2.11 Investigator's Final Report

Upon completion of the study, the investigator, where applicable, should inform the institution; the investigator/institution should provide the REC with a summary of the study's outcome and the sponsor and regulatory authority(ies) with any reports as required. Interim reports are expected to be provided to regulatory authorities to allow study vaccine development advancement given the pandemic situation. These reports are planned to be aggregate and at the study vaccine level unless the SMC deems additional data at the individual level (e.g., select listings of select participants) will be beneficial. In such a case, a firewall will be in place to maintain the blind for those individuals involved in the study conduct to ensure unbiased assessment continue.

##### 9.3.2.12 Records Retention

Essential documents should be retained until at least 2 years after the last approval of a marketing application in an ICH region and until there are no pending or contemplated marketing applications in an ICH region or until at least 2 years have elapsed since the formal discontinuation of clinical development of the study vaccine or per local regulation, whichever is longer. These documents should be retained for a longer period, however, if required by applicable regulatory requirements or by an agreement with the sponsor. It is the sponsor's responsibility to inform the investigator/institution as to when these documents no longer need to be retained.

##### 9.3.2.13 Publications

After completion of the study, the data may be considered for reporting at a scientific meeting or for publication in a scientific journal. In these cases, the sponsor will be responsible for these activities and will work with the investigators to determine how the manuscript is written and edited, the number and order of authors, the publication to which it will be submitted, and any other related issues. The sponsor has final approval authority over all such issues.

Data are the property of the sponsor and cannot be published without their prior authorisation, but data and any publication thereof will not be unduly withheld.

##### 9.3.3 Study Management

###### 9.3.3.1 Monitoring

###### 9.3.3.1.1 Monitoring of the Study

The clinical research organisation clinical monitor, as a representative of the sponsor, is obligated to follow the study closely. In doing so, the monitor will visit the investigator and study site at periodic intervals in addition to maintaining necessary telephone and email contact. The monitor will maintain current personal knowledge of the study through observation, review of study records and source documentation, and discussion of the conduct of the study with the investigator and personnel. The monitor will be blinded to study vaccine assignment. A separate unblinded study monitor will be responsible for drug accountability.

All aspects of the study will be carefully monitored by the sponsor or its designee for compliance with applicable government regulation with respect to current ICH E6(R2) guidelines and standard operating procedures.

###### 9.3.3.1.2 Inspection of Records

The investigator and institution involved in the study will permit study-related monitoring, audits, REC review, and regulatory inspections by providing direct access to all study records. In the event of an audit, the investigator agrees to allow the sponsor, their representatives, or the regulatory authority access to all study records.

The investigator should promptly notify the sponsor of any audits scheduled by any regulatory authorities and promptly forward copies of any audit reports received to the sponsor.

###### 9.3.3.2 Management of Protocol Amendments and Deviations

###### 9.3.3.2.1 Modification of the Protocol

This is a Phase 3 study to evaluate the efficacy, immunogenicity, and safety of SARS-CoV-2 rS with Matrix-M1 adjuvant. This protocol is written with some flexibility to accommodate the evolving pandemic and urgency for efficacious vaccine availability. Modifications to the dose, dosing regimen, and/or clinical or laboratory procedures currently outlined below may be required to achieve the scientific goals of the study objectives and/or to ensure appropriate safety monitoring of the study participants:

- The timing of procedures for assessment of safety procedures may be modified based on newly available safety and tolerability data or evolving COVID-19 data.

- Up to an additional 25 mL of blood may be drawn for safety or immunogenicity analyses. The total blood volume withdrawn from any single participant will not exceed the maximum allowable volume during his or her participation in the entire study.
- Additional database freezes may occur as the study evolves and should the ongoing epidemic progression warrant rapid decision-making on product manufacturing. The study will continue in a blinded fashion (at the participant level) until the EOS.
- Rapid diagnostic testing for SARS-CoV-2 by point-of-care tests may be available and substituted for centralised testing if accepted by regulatory authorities as a secondary endpoint in this study and hold validity for study vaccine advancement.

It is understood that the current study may employ some or none of the alterations described above. Any changes in this research activity, except those necessary to remove an apparent immediate hazard to the participant, must be reviewed and approved by the sponsor or designee. Amendments to the protocol must be approved by the REC, and regulatory authority where applicable, before participants can be enrolled into an amended protocol.

#### 9.3.3.2.2 Protocol Deviations

The investigator or designee must document and explain in the participant's source documentation any deviation from the approved protocol. The investigator may implement a deviation from, or a change to, the protocol to eliminate an immediate hazard to study participants without prior REC approval. As soon as possible after such an occurrence, the implemented deviation or change, the reasons for it, and any proposed protocol amendments should be submitted to the REC for review and approval, to the sponsor for agreement, and to the regulatory authorities, if required.

A protocol deviation is any change, divergence, or departure from the study design or procedures defined in the protocol. An important deviation (sometimes referred to as a major or significant deviation) is a subset of protocol deviations that leads to a participant being discontinued from the study or significantly affects the participant's rights, safety, or well-being and/or the completeness, accuracy, and reliability of the study data. An important deviation can include nonadherence to inclusion or exclusion criteria or nonadherence to regulatory authority including ICH E6(R2) guidelines.

Protocol deviations will be documented by the clinical monitor throughout the course of monitoring visits. The investigator will be notified in writing by the monitor of deviations. The REC should be notified of all protocol deviations, if appropriate, in a timely manner.

###### 9.3.3.3 Study Termination

Although the sponsor has every intention of completing the study, they reserve the right to discontinue it at any time for clinical or administrative reasons.

The end of the study is defined as the date on which the last participant completes the last study visit (including the EOS visit and any additional long-term follow-up). Any additional long-term follow-up that is required for monitoring of the resolution of an AE or finding may be appended to the clinical study report.

###### 9.3.3.4 Final Report

Regardless of whether the study is completed or prematurely terminated, the sponsor will ensure that clinical study reports are prepared and provided to regulatory agency(ies) as required by the applicable regulatory requirement(s). The sponsor will also ensure that clinical study reports in marketing applications meet the standards of the ICH E3: Structure and Content of Clinical Study Reports.

Where required by applicable regulatory requirements, an investigator signatory will be identified for the approval of the clinical study report. The investigator will be provided reasonable access to statistical tables, figures, and relevant reports and will have the opportunity to review complete study results.

#### 9.4 Appendix 4: FDA Toxicity Grading Scales

**Table 9-1 FDA Toxicity Grading Scale for Clinical Abnormalities (Local and General Systemic Reactogenicity)**

| **Local Reaction to Injectable Product** | | | | |
|---|---|---|---|---|
| **Clinical Abnormality** | **Mild (Grade 1)** | **Moderate (Grade 2)** | **Severe (Grade 3)** | **Potentially Life Threatening (Grade 4)** |
| Pain | Does not interfere with activity | Repeated use of nonnarcotic pain reliever >24 hours or interferes with activity | Any use of narcotic pain reliever or prevents daily activity | ER visit or hospitalization |
| Tenderness | Mild discomfort to touch | Discomfort with movement | Significant discomfort at rest | ER visit or hospitalization |
| Erythema/redness[a] | 2.5 – 5 cm | 5.1 – 10 cm | > 10 cm | Necrosis or exfoliative dermatitis |
| Induration/swelling[b] | 2.5 – 5 cm and does not interfere with activity | 5.1 – 10 cm or interferes with activity | > 10 cm or prevents daily activity | Necrosis |
| **Systemic (General)** | | | | |
| **Clinical Abnormality** | **Mild (Grade 1)** | **Moderate (Grade 2)** | **Severe (Grade 3)** | **Potentially Life Threatening (Grade 4)** |
| Fever (°C)<br>(°F) | 38.0 – 38.4<br>100.4 – 101.1 | 38.5 – 38.9<br>101.2 – 102.0 | 39.0 – 40<br>102.1 – 104 | > 40<br>> 104 |
| Nausea/vomiting | No interference with activity or 1 – 2 episodes/24 hours | Some interference with activity or >2 episodes/24 hours | Prevents daily activity, or requires outpatient IV hydration | ER visit or hospitalization for hypotensive shock |
| Headache | No interference with activity | Repeated use of nonnarcotic pain reliever >24 hours or some interference with activity | Significant; any use of narcotic pain reliever or prevents daily activity | ER visit or hospitalization |
| Fatigue/Malaise | No interference with activity | Some interference with activity | Significant; prevents daily activity | ER visit or hospitalization |
| Myalgia | No interference with activity | Some interference with activity | Significant; prevents daily activity | ER visit or hospitalization |
| Arthralgia | No interference with activity | Some interference with activity | Significant; prevents daily activity | ER visit or hospitalization |

Abbreviations: DHHS = Department of Health and Human Services; ER = emergency room; FDA = United States Food and Drug Administration.

[a] In addition to grading the measured local reaction at the greatest single diameter, the measurement should be recorded as a continuous variable.

[b] Induration/swelling should be evaluated and graded using the functional scale as well as the actual measurement.

[c] Oral temperature; no recent hot or cold beverages.

Source: DHHS 2007.

**Table 9-2 FDA Toxicity Grading Scale for Clinical Abnormalities (Vital Signs)**

| Vital Signs | Mild (Grade 1) | Moderate (Grade 2) | Severe (Grade 3) | Potentially Life Threatening (Grade 4) |
|---|---|---|---|---|
| Tachycardia (bpm) | 101 – 115 | 116 – 130 | > 130 | ER visit or hospitalization for arrhythmia |
| Bradycardia (bpm)[a] | 50 – 54 | 45 – 49 | < 45 | ER visit or hospitalization for arrhythmia |
| Hypertension (systolic) (mm Hg) | 141 – 150 | 151 – 155 | > 155 | ER visit or hospitalization for malignant hypertension |
| Hypertension (diastolic) (mm Hg) | 91 – 95 | 96 – 100 | > 100 | ER visit or hospitalization for malignant hypertension |
| Hypotension (systolic) (mm Hg) | 85 – 89 | 80 – 84 | < 80 | ER visit or hospitalization for hypotensive shock |
| Respiratory Rate (breaths per minute) | 17 – 20 | 21 – 25 | > 25 | Intubation |

Abbreviations: DHHS = Department of Health and Human Services; ER = emergency room; FDA = United States Food and Drug Administration.

Note: Participant should be at rest for all vital sign measurements, with toxicity scored on day of study vaccination (pre- and post-dose).

[a] When resting heart rate is between 60 – 100 bpm. Use clinical judgement when characterising bradycardia among some healthy participant populations (e.g., conditioned athletes).

Source: DHHS 2007.

#### 9.5 Appendix 5: Listings of Adverse Events of Special Interest

Because it has been hypothesised that immunisations with or without adjuvant may be associated with autoimmunity, regulatory authorities have requested that Novavax instruct investigators to be especially vigilant regarding the PIMMC listed in Table 9-3.

**Table 9-3 Potential Immune-Mediated Medical Conditions (PIMMC)**

| Categories | Diagnoses (as MedDRA Preferred Terms) |
|---|---|
| Neuro-inflammatory Disorders: | Acute disseminated encephalomyelitis (including site specific variants: e.g., noninfectious encephalitis, encephalomyelitis, myelitis, myeloradiculomyelitis), cranial nerve disorders including paralyses/paresis (e.g., Bell's palsy), generalised convulsion, Guillain-Barre syndrome (including Miller Fisher syndrome and other variants), immune-mediated peripheral neuropathies and plexopathies (including chronic inflammatory demyelinating polyneuropathy, multifocal motor neuropathy and polyneuropathies associated with monoclonal gammopathy), myasthenia gravis, multiple sclerosis, narcolepsy, optic neuritis, transverse myelitis, uveitis |
| Musculoskeletal and Connective Tissue Disorders: | Antisynthetase syndrome, dermatomyositis, juvenile chronic arthritis (including Still's disease), mixed connective tissue disorder, polymyalgia rheumatic, polymyositis, psoriatic arthropathy, relapsing polychondritis, rheumatoid arthritis, scleroderma (including diffuse systemic form and CREST syndrome), spondyloarthritis (including ankylosing spondylitis, reactive arthritis [Reiter's Syndrome] and undifferentiated spondyloarthritis), systemic lupus erythematosus, systemic sclerosis, Sjogren's syndrome |
| Vasculidities: | Large vessels vasculitis (including giant cell arteritis such as Takayasu's arteritis and temporal arteritis), medium sized and/or small vessels vasculitis (including polyarteritis nodosa, Kawasaki's disease, microscopic polyangiitis, Wegener's granulomatosis, Churg–Strauss syndrome [allergic granulomatous angiitis], Buerger's disease [thromboangiitis obliterans], necrotising vasculitis and anti-neutrophil cytoplasmic antibody [ANCA] positive vasculitis [type unspecified], Henoch-Schonlein purpura, Behcet's syndrome, leukocytoclastic vasculitis) |
| Gastrointestinal Disorders: | Crohn's disease, celiac disease, ulcerative colitis, ulcerative proctitis |
| Hepatic Disorders: | Autoimmune hepatitis, autoimmune cholangitis, primary sclerosing cholangitis, primary biliary cirrhosis |
| Renal Disorders: | Autoimmune glomerulonephritis (including IgA nephropathy, glomerulonephritis rapidly progressive, membranous glomerulonephritis, membranoproliferative glomerulonephritis, and mesangioproliferative glomerulonephritis. |
| Cardiac Disorders: | Autoimmune myocarditis/cardiomyopathy |
| Skin Disorders: | Alopecia areata, psoriasis, vitiligo, Raynaud's phenomenon, erythema nodosum, autoimmune bullous skin diseases (including pemphigus, pemphigoid and dermatitis herpetiformis), cutaneous lupus erythematosus, morphoea, lichen planus, Stevens-Johnson syndrome, Sweet's syndrome |
| Haematologic Disorders: | Autoimmune hemolytic anaemia, autoimmune thrombocytopenia, antiphospholipid syndrome, thrombocytopenia |
| Metabolic Disorders: | Autoimmune thyroiditis, Grave's or Basedow's disease, Hashimoto thyroiditis[a], diabetes mellitus type 1, Addison's disease |

| Categories | Diagnoses (as MedDRA Preferred Terms) |
|---|---|
| Other Disorders: | Goodpasture syndrome, idiopathic pulmonary fibrosis, pernicious anaemia, sarcoidosis |

Abbreviations: ANCA = anti-neutrophil cytoplasmic antibody; IgA = immunoglobulin A; MedDRA = Medical Dictionary for Regulatory Activities.

[a] For Hashimoto thyroiditis: new onset only.

AESIs relevant to COVID-19 are listed in Table 9-4. The list is not intended to be exhaustive, nor does it exclude the possibility that other diagnoses may be AESI. It is anticipated that additional AESI may be associated with COVID-19. Investigators should stay updated regarding such public health notifications.

**Table 9-4 Adverse Events of Special Interest Relevant to COVID-19[a]**

| Body System | Diagnoses[a] |
|---|---|
| Immunologic | Enhanced disease following immunisation,[b] cytokine release syndrome related to COVID-19[c], Multisystem inflammatory syndrome in children (MIS-C) |
| Respiratory | Acute respiratory distress syndrome (ARDS) |
| Cardiac | Acute cardiac injury including:<br>• Microangiopathy<br>• Heart failure and cardiogenic shock<br>• Stress cardiomyopathy<br>• Coronary artery disease<br>• Arrhythmia<br>• Myocarditis, pericarditis |
| Haematologic | Coagulation disorder<br>• Deep vein thrombosis<br>• Pulmonary embolus<br>• Cerebrovascular stroke<br>• Limb ischemia<br>• Hemorrhagic disease<br>• Thrombotic complications |
| Renal | Acute kidney injury |
| Gastrointestinal | Liver injury |
| Neurologic | Guillain-Barré Syndrome, anosmia, ageusia, meningoencephalitis |
| Dermatologic | Chilblain-like lesions, single organ cutaneous vasculitis, erythema multiforme |

Abbreviations: AESI = adverse event of special interest; COVID-19 = coronavirus disease 2019; DAIDS = Division of AIDS; PCR = polymerase chain reaction; SARS-CoV2 = severe acute respiratory syndrome coronavirus 2.

[a] To be recorded as AESIs relevant to COVID-19, these complications should be associated with a positive PCR test for SARS-CoV-2.

[b] COVID-19 manifestations associated with more severe presentation and decompensation with consideration of enhanced disease potential (SPEAC 2020).

[c] Cytokine release syndrome related to COVID-19 infection is a disorder characterised by nausea, headache, tachycardia, hypotension, rash, and/or shortness of breath (DAIDS 2017).

| Table of Amendments to Protocol | |
|---|---|
| **Version 3.0 (December 23, 2020) to Version 4.0 (25 February 2021)** | |
| **Location of change** | **Change/Modification** |
| General | • The primary purpose of this amendment is to add the blinded crossover design. All changes were made because of addition of crossover design except as noted below. |
| Synopsis; Secondary Objectives (Section 2.1.2) | • Deleted "in SARS CoV-2 seropositive" from the second objective to correct error.<br>• Added "in the initial set of vaccinations" to the 9th secondary objective per MHRA recommendation.<br>• Changed safety and reactogenicity objectives from 7 days after each study vaccination to "the initial set of study vaccinations."<br>• Added the following secondary objective: "To assess the duration of vaccine efficacy (measured by all efficacy endpoints) in initial active vaccine recipients vs. crossover (delayed) active vaccine recipients. |
| Synopsis; Exploratory Objectives (Section 2.1.3) | • Added "in the initial set of vaccinations" to the first and second exploratory objective.<br>• Added "unblinded before the crossover" to second exploratory objective. |
| Synopsis; Primary Endpoint (Section 2.2.1) and Secondary Endpoints (Section 2.2.2) | • Added "in the initial set of vaccinations" to the primary endpoint and the first 6 secondary endpoints.<br>• Changed "First occurrence of laboratory-confirmed COVID-19 to participants with negative serostatus at baseline. |
| Synopsis; Other Secondary Endpoints (Section 2.2.2.2) | • Added analysis of antibodies endpoint for Crossover Day 0 and Day 35.<br>• Deleted "for 7 days" and added "the initial set of vaccinations" to reactogenicity endpoint.<br>• Added "following the initial set of vaccinations" to MAAE and unsolicited AE endpoint.<br>• Added the following secondary endpoint: "Relative vaccine efficacy (measured by all efficacy endpoints) in initial active vaccine recipients vs. crossover (delayed) active vaccine recipients. |
| Synopsis; Exploratory Endpoints (Section 2.2.3) | • Added "in the initial set of vaccinations" to the first 2 exploratory endpoints.<br>• Added the following endpoint: "Any occurrence of serologic conversion (by serology to SARS-CoV-2 N protein) between Crossover Day 0 visit and the end of study visit in the second set of vaccinations in adult participants seronegative at baseline."<br>• Added "and Crossover Day 35 visit (14 days after the Crossover Day 21 visit" to seroconversion and VNA endpoint.<br>• Added the following endpoint: First occurrence of virologically confirmed (by PCR to SARS-CoV-2), symptomatic mild, moderate, or severe COVID-19, with onset at least 7 days after second study vaccination (e.g., Day 28) in the initial set of vaccinations in seronegative adult participants by age ($<65$ and $\geq 65$) in racial and ethnic minorities and in those with co-morbid conditions.<br>• Added "The occurrence, severity, and relationship to study vaccination of unsolicited AEs (in all adult participants) for 21 days after first study vaccination and 28 days after second study vaccination following the initial set of vaccinations with the adjustment to remove reactogenicity events which were recorded as unsolicited AEs within 7 days of each dose in the initial set of vaccinations." |

| Table of Amendments to Protocol | |
|---|---|
| **Version 3.0 (December 23, 2020) to Version 4.0 (25 February 2021)** | |
| **Location of change** | **Change/Modification** |
| Synopsis; Study Design (Section 3) | • Added description of crossover design.<br>• Added "and Crossover Day 0 visit" for nose/throat samples.<br>• Updated paragraph about participants receiving an approved vaccine to include crossover portion; added a sentence about participants who are unblinded and receive another vaccine.<br>• Added "Participants who are unblinded and ineligible for the blinded crossover will continue to follow the original study design featured in Figure 1b.<br>• Clarified that the duration of individual participation is 1 year from the initial set of vaccinations to correct error.<br>• Added information about blood draws and assessment of vaccine efficacy.<br>• Added information about unblinded participants who receive another vaccine |
| Synopsis; Study Vaccination Pause Rules (Section 3.1) | • Added "for the first set of vaccinations only" to pause rules based on reactogenicity. |
| Synopsis; Exclusion Criteria (Section 4.2) | • Added statement about inclusion/exclusion criteria at crossover. |
| Synopsis ; Study Vaccine Administration (Section 5.1) | • Added "At the time of the blinded crossover, participants will receive the alternate study material in 2 IM injections 21 + 7 days apart" to description of study vaccines.<br>• Deleted description of nose/throat samples and referred to SOE. |
| Day21 (Section 6.1.3) | • Added sentence about assessment of exclusion criteria prior to second dose. |
| COVID Surveillance Visits (Section 6.1.5.1 and 6.1.5.2) | • Added "as appropriate" to collection of concomitant medications and added collection of nose/throat samples. |
| Synopsis; Immunogenetic Assessments (Section 6.3) | • Added "Anti-S protein serology in the Anti-S Protein Serology Subset will be collected after the first and second set of vaccinations." |
| Synopsis; Safety Assessments (Section 6.4) and Local and General Systemic Reactogenicity Symptoms (Section 6.4.1.1.2) | • Added "after the first set of vaccinations" and "in the initial set of vaccinations to Safety Assessments.<br>• Added "Local and systemic reactogenicity events will not be recorded for the second set of vaccinations."<br>• Added statement about unsolicited AEs in participants who are not in the sub-study.<br>• Deleted 'virologically confirmed" to correct error.<br>• Added "who will be initially" to description of randomization.<br>• Added "unblinded participants should not be included in the blinded crossover." |
| MAAEs (Section 6.4.1.1.4) | • Added "for the first set of vaccinations only." |
| Synopsis; Analysis Sets (Section 7.2) | • Changed 7 to 6 days or less for positive SARS-CoV-2 test to correct error.<br>• Deleted serum IgG antibody and changed to antibody tests to correct error. |
| Synopsis; Efficacy Analyses (Section 7.3.1) | • Added a paragraph about primary analysis of primary and key secondary efficacy endpoints pre and post crossover. |
| Synopsis; Immunogenicity Analysis (Section 7.3.2) | • Added description of immunogenicity analysis after the second set of vaccinations.<br>• Deleted sentence about EOS analysis. |

| Table of Amendments to Protocol | |
|---|---|
| **Version 3.0 (December 23, 2020) to Version 4.0 (25 February 2021)** | |
| **Location of change** | **Change/Modification** |
| Synopsis; Safety Analysis (Section 7.3.3) | • Added "after the initial set of vaccinations only" to capture of solicited AEs.<br>• Added clarification about solicited AEs in participants who are not part of the sub-study. |
| Synopsis; Interim Analyses (Section 7.5) | • Added "Novavax will be unblinded at the participant level at the time of the primary 100-event analysis." |
| Synopsis; Pre- and Post-Blinded Crossover (Sections 7.6 and 7.7) | • Added sections on pre- and post-blinded crossover. |
| Clinical Experience (Section 1.3) | • Added sentence about 302 interim analysis. |
| Figure 1a: Trial Design | • Updated figure to portray crossover design (Figure 1a) and changed Figure 1 to Figure 1b.<br>• Added footnote to Figure 1a to describe possible consolidation of crossover visits. |
| Table 3-1: Schedule of Events | • Updated to include crossover design |
| Method of Assigning Participants (Section 5.3) | • Added sentence about the use of the IRT system. |
| Blinding Procedures (Section 5.3.1) | • Added sentence about keeping study participants and personnel blinded.<br>• Added information about reconsenting participants at crossover. |
| Concomitant Medications (Section 5.5.1) | • Added clarification about concomitant medications based on new study design. |
| Initial COVID-19 Surveillance Visit (Section 6.1.5) and Follow-Up Visit (Section 6.1.5.2) | • Added "as appropriate" to collection of prior and concomitant medications.<br>• Added paragraph about nose/throat sampling. |
| Unblinding Visit (Section 6.1.6) | • Changed instructions for unblinding visit.<br>• Added written informed consent<br>• Added "as appropriate" to collection of prior and concomitant medication.<br>• Deleted "If prior to participant's Day 21 and Day 35 visit." |
| 3 Month Visit (Section 6.1.7) | • Added a paragraph changing the requirement for the 3-month visit.<br>• Added "as appropriate" to collection of prior and concomitant medications. |
| Crossover Day 0 visit (Section 6.1.8) | • Added section |
| Crossover Day 21 visit | • Added section.<br>• Deleted 6-month visit. |
| Crossover Day 35 visit | • Added section |
| 12 months After Second Vaccination (Section 6.1.11) | • Updated instructions for EOS visit. |
| Day 35—Follow-up Visit (Section 6.1.4) | • Deleted "Visit may be skipped as per unblinding visit below." |
| Nose/Throat Samples (Section 6.2.1) | • Added Crossover Day 0 and 21 for Nose/Throat samples. |
| Virologic Confirmation of SARS-CoV-2 (Section 6.2.2 | • Deleted section |

| Table of Amendments to Protocol | |
|---|---|
| **Version 3.0 (December 23, 2020) to Version 4.0 (25 February 2021)** | |
| **Location of change** | **Change/Modification** |
| Initial COVID-19 Surveillance Visit | • Added "as appropriate" to ascertainment of new concomitant medications. |
| Safety Assessment (Section 6.4) | • Added "unblinded participants should not be included in the blinded crossover." |
| MAAEs (Section 6.4.1.1.4) | • Added "after the first set of vaccinations only" to MAAE collection. |
| Statistical analysis (Section 7.3) | • Changed information about management of unblinded participants. |
| Reference List (Section 8) | • Updated based on new text. |

| Table of Amendments to Protocol | |
|---|---|
| **Version 2.0 (23 October 2020) to Version 3.0 (23 December 2020)** | |
| **Location of change** | **Change/Modification** |
| Synopsis | • Updated number of study sites from 28 to 33. |
| Synopsis; Exploratory Objectives (Section 2.1.3) | • Added an objective to exploratory endpoint to explore the efficacy and safety of SARS-CoV-2rS with Matrix-M1 adjuvant with an approved or deployed vaccine to take into account the changing vaccine landscape. |
| Synopsis; Secondary Endpoints (Section 2.2.2) | • Added a secondary endpoint to measure severe COVID-19 separately. |
| Synopsis; Exploratory Endpoints (Section 2.2.3) | • Added 2 exploratory endpoints, one due to changes in vaccine landscape and one to address asymptomatic disease. |
| Synopsis; Study Design (Section 3) | • Added "approximately" to description of 400 participants in substudy to allow for greater flexibility. |
| Synopsis; Section 1 (Introduction); Section 3 (Study Design); Section 5.3.2 (Breaking the Blind) | • Added advice from regulatory agencies concerning unblinding of treatment assignment for participants receiving an approved or deployed SARS-CoV-2 vaccine during the study. |
| Synopsis, Schedule of Events, Section 6.1.5.1 (Initial COVID-19 Surveillance Visit); Follow-up COVID-19 Surveillance Visit (Section 6.1.5.2); and 6.2.3.2 (COVID-19 Surveillance Visit [Initial and Follow-up]) | • Added information about replacing in-person COVID-19 Surveillance Visits with phone calls if government restrictions are in place. Also added "with exception per protocol" to accommodate this new recommendation. |

| Table of Amendments to Protocol | |
|---|---|
| **Version 2.0 (23 October 2020) to Version 3.0 (23 December 2020)** | |
| **Location of change** | **Change/Modification** |
| Synopsis, Section 7.1 (Sample Size), Table 7-1 | • Reduced number of target endpoints from 152 to 100 based on higher than predicted vaccine efficacy observed in the recent Phase 3 trials by Pfizer and Moderna. This reduced number was chosen to provide > 95% power for 70% or higher vaccine efficacy.<br>• Reduced number of planned interim analyses from 2 to 1 and revised timing of the single interim and final analyses of the primary efficacy endpoint to 50 and 100 events, respectively. |
| Synopsis, Section 7.2 (Analysis Sets) | • Clarified the wording of the PP-EFF analysis set. |
| Synopsis, Section 7.3.1 (Efficacy Analysis) | • Clarified the wording of the primary efficacy analyses at the interim and final analyses of the primary efficacy endpoint. |
| Synopsis, Section 7.3.2 (Immunogenicity Analysis and Correlates of Risk) | • Changed HAI to ELISA for SARS-CoV-2 rS serum antibody levels to correct error and added influenza vaccine to later sentences for greater clarity. |
| Synopsis, Section 7.5 (Interim Analysis); Table 7-2 | • Reduced the number of interim analyses for the primary efficacy endpoint from 2 to 1, with the timing of the single interim analysis based on the accumulation of approximately 50% (50 events).<br>• Clarified that the unblinded Biostatistics and Programming team is from the CRO, PPD.<br>• Clarified the wording of the single interim and final analyses of the primary efficacy endpoint. |
| Synopsis (2 times); Section 6.4 (Safety Assessments); Section 7.3.3. (Safety Analyses) | • Added "Separate analyses of safety events received with respect to approved or deployed SARS-CoV-2 vaccines will be performed" to synopsis. Added specific information about safety reporting for participants who receive an approved or deployed SARS-CoV-2 vaccine. |
| Introduction | • Added a paragraph about changes due to approved or deployed SARS-CoV-2 vaccines. |
| Section 2.2.1 (Primary Endpoints); Table 2-1; Section 6.2.3 (Severity of COVID-19 Symptoms) | • Added the following footnote: "Participants with vital sign abnormalities in the moderate or severe categories must meet the criteria for mild COVID-19" and added the same information in text. |
| Section 2.2.1 (Primary Endpoints); Table 2-2 | • Removed duration of ≥ 48 hours for headache, nausea, vomiting, and diarrhea for consistency with other SARS-CoV-2 rS trials. |
| Section 4.4.1 (Reasons for Withdrawal) | • Added a statement that vaccination with an approved or deployed SARS-CoV-2 vaccine does not constitute a withdrawal, this is based on MHRA guidance. |
| Section 5.3.1 (Blinding Procedures); Section 5.3.2 (Breaking the Blind) | • Added specific information on unblinding if participants wish to receive an approved or deployed SARS-CoV-2 vaccine. |
| Section 5.5.1 (Concomitant Medications) | • Added that participants will be asked to record the date and brand of the approved or deployed vaccine that they receive. |

| Table of Amendments to Protocol | |
|---|---|
| **Version 2.0 (23 October 2020) to Version 3.0 (23 December 2020)** | |
| **Location of change** | **Change/Modification** |
| Section 5.5.2 (Prohibitive Therapy) | • Added an exception to vaccine restrictions for participants who wish to receive an approved or deployed vaccine at least 21 days after study vaccine. |
| Section 6.1 (Study Visit Procedures) | • Added "Visit may be skipped as per the Unblinding Visit described below" in Sections 6.1.3 and 6.1.4.<br>• Clarified that a targeted physical examination may be performed if participant has any ongoing complaints in Section 6.1.4.<br>• Clarified that blood sampling for SARS-CoV-2 (ELISA for anti-N-protein serology) in all participants "unless the participant has had an Unblinding Visit in the prior 14 days where anti-N serology was drawn" in Sections 6.1.4, 6.1.7, and 6.1.8. |
| Section 6.1.5.1 (Initial COVID-19 Surveillance Visit) | • Added "including those who have been unblinded" to list of requirements at this visit. |
| Section 6.1.6 (Unscheduled Blinding Visit) | • Added section to describe unblinding process for people who wish to receive approved or deployed SARS-CoV-2 vaccine. |
| Section 6.2.2 (Virologic Confirmation of SARS-CoV-2) | • Clarified that nose/throat self-sampling may be skipped is a participant is known to be PCR positive to SARS-CoV-2 from the Screening or Day 0 PCR within 14 days of the positive test.<br>• Clarified that if a participant obtains a PCR test for SARS-CoV-2 outside of the study for any other reason than having suspected COVID-19 symptoms, then this result should not be noted or reported.<br>• Clarified that if a participant obtains a PCR test for SARS-CoV-2 outside of the study for suspected COVID-19 symptoms, then the participant should begin all protocol-required assessments for participants with suspected COVID-19 symptoms. |
| Section 6.2.3 (Monitoring for Suspected COVID-19) | • Added "This is the case for blinded and unblinded participants." |
| Section 6.2.3.2 (COVID-19 Surveillance Visit) | • Added "See Section 6.1.5.1 for exceptions for participants affected by extenuating circumstances (e.g., local government restrictions). |
| Section 6.2.3.2.1 (Initial COVID-19 Surveillance Visit) | • Added "Proper documentation in the eCRF of the dates, bar codes, and results of all self-swabs taken in proximity of this visit." |
| Section 6.4 (Safety Assessments) | • Added safety requirements for unblinded patients. |
| Section 6.4.1.1.2 (Local and General Systemic Reactogenicity Symptoms) | • Added "Solicited AEs will not be captured for any approved or deployed SARS-CoV-2 vaccine." |
| Section 6.4.1.3 (Reporting Adverse Events) | • Added information about SAE reporting for participants who receive an approved or deployed vaccine. |

| Table of Amendments to Protocol | |
|---|---|
| **Version 2.0 (23 October 2020) to Version 3.0 (23 December 2020)** | |
| **Location of change** | **Change/Modification** |
| Section 7.3 (Statistical Analysis) | • Added "In order to manage unblinded participants, unblinding will result in censoring of all efficacy and immunogenicity endpoints. The main presentation of data based on the Safety Analysis set will include all participant data, with supporting presentations to exclude the data post unblinding. Sensitivity analysis of reactogenicity and unsolicited AEs will be conducted for the Safety analysis subset of participants not unblinded. Additional analysis of AEs collected after unblinding may be undertaken as exploratory safety analyses. ." |

Abbreviations: COVID-19 = coronavirus disease 2019; CRO = contract research organisation; ELISA = enzyme-linked immunosorbent assay; HAI = hemagglutination inhibition assay; PCR = polymerase chain reaction; PP-EFF = per-protocol efficacy; SARS-CoV-2 = severe acute respiratory syndrome coronavirus 2; SARS-CoV-2 rS = severe acute respiratory syndrome coronavirus 2 recombinant spike nanoparticle vaccine.

| Table of Amendments to Protocol | |
|---|---|
| **Changes Made from Version 1.0 (17 September 2020) to Version 2.0 (23 October 2020)** | |
| **Location of Change** | **Change/Modification** |
| **Changes Made from Version 1.0 (17 September 2020) to Version 1.1 (17 September 2020)** | |
| Section 4.4.1 (Reasons for Withdrawal) | The following changes were made based on MHRA recommendations:<br>• Changed "may withhold" to "will withhold" further vaccinations.<br>• Deleted "The investigator will withhold further study vaccination from a participant in the study if the participant" prior to the bullet regarding pregnancy to indicate that all numbered items are required to withhold vaccination.<br>• Deleted "Upon the occurrence of an SAE or intolerable AE, the investigator may confer with the sponsor before future study vaccination" for more restrictive requirements. |
| **Changes Made from Version 1.1 (17 September 2020) to Version 1.2 (21 September 2020)** | |
| Synopsis (Study Vaccination Pause Rules); Section 3.1 | • Added: "for example, any SAE for which causality is at least possibly related" for greater clarity/MHRA recommendation. |
| Section 6.1.4 [Day 35 – Follow-up Visit (+ 7 days)] | • Deleted "prior to study vaccination" from the blood sampling procedures on Day 35 since vaccination is only given on Days 0 and 21 (MHRA recommendation). |
| Section 6.1.6 [3 Months (± 15 days) After Second Study Vaccination] | • Deleted "prior to study vaccination" from the blood sampling procedure on Month 3 since vaccination is only given on Days 0 and 21 (MHRA recommendation). |
| Section 6.1.7 [6 Months (± 15 days) After Second Study Vaccination] | • Deleted "prior to study vaccination" from the blood sampling procedure on Month 6 since vaccination is only given on Days 0 and 21 (MHRA recommendation). |
| Section 6.1.8 [12 Months (± 15 days) After Second Study Vaccination] | • Deleted "prior to study vaccination" from the blood sampling procedure on Month 12 since vaccination is only given on Days 0 and 21 (MHRA recommendation). |

| Table of Amendments to Protocol | |
|---|---|
| **Changes Made from Version 1.0 (17 September 2020) to Version 2.0 (23 October 2020)** | |
| **Location of Change** | **Change/Modification** |
| **Changes Made from Version 1.2 (21 September 2020) to Version 2.0 (23October 2020)** | |
| Synopsis | • Changed “up to 18 regions” to “28 sites” across the United Kingdom (UK) based on updated information. |
| Synopsis (Secondary Objectives); Section 2.1.2 | • Added “related to study vaccination” to 6th secondary objective for clarity.<br>• Deleted “and in terms of unsolicited AEs for 21 days after first study vaccination and 28 days after second study vaccination” and added a separate bullet for MAAEs for greater clarity. |
| Synopsis (Primary Endpoints); Section 2.2.1 | • Changed to only one primary endpoint and moved the second primary endpoint to key secondary endpoint<br>• Added “mild, moderate, or severe” to COVID-19 description in primary endpoint for clarity |
| Synopsis (Secondary Endpoints); Section 2.2.2 | • Added “mild, moderate, or severe” to COVID-19 description to 2nd secondary endpoint for clarity.<br>• Added “symptomatic” to description of mild COVID19 to 4th primary endpoint for clarity.<br>• Added N-protein to serology endpoint for greater clarity.<br>• Eliminated ELISA testing from Day 21, as this was an oversight from a prior version.<br>• Added “related to study vaccination” to endpoint for SAEs and MAAEs for greater clarity.<br>• Added 3 safety endpoints (i.e., AESIs/PIMMCs, solicited AEs, and unsolicited AEs) to correlate with secondary objectives. |
| Synopsis (Exploratory Endpoints); Section 2.2.3 | • Added “mild, moderate, or severe” to COVID-19 description to first exploratory endpoint |
| Synopsis (Study Design); Section 3 | • Changed “regions” to “sites” in 2 places for clarity.<br>• Changed mucosal to nose/throat and eliminated saliva samples for COVID-19 at Screening and Day 0 due to lack of saliva test availability in UK.<br>• Changed study population from 9000 to 15,000 to increase accrual rate of COVID-19 endpoints.<br>• Deleted information about enrolment of participants ≥age 65 and operational cutoff based on updated information. |
| Synopsis (Table S1-1) | • Changed both groups from 4,500 to 7,500 to accommodate increased enrolment. |
| Synopsis (Study Vaccination Pause Rules); Section 3.1 | • Allowed the SMC chair to review SAEs and decide on the need for a full SMC meeting before starting a study enrolment pause.<br>• Expanded requirement for solicited and unsolicited AEs to also include Grade 4 (potentially life-threatening), in addition to Grade 3, for greater clarity.<br>• Added “after a minimum of 100 participants were enrolled” to 5% and 10% cutoffs for solicited and unsolicited AEs to prevent a pause being triggered due to an early cluster of grade 3 events.<br>• Changed grade 3 (severe) to severe and added “after a minimum of 100 subjects are enrolled” to third bullet for clarity.<br>• Added “for example, any SAE for which causality is at least possibly related” to provide more clarity to pause rule. |

| Table of Amendments to Protocol | |
|---|---|
| **Changes Made from Version 1.0 (17 September 2020) to Version 2.0 (23 October 2020)** | |
| **Location of Change** | **Change/Modification** |
| | • Added specific text about analysis of grade 3 or higher solicited AEs in 400-subject influenza substudy for greater clarity. |
| Synopsis (Inclusion Criteria); Section 4.1 | • Deleted "with spermicide" from condom criteria due to input from the UK ethics review.<br>• Added "Other approaches to abstinence are not acceptable" to clarify how this method can be used as contraception due to input from the UK ethics review. .<br>• Added "Room saturation of > 95% at Screening/Day 0 to inclusion criteria as this is aligned with the severity grading criteria which assigns a greater level of severity below 95%. |
| Synopsis (Exclusion Criteria); Section 4.2 | • Changed participation in serologic surveys to future participation to allow participation to those who participated prior to enrollment.<br>• Added text that allowed participants with controlled HIV to participate per investigator's suggestion.<br>• Added "within 3 months following the last study vaccine" to pregnancy/lactation criteria participate per investigator's suggestion.<br>• Added additional information to restrictions on administration of influenza vaccine for greater clarity.<br>• Added "including hepatitis B and C" to hepatic exclusion for greater clarity per investigator's suggestion.<br>• Deleted "and epilepsy" from neurological disorders per investigator's suggestion.<br>• Deleted "or immunodeficiency" and added Table 9-3 (specific conditions) and the need for biologic therapy for greater clarity as it was repetitious of a prior exclusion.<br>• Added "The Skin and Metabolic disorders listed in Table 9-3 are eligible at the discretion of the investigator" and eliminated caveat about endocrine disorders.<br>• Added the use of continuous oxygen therapy as an Exclusion Criteria (allowing for nocturnal oxygen) as it would not be possible to grade severity levels based on oxygen saturation if someone is on continuous oxygen.<br>Other Considerations:<br>Added a bullet point addressing participants with hypertension. |
| Synopsis (Study Vaccine Administration); Section 5.1 | • Added information about specific flu vaccines for participants 18-64 years of age and those ≥ 65 years of age.<br>• Added instructions for using the right deltoid for flu vaccine and the left deltoid for study vaccine on Day 0 whenever possible to allow for the majority of right handed people to more easily access the study vaccine injection site. |
| Synopsis (Efficacy Assessments); Section 6.1.1, 6.1.2, and 6.1.3 | • Replaced mucosal with nose/throat and removed "or saliva" from testing method for efficacy assessments due to lack of saliva test availability in UK. |
| Synopsis (Safety Assessments); Section 6.4 | • Added "Participants in the seasonal influenza vaccine co-administration substudy will record local reactogenicity for the study vaccine injection site only" as the symptom diary could not accommodate the measurement of 2 injection sites. |

| Table of Amendments to Protocol | |
|---|---|
| **Changes Made from Version 1.0 (17 September 2020) to Version 2.0 (23 October 2020)** | |
| **Location of Change** | **Change/Modification** |
| Synopsis (Statistical Analysis Plan); Section 7.1 | • Changed sample size from 9000 to 15,000 to increase the rate of endpoint accumulation.<br>• Updated the statistical analysis plan based on increased enrolment.<br>• Deleted information designed about VE—information moved to Efficacy Section 7.3.1.<br>• Deleted information about sample size based on updated information. |
| Synopsis (Analysis Sets); Section 7.2 | • Defined the all randomized set and ITT set—deleted ITT-EFF and ITT-IMM analysis sets for further clarity.<br>• Added “baseline” to describe seronegative participants in the PP-EFF set.<br>• Added “that occur before the first COVID-19 episode” to the restriction for no major protocol deviations for the PP-EFF population for greater clarity.<br>• Changed 14-day exclusion to 7 days for positive SARS-CoV-2 illness episodes occurring after the second vaccination (i.e., Day 35 to Day 28) for PP-EFF population analysis as this was a correction.<br>• Changed Day 21 or Day 35 to Day 35 only for PP-IMM analysis as this was a correction.<br>• Added “Both PP populations will be analysed according to the study vaccine group as randomized” for further clarity. |
| Synopsis (Efficacy Analysis); Section 7.3.1 | • Changed 2 key decision points to 4 potential decision points and described them for greater clarity.<br>• Changed ITT-EFF population to ITT analysis group.<br>• Added extensive statistical information about planned analyses for greater clarity.<br>• Moved EOS analysis to Immunogenicity section<br>• Deleted reference to SAP because of expanded text.<br>• VE definition was moved here. |
| Synopsis (Immunogenicity Analysis); Section 7.3.2 | • Removed reference to ITT-IMM population—changed to ITT. Added further statistical details to the immunogenicity analysis for the influenza sub-study for greater clarity.<br>• Moved EOS analysis in Neutralisation Assay subset to this section |
| Synopsis (Safety Analysis); 7.3.3 | • Changed AE reporting window to 49 days from 35.<br>• Added “through 35 days after first study vaccination” for greater clarity. |
| Synopsis (Interim Analysis); Section 7.5 | • Added new section describing statistical information about planned interim analysis. These were added to potentially increase the speed of detecting vaccine efficacy during this global pandemic. |
| Section 1.2.1 (Nonclinical Data) | • Updated study data. |
| Section 1.3 (Clinical Experience) | • Updated Part 1 data from Phase 1/2 study and added information about Part 2.<br>• Specified number of healthy participants as 131. |
| Section 1.4 (Rationale for Study) | • Added information about results of 5-day reactogenicity data from the Phase 2 (Part 2) study.<br>• Deleted information about enrolment of participants ≥ age 65. |
| Section 2.2.1 (Table 2-1) | • Deleted reference to virologically confirmed COVID-19. |

| Table of Amendments to Protocol | |
|---|---|
| **Changes Made from Version 1.0 (17 September 2020) to Version 2.0 (23 October 2020)** | |
| **Location of Change** | **Change/Modification** |
| Section 2.2.2 (Table 2-2) | • Updated information about qualifying symptoms of COVID-19 to clarify trigger should be based on new onset of symptoms. |
| Section 3 (Study Design); Figure 1 | • Updated trial schema to include 15,000 participants to reflect increased enrolment. |
| Table 3-1 (Schedule of Events) | • Changed mucosal to nose/throat for greater clarity.<br>• Footnote c: Changed EOS telephone call to visit to correct an error.<br>• Footnote l: Added to provide further clarity on nose/throat testing.<br>• Footnote z: Edited for clarity that swabbing refers to self collection by participants. |
| Section 4 (Study Population) | • Updated introduction from 9000 to 15,000 participants to increase the rate of endpoint accumulation. |
| Section 5.2 (Investigational Products) | • Added 2 choices of flu vaccine depending on age group to table and bullet points. This information was just made available. |
| Section 5.4 (Study Vaccine Compliance) | • Deleted information about home study vaccinations, as study sites were not able to perform home vaccinations. |
| Section 5.5.2 Prohibitive Therapy | • Added information to bullet point about administration of influenza vaccine for greater clarity. |
| Section 6 (Study Procedures) | • Deleted information about home vaccinations as study sites were not able to perform home vaccinations. |
| Section 6.1.5.1 (Initial COVID-19 Surveillance Visit) | • Deleted requirement for mucosal samples at this visit.<br>• Changed target physical examination to respiratory from lymphatic and thus removed "of injected upper extremity." This was done at Surveillance visits to stress the importance of the respiratory exam for participants with possible COVID and not the need to assess the vaccination site unless symptomatic. |
| Section 6.1.5.2 (Follow-up Surveillance Visit) | • Deleted requirements for mucosal samples at this visit as these samples have a very high capture rate of RNA when shedding.<br>• Changed target physical examination to respiratory from lymphatic and thus removed "of injected upper extremity." This was done at Surveillance visits to stress the importance of the respiratory exam for participants with possible COVID and not the need to assess the vaccination site unless symptomatic. |
| Section 6.2.1 (Nose/Throat Samples for Virus Detection) | • Changed mucosal to nose/throat and removed option for saliva testing for virus detection based on test availability in UK. |
| Section 6.2.2 (Virologic Confirmation) | • Changed mucosal to nose/throat and removed option for saliva testing for virus detection.<br>• Deleted HCP sampling option for mucosal sampling as these samples have a very high capture rate of RNA when shedding. |
| Section 6.2.3.2.1 (Initial COVID-19 Surveillance Visit) | • Deleted "or the participant's basal level of chronic supplemental oxygen use" from resting respiratory rate, eliminating the use of supplemental oxygen. This is aligned with the exclusion of those on basal oxygen treatment.<br>• Deleted reference to nasal turbinate swab. |

| Table of Amendments to Protocol | |
|---|---|
| **Changes Made from Version 1.0 (17 September 2020) to Version 2.0 (23 October 2020)** | |
| **Location of Change** | **Change/Modification** |
| Section 6.2.3.2.2 (Follow-up COVID-19 Surveillance Visit) | • Changed mucosal to nose/throat and removed option for saliva testing for virus detection due to saliva test availability in UK.<br>• Deleted "however, a repeat mucosal sampling will NOT be obtained since the participant has already tested positive." This was no longer needed, as sampling for this visit was removed.<br>• Deleted paragraph reference nose/throat samples |
| Section 6.4.1.1.2 (Local and General Systemic Reactogenicity Symptoms) | • Added "of study vaccine" after injection for greater clarity.<br>• Added "Local reactogenicity symptoms should not be recorded for the influenza vaccine injection site." |
| Section 6.4.14 (Medically Attended Adverse Events) | • Added "potentially life threatening (Grade 3)" to AE criteria |
| Section 6.4.2 (Vital Sign Measurements) | • Deleted text referring to supplemental oxygen due to new exclusion criteria. |
| Section 6.4.3 (Physical Examinations) | • Added Day 0 to physical exam.<br>• Special attention should made to examine the lymph nodes of the upper extremities on vaccination days and the respiratory system at all Surveillance visits. |
| Section 7.1 (Sample Size Calculations and Table 7-1) | • Updated enrolment from 9000 to 15,000 participants.<br>• Changed number of events based on increased enrollment.<br>• Updated text to match new enrolment and revised statistical analysis. |
| Section 7.1 (Table 7-2) | • Revised table based on revised enrolment and statistical analysis.<br>• Deleted information about increasing precision around VE |
| Section 7.5 (Interim Analysis; Tables 7-3 and 7-4) | • Revised numbers in tables and added text about an interim analysis. |
| Appendix 1 | • Added Protocol Change History |
| Appendix 2 | • Updated List of Abbreviations to accommodate text changes |
| Table 9-4 | • Added a footnote: "To be recorded as AESIs relevant to COVID-19, these complications should be associated with a positive PCR test for SARS-CoV-2" for greater clarity. |

Abbreviations: AE = adverse event; AESI = adverse event of special interest; COVID-19 = coronavirus disease 2019; ELISA = enzyme-linked immunosorbent assay; EOS = end of study; HIV = human immunodeficiency virus; IM = intramuscular; ITT = intent-to-treat; ITT-EFF = intent-to-treat efficacy; ITT-IMM = intent-to-treat immunogenicity; MAAE = medically attended adverse event; MHRA = Medicines and Healthcare products Regulatory Agency; N-protein = nucleocapsid; PCR = polymerase chain reaction; PIMMC = potential immune-mediated medical conditions; PP = per-protocol; PP-EFF = per-protocol efficacy; PP-IMM = per-protocol immunogenicity; SAE = serious adverse event; SAP = statistical analysis plan; SARS-CoV-2 = severe acute respiratory syndrome coronavirus 2; SMC = safety monitoring committee; UK = United Kingdom; VE = vaccine efficacy.

**Novavax, Inc.**

**2019nCoV-302**

### A PHASE 3, RANDOMISED, OBSERVER-BLINDED, PLACEBO-CONTROLLED TRIAL TO EVALUATE THE EFFICACY AND SAFETY OF A SARS-COV-2 RECOMBINANT SPIKE PROTEIN NANOPARTICLE VACCINE (SARS-COV-2 RS) WITH MATRIX-M1™ ADJUVANT IN ADULT PARTICIPANTS 18-84 YEARS OF AGE IN THE UNITED KINGDOM

**22JAN2021**

Statistical Analysis Plan

**SAP Version 1.0**

Prepared by:

PPD
[redacted]

**Approval Signatures**

| | |
|---|---|
| Date | Prepared by: [redacted]<br>[redacted] PPD |
| Date | Reviewed by: [redacted]<br>[redacted] PPD |
| Date | Approved by: [redacted]<br>[redacted] Novavax |

#### Summary of Changes

##### Statistical Analysis Plan Revision History

| Version | Date | Summary of Changes |
|---|---|---|

#### List of Abbreviations

| | |
|---|---|
| AE | adverse event |
| AESI | adverse event of special interest |
| CI | confidence interval |
| COVID-19 | coronavirus disease 2019 |
| eCRF | electronic case report form |
| ELISA | enzyme-linked immunosorbent assay |
| ELISpot | enzyme-linked immune absorbent spot |
| EOS | end of study |
| FDA | Food and Drug Administration |
| GMEU | Geometric mean ELISA Units |
| GMFR | Geometric mean fold rise |
| HAI | Haemagglutination Inhibition Assay |
| HR | Hazard Ratio |
| IgG | immunoglobulin G |
| IM | Intramuscular |
| IRT | interactive response technology |
| ITT | Intent-to-treat |
| LBCI | Lower Bound Confidence Interval |
| LLOQ | lower limit of quantification |
| MedDRA | Medical Dictionary for Regulatory Activities |
| MAAE | medically-attended adverse event |
| MHRA | Medicines and healthcare products regulatory agency |
| PCR | polymerase chain reaction |
| PIMMC | potential immune-mediated medical conditions |
| PP-EFF | Per-Protocol efficacy |
| PP-IMM | Per-Protocol immunogenicity |
| PT | preferred term |
| SAE | serious adverse event |
| SARS-CoV-2 | severe acute respiratory syndrome coronavirus 2 |
| SARS-CoV-2 rS | SARS-CoV-2 Recombinant Spike Protein Nanoparticle Vaccine |
| SCR | seroconversion rate |
| SMC | safety monitoring committee |
| SOC | system organ class |
| TEAE | treatment-emergent adverse event |
| ULOQ | upper limit of quantification |
| VE | vaccine efficacy |
| WHODrug | World Health Organisation Drug Dictionary |

### 1. Introduction

This document outlines the statistical methods to be implemented in the analysis of data collected within the scope of Novavax, Inc., protocol 2019nCoV-302 (A Phase 3, Randomised, Observer-Blinded, Placebo-Controlled Trial to Evaluate the Efficacy and Safety of a SARS-CoV-2 Recombinant Spike Protein Nanoparticle Vaccine (SARS-CoV-2 rS) with Matrix-M1™ Adjuvant in Adult Subjects 18 to 84 Years of Age in the United Kingdom), Version 2.0, dated 23 October 2020. At the time of release of this SAP, version 3.0 dated 23Dec2020 of the study protocol had also been released. Only those parts of protocol version 3.0 that are relevant to the interim analysis of this study are incorporated into this SAP (the remaining impact of protocol version 3.0 will be addressed in a future SAP update).

Novavax, Inc. is developing a recombinant vaccine adjuvanted with the saponin-based Matrix-M1™ for active immunisation for the prevention of coronavirus disease 2019 (COVID-19) caused by severe acute respiratory syndrome coronavirus 2 (SARS-CoV-2).

The purpose of this study is to evaluate the efficacy, safety, and immunogenicity of SARS-CoV-2 rS with Matrix-M1 adjuvant in adults 18 to 84 years of age (inclusive). The study will be conducted at anticipated high COVID-19 transmission areas in the United Kingdom. The information provided in this study will inform progression of the study vaccine, to determine efficacy of the study vaccine to prevent COVID-19 in the general population, in subjects regardless of serostatus, in subjects who have required medical intervention, and in subjects with mild or asymptomatic infections. The study will determine the safety of the study vaccine to use in the general population and to ensure that it elicits a robust immune response.

The purpose of this statistical analysis plan is to define the planned statistical analysis of the study data consistent with the study objectives. This document does not fully cover the details of the planned analyses for the Safety Monitoring Committee (SMC). The SMC charter and a SMC Table, Listing, and Figure Shells document will outline the sequential nature of these reviews.

### 2. Objectives, Endpoints and Estimands

#### Table 1 Primary Objectives, Endpoints and Estimands

| Objectives | Endpoints | Estimands |
|---|---|---|
| To demonstrate the efficacy of SARS-CoV-2 rS with Matrix-M1 adjuvant in the prevention of PCR confirmed(by polymerase chain reaction [PCR] to SARS-CoV-2), symptomatic COVID-19, when given as a 2-dose vaccination regimen, as compared to placebo, in serologically negative (to SARS-CoV-2) adult subjects. | **PRIMARY ENDPOINT:** First occurrence of PCR confirmed **mild, moderate, or severe** COVID-19 (Appendix 12.3 and 12.4) with onset at least 7 days after second study vaccination (e.g., Day 28) in serologically negative (to SARS-CoV-2) adult subjects at baseline until the endpoint-driven efficacy analysis is triggered by the occurrence of a prespecified number of blinded endpoints. | **Primary Estimand 1a** (See Table 4 for estimand attributes and rationale for strategies)<br><br>Vaccine efficacy measured as VE (%) =100 × (1-RR) in SARS-CoV-2-naiive (confirmed serologically negative) adults **who receive both initial vaccination and booster after 3 weeks**. RR=Relative risk (SARS-CoV-2 Matrix-M1/placebo) of first occurrence of PCR-confirmed **mild, moderate or severe** COVID-19 disease with onset during a surveillance period from 7 days after second vaccination and up to 12 months after the 2nd vaccination or up to death from other causes or future use of an alternative COVID-19 vaccine or prohibited medication. |
| | | **Supportive Estimand 1b:** Vaccine efficacy measured as VE (%) =100 × (1-RR) in COVID-19 disease naiive adults **irrespective of compliance with second vaccination and actual serostatus at baseline**. RR=Relative risk ([SARS-CoV-2 Matrix-M1]/placebo) of first occurrence of PCR-confirmed **mild, moderate or severe** COVID-19 disease with onset during a surveillance period from **first vaccination** and up to 12 months after last vaccination or up to death from other causes or future use of an alternative COVID-19 vaccine, irrespective of use of prohibited medications. |

#### Table 2 Secondary and Exploratory Objective, Endpoint and Estimands

| Objectives | Endpoints | Estimands |
|---|---|---|
| To demonstrate the efficacy of SARS-CoV-2 rS with Matrix-M1 adjuvant in the prevention of PCR confirmed(by polymerase chain reaction [PCR] to SARS-CoV-2), symptomatic COVID-19, when given as a 2-dose vaccination | **KEY SECONDARY ENDPOINT:** First occurrence of PCR confirmed symptomatic **moderate or severe** COVID-19 (Appendix 12.3) with onset at least 7 days after second study | **Secondary Estimand 2a** (See Table 4 for estimand attributes and rationale for strategies)<br><br>Vaccine efficacy measured as VE (%) =100 × (1-RR) in SARS-CoV-2-naiive (confirmed serologically negative) adults **who receive both initial vaccination** |

| Objectives | Endpoints | Estimands |
|---|---|---|
| regimen, as compared to placebo, in adult subjects regardless of their serostatus at baseline | vaccination (e.g., Day 28) in serologically negative (to SARS-CoV-2) adult subjects at baseline until the endpoint-driven efficacy analysis is triggered by the occurrence of a prespecified number of blinded endpoints. | **and booster after 3 weeks** . RR=Relative risk ([SARS-CoV-2 Matrix-M1]/placebo) of first occurrence of PCR-confirmed **moderate or severe** COVID-19 disease with onset during a surveillance period from **7 days after second vaccination** and up to 12 months after the 2nd vaccination or up to death from other causes or future use of an alternative COVID-19 vaccine or prohibited medication. |
| | | **Supportive Estimand 2b:** Vaccine efficacy measured as VE (%) =100 × (1-RR) in COVID-19 disease naiive adults **irrespective of compliance with second vaccination and actual serostatus at baseline**. RR=Relative risk ([SARS-CoV-2 Matrix-M1]/placebo) of first occurrence of PCR-confirmed **moderate or severe** COVID-19 disease with onset during a surveillance period from **first vaccination** and up to 12 months after last vaccination or up to death from other causes or future use of an alternative COVID-19 vaccine, irrespective of use of other prohibited medications. |
| To demonstrate the efficacy of SARS-CoV-2 rS with Matrix-M1 adjuvant in the prevention of PCR confirmed(by polymerase chain reaction [PCR] to SARS-CoV-2), symptomatic COVID-19, when given as a 2-dose vaccination regimen, as compared to placebo, in adult subjects regardless of their serostatus at baseline | First occurrence of PCR confirmed symptomatic **severe** COVID-19 (Appendix 12.3) with onset at least 7 days after second study vaccination (e.g., Day 28) in serologically negative (to SARS-CoV-2) adult subjects at baseline until the endpoint-driven efficacy analysis is triggered by the occurrence of a prespecified number of blinded endpoints. | **Secondary Estimand 3a** (See Table 4 for estimand attributes and rationale for strategies)<br><br>Vaccine efficacy measured as VE (%) =100 × (1-RR) in SARS-CoV-2-naiive (confirmed serologically negative) adults **who receive both initial vaccination and booster after 3 weeks** . RR=Relative risk ([SARS-CoV-2 Matrix-M1]/placebo) of first occurrence of PCR-confirmed symptomatic **severe** COVID-19 disease with onset during a surveillance period from **7 days after second vaccination** and up to 12 months after the 2nd vaccination or up to death from other causes or future use of an alternative COVID-19 vaccine or prohibited medication. |
| | | **Supportive Estimand 3b:** Vaccine efficacy measured as VE (%) =100 × (1-RR) in COVID-19 disease naiive adults **irrespective of compliance with second vaccination and actual serostatus at baseline**. |

| **Objectives** | **Endpoints** | **Estimands** |
|---|---|---|
| | | RR=Relative risk ([SARS-CoV-2 Matrix-M1]/placebo) of first occurrence of PCR- confirmed **severe** COVID-19 disease with onset during a surveillance period from **first vaccination** and up to 12 months after last vaccination or up to death from other causes or future use of an alternative COVID-19 vaccine, irrespective of use of other prohibited medications. |
| To demonstrate the efficacy of SARS-CoV-2 rS with Matrix-M1 adjuvant in the prevention of PCR confirmed[by PCR to SARS-CoV-2], symptomatic COVID-19, when given as a 2-dose vaccination regimen, as compared to placebo, in adult subjects regardless of their serostatus at baseline | First occurrence of PCR confirmed symptomatic **mild**, **moderate, or severe** COVID-19, with onset at least 7 days after second study vaccination (e.g., Day 28) in adult subjects regardless of their serostatus at baseline. | No estimand specified. Summaries of confirmed **mild**, **moderate, or severe** COVID-19 disease emerging will be split by severity. In ITT, this will be further split **by serostatus at baseline and time of onset (prior to 14 days after first vaccination**, from 14 days after first vaccination to 6 days after 2nd vaccination or after 7 days after 2nd vaccination) and severity. |
| To assess the efficacy of SARS-CoV-2 rS with Matrix-M1 adjuvant on SARS-CoV-2 seropositive adult subjects requiring specific medical interventions as compared to placebo. | First occurrence of COVID-19 requiring hospitalisation, intensive care unit (ICU) admission or mechanical ventilation linked to any PCR confirmed(by PCR to SARS-CoV-2) COVID-19 with onset at least 7 days after second vaccination. | **Secondary Estimand 4a/**<br><br>**Supportive Estimand 4b** (Similar to 1a and 1b)<br><br>Vaccine efficacy measured as VE (%) =100 × (1-RR) where RR=Relative risk ([SARS-CoV-2 Matrix-M1]/placebo) of first occurrence of PCR- confirmed moderate or severe COVID-19 disease **requiring hospitalisation, intensive care unit (ICU) admission or mechanical ventilation.**<br><br>See Estmand 1a and 1b in Table 4 for estimand attributes and rationale for strategies. |
| To assess the efficacy of SARS-CoV-2 rS with Matrix-M1 adjuvant on mild COVID-19 symptoms as compared to placebo. | First occurrence of PCR confirmed **symptomatic mild** COVID-19 (with no progression to moderate or severe COVID-19 during the course of the COVID-19 episode) with onset at least 7 days after second study vaccination (e.g., Day 28) in adult subjects regardless of their serostatus at baseline. | No estimand specified. Summaries of confirmed symptomatic COVID-19 disease emerging will be split by serostatus at baseline and time of onset (prior to or after 7 days after 2nd vaccination) and severity. |

| **Objectives** | **Endpoints** | **Estimands** |
|---|---|---|
| To assess the efficacy of SARS-CoV-2 rS with Matrix-M1 adjuvant on mild COVID-19 symptoms as compared to placebo. | First occurrence of PCR confirmed **symptomatic or asymptomatic** COVID-19 with onset at least 7 days after second study vaccination (e.g., Day 28) in adult subjects regardless of their serostatus at baseline. | No estimand specified. Summaries of confirmed **asymptotic or symptomatic** COVID-19 disease emerging will be split by serostatus at baseline and time of onset (prior to or after 7 days after 2nd vaccination) and severity. |
| To assess the efficacy of SARS-CoV-2 rS with Matrix-M1 adjuvant on occurrence of asymptomatic or undetected infections with SARS-CoV-2 as compared to placebo. | First occurrence of laboratory-confirmed (by PCR or nucleocapsid (N)-protein serology to SARS-CoV-2) **symptomatic or asymptomatic** COVID-19 with onset at least 7 days after second study vaccination (e.g., Day 28) in adult subjects with serostatus negative at baseline. | **Secondary Estimand 5a/**<br><br>**Supportive Estimand 5b** (Similar to 1a/1b)<br><br>Vaccine efficacy measured as VE (%) =100 × (1-RR) where RR=Relative risk (SARS-CoV-2 Matrix-M1/placebo) of laboratory confirmed **symptomatic or asymptomatic** infection with SARS-CoV-2.<br><br>Full descriptions as per Estmand 1a and 1b in Table 4 for this endpoint with the same rationales for strategies of handling intercurrent events. Note that target population for 5b differs from 1b and is in SARS-CoV-2-naive adults (confirmed serologically negative) irrespective of compliance with second vaccination. |
| | First occurrence of PCR confirmed **symptomatic mild, moderate, or severe** COVID-19 with onset at least 14 days after first study vaccination (e.g., Day 14) in serologically negative (to SARS-CoV-2) adult subjects at baseline until the endpoint-driven efficacy analysis is triggered by the occurrence of a prespecified number of blinded endpoints | **Supportive Estimand 6** (Similar 2b)<br><br>Vaccine efficacy measured as VE (%) =100 × (1-RR) in COVID-19 disease-naiive adults **irrespective of compliance with second vaccination and actual serostatus at baseline**. RR=Relative risk ([SARS-CoV-2 Matrix-M1]/placebo) of first occurrence of PCR-confirmed **mild, moderate, or severe** COVID-19 disease with onset during a surveillance period from 14 days after **first vaccination** and up to 12 months after last vaccination or up to death from other causes or future use of an alternative COVID-19 vaccine, irrespective of use of prohibited medications |
| In a subset of adult subjects, to evaluate the immunogenicity of SARS-CoV-2 rS | Analysis of antibodies binding to the SARS-CoV-2 spike (S) protein by enzyme linked immunosorbent assay (ELISA) | Not defined |

| Objectives | Endpoints | Estimands |
|---|---|---|
| with Matrix-M1 adjuvant as compared to placebo. | at Day 0 (baseline) and Day 35 (14 days after second study vaccination).<br><br>Any occurrence of serologic conversion (by serology to SARS-CoV-2 nucleocapsid (N) protein) between baseline and 1 year after last study vaccination in adult subjects seronegative at baseline.<br><br>Evidence of seroconversion as demonstrated by antibodies to the SARS-CoV-2 S protein by ELISA at Day 35 (14 days after second study vaccination).<br><br>Cell-mediated immune responses (for example, as measured by enzyme-linked immune absorbent spot (ELISpot) ± intracellular cytokine staining) on Days 0 (baseline) and 35 (14 days after second study vaccination).<br><br>SARS-CoV-2 neutralisation as measured by virus neutralisation assay (VNA; wildtype virus and/or pseudovirion expressing SARS-CoV-2 S protein) at Day 0 (baseline) and Day 35 (14 days after second study vaccination). | |
| To evaluate safety in terms of Serious Adverse Events (SAEs) and medically-attended adverse events (MAAEs) related to study vaccination in all adult subjects during the entire study period. | The occurrence and relationship to study vaccination of SAEs and MAAEs related to study vaccination (in all adult subjects) during the entire study period. | **Estimand 7**<br><br>Percentage of vaccinated healthy adults who would develop MAAEs, etc, within timeframe. A treatment policy strategy is used for assessing safety irrespective |

| Objectives | Endpoints | Estimands |
|---|---|---|
| | | of a current (or prior) infection at time of first vaccination or missed second vaccination. |
| To evaluate safety in terms of adverse events of special interest (AESI), which encompasses potential immune-mediated medical conditions (PIMMCs) and AESIs relevant to COVID-19 including possible vaccine-enhanced disease, in all adult subjects at any time after the first dose. | As above with specific interest in AESI and PIMMCs. | Not defined. |
| In a subset of adult subjects, to evaluate the safety and reactogenicity in terms of solicited local and systemic adverse events (AEs) for 7 days after each study vaccination. | The occurrence and severity of reactogenicity in terms of solicited local and systemic<br><br>AEs (in all sub-study participants) for 7 days after each study vaccination | Not defined. |
| To evaluate safety in terms of all MAAEs for 14 days after second vaccination and unsolicited AEs for 21 days after first study vaccination and 28 days after second study vaccination. | The occurrence, severity, and relationship to study vaccination of all MAAEs for<br><br>14 days after second vaccination and unsolicited AEs (in all adult participants) for<br><br>21 days after first study vaccination and 28 days after second study vaccination. | Not defined. |
| In a subset of adult subjects, to evaluate the safety and immunogenicity of SARS-CoV-2 rS with Matrix-M1 adjuvant when co-administered with a licensed seasonal influenza vaccine. | Analysis of the safety and immunogenicity of SARS-CoV-2 rS with Matrix-M1 adjuvant when co-administered with seasonal influenza vaccine in a subset population | Not defined – See comment |
| Analysis of the efficacy of SARS-CoV-2 rS with Matrix-M1 adjuvant in participants with asymptomatic COVID-19 who test positive for the disease by | | Not defined |

| Objectives | Endpoints | Estimands |
|---|---|---|
| SARS-CoV-2 N protein serology but have no accompanying symptoms. | | |

### 3. Investigational Plan

#### 3.1. Overall Study Design and Plan

This is a Phase 3, multi-centre, randomised, observer-blind, placebo-controlled study evaluating the efficacy, safety, and immunogenicity of SARS-CoV-2 rS with Matrix-M1adjuvant in adult subjects 18 to 84 years of age (inclusive) in the UK. Every effort will be made to identify regions of high SARS-CoV-2 activity, and populations within these regions who are at high risk of exposure to the virus will be enrolled such that the incidence of COVID-19 in the study will be higher than 1% and the sample size and enrolment period may be reduced.

After signing informed consent, subjects may be screened within a window of up to approximately 30 days. Nose/throat samples may be taken during the screening period to detect SARS-CoV-2 by PCR, if the subject has any COVID-19 symptoms or significant exposure history. Approximately 15,000 male and female adult subjects 18 to 84 years of age (inclusive) with and without relevant comorbidities is planned for the study. An effort will be made to enrol a target of at least 25% of subjects who are ≥ 65 years of age as well as prioritising other groups that are most affected by COVID-19, including racial and ethnic minorities. This will be under the assumption that the annual incidence of symptomatic COVID-19 will be approximately 1% to 5%.

The study will consist of the screening period (Days -30 to 0); study vaccination Days 0 and 21 (+7-day window); and outpatient study visits on Day 0, Day 21 (+ 7 days), and Day 35 (14 days minimum after second study vaccination [+ 7 days]); and at 3, 6, and 12 months (± 15 days) after last study vaccination.

The duration of individual participation, including screening, will be a maximum of 1 year after second study vaccination (Day 386 ± 15 days). This will be an event-driven study for the assessment of vaccine efficacy (VE) and will end when a sufficient number of events have been observed, yet all subjects will be followed for the entire study duration for safety endpoints. The EOS analysis will be performed when the last subject reaches 12 months after the last study vaccination.

A licensed seasonal influenza co-administration sub-study will be conducted in the first 400 subjects who meet the additional inclusion criteria for this study. Subjects may be enrolled at select study sites due to the availability of seasonal influenza vaccine. After being randomised to receive intramuscular (IM) injections of SARS-CoV-2 rS with Matrix-M1 adjuvant or placebo, sub-study subjects will receive a licensed seasonal influenza vaccine on Day 0 in the opposite deltoid. These subjects will be part of the solicited AE safety subset analysis.

Given that this study and others will enrol during the influenza virus season and that the highest at-risk population, namely the elderly and adults with comorbid conditions for influenza mirrors that of SARS-CoV-2, a co-vaccination sub-group is included. This sub-group will undergo the same safety and efficacy evaluations as the entire study population. In addition, the entire sub-study will have reactogenicity assessed. This group must not have had a current season licensed influenza vaccine and have no prior history of allergy or severe reaction to seasonal influenza vaccine. To assess the possible impact of the study vaccine on the immunogenicity of the influenza vaccine, this group will also have a haemagglutination inhibition assay (HAI) performed.

Subjects will be monitored for COVID-19 throughout the study, a COVID-19 Surveillance Visit will be triggered by every episode of a new onset of symptoms of suspected COVID-19.

**Study Vaccines**

Study vaccinations (5-µg SARS-CoV-2 rS with 50-µg Matrix-M1 adjuvant or placebo [saline]) will comprise 2 intramuscular (IM) injections on Days 0 and 21, ideally in alternating deltoids. For blinding purposes, all subjects will be vaccinated using the same injection volume (i.e., 0.5 mL).

The seasonal influenza vaccine co-administration sub-study will comprise a single IM injection (0.5 mL) of a licensed influenza vaccine on Day 0, ideally in the opposite deltoid of the study vaccine. Flucelvax Quadrivalent will be given to those 18 to 64 years of age, and an adjuvanted trivalent influenza vaccine will be given to those ⩾ 65 years of age. Whenever possible the right deltoid will be used for the influenza vaccine and the left deltoid for the study vaccine.

All study vaccinations will be administered on an outpatient basis by designated site personnel in a way to maintain the blind. The influenza vaccine will not require blinding. Any pharmacy preparation with unblinded product will require unblinded site personnel who will not otherwise be involved in the study procedures or observation of subjects.

The schedule of events is provided in Appendix 1

**Vaccination Pause Rules**

Vaccination pause rules based on reactogenicity, AEs, and SAEs related to study participation will be in place to monitor subject safety during the study.

AEs meeting any one of the following criteria will result in a hold being placed on subsequent study vaccinations pending further review by the SMC and the sponsor:

- Any toxicity grade 3 or higher (severe or potentially life-threatening) solicited (local or systemic) single AE term occurring in ≥ 10% of subjects (after a minimum of 100 subjects are enrolled) in the SARS-CoV-2 rS with Matrix-M1 adjuvant group within the first 7 days after study vaccination.

- Any severe unsolicited single AE preferred term for which the investigator assesses as related that occurs in ≥ 5% of subjects (after a minimum of 100 subjects are enrolled) in the SARS-CoV-2 rS with Matrix-M1 adjuvant group, within the first 7 days after study vaccination.

In addition, any SAE assessed as related to vaccine (final assessment by the sponsor) will be reported by the sponsor to the SMC Chair as soon as possible, and within 24 hours of the sponsor's awareness of the event. Based on this initial report of the event to the SMC Chair, the Chair may advise the sponsor to immediately pause enrolment and further dosing in either some or all subjects in the study and to convene an ad hoc meeting, or make alternative recommendations. The SMC Charter defines processes for how this review will occur and how the Chair's recommendations will be documented.

The sponsor, along with medical monitor, may request an SMC review for any safety concerns that may arise in the study, even if they are not associated with any specific pause rule.

The subject influenza vaccine co-administration study will utilise the same pause rules with one difference due to the expectation of greater reactogenicity and the small size:

- Any grade 3 or higher (severe or potentially life-threatening) solicited (local or systemic) single AE term occurring in ⩾ 20% of participants in the SARS-CoV-2 rS with Matrix-M1 adjuvant plus seasonal influenza vaccine group, within the first 7 days after study vaccination.

##### 3.1. Individual Unblinding

Unblinding of treatment assignment may occur in order to allow a participant to make an informed decision regarding receipt of an approved or deployed SARS-CoV-2 vaccine. Participants who choose to receive an approved or deployed SARS-CoV-2 vaccine as per UK government guidance will be encouraged to remain in the study for scheduled safety assessments.

In order to manage unblinded participants, unblinding will result in censoring of all efficacy and immunogenicity endpoints. Unblinding will also result in censoring from reactogenicity analyses and analyses of unsolicited AEs. This will be addressed via sensitivity analyses - i.e. the main presentations of data based on the Safety Analysis set will include all patient data, with supporting presentations to exclude the data post unbinding.

See Sections 4.4 of handling of unblinded subjects in the analysis sets and Section 10 for handling unblinded subjects in the Interim Analyses.

#### 4. General Statistical Considerations

All data collected will be presented in data listings. Data from subjects excluded from an analysis set will be presented in the data listings but not included in the calculation of summary statistics for that analysis set. Data used for summaries will be indicated in the listings.

For categorical variables, counts and percentages of subjects will be presented. Continuous variables will be summarised using descriptive statistics (number of subjects, mean, standard deviation, median, minimum, and maximum).

When count data are presented, the percentage will be suppressed when the count is zero in order to draw attention to the non-zero counts. A row denoted “Missing” will be included in count tabulations where specified on the shells to account for missing values. The denominator for all percentages will be the number of subjects in that study vaccine within the population of interest, unless otherwise specified. Non-zero percentages will be rounded to one decimal place, except 100% will be displayed without any decimal places.

For the summary statistics of all continuous variables unless otherwise specified, minimum and maximum will be displayed to the same level of precision as reported. Mean, LSMeans and median will be displayed to one level of precision greater than the data collected. Standard deviation and standard error will be displayed to two levels of precision greater than the data collected.

All confidence intervals (CI) will be 2-sided and performed using a 5% significance level unless stated otherwise. All p-values will be presented to 3 decimal places and values less than 0.001 or greater than 0.999 will be presented as <0.001 and >0.999, respectively.

Unless specified otherwise, baseline will be defined as the last non-missing assessment prior to the first study vaccine administration. Both scheduled and unscheduled visits and assessments will be used in determining baseline.

Baseline serostatus will be defined based on baseline anti N-protein results only.

Summaries by treatment group and overall will be presented for tables, for all subgroups analyses see Section 8.10.

Study day will be calculated relative to the first vaccination date as:

- Day 0 is defined as the first vaccination dose.
- Study Day = Assessment Date – First Vaccination Date.

For Geometric Mean Fold Rise (GMFR), seroconversion rate (SCR) calculations, antibody values reported as below the lower limit of quantification (LLOQ) will be replaced by 0.5 × LLOQ as applicable. Values that are greater than the upper limit of quantification

(ULOQ) will be replaced by the ULOQ as applicable. Missing results will not be imputed. No other imputations will be performed.

For HAI, seroconversion is defined as either a baseline reciprocal (Day 0) titre of < 10 and a post-vaccination reciprocal titre >= 40, or a baseline titre of >= 10 and a post-vaccination titre >= 4-fold higher. For all other immunogencity data, seroconversion is defined as post vaccination >= 4-fold higher than baseline.

##### 4.1. Sample Size

This study is designed to enrol approximately 15,000 subjects, randomised 1:1 into the 2 treatment groups. The sample size is driven by the total number of events expected to achieve statistical significance for the primary efficacy endpoint – a target of 100 mild, moderate, or severe COVID-19 cases. The target number of events of 100 was chosen to provide 95% power for 70%  or higher vaccine efficacy (VE). A single interim analysis of efficacy will be conducted based on the accumulation of approximately 50% (50 events) of the total anticipated primary endpoint using Pocock boundary conditions. Power calculations were performed using 10,000 simulated trials that were created under various assumptions of VEs and analysed using methods described in the "Primary efficacy analysis" section without covariates. A 90% evaluability rate for the per-protocol (PP) efficacy (PP-EFF) population (assuming 10% unevaluable due to attrition and/or baseline-seropositive participants) was used for the power calculations. All simulations were performed in SAS V9.4 and the power under various vaccine efficacy assumptions is shown in Appendix 12.2.

##### 4.2. Randomisation, Stratification and Blinding

Subjects will be randomly assigned in a blinded manner using the centralised Interactive Response Technology (IRT) according to pre-generated randomisation schedules. Subjects will be randomised in a 1:1 ratio via block randomisation to receive 2 IM injections of the SARS-CoV-2 rS with Matrix-M1 adjuvant or placebo. Randomisation will be stratified by site and by age ≥ 65 years. The first approximately 400 subjects who qualify for the seasonal influenza vaccine co-administration sub-study (which may be at select study locations only) will be assigned prior to randomisation. These subjects will be part of the Solicited AE Safety Subset Analysis Set. Details regarding the IRT process will be provided separately to the sites.

This is an observer-blinded study. To maintain the blind, placebo vaccination via IM route will be included and unblinded site personnel will manage study vaccine logistics, preparation, and administration so as to maintain the blind from the remainder of the site personnel and subjects. The unblinded site personnel will not be involved in study related assessments or have subject contact for data collection following study vaccine administration.

Seasonal influenza vaccine will be administered in an open-label manner.

#### 4.3. Intercurrent Event and Estimands

**Table 3 Intercurrent Event**

| Label | Intercurrent Event Type |
|---|---|
| IcEv1a (COVID-19 death) | Death due to COVID-19 disease or complications of SARS-CoV-2 infection. |
| IcEv1b (Unrelated death) | Death due to other cause, unrelated to SARS-CoV-2 infection or COVID-19 disease |
| IcEv3 (Missed 2nd Vaccine) | Second scheduled vaccination at Day 21 (+7 days) not received. |
| IcEv4a (Alternative vaccine) | Use of alternative SARS-CoV-2 vaccine. |
| IcEv4b (Vaccine interference) | Use of any live vaccine within 4 weeks of any vaccination or any vaccine (excluding flu) within 2 weeks prior to first vaccine and 4 weeks after 2nd vaccination. |
| IcEv4c (Prohibited medications) | Use of prohibited medications deemed to impact on efficacy. |
| IcEv4d (Other Immune modifying) | Use of any other (non-prohibited) immune modifying drugs in treatment of emerging conditions. |
| IcEv5 (Prior Infection) | Laboratory confirmed SARS-CoV-2 infection or antibodies to SARS-CoV-2 on or prior to Day 0. This is added for clarity since it is possible errors might be made (which come to light after vaccination) in vaccinating people who are not naiive to SAR-CoV-2 as per the intended target population for Estimands 1a-5a. |
| IcEv6a (Day 0-27 infection) | Develops a new positive PCR-confirmed SARS-CoV-2 infection occurring between first vaccination and Second dose of Vaccination +6 days (i.e. prior to when the vaccination series is expected to be fully protective). |
| IcEv6b (Day 28-12 month infection) | Develops a new positive PCR-confirmed SARS-CoV-2 infection occurring from 2nd dose of vaccination +7 days to end of study. |

#### Table 4 Summary of Estimands 1a/1b

| **Estimand Label** | **Primary Estimand 1a** | **Supportive Estimand 1b** |
|---|---|---|
| **Estimand Description** | Vaccine efficacy measured as VE (%) =100 × (1-RR) in SARS-CoV-2-naiive (confirmed serologically negative) adults who receive both initial vaccination and booster after 3 weeks. RR=Relative risk ([SARS-CoV-2 Matrix-M1]/placebo) of PCR confirmed **mild, moderate or severe** COVID-19 disease with onset during a surveillance period from 7 days after second vaccination and up to 12 months after the 2nd vaccination or up to death from other causes or future use of an alternative COVID-19 vaccine or prohibited medication. | Vaccine efficacy measured as VE (%) =100 × (1-RR) in COVID-19 disease naiive adults **irrespective of compliance with second vaccination and actual serostatus at baseline**. RR=Relative risk ([SARS-CoV-2 Matrix-M1]/placebo) of first occurrence of PCR-confirmed **mild, moderate or severe** COVID-19 disease with onset during a surveillance period from **first vaccination** and up to 12 months after last vaccination or up to death from other causes or future use of an alternative COVID-19 vaccine, irrespective of use of prohibited medications. |
| **Target Population** | Adults aged 18 to 84 years who are naiive to SARS-COV-2 prior to vaccination, receive both vaccinations and without confirmed SARS-Cov-2 infection up to 6 days after 2$^{nd}$ vaccination (expected to be Day 27) | Adults aged 18 to 84 years naiive to known COVID-19 disease prior to vaccination. |
| **Endpoint** | Occurrence of PCR confirmed(by PCR to SARS-CoV-2) mild, moderate or severe COVID-19 disease with event onset between 7 days and 12 months after second study vaccination denoted as 0 (no infection) or 1 (one or more infections) where the surveillance time is from 7 days following last vaccination up to onset of event, EOS or censoring. See Section 4.4.1. | Occurrence of PCR confirmed(by PCR to SARS-CoV-2) COVID-19 disease with event onset after first vaccination denoted as 0 (no infection) or 1 (one or more infections) where the surveillance time is from first vaccination to EOS or censoring. See Section 4.4.1. |
| **Treatment Conditions** | SARS-CoV-2 rS with Matrix-M1 vs Placebo. | SARS-CoV-2 rS with Matrix-M1 vs Placebo. |
| **Population-Level Summary** | Vaccine efficacy defined as 100 x (1-RR) where RR = Relative Risk calculated as ratio of incidence rates | As per Estimand 1a. |

| | | |
|---|---|---|
| | (SARS CoV-2 Rs with Matrix-M1 / Placebo). | |
| **Intercurrent Event Strategy** | | |
| **IcEv1a (COVID-19 death)** | Composite | Composite |
| **IcEv1b (Unrelated death)** | Hypothetical | Hypothetical |
| **IcEv3 (Missed 2nd Vaccine)** | Principal stratum | Treatment policy |
| **IcE4a (Alternative vaccine)** | Hypothetical | Hypothetical |
| **IcE4b (Vaccine interference)** | Principal stratum | Treatment policy |
| **IcE4c (Prohibited medications)** | Hypothetical | Treatment policy |
| **IcE4d (Other immune modifying)** | Treatment policy | Treatment policy |
| **IcEv5 (Prior infection)** | Principal stratum | Treatment Policy |
| **IcEv6a (Day 0-27 infection)** | Principal stratum | Composite |
| **IcEv6b (Day 28-365 infection)** | Compoiste | Composite |
| **Rationale for Strategies** | This estimand seeks to understand efficacy during a surveillance period (7 days to 12 months after 2nd vaccination) which starts after the vaccine is considered to have stimulated an immune response in adults naiive to SARS-COV-2 infection (confirmed seronegative) who comply with dosing schedule and do not start an infection prior to 7 days after 2nd vaccination.<br>A hypothetical strategy is used for unrelated deaths and significant deviations (such as use of alternative vaccines and prohibited medications) so that interest lies in the hypothetical situation that these do not occur. | A treatment policy strategy is used for following up efficacy irrespective of whether they missed second vaccination and also including all subjects vaccinated irrespective of whether they subsequently were found not to strictly meet the criteria of the target population (i.e. found to be seropositive at vaccination). There is interest in understand efficacy in the light of poor compliance which may happen in clinically practice and may reflect reactions to the first vaccination as well as poor compliance for unrelated reasons.<br>The surveillance period starts from Day 0 because whilst the vaccine will not |

| | | |
|---|---|---|
| | | protect against early infections, the risk of early infection should be balanced between groups.<br>The hypothetical strategy is employed for other intercurrent events (unrelated deaths, alternative vaccine) which match Estimand1a. |

#### 4.4. Censoring and Surveillance Time

##### 4.4.1. Censoring

Subjects are censored, in efficacy analyses, at the earliest of

(a) follow up contact at 12 months after the second vaccination
(b) data cut-off date (for interim analysis – see Section **Error! Reference source not found.**),
(c) unblinding for any reason (including intended receipt of alternative COVID-19 vaccine)
(d) early withdrawal from study
(e) death

In addition, in the analyses of the PP-EFF set, there is additional censoring at the date of a major protocol deviations (agreed prior to breaking the study blind).

Events with onset after the censoring date are excluded.

##### 4.4.2. Surveillance Time

The surveillance time will vary according to the surveillance period defined in the estimand which will have a surveillance start date of:

- 7 days after the 2$^{nd}$ dose of vaccination (e.g. Estimands 1a, 2a, 3a, 4a, 5a), or
- the first vaccination (e.g. Estimands 1b, 2b, 3b, 4b, 5b), or
- 14 days after the first vaccination (e.g. Estimand 6).

Surveillance will have an end date of the censoring date, for those without an event.

For those with an uncensored event, surveillance end date will depend on the estimand definition as either:

- the onset of illness (any symptom irrespective of severity level) for those estimands evaluating first occurrence of PCR confirmed symptomatic COVID-19 disease events is defined as the start of illness episode symptoms (irrespective of severity level)

- the onset of event for those estimands evaluating asymptomatic and symptomatic infections is defined as the earliest post-vaccination date of sample which returned a positive result (PCR or anti-N).

Surveillance time is calculated as surveillance end date - surveillance start date+1.

#### 4.5. Analysis Sets

The analysis sets that will be analysed in this study are:

- All Screened Subjects
- All Randomised Subjects
- Intent-to-Treat (ITT) Analysis Set
    - Anti-S Protein Serology Subset
    - Neutralisation Assay Subset
    - Cell-mediated Assay Subset
    - Seasonal Influenza Vaccine Sub-study
- Per-Protocol Efficacy (PP-EFF) Analysis Set
- Per-Protocol Immunogenicity (PP-IMM) Analysis Set
    - Anti-S Protein Serology Subset
    - Neutralisation Assay Subset
    - Cell-mediated Assay Subset
    - Seasonal Influenza Vaccine Sub-study
- Safety Analysis Set
    - Solicited AE Safety Subset Analysis Set

The safety analysis will be based on the Safety Analysis Set, with exception of the data specific to each of the above subsets/sub-study. The primary efficacy analyses will be based on the PP-EFF and supported by ITT Analysis Sets respectively. The secondary immunogenicity analyses and efficacy analyses will be based on PP-IMM and PP-EFF, respectively, and supported by ITT Analysis Sets. The exploratory immunogenicity and exploratory efficacy analyses will be based on ITT.

##### 4.5.1. All Screened Subjects Set

All Screened Subject Analysis Set includes all subjects who are screened.

##### 4.5.2. All Randomised Subjects Set

The all-randomised set will include all participants who were randomised regardless of the receipt of any study vaccine (SARS-CoV-2 rS with Matrix-M1 adjuvant or placebo). The all-randomised set will be used for the subject disposition summaries.

##### 4.5.3. Intent to Treat Analysis Set (ITT)

The Intent-To-Treat (ITT) Analysis Set will include all subjects who are randomised and receive at least 1 dose study vaccine (SARS-CoV-2 rS with Matrix-M1 adjuvant or placebo), regardless of protocol violations or missing data. Data may be censored (see Section 4.4.1).

Data will be analysed according to the treatment group randomised. Within the ITT Analysis Set there are three subsets defined:

###### 4.5.3.1. Anti-S Protein Serology Subset

All subjects in the ITT Analysis Set who were tested for Anti-S protein serology using ELISA prior to study vaccination will be included in this subset.

###### 4.5.3.2. Neutralisation Assay Subset

All subjects in the ITT Analysis Set who were tested for neutralisation prior to study vaccination will be included in this subset.

###### 4.5.3.3. Cell-mediated Assay Subset

All subjects in the ITT Analysis Set who have their cell-mediated immune response assessed by ELISpot ± intracellular cytokine staining prior to study vaccination will be included in this subset.

###### 4.5.3.4. Seasonal Influenza Vaccine Sub-study

All subjects in the ITT Analysis Set who have been randomised and vaccinated subjects receiving co-administered licensed seasonal influenza vaccine and the study vaccine will be included in the sub-study. This will be the initial approximate 400 subjects who meet additional inclusion criteria for this sub-study.

##### 4.5.4. Per-Protocol Efficacy Analysis Set (PP-EFF)

Subjects will be excluded from the Per-Protocol Efficacy (PP-EFF) Analysis Set if

- they have a laboratory confirmed prior SAR-COV-2 infection by Anti-N antibody test (note that Anti-S results will not be considered as these are only measured in a subset of subjects).
- they have a laboratory confirmed current SAR-COV-2 infection by any validated or licensed PCR test with symptom onset or positive PCR swab occurring up to 6 days after second study vaccination (i.e. exclusion depends on the earlier of the symptom onset date or positive swab date rather than just date of laboratory result).
- they do not receive both doses of study vaccine (SARS-CoV-2 rS with Matrix-M1 adjuvant or placebo).

- they are mis-dosed (i.e. received a different study vaccine to that which they were randomised to receive).
- they have a major study deviation (e.g. incorrect volume of vaccine administered) affecting the primary efficacy outcome.

In addition, data for subjects included in PP-EFF will be censored at the date of major protocol deviations. The review and determination of major protocol deviations, which lead to exclusion or censoring, will be carried out in a blinded fashion by the study clinician prior to unblinding. See Section 4.4.1 for more detail on censoring.

Data will be analysed according to the study vaccine group as randomised.

##### 4.5.5. Per-Protocol Immunogenicity Analysis Set (PP-IMM)

The Per-Protocol Immunogenicity (PP-IMM) Analysis Set will include subjects who receive both scheduled dose(s) of study vaccine (SARS-CoV-2 rS with Matrix-M1 adjuvant or placebo).

For each visit, the SARS-CoV-2 unexposed population will also exclude any illness episodes with positive SARS-CoV-2 by any validated PCR or antibody test prior to each visit. Prior exposed subjects will be determined using baseline SARS-CoV-2 immunity defined as positive SARS-CoV-2 by PCR or serum IgG antibody at baseline, or positive SARS-CoV-2 by qualitative PCR until Day 35, according to the specified analysis. Immune measurements (ELISA) will be conducted on serum (immunoglobulin G [IgG]) for SARS-CoV-2 anti-S protein serology in approximately 900 participants in the Anti-S Protein Serology Subset and on anti-N protein serology (immunoglobulin G [IgG] for SARS-CoV-2 anti-N protein) in all participants.

###### 4.5.5.1. Anti-S Protein Serology Subset

All subjects in the PP-IMM Analysis Set who were tested for Anti-S protein serology using ELISA prior to study vaccination will be included in this subset.

###### 4.5.5.2. Neutralisation Assay Subset

All subjects in the PP-IMM Analysis Set who were tested for neutralisation prior to study vaccination will be included in this subset.

###### 4.5.5.3. Cell-mediated Assay Subset

All subjects in the PP-IMM Analysis Set who have their cell-mediated immune response assessed by ELISpot ± intracellular cytokine staining prior to study vaccination will be included in this subset.

###### 4.5.5.4. Seasonal Influenza Vaccine Sub-study

All subjects in the PP-IMM Analysis Set who have been randomised and vaccinated subjects receiving co-administered licensed seasonal influenza vaccine and the study vaccine will be included in the sub-study.

##### 4.5.6. Safety Analysis Set

The Safety Analysis Set will include all subjects who receive at least 1 dose of study vaccine (SARS-CoV-2 rS with Matrix-M1 adjuvant or placebo). Safety Analysis Set Data will be analysed according to the study vaccine received. Subjects who are mis-dosed (i.e. receive a different study vaccine(s) to that which they were randomised to receive) will be analysed according to the first study vaccine received (regardless of whether this is the same as the second study vaccine received).

##### 4.5.7. Solicited AE Safety Subset Analysis Set

The Solicited AE Safety Subset Analysis Set will be a subset of the Safety Analysis Set and will be analysed according to the study vaccine actually received. It will comprise the following: the Seasonal Influenza Vaccine Sub-study subjects and the initial approximate 2,000 subjects randomised who receive a vaccine.

### 5. Subject Disposition and Protocol Deviations

#### 5.1. Disposition

Subject disposition will be summarised for each treatment group and overall for the All Randomised Subjects Set.

The number of subjects who are randomised in the study to each treatment group and the counts and percentage of subjects who complete the study (i.e. study participation until the End of Study (EOS) Visit without early termination) will be presented. Counts and percentage of subjects who have completed, are ongoing, withdraw or discontinue from the study, and the reason for withdrawal or discontinuation, will also be summarised. In addition, when an analysis is requested or for the planned SMC's, the counts and percentage of subjects who are ongoing at the time of the data extraction will also be summarised. The counts and percentages of subjects in each analysis set (excluding All Screened Subject Analysis Set) will be presented in a summary table.

Subject disposition data, analysis sets, and randomisation data will be presented in data listings. Screen failure data will be summarised and will be presented in data listings.

#### 5.2. Protocol Deviations

A protocol deviation is any change, divergence, or departure from the study design or procedures defined in the protocol. A significant deviation is a subset of protocol deviations that leads to a subject being discontinued from the study or significantly affects the subject's rights, safety, or well-being and/or the completeness, accuracy, and reliability of the study data, especially as it pertains to elicited immune response to planned study vaccination. A significant deviation can include non-adherence to inclusion or exclusion criteria or non-adherence to regulatory authority including International Council for Harmonisation E6 (R2) guidelines.

Protocol deviations will be documented by the clinical monitor throughout the course of monitoring visits. The investigator will be notified in writing by the monitor of deviations. The Health Research Ethics Committee (HREC) should be notified of all protocol deviations, if appropriate, in a timely manner.

All protocol deviations will be presented in a data listing with another displaying only significant deviations. This latter listing will include a column to indicate whether each significant deviation has led to exclusion from any analysis sets or censoring (for PP-EFF analysis set only). Inclusion/Exclusion for analysis sets will be presented in a data listing. Details of study entry criteria deviations will be presented in a separate data listing. Significant deviations will be summarised by treatment group for each site for the ITT Analysis Set. An additional summary will be presented for major protocol deviations leading to exclusion from analysis sets, or leading to censoring of study data for analysis.

### 6. Demographics and Baseline Characteristics

#### 6.1. Demographics

Demographic and baseline characteristics will be summarised for each treatment group and overall for the ITT Analysis Set, Safety Analysis Set, PP-EFF Analysis Set, PP-IMM Anti-S Protein Serology Subset, PP-IMM Cell-Mediated Assay Subset and PP-IMM Analysis Set (Seasonal Influenza Vaccine Subset).

The demographic characteristics consist of age (years), age-group (<65 yrs, >=65 yrs) sex, child-bearing potential, birth control method, race, and ethnicity. The baseline characteristics consist of baseline weight (kg), baseline height (cm), baseline body mass index (BMI, $kg/m^2$), PCR (+/−) at Day 0 and 21, SARS-CoV-2 serostatus based on Anti-N only (positive/negative/missing sample) at Day 0.

Subject demographic and baseline characteristics will be presented in a data listing.

#### 6.2. Medical History

Medical history will be classified by system organ class (SOC) and preferred term (PT) using MedDRA (Version 23.1 or later).

Medical history will be summarised by SOC and PT will be summarised for each treatment group and overall for the Safety Analysis Set. Medical history will be presented in a data listing.

### 7. Treatments, Medications, and Recent Vaccinations

#### 7.1. Recent Vaccinations and Concomitant Medications

Administration of medications, therapies, or vaccines will be recorded in the electronic case report form (eCRF). Recent vaccinations will include vaccinations taken ≤90 days prior to taking study vaccination. Concomitant medications will include all medications (including vaccines) taken by the subject from the time of signing the informed consent form until EOS (or until the early termination visit if prior to that time). Prescription and over the counter drugs, as well as herbals, vitamins, and supplements, will be included.

Recent vaccinations and concomitant medications and therapies will be summarised for the Safety Analysis Set by treatment group, anatomical therapeutic chemical (ATC Levels 1-2) and preferred drug name as coded using the World Health Organisation Drug Dictionary (currently WHODrug Global-B3 Sep 2020 however this will be updated during the course of the study).

For the purpose of inclusion in recent vaccinations and concomitant medications tables, incomplete start and stop dates will be imputed according to the below rules (where UK, UKN, and UNKN indicate unknown or missing day, month, and year, respectively):

<u>Missing Start Dates</u>

- UK-MMM-YYYY: Assume 01-MMM-YYYY, but if month and year are the same as the first study vaccination month and year, then assume the date of first vaccination
- UK-UKN-YYYY: Assume 01-JAN-YYYY, but if year is the same as the first study vaccination year, then assume the date of first study vaccination
- UK-UKN-UNKN: Assume date of first study vaccination

<u>Missing Stop Dates</u>

- UK-MMM-YYYY: Assume the last day of the month
- UK-UKN-YYYY: Assume 31-DEC-YYYY
- UK-UKN-UNKN: Do not impute and assume ongoing

All recent vaccinations and concomitant medications will be presented in a data listing.

#### 7.2. Medical and Surgical Treatment Procedures

All medical and surgical treatment procedures (conducted during the study period) will be coded using MedDRA (Version 23.1 or later). Medical and surgical treatment procedures will be summarised by SOC, PT and treatment group for the Safety Analysis Set with all procedures presented in a data listing.

#### 7.3. Study Vaccine Administration

Study vaccinations (SARS-CoV-2 rS with Matrix-M1 adjuvant or placebo) will comprise 2 IM injections (Day 0 and Day 21), ideally in alternating deltoids. For blinding purposes, all subjects will be vaccinated using the same injection volume (i.e., 0.5 mL). The dose level will be 5 µg SARS-CoV-2 rS with 50 µg Matrix-M1 adjuvant (co-formulated in a single vial); placebo will be saline for injection. The subjects in the Seasonal Influenza Vaccine Sub-study will be vaccinated using a single IM injection (0.5 mL) of a licensed influenza vaccine on Day 0, ideally in the opposite deltoid of the study vaccine.

Study vaccinations will be summarised for the Safety Analysis Set including the counts and percentage of subjects receiving each dose, and if administered per protocol (yes, no), as well as a duration of follow-up for subjects receiving the 2$^{nd}$ vaccination. Similarly, a summary table for the Seasonal Influenza Vaccine Sub-study will present the counts and percentages of subjects receiving the influenza vaccine at Day 0, and whether trivalent or quadrivalent.

All study drug administration data will be presented in a data listing.

### 8. Efficacy Analysis and Immunogenicity Analysis

#### 8.1. Symptoms of Suspected COVID-19

Subjects with symptoms of suspected COVID-19 will

- take their temperature daily for 10 days
- self swab themselves daily for 3 days
- and complete a FLU-PRO diary that will collect their symptoms See Appendix C Severity of COVID-19 Symptoms

Every episode of a "new onset" of symptoms of suspected COVID-19 will trigger a COVID-19 Surveillance Visit (Initial and Follow-Up) or phone call. A "New onset" will require at least a 7-day period symptom-free prior to the event to differentiate a specific episode from any prior illness. For the definitions around the primary estimand 1a and supportive endpoint See Table 4.

#### 8.2. Asymptomatic Infections

**Asymptomatic infection** will be defined as a SARS-CoV-2 infection confirmed by N protein serology without meeting the criteria for PCR confirmed mild, moderate or severe COVID disease. This can only be assessed in those who have serostatus negative at baseline.

The event onset date will not be known exactly but for the purposes of calculating the surveillance time, the event onset date will be from the date of laboratory sample taken.

#### 8.3. Summary of Statistical Methods for Estimation of Primary and Secondary Estimands

**Table 5 Overview of Estimation Methods and Sensitivity Analyses for Primary and Secondary Estimands**

| Estimand Label | Estimand Description | Main Estimation | | | Sensitivity/ Supplementary Analyses. |
|---|---|---|---|---|---|
| | | Analysis Set | Imputation/ Data/ Censoring Rules | Analysis Model/Method | |
| Estimand 1a | Vaccine efficacy measured as VE (%) =100 × (1-RR) in SARS-CoV-2-naiive (confirmed serologically negative) adults **who receive both initial vaccination and booster after 3 weeks**. RR (SARS-CoV-2 Matrix-M1/placebo) of first occurrence of PCR- confirmed **mild, moderate or severe** COVID-19 disease with onset during a surveillance period from 7 days after second vaccination and up to 12 months after the 2nd vaccination or up to death from other causes or future use of an alternative COVID-19 vaccine or prohibited medication. | PP-EFF | Surveillance time is from 7 days after the date of $2^{nd}$ vaccination to the time of the first occurrence of illness episode onset or end of study, with appropriate censoring rules in Section 4.5.4<br><br>For IA reporting Surveillance time for subjects | A modified Poisson regression model will be fitted to the occurrence of PCR confirmed COVID-19 disease denoted as either 0 (no infection) or 1 (one or more infections) with onset between 7 days after second study vaccination and end of study at 12 months (+15 days). The model will include stratification factors and treatment group as fixed effects and robust error variances (Zou, 2004) as well as the natural log of the surveillance time as an offset. See Section 8.4 for further details. VE (%) will be presented with 95% CIs, (or adjusted CI where appropriate) and unadjusted one-sided p-value.<br><br>In the case of sparse events in one given group the Poisson regression model may fail, an Exact binomial CI will be used to summarise the Estimand. | Sensitivity 1: Additional 95% CIs for the percentage with confirmed symptomatic COVID-19 disease based on Cox Proportional Hazard model method with Efron's method of tie handling and with the treatment group as a covariate, adjusting for stratification factor will be presented on PP-EFF. 95% CI for relative risk is calculated using an Exact binomial confidence interval. Where Hazard Ratio = |

| | | | | | |
|---|---|---|---|---|---|
| | | | with events see Section 10. | See Section 8.4.1<br><br>***NOTE: Same methods and sensitivity (1) apply to Estimands 2a 3a and 4a excluding adjusted CIs | Relative Risk in the estimand.<br><br>Sensitivity 2: A tipping point analysis as described in Section 8.3.1 investigating potential events with missing data components.<br><br>Supplementary: See Estimand 1b. |
| Estimand 1b- Treatment Policy | **Supportive Estimand 1b:** Vaccine efficacy measured as VE (%) =100 × (1-RR) in COVID-19 disease -naiive adults **irrespective of compliance with second vaccination and actual serostatus at baseline**. RR=Relative risk ([SARS-CoV-2 Matrix-M1]/placebo) of first occurrence of PCR-confirmed **mild, moderate or severe** COVID-19 disease with onset during a surveillance period from **first vaccination** and up to 12 months after last vaccination or up to death from other causes or future use of an alternative COVID-19 vaccine, irrespective of use of prohibited medications | ITT | Surveillance time is from 1st vaccination to the time of onset of first occurrence of illness episode or end of study, censored at use of alternative COVID-19 prophylactics and death from other causes. | A modified Poisson regression model (as above and explained in Section 8.4) will be fitted to first occurrence of PCR confirmed mild, moderate or severe COVID-19 disease. Surveillance time is calculated **from the first study vaccination.** Note this estimand is estimated using ITT which includes subjects who do not comply with the 2nd vaccination and may have been inadvertently vaccinated despite evidence of prior or current infection at baseline.<br>VE (%) will be presented with 95% CIs<br><br>***NOTE: Same methods and sensitivity apply to Estimands 2b 3b and 4b | Sensitivity: Additional 95% CIs for the percentage with confirmed symptomatic based on Cox Proportional Hazard model with Efron's method of tie handling and with the treatment group as a covariate, adjusting for stratification factor will be presented on ITT. 95% CI for relative risk is calculated using an Exact binomial confidence interval. |

| | | | | | |
|---|---|---|---|---|---|
| | | | | Supplementary to this, cumulative incidence rate summary and plot will be reported. | Where Hazard Ratio = Relative Risk in the estimand.<br><br>Note this is supplementary to Estimand 1a. |
| Estimand 5a | Vaccine efficacy measured as VE (%) =100 × (1-RR) in SARS-CoV-2-naiive (confirmed serologically negative) adults who receive both initial vaccination and booster after 3 weeks. RR of (SARS-CoV-2 Matrix-M1/placebo) of laboratory confirmed symptomatic or asymptomatic infection with SARS-CoV-2 with onset during a surveillance period from 7 days after second vaccination and up to 12 months after the second vaccination or up to death from other causes or future use of an alternative COVID-19 vaccine or prohibited medication. | PP-EFF | Surveillance time is from 7 days after the date of 2nd vaccination to the time of sample taken with positive result in Anti-N or PCR-confirmed end of treatment or with appropriate censoring rules in Section 4.5.4 | ***NOTE: Same methods and sensitivity apply as Estimand 1a without adjusted CI's | Sensitivity 1: Additional 95% CIs for the percentage with confirmed symptomatic COVID-19 disease based on Cox Proportional Hazard model method with Efron's method of tie handling and with the treatment group as a covariate, adjusting for stratification factor will be presented on PP-EFF. 95% CI for relative risk is calculated using exact confidence interval. Where Hazard Ratio = |

| | | | | | |
|---|---|---|---|---|---|
| | | | | | Relative Risk in the estimand.<br><br>Supplementary: See Estimand 1b. |
| Estimand 5b | Vaccine efficacy measured as VE (%) =100 × (1-RR) in SARS-CoV-2-naiive adults (confirmed serologically negative ) irrespective of compliance with second vaccination. RR (SARS-CoV-2 Matrix-M1/placebo) of laboratory confirmed symptomatic or asymptomatic infection with SARS-CoV-2 with laboratory sample taken during a surveillance period from first vaccination and up to 12 months after last vaccination or up to death from other causes or future use of an alternative COVID-19 vaccine, irrespective of use of prohibited medications. | ITT (subset removing DAY 0 PCR positive or Anti-N positive test) | Surveillance time is from 1st vaccination to the time of positive (by PCR or Anti-N) laboratory sample taken or end of study, censored at use of alternative COVID-19 prophylactics and death from other causes. | ***NOTE: Same methods and sensitivity apply as Estimand 1b without adjusted CI's | Sensitivity 1: Additional 95% CIs for the percentage with confirmed symptomatic COVID-19 disease based on Cox Proportional Hazard model method with Efron's method of tie handling and with the treatment group as a covariate, adjusting for stratification factor will be presented on PP-EFF. 95% CI for relative risk is calculated using exact binomial confidence interval. Where Hazard |

| | | | | | |
|---|---|---|---|---|---|
| | | | | | Ratio = Relative Risk in the estimand.<br><br>Supplementary: See Estimand 1b. |
| Estimand 6 | Vaccine efficacy measured as VE (%) =100 × (1-RR) in COVID19 disease-naiive adults **irrespective of compliance with second vaccination and actual serostatus at baseline**. RR=Relative risk ([SARS-CoV-2 Matrix-M1]/placebo) of first occurrence of PCR- confirmed **mild, moderate or severe** COVID-19 disease with onset during a surveillance period from 14 days after **first vaccination** and up to 12 months after last vaccination or up to death from other causes or future use of an alternative COVID-19 vaccine, irrespective of use of prohibited medications | ITT | Surveillance time is from 14 days after the date of 1st vaccination to end of study, censored at use of alternative prophylactics COVID-19 and death from other causes. | A similar approach to Estimand 1b will be repeated. | |
| Estimand 7 | Percentage of vaccinated healthy adults who would develop MAAEs, etc, within timeframe.<br>A treatment policy strategy is used for assessing safety irrespective of a current | Safety<br><br>and Seasonal Influenza | Infections and death (if they meet the AE and time window criteria) are included in | Summaries of number of participants (%) with MAAEs, SAEs, and grade 3 or higher TEAEs (solicited or unsolicited) possibly or probably related to vaccine administration will be presented. The Clopper-Pearson 95% Cis will | |

| | | | | |
|---|---|---|---|---|
| | (or prior) infection at time of first vaccination or missed second vaccination. | Vaccine Sub-study | the endpoint (composite strategy). | be presented for the incidence rate of MAAEs (similarly for SAEs). |

#### 8.4. Primary Efficacy Analysis

The primary endpoint will be analysed on the PP- EFF Analysis Set in order to estimate Estimand 1a and 2a. In addition, this will be supported by estimation of a treatment policy estimands (Estimands 1b and 2b) on the ITT Set as described in Section 4.4 and overview of summary statistical methods in Section 8.1. Severity categories are defined in Appendix 12.3.

The number of seronegative subjects at baseline will be presented by treatment group and overall. This will be used as the denominator in the relevant analysis population for the statistics below.

##### 8.4.1. Modified Poisson Regression Model

Summary statistics of a log-linear model using Zou, 2004 modified Poisson Regression approach on EFF will be presented for Relative risk (RR). The Vaccine efficacy (VE) is defined as:

$$VE\ (\%) = 100 \times (1 - RR)$$

where RR = Relative Risk of incidence rates between the two treatment groups (SARS-CoV-2 rS / Placebo). Mean disease incidence rate will be reported as incidence rate per year in 1,000 people.

The one sided p-value will be presented with VE, two-sided 95% CI, calculated by modified Poisson regression with robust error variance (Zou, 2004) using SAS PROC GENMOD, where LSMEANS will be used to obtain the log relative risk (estimates) and relative risk (exponentiated). The one-sided alpha % will be calculated using the Lan-DeMets alpha-spending for Pocock boundary conditions.

The main (hypothesis testing) analysis (i.e., event-driven) for the interim and final analyses for the primary objective (Estimand 1a in the PP-EFF Analysis Set) will be carried out at an overall one-sided Type I error rate of 0.025 for the primary endpoint.

The explanatory variables in the modified Poisson regression model will include the treatment group and the stratification variables (region [pooled sites] and age group <65/≥ 65 years). The pooling of sites into regions (See Appendix H Pooling of Site) will be determined and documented prior to breaking the blind. In the case of convergence problems caused by sparse events in one group, the region term may be dropped from the model. If the model still fails then the Clopper Pearson approach will be used (see Section 8.4.2). The dependent variable will be the occurrence of the endpoint of interest. The robust error variances will be estimated using repeated statement and the subject identifier as well as the log of the surveillance time as an offset. Surveillance time is defined as per Section 4.4.2.

Subject occurrence status will be denoted as either 0 (no infection) or 1 (one or more infections). The Poisson distribution will be used with a logarithmic link function.

Hypothesis testing for the primary efficacy endpoint will be carried out against

H0: VE ≤ 30%
HI: VE > 30%

Rejection of the null hypothesis demonstrates a statistically significant vaccine effect for either primary endpoint, (i.e., alpha-adjusted lower bound confidence interval [LBCI] > 30%) will be considered meeting the pre-specified study success criterion.

All efficacy data will be presented in listings.

##### 8.4.2. Clopper Pearson Adjusted for Surveillance Time

This exact method will be considered if the Poisson regression model does not converge (e.g the total number of events in a given treatment arm are too low) but otherwise will be considered as sensitivity as the method used in the tipping point approach (see Section 8.4.4).

This method is based on the number of events in the SARS-CoV-2 rS with Matrix-M1 adjuvant group (s) as a proportion of the total number of events observed in both treatment groups ($n_e$). The first step is to calculate the Clopper Pearson CI for the proportion of events ($s/ n_e$) that occur in the active group out of all events (not subjects).

Relative risk (active/placebo) is then defined as $(s/ n_e)/(1-s/ n_e)$ and can be adjusted for total surveillance time by dividing by ratio of the total surveillance times (active/placebo). Note that the total surveillance times use information on all subjects in each group. This transformation of s/ne can be applied to the confidence limits as well as the point estimate.

##### 8.4.3. Sensitivity: Cox Proportional Hazard

The number of subjects who had an event and subjects who were censored will be summarized.

Vaccine Efficacy (Hazard Ratio(HR)) will be defined as 1- HR the primary endpoint where the hazard ratio is obtained using the Cox Proportional Hazard model.

A sensitivity analysis to explore the robustness of the model will be conducted using a stratified Cox Proportional Hazard model with Efron's method of tie handling. Treatment group will be used as covariate. A two-sided p-value and 95% CI will be presented on the PP-EFF analysis.

Counts and percentages (with 95% CIs based on the stratified Cox Proportional Hazard model) of first occurrence of PCR-Confirmed Mild, Moderate or Severe SARS-CoV-2 with an onset from at least 7 days after the second study vaccination regardless of symptoms will be presented by severity, treatment group and overall based on PP-EFF, similarly the first occurrence of symptomatic SARS-CoV-2 that is triggered by the symptomatic algorithm, as well as for moderate and severe categories for the second primary endpoint.

##### 8.4.4. Sensitivity: Tipping Point

A tipping point sensitivity analysis will be implemented for primary endpoint on the PP-EFF analysis set (Estimand 1a) to explore the influence of missing data on the overall conclusion of the efficacy analysis results and specifically to find the "tipping" point . Conclusions will change from being favourable towards SARS-CoV-2 rS with Matrix-M1 adjuvant to being unfavourable i.e. Lowest Bound Confidence Interval (LBCI) falls below VE ≤ 30%. After such a tipping point is determined, clinical judgment can be applied as to the plausibility of the assumptions underlying this tipping point.

The tipping point analysis will follow the following strategy:

- Assuming all potential events in placebo group are classed as non-event
    - All potential events in active group will be initially assumed as non-events (as per primary analysis). Further analyses will be performed incrementing the number of events until all potential events are classes as actual events. This will proceed until the tipping point is reached (LCBI ≤ 30%).

Results will be presented prior to and after the tipping point if there is a tipping point or else at the worst case and best case scenario for the active group.

Results will include VE adjusted for surveillance time with 95% Clopper Pearson CI (see Section 8.4.4) and p-value, with the number of subjects with an actual event, potential event and the number of potential events taken as events.

A subject with a potential event is defined as a subject who did not meet the primary endpoint definition with either

(1) at least one illness episode meeting mild, moderate, or severe definition with no PCR swab taken within the illness episode or

(2) at least one PCR positive swab with no corresponding FluPRO or hospitalization data within 7 days of the PCR positive swab.

Note that all Per Protocol analyses only include events with onset at least 7 days after second dose; in case of missing FluPRO data and positive PCR swabs, these will be counted as potential events if the PCR swab was taken at least 7 days after second dose.

###### 8.4.5. Sensitivity: Cumulative Incidence Events

Cumulative event rates will be summarised for the ITT analysis set at 7, 14, 21, 28 days and 3,6,9,12 months after first vaccination for both treatment groups. A corresponding cumulative incidence curve will also be reported.

##### 8.5. Secondary Endpoints Efficacy Analysis

Secondary efficacy endpoints will be analysed based on the ITT analysis set using the same method as the primary efficacy endpoint as described in Section 8.1 and Section 8.4 without adjustment for multiple comparisons (i.e. two-sided alpha 0.05).

##### 8.6. Additional Efficacy Analyses

Qualitative PCR tests will be summarised, by the number of swabs and tested positive at these timepoints: prior vaccination dose 1, prior vaccination dose 2 (Day 1- Day 21) and post-vaccination dose 2 (Day 22 +), who tested positive on the ITT Analysis Set, by overall and by site.

First occurrence of PCR-Confirmed COVID-19 Disease with onset from first vaccination, after dose 1, after dose 1 to before dose 2, dose 2 to 7 days after dose 2 and ≥7 days after dose 2 will have surveillance time summarised for both treatment groups on ITT Analysis Set.

##### 8.7. Immunogenicity Analysis

Blood will be collected from all subjects for humoral immunogenicity at Day 0, Day 35, Month 3, 6, and 12. The immunogenicity data will be analysed primarily using the PP-IMM analysis subsets, with supportive analyses performed using the ITT analysis subsets as specified in each section.

All the summaries will be conducted regardless of and stratified by age group (< 65 years or ≥ 65 years).

###### 8.7.1. Serum IgG Antibody Levels Specific for SARS-CoV-2 rS Protein Antigen

The evaluations of the serum IgG antibodies specific for the SARS-CoV-2 protein antigen(s), i.e. anti-N protein and anti-S protein as detected by ELISA, across study visits, will be performed based respectively on PP-IMM and ITT analysis sets, and PP-IMM and ITT anti-S protein serology subsets. For analysis performed on ITT analysis set and ITT anti-S protein serology subset, summaries will also be stratified by baseline serostatus (negative or positive).

All anti-N protein data will be listed for the ITT analysis set where all anti-S protein data will be listed for the ITT anti-S protein serology subset. Subjects included in the PP-IMM analysis set and PP-IMM anti-S protein serology subset will be flagged in the listings.

- GMTs (reported in Geometric mean ELISA Units (GMEUs)) by treatment and overall with 95% CI. The 95% CI will be calculated based on the t-distribution of the log-transformed values for GMTs, then back transformed to the original scale (at baseline [Day 0] and at each post-vaccination visit). Plots of the reverse cumulative distribution curves also will be provided by treatment and overall.
- Geometric Mean Fold Rise (GMFR) (at each post-vaccination visit compared to baseline [Day 0]). The 95% CI will be calculated based on the t-distribution of the log-transformed fold-rise values for GMFRs, then back transformed to the original scale.
- Seroconversion Percentage (defined as percentage of subjects at each post vaccination visit with a titre ≥ 4-fold rise).naiive The corresponding two-sided exact binomial 95% CIs will be calculated by treatment group using the Clopper-Pearson method.
- For the calculation of strain-specific GMTs in each treatment group, titres reported below the lower limit of quantification (LLOQ; i.e., below the starting dilution of assay reported as "< 10") will be set to half that limit (i.e., 10 / 2 = 5).
- The GMT will be calculated using the following formula:

$$10^{\left\{\frac{\sum_{i=1}^{n} log_{10}(t_i)}{n}\right\}}$$

where $t_1$, $t_2$, … $t_n$ are observed immunogenicity titres for n subjects.

The GMFR measures the changes in immunogenicity titres within subjects. The GMFR will be calculated using the following formula:

$$10^{\left\{\frac{\sum_{i=1}^{n} \log_{10} {}^{v_{ij}}/_{v_{ik}}}{n}\right\}} = 10^{\frac{\{\sum_{i=1}^{n}(\log_{10} v_{ij} - \log_{10} v_{ik})\}}{n}}$$

where, for n subjects, $v_{jj}$ and $v_{ik}$ are observed immunogenicity titres for subject i at time points j and k=0 (Baseline, Day 0).

Antibody titres will be summarised at baseline and each post-vaccination visit (the number of subjects with non-missing data, median, min, max, GMT and 95% CI). GMFR and the corresponding 95% CI for the GMFR will be presented by treatment group and by post-baseline visit.

Box plots of titre and fold rise by treatment group and visit will be provided. Figures based on ITT analysis set and ITT anti-S protein serology subset will be presented regardless of and stratified by baseline serostatus.

##### 8.7.2. Virus Neutralisation Assay Specific for SARS-CoV-2 Wildtype (or Variant)

An analysis similar to the serum IgG antibody levels described in Section 8.7.1 will be performed based on a neutralisation assay subset. All neutralisation assay data will also be listed for the ITT neutralisation assay subset (with a flag to identify subjects included in the PP-IMM).

##### 8.7.3. Cell-Mediated Response ± Intracellular Cytokine

Cell-mediated response (immunity) (counts per 1 Million Cells) for both Type 1 T Helper (Th1) and Type 2 T Helper (Th2) pathways will be assessed for each peptide pool by cytokine profiling and summarised by treatment group as mean, standard deviation (SD), median, min, max, and geometric mean. Change from baseline at Day 35 will also be summarised by mean, SD, median, min, and max GMFR at Day 35 will also be included. Summaries will be based on the PP-IMM and ITT cell-mediated assay subsets and will include the following cytokines: IFNg+, TNFa+, IFNg+TNFa+ (double positive), and IL-5+.

Box plots of cell-mediated response for cytokines IFNg+, TNFa+, IFNg+TNFa+ (double positive), and IL-5+ as by treatment group and visit will be provided. All cytokine data will also be presented in a data listing for the ITT cell-mediated assay subset.

#### 8.8. HAI Assay

For all subjects in the Seasonal Influenza Vaccine Sub-study, blood sampling for haemagglutination inhibition assay will be carried out on Day 0 and Day 21.

In addition to by treatment group summaries, treatment comparison will be made by comparing the strain-specific GMTs and the SCRs. The SCR for HAI assay is defined as the proportion of subjects with either a baseline reciprocal (Day 0) titre of < 10 and a post-vaccination reciprocal titre ≥ 40, or a baseline titre of ≥ 10 and a post-vaccination titre ≥ 4-fold higher.

For strain-specific GMTs in each treatment group, titres reported below the lower limit of quantitation (LLOQ; i.e., below the starting dilution of assay reported as "< 10") will be set to half that limit (i.e., 10 / 2 = 5). Strain-specific GMTs will be summarised by treatment group and visit day along with the corresponding two-sided 95% CIs, by exponentiating the corresponding log-transformed means and their 95% CIs. The ratio of GMTs between treatment groups post-vaccination and the corresponding two-sided 95% CI will be calculated on log-transformed titres using the analysis of covariance (ANCOVA) with treatment group and baseline (Day 0) measurement as the covariate.

Plots of the reverse cumulative distribution curves also will be provided by treatment and overall

For strain-specific seroconversion, the rate in percentage and the corresponding two-sided exact binomial 95% CIs will be calculated using the Clopper-Pearson method. The two-sided 95% CIs for the absolute rate difference between the two treatment groups will be constructed using the Newcombe method.

HAI assays will be summarised under the PP-IMM and ITT-IMM (influenza subset only) analysis populations.

All HAI assay data will be listed under the seasonal influenza vaccine sub-study.

##### 8.9. COVID Symptom Diary

COVID-19 symptom diary data will be captured in the FLU-PRO Plus questionnaire and FLU-PRO Plus Global Additional Daily Diary. This data will be listed only.

##### 8.10. Subgroup Analyses

The primary endpoint will be analysed by the following subgroups: age group (<65, ≥65 yrs), gender, race and ethnicity on the PP-EFF Analysis Set. Additionally age group analysis will be performed on the primary endpoint for ITT Analysis Set and key secondary endpoint for PP-EFF and ITT Analysis Sets.

#### 9. Safety Analysis

All safety summaries and analyses will be conducted for the Safety Analysis Set, except for the reactogenicity analyses that will be done on the Solicited AE Safety Subset Analysis Set.

##### 9.1. Reactogenicity

Subjects in the Solicited AE Safety Subset Analysis Set will be issued with an electronic diary to collect solicited reactogenicity, to be recorded from the time of study vaccination until 7 days after study vaccination, for each dose. Solicited local and general systemic reactogenicity will be assessed for occurrence and intensity (toxicity grade) of selected signs and symptoms from the subject during a specific post-vaccination follow-up period (day of vaccination [Day 0] and 6 subsequent days), using a pre-defined checklist in their diary. Participants in the licensed Seasonal Influenza Vaccine Sub-study will record local reactogenicity for the study vaccine injection site only. Toxicity grading will be standardised according to the FDA toxicity grading scale in Table 1 below. Any reactogenicity event extending beyond 7 days after vaccination (toxicity grade ≥1) will be recorded as an AE with a start date as date of vaccination + 7 days and followed to resolution.

The following local AEs (injection site: pain, tenderness, erythema/redness, and induration/swelling) and systemic AEs (fever, nausea/vomiting, headache, fatigue, malaise, myalgia, arthralgia) as displayed in Table 6 will be used.

**Table 6 FDA Toxicity Grading Scale for Clinical Abnormalities (Local and General Systemic Reactogenicity)**

| **Local Reaction to Injectable Product** | | | | |
|---|---|---|---|---|
| **Clinical Abnormality** | **Mild (Grade 1)** | **Moderate (Grade 2)** | **Severe (Grade 3)** | **Potentially Life Threatening (Grade 4)** |
| Pain | Does not interfere with activity | Repeated use of non-narcotic pain reliever >24 hours or interferes with activity. | Any use of narcotic pain reliever or prevents daily activity | ER visit or hospitalisation |
| Tenderness | Mild discomfort to touch | Discomfort with movement | Significant discomfort at rest | ER visit or hospitalisation |
| Erythema/redness | 2.5 − 5 cm | 5.1 − 10 cm | > 10 cm | Necrosis or exfoliative dermatitis |
| Induration/swelling | 2.5 − 5 cm and does not interfere with activity | 5.1 − 10 cm or interferes with activity | > 10 cm or prevents daily activity | Necrosis |
| **Systemic (General)** | | | | |
| **Clinical Abnormality** | **Mild (Grade 1)** | **Moderate (Grade 2)** | **Severe (Grade 3)** | **Potentially Life Threatening (Grade 4)** |
| Fever (°C)<br>(°F) | 38.0 − 38.4<br>100.4 − 101.1 | 38.5 − 38.9<br>101.2 − 102.0 | 39.0 − 40<br>102.1 − 104 | > 40<br>> 104 |
| Nausea/vomiting | No interference with activity or 1 − 2 episodes/24 hours | Some interference with activity or >2 episodes/24 hours | Prevents daily activity, or requires outpatient IV hydration | ER visit or hospitalisation for hypotensive shock |
| Headache | No interference with activity | Repeated use of nonnarcotic pain reliever >24 hours or some interference with activity | Significant; any use of narcotic pain reliever or prevents daily activity | ER visit or hospitalisation |
| Fatigue/Malaise | No interference with activity | Some interference with activity | Significant; prevents daily activity | ER visit or hospitalisation |
| Myalgia | No interference with activity | Some interference with activity | Significant; prevents daily activity | ER visit or hospitalisation |
| Arthralgia | No interference with activity | Some interference with activity | Significant; prevents daily activity | ER visit or hospitalisation |

The counts and percentage (with 95% CIs based on the exact Clopper-Pearson method) of subjects who report each solicited AE immediately post study vaccination, on the day of vaccination [Day 0] and reported up to and including each of the 6 subsequent days and overall (i.e. Day 0 – 6, and Day 21 – 27 for the respective vaccine) will be summarised by maximum toxicity grade and treatment group for each vaccination dose. The 95% CI will be calculated based on subjects who report each solicited AE immediately post study vaccination and recorded a grade greater than 0. Percentages will be based upon the number of subjects in the Solicited AE Safety Subset Analysis Set within each vaccination group who reported data for the respective category, relative to the given vaccination dose.

The summary will be presented for the Seasonal Influenza Vaccine Sub-study, separately for the subjects not included in the sub-study and overall.

The duration (in days) of solicited AEs after each vaccination will be summarised by treatment group for each vaccination for the Solicited AE Safety Subset Analysis Set, separately for those subjects in the Seasonal Influenza Vaccine Sub-study, those not included, and overall. Duration will be calculated as the number of days the solicited AE was greater than Grade 0 during the Day 0-6 assessment period.

All solicited local and systemic AEs will be presented in a data listing for the Solicited AE Safety Subset Analysis Set, separately for the subjects in the Seasonal Influenza Vaccine Sub-study and those not included. Additionally, a separate listing of solicited AEs that continued beyond 7 days after vaccination will be presented. A listing will be presented for treatment-related TEAEs.

#### 9.2. Unsolicited Adverse Events

An adverse events (AE) is defined as any untoward medical occurrence in a subject enrolled into this study regardless of its causal relationship to study vaccination.

A treatment-emergent AE (TEAE) is defined as any event not present before exposure to study vaccination or any event already present that worsens in intensity or frequency after exposure. Adverse events will also be evaluated by the investigator for the coexistence of MAAE which is defined as an AE that leads to an unscheduled visit to a healthcare practitioner.

Subjects will be assessed for diagnosis of an AESI at all study visits. AESI includes potential immune-mediated medical conditions (PIMMC) and AEs relevant to COVID-19.

AEs will be classified by system organ class (SOC) and preferred term (PT) using MedDRA (Version 23.1 or later). Only TEAEs will be included in summary tables and will be summarised by treatment group and vaccination (first dose, second dose, and overall) for the Safety Analysis Set. Summary tables for unsolicited AEs within the 21 days after first study vaccination and within the 28 days after second study vaccination will also be presented. Percentages will be based upon the number of subjects in the Safety Analysis Set, unless otherwise stated. A subject may have more than 1 AE for an SOC or PT. A subject with 2 or more AEs within the same level of summarisation will be counted only once in that level.

All subjects will be assessed for unsolicited AEs from the time of first study vaccination until Day 49; SAEs will be assessed from the time the informed consent is signed until completion of the participant’s last study-related procedure; all MAAEs will be reported from the time of first study vaccination until Day 35; MAAEs related to study vaccination and AESIs will be reported from the time of first study vaccination until completion of the participant’s last study-related procedure. All AEs will be followed until resolution or until clinically stable.

If a solicited adverse event extended beyond 6 days following vaccination, then that event will be captured as an adverse event. The onset of that event will be noted to be Day 7 (i.e. the 7th day following study vaccination).

For the purpose of inclusion in TEAE tables, incomplete AE onset and end dates will be imputed according to the below rules (where UK, UKN, and UNKN indicate unknown or missing day, month, and year, respectively):

Missing Onset Dates

- UK-MMM-YYYY: Assume 01-MMM-YYYY, but if month and year are the same as the first study vaccination month and year, then assume the date of first vaccination
- UK-UKN-YYYY: Assume 01-JAN-YYYY, but if year is the same as the first study vaccination year, then assume the date of first study vaccination
- UK-UKN-UNKN: Assume date of first study vaccination

Missing End Dates

- UK-MMM-YYYY: Assume the last day of the month
- UK-UKN-YYYY: Assume 31-DEC-YYYY
- UK-UKN-UNKN: Do not impute and assume ongoing

All unsolicited AEs will be presented in a data listing.

##### 9.2.1. Incidence for Adverse Events

An overview of TEAEs will be presented by treatment group and overall, including number of TEAEs, counts and percentage of subjects with any:

- TEAEs
- Severe TEAEs
- Treatment-related TEAEs
- Severe treatment-related TEAEs
- MAAEs
- Serious TEAEs
- TEAEs leading to vaccination discontinuation
- TEAEs leading to study discontinuation
- AESIs: PIMMC
- AESIs: relevant to COVID-19
- Serious Treatment-related MAAEs
- Treatment-related MAAEs
- Treatment-related TEAEs leading to vaccination discontinuation
- Treatment-related TEAEs leading to study discontinuation
- Treatment-related AESIs: PIMMC
- Treatment-related AESIs: relevant to COVID-19

An overall summary table of unsolicited AEs will be presented along with a further table that excludes events reported after unblinding . A summary of TEAEs will be presented by SOC and PT by treatment group and overall.

##### 9.2.2. Relationship of Adverse Events to Study Vaccine

The relationship or association of the study vaccine in causing or contributing to the AE will be characterised by the investigator as "Related" or "Not Related". All TEAEs will be presented in a summary table for each treatment group and overall by SOC, PT, and relationship to study vaccine. If a subject has 2 or more TEAEs in the same SOC (or with the same PT) with a different relationship to study vaccine, then the subject will be counted under "Related". If the relationship information is missing, the AE will be considered "Related" in the summary but will be presented as missing in the data listings.

A summary table for AEs with a start date up to an including 21 days after first study vaccination; and similarly for those unsolicited AEs with a start date up to and including 28 days after second study vaccination will also be presented by relationship to study vaccine.

Additionally a summary table of treatment-related TEAEs only will be presented.

##### 9.2.3. Severity of Adverse Event

The severity (or intensity) of an AE refers to the extent to which it affects the subject's daily activities and will be classified as mild, moderate, or severe. A summary of TEAEs and another for treatment-related TEAEs will be presented by each treatment group and overall by maximum severity, SOC, and PT. At each level of subject summarisation (SOC or PT), a subject will be counted once at the maximal severity if the subject reported one or more events. If the severity information is missing, the AE will be considered severe in the summary but will be presented as missing in the data listings.

A summary table for AEs with a start date up to an including 21 days after first study vaccination; and similarly for those unsolicited AEs with a start date up to and including 28 days after second study vaccination will also be presented by severity.

##### 9.2.4. Serious Adverse Events

A summary table for all serious TEAEs and another for treatment-related serious TEAEs will be presented for each treatment group and overall by SOC and PT. All SAEs will be presented in a data listing.

##### 9.2.5. Medically-Attended Adverse Events

MAAEs are treatment-emergent medically-attended adverse events. All MAAEs, treatment-related MAAEs and all MAAEs by maximum severity will be presented in separate summary tables for each treatment group and overall by SOC and PT. A further table will be presented for all MAAEs with a start date up to an including 14 days after second vaccination. All MAAEs will be presented in a data listing.

###### 9.2.6. Adverse Events of Special Interest

An AESI is defined as follows:

- Treatment-emergent Potential Immune-Mediated Medical Conditions (PIMMC)
- Treatment-emergent Adverse Events of Special Interest Relevant to COVID-19

All treatment-related AESIs will be presented in a summary table for each treatment group and overall by SOC and PT. All treatment-emergent PIMMC will be presented in data listings. Similarly, treatment-emergent AESIs that are relevant to COVID-19 will be presented in data listings.

###### 9.2.7. Adverse Events Leading to Study Vaccine Discontinuation

All AEs leading to study vaccine discontinuation will be presented in a data listing.

###### 9.2.8. Adverse Events Leading to Discontinuation

All AEs leading to discontinuation from study will be presented in a data listing.

##### 9.3. Clinical Laboratory Evaluations

All pregnancy test results will be presented in a listing.

##### 9.4. Vital Sign Measurements

Vital sign measurements will be taken at Screening, Day 0 (pre-injection), Day 0 (15-30 minutes post-injection), Day 21 (pre-injection), Day 21 (15-30 minutes post-injection). The measurements respiratory rate, systolic/diastolic blood pressure, pulse rate, pulse oximetry and oral temperature (or via forehead/ear reader) will be recorded. Actual values and changes from baseline for vital sign data will be summarised by nominal visit, timepoint (predose and postdose) on vaccination days, and by treatment group on the Safety Analysis Set. All vital signs measurements will be presented in a data listing.

A summary of the toxicity grades (according to the FDA toxicity grading scale, See Appendix 12.5) will be summarised by nominal visit, timepoint (predose and postdose) on vaccination days for each treatment group and overall using the count and percentage of subjects in each category.

##### 9.5. Physical Examination

Physical examination results will be summarised for each scheduled visit and examination by treatment group and overall for the Safety Analysis Set. Physical examination results for all subjects will be presented in a listing.

##### 9.6. Enhanced Emergency Room and/ or Hospitalisation Record

Enhanced Emergency Room and/ or Hospitalisation Records results for all subjects in the Safety Analysis Set will be presented in a listing.

#### 10. Interim Analyses

Safety data will be generated for two Safety Monitoring Committee (SMC) Day 7 reviews and four further SMC reviews thereafter, approximately every 3 months, when the last enrolled subject is projected to have completed 3, 6, 9 and 12 months of follow-up. Blinded outputs will be reviewed first, and when deemed necessary, unblinded tables by treatment group and unblinded subject-level listings can be reviewed to facilitate the decision. RR with 90% CI for severe COVID-19 will be generated approximately for SMC reviews at 3, 6, 9, and 12 months.

Prior to the final analysis, a single interim analysis of efficacy will be conducted based on the accumulation of approximately 50% (50 events) of the total anticipated target number of the primary endpoint (100 events). See Section 10.1 for more detail on how the primary endpoint events will be established for the interim analysis.

The interim analysis will follow standard group-sequential design using the Lan-DeMets alpha-spending function for Pocock boundary conditions. Appendix F  Interim and Final Boundaries Using Pocock Spending Function**Error! Reference source not found.** summarizes the timing, number of endpoints, and statistical success boundaries at the planned interim and final analyses.

If an unplanned additional interim analysis is to be added or the timing of a planned analysis is modified, the Lan-DeMets alpha-spending function will be used to adjust the nominal alphas to maintain the pre-specified overall one-sided type I error at 0.025.

##### 10.1. Blinded Review of Events

For the interim analysis of efficacy, data will be extracted from the clinical datatbase (EDC) when there is confidence that a minimum of 50 PCR-confirmed mild, moderate or severe events have occurred.

The illness episode algorithm (See Appendix G Algorithm for Determining Illness Episode) will be applied to this data extract, to generate a list of the subjects who have a PCR-confirmed mild/moderate/severe event. This list will be known as the *confirmed event list* and will clearly state the data extraction date and will be signed off by a member of the PPD and Novavax BIOS teams, prior to breaking the blind.

PPD DM and PPD clinical will work to clean the data for all patients included on the *confirmed event list*. Once these data cleaning activities are complete, a further data extract will be taken. This later extract will be used to generate the interim analysis of efficacy deliverables.

#### 10.2. Study Unblinding Steps

The interim analysis will be performed by an unblinded Biostatistics and Programming team (PPD), and the unblinded statistician will communicate the results of the analyses to the Sponsor in terms of fulfillment or nonfulfillment of the predefined success criterion (yes/no).

The pre-defined success criterion requires that the lower limit of the alpha-adjusted confidence interval for vaccine efficacy of the primary efficacy endpoint (Estimand 1a) >30%.

If the pre-defined success criterion of the interim analysis is unfulfilled (no), then the Sponsor will remain blinded to treatment assignment until the final analysis. If the pre-defined success criterion of the interim analysis is fulfilled (yes), then the Sponsor will sign an Unblinding Authorization Form and subsequently receive selected accrued unblinded data at the treatment group level (see Section 10.3) and continue the study while maintaining the blind to achieve a more robust safety and efficacy data package.

The unblinded analyses will be conducted in a separated area and the unblinded team will remain isolated from the study personnel and Sponsor. They will complete a review independent of the study team and Sponsor.

Summaries produced for the interim analyses will not unblind at subject level and only information at treatment level will be presented.

#### 10.3. Interim Analysis Reporting

If the results of the interim analysis for efficacy fulfil the predefined success criterion, then the following outputs will be provided to the Sponsor per the process described in Section 10.2:

- Poisson regression analysis of the primary efficacy endpoint (Estimand 1a). Note: if insufficient events occur to enable Poisson model convergence an alternative analysis will be presented based on Clopper Pearson adjusted for surveillance time (see Section 8.4.2)
- Supporting summary table for the primary efficacy endpoint (Estimand 1a)
- Overall Summary of Unsolicited Adverse Events

- Overall Summary of Unsolicited Adverse Events (excluding events reported post unblinding).

Of note, for the primary efficacy endpoint (Estimand 1a), subjects will only be classified as an event if they were included on the *confirmed event list* (generated based on the first data extract and subjected to focused data cleaning activities) and if they still meet the event criteria per the later, interim analysis data extract. This means that subjects on the *confirmed event list* may be downgraded to a non-event (owing to data cleaning outcomes); however, all subjects who were not included on the *confirmed event list* will be considered a non-event, regardless of whether they meet the event criteria per the later, interim analysis data extract (because the subset of subjects who met the primary endpoint definition only in the later data extract will not be considered sufficiently clean).

Among the patients classified as an event for the interim analysis, the onset date of the associated illness episodes will be ordered chronologically and the date of the latest of these will be established. This will be used as the *cut-off* date for the purposes of deriving surveillance time, for the primary efficacy endpoint (Estimand 1a).

#### 11. Changes from Protocol

- Section 7.2 of study protocol asserts that the interim analysis will be based on a 'locked database'. This is an error - the database will not be locked for the interim analysis. Data used for the interim analysis will be cleaned per the detail provided in Section 10.1.

- The protocol defines the PP-EFF as follows:

  *The PP-EFF will include baseline seronegative participants who receive both doses of study vaccine (SARS-CoV-2 rS with Matrix-M1 adjuvant or placebo) and have no major protocol deviations that occur before the first COVID-19 episode affecting the primary efficacy outcome (i.e., participants will be censored at the time of the protocol deviation) as assessed by the sponsor prior to unblinding. All analyses of the PP-EFF population will also exclude any illness episodes with positive SARS-CoV-2 by any validated PCR or antibody test occurring 7 days or less after the second study vaccination (e.g., Day 28*).

  This text includes some errors: Subjects with illness episodes occurring 6 days or less (rather than 7 days or less) after second study vaccination will be excluded from the PP-EFF (rather than exclusion of episodes alone).

This SAP also further qualifies that anti-s antibody test results will NOT be used to determine PP-EFF eligibility . This decision was made because anti-s testing is performed for only a subset of patients (<10%).

- The Protocol states that unblinding will result in censoring from reactogenicity analyses and analyses of unsolicited AEs. The SAP clarifies that this will be achieved via sensitivity analyses.

- The protocol defines the following endpoint:

  *First occurrence of laboratory-confirmed (by PCR or nucleocapsid [N]-protein serology to SARS-CoV-2) symptomatic or asymptomatic COVID-19 with onset at least 7 days after second study vaccination (e.g., Day 28) in adult participants regardless of their serostatus at baseline.*

  The reference to 'adult participants regardless of their serostatus at baseline' is an error. This should refer to negative baseline serostatus.

#### 12. References

Zou G. A modified poisson regression approach to prospective studies with binary data. Am J Epidemiol. 2004 Apr 1;159(7):702-6. doi: 10.1093/aje/kwh090. PMID: 15033648.

### Appendices

#### 12.1. Appendix A Schedule of Events

| Study Period: | Screening Period [a] | Clinic Visits | | | | Months After Last Study Vaccination | | |
|---|---|---|---|---|---|---|---|---|
| Study Day: | -30 to 0 | 0 [a] | 21 | 35 | COVID-19 Surveillance Visits (Unscheduled) | 3 | 6 | 12 |
| Window (days): [b] | – | 0 | + 7 | + 7 | | ± 15 | ± 15 | ± 15 |
| Minimum days following most recent study vaccination: [b] | – | 0 | 21 | 14 | | – | – | – |
| Study Visit: | Screening | 1 | 2 | 3 | | 4 | 5 | EOS [c] |
| Informed consent | X | | | | | | | |
| Medical history [d] | X | | | | X | | | |
| Inclusion/exclusion criteria [e] | X | X [f] | X [f] | | | | | |
| Demographics [g] | X | | | | | | | |
| Prior/concomitant medications [h] | X | X [f] | X [f] | X | X | X | X | X |
| Vital sign measurements [i] | X | X | X | | X | | | |
| Urine pregnancy test (WOCBP) [j] | X | X [f] | X [f] | | | | | |
| Physical examination (targeted) [k] | X | X [f] | X [f] | X | X | | | |
| Nose/throat testing for SARS-CoV-2 (PCR) [l] | X | X [f] | X [f] | | X [g] | | | |
| Blood sampling for SARS-CoV-2 (ELISA for anti N-protein serology) | | X [f] | | X | | X | X | X |
| Blood sampling for SARS-CoV-2 (ELISA for anti S-protein serology – subset of participants) [m] | | X [f] | | X | | | | |
| Blood sampling for SARS-CoV-2 neutralisation assay (subset) [n] | | X [f] | | X | | | | |
| Blood sampling for HAI (influenza co-administration subset) [o] | | X [f] | X | | | | | |
| Cell-mediated assessments (subset of participants) [p] | | X [f] | | X | | | | |
| Randomisation | | X | | | | | | |
| Study vaccination [q] | | X | X | | | | | |
| Reactogenicity (subset of participants) [r] | | X | X | | | | | |
| Monitoring for COVID-19 [s] | | COVID-19 case ascertainment will commence from Day 0 until EOS | | | | | | |
| COVID-19 Symptom Diary [t] | | | | | X | | | |

| Study Period: | Screening Period [a] | Clinic Visits | | | | Months After Last Study Vaccination | | |
|---|---|---|---|---|---|---|---|---|
| Study Day: | −30 to 0 | 0[a] | 21 | 35 | COVID-19 Surveillance Visits (Unscheduled) | 3 | 6 | 12 |
| Window (days):[b] | – | 0 | + 7 | + 7 | | ± 15 | ± 15 | ± 15 |
| Minimum days following most recent study vaccination:[b] | – | 0 | 21 | 14 | | – | – | – |
| Study Visit: | Screening | 1 | 2 | 3 | | 4 | 5 | EOS [c] |
| All unsolicited AEs [t] | | X | X | X | | | | |
| MAAEs [u] | | X | X | X | X | X | X | X |
| SAEs [v] | X | X | X | X | X | X | X | X |
| AESI [w] | | X | X | X | X | X | X | X |
| EOS form [x] | | | | | | | | X |

Abbreviations: AE = adverse event; AESI = adverse event(s) of special interest; COVID-19 = coronavirus disease 2019; ELISA = enzyme-linked immunosorbent assay; ELISpot = enzyme-linked immune sorbent spot; EOS = end of study; HAI = hemagglutinin assay inhibition; HEENT = head, eye, ear, nose, and throat (exam); ID = identification; MAAE = medically attended adverse event; N = nucleocapsid; PCR = polymerase chain reaction; PIMMC = potential immune-mediated medical conditions; S = spike protein; SAE = serious adverse event; SARS-CoV-2 = severe acute respiratory syndrome coronavirus 2; WOCBA = women of childbearing age.

a The Screening visit and Day 0 visit may be combined if feasible at any given study site.

b Days relative to study vaccination are only estimates because the window allowance is not inclusive. Should a study pause occur, visits/windows will be adjusted to allow participants to continue without protocol deviation. Visit schedules after second study vaccination are calculated relative to the day the study vaccinations were received.

c EOS visit. Should participants decide to terminate early, an EOS telephone call will occur to collect the maximum safety data possible.

d Including prior and concomitant medical conditions, recent vaccinations (≤ 90 days), and significant surgical procedures.

e Specific exclusions to study vaccination will be assessed before any vaccination. Waivers to enrolling participants with exclusions will not be given.

f Performed prior to study vaccination.

g Screening only. Including date of birth (day, month, and year), sex, race, ethnicity, weight, and height.

h Recent and current medications at the time of screening to be reviewed to ensure eligibility criteria are fulfilled. Concomitant medications include all medications (including vaccines) taken by the participant.

i Including respiratory rate, blood pressure, pulse rate, pulse oximetry and oral temperature (or via forehead/ear reader). On study vaccination days, vital sign measurements will be collected once before study vaccination to ensure participant has no evidence of fever prior to study vaccination. Vital sign measurements will also be collected once again, approximately 15 to 30 minutes after study vaccination, to check for any reactions to the vaccine.

j Women of childbearing potential only. A urine pregnancy test will be performed at Screening and prior to each study vaccination. A positive urine pregnancy test at any time will result in the participant not receiving any further study vaccination. A positive urine pregnancy test at Screening will result in screen failure.

k Examination at screening to include height and weight; HEENT, neck, thyroid, lungs, heart, cardiovascular, abdomen, lymph nodes of the upper extremities and neck; and musculoskeletal system/extremities to allow for study vaccination; symptom-directed (targeted) physical examination at all other scheduled time points but always to include lymphatic assessment of injected upper extremity on study vaccination days. Physical examination on study vaccination visits must be done prior to vaccination. Interim physical examinations will be performed at any unscheduled visit at the discretion of the investigator, if necessary.

l Samples will be collected at Screening only if the participant has any COVID-19 symptoms or significant exposure history. If a participant has a positive PCR prior to enrolment, they will be considered a screen failure. Samples will be collected on Day 0 and the method of collection will be taught. Participants found to be SARS-CoV-2 positive on Day 0 will be excluded from some analyses of the study as per the SAP. Samples may be collected on Day 21 only if the participant has any

COVID-19 symptoms or significant exposure history between Days 0 and 21. Results of that test are not required for study vaccination, but participants who are symptomatic may have the second study vaccination delayed per protocol. Participants found to be SARS-CoV-2 positive on Day 21 will be excluded from some analyses of the study as per the SAP.

m The ELISA for anti-S protein serology will be performed in the approximately 900 participants in the Anti-S Protein Serology Subset.

n The neutralising antibody assay will be performed in the approximately 900 participants in the Neutralisation Assay Subset.

o The HAI assay will be performed in the approximately 400 participants in the licensed seasonal influenza co-administration sub-study.

p Cell-mediated immune responses, as assessed by ELISpot ± intracellular cytokine staining, will be performed in approximately 450 participants in the Cell-mediated Assay Subset.

q Study vaccination on Day 0 will consist of study vaccine plus, in the seasonal influenza vaccine co-administration sub-study, a single dose of licensed influenza vaccine. Study vaccination on Day 21 will consist of study vaccine.

r Solicited AEs (e.g., local and general systemic reactogenicity symptoms) will be captured in the Safety Subset of approximately 2,000 participants and in the first approximately 400 participants who qualify for the seasonal influenza vaccine co-administration sub-study. On study vaccination days, participants will remain in clinic for at least 30 minutes to be monitored for any severe reactogenicity. Severe reactions will be noted as AEs on day of study vaccination.

s Participants will monitor for COVID-19 symptoms. This will include provision of a Study ID Card that provides details on study participation, study site contact information and assessment of symptoms of suspected COVID-19 (see Table 2-2). Instructions to participants to contact the study site within 24 hours for symptoms of suspected COVID-19 will be given. A participant with suspected or confirmed COVID-19 will be asked to complete a COVID-19 symptom diary for 10 days. The Study ID Card should be presented to healthcare providers not affiliated with the study who encounter participants with symptoms of suspected COVID-19.

t A participant with suspected or confirmed COVID-19 will be asked to complete the COVID-19 symptom diary. Participants should start the diary on their first day of symptoms and complete this diary daily for a minimum of 10 days even if their symptoms have resolved and regardless of any SARS-CoV-2 PCR results. They should continue the diary beyond 10 days if they have any ongoing symptoms related to the episode and should continue to do so until all symptoms have resolved.

u All unsolicited AEs are to be reported from the time of first study vaccination until 21 days after first study vaccination and 28 days after second study vaccination in all participants.

v MAAEs are to be collected from the time of first study vaccination until Day 35, and MAAEs assessed as related to study vaccination from the time of first study vaccination until completion of the participant's last study-related procedure.

w SAEs are to be collected from the time of informed consent until completion of the participant's last study-related procedure.

x AESI: To include PIMMC and AEs related to COVID-19 are to be collected from the time of first study vaccination until completion of the participant's last studyrelated procedure.

y EOS form will be completed for all participants, including participants who are terminated early.

z Samples will be self-collected by the participants in an effort to determine if the current symptoms are due to SARS-CoV-2 infection. Approximately 24 hours after the onset of symptoms the participants will swab themselves daily for up to 3 days but only until the point that 1 of the samples tests positive for SARS-CoV-2.

#### 12.2. Appendix B Power Under Various Vaccine Efficacy Assumptions

| **Assumed Vaccine Efficacy** | **Estimated Power** | | |
|---|---|---|---|
| **Symptomatic COVID-19 Illness PCR-Confirmed SARS-CoV-2 Infection** | **At Planned Interim Analysis with 50 Events** | **At Final Analysis with 100 Events** | **Overall (At Interim Analysis or Final Analysis)** |
| 60% | 29% | 39% | 68% |
| 65% | 45% | 41% | 87% |
| 70% | 64% | 32% | 96% |
| 75% | 81% | 18% | >99% |
| 80% | 94% | 6% | >99% |
| 85% | 99% | 1% | >99% |
| 90% | >99% | <10% | >99% |

Abbreviations: COVID-19 = coronavirus disease 2019; PCR = polymerase chain reaction; SARS-CoV-2 = severe acute respiratory syndrome coronavirus 2

#### 12.3. Appendix C Severity of COVID-19 Symptoms

| **COVID-19 Severity** | **Endpoint Definitions** |
|---|---|
| Mild | ≥ 1 of:<br>• Fever (defined by subjective or objective measure, regardless of use of anti-pyretic medications)<br>• New onset cough<br>• ≥ 2 COVID-19 respiratory/non-respiratory symptoms in Appendix 12.4<br>AND<br>• Does not meet criteria for moderate or severe disease |
| Moderate | ≥ 1 of:<br>• Fever (defined by subjective or objective measure, regardless of use of anti-pyretic medications) + any 2 COVID-19 symptoms in Appendix 12.4 for ≥ 3 days (need not be contiguous days)<br>• High fever (≥ 38.4 C) for ≥ 3 days (need not be contiguous days)<br>• Any evidence of significant LRTI:<br>− Shortness of breath (or breathlessness or difficulty breathing) with or without exertion (greater than baseline)<br>− Tachypnea: 20 to 29 breaths per minute at rest<br>− SpO2: 94% to 95% on room air<br>− Abnormal chest x-ray or chest CT consistent with pneumonia or LRTI<br>− Adventitious sounds on lung auscultation (e.g., crackles/rales, wheeze, rhonchi, pleural rub, stridor)<br>**AND**<br>• Does not meet criteria for severe disease |
| Severe | ≥ 1 of:<br>• Tachypnea: ≥ 30 breaths per minute at rest<br>• Resting heart rate ≥ 125 beats per minute<br>• SpO2: ≤ 93% on room air or PAO2/FiO2 < 300<br>• High flow oxygen therapy or NIV/NIPPV (e.g., CPAP or BiPAP)<br>• Mechanical ventilation or ECMO<br>• One or more major organ system dysfunction or failure (e.g., cardiac/circulatory, pulmonary, renal, hepatic, and/or neurological, to be defined by diagnostic testing/clinical syndrome/interventions), including any of the following:<br>o ARDS<br>o Acute renal failure<br>o Acute hepatic failure<br>o Acute right or left heart failure<br>o Septic or cardiogenic shock (with shock defined as SBP < 90 mm Hg OR DBP < 60 mm Hg<br>o Acute stroke (ischemic or hemorrhagic)<br>o Acute thrombotic event: AMI, DVT, PE<br>o Requirement for: vasopressors, systemic corticosteroids, or hemodialysis.<br>• Admission to an ICU<br>• Death |

Abbreviations: AMI = acute myocardial infarction; ARDS = acute respiratory distress syndrome; BiPAP = bi-level positive airway pressure; CPAP = continuous positive air pressure; CT = computed tomography; DBP = diastolic blood pressure; DVT = deep vein thrombosis; ECMO = extracorporeal membrane oxygenation; $FiO_2$ = fraction of inspired oxygen; ICU = intensive care unit; LRTI = lower respiratory tract infection; NIV = non-invasive ventilation; NIPPV = non-invasive positive pressure ventilation; $PAO_2$ = partial pressure of oxygen in the alveolus; PE = pulmonary embolism; SBP = systolic blood pressure; $SpO_2$ = oxygen saturation.

#### 12.4. Appendix D Qualifying Symptoms of Suspected COVID-19

| Respiratory Symptoms |
| --- |
| Fever |
| New onset cough |
| New onset or worsening of shortness of breath or difficulty breathing compared to baseline |
| New onset fatigue |
| New onset generalised muscle or body aches |
| New onset headache lasting ⩾ 48 hours |
| New loss of taste or smell |
| Acute onset of sore throat, congestion, and runny nose |
| New onset nausea, vomiting, or diarrhea lasting ⩾ 48 hours |

Abbreviations: COVID-19 = coronavirus disease 2019.

#### 12.5. Appendix E FDA Toxicity Grades Scale for Clinical Abnormalities (Vital Signs)

| Vital Signs | Mild (Grade 1) | Moderate (Grade 2) | Severe (Grade 3) | Potentially Life Threatening (Grade 4) |
|---|---|---|---|---|
| Tachycardia (bpm) | 101-115 | 116-130 | >30 | ER visit or hospitalisation for arrhythmia |
| Bradycardia (bpm) | 50-54 | 45-49 | <45 | ER visit or hospitalisation for arrhythmia |
| Hypertension (systolic) (mm Hg) | 141-150 | 151-155 | >155 | ER visit or hospitalisation for arrhythmia |
| Hypertension (diastolic) (mm Hg) | 91-95 | 96-100 | >100 | ER visit or hospitalisation for arrhythmia |
| Hypotension (systolic) (mm Hg) | 85-89 | 80-84 | <80 | ER visit or hospitalisation for shock |
| Respiratory Rate (breaths per minute) | 17-20 | 21-25 | >25 | Intubation |

Abbreviations: DHHS = Department of Health and Human Services; ER = emergency room; FDA = United States Food and Drug Administration.
Note: Participant should be at rest for all vital sign measurements, with toxicity scored on day of study vaccination (pre- and post-dose).
[a] When resting heart rate is between 60 – 100 bpm. Use clinical judgement when characterising bradycardia among some healthy participant populations (e.g., conditioned athletes).

#### 12.6. Appendix F Interim and Final Boundaries Using Pocock Spending Function

| Planned Information Fraction (% of total endpoints) | Planned Blinded Total Number of Endpoints | Planned One-Sided Nominal Alpha | VE Boundary for LBCI > 30% |
|---|---|---|---|
| Interim analysis at 50% | 50 | 0.01550 | ~68% |
| Final analysis at 100% | 100 | 0.01387 | ~57% |

Abbreviations: LBCI = lower bound confidence interval; VE = vaccine efficacy.

#### 12.7. Appendix G Algorithm for Determining Illness Episode

Note: This algorithm only applies to subjects who have received at least one dose of study drug.

Definitions:

Illness episode – Range of dates, inclusive, from when a subject first reports the onset of COVID-19 symptoms (irrespective of whether they meet the criteria for mild, moderate or severe COVID-19 disease) to the day before when the subject reports that the symptoms are back to normal (or if date of return to normal is missing, then the last day subject reports a symptom).

Infection episode – Range of dates, inclusive defined by a series of PCR+ swabs with details given below in Step 6,

I. Steps for subjects with evidence of suspected COVID-19 symptoms (as defined in Table 2-2 of the protocol):

1. Determine start and stop dates of each subject's illness episode.
    a. Source CRFs are FLU-PRO, COVID-19 New Symptoms Intake Evaluation, COVID-19 Surveillance PHONE Contact (Initial & Follow-up), COVID-19 Surveillance Visits (Initial & Follow-up), Vital Signs (temperature ≥ 37.8°C), and Enhanced ER/Hospitalization.

b. If there is at least a 7-day period free of symptoms, split into separate illness episodes.

2. After establishing Step 1 illness episode start and stop dates, the illness episode may be extended using the following symptoms if present no more than 7 days after Step 1 illness episode stop date:
    a. Moderate or Severe Pulse Oximetry (as defined in Table 2-1 of the protocol); if more than 1 occurs on the same day, select the higher pulse oximetry.
    b. Severe Heart Rate (as defined in Table 2-1 of the protocol); if more than 1 occurs on the same day, select the lower heart rate.
    c. Moderate or Severe Respiratory Rate (as defined in Table 2-1 of the protocol); if more than 1 occurs on the same day, select the lower respiratory rate.
    d. Adventitious sounds on lung auscultation (eg, crackles/rales, wheeze, rhonchi, pleural rub, stridor)
3. If the illness episode corresponds with a negative PCR swab test result, then the illness episode will be the dates determined in Steps 1 & 2.
4. If the illness episode corresponds with at least 1 PCR+ swab test result from Day 0 onward, then the illness episode may be further extended using Steps 5-7 below.
5. Flag date(s) from Day 0 onward where the subject has a PCR+ swab test result.
6. Determine start and stop dates of each <u>infection episode</u> based on PCR+ swab test results found in Step 5.
    a. If there is only 1 PCR+ swab test result, the start and stop dates of the infection episode = date of the PCR+ swab test result.
    b. If there are 2 PCR+ swab test results and they occur <=35 days apart, then these should be combined into 1 infection episode (i.e. start date = date of PCR+ swab test #1, end date = date of PCR+ swab test #2); otherwise, leave as 2 individual infection episodes (i.e. for 2 PCR+ swab test results occurring >35 days apart, start and stop dates for each episode = date of each PCR+ swab test).
    c. If there are 3 or more PCR+ swab test results, apply Step 3b pairwise, starting with the 2 PCR+ swab test results that are closest to each other for that subject.

II. Steps for any PCR+ swab test results that do not fall within an illness episode:
1. Use the Enhanced ER/Hospitalization CRFs to see if at least 1 of the following occurs on the same day as the PCR swab or no more than 7 days afterwards:
    a. Any evidence of significant LRTI

b. Except for Death (Considered in Step IV), any Moderate or Severe COVID-19 endpoint (as defined in Table 2-1 of the protocol); if more than 1 measurement occurs on the same day, select the less severe result before comparing to the criteria (i.e. lower heart rate, lower respiratory rate, higher pulse oximetry).

2. If Step 1 is met, set the start and stop dates of the illness episode as the hospital admission and discharge dates (respectively). If the discharge date is missing and the Adverse Events CRF page confirms that the subject was hospitalized due to an event related to COVID-19, then set the discharge date as the AE end date. If the discharge date is missing and there is no evidence of hospitalization due to a COVID-19-related event, then this is considered to be a 1-day event (i.e. discharge date = admission date).

III. Steps for determining if illness episodes from Sections I and II should be combined:

1. If the number of days (based on the difference between start date of episode and stop date of previous episode) between the illness episodes determined in Sections I and II is more than 7 days, then the 2 illness episodes remain separate.
2. If the number of days between the illness episodes determined in Sections I and II is <=7 days, then 2 illness episodes should be combined; set the start date as the earliest of the Section I or II illness episode start date and set the stop date as the latest of the Section I or II illness episode end date.
3. Extend illness episode start and stop dates by factoring in infection episode start and stop dates.
   a. If an illness episode has no dates in common with an infection episode, then the illness episode remains unchanged.
   b. If an illness episode overlaps by at least 1 day with an infection episode, or if an infection episode occurs <=7 days before an illness episode, or if an illness episode occurs <=7 days before an infection episode, then the final illness episode start and stop dates are the minimum of the start dates from Sections I/II/III and the maximum of the stop dates from Sections I/II/III, respectively. The start and stop dates of the combined illness episode will only use start and stop dates of the illness episodes to be merged – infectious episode start and stop dates are used to verify if the event is PCR-confirmed.

IV. Special consideration for COVID-19 related deaths:

If an Adverse Event leading to a COVID-19-related death occurs on or after a PCR+ swab test result, this is assumed to be severe illness even in the absence of symptoms recorded and is also eligible to be counted as a severe endpoint case. but no assumptions for illness episode start and stop dates are made.

Any data issues found whilst working on the algorithm will be raised with data management for data cleaning. In order for the algorithm to run the following rule will be applied:

- If there is an impossible onset date reported at a surveillance visit (i.e. which is after the date of the visit), the onset date is set to the visit date.

#### 12.8. Appendix H Pooling of Site

| Location | Site # | Region |
|---|---|---|
| London | UK001 | England South East |
| Corby | UK005 | England Other |
| London | UK006 | England South East |
| Aberdeen | UK007 | Scotland, Wales, Northern Ireland |
| Glasgow | UK008 | Scotland, Wales, Northern Ireland |
| Stockport | UK009 | England Other |
| Blackpool | UK010 | England Other |
| Belfast | UK011 | Scotland, Wales, Northern Ireland |
| London | UK012 | England South East |
| Exeter | UK013 | England Other |
| Bournemouth | UK014 | England Other |
| Norwich | UK015 | England Other |
| Oxford | UK016 | England South East |
| Stoke-on-trent | UK017 | England Other |
| Bradford | UK018 | England Other |
| Leeds | UK019 | England Other |
| London | UK020 | England South East |
| Hartlepool | UK021 | England Other |
| Lancashire | UK022 | England Other |
| Hexham | UK023 | England Other |
| Midlands | UK024 | England Other |
| Wales | UK025 | Scotland, Wales, Northern Ireland |
| Merseyside | UK026 | England Other |
| Wrexham | UK027 | Scotland, Wales, Northern Ireland |

| | | |
|---|---|---|
| Kent | UK028 | England South East |
| Lancaster | UK029 | England Other |
| Salford | UK030 | England Other |
| Thames Valley | UK031 | England South East |
| Manchester | UK032 | England Other |
| Glasgow | UK033 | Scotland, Wales, Northern Ireland |
| Ipswich & Colchester | UK034 | England Other |
| Warnsford | UK035 | England South East |
| Cornwall | UK036 | England Other |

**Novavax, Inc.**

**2019nCoV-302**

**A PHASE 3, RANDOMISED, OBSERVER-BLINDED, PLACEBO-CONTROLLED TRIAL TO EVALUATE THE EFFICACY AND SAFETY OF A SARS-COV-2 RECOMBINANT SPIKE PROTEIN NANOPARTICLE VACCINE (SARS-COV-2 RS) WITH MATRIX-M1™ ADJUVANT IN ADULT PARTICIPANTS 18-84 YEARS OF AGE IN THE UNITED KINGDOM**

**05MAR2021**

Statistical Analysis Plan

**SAP Version 4.0**

Prepared by:

PPD
[redacted]

**Approval Signatures**

| | |
|---|---|
| Date | Prepared by: [redacted] PPD |
| Date | Reviewed by: [redacted] PPD |
| Date | Approved by: [redacted] Novavax |

#### SAP Revision History

| Version | Date | Summary of Changes |
|---|---|---|
| 1.0 | 21JAN21 | First version |
| 2.0 | 15FEB21 | ✓ Incorporation of changes introduced by protocol version 3.0 other than those relevant to the interim analysis (addressed in v1.0)<br>✓ Additional condition for excluding subjects from PP-EFF and PP-IMM who received both doses of study vaccine (SARS-CoV-2 rS with Matrix-M1 adjuvant or placebo) more than 45 days apart<br>✓ Estimand 6 extended to analyse endpoint during a surveillance period from i) 7, ii) 10 in addition to iii) 14 days after first vaccination<br>✓ Further consideration for impact of individual unblinding on efficacy and immugenicity analyses<br>✓ Appendix G -- definition of the start and stop dates of the illness episode when consideration for COVID-19 related deaths<br>✓ Subject co-morbidity status (Yes/No) as an additional demographic characteristic |
| 2.1 | 19FEB21 | ✓ Clarification of the Baseline [Day 0] notion when appropriate<br>✓ Section 4.4.5 -- 4th bullet point -- Precision that post-baseline serum samples must be assessed in the specified visit window from second study vaccine<br>✓ Extension of the definition of the duration of solicited AEs |
| 3.0 | 22FEB21 | ✓ Added provision for subjects who receive approved or deployed SARS-CoV-2 vaccine prior to, or without unblinding. Such subjects should be handled in the same way as subjects who are unblinded owing to the offer of approved or deployed SARS-CoV-2 vaccine (i.e. censored/excluded from date of receipt of the deployed SARS-CoV-2 vaccine).<br>✓ Remove sentence regarding sensitivity analysis of reactogenicity and unsolicited AEs conducted for the Safety Analysis subset of subjects not unblinded as this is not planned. |
| 4.0 | 05MAR21 | ✓ In section 12, acknowledgment of any corrections done in protocol version 4.0 that were highlighted in the previous version of the SAP.<br>✓ Update of the last bullet point in Per-Protocol Immunogenicity Analysis Set (PP-IMM) definition to *not* plan to censor data after death or end of study, both events censoring de facto the subject' data.<br>✓ Precision for concomitant medications and adverse events that start dates are imputed if the imputed end date is missing or after first vaccination date.<br>✓ Clarification of the use of Clopper (vs Poisson) in sections 8.4.2 and 8.10.<br>✓ In section 4.5.1, additional consideration for screen failures.<br>✓ Precision primarily in section 4.4.1 that events with onset on the censoring date are still considered as an event (i.e. not censored).<br>✓ Additional considerations for mis-randomisation (e.g. section 4.5.4). |

#### List of Abbreviations

| | |
|---|---|
| AE | Adverse Event |
| AESI | Adverse Event of Special Interest |
| BMI | Body Mass Index |
| CI | Confidence Interval |
| COVID-19 | Coronavirus Disease 2019 |
| eCRF | Electronic Case Report Form |
| ELISA | Enzyme-Linked Immunosorbent Assay |
| ELISpot | Enzyme-Linked Immune Absorbent Spot |
| EOS | End of Study |
| ER | Emergency Room |
| EUA | Emergency Use Authorization |
| FDA | Food and Drug Administration |
| GMEU | Geometric Mean ELISA Units |
| GMFR | Geometric Mean Fold Rise |
| HAI | Haemagglutination Inhibition Assay |
| HR | Hazard Ratio |
| IgG | Immunoglobulin G |
| IM | Intramuscular |
| IRT | Interactive Response Technology |
| ITT | Intent-to-Treat |
| LBCI | Lower Bound Confidence Interval |
| LLOQ | Lower Limit of Quantification |
| LS | Least Square |
| MedDRA | Medical Dictionary for Regulatory Activities |
| MAAE | Medically-Attended Adverse Event |
| MHRA | Medicines and Healthcare Products Regulatory Aagency |
| PCR | Polymerase Chain Reaction |
| PIMMC | Potential Immune-Mediated Medical Conditions |
| PP-EFF | Per-Protocol Efficacy |
| PP-IMM | Per-Protocol Immunogenicity |
| PT | Preferred Term |
| SAE | Serious Adverse Event |
| SAP | Statistical Analysis Plan |
| SARS-CoV-2 | Severe Acute Respiratory Syndrome Coronavirus 2 |
| SARS-CoV-2 rS | SARS-CoV-2 Recombinant Spike Protein Nanoparticle Vaccine |
| SCR | Seroconversion Rate |
| SMC | Safety Monitoring Committee |
| SOC | System Organ Class |
| TEAE | Treatment-Emergent Adverse Event |
| UK | United Kingdom |
| ULOQ | Upper Limit of Quantification |
| VE | Vaccine Efficacy |
| WHODrug | World Health Organisation Drug Dictionary |

#### 1. Introduction

This document outlines the statistical methods to be implemented in the analysis of data collected within the scope of Novavax, Inc., protocol 2019nCoV-302 (A Phase 3, Randomised, Observer-Blinded, Placebo-Controlled Trial to Evaluate the Efficacy and Safety of a SARS-CoV-2 Recombinant Spike Protein Nanoparticle Vaccine (SARS-CoV-2 rS) with Matrix-M1™ Adjuvant in Adult Subjects 18 to 84 Years of Age in the United Kingdom). This statistical analysis plan (SAP) was written under protocol version 3.0 (23 December 2020) to support the final efficacy analysis and Emergency Use Authorisation (EUA) submission. Protocol version 4.0 (25 February 2021) has since been released to outline primarily changes to the study design. The changes based on this version has not been implemented in this SAP apart from the acknowledgment of any corrections that were highlighted in the previous version of the SAP and prior to the release of Protocol version 4.0.

Novavax, Inc. is developing a recombinant vaccine adjuvanted with the saponin-based Matrix-M1™ for active immunisation for the prevention of coronavirus disease 2019 (COVID-19) caused by severe acute respiratory syndrome coronavirus 2 (SARS-CoV-2).

The purpose of this study is to evaluate the efficacy, safety, and immunogenicity of SARS-CoV-2 rS with Matrix-M1 adjuvant in adults 18 to 84 years of age (inclusive). The study will be conducted at anticipated high COVID-19 transmission areas in the United Kingdom (UK). The information provided in this study will inform progression of the study vaccine, to determine efficacy of the study vaccine to prevent COVID-19 in the general population, in subjects regardless of serostatus, in subjects who have required medical intervention, and in subjects with mild or asymptomatic infections. The study will determine the safety of the study vaccine to use in the general population and to ensure that it elicits a robust immune response.

The purpose of this statistical analysis plan is to define the planned statistical analysis of the study data consistent with the study objectives. This document does not fully cover the details of the planned analyses for the Safety Monitoring Committee (SMC). The SMC charter and a SMC Table, Listing, and Figure Shells document will outline the sequential nature of these reviews.

#### 2. Objectives, Endpoints and Estimands

##### Table 1 Primary Objectives, Endpoints and Estimands

| Objectives | Endpoints | Estimands |
|---|---|---|
| To demonstrate the efficacy of SARS-CoV-2 rS with Matrix-M1 adjuvant in the prevention of PCR confirmed(by polymerase chain reaction [PCR] to SARS-CoV-2), symptomatic COVID-19, when given as a 2-dose vaccination regimen, as compared to placebo, in serologically negative (to SARS-CoV-2) adult subjects. | **PRIMARY ENDPOINT:** First occurrence of PCR confirmed **mild, moderate, or severe** COVID-19 (Appendices C and D) with onset at least 7 days after second study vaccination (e.g., Day 28) in serologically negative (to SARS-CoV-2) adult subjects at baseline until the endpoint-driven efficacy analysis is triggered by the occurrence of a prespecified number of blinded endpoints. | **Primary Estimand 1a** (See Table 5 for estimand attributes and rationale for strategies)<br>Vaccine efficacy measured as VE (%) =100 × (1-RR) in SARS-CoV-2-naiive (confirmed serologically negative) adults **who receive both initial vaccination and booster after 3 weeks**. RR=Relative risk (SARS-CoV-2 Matrix-M1/placebo) of first occurrence of PCR-confirmed **mild, moderate or severe** COVID-19 disease with onset during a surveillance period from 7 days after second vaccination and up to 12 months after the 2nd vaccination or up to death from other causes or future use of an alternative COVID-19 vaccine or prohibited medication. |
| | | **Supportive Estimand 1b:** Vaccine efficacy measured as VE (%) =100 × (1-RR) in COVID-19 disease naiive adults **irrespective of compliance with second vaccination and actual serostatus at baseline**. RR=Relative risk ([SARS-CoV-2 Matrix-M1]/placebo) of first occurrence of PCR-confirmed **mild, moderate or severe** COVID-19 disease with onset during a surveillance period from **first vaccination** and up to 12 months after last vaccination or up to death from other causes or future use of an alternative COVID-19 vaccine, irrespective of use of prohibited medications. |

##### Table 2 Secondary and Exploratory Objective, Endpoint and Estimands

| Objectives | Endpoints | Estimands |
|---|---|---|
| To demonstrate the efficacy of SARS-CoV-2 rS with Matrix-M1 adjuvant in the prevention of PCR confirmed(by polymerase chain reaction [PCR] to SARS-CoV-2), symptomatic COVID-19, when given as a 2-dose vaccination regimen, as compared to placebo, in adult subjects regardless of their serostatus at baseline | **KEY SECONDARY ENDPOINT:** First occurrence of PCR confirmed symptomatic **moderate or severe** COVID-19 (Appendix C) with onset at least 7 days after second study vaccination (e.g., Day 28) in serologically negative (to SARS-CoV-2) adult subjects at baseline until the endpoint-driven efficacy analysis is triggered by | **Secondary Estimand 2a** (See Table 5 for estimand attributes and rationale for strategies)<br>Vaccine efficacy measured as VE (%) =100 × (1-RR) in SARS-CoV-2-naiive (confirmed serologically negative) adults **who receive both initial vaccination and booster after 3 weeks**. RR=Relative risk ([SARS-CoV-2 Matrix-M1]/placebo) of first occurrence of PCR-confirmed **moderate or severe** COVID-19 disease with onset during a surveillance period from **7 days after second vaccination** and up to 12 months after the 2nd vaccination or up to death |

| Objectives | Endpoints | Estimands |
|---|---|---|
| | the occurrence of a prespecified number of blinded endpoints. | from other causes or future use of an alternative COVID-19 vaccine or prohibited medication. |
| | | **Supportive Estimand 2b:** Vaccine efficacy measured as VE (%) =100 × (1-RR) in COVID-19 disease naiive adults **irrespective of compliance with second vaccination and actual serostatus at baseline**. RR=Relative risk ([SARS-CoV-2 Matrix-M1]/placebo) of first occurrence of PCR-confirmed **moderate or severe** COVID-19 disease with onset during a surveillance period from **first vaccination** and up to 12 months after last vaccination or up to death from other causes or future use of an alternative COVID-19 vaccine, irrespective of use of other prohibited medications. |
| To demonstrate the efficacy of SARS-CoV-2 rS with Matrix-M1 adjuvant in the prevention of PCR confirmed(by polymerase chain reaction [PCR] to SARS-CoV-2), symptomatic COVID-19, when given as a 2-dose vaccination regimen, as compared to placebo, in adult subjects regardless of their serostatus at baseline | First occurrence of PCR confirmed symptomatic **severe** COVID-19 (Appendix C) with onset at least 7 days after second study vaccination (e.g., Day 28) in serologically negative (to SARS-CoV-2) adult subjects at baseline until the endpoint-driven efficacy analysis is triggered by the occurrence of a prespecified number of blinded endpoints. | **Secondary Estimand 3a** (See Table 5 for estimand attributes and rationale for strategies)<br>Vaccine efficacy measured as VE (%) =100 × (1-RR) in SARS-CoV-2-naiive (confirmed serologically negative) adults **who receive both initial vaccination and booster after 3 weeks** . RR=Relative risk ([SARS-CoV-2 Matrix-M1]/placebo) of first occurrence of PCR-confirmed symptomatic **severe** COVID-19 disease with onset during a surveillance period from **7 days after second vaccination** and up to 12 months after the 2nd vaccination or up to death from other causes or future use of an alternative COVID-19 vaccine or prohibited medication. |
| | | **Supportive Estimand 3b:** Vaccine efficacy measured as VE (%) =100 × (1-RR) in COVID-19 disease naiive adults **irrespective of compliance with second vaccination and actual serostatus at baseline**. RR=Relative risk ([SARS-CoV-2 Matrix-M1]/placebo) of first occurrence of PCR- confirmed **severe** COVID-19 disease with onset during a surveillance period from **first vaccination** and up to 12 months after last vaccination or up to death from other causes or future use of an alternative COVID-19 vaccine, irrespective of use of other prohibited medications. |

| Objectives | Endpoints | Estimands |
| --- | --- | --- |
| To demonstrate the efficacy of SARS-CoV-2 rS with Matrix-M1 adjuvant in the prevention of PCR confirmed[by PCR to SARS-CoV-2], symptomatic COVID-19, when given as a 2-dose vaccination regimen, as compared to placebo, in adult subjects regardless of their serostatus at baseline. | First occurrence of PCR confirmed symptomatic **mild, moderate, or severe** COVID-19, with onset at least 7 days after second study vaccination (e.g., Day 28) in adult subjects regardless of their serostatus at baseline. | No estimand specified. Summaries of confirmed **mild, moderate, or severe** COVID-19 disease emerging will be split by severity.<br>In ITT, this will be further split **by serostatus at baseline and time of onset (prior to 14 days after first vaccination**, from 14 days after first vaccination to 6 days after 2nd vaccination or after 7 days after 2$^{nd}$ vaccination) and severity. |
| To assess the efficacy of SARS-CoV-2 rS with Matrix-M1 adjuvant in adult subjects requiring specific medical interventions as compared to placebo. | First occurrence of COVID-19 requiring hospitalisation, intensive care unit (ICU) admission or mechanical ventilation linked to any PCR confirmed(by PCR to SARS-CoV-2) COVID-19 with onset at least 7 days after second vaccination. | **Secondary Estimand 4a/**<br>**Supportive Estimand 4b** (Similar to 1a and 1b)<br>Vaccine efficacy measured as VE (%) =100 × (1-RR) where RR=Relative risk ([SARS-CoV-2 Matrix-M1]/placebo) of first occurrence of PCR- confirmed moderate or severe COVID-19 disease **requiring hospitalisation, intensive care unit (ICU) admission or mechanical ventilation.**<br>See Estmand 1a and 1b in Table 5 for estimand attributes and rationale for strategies. |
| To assess the efficacy of SARS-CoV-2 rS with Matrix-M1 adjuvant on mild COVID-19 symptoms as compared to placebo. | First occurrence of PCR confirmed **symptomatic mild** COVID-19 (with no progression to moderate or severe COVID-19 during the course of the COVID-19 episode) with onset at least 7 days after second study vaccination (e.g., Day 28) in adult subjects regardless of their serostatus at baseline. | No estimand specified. Summaries of confirmed symptomatic COVID-19 disease emerging will be split by serostatus at baseline and time of onset (prior to or after 7 days after 2$^{nd}$ vaccination) and severity. |
| To assess the efficacy of SARS-CoV-2 rS with Matrix-M1 adjuvant on occurrence of asymptomatic or undetected infections with SARS-CoV-2 as compared to placebo. | First occurrence of laboratory-confirmed (by PCR or nucleocapsid (N)-protein serology to SARS-CoV-2) **symptomatic or asymptomatic** COVID-19 with onset at least 7 days after second study vaccination (e.g., Day 28) in adult subjects with serostatus negative at baseline. | **Secondary Estimand 5a/**<br>**Supportive Estimand 5b** (Similar to 1a/1b)<br>Vaccine efficacy measured as VE (%) =100 × (1-RR) where RR=Relative risk (SARS-CoV-2 Matrix-M1/placebo) of laboratory confirmed **symptomatic or asymptomatic** infection with SARS-CoV-2.<br>Full descriptions as per Estmand 1a and 1b in Table 5 for this endpoint with the same rationales for strategies of handling intercurrent events. Note that target population for 5b differs from 1b and is in SARS- |

| Objectives | Endpoints | Estimands |
|---|---|---|
| | | CoV-2-naive adults (confirmed serologically negative) irrespective of compliance with second vaccination. |
| | First occurrence of PCR confirmed **symptomatic mild, moderate, or severe** COVID-19 with onset at least 7, 10 and 14 days after first study vaccination (e.g., Days 7, 10, 14) in serologically negative (to SARS-CoV-2) adult subjects at baseline until the endpoint-driven efficacy analysis is triggered by the occurrence of a prespecified number of blinded endpoints | **Supportive Estimand 6** (Similar 1b)<br><br>Vaccine efficacy measured as VE (%) =100 × (1-RR) in COVID-19 disease-naiive adults **irrespective of compliance with second vaccination and actual serostatus at baseline**. RR=Relative risk ([SARS-CoV-2 Matrix-M1]/placebo) of first occurrence of PCR-confirmed **mild, moderate, or severe** COVID-19 disease with onset during a surveillance period from i) 7, ii) 10 and iii) 14 days after **first vaccination** and up to 12 months after last vaccination or up to death from other causes or future use of an alternative COVID-19 vaccine, irrespective of use of prohibited medications. |
| In a subset of adult subjects, to evaluate the immunogenicity of SARS-CoV-2 rS with Matrix-M1 adjuvant as compared to placebo. | Analysis of antibodies binding to the SARS-CoV-2 spike (S) protein by enzyme linked immunosorbent assay (ELISA) at baseline [Day 0] and Day 35 (14 days after second study vaccination).<br><br>Any occurrence of serologic conversion (by serology to SARS-CoV-2 nucleocapsid (N) protein) between baseline and 1 year after last study vaccination in adult subjects seronegative at baseline.<br><br>Evidence of seroconversion as demonstrated by antibodies to the SARS-CoV-2 S protein by ELISA at Day 35 (14 days after second study vaccination).<br><br>Cell-mediated immune responses (for example, as measured by enzyme-linked immune absorbent spot (ELISpot) ± intracellular cytokine staining) on Days 0 (baseline) and 35 (14 | No estimand specified. |

| Objectives | Endpoints | Estimands |
|---|---|---|
| | days after second study vaccination).<br>SARS-CoV-2 neutralisation as measured by virus neutralisation assay (VNA; wildtype virus and/or pseudovirion expressing SARS-CoV-2 S protein) at baseline [Day 0] and Day 35 (14 days after second study vaccination). | |
| To evaluate safety in terms of Serious Adverse Events (SAEs) and medically-attended adverse events (MAAEs) related to study vaccination in all adult subjects during the entire study period. | The occurrence and relationship to study vaccination of SAEs and MAAEs related to study vaccination (in all adult subjects) during the entire study period. | **Estimand 7**<br>Percentage of vaccinated healthy adults who would develop MAAEs, etc, within timeframe. A treatment policy strategy is used for assessing safety irrespective of a current (or prior) infection at time of first vaccination or missed second vaccination. |
| To evaluate safety in terms of adverse events of special interest (AESI), which encompasses potential immune-mediated medical conditions (PIMMCs) and AESIs relevant to COVID-19 including possible vaccine-enhanced disease, in all adult subjects at any time after the first dose. | As above with specific interest in AESI and PIMMCs. | No estimand specified. |
| In a subset of adult subjects, to evaluate the safety and reactogenicity in terms of solicited local and systemic adverse events (AEs) for 7 days after each study vaccination. | The occurrence and severity of reactogenicity in terms of solicited local and systemic<br>AEs (in all sub-study participants) for 7 days after each study vaccination | No estimand specified. |
| To evaluate safety in terms of all MAAEs for 14 days after second vaccination and unsolicited AEs for 21 days after first study vaccination and 28 days after second study vaccination. | The occurrence, severity, and relationship to study vaccination of all MAAEs for<br>14 days after second vaccination and unsolicited AEs (in all adult participants) for | No estimand specified. |

| Objectives | Endpoints | Estimands |
|---|---|---|
| | 21 days after first study vaccination and 28 days after second study vaccination. | |
| In a subset of adult subjects, to evaluate the safety and immunogenicity of SARS-CoV-2 rS with Matrix-M1 adjuvant when co-administered with a licensed seasonal influenza vaccine. | Analysis of the safety and immunogenicity of SARS-CoV-2 rS with Matrix-M1 adjuvant when co-administered with seasonal influenza vaccine in a subset population | No estimand specified. |
| In a subset of unblinded adult participants, to explore the efficacy and safety of SARS-CoV-2 rS with Matrix-M1 adjuvant and/or an approved or deployed SARS-CoV-2 vaccine. | Analysis of the efficacy and safety of SARS-CoV-2 rS with Matrix-M1 adjuvant and/or an approved or deployed SARS-CoV-2 vaccine in a subset population | No estimand specified. The estimand is deemed assessable based on long term study data (i.e. 3, 6 and months after last study vaccination) and the level of data collected was judged insufficient at the time of release of this SAP for permitting a proper analysis of the endpoint. Further considerations will be addressed in a future SAP update. |
| To assess the efficacy of SARS-CoV-2 rS with Matrix-M1 adjuvant on occurrence of asymptomatic or undetected infections with SARS-CoV-2 as compared to placebo. | Analysis of the efficacy of SARS-CoV-2 rS with Matrix-M1 adjuvant in participants with asymptomatic COVID-19 who test positive for the disease by SARS-CoV-2 N protein serology but have no accompanying symptoms. | No estimand specified. Summaries of confirmed serology will show the breakdown of how many of these are considered asymptomatic, alongside those accompanied by mild, moderate and severe symptoms. |

### 3. Investigational Plan

#### 3.1. Overall Study Design and Plan

This is a Phase 3, multi-centre, randomised, observer-blind, placebo-controlled study evaluating the efficacy, safety, and immunogenicity of SARS-CoV-2 rS with Matrix-M1 adjuvant in adult subjects 18 to 84 years of age (inclusive) in the UK. Every effort will be made to identify regions of high SARS-CoV-2 activity, and populations within these regions who are at high risk of exposure to the virus will be enrolled such that the incidence of COVID-19 in the study will be higher than 1% and the sample size and enrolment period may be reduced.

After signing informed consent, subjects may be screened within a window of up to approximately 30 days. Nose/throat samples may be taken during the screening period to detect SARS-CoV-2 by PCR, if the subject has any COVID-19 symptoms or significant exposure history. Approximately 15,000 male and female adult subjects 18 to 84 years of age (inclusive) with and without relevant comorbidities is planned for the study. An effort will be made to enrol a target of at least 25% of subjects who are ≥ 65 years of age as well as prioritising other groups that are most affected by COVID-19, including racial and ethnic minorities. This will be under the assumption that the annual incidence of symptomatic COVID-19 will be approximately 1% to 5%.

The study will consist of the screening period (Days -30 to 0); study vaccination Days 0 and 21 (+ 7-day window); and outpatient study visits on Day 0, Day 21 (+ 7 days), and Day 35 (14 days minimum after second study vaccination [+ 7 days]); and at 3, 6, and 12 months (± 15 days) after last study vaccination.

The duration of individual participation, including screening, will be a maximum of 1 year after second study vaccination (Day 386 ± 15 days). This will be an event-driven study for the assessment of vaccine efficacy (VE) and will end when a sufficient number of events have been observed, yet all subjects will be followed for the entire study duration for safety endpoints. The End of Study (EOS) analysis will be performed when the last subject reaches 12 months after the last study vaccination.

A licensed seasonal influenza co-administration sub-study will be conducted in the first 400 subjects who meet the additional inclusion criteria for this study. Subjects may be enrolled at select study sites due to the availability of seasonal influenza vaccine. After being randomised to receive intramuscular (IM) injections of SARS-CoV-2 rS with Matrix-M1 adjuvant or placebo, sub-study subjects will receive a licensed seasonal influenza vaccine on Day 0 in the opposite deltoid. These subjects will be part of the solicited AE safety subset analysis.

Given that this study and others will enrol during the influenza virus season and that the highest at-risk population, namely the elderly and adults with comorbid conditions for influenza mirrors that of SARS-CoV-2, a co-vaccination sub-group is included. This sub-group will undergo the same safety and efficacy evaluations as the entire study population. In addition, the entire sub-study will have reactogenicity assessed. This group must not have had a current season licensed influenza vaccine and have no prior history of allergy or severe reaction to seasonal influenza vaccine. To assess the possible impact of the study vaccine on the immunogenicity of the influenza vaccine, this group will also have a haemagglutination inhibition assay (HAI) performed.

Subjects will be monitored for COVID-19 throughout the study, a COVID-19 Surveillance Visit will be triggered by every episode of a new onset of symptoms of suspected COVID-19.

##### Study Vaccines

Study vaccinations (5-µg SARS-CoV-2 rS with 50-µg Matrix-M1 adjuvant or placebo [saline]) will comprise 2 IM injections on Days 0 and 21, ideally in alternating deltoids. For blinding purposes, all subjects will be vaccinated using the same injection volume (i.e., 0.5 mL).

The seasonal influenza vaccine co-administration sub-study will comprise a single IM injection (0.5 mL) of a licensed influenza vaccine on Day 0, ideally in the opposite deltoid of the study vaccine. Flucelvax Quadrivalent will be given to those 18 to 64 years of age, and an adjuvanted trivalent influenza vaccine will be given to those ≥ 65 years of age. Whenever possible the right deltoid will be used for the influenza vaccine and the left deltoid for the study vaccine.

All study vaccinations will be administered on an outpatient basis by designated site personnel in a way to maintain the blind. The influenza vaccine will not require blinding. Any pharmacy preparation with unblinded product will require unblinded site personnel who will not otherwise be involved in the study procedures or observation of subjects.

The schedule of events is provided in Appendix A.

##### Vaccination Pause Rules

Vaccination pause rules based on reactogenicity, AEs, and SAEs related to study participation will be in place to monitor subject safety during the study.

AEs meeting any one of the following criteria will result in a hold being placed on subsequent study vaccinations pending further review by the SMC and the sponsor:

- Any toxicity grade 3 or higher (severe or potentially life-threatening) solicited (local or systemic) single AE term occurring in ≥ 10% of subjects (after a minimum of 100 subjects are enrolled) in the SARS-CoV-2 rS with Matrix-M1 adjuvant group within the first 7 days after study vaccination.
- Any severe unsolicited single AE preferred term for which the investigator assesses as related that occurs in ≥ 5% of subjects (after a minimum of 100 subjects are enrolled) in the SARS-CoV-2 rS with Matrix-M1 adjuvant group, within the first 7 days after study vaccination.

In addition, any SAE assessed as related to vaccine (final assessment by the sponsor) will be reported by the sponsor to the SMC Chair as soon as possible, and within 24 hours of the sponsor's awareness of the event. Based on this initial report of the event to the SMC Chair, the Chair may advise the sponsor to immediately pause enrolment and further dosing in either some or all subjects in the study and to convene an ad hoc meeting, or make alternative recommendations. The SMC Charter defines processes for how this review will occur and how the Chair's recommendations will be documented.

The sponsor, along with medical monitor, may request an SMC review for any safety concerns that may arise in the study, even if they are not associated with any specific pause rule.

The subject influenza vaccine co-administration study will utilise the same pause rules with one difference due to the expectation of greater reactogenicity and the small size:

- Any grade 3 or higher (severe or potentially life-threatening) solicited (local or systemic) single AE term occurring in ≥ 20% of participants in the SARS-CoV-2 rS with Matrix-M1 adjuvant plus seasonal influenza vaccine group, within the first 7 days after study vaccination.

##### 3.2. Individual Unblinding

Starting from December 2020, approved SARS-CoV-2 vaccines are deployed in the UK and regulatory agencies have issued advice concerning unblinding of treatment assignment. Those who are eligible (as per the UK government prioritisation strategy) can be invited to receive an approved or deployed SARS-CoV-2 vaccine may request to be unblinded. Participants who choose to receive an approved or deployed SARS-CoV-2 vaccine will be encouraged to remain in the study for scheduled safety assessments, regardless of the UK government approved combination of SARS-CoV-2 vaccines received.

Reasons for individual unblinding other than due to participant wished to receive approved vaccine are captured on the Unblinding eCRF page - accidental unblinding, safety or efficacy concern, emergency treatment, other.

Note that the SAP acknowledges that subjects might not have been unblinded prior to receiving an approved or deployed SARS-CoV-2 vaccine; Such subjects should be handled in the same way as subjects who are unblinded owing to the offer of approved or deployed SARS-CoV-2 vaccine (i.e. censored/excluded from date of receipt of the deployed SARS-CoV-2 vaccine).

In order to manage unblinded participants and participants who received an approved or deployed SARS-CoV-2 vaccine, unblinding and approved or deployed SARS-CoV-2 vaccine receipt will result in censoring of all efficacy and immunogenicity endpoints (i.e. censoring at the earliest of these two dates if both are available). The main presentations of data based on the Safety Analysis set will include all subject data, with supporting presentations to exclude the data post unblinding/post approved or deployed SARS-CoV-2 vaccine receipt (whichever comes first). Additional analysis of AEs collected after unblinding or after an approved or deployed SARS-CoV-2 vaccine receipt may be undertaken as exploratory safety analysis.

Table 3 provides an overview of the impacts of individual unblinding and, approved or deployed SARS-CoV-2 vaccine receipt on each analysis sets. Further details are provided in Section 4.4.1 and Section 10.

**Table 3 Impact of Unblinding and Approved or Deployed SARS-CoV-2 Vaccine Receipt on Analysis Sets**

| Analysis Sets | Impact of Unblinding and Approved or Deployed SARS-CoV-2 Vaccine receipt |
|---|---|
| Intent to Treat when Efficacy [a] | Censor events[b] after the earliest of unblinding date for any unblinded subjects and, approved or deployed SARS-CoV-2 vaccine receipt date |
| Per-Protocol Efficacy [a] | Censor events[b] after the earliest of unblinding date for any unblinded subjects and, approved or deployed SARS-CoV-2 vaccine receipt date |
| Intent to Treat subsets | None |
| Per-Protocol Immunogenicity [a] | Censor events[b] after the earliest of unblinding date for any unblinded subjects and, approved or deployed SARS-CoV-2 vaccine receipt date |
| Safety | Overall and additional sensitivity analysis excluding events post unblinding or after receipt of an approved or deployed SARS-CoV-2 vaccine |

[a] Individual unblinding is not a protocol deviation
[b] Events with onset on the censoring date are still considered as an event (i.e. not censored).

#### 4. General Statistical Considerations

All data collected will be presented in data listings. Data from subjects excluded from an analysis set will be presented in the data listings but not included in the calculation of summary statistics for that analysis set. Data used for summaries will be indicated in the listings.

For categorical variables, counts and percentages of subjects will be presented. Continuous variables will be summarised using descriptive statistics (number of subjects, mean, standard deviation, median, minimum, and maximum).

When count data are presented, the percentage will be suppressed when the count is zero in order to draw attention to the non-zero counts. A row denoted “Missing” will be included in count tabulations where specified on the shells to account for missing values. The denominator for all percentages will be the number of subjects in that study vaccine within the population of interest, unless otherwise specified. Non-zero percentages will be rounded to one decimal place, except 100% will be displayed without any decimal places.

For the summary statistics of all continuous variables unless otherwise specified, minimum and maximum will be displayed to the same level of precision as reported. Mean, least square (LS) means where applicable and, median will be displayed to one level of precision greater than the data collected. Standard deviation and standard error will be displayed to two levels of precision greater than the data collected.

All confidence intervals (CI) will be 2-sided and performed using a 5% significance level unless stated otherwise. All p-values will be presented to 3 decimal places and values less than 0.001 or greater than 0.999 will be presented as <0.001 and >0.999, respectively.

Unless specified otherwise, baseline will be defined as the last non-missing assessment prior to the first study vaccine administration. Both scheduled and unscheduled visits and assessments will be used in determining baseline.

Baseline serostatus will be defined based on baseline anti N-protein results only.
Summaries by treatment group and overall will be presented for tables, for all subgroups analyses see Section 8.10.

Study day will be calculated relative to the first vaccination date as:

- Day 0 is defined as the first vaccination dose.
- Study Day = Assessment Date – First Vaccination Date.

For immunogenicity analysis calculations, antibody values reported as below the lower limit of quantification (LLOQ) will be replaced by 0.5 × LLOQ as applicable. Values that are greater than the upper limit of quantification (ULOQ) will be replaced by the ULOQ as applicable. Missing results will not be imputed.

For HAI, seroconversion is defined as either a baseline reciprocal (Day 0) titre of < 10 and a post-vaccination reciprocal titre >= 40, or a baseline titre of >= 10 and a post-vaccination titre >= 4-fold higher. For all other immunogencity data, seroconversion is defined as post vaccination >= 4-fold higher than baseline.

##### 4.1. Sample Size

This study is designed to enrol approximately 15,000 subjects, randomised 1:1 into the 2 treatment groups. The sample size is driven by the total number of events expected to achieve statistical significance for the primary efficacy endpoint – a target of 100 mild, moderate, or severe COVID-19 cases. The target number of events of 100 was chosen to provide 95% power for 70% or higher VE.

A single interim analysis of efficacy will be conducted based on the accumulation of approximately 50% (50 events) of the total anticipated primary endpoint using Pocock boundary conditions. Power calculations were performed using 10,000 simulated trials that were created under various assumptions of VEs and analysed using methods described in the “Primary efficacy analysis” section without covariates. A 90% evaluability rate for the per-protocol (PP) efficacy (PP-EFF) population (assuming 10% unevaluable due to attrition and/or baseline-seropositive participants) was used for the power calculations. All simulations were performed in SAS V9.4 and the power under various VE assumptions is shown in Appendix B.

##### 4.2. Randomisation, Stratification and Blinding

Subjects will be randomly assigned in a blinded manner using the centralised Interactive Response Technology (IRT) according to pre-generated randomisation schedules. Subjects will be randomised in a 1:1 ratio via block randomisation to receive 2 IM injections of the SARS-CoV-2 rS with Matrix-M1 adjuvant or placebo. Randomisation will be stratified by site and by age (< 65 years, ≥ 65 years). The first approximately 400 subjects who qualify for the seasonal influenza vaccine co-administration sub-study (which may be at select study locations only) will be assigned prior to randomisation. These subjects will be part of the Solicited AE Safety Subset Analysis Set. Details regarding the IRT process will be provided separately to the sites.

This is an observer-blinded study. To maintain the blind, placebo vaccination via IM route will be included and unblinded site personnel will manage study vaccine logistics, preparation, and administration so as to maintain the blind from the remainder of the site personnel and subjects. The unblinded site personnel will not be involved in study related assessments or have subject contact for data collection following study vaccine administration.

Seasonal influenza vaccine will be administered in an open-label manner.

#### 4.3. Intercurrent Event and Estimands

##### Table 4 Intercurrent Event

| Label | Intercurrent Event Type |
|---|---|
| IcEv1a (COVID-19 death) | Death due to COVID-19 disease or complications of SARS-CoV-2 infection. |
| IcEv1b (Unrelated death) | Death due to other cause, unrelated to SARS-CoV-2 infection or COVID-19 disease. |
| IcEv3 (Missed 2nd Vaccine) | Second scheduled vaccination at Day 21 (+7 days) not received. |
| IcEv4a (Alternative vaccine) | Use of alternative SARS-CoV-2 vaccine. |
| IcEv4b (Vaccine interference) | Use of any live vaccine within 4 weeks of any vaccination or any vaccine (excluding flu) within 2 weeks prior to first vaccine and 4 weeks after 2nd vaccination. |
| IcEv4c (Prohibited medications) | Use of prohibited medications deemed to impact on efficacy. |
| IcEv4d (Other Immune modifying) | Use of any other (non-prohibited) immune modifying drugs in treatment of emerging conditions. |
| IcEv5 (Prior Infection) | Laboratory confirmed SARS-CoV-2 infection or antibodies to SARS-CoV-2 on or prior to Day 0. This is added for clarity since it is possible errors might be made (which come to light after vaccination) in vaccinating people who are not naiive to SAR-CoV-2 as per the intended target population for Estimands 1a-5a. |
| IcEv6a (Day 0-27 infection) | Develops a new positive PCR-confirmed SARS-CoV-2 infection occurring between first vaccination and Second dose of Vaccination +6 days (i.e. prior to when the vaccination series is expected to be fully protective). |
| IcEv6b (Day 28-12 month infection) | Develops a new positive PCR-confirmed SARS-CoV-2 infection occurring from 2nd dose of vaccination +7 days to end of study. |

#### Table 5 Summary of Estimands 1a/1b

| Estimand Label | Primary Estimand 1a | Supportive Estimand 1b |
|---|---|---|
| **Estimand Description** | Vaccine efficacy measured as VE (%) =100 × (1-RR) in SARS-CoV-2-naiive (confirmed serologically negative) adults who receive both initial vaccination and booster after 3 weeks. RR=Relative risk ([SARS-CoV-2 Matrix-M1]/placebo) of PCR confirmed **mild, moderate or severe** COVID-19 disease with onset during a surveillance period from 7 days after second vaccination and up to 12 months after the 2nd vaccination or up to death from other causes or future use of an alternative COVID-19 vaccine or prohibited medication. | Vaccine efficacy measured as VE (%) =100 × (1-RR) in COVID-19 disease naiive adults **irrespective of compliance with second vaccination and actual serostatus at baseline**. RR=Relative risk ([SARS-CoV-2 Matrix-M1]/placebo) of first occurrence of PCR-confirmed **mild, moderate or severe** COVID-19 disease with onset during a surveillance period from **first vaccination** and up to 12 months after last vaccination or up to death from other causes or future use of an alternative COVID-19 vaccine, irrespective of use of prohibited medications. |
| **Target Population** | Adults aged 18 to 84 years who are naiive to SARS-COV-2 prior to vaccination, receive both vaccinations and without confirmed SARS-Cov-2 infection up to 6 days after 2$^{nd}$ vaccination (expected to be Day 27) | Adults aged 18 to 84 years naiive to known COVID-19 disease prior to vaccination. |
| **Endpoint** | Occurrence of PCR confirmed (by PCR to SARS-CoV-2) mild, moderate or severe COVID-19 disease with event onset between 7 days and 12 months after second study vaccination denoted as 0 (no infection) or 1 (one or more infections) where the surveillance time is from 7 days following last vaccination up to onset of event, EOS or censoring. See Section 4.4.1. | Occurrence of PCR confirmed (by PCR to SARS-CoV-2) COVID-19 disease with event onset after first vaccination denoted as 0 (no infection) or 1 (one or more infections) where the surveillance time is from first vaccination to EOS or censoring. See Section 4.4.1. |
| **Treatment Conditions** | SARS-CoV-2 rS with Matrix-M1 vs Placebo. | SARS-CoV-2 rS with Matrix-M1 vs Placebo. |
| **Population-Level Summary** | Vaccine efficacy defined as 100 x (1-RR) where RR = Relative Risk calculated as ratio of incidence rates | As per Estimand 1a. |

| | | |
|---|---|---|
| | (SARS CoV-2 Rs with Matrix-M1 / Placebo). | |
| **Intercurrent Event Strategy** | | |
| **IcEv1a (COVID-19 death)** | Composite | Composite |
| **IcEv1b (Unrelated death)** | Hypothetical | Hypothetical |
| **IcEv3 (Missed 2nd Vaccine)** | Principal stratum | Treatment policy |
| **IcE4a (Alternative vaccine)** | Hypothetical | Hypothetical |
| **IcE4b (Vaccine interference)** | Principal stratum | Treatment policy |
| **IcE4c (Prohibited medications)** | Hypothetical | Treatment policy |
| **IcE4d (Other immune modifying)** | Treatment policy | Treatment policy |
| **IcEv5 (Prior infection)** | Principal stratum | Treatment Policy |
| **IcEv6a (Day 0-27 infection)** | Principal stratum | Composite |
| **IcEv6b (Day 28-365 infection)** | Compoiste | Composite |
| **Rationale for Strategies** | This estimand seeks to understand efficacy during a surveillance period (7 days to 12 months after 2nd vaccination) which starts after the vaccine is considered to have stimulated an immune response in adults naiive to SARS-COV-2 infection (confirmed seronegative) who comply with dosing schedule and do not start an infection prior to 7 days after 2nd vaccination.<br><br>A hypothetical strategy is used for unrelated deaths and significant deviations (such as use of alternative vaccines and prohibited medications) so that interest lies in the hypothetical situation that these do not occur. | A treatment policy strategy is used for following up efficacy irrespective of whether they missed second vaccination and also including all subjects vaccinated irrespective of whether they subsequently were found not to strictly meet the criteria of the target population (i.e. found to be seropositive at vaccination). There is interest in understand efficacy in the light of poor compliance which may happen in clinically practice and may reflect reactions to the first vaccination as well as poor compliance for unrelated reasons.<br><br>The surveillance period starts from Day 0 because whilst the vaccine will not |

| | | |
|---|---|---|
| | | protect against early infections, the risk of early infection should be balanced between groups.<br><br>The hypothetical strategy is employed for other intercurrent events (unrelated deaths, alternative vaccine) which match Estimand1a. |

#### 4.4. Censoring and Surveillance Time

##### 4.4.1. Censoring

Subjects are censored, in efficacy analyses, at the earliest of

(a) follow up contact at 12 months after the second vaccination

(b) data cut-off date (see Sections 10.3 and 11),

(c) (i) unblinding for any reason (including intended receipt of approved or deployed SARS-CoV-2 vaccine)

(c) (ii) Receipt of an approved or deployed SARS-CoV-2 vaccine
Details of approved or deployed SARS-CoV-2 vaccine are recorded into Concomitant Medications eCRF page. All concomitant medication data will be reviewed by the Sponsor prior to reporting in order to identify all occurrences of approved or deployed SARS-CoV-2 vaccine receipt. Sponsor' identification of occurrences of approved or deployed SARS-CoV-2 vaccine will be integrated into the ADAM-level for programming purpose.

(d) early withdrawal from study

(e) death

In addition, in the analyses of the PP-EFF set, there is additional censoring at the date of a major protocol deviations (agreed prior to breaking the study blind).

Events with onset on the censoring date are still considered as an event (i.e. not censored); events after the censoring date are excluded.

Additional subject data censoring may be agreed, prior to breaking the study blind, following Study Deviations Rules document and Protocol Deviations review meeting.

##### 4.4.2. Surveillance Time

The surveillance time will vary according to the surveillance period defined in the estimand which will have a surveillance start date of:

- 7 days after the 2nd dose of vaccination (e.g. Estimands 1a, 2a, 3a, 4a, 5a), or
- the first vaccination (e.g. Estimands 1b, 2b, 3b, 4b, 5b), or
- 7, 10, or 14 days after the first vaccination, as applicable (i.e. Estimand 6 i, ii, and iii).

Surveillance will have an end date of the censoring date, for those without an event.

For those with an uncensored event, surveillance end date will depend on the estimand definition as either:

- the onset of event for those estimands evaluating first occurrence of PCR confirmed symptomatic COVID-19 disease events is defined as the earliest of
    - the start of the first post-vaccination illness episode symptoms (irrespective of severity level)
    - the corresponding infectious episode start (date of first positive PCR swab associated with illness episode meeting the event criteria)
- the onset of event for those estimands evaluating asymptomatic and symptomatic infections is defined as the earliest post-vaccination date of sample which returned a positive result (PCR or anti-N).

In intent-to-treat analyses, for estimands evaluating first occurrence of an event after first study vaccination (i.e. Estimands 1b, 2b, 3b, 4b, 5b), an event can be considered as the endpoint if the start of illness episode symptoms date is on or after Day 0 and, the corresponding infectious episode start after Day 0 (date of first positive PCR swab associated with illness episode meeting the event criteria).

For Estimand 6 (i, ii and iii), if the first event is prior to the start of the surveillance period, the subject will not be evaluable for the estimand and excluded from the associated analysis.

Surveillance time is calculated as surveillance end date - surveillance start date + 1.

#### 4.5. Analysis Sets

The analysis sets that will be analysed in this study are:

- All Screened Subjects
- All Randomised Subjects

- Intent-to-Treat (ITT) Analysis Set
    - Anti-S Protein Serology Subset
    - Neutralisation Assay Subset
    - Cell-mediated Assay Subset
    - Seasonal Influenza Vaccine Sub-study
- Per-Protocol Efficacy (PP-EFF) Analysis Set
- Per-Protocol Immunogenicity (PP-IMM) Analysis Set
    - Anti-S Protein Serology Subset
    - Neutralisation Assay Subset
    - Cell-mediated Assay Subset
    - Seasonal Influenza Vaccine Sub-study
- Safety Analysis Set
    - Seasonal Influenza Vaccine Sub-study
    - Solicited AE Safety Subset Analysis Set

The safety analysis will be based on the Safety Analysis Set, with exception of the data specific to each of the above subsets/sub-study. The primary efficacy analyses will be based on the PP-EFF and supported by ITT Analysis Sets respectively. The secondary immunogenicity analyses and efficacy analyses will be based on PP-IMM subsets and PP-EFF, respectively, and supported by ITT Analysis Sets. The exploratory immunogenicity and exploratory efficacy analyses will be based on ITT.

##### 4.5.1. All Screened Subjects Analysis Set

All Screened Subject Analysis Set includes all subjects who are screened. This includes screen failures. Screen failures are subjects who are identified as such on the Screen Failure CRF page and received no study vaccine.

##### 4.5.2. All Randomised Subjects Analysis Set

The All Randomised Subjects Analysis Set will include all participants who were randomised regardless of whether they actually received any study vaccine (SARS-CoV-2 rS with Matrix-M1 adjuvant or placebo), whereas the Intent-To-Treat and Per-Protocol Efficacy Analysis Sets do require subjects received at least one dose of study vaccine.

##### 4.5.3. The All Randomised Subjects Analysis Set will be used for the subject disposition summaries.Intent to Treat Analysis Set (ITT)

The Intent-To-Treat (ITT) Analysis Set will include all subjects who are randomised and receive at least 1 dose study vaccine (SARS-CoV-2 rS with Matrix-M1 adjuvant or placebo), regardless of protocol violations or missing data. Subjects will be censored in efficacy analyses (see Section 4.4.1).

Data will be analysed according to the treatment group randomised.

Within the ITT Analysis Set there are four subsets defined:

###### 4.5.3.1. Anti-S Protein Serology Subset

All subjects in the ITT Analysis Set who were tested for Anti-S protein serology using ELISA prior to study vaccination will be included in this subset.

###### 4.5.3.2. Neutralisation Assay Subset

All subjects in the ITT Analysis Set who were tested for neutralisation prior to study vaccination will be included in this subset.

###### 4.5.3.3. Cell-mediated Assay Subset

All subjects in the ITT Analysis Set who have their cell-mediated immune response assessed by ELISpot ± intracellular cytokine staining prior to study vaccination will be included in this subset.

###### 4.5.3.4. Seasonal Influenza Vaccine Sub-study

All subjects in the ITT Analysis Set who have been randomised and vaccinated subjects receiving co-administered licensed seasonal influenza vaccine and the study vaccine will be included in the sub-study. This will be the initial approximate 400 subjects who meet additional inclusion criteria for this sub-study.

###### 4.5.4. Per-Protocol Efficacy Analysis Set (PP-EFF)

Subjects will be excluded from the Per-Protocol Efficacy (PP-EFF) Analysis Set if

- they have a laboratory confirmed prior SAR-COV-2 infection by anti-N antibody test occurring up to 6 days after second study vaccination (note that Anti-S results will not be considered as these are only measured in a subset of subjects);
- they have a laboratory confirmed current SAR-COV-2 infection by any validated or licensed PCR test with symptom onset or positive PCR swab occurring up to 6 days after second study vaccination (i.e. exclusion depends on the earlier of the symptom onset date or positive swab date rather than just date of laboratory result);
- they do not receive both doses of study vaccine (SARS-CoV-2 rS with Matrix-M1 adjuvant or placebo);
- they receive both doses of study vaccine (SARS-CoV-2 rS with Matrix-M1 adjuvant or placebo) more than 45 days apart;
- they are mis-dosed (i.e. received a different study vaccine to that which they were randomised to receive);

- they have a major study deviation (e.g. incorrect volume of vaccine administered) affecting the primary efficacy outcome;
- they have a censoring event (i.e., unblinding, receipt of an approved or deployed SARS-CoV-2 vaccine, withdrew from study, died) occurring before 7 days after second study vaccination. Note that events with onset on the censoring date are still considered as an event (i.e. not censored).

In addition, data for subjects included in PP-EFF will be censored at the date of major protocol deviations. The review and determination of major protocol deviations, which lead to exclusion or censoring, will be carried out in a blinded fashion by the study clinician prior to unblinding. See Section 4.4.1 for more detail on censoring.

Mis-randomisations capture subjects who were randomised when they should not have been (e.g. screen failures who were never dosed) and subjects who were allocated to the wrong randomisation strata. Subjects will not be excluded from the Per-Protocol Analysis Sets on the basis of mis-randomisation alone - though associated study deviations may result in Per-Protocol Analysis Sets exclusion (i.e. based upon the outcomes of the blinded review of study deviations, which is described in this section 4.5.4).

Data will be analysed according to the study vaccine group as randomised.

##### 4.5.5. Per-Protocol Immunogenicity Analysis Set (PP-IMM)

Subjects will be excluded from the Per-Protocol Immunogenicity (PP-IMM) Analysis Set for each post-randomisation visit if

- they do not receive both doses of study vaccine (SARS-CoV-2 rS with Matrix-M1 adjuvant or placebo) (patients excluded from all visits);
- they receive both doses of study vaccine (SARS-CoV-2 rS with Matrix-M1 adjuvant or placebo) more than 45 days apart (except for Seasonal Influenza Vaccine Sub-study when analysing HAI assay; subjects assessed at post-baseline Day 21 only);
- they are mis-dosed (i.e. received a different study vaccine to that which they were randomised to receive) (patients excluded from all visits);
- they do not have at least a baseline and, a post-baseline serum sample result available in the specified visit window (from second study vaccine);
- they have a major protocol deviation that are considered clinically likely to impact the immunogenicity response at (or prior to) the corresponding study visit;
- they have a laboratory confirmed prior SARS-CoV-2 infection by any validated or licenced PCR or anti-N antibody test prior to each visit;
- they have a censoring event (i.e., unblinding, receipt of an approved or deployed SARS-CoV-2 vaccine) occurring at (or prior to) the corresponding study visit. Note

that events with onset on the censoring date are still considered as an event (i.e. not censored).

Immune measurements (ELISA) will be conducted on serum (immunoglobulin G [IgG]) for SARS-CoV-2 anti-S protein serology in approximately 900 participants in the Anti-S Protein Serology Subset and on anti-N protein serology (immunoglobulin G [IgG] for SARS-CoV-2 anti-N protein) in all participants.

###### 4.5.5.1. Anti-S Protein Serology Subset

All subjects in the PP-IMM Analysis Set who were tested for anti-S protein serology using ELISA prior to study vaccination will be included in this subset. Subjects with a serum sample at Day 35 assessed prior to 14 days or more than 28 days after second study vaccine will be excluded for this subset.

###### 4.5.5.2. Neutralisation Assay Subset

All subjects in the PP-IMM Analysis Set who were tested for neutralisation prior to study vaccination will be included in this subset.

###### 4.5.5.3. Cell-mediated Assay Subset

All subjects in the PP-IMM Analysis Set who have their cell-mediated immune response assessed by ELISpot ± intracellular cytokine staining prior to study vaccination will be included in this subset.

###### 4.5.5.4. Seasonal Influenza Vaccine Sub-study

All subjects in the PP-IMM Analysis Set who have been randomised and vaccinated subjects receiving co-administered licensed seasonal influenza vaccine and the study vaccine will be included in the sub-study.

##### 4.5.6. Safety Analysis Set

The Safety Analysis Set will include all subjects who receive at least 1 dose of study vaccine (SARS-CoV-2 rS with Matrix-M1 adjuvant or placebo).

Safety data will be analysed according to the study vaccine received. Subjects who are mis-dosed (i.e. receive a different study vaccine(s) to that which they were randomised to receive) will be analysed according to the first study vaccine received (regardless of whether this is the same as the second study vaccine received).

###### 4.5.6.1. Seasonal Influenza Vaccine Sub-study

All subjects in the Safety Analysis Set who have been randomised and vaccinated subjects receiving co-administered licensed seasonal influenza vaccine and the study vaccine will be included in the sub-study.

###### 4.5.6.2. Solicited AE Safety Subset Analysis Set

The Solicited AE Safety Subset Analysis Set will be a subset of the Safety Analysis Set and will be analysed according to the study vaccine actually received. It will comprise the following: the Seasonal Influenza Vaccine Sub-study subjects and the initial approximate 2,000 subjects randomised who receive a vaccine.

### 5. Subject Disposition and Protocol Deviations

#### 5.1. Disposition

Subject disposition will be summarised for each treatment group and overall for the All Randomised Subjects Set.

The number of subjects who are randomised in the study to each treatment group and the count and percentage of subjects who complete the study (i.e. study participation until the EOS Visit without early termination) will be presented. Count and percentage of subjects who have completed, are ongoing, withdraw or discontinue from the study, and the reason for withdrawal or discontinuation, will also be summarised.

In addition, when an analysis is requested or for the planned SMC's, the count and percentage of subjects who are ongoing at the time of the data extraction will also be summarised.

The count and percentage of subjects in each analysis set (excluding All Screened Subject Analysis Set) will be presented in a summary table.

Subject disposition data including reason for individual unblinding, analysis sets, and randomisation data will be presented in data listings. Screen failure data will be summarised and will be presented in data listings.

#### 5.2. Protocol Deviations

A protocol deviation is any change, divergence, or departure from the study design or procedures defined in the protocol. A significant deviation is a subset of protocol deviations that leads to a subject being discontinued from the study or significantly affects the subject's rights, safety, or well-being and/or the completeness, accuracy, and reliability of the study data, especially as it pertains to elicited immune response to planned study vaccination. A significant deviation can include non-adherence to inclusion or exclusion criteria or non-adherence to regulatory authority including International Council for Harmonisation E6 (R2) guidelines.

Protocol deviations will be documented by the clinical monitor throughout the course of monitoring visits. The investigator will be notified in writing by the monitor of deviations. The Health Research Ethics Committee (HREC) should be notified of all protocol deviations, if appropriate, in a timely manner.

All protocol deviations will be presented in a data listing with another displaying only significant deviations. This latter listing will include a column to indicate whether each significant deviation has led to exclusion from any analysis sets or censoring (for PP-EFF analysis set only). Inclusion/Exclusion for analysis sets will be presented in a data listing.

Details of study entry criteria deviations will be presented in a separate data listing.

Significant deviations will be summarised by treatment group for each site for the ITT Analysis Set. An additional summary will be presented for major protocol deviations leading to exclusion from analysis sets, or leading to censoring of study data for analysis.

### 6. Demographics and Baseline Characteristics

#### 6.1. Demographics

Demographic and baseline characteristics will be summarised for each treatment group and overall for the ITT Analysis Set, Safety Analysis Set, PP-EFF Analysis Set, PP-IMM Anti-S Protein Serology Subset, PP-IMM Cell-Mediated Assay Subset and PP-IMM Analysis Set (Seasonal Influenza Vaccine Subset).

A further summary of demographic and baseline characteristics will be presented for subjects in PP-EFF Analysis Set with COVID-19 disease attributable to the UK variant.

The demographic characteristics consist of age (years), age-group (<65 yrs, >=65 yrs), sex, child-bearing potential, birth control method, race and, ethnicity. The baseline characteristics consist of baseline weight (kg), baseline height (cm), baseline body mass index (BMI, $kg/m^2$), PCR (+/−) at baseline and Day 21, SARS-CoV-2 serostatus based on anti-N antibody test only (positive/negative) at baseline and, co-morbidity status (Yes/No).

The sponsor will review a list of all preferred terms amongst the medical history data (prior to study unblinding) and identify those that are to be considered comorbid conditions. Comorbid subjects will then be identified as those who have at least one of these comorbid conditions reported as a medical or, have a screening BMI value greater than 30 $kg/m^2$.

Subject demographic and baseline characteristics will be presented in a data listing.

#### 6.2. Medical History

Medical history will be classified by system organ class (SOC) and preferred term (PT) using MedDRA (Version 23.1 or later).

Medical history will be summarised by SOC and PT will be summarised for each treatment group and overall for the Safety Analysis Set. Medical history will be presented in a data listing.

### 7. Treatments, Medications, and Recent Vaccinations

#### 7.1. Recent Vaccinations and Concomitant Medications

Administration of medications, therapies, or vaccines will be recorded in the electronic case report form (eCRF). Recent vaccinations will include vaccinations taken ≤90 days prior to taking study vaccination. Concomitant medications will include all medications (including vaccines) taken by the subject from the time of signing the informed consent form until EOS (or until the early termination visit if prior to that time). Prescription and over the counter drugs, as well as herbals, vitamins, and supplements, will be included.

Recent vaccinations and concomitant medications and therapies will be summarised for the Safety Analysis Set by treatment group, anatomical therapeutic chemical (ATC Levels 1-2) and preferred drug name as coded using the World Health Organisation Drug Dictionary (currently WHODrug Global-B3 Sep 2020 however this will be updated during the course of the study).

For the purpose of inclusion in recent vaccinations and concomitant medications tables, incomplete start and stop dates will be imputed according to the below rules (where UK, UKN, and UNKN indicate unknown or missing day, month, and year, respectively):

Missing Start Dates

- UK-MMM-YYYY: Assume 01-MMM-YYYY, but if month and year are the same as the first study vaccination month and year, then assume the date of first vaccination
- UK-UKN-YYYY: Assume 01-JAN-YYYY, but if year is the same as the first study vaccination year, then assume the date of first study vaccination
- UK-UKN-UNKN: Assume date of first study vaccination

Note that for concomitant medications, start dates will only be imputed, if the imputed end date is missing or after the first vaccination date.

Missing Stop Dates

- UK-MMM-YYYY: Assume the last day of the month
- UK-UKN-YYYY: Assume 31-DEC-YYYY
- UK-UKN-UNKN: Do not impute and assume ongoing

All recent vaccinations and concomitant medications will be presented in a data listing.

#### 7.2. Medical and Surgical Treatment Procedures

All medical and surgical treatment procedures (conducted during the study period) will be coded using MedDRA (Version 23.1 or later). Medical and surgical treatment procedures will be summarised by SOC, PT and treatment group for the Safety Analysis Set with all procedures presented in a data listing.

##### 7.3. Study Vaccine Administration

Study vaccinations (SARS-CoV-2 rS with Matrix-M1 adjuvant or placebo) will comprise 2 IM injections (Day 0 and Day 21), ideally in alternating deltoids. For blinding purposes, all subjects will be vaccinated using the same injection volume (i.e., 0.5 mL). The dose level will be 5 µg SARS-CoV-2 rS with 50 µg Matrix-M1 adjuvant (co-formulated in a single vial); placebo will be saline for injection. The subjects in the Seasonal Influenza Vaccine Sub-study will be vaccinated using a single IM injection (0.5 mL) of a licensed influenza vaccine on Day 0, ideally in the opposite deltoid of the study vaccine.

Study vaccinations will be summarised for the Safety Analysis Set including the count and percentage of subjects receiving each dose, and if administered per protocol (yes, no), as well as a duration of follow-up for subjects receiving the 2$^{nd}$ vaccination. Similarly, a summary table for the Seasonal Influenza Vaccine Sub-study will present the count and percentage of subjects receiving the influenza vaccine at Day 0, and whether trivalent or quadrivalent.

All study drug administration data will be presented in a data listing.

#### 8. Efficacy Analysis and Immunogenicity Analysis

##### 8.1. Symptoms of Suspected COVID-19

Subjects with symptoms of suspected COVID-19 will

- take their temperature daily for 10 days
- self swab themselves daily for 3 days
- and complete a FLU-PRO diary that will collect their symptoms See Appendix C

Every episode of a "new onset" of symptoms of suspected COVID-19 will trigger a COVID-19 Surveillance Visit (Initial and Follow-Up) or phone call. A "New onset" will require at least a 7-day period symptom-free prior to the event to differentiate a specific episode from any prior illness.

For the definitions around the primary estimand 1a and supportive endpoint, see Table 5.

##### 8.2. Asymptomatic Infections

Asymptomatic infection will be defined as a SARS-CoV-2 infection confirmed by N protein serology without meeting the criteria for PCR confirmed mild, moderate or severe COVID disease. This can only be assessed in those who have serostatus negative at baseline.

The event onset date will not be known exactly but for the purposes of calculating the surveillance time, the event onset date will be the date of laboratory sample taken.

#### 8.3. Summary of Statistical Methods for Estimation of Primary and Secondary Estimands

**Table 6 Overview of Estimation Methods and Sensitivity Analyses for Primary and Secondary Estimands**

| Estimand Label | Estimand Description | Main Estimation | | | Sensitivity/ Supplementary Analyses. |
|---|---|---|---|---|---|
| | | Analysis Set | Imputation/ Data/ Censoring Rules | Analysis Model/Method | |
| Estimand 1a | Vaccine efficacy measured as VE (%) =100 × (1-RR) in SARS-CoV-2-naiive (confirmed serologically negative) adults **who receive both initial vaccination and booster after 3 weeks**. RR (SARS-CoV-2 Matrix-M1/placebo) of first occurrence of PCR- confirmed **mild, moderate or severe** COVID-19 disease with onset during a surveillance period from 7 days after second vaccination and up to 12 months after the 2nd vaccination or up to death from other causes or future use of an alternative COVID-19 vaccine or prohibited medication. | PP-EFF | Surveillance time is from 7 days after the date of 2$^{nd}$ vaccination to the time of the first occurrence of illness episode onset or end of study, with appropriate censoring rules in Section 4.5.4<br><br>For IA reporting Surveillance time for subjects with events see Section 10. | A modified Poisson regression model will be fitted to the occurrence of PCR confirmed COVID-19 disease denoted as either 0 (no infection) or 1 (one or more infections) with onset between 7 days after second study vaccination and end of study at 12 months. The model will include stratification factors and treatment group as fixed effects and robust error variances (Zou, 2004) as well as the natural log of the surveillance time as an offset. See Section 8.4 for further details. VE (%) will be presented with 95% CIs, (or adjusted CI where appropriate) and unadjusted one-sided p-value.<br><br>In the case of sparse events in one given group the Poisson regression model may fail, an Exact binomial CI will be used to summarise the Estimand.<br><br>See Section 8.4.1 | Sensitivity 1: Additional 95% CIs for the percentage with confirmed symptomatic COVID-19 disease based on Cox Proportional Hazard model method with Efron's method of tie handling and with the treatment group as a covariate, adjusting for stratification factor will be presented on PP-EFF. 95% CI for relative risk is calculated using an Exact binomial confidence interval. Where Hazard Ratio = |

| | | | | | |
|---|---|---|---|---|---|
| | | | | ***NOTE:<br>Same methods and sensitivity (1) apply to Estimands 2a 3a and 4a excluding adjusted CIs and unadjusted one-sided p-value.<br>For Estimands 3a and 4a, the exact method will be undertaken regardless of the convergence status of the Poisson regression model. | Relative Risk in the estimand.<br><br>Sensitivity 2: A tipping point analysis as described in Section 8.3.1 investigating potential events with missing data components.<br><br>Supplementary: See Estimand 1b. |
| Estimand 1b-Treatment Policy | **Supportive Estimand 1b:** Vaccine efficacy measured as VE (%) =100 × (1-RR) in COVID-19 disease -naiive adults **irrespective of compliance with second vaccination and actual serostatus at baseline**. RR=Relative risk ([SARS-CoV-2 Matrix-M1]/placebo) of first occurrence of PCR-confirmed **mild, moderate or severe** COVID-19 disease with onset during a surveillance period from **first vaccination** and up to 12 months after last vaccination or up to death from other causes or future use of an alternative COVID-19 vaccine, irrespective of use of prohibited medications. | ITT | Surveillance time is from 1st vaccination to the time of onset of first occurrence of illness episode or end of study, censored at use of alternative COVID-19 prophylactics and death from other causes. | A modified Poisson regression model (as above and explained in Section 8.4) will be fitted to first occurrence of PCR confirmed mild, moderate or severe COVID-19 disease. Surveillance time is calculated **from the first study vaccination.**<br>Note this estimand is estimated using ITT which includes subjects who do not comply with the 2nd vaccination and may have been inadvertently vaccinated despite evidence of prior or current infection at baseline.<br>VE (%) will be presented with 95% CIs and unadjusted one-sided p-value. | Sensitivity: Additional 95% CIs for the percentage with confirmed symptomatic based on Cox Proportional Hazard model with Efron's method of tie handling and with the treatment group as a covariate, adjusting for stratification factor will be presented on ITT. 95% CI for relative risk is calculated using an Exact binomial confidence interval. Where Hazard Ratio = |

| | | | | | |
|---|---|---|---|---|---|
| | | | | ***NOTE:<br>Same methods and sensitivity apply to Estimands 2b 3b and 4b excluding unadjusted one-sided p-value.<br>For Estimands 3b and 4b, the exact method will be undertaken regardless of the convergence status of the Poisson regression model.<br>Supplementary to this, cumulative incidence rate summary and plot will be reported. | Relative Risk in the estimand.<br>Note this is supplementary to Estimand 1a. |
| Estimand 5a | Vaccine efficacy measured as VE (%) =100 × (1-RR) in SARS-CoV-2-naiive (confirmed serologically negative) adults who receive both initial vaccination and booster after 3 weeks. RR of (SARS-CoV-2 Matrix-M1/placebo) of laboratory confirmed symptomatic or asymptomatic infection with SARS-CoV-2 with onset during a surveillance period from 7 days after second vaccination and up to 12 months after the second vaccination or up to death from other causes or future use of an alternative COVID-19 vaccine or prohibited medication. | PP-EFF | Surveillance time is from 7 days after the date of 2$^{nd}$ vaccination to the time of sample taken with positive result in Anti-N or PCR-confirmed end of treatment or with appropriate censoring rules in Section 4.5.4 | ***NOTE: Same methods and sensitivity apply as Estimand 1a without adjusted CIs and unadjusted one-sided p-value. | Sensitivity 1: Additional 95% CIs for the percentage with confirmed symptomatic COVID-19 disease based on Cox Proportional Hazard model method with Efron’s method of tie handling and with the treatment group as a covariate, adjusting for stratification factor will be presented on PP-EFF. 95% CI for relative risk is calculated using exact confidence interval. Where Hazard Ratio = |

| | | | | | |
|---|---|---|---|---|---|
| | | | | | Relative Risk in the estimand.<br><br>Supplementary: See Estimand 1b. |
| Estimand 5b | Vaccine efficacy measured as VE (%) =100 × (1-RR) in SARS-CoV-2-naiive adults (confirmed serologically negative ) irrespective of compliance with second vaccination. RR (SARS-CoV-2 Matrix-M1/placebo) of laboratory confirmed symptomatic or asymptomatic infection with SARS-CoV-2 with laboratory sample taken during a surveillance period from first vaccination and up to 12 months after last vaccination or up to death from other causes or future use of an alternative COVID-19 vaccine, irrespective of use of prohibited medications. | ITT (subset removing subjects Day 0 PCR positive or anti-N positive test at baseline) | Surveillance time is from 1st vaccination to the time of positive (by PCR or Anti-N) laboratory sample taken or end of study, censored at use of alternative COVID-19 prophylactics and death from other causes. | ***NOTE: Same methods and sensitivity apply as Estimand 1b without unadjusted one-sided p-value | Sensitivity 1: Additional 95% CIs for the percentage with confirmed symptomatic COVID-19 disease based on Cox Proportional Hazard model method with Efron's method of tie handling and with the treatment group as a covariate, adjusting for stratification factor will be presented on PP-EFF. 95% CI for relative risk is calculated using exact binomial confidence interval. Where Hazard Ratio = Relative Risk in the estimand.<br><br>Supplementary: See Estimand 1b. |

| | | | | |
|---|---|---|---|---|
| Estimand 6 | Vaccine efficacy measured as VE (%) =100 × (1-RR) in COVID19 disease-naiive adults **irrespective of compliance with second vaccination and actual serostatus at baseline**. RR=Relative risk ([SARS-CoV-2 Matrix-M1]/placebo) of first occurrence of PCR- confirmed **mild, moderate or severe** COVID-19 disease with onset during a surveillance period from i) 7, ii) 10 and iii) 14 days after **first vaccination** and up to 12 months after last vaccination or up to death from other causes or future use of an alternative COVID-19 vaccine, irrespective of use of prohibited medications | ITT (subset excluding subjects with the first event prior to the start of the surveillance period) | Surveillance time is from i) 7, ii) 10 and iii) 14 days after the date of 1st vaccination to end of study, censored at use of alternative prophylactics COVID-19 and death from other causes. | A similar approach to Estimand 1b will be repeated excluding unadjusted one-sided p-value.. |
| Estimand 7 | Percentage of vaccinated healthy adults who would develop MAAEs, etc, within timeframe.<br><br>A treatment policy strategy is used for assessing safety irrespective of a current (or prior) infection at time of first vaccination or missed second vaccination. | Safety and Seasonal Influenza Vaccine Sub-study | Infections and death (if they meet the AE and time window criteria) are included in the endpoint (composite strategy). | Summaries of number of participants (%) with MAAEs, SAEs, and grade 3 or higher TEAEs (solicited or unsolicited) possibly or probably related to vaccine administration will be presented. The Clopper-Pearson 95% Cis will be presented for the incidence rate of MAAEs (similarly for SAEs). |

#### 8.4. Primary Efficacy Analysis

The primary endpoint will be analysed on the PP- EFF Analysis Set in order to estimate Estimand 1a and 2a. In addition, this will be supported by estimation of a treatment policy estimands (Estimands 1b and 2b) on the ITT Set as described in Section 4.4 and overview of summary statistical methods in Section 8.1. Severity categories are defined in Appendix C.

The number of seronegative subjects at baseline will be presented by treatment group and overall. This will be used as the denominator in the relevant analysis population for the subsequent statistics.

##### 8.4.1. Modified Poisson Regression Model

Summary statistics of a log-linear model using Zou, 2004 modified Poisson Regression approach on EFF will be presented for Relative risk (RR). The Vaccine efficacy (VE) is defined as:

$$VE\ (\%) = 100 \times (1 - RR)$$

where RR = Relative Risk of incidence rates between the two treatment groups (SARS-CoV-2 rS / Placebo). Mean disease incidence rate will be reported as incidence rate per year in 1,000 people.

The main (hypothesis testing) analysis (i.e., event-driven) for the interim and final analyses for the primary objective (Estimand 1a in the PP-EFF Analysis Set) will be carried out at an overall one-sided Type I error rate of 0.025 for the primary endpoint.

Hypothesis testing for the primary efficacy endpoint will be carried out against

H0: $VE \leq 30\%$

HI: $VE > 30\%$

Rejection of the null hypothesis demonstrates a statistically significant vaccine effect for either primary endpoint, (i.e., alpha-adjusted lower bound confidence interval [LBCI] > 30%) will be considered meeting the pre-specified study success criterion.

The one sided p-value will be presented with VE along with two-sided 95% CI, calculated by modified Poisson regression with robust error variance (Zou, 2004) using SAS PROC GENMOD, where LSMEANS will be used to obtain the log relative risk (estimates) and relative risk (exponentiated). The one-sided alpha % will be calculated using the Lan-DeMets alpha-spending for Pocock boundary conditions.

The explanatory variables in the modified Poisson regression model will include the treatment group and the stratification variables (region [pooled sites] and age group (<65/≥ 65 years). Note that strata will be corrected for mis-allocations at randomisation. The pooling

of sites into regions (See Appendix H) will be determined and documented prior to breaking the blind. In the case of convergence problems caused by sparse events in one group, the region term may be dropped from the model. If the model still fails then the Clopper Pearson approach will be used (see Section 8.4.2). The dependent variable will be the occurrence of the endpoint of interest. The robust error variances will be estimated using repeated statement and the subject identifier as well as the log of the surveillance time as an offset. Surveillance time is defined as per Section 4.4.2.
Subject occurrence status will be denoted as either 0 (no infection) or 1 (one or more infections). The Poisson distribution will be used with a logarithmic link function.

All efficacy data will be presented in listings.

##### 8.4.2. Clopper Pearson Adjusted for Surveillance Time

This exact method will be considered in case of low numbers of events (e.g. Estimands 3a, 3b, 4a and 4b) or, if the modified Poisson regression model does not converge (i.e. zero counts in one treatment group or stratum) but, otherwise will be considered as sensitivity as the method used in the tipping point approach (see Section 8.4.4).

This method is based on the number of events in the SARS-CoV-2 rS with Matrix-M1 adjuvant group (s) as a proportion of the total number of events observed in both treatment groups ($n_e$). The first step is to calculate the Clopper Pearson CI for the proportion of events ($s/ n_e$) that occur in the active group out of all events (not subjects).

Relative risk (active/placebo) is then defined as $(s/ n_e)/(1-s/ n_e)$ and can be adjusted for total surveillance time by dividing by ratio of the total surveillance times (active/placebo). Note that the total surveillance times use information on all subjects in each group. This transformation of $s/n_e$ can be applied to the confidence limits as well as the point estimate.

##### 8.4.3. Sensitivity: Cox Proportional Hazard

A sensitivity analysis to explore the robustness of the modified Poisson model will be conducted using a stratified Cox Proportional Hazard model with Efron's method of tie handling. Treatment group will be used as covariate. The one-sided p-value and 95% CI will be presented on the PP-EFF analysis.

Vaccine Efficacy will be defined as 1- HR where the HR (Hazard Ratio) is obtained using the Cox Proportional Hazard model.

Count and percentage (with 95% CIs based on the stratified Cox Proportional Hazard model) of first occurrence of PCR-Confirmed Mild, Moderate or Severe SARS-CoV-2 with an onset from at least 7 days after the second study vaccination regardless of symptoms will be presented by severity, treatment group and overall based on PP-EFF, similarly the first occurrence of symptomatic SARS-CoV-2 that is triggered by the symptomatic algorithm, as well as for moderate and severe categories for the second primary endpoint.

##### 8.4.4. Sensitivity: Tipping Point

A tipping point sensitivity analysis will be implemented for primary endpoint on the PP-EFF analysis set (Estimand 1a) to explore the influence of missing PCR data on the overall conclusion of the efficacy analysis results and specifically to find the "tipping" point . Conclusions will change from being favourable towards SARS-CoV-2 rS with Matrix-M1 adjuvant to being unfavourable i.e. Lowest Bound Confidence Interval (LBCI) falls below VE ≤ 30%. After such a tipping point is determined, clinical judgment can be applied as to the plausibility of the assumptions underlying this tipping point.

The tipping point analysis will follow the following strategy:

- Assuming all potential events in placebo group are classed as non-event
- All potential events in active group will be initially assumed as non-events (as per primary analysis). Further analyses will be performed incrementing the number of events until all potential events are classes as actual events. This will proceed until the tipping point is reached (LCBI ≤ 30%).

Results will be presented prior to and after the tipping point if there is a tipping point or else at the worst case and best case scenario for the active group.

Results will include VE adjusted for surveillance time with 95% Clopper Pearson CI (see Section 8.4.4) and p-value, with the number of subjects with an actual event, potential event and the number of potential events taken as events.

A subject with a potential event is defined as a subject who did have at least one illness episode meeting mild, moderate, or severe definition but with no PCR swab taken.

Instances of subjects with at least one PCR positive swab with no corresponding FluPRO or hospitalisation data within 7 days of the PCR positive swab will be explored further via the estimands targeting asymptomatic and symptomatic infections.

Note that all Per-Protocol analyses only include events with onset at least 7 days after second dose.

##### 8.4.5. Sensitivity: Cumulative Incidence Events

Cumulative event rates will be summarised for the ITT analysis set at 7, 14, 21, 28 days and 3,6,9,12 months after first vaccination for both treatment groups. A corresponding cumulative incidence curve will also be reported.

#### 8.5. Secondary Endpoints Efficacy Analysis

Secondary efficacy endpoints will be analysed based on the ITT analysis set using the same method as the primary efficacy endpoint as described in Section 8.1 and Section 8.4 without adjustment for multiple comparisons (i.e. two-sided alpha 0.05).

#### 8.6. Additional Efficacy Analyses

Estimation of estimands 1a, 1b, 2a and 2b and, associated subgroup analysis will be also conducted for event (see Sections 4.3 and 4.4.2) attributable to the UK variant of SARS-CoV-2.

Qualitative PCR tests will be summarised, by the number of swabs and tested positive at these timepoints: prior vaccination dose 1, prior vaccination dose 2 (Day 1- Day 21) and post-vaccination dose 2 (Day 22 +), who tested positive on the ITT Analysis Set, by overall and by site.

First occurrence of PCR-Confirmed COVID-19 Disease with onset from first vaccination, after dose 1, after dose 1 to before dose 2, dose 2 to 7 days after dose 2 and ≥7 days after dose 2 will have surveillance time summarised for both treatment groups on ITT Analysis Set.

#### 8.7. Immunogenicity Analysis

Blood will be collected from all subjects for humoral immunogenicity at Day 0, Day 35, Month 3, 6, and 12. The immunogenicity data will be analysed primarily using the PP-IMM analysis subsets, with supportive analyses performed using the ITT analysis subsets as specified in each section.

All the summaries will be conducted regardless of and stratified by age group (< 65 years or ≥ 65 years).

Serum IgG antibody levels specific for SARS-CoV-2 rS protein antigen and virus neutralisation assay specific for SARS-CoV-2 wildtype (or variant) will be additionally summarised for the the UK variant of SARS-CoV-2.

A visit-windowing approach may be adopted to account for samples obtained at both scheduled and unscheduled (e.g. unblinding) visit assessments.

##### 8.7.1. Serum IgG Antibody Levels Specific for SARS-CoV-2 rS Protein Antigen

The evaluations of the serum IgG antibodies specific for the SARS-CoV-2 protein antigen, i.e. anti-S protein as detected by ELISA, across study visits, will be performed based on PP-IMM and ITT anti-S protein serology subsets. For analysis performed on ITT anti-S protein serology subset, summaries will also be stratified by baseline serostatus (negative or positive).

All anti-S protein data will be listed for the ITT anti-S protein serology subset. Subjects included in the PP-IMM anti-S protein serology subset will be flagged in the listings.

- GMTs (reported in Geometric mean ELISA Units (GMEUs)) by treatment and overall with 95% CI. The 95% CI will be calculated based on the t-distribution of the log-transformed values for GMTs, then back transformed to the original scale (at baseline [Day 0] and at each post-vaccination visit). Plots of the reverse cumulative distribution curves also will be provided by treatment and overall.
- Geometric Mean Fold Rise (GMFR) (at each post-vaccination visit compared to baseline [Day 0]). The 95% CI will be calculated based on the t-distribution of the log-transformed fold-rise values for GMFRs, then back transformed to the original scale.
- Seroconversion Percentage (defined as percentage of subjects at each post vaccination visit with a titre ≥ 4-fold rise).naiive The corresponding two-sided exact binomial 95% CIs will be calculated by treatment group using the Clopper-Pearson method.
- For the calculation of strain-specific GMTs in each treatment group, titres reported below the lower limit of quantification (LLOQ; i.e., below the starting dilution of assay reported as "< 10") will be set to half that limit (i.e., 10 / 2 = 5).
- The GMT will be calculated using the following formula:

where $t_1, t_2, \ldots t_n$ are observed immunogenicity titres for n subjects.

The GMFR measures the changes in immunogenicity titres within subjects. The GMFR will be calculated using the following formula:

where, for n subjects, $v_{ij}$ and $v_{ik}$ are observed immunogenicity titres for subject i at time points j and k=0 (baseline [Day 0]).

Antibody titres will be summarised at baseline and each post-vaccination visit (the number of subjects with non-missing data, median, min, max, GMT and 95% CI). GMFR and the corresponding 95% CI for the GMFR will be presented by treatment group and by post-baseline visit.

Box plots of titre and fold rise by treatment group and visit will be provided. Figures based on ITT anti-S protein serology subset will be presented regardless of and stratified by baseline serostatus.

In addition, anti-N protein as detected by ELISA will be listed for ITT analysis set.

###### 8.7.2. Virus Neutralisation Assay Specific for SARS-CoV-2 Wildtype (or Variant)

An analysis similar to the serum IgG antibody levels described in Section 8.7.1 will be performed based on a neutralisation assay subset. All neutralisation assay data will also be listed for the ITT neutralisation assay subset (with a flag to identify subjects included in the PP-IMM subset).

##### 8.7.3. Cell-Mediated Response ± Intracellular Cytokine

Cell-mediated response (immunity) (counts per 1 Million Cells) for both Type 1 T Helper (Th1) and Type 2 T Helper (Th2) pathways will be assessed for each peptide pool by cytokine profiling and summarised by treatment group as mean, standard deviation (SD), median, min, max, and geometric mean. Change from baseline at Day 35 will also be summarised by mean, SD, median, min, and max GMFR at Day 35 will also be included. Summaries will be based on the PP-IMM and ITT cell-mediated assay subsets and will include the following cytokines: IFNg+, TNFa+, IFNg+TNFa+ (double positive), and IL-5+.

Box plots of cell-mediated response for cytokines IFNg+, TNFa+, IFNg+TNFa+ (double positive), and IL-5+ as by treatment group and visit will be provided. All cytokine data will also be presented in a data listing for the ITT cell-mediated assay subset.

#### 8.8. HAI Assay

For all subjects in the Seasonal Influenza Vaccine Sub-study, blood sampling for haemagglutination inhibition assay will be carried out on baseline [Day 0] and Day 21.

In addition to by treatment group summaries, treatment comparison will be made by comparing the strain-specific GMTs and the SCRs. The SCR for HAI assay is defined as the proportion of subjects with either a baseline reciprocal [Day 0] titre of < 10 and a post-vaccination reciprocal titre ≥ 40, or a baseline titre of ≥ 10 and a post-vaccination titre ≥ 4-fold higher.

For strain-specific GMTs in each treatment group, titres reported below the lower limit of quantitation (LLOQ; i.e., below the starting dilution of assay reported as "< 10") will be set to half that limit (i.e., 10 / 2 = 5). Strain-specific GMTs will be summarised by treatment group and visit day along with the corresponding two-sided 95% CIs, by exponentiating the corresponding log-transformed means and their 95% CIs. The ratio of GMTs between treatment groups post-vaccination and the corresponding two-sided 95% CI will be calculated on log-transformed titres using the analysis of covariance (ANCOVA) with treatment group and baseline [Day 0] measurement as the covariate.

Plots of the reverse cumulative distribution curves also will be provided by treatment and overall.

For strain-specific seroconversion, the rate in percentage and the corresponding two-sided exact binomial 95% CIs will be calculated using the Clopper-Pearson method. The two-sided 95% CIs for the absolute rate difference between the two treatment groups will be constructed using the Newcombe method.

HAI assays will be summarised for the PP-IMM and ITT-IMM seasonal influenza sub-study. All HAI assay data will be listed under the seasonal influenza vaccine sub-study.

##### 8.9. COVID Symptom Diary

COVID-19 symptom diary data will be captured in the FLU-PRO Plus questionnaire and FLU-PRO Plus Global Additional Daily Diary. This data will be listed only.

##### 8.10. Subgroup Analyses

The primary endpoint will be analysed by the following subgroups: age group (<65, ≥65 yrs), gender and, race on the PP-EFF Analysis Set. Unknown or not reported race would be omitted for subgroup analysis.

Stratification variables (region [pooled sites] and age group (<65/≥65 years) will be retained for subgroup analyses. Age group factor will be dropped for the age subgroup analysis. Note that strata will be corrected for mis-allocations at randomisation.

In the case the modified Poisson regression does not converge for a subgroup (e.g. zero counts in one treatment group), Clopper-Pearson method will be undertaken for that subgroup.

Additionally, analyses will be performed for estimands 1b, 2a and 2b in the age subgroups for PP-EFF Analysis Set.

#### 9. Safety Analysis

All safety summaries and analyses will be conducted for the Safety Analysis Set, except for the reactogenicity analyses that will be done on the Solicited AE Safety Subset Analysis Set.

The main presentations of data based on the Safety Analysis set will include all subject data, with supporting presentations to exclude the data post unblinding/post approved or deployed SARS-CoV-2 vaccine receipt (whichever comes first). Additional analysis of AEs collected after unblinding and after receipt of an approved and deployed SARS-CoV-2 vaccine receipt may be undertaken as exploratory safety analysis.

##### 9.1. Reactogenicity

Subjects in the Solicited AE Safety Subset Analysis Set will be issued with an electronic diary to collect solicited reactogenicity, to be recorded from the time of study vaccination until 7 days after study vaccination, for each dose. Solicited local and general systemic reactogenicity will be assessed for occurrence and intensity (toxicity grade) of selected signs and symptoms from the subject during a specific post-vaccination follow-up period (day of vaccination [Day 0] and 6 subsequent days), using a pre-defined checklist in their diary.

Participants in the licensed Seasonal Influenza Vaccine Sub-study will record local reactogenicity for the study vaccine injection site only. Toxicity grading will be standardised according to the FDA toxicity grading scale in Table 7 below. Any reactogenicity event extending beyond 7 days after vaccination (toxicity grade ≥1) will be recorded as an AE with a start date as date of vaccination + 7 days and followed to resolution.

The following local AEs (injection site: pain, tenderness, erythema/redness, and induration/swelling) and systemic AEs (fever, nausea/vomiting, headache, fatigue, malaise, myalgia, arthralgia) as displayed in Table 7 will be used.

**Table 7 FDA Toxicity Grading Scale for Clinical Abnormalities (Local and General Systemic Reactogenicity)**

| Clinical Abnormality | Mild (Grade 1) | Moderate (Grade 2) | Severe (Grade 3) | Potentially Life Threatening (Grade 4) |
|---|---|---|---|---|
| **Local Reaction to Injectable Product** | | | | |
| Pain | Does not interfere with activity | Repeated use of non-narcotic pain reliever >24 hours or interferes with activity. | Any use of narcotic pain reliever or prevents daily activity | ER visit or hospitalisation |
| Tenderness | Mild discomfort to touch | Discomfort with movement | Significant discomfort at rest | ER visit or hospitalisation |
| Erythema/redness | 2.5 − 5 cm | 5.1 − 10 cm | > 10 cm | Necrosis or exfoliative dermatitis |
| Induration/swelling | 2.5 − 5 cm and does not interfere with activity | 5.1 − 10 cm or interferes with activity | > 10 cm or prevents daily activity | Necrosis |
| **Systemic (General)** | | | | |
| Fever (°C)<br>(°F) | 38.0 − 38.4<br>100.4 − 101.1 | 38.5 − 38.9<br>101.2 − 102.0 | 39.0 − 40<br>102.1 − 104 | > 40<br>> 104 |
| Nausea/vomiting | No interference with activity or 1 − 2 episodes/24 hours | Some interference with activity or >2 episodes/24 hours | Prevents daily activity, or requires outpatient IV hydration | ER visit or hospitalisation for hypotensive shock |
| Headache | No interference with activity | Repeated use of nonnarcotic pain reliever >24 hours or some interference with activity | Significant; any use of narcotic pain reliever or prevents daily activity | ER visit or hospitalisation |
| Fatigue/Malaise | No interference with activity | Some interference with activity | Significant; prevents daily activity | ER visit or hospitalisation |
| Myalgia | No interference with activity | Some interference with activity | Significant; prevents daily activity | ER visit or hospitalisation |
| Arthralgia | No interference with activity | Some interference with activity | Significant; prevents daily activity | ER visit or hospitalisation |

The count and percentage (with 95% CIs based on the exact Clopper-Pearson method) of subjects who report each solicited AE immediately post study vaccination, on the day of vaccination (Day 0) and reported up to and including each of the 6 subsequent days and overall (i.e. Day 0 – 6, and Day 21 – 27 for the respective vaccine) will be summarised by maximum toxicity grade and treatment group for each vaccination dose. The 95% CI will be calculated based on subjects who report each solicited AE immediately post study vaccination and recorded a grade greater than 0. Percentages will be based upon the number of subjects in the Solicited AE Safety Subset Analysis Set within each vaccination group who reported data for the respective category, relative to the given vaccination dose. The summary will be presented for the Seasonal Influenza Vaccine Sub-study, separately for the subjects not included in the sub-study and overall.

The duration (in days) of solicited AEs after each vaccination will be summarised by treatment group for each vaccination for the Solicited AE Safety Subset Analysis Set, separately for those subjects in the Seasonal Influenza Vaccine Sub-study, those not included, and overall. Duration will be calculated as the number of days the solicited AE was greater than Grade 0 during the Day 0-6 assessment period plus any continuation beyond Day 6 post-vaccination, if recorded on the AE eCRF page.

All solicited local and systemic AEs will be presented in a data listing for the Solicited AE Safety Subset Analysis Set, separately for the subjects in the Seasonal Influenza Vaccine Sub-study and those not included. Additionally, a separate listing of solicited AEs that continued beyond 7 days after vaccination will be presented. A listing will be presented for treatment-related TEAEs.

##### 9.2. Unsolicited Adverse Events

An AE is defined as any untoward medical occurrence in a subject enrolled into this study regardless of its causal relationship to study vaccination.

A treatment-emergent AE (TEAE) is defined as any event not present before exposure to study vaccination or any event already present that worsens in intensity or frequency after exposure. Adverse events will also be evaluated by the investigator for the coexistence of MAAE which is defined as an AE that leads to an unscheduled visit to a healthcare practitioner.

Subjects will be assessed for diagnosis of an AESI at all study visits. AESI includes potential immune-mediated medical conditions (PIMMC) and AEs relevant to COVID-19.

AEs will be classified by system organ class (SOC) and preferred term (PT) using MedDRA (Version 23.1 or later). Only TEAEs will be included in summary tables and will be summarised by treatment group and vaccination (first dose, second dose, and overall) for the Safety Analysis Set. Summary tables for unsolicited AEs within the 21 days after first study vaccination and within the 28 days after second study vaccination will also be presented.

Percentages will be based upon the number of subjects in the Safety Analysis Set, unless otherwise stated. A subject may have more than 1 AE for an SOC or PT. A subject with 2 or more AEs within the same level of summarisation will be counted only once in that level.

All subjects will be assessed for unsolicited AEs from the time of first study vaccination until Day 49; SAEs will be assessed from the time the informed consent is signed until completion of the participant's last study-related procedure; all MAAEs will be reported from the time of first study vaccination until Day 35; MAAEs related to study vaccination and AESIs will be reported from the time of first study vaccination until completion of the participant's last study-related procedure. All AEs will be followed until resolution or until clinically stable.

If a solicited adverse event extended beyond 6 days following vaccination, then that event will be captured as an adverse event. The onset of that event will be noted to be Day 7 (i.e. the 7$^{th}$ day following study vaccination).

For the purpose of inclusion in TEAE tables, incomplete AE onset and end dates will be imputed according to the below rules (where UK, UKN, and UNKN indicate unknown or missing day, month, and year, respectively):

Missing Onset Dates

- UK-MMM-YYYY: Assume 01-MMM-YYYY, but if month and year are the same as the first study vaccination month and year, then assume the date of first vaccination
- UK-UKN-YYYY: Assume 01-JAN-YYYY, but if year is the same as the first study vaccination year, then assume the date of first study vaccination
- UK-UKN-UNKN: Assume date of first study vaccination

Note that start dates will only be imputed, if the imputed end date is missing or after the first vaccination date.

Missing End Dates

- UK-MMM-YYYY: Assume the last day of the month
- UK-UKN-YYYY: Assume 31-DEC-YYYY
- UK-UKN-UNKN: Do not impute and assume ongoing

All unsolicited AEs will be presented in a data listing.

##### 9.2.1. Incidence for Adverse Events

An overview of TEAEs will be presented by treatment group and overall, including number of TEAEs, count and percentage of subjects with any:

- TEAEs
- Severe TEAEs
- Treatment-related TEAEs
- Severe treatment-related TEAEs
- MAAEs
- Serious TEAEs
- TEAEs leading to vaccination discontinuation
- TEAEs leading to study discontinuation
- AESIs: PIMMC
- AESIs: relevant to COVID-19
- Serious Treatment-related MAAEs
- Treatment-related MAAEs
- Treatment-related TEAEs leading to vaccination discontinuation
- Treatment-related TEAEs leading to study discontinuation
- Treatment-related AESIs: PIMMC
- Treatment-related AESIs: relevant to COVID-19

An overall summary table of unsolicited AEs will be presented along with a further table that excludes events reported after unblinding. A summary of TEAEs will be presented by SOC and PT by treatment group and overall.

##### 9.2.2. Relationship of Adverse Events to Study Vaccine

The relationship or association of the study vaccine in causing or contributing to the AE will be characterised by the investigator as “Related” or “Not Related”. All TEAEs will be presented in a summary table for each treatment group and overall by SOC, PT, and relationship to study vaccine. If a subject has 2 or more TEAEs in the same SOC (or with the same PT) with a different relationship to study vaccine, then the subject will be counted under “Related”. If the relationship information is missing, the AE will be considered “Related” in the summary but will be presented as missing in the data listings.

A summary table for AEs with a start date up to an including 21 days after first study vaccination; and similarly for those unsolicited AEs with a start date up to and including 28 days after second study vaccination will also be presented by relationship to study vaccine. Additionally a summary table of treatment-related TEAEs only will be presented.

##### 9.2.3. Severity of Adverse Event

The severity (or intensity) of an AE refers to the extent to which it affects the subject’s daily activities and will be classified as mild, moderate, or severe. A summary of TEAEs and another for treatment-related TEAEs will be presented by each treatment group and overall by maximum severity, SOC, and PT. At each level of subject summarisation (SOC or PT),

a subject will be counted once at the maximal severity if the subject reported one or more events. If the severity information is missing, the AE will be considered severe in the summary but will be presented as missing in the data listings.
A summary table for AEs with a start date up to an including 21 days after first study vaccination; and similarly for those unsolicited AEs with a start date up to and including 28 days after second study vaccination will also be presented by severity.

###### 9.2.4. Serious Adverse Events

A summary table for all serious TEAEs and another for treatment-related serious TEAEs will be presented for each treatment group and overall by SOC and PT. All SAEs will be presented in a data listing.

###### 9.2.5. Medically-Attended Adverse Events

MAAEs are treatment-emergent medically-attended adverse events. All MAAEs, treatment-related MAAEs and all MAAEs by maximum severity will be presented in separate summary tables for each treatment group and overall by SOC and PT. A further table will be presented for all MAAEs with a start date up to an including 14 days after second vaccination. All MAAEs will be presented in a data listing.

###### 9.2.6. Adverse Events of Special Interest

An AESI is defined as follows:

- Treatment-emergent Potential Immune-Mediated Medical Conditions (PIMMC)
- Treatment-emergent Adverse Events of Special Interest Relevant to COVID-19

All treatment-related AESIs will be presented in a summary table for each treatment group and overall by SOC and PT. All treatment-emergent PIMMC will be presented in data listings. Similarly, treatment-emergent AESIs that are relevant to COVID-19 will be presented in data listings.

###### 9.2.7. Adverse Events Leading to Study Vaccine Discontinuation

All AEs leading to study vaccine discontinuation will be presented in a data listing.

###### 9.2.8. Adverse Events Leading to Discontinuation

All AEs leading to discontinuation from study will be presented in a data listing.

##### 9.3. Clinical Laboratory Evaluations

All pregnancy test results will be presented in a listing.

##### 9.4. Vital Sign Measurements

Vital sign measurements will be taken at Screening, Day 0 (pre-injection), Day 0 (15-30 minutes post-injection), Day 21 (pre-injection), Day 21 (15-30 minutes post-injection). The measurements respiratory rate, systolic/diastolic blood pressure, pulse rate, pulse oximetry and oral temperature (or via forehead/ear reader) will be recorded. Actual values and changes from baseline for vital sign data will be summarised by nominal visit, timepoint (predose and postdose) on vaccination days, and by treatment group on the Safety Analysis Set. All vital signs measurements will be presented in a data listing.

A summary of the toxicity grades (according to the FDA toxicity grading scale, See Appendix E) will be summarised by nominal visit, timepoint (predose and postdose) on vaccination days for each treatment group and overall using the count and percentage of subjects in each category.

##### 9.5. Physical Examination

Physical examination results will be summarised for each scheduled visit and examination by treatment group and overall for the Safety Analysis Set. Physical examination results for all subjects will be presented in a listing.

##### 9.6. Enhanced Emergency Room and/ or Hospitalisation Record

Enhanced Emergency Room and/ or Hospitalisation Records results for all subjects in the Safety Analysis Set will be presented in a listing.

#### 10. Interim Analyses

Safety data will be generated for two Safety Monitoring Committee (SMC) Day 7 reviews and four further SMC reviews thereafter, approximately every 3 months, when the last enrolled subject is projected to have completed 3, 6, 9 and 12 months of follow-up. Blinded outputs will be reviewed first, and when deemed necessary, unblinded tables by treatment group and unblinded subject-level listings can be reviewed to facilitate the decision. RR with 90% CI for severe COVID-19 will be generated approximately for SMC reviews at 3, 6, 9, and 12 months.

Prior to the final analysis, a single interim analysis of efficacy will be conducted based on the accumulation of approximately 50% (50 events) of the total anticipated target number of the primary endpoint (100 events). See Section 10.1 for more detail on how the primary endpoint events will be established for the interim analysis.

The interim analysis will follow standard group-sequential design using the Lan-DeMets alpha-spending function for Pocock boundary conditions. Appendix F summarises the timing, number of endpoints, and statistical success boundaries at the planned interim and final analyses.

If an unplanned additional interim analysis is to be added or the timing of a planned analysis is modified, the Lan-DeMets alpha-spending function will be used to adjust the nominal alphas to maintain the pre-specified overall one-sided type I error at 0.025.

#### 10.1. Blinded Review of Events

For the interim analysis of efficacy, data will be extracted from the clinical database (EDC) when there is confidence that a minimum of 50 PCR-confirmed mild, moderate or severe events have occurred.

The illness episode algorithm (See Appendix G) will be applied to this data extract, to generate a list of the subjects who have a PCR-confirmed mild/moderate/severe event. This list will be known as the *confirmed event list* and will clearly state the data extraction date and will be signed off by a member of the PPD and Novavax BIOS teams, prior to breaking the blind.

PPD DM and PPD clinical will work to clean the data for all subjects included on the *confirmed event list*. Once these data cleaning activities are complete, a further data extract will be taken. This later extract will be used to generate the interim analysis of efficacy deliverables.

#### 10.2. Study Unblinding Steps

The interim analysis will be performed by an unblinded Biostatistics and Programming team (PPD), and the unblinded statistician will communicate the results of the analyses to the Sponsor in terms of fulfillment or nonfulfillment of the predefined success criterion (yes/no).

The pre-defined success criterion requires that the lower limit of the alpha-adjusted confidence interval for vaccine efficacy of the primary efficacy endpoint (Estimand 1a) >30%. If the pre-defined success criterion of the interim analysis is unfulfilled (no), then the Sponsor will remain blinded to treatment assignment until the final analysis. If the pre-defined success criterion of the interim analysis is fulfilled (yes), then the Sponsor will sign an Unblinding Authorisation Form and subsequently receive selected accrued unblinded data at the treatment group level (see Section 10.3) and continue the study while maintaining the blind to achieve a more robust safety and efficacy data package.

The unblinded analyses will be conducted in a separated area and the unblinded team will remain isolated from the study personnel and Sponsor. They will complete a review independent of the study team and Sponsor.

Summaries produced for the interim analyses will not unblind at subject level and only information at treatment level will be presented.

#### 10.3. Interim Analysis Reporting

If the results of the interim analysis for efficacy fulfil the predefined success criterion, then the following outputs will be provided to the Sponsor per the process described in Section 10.2:

- Poisson regression analysis of the primary efficacy endpoint (Estimand 1a). Note: if insufficient events occur to enable Poisson model convergence an alternative analysis will be presented based on Clopper Pearson adjusted for surveillance time (see Section 8.4.2)

- Supporting summary table for the primary efficacy endpoint (Estimand 1a)
- Overall Summary of Unsolicited Adverse Events
- Overall Summary of Unsolicited Adverse Events (excluding events reported post unblinding).

Of note, for the primary efficacy endpoint (Estimand 1a), subjects will only be classified as an event if they were included on the *confirmed event list* (generated based on the first data extract and subjected to focused data cleaning activities) and if they still meet the event criteria per the later, interim analysis data extract. This means that subjects on the *confirmed event list* may be downgraded to a non-event (owing to data cleaning outcomes); however, all subjects who were not included on the *confirmed event list* will be considered a non-event, regardless of whether they meet the event criteria per the later, interim analysis data extract (because the subset of subjects who met the primary endpoint definition only in the later data extract will not be considered sufficiently clean).

Among the subjects classified as an event for the interim analysis, the onset date of the associated illness episodes will be ordered chronologically and the date of the latest of these will be established. This will be used as the *cut-off* date for the purposes of deriving surveillance time, for the primary efficacy endpoint (Estimand 1a).

#### 11. Final Analysis and EUA

For the final analysis of efficacy, data will be extracted from the clinical database (EDC) when there is confidence that a minimum of 100 PCR-confirmed mild, moderate or severe events have occurred (per primary estimand definition, 1a).

The illness episode algorithm (see Appendix G) will be applied to this data extract, to generate a list of the subjects who have a PCR-confirmed mild, moderate or severe event. Provided that this list includes the minimum required number of required events, the date of the associated data extract will be known as the Final Analysis Trigger date.

As part the Data Delivery Plan, PPD DM and PPD Clinical will ensure that those CRF data that are used by the illness episode algorithm are 100% cleaned and Source Data Verified through to the Final Analysis Trigger date, for all subjects with a positive PCR result (post-Day 0) per this initial data extract.

Once these data cleaning activities are complete, a further data extract will be taken. This later extract will be used to generate the Final Analysis and EUA deliverables.

For the Final Analysis and EUA, subjects will only be classified as an event if they meet the event criteria per the later data extract and if they were subjected to the focused data cleaning activities described above (i.e. had a positive PCR result (post- Day 0) on the earlier data extract).

The Final Analysis Trigger Date, will be used as the 'cut-off date' for the purposes of deriving surveillance times.

Note: subjects who were not part of the focused data cleaning activities and who have any newly appearing PCR positive results (on the Final analysis/EUA data extract) which are dated prior to the Final Analysis Trigger Date will be considered a non-event (as they will not be considered sufficiently clean).

#### 12. Changes from Protocol

- Section 7.2 of study protocol asserts that the interim analysis will be based on a 'locked database'. This is an error - the database will not be locked for the interim analysis. Data used for the interim analysis will be cleaned per the detail provided in Section 10.1.
- The protocol defines the PP-EFF as follows:

  *The PP-EFF will include baseline seronegative participants who receive both doses of study vaccine (SARS-CoV-2 rS with Matrix-M1 adjuvant or placebo) and have no major protocol deviations that occur before the first COVID-19 episode affecting the primary efficacy outcome (i.e., participants will be censored at the time of the protocol deviation) as assessed by the sponsor prior to unblinding. All analyses of the PP-EFF population will also exclude any illness episodes with positive SARS-CoV-2 by any validated PCR or antibody test occurring 6 days or less after the second study vaccination (e.g., Day 28*).

  However, it must be read that the PP-EFF population will also exclude *any subjects with an illness episodes* occurring 6 days or less after second study vaccination (rather than exclusion of episodes alone).

  This SAP also further qualifies that anti-s antibody test results will NOT be used to determine PP-EFF eligibility. This decision was made because anti-s testing is performed for only a subset of subjects (<10%).
- The Protocol states that unblinding will result in censoring from reactogenicity analyses and analyses of unsolicited AEs. The SAP clarifies that this will be achieved via sensitivity analyses.
- The protocol assumes that all subjects who received approved or deployed SARS-CoV-2 vaccine will be unblinded prior to administration. The SAP acknowledges that this is not always the case, and includes details on the handling of such scenarios (i.e. censoring at the earliest of unblinding and receipt of approved or deployed SARS-CoV-2 vaccine date).

#### 13. References

Zou G. A modified poisson regression approach to prospective studies with binary data. Am J Epidemiol. 2004 Apr 1;159(7):702-6. doi: 10.1093/aje/kwh090. PMID: 15033648.

### Appendices

#### Appendix A Schedule of Events

| **Study Period:** | **Screening Period [a]** | **Clinic Visits** | | | | | **Months After Last Study Vaccination** | | |
|---|---|---|---|---|---|---|---|---|---|
| **Study Day:** | **–30 to 0** | **0[a]** | **21** | **35** | **COVID-19 Surveillance Visits (Unscheduled)[c]** | **Unblinding Visit[d]** | **3** | **6** | **12** |
| **Window (days):[b]** | **–** | **0** | **+ 7** | **+ 7** | | | **± 15** | **± 15** | **± 15** |
| **Minimum days following most recent study vaccination:[b]** | **–** | **0** | **21** | **14** | | | **–** | **–** | **–** |
| **Study Visit:** | **Screening** | **1** | **2** | **3** | | | **4** | **5** | **EOS[e]** |
| Informed consent | X | | | | | | | | |
| Medical history [f] | X | | | | X | | | | |
| Inclusion/exclusion criteria [g] | X | X [h] | X [h] | | | | | | |
| Demographics [i] | X | | | | | | | | |
| Prior/concomitant medications [j] | X | X [h] | X [h] | X | X | X | X | X | X |
| Vital sign measurements [k] | X | X | X | | X | | | | |
| Urine pregnancy test (WOCBP) [l] | X | X [h] | X [h] | | | | | | |
| Physical examination (targeted) [m] | X | X [h] | X [h] | X | X | | | | |
| Nose/throat testing for SARS-CoV-2 (PCR) [n] | X | X [h] | X [h] | | X | | | | |
| Blood sampling for SARS-CoV-2 (ELISA for anti N-protein serology) | | X [h] | | X | | X[d] | X | X | X |
| Blood sampling for SARS-CoV-2 (ELISA for anti S-protein serology – subset of participants) [o] | | X [h] | | X | | X[d] | | | |
| Blood sampling for SARS-CoV-2 neutralisation assay (subset) [p] | | X [h] | | X | | X[d] | | | |
| Blood sampling for HAI (influenza co-administration subset) [q] | | X [h] | X | | | X[d] | | | |
| Cell-mediated assessments (subset of participants) [r] | | X [h] | | X | | | | | |
| Randomisation | | X | | | | | | | |
| Study vaccination [s] | | X | X | | | | | | |
| Reactogenicity (subset of participants) [t] | | X | X | | | | | | |
| Monitoring for COVID-19 [u] | | | COVID-19 case ascertainment will commence from Day 0 until EOS [v] | | | | | | |
| COVID-19 Symptom Diary [w] | | | | | X | | | | |
| All unsolicited AEs [x] | | X | X | X | | | | | |
| MAAEs [y] | | X | X | X | X | X | X | X | X |

| Study Period: | Screening Period [a] | Clinic Visits | | | | | Months After Last Study Vaccination | | |
|---|---|---|---|---|---|---|---|---|---|
| Study Day: | –30 to 0 | 0[a] | 21 | 35 | COVID-19 Surveillance Visits (Unscheduled)[c] | Unblinding Visit[d] | 3 | 6 | 12 |
| Window (days):[b] | – | 0 | + 7 | + 7 | | | ± 15 | ± 15 | ± 15 |
| Minimum days following most recent study vaccination:[b] | – | 0 | 21 | 14 | | | – | – | – |
| Study Visit: | Screening | 1 | 2 | 3 | | | 4 | 5 | EOS[e] |
| SAEs [z] | X | X | X | X | X | X | X | X | X |
| AESI [aa] | | X | X | X | X | X | X | X | X |
| EOS form [bb] | | | | | | | | | X |

Abbreviations: AE = adverse event; AESI = adverse event(s) of special interest; COVID-19 = coronavirus disease 2019; ELISA = enzyme-linked immunosorbent assay; ELISpot = enzyme-linked immune sorbent spot; EOS = end of study; HAI = hemagglutinin assay inhibition; HEENT = head, eye, ear, nose, and throat (exam); ID = identification; MAAE = medically attended adverse event; N = nucleocapsid; PCR = polymerase chain reaction; PIMMC = potential immune-mediated medical conditions; S = spike protein; SAE = serious adverse event; SAP = statistical analysis plan; SARS-CoV-2 = severe acute respiratory syndrome coronavirus 2; WOCBA = women of childbearing age.

a) The Screening visit and Day 0 visit may be combined if feasible at any given study site.
b) Days relative to study vaccination are only estimates because the window allowance is not inclusive. Should a study pause occur, visits/windows will be adjusted to allow participants to continue without protocol deviation. Visit schedules after second study vaccination are calculated relative to the day the study vaccinations were received.
c) If the participant is known to be COVID-19 positive at the time of scheduling for the Initial Surveillance Visit and cannot attend the visit due to extenuating circumstances (e.g., local government restrictions), then a phone call or phone calls can be substituted to assess the participant's status. However, the participant should make every effort to attend the Follow-up Surveillance Visit (as per protocol timeline) when government restrictions or other circumstances allow. If the participant has made an Initial Surveillance phone contact or visit and is found to be COVID-19 negative on all swabs taken for this episode, then a phone call can be substituted for the Follow-up Surveillance Visit.
d) An Unscheduled Unblinding Visit should be arranged (where possible and practical to do so) for those who are offered and are willing to accept an approved or deployed SARS-CoV-2 approved or deployed vaccine. Serology will be obtained as per Section 6.1.6. Visits on Days 21 and 35 may be skipped for those who have been unblinded as per Section 6.1.6.
e) EOS visit. Should participants decide to terminate early, an EOS telephone call will occur to collect the maximum safety data possible.
f) Including prior and concomitant medical conditions, recent vaccinations (≤ 90 days), and significant surgical procedures.
g) Specific exclusions to study vaccination will be assessed before any vaccination. Waivers to enrolling participants with exclusions will not be given.
h) Performed prior to study vaccination.
i) Screening only. Including date of birth (day, month, and year), sex, race, ethnicity, weight, and height.
j) Recent and current medications at the time of screening to be reviewed to ensure eligibility criteria are fulfilled. Concomitant medications include all medications (including vaccines) taken by the participant.
k) Including respiratory rate, blood pressure, pulse rate, pulse oximetry and oral temperature (or via forehead/ear reader). On study vaccination days, vital sign measurements will be collected once before study vaccination to ensure participant has no evidence of fever prior to study vaccination. Vital sign measurements will also be collected once again, approximately 15 to 30 minutes after study vaccination, to check for any reactions to the vaccine.
l) Women of childbearing potential only. A urine pregnancy test will be performed at Screening and prior to each study vaccination. A positive urine pregnancy test at any time will result in the participant not receiving any further study vaccination. A positive urine pregnancy test at Screening will result in screen failure.
m) Examination at screening to include height and weight; HEENT, neck, thyroid, lungs, heart, cardiovascular, abdomen, lymph nodes of the upper extremities and neck; and musculoskeletal system/extremities to allow for study vaccination; symptom-directed (targeted) physical examination at all other scheduled time points but always to include lymphatic assessment of injected upper extremity on study vaccination days. Physical examination on study vaccination visits must be done prior to vaccination. Interim physical examinations will be performed at any unscheduled visit at the discretion of the investigator, if necessary.
n) Samples will be collected at Screening only if the participant has any COVID-19 symptoms or significant exposure history. If a participant has a positive PCR prior to enrolment, they will be considered a screen failure. Samples will be collected on Day 0 and the method of collection will be taught. Participants found to be SARS-CoV-2 positive on Day 0 will be excluded from some analyses of the study as per the SAP. Samples may be collected on Day 21 only if the participant has any COVID-19 symptoms or significant exposure history

| Study Period: | Screening Period [a] | Clinic Visits | | | | Unblinding Visit[d] | Months After Last Study Vaccination | | |
|---|---|---|---|---|---|---|---|---|---|
| Study Day: | –30 to 0 | 0[a] | 21 | 35 | COVID-19 Surveillance Visits (Unscheduled)[c] | | 3 | 6 | 12 |
| Window (days):[b] | – | 0 | + 7 | + 7 | | | ± 15 | ± 15 | ± 15 |
| Minimum days following most recent study vaccination:[b] | – | 0 | 21 | 14 | | | – | – | – |
| Study Visit: | Screening | 1 | 2 | 3 | | | 4 | 5 | EOS[e] |

between Days 0 and 21. Results of that test are not required for study vaccination, but participants who are symptomatic may have the second study vaccination delayed per protocol. Participants found to be SARS-CoV-2 positive on Day 21 will be excluded from some analyses of the study as per the SAP.

o) The ELISA for anti-S protein serology will be performed in the approximately 900 participants in the Anti-S Protein Serology Subset.
p) The neutralising antibody assay will be performed in the approximately 900 participants in the Neutralisation Assay Subset.
q) The HAI assay will be performed in the approximately 400 participants in the licensed seasonal influenza co-administration sub-study.
r) Cell-mediated immune responses, as assessed by ELISpot ± intracellular cytokine staining, will be performed in approximately 450 participants in the Cell-mediated Assay Subset.
s) Study vaccination on Day 0 will consist of study vaccine plus, in the seasonal influenza vaccine co-administration sub-study, a single dose of licensed influenza vaccine. Study vaccination on Day 21 will consist of study vaccine.
t) Solicited AEs (e.g., local and general systemic reactogenicity symptoms) will be captured in the Safety Subset of approximately 2,000 participants and in the first approximately 400 participants who qualify for the seasonal influenza vaccine co-administration sub-study. On study vaccination days, participants will remain in clinic for at least 30 minutes to be monitored for any severe reactogenicity. Severe reactions will be noted as AEs on day of study vaccination.
u) Participants will monitor for COVID-19 symptoms. This will include provision of a Study ID Card that provides details on study participation, study site contact information and assessment of symptoms of suspected COVID-19 (see Table 2-2). Instructions to participants to contact the study site within 24 hours for symptoms of suspected COVID-19 will be given. A participant with suspected or confirmed COVID-19 will be asked to complete a COVID-19 symptom diary for 10 days. The Study ID Card should be presented to healthcare providers not affiliated with the study who encounter participants with symptoms of suspected COVID-19.
v) Samples will be self-collected by the participants in an effort to determine if the current symptoms are due to SARS-CoV-2 infection. Approximately 24 hours after the onset of symptoms the participants will swab themselves daily for up to 3 days but only until the point that 1 of the samples tests positive for SARS-CoV-2.
w) A participant with suspected or confirmed COVID-19 will be asked to complete the COVID-19 symptom diary. Participants should start the diary on their first day of symptoms and complete this diary daily for a minimum of 10 days even if their symptoms have resolved and regardless of any SARS-CoV-2 PCR results. They should continue the diary beyond 10 days if they have any ongoing symptoms related to the episode and should continue to do so until all symptoms have resolved.
x) All unsolicited AEs are to be reported from the time of first study vaccination until 21 days after first study vaccination and 28 days after second study vaccination in all participants.
y) MAAEs are to be collected from the time of first study vaccination until Day 35, and MAAEs assessed as related to study vaccination from the time of first study vaccination until completion of the participant's last study-related procedure.
z) SAEs are to be collected from the time of informed consent until completion of the participant's last study-related procedure.
aa) AESI: To include PIMMC and AEs related to COVID-19 are to be collected from the time of first study vaccination until completion of the participant's last study-related procedure.
bb) EOS form will be completed for all participants, including participants who are terminated early.

### Appendix B Power Under Various Vaccine Efficacy Assumptions

| Assumed Vaccine Efficacy | Estimated Power | | |
|---|---|---|---|
| Symptomatic COVID-19 Illness PCR-Confirmed SARS-CoV-2 Infection | At Planned Interim Analysis with 50 Events | At Final Analysis with 100 Events | Overall (At Interim Analysis or Final Analysis) |
| 60% | 29% | 39% | 68% |
| 65% | 45% | 41% | 87% |
| 70% | 64% | 32% | 96% |
| 75% | 81% | 18% | >99% |
| 80% | 94% | 6% | >99% |
| 85% | 99% | 1% | >99% |
| 90% | >99% | <10% | >99% |

Abbreviations: COVID-19 = coronavirus disease 2019; PCR = polymerase chain reaction; SARS-CoV-2 = severe acute respiratory syndrome coronavirus 2

#### Appendix C Severity of COVID-19 Symptoms

| COVID-19 Severity | Endpoint Definitions |
|---|---|
| Mild | ≥ 1 of:<br>• Fever (defined by subjective or objective measure, regardless of use of anti-pyretic medications)<br>• New onset cough<br>• ≥ 2 COVID-19 respiratory/non-respiratory symptoms in Appendix D<br>AND<br>• Does not meet criteria for moderate or severe disease |
| Moderate | ≥ 1 of:<br>• Fever (defined by subjective or objective measure, regardless of use of anti-pyretic medications) + any 2 COVID-19 symptoms in Appendix D for ≥ 3 days (need not be contiguous days)<br>• High fever (≥ 38.4°C) for ≥ 3 days (need not be contiguous days)<br>• Any evidence of significant LRTI:<br>− Shortness of breath (or breathlessness or difficulty breathing) with or without exertion (greater than baseline)<br>− Tachypnea: 20 to 29 breaths per minute at rest<br>− SpO2: 94% to 95% on room air<br>− Abnormal chest x-ray or chest CT consistent with pneumonia or LRTI<br>− Adventitious sounds on lung auscultation (e.g., crackles/rales, wheeze, rhonchi, pleural rub, stridor)<br>**AND**<br>• Does not meet criteria for severe disease |

| **COVID-19 Severity** | **Endpoint Definitions** |
|---|---|
| Severe | ≥ 1 of:<br>• Tachypnea: ≥ 30 breaths per minute at rest<br>• Resting heart rate ≥ 125 beats per minute<br>• SpO2: ≤ 93% on room air or PAO2/FiO2 < 300<br>• High flow oxygen therapy or NIV/NIPPV (e.g., CPAP or BiPAP)<br>• Mechanical ventilation or ECMO<br>• One or more major organ system dysfunction or failure (e.g., cardiac/circulatory, pulmonary, renal, hepatic, and/or neurological, to be defined by diagnostic testing/clinical syndrome/interventions), including any of the following:<br>o ARDS<br>o Acute renal failure<br>o Acute hepatic failure<br>o Acute right or left heart failure<br>o Septic or cardiogenic shock (with shock defined as SBP < 90 mm Hg OR DBP < 60 mm Hg<br>o Acute stroke (ischemic or hemorrhagic)<br>o Acute thrombotic event: AMI, DVT, PE<br>o Requirement for: vasopressors, systemic corticosteroids, or hemodialysis.<br>• Admission to an ICU<br>• Death |

Abbreviations: AMI = acute myocardial infarction; ARDS = acute respiratory distress syndrome; BiPAP = bi-level positive airway pressure; CPAP = continuous positive air pressure; CT = computed tomography; DBP = diastolic blood pressure; DVT = deep vein thrombosis; ECMO = extracorporeal membrane oxygenation; FiO2 = fraction of inspired oxygen; ICU = intensive care unit; LRTI = lower respiratory tract infection; NIV = non-invasive ventilation; NIPPV = non-invasive positive pressure ventilation; PAO2 = partial pressure of oxygen in the alveolus; PE = pulmonary embolism; SBP = systolic blood pressure; SpO2 = oxygen saturation.

### Appendix D Qualifying Symptoms of Suspected COVID-19

| **Respiratory Symptoms** |
|---|
| Fever |
| New onset cough |
| New onset or worsening of shortness of breath or difficulty breathing compared to baseline |
| New onset fatigue |
| New onset generalised muscle or body aches |
| New onset headache lasting ≥ 48 hours |
| New loss of taste or smell |
| Acute onset of sore throat, congestion, and runny nose |
| New onset nausea, vomiting, or diarrhea lasting ≥ 48 hours |

Abbreviations: COVID-19 = coronavirus disease 2019.

### Appendix E FDA Toxicity Grades Scale for Clinical Abnormalities (Vital Signs)

| Vital Signs | Mild (Grade 1) | Moderate (Grade 2) | Severe (Grade 3) | Potentially Life Threatening (Grade 4) |
|---|---|---|---|---|
| Tachycardia (bpm) | 101-115 | 116-130 | >30 | ER visit or hospitalisation for arrhythmia |
| Bradycardia (bpm) | 50-54 | 45-49 | <45 | ER visit or hospitalisation for arrhythmia |
| Hypertension (systolic) (mm Hg) | 141-150 | 151-155 | >155 | ER visit or hospitalisation for arrhythmia |
| Hypertension (diastolic) (mm Hg) | 91-95 | 96-100 | >100 | ER visit or hospitalisation for arrhythmia |
| Hypotension (systolic) (mm Hg) | 85-89 | 80-84 | <80 | ER visit or hospitalisation for shock |
| Respiratory Rate (breaths per minute) | 17-20 | 21-25 | >25 | Intubation |

Abbreviations: DHHS = Department of Health and Human Services; ER = emergency room; FDA = United States Food and Drug Administration.
Note: Participant should be at rest for all vital sign measurements, with toxicity scored on day of study vaccination (pre- and post-dose).
a When resting heart rate is between 60 – 100 bpm. Use clinical judgement when characterising bradycardia among some healthy participant populations (e.g., conditioned athletes).

### Appendix F Interim and Final Boundaries Using Pocock Spending Function

| Planned Information Fraction (% of total endpoints) | Planned Blinded Total Number of Endpoints | Planned One-Sided Nominal Alpha | VE Boundary for LBCI > 30% |
|---|---|---|---|
| Interim analysis at 50% | 50 | 0.01550 | ~68% |
| Final analysis at 100% | 100 | 0.01387 | ~57% |

Abbreviations: LBCI = lower bound confidence interval; VE = vaccine efficacy.

#### Appendix G Algorithm for Determining Illness Episode

Note: This algorithm only applies to subjects who have received at least one dose of study drug.

Definitions:

Illness episode – Range of dates, inclusive, from when a subject first reports the onset of COVID-19 symptoms (irrespective of whether they meet the criteria for mild, moderate or severe COVID-19 disease) to the day before when the subject reports that the symptoms are back to normal (or if date of return to normal is missing, then the last day subject reports a symptom).

Infection episode – Range of dates, inclusive defined by a series of PCR+ swabs with details given below in Section I Step 6,

I Steps for subjects with evidence of suspected COVID-19 symptoms (as defined in Table 2-2 of the protocol):

1 Determine start and stop dates of each subject's illness episode.

a Source CRFs are FLU-PRO, COVID-19 New Symptoms Intake Evaluation, COVID-19 Surveillance PHONE Contact (Initial & Follow-up), COVID-19 Surveillance Visits (Initial & Follow-up), Vital Signs (temperature ≥ 37.8°C), and Enhanced ER/Hospitalization.

b If there is at least a 7-day period free of symptoms, split into separate illness episodes.

2 After establishing Step 1 illness episode start and stop dates, the illness episode may be extended using the following symptoms if present no more than 7 days after Step 1 illness episode stop date:

a Moderate or Severe Pulse Oximetry (as defined in Table 2-1 of the protocol); if more than 1 occurs on the same day, select the higher pulse oximetry.

b Severe Heart Rate (as defined in Table 2-1 of the protocol); if more than 1 occurs on the same day, select the lower heart rate.

c Moderate or Severe Respiratory Rate (as defined in Table 2-1 of the protocol); if more than 1 occurs on the same day, select the lower respiratory rate.

d Adventitious sounds on lung auscultation (eg, crackles/rales, wheeze, rhonchi, pleural rub, stridor)

3 If the illness episode corresponds with a negative PCR swab test result, then the illness episode will be the dates determined in Steps 1 & 2.

4 If the illness episode corresponds with at least 1 PCR+ swab test result from Day 0 onward, then the illness episode may be further extended using Steps 5-7 below.

5 Flag date(s) from Day 0 onward where the subject has a PCR+ swab test result.

6 Determine start and stop dates of each infection episode based on PCR+ swab test results found in Step 5.

  a If there is only 1 PCR+ swab test result, the start and stop dates of the infection episode = date of the PCR+ swab test result.

  b If there are 2 PCR+ swab test results and they occur <=35 days apart, then these should be combined into 1 infection episode (i.e. start date = date of PCR+ swab test #1, end date = date of PCR+ swab test #2); otherwise, leave as 2 individual infection episodes (i.e. for 2 PCR+ swab test results occurring >35 days apart, start and stop dates for each episode = date of each PCR+ swab test).

  c If there are 3 or more PCR+ swab test results, apply Step 3b pairwise, starting with the 2 PCR+ swab test results that are closest to each other for that subject.

II Steps for any PCR+ swab test results that do not fall within an illness episode:

1 Use the Enhanced ER/Hospitalization CRFs to see if at least 1 of the following occurs on the same day as the PCR swab or no more than 7 days afterwards:

  a Any evidence of significant LRTI

  b Except for Death (Considered in Section IV), any Severe COVID-19 endpoint (as defined in Table 2-1 of the protocol); if more than 1 occurs on the same day, select the less severe result (i.e. lower heart rate, lower respiratory rate, higher pulse oximetry).

2 If Step 1 is met, set the start and stop dates of the illness episode as the hospital admission and discharge dates (respectively). If the discharge date is missing and the Adverse Events CRF page confirms that the subject was hospitalized due to an event related to COVID-19, then set the discharge date as the AE end date. If the discharge date is missing and there is no evidence of hospitalization due to a COVID-19-related event, then this is considered to be a 1-day event (i.e. discharge date = admission date).

III Steps for determining if illness episodes from Sections I and II should be combined:

1 If the number of days (based on the difference between start date of episode and stop date of previous episode) between the illness episodes determined in Sections I and II is more than 7 days, then the 2 illness episodes remain separate.

2. If the number of days between the illness episodes determined in Sections I and II is <=7 days, then the 2 illness episodes should be combined; set the start date as the earliest of the Section I or II illness episode start date and set the stop date as the latest of the Section I or II illness episode end date.
3. Extend illness episode start and stop dates by factoring in infection episode start and stop dates.
    a. If an illness episode has no dates in common with an infection episode, then the illness episode remains unchanged.
    b. If an illness episode overlaps by at least 1 day with an infection episode, or if an infection episode occurs <=7 days before an illness episode, or if an illness episode occurs <=7 days before an infection episode, then the final illness episode start and stop dates are the minimum of the start dates from Sections I/II/III and the maximum of the stop dates from Sections I/II/III, respectively. The start and stop dates of the combined illness episode will only use start and stop dates of the illness episodes to be merged; infectious episode start and stop dates are used to verify if the event is PCR-confirmed.

IV Special consideration for COVID-19 related deaths:

1. Obtain the COVID-19 related death date from the Adverse Events CRF.
2. Extend the illness episode from Section III by setting the stop date as the date of the COVID-related death
3. If the COVID-19 related death can only be linked to a PCR+ swab test result, then the illness episode start date is the PCR+ swab test result date and the illness episode end date is the COVID-19 related death date.

#### Appendix H Pooling of Site

| Location | Site # | Region |
|---|---|---|
| London | UK001 | England South East |
| Corby | UK005 | England Other |
| London | UK006 | England South East |
| Aberdeen | UK007 | Scotland, Wales, Northern Ireland |
| Glasgow | UK008 | Scotland, Wales, Northern Ireland |
| Stockport | UK009 | England Other |
| Blackpool | UK010 | England Other |
| Belfast | UK011 | Scotland, Wales, Northern Ireland |
| London | UK012 | England South East |
| Exeter | UK013 | England Other |
| Bournemouth | UK014 | England Other |
| Norwich | UK015 | England Other |
| Oxford | UK016 | England South East |
| Stoke-on-trent | UK017 | England Other |
| Bradford | UK018 | England Other |
| Leeds | UK019 | England Other |
| London | UK020 | England South East |
| Hartlepool | UK021 | England Other |
| Lancashire | UK022 | England Other |
| Hexham | UK023 | England Other |
| Midlands | UK024 | England Other |
| Wales | UK025 | Scotland, Wales, Northern Ireland |
| Merseyside | UK026 | England Other |
| Wrexham | UK027 | Scotland, Wales, Northern Ireland |
| Kent | UK028 | England South East |
| Lancaster | UK029 | England Other |
| Salford | UK030 | England Other |
| Thames Valley | UK031 | England South East |
| Manchester | UK032 | England Other |
| Glasgow | UK033 | Scotland, Wales, Northern Ireland |
| Ipswich & Colchester | UK034 | England Other |
| Warnsford | UK035 | England South East |
| Cornwall | UK036 | England Other |

| Table of Amendments to the Statistical Analysis Plan | | |
|---|---|---|
| **Version** | **Date** | **Summary of Changes** |
| 1.0 | 21Jan2021 | First version |
| 2.0 | 15Feb2021 | • Incorporation of changes introduced by protocol version 3.0 other than those relevant to the interim analysis (addressed in v1.0)<br>• Additional condition for excluding subjects from PP-EFF and PP-IMM who received both doses of study vaccine (SARS-CoV-2 rS with Matrix-M1 adjuvant or placebo) more than 45 days apart<br>• Estimand 6 extended to analyse endpoint during a surveillance period from i) 7, ii) 10 in addition to iii) 14 days after first vaccination<br>• Further consideration for impact of individual unblinding on efficacy and immugenicity analyses<br>• Appendix G -- definition of the start and stop dates of the illness episode when consideration for COVID-19 related deaths<br>• Subject co-morbidity status (Yes/No) as an additional demographic characteristic |
| 2.1 | 19Feb2021 | • Clarification of the Baseline [Day 0] notion when appropriate<br>• Section 4.4.5 -- 4$^{th}$ bullet point -- Precision that post-baseline serum samples must be assessed in the specified visit window from second study vaccine<br>• Extension of the definition of the duration of solicited AEs |
| 3.0 | 22Feb2021 | • Added provision for subjects who receive approved or deployed SARS-CoV-2 vaccine prior to, or without unblinding. Such subjects should be handled in the same way as subjects who are unblinded owing to the offer of approved or deployed SARS-CoV-2 vaccine (i.e. censored/excluded from date of receipt of the deployed SARS-CoV-2 vaccine).<br>• Remove sentence regarding sensitivity analysis of reactogenicity and unsolicited AEs conducted for the Safety Analysis subset of subjects not unblinded as this is not planned. |
| 4.0 | 05Mar2021 | • In section 12, acknowledgment of any corrections done in protocol version 4.0 that were highlighted in the previous version of the SAP.<br>• Update of the last bullet point in Per-Protocol Immunogenicity Analysis Set (PP-IMM) definition to *not* plan to censor data after death or end of study, both events censoring de facto the subject' data.<br>• Precision for concomitant medications and adverse events that start dates are imputed if the imputed end date is missing or after first vaccination date.<br>• Clarification of the use of Clopper (vs Poisson) in sections 8.4.2 and 8.10.<br>• In section 4.5.1, additional consideration for screen failures.<br>• Precision primarily in section 4.4.1 that events with onset on the censoring date are still considered as an event (i.e. not censored).<br>• Additional considerations for mis-randomisation (e.g. section 4.5.4). |
